# Dopaminergic Medication Accentuates Fecal Gut Microbiome Changes in Parkinson’s Disease

**DOI:** 10.1101/2022.12.23.22283907

**Authors:** Jeffrey M Boertien, Pedro AB Pereira, Pia Laine, Lars Paulin, Sygrid van der Zee, Petri Auvinen, Filip Scheperjans, Teus van Laar

## Abstract

Fecal gut microbiota changes are associated with Parkinson’s disease (PD). However, disease related changes cannot readily be discerned from medication effects, as almost all participants in previous studies were using PD medication, and conclusive longitudinal data related to treatment initiation is lacking. Here, fecal gut microbiota composition was assessed in 62 *de novo* PD participants who were untreated at baseline and used PD medication at one-year follow-up, by means of 16S-sequencing. In addition, participants were stratified for the type of dopaminergic medication.

Overall gut microbiota composition did not differ between baseline and one-year follow-up, but was associated with levodopa dose and levodopa equivalent daily dose (LEDD). Several differentially abundant taxa are in line with previously described changes in PD. These included reduced levels of amplicon sequence variants (ASVs) belonging to *Faecalibacterium prausnitzii* and Lachnospiraceae in all participants at follow-up, and increased levels of an ASV belonging to *Bifidobacterium* in dopamine agonist users. The family Bifidobacteriaceae was increased in dopamine agonist users who only used pramipexole. Levodopa dose was inversely related to the abundance of the families Ruminococcaceae and Lachnospiraceae, and the genus *Lachnospiraceae ND3007 group*. PD medications exert a measurable and dose-dependent effect on gut microbiota composition and accentuate several previously described gut microbiota changes in PD. Detailed knowledge of medication effects should be part of future trial designs of gut microbiome studies in PD and are necessary to interpret previously published data.

## Introduction

Parkinson’s disease (PD) is characterized by alpha-synuclein (aSyn) aggregation and subsequent Lewy body pathology, which is hypothesized to spread along neuro-anatomical connections in a prion-like manner.^1,2^ Although the clinical diagnosis of PD largely depends on its cardinal motor symptoms,^3^ non-motor symptomatology often precedes the clinical diagnosis by several years.^4^ In particular, constipation is recognized as one of the earliest prodromal symptoms of PD.^5^ In concordance, aSyn pathology has been found in the enteric nervous system of prodromal PD subjects and autonomic denervation along the vagal nerve is described in a subgroup of PD subjects before and at the time of diagnosis.^6^ This suggests PD pathology might spread along the vagal nerve in a so-called body-first subtype with a possible gastrointestinal origin of the disease.^7,8^

Gut microbiota might contribute to PD pathology through reduced production of anti-inflammatory short chain fatty acids (SCFA), increased mucin degradation, and cross-seeding of aSyn through bacterial amyloid proteins.^9–12^ Several studies have found differences in gut microbiome composition between PD and healthy controls (HC).^13–16^ Among the most replicated findings are increased relative abundances of the genera *Bifidobacterium, Lactobacillus* and *Akkermansia*, whereas *Faecalibacterium, Prevotella* and several genera belonging to the family Lachnospiraceae tend to be decreased. However, most studies interrogated the gut microbiome of PD subjects using dopaminergic medication.^13^ Consequently, the effects of PD and PD medication on gut microbiome composition cannot adequately be disentangled.

Recently, we published the results of two independent case-control cohorts of treatment-naïve *de novo* PD subjects, with taxonomic differences mainly limited to reduced SCFA-producing bacteria, in particular including taxa belonging to the family of Lachnospiraceae.^17^ Similarly, a study with a subpopulation of untreated PD subjects concluded that most differences in gut microbiome composition between PD and HC disappeared after statistical adjustment for treatment and disease duration.^18^ The untreated subpopulation was still characterized by lower levels of the family Lachnospiraceae and two of its genera. Moreover, a mendelian randomization study suggests that increased levels of *Bifidobacterium* are due to reverse causation.^19^ Also, several taxa have been correlated to levodopa dosage, including *Bifidobacterium, Lactobacillus, Faecalibacterium* and several taxa belonging to the family of Lachnospiraceae.^14,20^ Therefore, several gut microbiome changes in PD might be attributable to the use of dopaminergic medication.

The relevance of the relationship between PD medication and gut microbiota extends beyond the identification of gut microbiota that might contribute to PD pathology. Taxa like *Enterococcus faecalis* are known to decarboxylate levodopa to dopamine in the intestine, limiting bioavailability in the central nervous system, thereby contributing to dose-response fluctuations.^21–23^ In addition, probiotics supplementation containing strains belonging to *Bifidobacterium* or *Lactobacillus* relieves gastro-intestinal discomfort in PD, whereas the relative abundance of these genera is increased in PD.^24,25^ Disentangling the relationship between PD medication and *Bifidobacterium* and *Lactobacillus* can add to their safety profile for treating gastro-intestinal discomfort, as a counterproductive effect on disease progression becomes more unlikely.

One study interrogated the gut microbiome of nineteen PD subjects before and 90 days after levodopa initiation.^26^ A decrease in *Clostridium group IV* was observed in those with a good response to therapy. Since PD medication is not known to have bacteriostatic or bactericidal properties, their putative influence might be exerted over longer periods of exposure through other selective pressures such as medication metabolism. Four longitudinal studies investigated the gut microbiome of already treated PD subjects at baseline and after one to two years.^27–30^ Gut microbiome composition apparently remained stable in studies using 16S rRNA gene sequencing, whereas total fecal bacterial counts were decreased after two years in one study using quantitative reverse-transciption polymerase chain reaction (qRT-PCR) of selected taxa. Therefore, an exposure period of more than 90 days, but less than two years, seems appropriate to investigate the effect of PD medication on gut microbiome composition. Here, the gut microbiome of 62 *de novo* PD subjects was investigated at baseline and after one year, during which treatment with dopaminergic medication was initiated.

## Results

### Clinical characteristics

Participants of the Dutch Parkinson Cohort of de novo PD subjects (DUPARC, NCT04180865), who did not use PD medication at baseline, but did use PD medication at one year follow-up, were included.^31^ Participants with major confounders of gut microbiome composition were excluded (e.g., antibiotics usage within one month before samples collection, Supplementary Table 1). 62 participants could be included with a successfully sequenced stool sample at baseline and follow-up. On average, participants collected their follow-up stool sample 55 weeks after the baseline stool sample. No differences between the two visits were found in non-motor symptoms measured by NMSQ, MoCA, stool-frequency and stool consistency. A lower burden of motor-symptoms was observed at follow-up (MDS-UPDRS-III, Hoehn and Yahr), indicating a clinically measurable effect of the dopaminergic medication. The vast majority of participants used levodopa at follow-up (87%), with only eight participants (13%) using only a dopamine agonist. No COMT-inhibitors, MAO-B-inhibitors, advanced therapies or amantadine were used. The mean LEDD was 466mg, suggesting that a large proportion of participants had at least one increase in the dose of dopaminergic medication, which typically starts at 50 or 100mg three times daily. An overview of the clinical characteristics is provided in table 1.

**Table 1.** Cohort characteristics

|  | Baseline |  | Follow-up | p-value |
| --- | --- | --- | --- | --- |
|  | n | Descriptive statistics | Descriptive statistics |  |
| Weeks since baseline, mean ±sd | 60 | - | 55.14 ±5.55 | - |
| Number of reads, mean ±sd | 62 | 42058 ±9295 | 40577 ±11040 | 0.43 |
| Age, mean ±sd | 62 | 66.81 ±8.16 | 67.89 ±8.19 | - |
| Female, n(%) | 62 | 21 (33.9%) | 21 (33.9%) | 1 |
| BMI, mean ±sd | 58 | 26.18 ±4.60 | 26.28 ±4.62 | 0.53 |
| Stool consistency, mean ±sd | 61 | 3.36 ±0.95 | 3.47±1.11 | 0.44 |
| Stool frequency, mean ±sd | 61 | 1.19 ±0.59 | 1.20 ±0.65 | 0.83 |
| MDS-UPDRSIII total, mean ±sd | 62 | 31.96 ±11.91 | 26.77 ±11.69 | **6.99E-5** |
| Motor subtype, n IND-PIGD-TD (%) | 62 | 10-35-17<br>(16.1%-56.5%-27.4%) | 15-25-22<br>(24.2%-40.3%-35.5%) | 8.54E-2 |
| Hoehn and Yahr, median [IQR] | 62 | 2 [2-2] | 2 [1-2] | **7.38E-3** |
| MoCA, median [IQR] | 61 | 25 [23-27] | 26 [24-27] | 0.80 |
| NMSQ total, median [IQR] | 58 | 7 [4-8] | 6.5 [4-8.75] | 0.30 |
| Users of PD medication, n (%) |  |  |  |  |
| Levodopa | 62 | 0 (0%) | 54 (87.1%) | - |
| Carbidopa | 62 | 0 (0%) | 40 (64.5%) | - |
| Benserazide | 62 | 0 (0%) | 15 (24.2%) | - |
| Levodopa CR | 62 | 0 (0%) | 10 (16.1%) | - |
| Ropinirole | 62 | 0 (0%) | 5 (8.1%) | - |
| Pramipexole | 62 | 0 (0%) | 8 (12.9%) | - |
| Rotigotine | 62 | 0 (0%) | 4 (6.5%) | - |
| COMT-inhibitors | 62 | 0 (0%) | 0 (0%) | - |
| MAO-B inhibitors | 62 | 0 (0%) | 0 (0%) | - |
| Amantadine | 62 | 0 (0%) | 0 (0%) | - |
| Advanced therapies | 62 | 0 (0%) | 0 (0%) | - |
| Only levodopa | 62 | 0 (0%) | 45 (72.6%) |  |
| Only dopamine agonist | 62 | 0 (0%) | 8 (12.9%) |  |
| LEDD, mean ±sd | 62 | - | 465.97 ±238.86 | - |

BMI, body mass index; COMT, catechol-O-methyltransferase; CR, controlled-release; IND, indeterminate subtype; IQR, interquartile range; LEDD, levodopa equivalent daily dose; MAO-B, monoamineoxidase-B; MDS-UPDRS, Movement Disorders Society Unified Parkinson's Disease Rating Scale; MoCA, Montreal Cognitive Assessment; n, number of participants with available data, missing data was deleted in a pairwise manner to ensure complete paired baseline and follow-up data; NMSQ, Non-Motor Symptom Questionnaire; PD, Parkinson's disease; PIGD, postural instability and gait disorder subtype; p-value, nominal p-value calculated by paired t-test or Wilcoxon for parametric and non-parametric continuous variables respectively, calculated by McNemar's test for categorical variables; sd, standard deviation; TD, tremor dominant subtype

### Variable selection

All clinical and technical variables were screened for their potential effect on the difference between baseline and follow-up visits using a PERMANOVA in the following model:

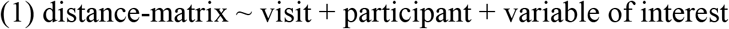

Variables that shifted the explained variance of the visit variable (R^2^) the most compared to its explained variance without the variable (distance-matrix ∼ visit + participant), were added to the list of relevant model covariates (Supplementary Table 2). Variables with a generalized variance inflation factor (GVIF) ≥5 were excluded to avoid (multi)collinearity (Supplementary Table 3). This resulted in the following model for the multivariate analysis of overall gut microbiome composition in all participants:

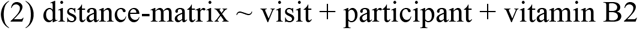

For the differential abundance analysis, variables with a significant relationship (p < 0.1) in the final model of overall gut microbiome composition would have been selected. However, no variables had a p-value below the selection threshold.

### Sequencing

The total library size contained 5,123,375 sequences, with a similar mean library size of 42,058 reads at baseline and 40,577 reads at follow-up (*p*=0.43, Table 1). The low number of sequences of the blank control samples at the DNA extraction and PCR steps, indicate very minimal to no contamination during the wet lab work (Supplementary Table 4).

### Overall microbiome composition

Alpha diversity (i.e. within-sample diversity) did not differ between timepoints for any of the four investigated indices: Chao1, Shannon, Inverse Simpson and observed richness (Figure 1). Multivariate analyses of overall community structure were performed on Aitchinson distances and Bray-Curtis dissimilarities.^32^ Statistical significance was calculated using a PERMANOVA, with adjustment for selected variables using marginal testing.^33^ The multivariate analysis of between-sample diversity in overall gut microbiota composition showed no statistically significant differences between baseline and follow-up based on either Aitchinson distances or Bray-Curtis dissimilarities (*p=*0.068 and *p=*0.19, respectively, Figure 1, Table 2). Also, when stratifying for dopaminergic monotherapy, no differences were found between baseline and follow-up (Table 2).

**Figure 1.**
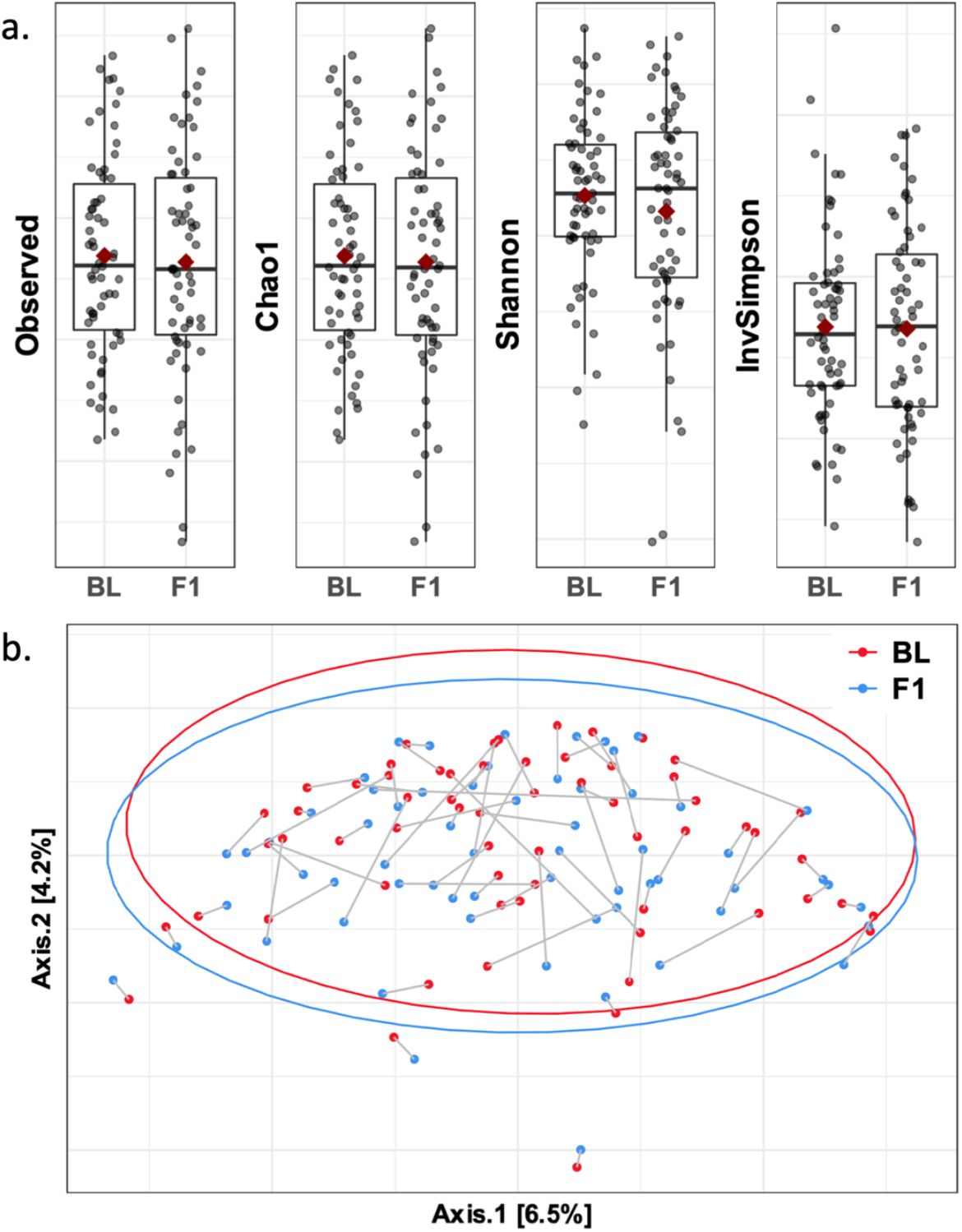
Differences in alpha-diversity (intra-sample) and overall microbiome composition (inter-sample) between baseline (n=62) and follow-up (n=62) samples. Each point represents one sample. (a) All four alpha-diversity indices showed no statistically significant differences beteen baseline and follow-up samples (Wilcoxon test, paired, univariable: p =0.57, 0.59, 0.46 and 0.74 for observed richness, Chao1, Shannon and Inverse Simpson, respectively). Boxplots represent the first quartile, median and third quartile at the lower, middle and upper boundaries, with the mean represented by the red diamond. The whiskers represent 1.5 times the interquartile range. (b) Principal component plot of Aitchinson distances of overall microbiome composition showing a large overlap between baseline (red) and follow-up (blue) samples (PERMANOVA adjusted for participant, to create a paired analysis: p = 0.068). Gray lines connect the baseline and follow-up samples of the same participant. BL, baseline samples; F1, one-year follow-up samples.

**Table 2.** Comparison of gut microbial community structures at baseline and follow-up

|  | Aitchinson distance |  | Bray-Curtis dissimilarity |  |
| --- | --- | --- | --- | --- |
|  | % variance | p-value | % variance | p-value |
| **a. Paired analysis all participants** |  |  |  |  |
|  | *n*=124 |  | *n*=114 |  |
| Visit | 0.41 | 0.068 | 0.39 | 0.19 |
| Participant | 78.7 | <1E-04 | 81.1 | <1E-04 |
| **b. Paired analysis all participants, adjusted for selected variables** |  |  |  |  |
|  | *n*=106 |  | *n*=98 |  |
| Visit | 0.45 | 0.62 | 0.40 | 0.43 |
| Participant | 73.4 | <1E-04 | 80.0 | <1E-04 |
| Vitamin B2 | 0.66 | 0.15 | 0.21 | 0.999 |
| **c. Paired analysis stratified for levodopa usage** |  |  |  |  |
|  | *n*=90 |  | *n*=82 |  |
| Visit | 0.53 | 0.20 | 0.56 | 0.12 |
| Participant | 78.6 | <1E-04 | 81.4 | <1E-04 |
| **d. Paired analysis stratified for dopamine agonist usage** |  |  |  |  |
|  | *n*=16 |  | *n*=14 |  |
| Visit | 2.71 | 0.57 | 3.84 | 0.27 |
| Participant | 77.1 | <1E-04 | 77.2 | <1E-04 |
| **e. Paired analysis stratified for pramipexole usage** |  |  |  |  |
|  | *n*=10 |  | *n*=10 |  |
| Visit | 5.18 | 0.43 | 6.31 | 0.21 |
| Participant | 74.8 | 6.0E-04 | 75.0 | 3.0E-04 |
| **f. Paired analysis stratified for ropinirole usage** |  |  |  |  |
|  | *n*=6 |  | *n*=4 |  |
| Visit | 8.60 | 0.67 | 16.2 | 0.67 |
| Participant | 72.8 | <1E-04 | 67.0 | 0.33 |

Multivariate analysis of differences in gut microbial community structures between baseline and follow-up samples. Six different analyses were performed: a. a paired comparison in all participants, b. a paired comparison adjusted for selected variables, and four paired comparisons in participants using only c. levodopa, d. a dopamine agonist, e. pramipexole, f. ropinirole as dopaminergic medication. Paired analyses were performed through adjusting for participant as pairing variable. All models were assessed using both Aitchinson distances and Bray-Curtis dissimilarities. Removal of samples due to missing data and/or rarefaction was done in a pair-wise manner for the paired analyses, to ensure a balanced design with a baseline and follow-up sample of each remaining participant. Differences were assessed for statistical significance with a PERMANOVA based on 10,000 permutations, setting the lowest p-value at 9.999E-05 (<1E-04), with the exception of f. (ropinirole stratification) due to low sample size, resulting in 719 and 23 permutations for Aitchinson distances and Bray-Curtis dissimilarities, respectively. Multivariable models were assessed using marginal testing, meaning each variable was adjusted for all other variables in the model.
% variance, percentage explained variance ( $R^2$ ); n, number of samples

### Differential abundance

Differential abundance analyses were performed at the ASV, genus and family level with ANCOM (v2.1) and DESeq2 (v1.30.1).^34–36^ Although several differentially abundant taxa were identified with statistical significance using DESeq2 and ANCOM, no taxa were identified as differentially abundant by both DESeq2 and ANCOM (Table 3). Two taxa belonging to the family Lachnospiraceae were reduced at follow-up in the analysis of all participants, including the genus *Hungatella*. In addition, an ASV belonging to the species *Faecalibacterium prausnitzii* was reduced (Table 3, Figure 2, Supplementary Table 5).

**Table 3.**
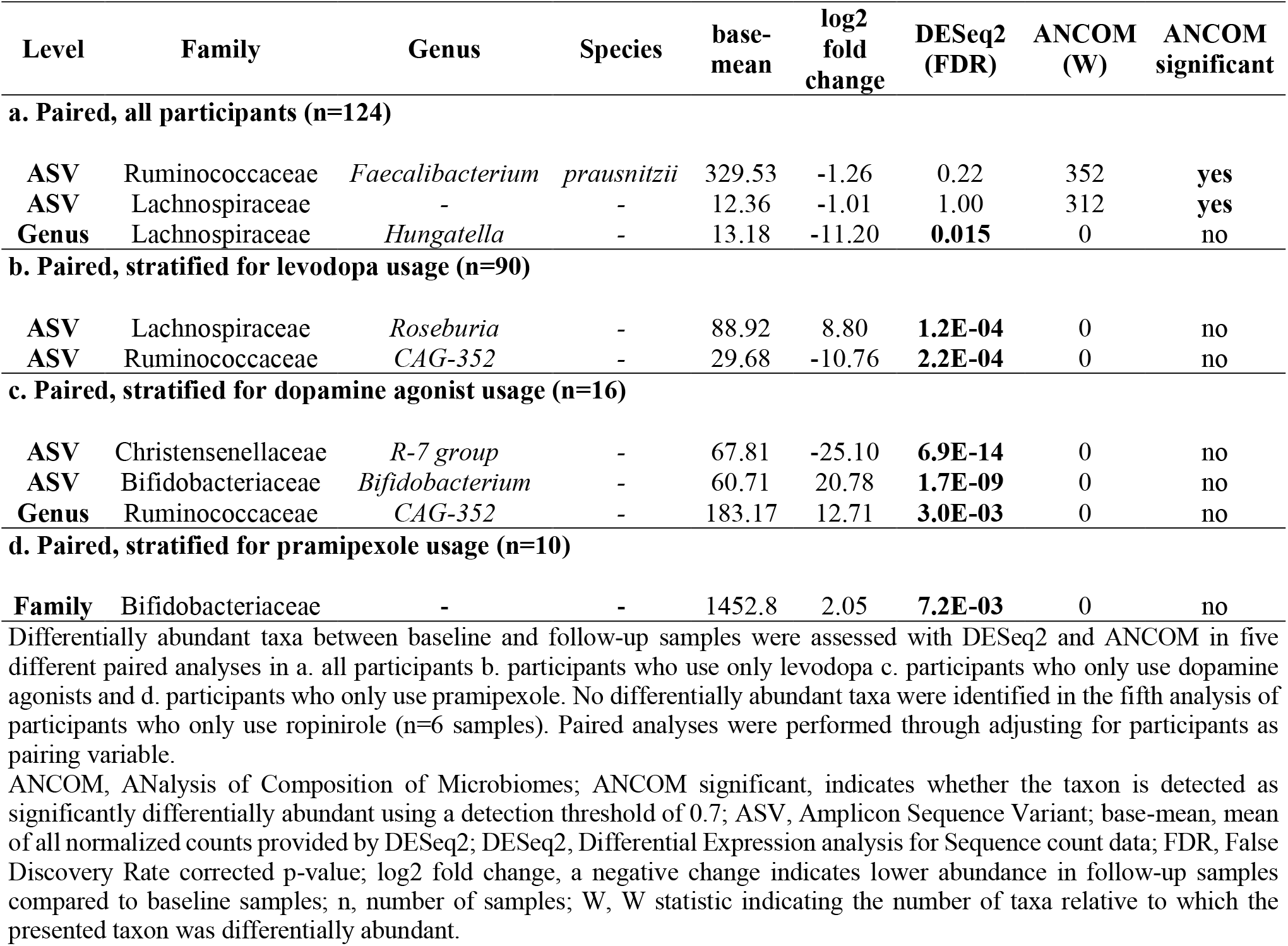
Differentially abundant taxa between baseline and follow-up samples

| Level | Family | Genus | Species | base-mean | log2 fold change | DESeq2 (FDR) | ANCOM (W) | ANCOM significant |
| --- | --- | --- | --- | --- | --- | --- | --- | --- |
| **a. Paired, all participants (n=124)** |  |  |  |  |  |  |  |  |
| ASV | Ruminococcaceae | *Faecalibacterium* | *prausnitzii* | 329.53 | -1.26 | 0.22 | 352 | yes |
| ASV | Lachnospiraceae | - | - | 12.36 | -1.01 | 1.00 | 312 | yes |
| Genus | Lachnospiraceae | *Hungatella* | - | 13.18 | -11.20 | **0.015** | 0 | no |
| **b. Paired, stratified for levodopa usage (n=90)** |  |  |  |  |  |  |  |  |
| ASV | Lachnospiraceae | *Roseburia* | - | 88.92 | 8.80 | **1.2E-04** | 0 | no |
| ASV | Ruminococcaceae | *CAG-352* | - | 29.68 | -10.76 | **2.2E-04** | 0 | no |
| **c. Paired, stratified for dopamine agonist usage (n=16)** |  |  |  |  |  |  |  |  |
| ASV | Christensenellaceae | *R-7 group* | - | 67.81 | -25.10 | **6.9E-14** | 0 | no |
| ASV | Bifidobacteriaceae | *Bifidobacterium* | - | 60.71 | 20.78 | **1.7E-09** | 0 | no |
| Genus | Ruminococcaceae | *CAG-352* | - | 183.17 | 12.71 | **3.0E-03** | 0 | no |
| **d. Paired, stratified for pramipexole usage (n=10)** |  |  |  |  |  |  |  |  |
| Family | Bifidobacteriaceae | - | - | 1452.8 | 2.05 | **7.2E-03** | 0 | no |

Differentially abundant taxa between baseline and follow-up samples were assessed with DESeq2 and ANCOM in five different paired analyses in a. all participants b. participants who use only levodopa c. participants who only use dopamine agonists and d. participants who only use pramipexole. No differentially abundant taxa were identified in the fifth analysis of participants who only use ropinirole (n=6 samples). Paired analyses were performed through adjusting for participants as pairing variable.
ANCOM, ANALysis of Composition of Microbiomes; ANCOM significant, indicates whether the taxon is detected as significantly differentially abundant using a detection threshold of 0.7; ASV, Amplicon Sequence Variant; base-mean, mean of all normalized counts provided by DESeq2; DESeq2, Differential Expression analysis for Sequence count data; FDR, False Discovery Rate corrected p-value; log2 fold change, a negative change indicates lower abundance in follow-up samples compared to baseline samples; n, number of samples; W, W statistic indicating the number of taxa relative to which the presented taxon was differentially abundant.

**Figure 2.**
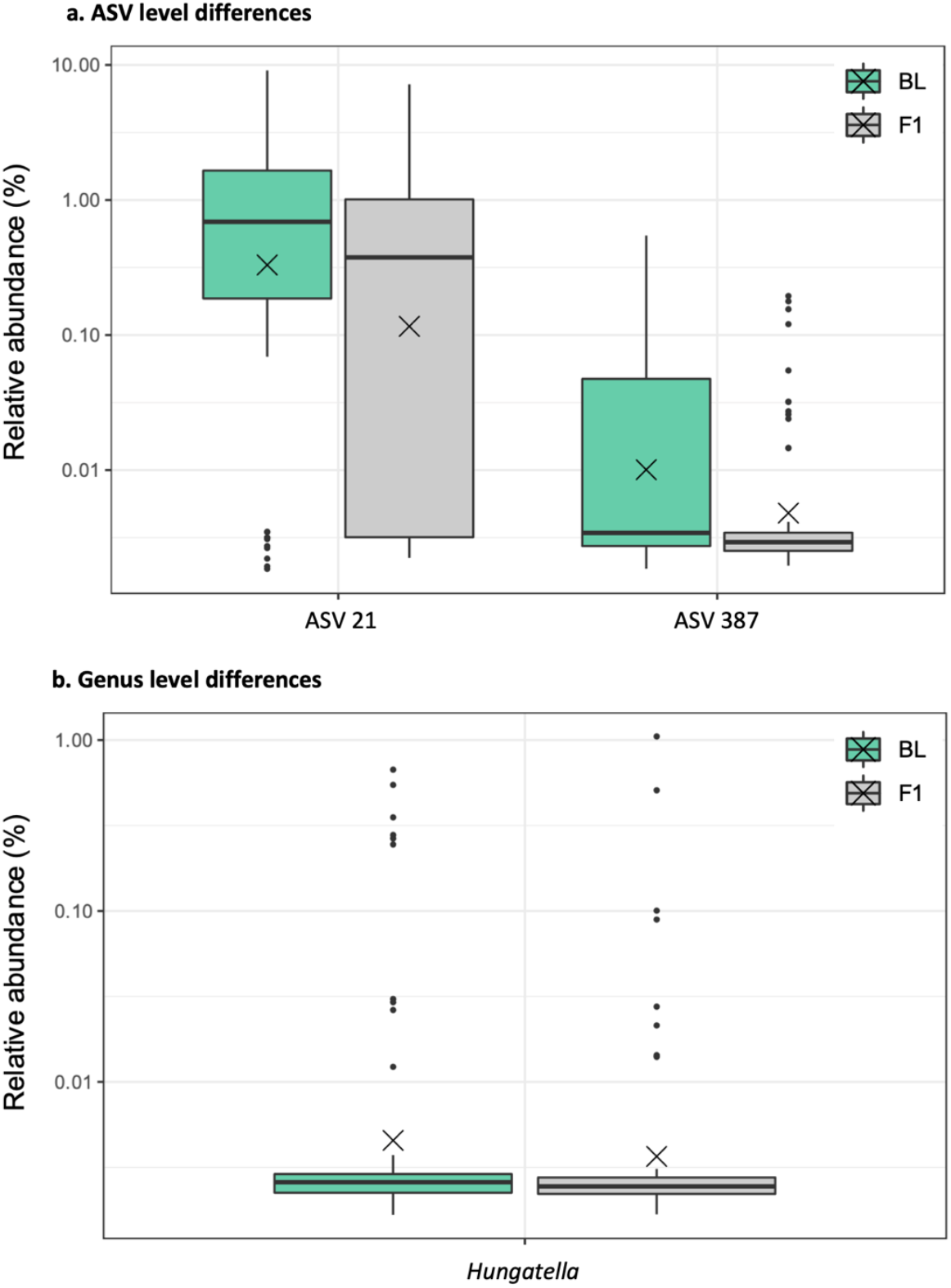
Relative abundances of taxa identified as differentially abundant with DESeq and/or ANCOM in all samples (n=124). Analyses were paired by adjusting for participants as the pairing variable. (a) Two ASVs belonging to the species *Faecalibacterium prausnitzii* and family Lachnospiraceae were reduced at follow-up in the paired analysis. (b) The genera Lachnospiraceae *Hungatella* was reduced at follow-up in the paired analysis. Each box represents the first quartile, median and third quartile at the lower, middle and upper boundaries, with the whiskers representing points within 1.5 the interquartile range and the X representing the mean. Relative abundances are presented on a log10 scale. ANCOM, ANalysis of Composition of Microbiomes; ASV, Amplicon Sequence Variant; BL, baseline samples; DESeq, Differential Expression analysis for Sequence count data; F1, follow-up samples one year after baseline.

Complementary to the analysis of all participants, participants were stratified for dopaminergic monotherapy. In levodopa users, ASVs belonging to the genera Lachnospiraceae *Roseburia* and Ruminococcaceae *CAG-352* were respectively increased and reduced at follow-up (Table 3, Figure 3, Supplementary Table 6). In contrast the genus Ruminococcaceae *CAG-352* was increased at follow-up in dopamine agonist users. One ASV belonging to the genus Christensenellaceae *R-7 group* was reduced in dopamine agonist users, whereas an ASV belonging to the genus *Bifidobacterium* was increased at follow-up. When stratifying only for pramipexole usage, the entire family Bifidobacteriaceae was increased at follow-up (Table 3, Figure 2, Supplementary Table 7).

**Figure 3.**
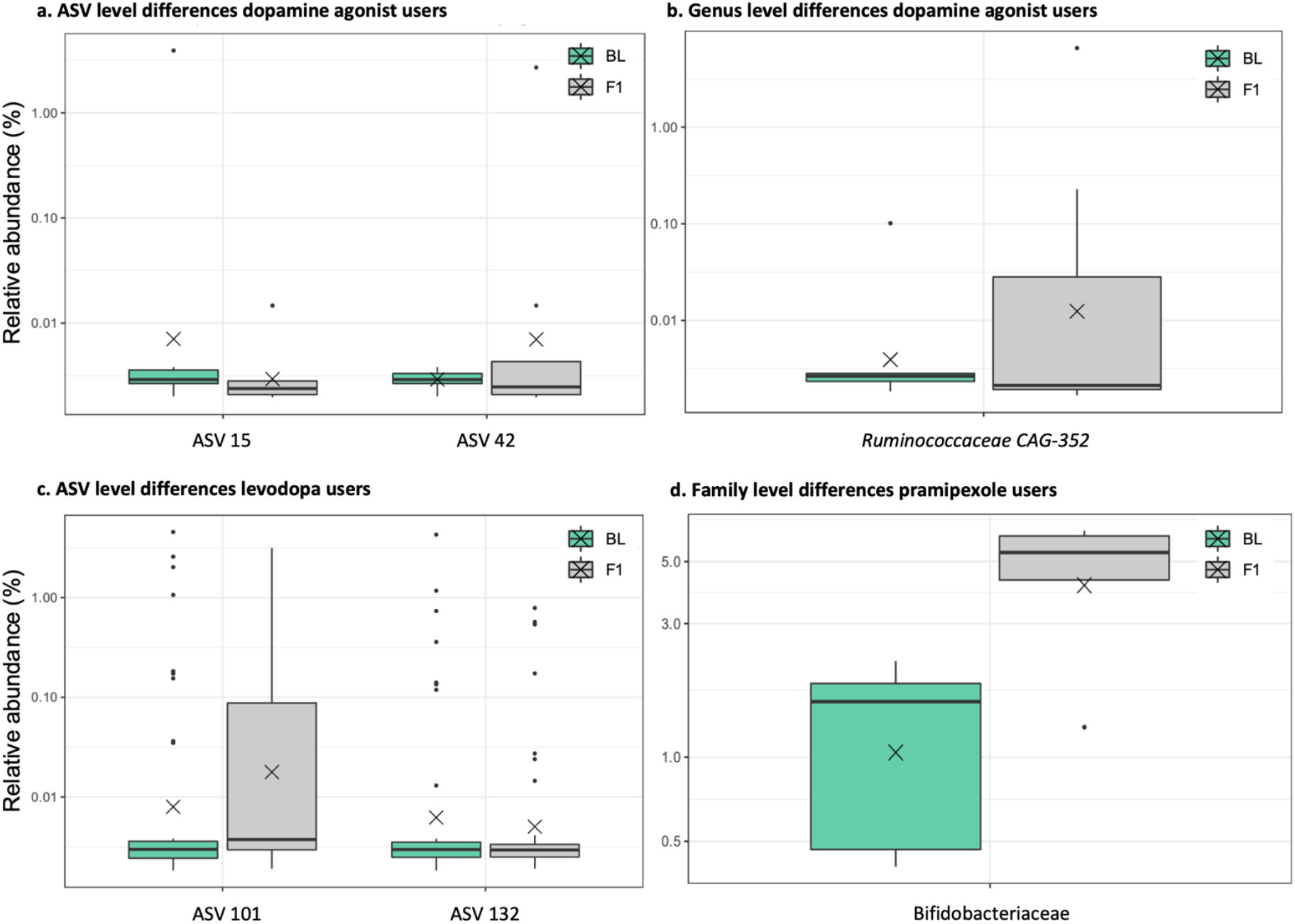
Relative abundances of taxa identified as differentially abundant with DESeq and/or ANCOM in samples stratified for dopaminergic monotherapy at follow-up. Analyses were paired by adjusting for participants as pairing variable. (a) At ASV-level, two ASVs belonging to the genera Christensenelleceae *R-7 group* and Bifidobacteriaceae *Bifidobacterium* were respectively decreased and increased in dopamine agonist users at follow-up (n=16 samples). (b) At genus level, Ruminococcaceae *CAG-352* was increased in dopamine agonists users at follow-up. (c) Two ASVs belonging to the genera Lachnospiraceae *Roseburia* and Ruminococcaceae *CAG-352* were respectively increased and decreased in levodopa users at follow-up (n=90 samples). (d) Bifidobacteriaceae were increased in pramipexole users at follow-up (n=10 samples). Each box represents the first quartile, median and third quartile at the lower, middle and upper boundaries, with the whiskers representing points within 1.5 the interquartile range and the X representing the mean. Relative abundances are presented on a log10 scale. ANCOM, ANalysis of Composition of Microbiomes; ASV, Amplicon Sequence Variant; BL, baseline samples; DESeq, Differential Expression analysis for Sequence count data; F1, follow-up samples one year after baseline.

*Bifidobacterium* and *Lactobacillus* showed a statistically significant increase at follow-up in dopamine agonist users before correction for multiple testing (*p*=0.035, *p*=0.042, respectively). A similar trend was found for their respective families. No associations were found between *Bifidobacterium, Lactobacillus* and levodopa usage. In contrast, the genera *Agathobacter, Lachnospira* and *Lachnospiraceae ND3007 group*, all belonging to the family Lachnospiraceae, as well as the genus *Prevotella*, showed a statistically significant decrease in levodopa users before correction for multiple testing (nominal *p*=0.0018, *p*=0.044, *p*=0.016, *p*=0.023, respectively). No associations were found between these taxa and dopamine agonist usage (Supplementary Tables 6-7)

### Dose-effect relationship of medication and microbiota

In addition, the effect of the medication dose as a continuous variable on overall community structure and differential abundance within the stratified groups were assessed using a PERMANOVA and DESeq2, respectively. Statistically significant correlations between medication dosage and individual taxa were excluded if the association was based on the presence of a taxon in less than three biological samples or could not be replicated after putative outlier removal (Supplementary Figure 2).

Both levodopa dosage and LEDD were associated with overall gut microbial community structure (Table 4). No associations for dopamine agonist dosage were found. However, smaller sample sizes must be noted for dopamine agonist usage, in particular for ropinirole, for which the number of permutations for the PERMANOVA had to be lowered. Indeed, the analysis seems underpowered, since the participant variable, merely added to pair samples from the same subject, showed no statistically significant association with overall gut microbiome composition, despite explaining more than two-thirds of variance.

**Table 4.** Effect of dopaminergic medication dosage on gut microbiota

**a. Multivariate analysis of gut microbial community structure**
|  | Aitchinson distance |  |  |  |  | Bray-Curtis dissimilarity |  |  |  |  |
| --- | --- | --- | --- | --- | --- | --- | --- | --- | --- | --- |
|  | Participant |  |  | Medication (mg) |  | Participant |  |  | Medication (mg) |  |
|  | n | % variance | p-value | % variance | p-value | n | % variance | p-value | % variance | p-value |
| **Levodopa** | 90 | 78.3 | **<1E-04** | 0.64 | **0.017** | 82 | 81.1 | **<1E-04** | 0.88 | **1.3E-03** |
| **Dopamine agonists<sup>1</sup>** | 16 | 75.5 | **<1E-04** | 2.60 | 0.62 | 14 | 75.7 | **<1E-04** | 4.20 | 0.16 |
| **Pramipexole** | 10 | 72.3 | **1.1E-03** | 4.60 | 0.59 | 10 | 73.7 | **3.0E-04** | 6.49 | 0.18 |
| **Ropinirole<sup>2</sup>** | 6 | 67.7 | 0.067 | 8.28 | 0.57 | 4 | 60.0 | 0.25 | 14.4 | 0.67 |
| **LEDD** | 124 | 78.5 | **<1E-04** | 0.47 | **0.013** | 114 | 80.8 | **<1E-04** | 0.63 | **1.0E-03** |

**b. Differential abundance**
|  | n | Level | Family | Genus | Species | base-mean | log2 fold change | DESeq2 (FDR) |
| --- | --- | --- | --- | --- | --- | --- | --- | --- |
| **Levodopa** | 90 | Genus | Lachnospiraceae | *ND3007 group* | - | 70.43 | -3.07E-03 | 0.021 |
| **Levodopa** | 90 | Family | Lachnospiraceae | - | - | 10353.20 | -9.13E-04 | 0.037 |
| **Levodopa** | 90 | Family | Erysipelatoclostridiaceae | - | - | 261.68 | -1.17 E-04 | 0.047 |
| **Levodopa** | 90 | Family | Ruminococcaceae | - | - | 6758.92 | -6.63E-04 | 0.047 |
| **Pramipexole** | 10 | ASV | Lachnospiraceae | *Fusicatenibacter* | *saccharivorans* | 902.91 | 2.09 | 0.031 |
| **Pramipexole** | 10 | Genus | Lachnospiraceae | *Fusicatenibacter* | - | 899.36 | 1.88 | 2.9E-03 |
| **LEDD** | 124 | Genus | Lachnospiraceae | *ND3007 group* | - | 59.65 | -2.68E-03 | 7.3E-03 |

Analysis of medication dosage effect as a continuous variable between baseline and follow-up samples of participants stratified for the usage of a given dopaminergic medication at follow-up. All analyses were paired through adjusting for participant as pairing variable. a. Multivariate analysis of gut microbial community structures using both Aitchinson distances and Bray-Curtis dissimilarities. Differences were assessed for statistical significance with a PERMANOVA based on 10,000 permutations, setting the lowest p-value at 9.999E-05 (<1E-04). b. Differential abundance analysis using DESeq2. Taxa identified for dopamine agonists (LEDD) and ropinirole were removed as the association was based on one outlier with all other observations being zero (supplementary figure 2). 1. Dopamine agonist dosages were converted to LEDD to create a composite dosage. 2. Due to low sample size, 719 and 23 permutations were used for the PERMANOVA of ropinirole dosage for Aitchinson distances and Bray-Curtis dissimilarities, respectively. % variance, percentage explained variance (R<sup>2</sup>); CR, controlled-release; LEDD, levodopa equivalent daily dosage; mg, milligram; n, number of samples.

The differential abundance analysis revealed an inverse association between levodopa dosage and the abundances of Lachnospiraceae, Erysipelatoclostridiaceae and Ruminococcaceae at the family level. The genus Lachnospiraceae *ND3007 group* was inversely correlated with both levodopa dosage and LEDD. Pramipexole dosage was positively correlated to the genus Lachnospiraceae *Fusicatenbacter* and one ASV belonging to its species *Fusicatenbacter saccharivorans* (Supplementary Table 8).

## Discussion

Longitudinal follow-up of *de novo* PD subjects, after initiation of dopaminergic treatment, confirms previously described cross-sectional alterations in treated PD subjects. No differences in alpha diversity and overall microbial community structure were found between baseline and follow-up, but overall microbial community structure was associated with levodopa dosage and LEDD. Levodopa and dopamine agonists seem to exert differential effects on gut microbiome composition. Levodopa was inversely related to the abundance of the families Ruminococcaceae and Lachnospiraceae, whereas Bifidobacteriaceae and an ASV belonging to *Bifidobacterium* were increased at follow-up in pramipexole users and overall dopamine agonist users, respectively.

Initiation of dopaminergic treatment was associated with reduced levels of several taxa belonging to the families Ruminococcaceae and Lachnospiraceae, including an ASV belonging to *Faecalibacterium prausnitzii* and the genus *Hungatella*. Reduced levels of Ruminococcaceae and Lachnospiraceae associated taxa are amongst the most replicated results in PD microbiome studies.^13,14,18,27^ Lower levels of *Faecalibacterium, Lachnospiraceae ND3007, Agathobacter, Lachnospira, Roseburia, Fusicatenbacter, Lachnospiraceae UCG-004* and *Blautia* have also been associated with levodopa dose.^14^ Here, levels of Ruminococcaceae and Lachnospiraceae indeed show an inverse correlation with levodopa dosage. Moreover, the inverse correlation between levodopa dose and *Lachnospiraceae ND3007 group* is a direct replication of previous findings.^14^ On the contrary, an ASV belonging to *Roseburia* was increased at follow-up in levodopa users.

However, the abundance of the ASV was rather low and does not seem to be representative of the entire genus, which was reduced at follow-up, although without statistical significance (*p*=0.11, *padj*=0.87, Supplementary Table 6). No other associations between genera belonging to Ruminococcaceae and Lachnospiraceae described in previous studies, were identified by ANCOM or DESeq2 as differentially abundant between baseline and follow-up or related to levodopa dosage after correction for multiple testing. However, *Agathobacter* and *Lachnospira* were decreased at follow-up in levodopa users with statistical significance before correction for multiple testing. Despite the strong association between Lachnospiraceae and levodopa in the current and previous studies, reduced levels of Lachnospiraceae and other SCFA producing taxa were already found in treatment-naïve *de novo* PD subjects independently of PD medication.^17,18^ PD medication might therefore initiate reductions in certain Lachnospiraceae taxa or aggravate already existing deficits.

The genera *Bifidobacterium* and *Lactobacillus* have been positively correlated with levodopa dosage, and both genera and their respective families, Bifidobacteriaceae and Lactobacillaceae, have been robustly replicated in several PD microbiome studies.^13–15^ Interestingly, no relation with levodopa usage or dosage could be established in the current dataset, but both families and genera were statistically significant increased at follow-up in dopamine agonist users before correction for multiple testing. In addition, one ASV belonging to *Bifidobacterium* was increased with statistical significance after correction for multiple testing in dopamine agonist users at follow-up, as was the family Bifidobacteriaceae in pramipexole users. The previously reported association between levodopa and *Bifidobacterium* and *Lactobacillus* might be explained by the concurrent use of dopamine agonists, as it is unclear what proportion of participants only used levodopa preparations.^14^ Levodopa and dopamine agonists groups in the current study were defined based on monotherapy, allowing the study of differential effects of both medications.

A differential effect between dopamine agonists and levodopa on gut microbiome composition also seems apparent from the positive correlation between pramipexole dosage and *Fusicatenibacter* and one of its ASVs. Previous publications suggest a negative correlation between already treated PD, levodopa and *Fusicatenibacter*.^14,20,30^ This is a trend also seen in the current dataset, although without statistical significance and based only on estimated effect size and effect direction. Also, the increased abundance of *Ruminococcaceae CAG-352* in dopamine agonist users at follow-up is in contrast with the reduced levels of one of its ASVs in levodopa users. The decreased levels at follow-up of an ASV belonging to Christensenellaceae in dopamine agonist users do not seem to be in contrast with previous studies that reported increased abundances of the entire family Christensellaceae in already treated subjects.^18,37^ The family Christensellaceae was not differentially abundant between baseline and follow-up in dopamine agonist users (*p*=0.51, *padj*=0.99, Supplementary Table 7).

Other expected findings include reduced levels of *Prevotella* and its family Prevotellaceae, which have been replicated in several PD microbiome studies, but not in treatment-naïve cohorts.^13,16,27^ In the entire study population, without stratification for dopaminergic monotherapy, *Prevotella* and Prevotellaceae showed a statistically significant decrease at follow-up before correction for multiple testing. Possibly, reduced levels of *Prevotella* and Prevotellaceae in PD can be influenced by medication effects. Previously reported decreases in *Clostridium group IV* after levodopa initiation could not be assessed as this taxon was not present in the current dataset.^26^ Similarly, no taxa belonging to *Enterococcus faecalis* and *Eggerthella lenta*, capable of intestinal decarboxylation of levodopa to dopamine and subsequent transformation to m-tyramine, respectively, were identified in the current dataset.^21^ However, presence of these species cannot be excluded, as DADA2 performs species assignment based on a 100% match against the reference sequence and might not have assigned ASVs to these species. Taxa belonging to *Lactobacillus* also contain the tyrosine decarboxylating gene (*tdc*) necessary for the bacterial conversion of levodopa to dopamine. Interestingly, *Lactobacillus* showed a statistically significant increase before correction for multiple testing, but only in dopamine agonist users. Two previous studies found associations of *tdc* abundance with levodopa dosage, and other PD medications, respectively.^22,23^ The association with PD medications other than levodopa is probably due to a more pronounced effect on gastro-intestinal motility of other medications.^22^ Therefore, studies aimed at investigating or modulating the effect of gut microbiota on the therapeutic availability of levodopa might benefit from participants using non-levodopa PD medications and a direct measurement of the tyrosine decarboxylation gene rather than surrogates derived from taxonomy. To conclude, Erysipelatoclostridiaceae was inversely correlated with levodopa dosage in the current dataset. This seems to be a novel finding as to our knowledge no previous PD microbiome study has reported on Erysipelatoclostridiaceae.

The current study has several clear advantages over previous studies and clearly addresses the effect of PD medication on gut microbiota. The advantages of this study include the longitudinal design with pre- and post-treatment measurements, the larger sample size compared to the only other treatment initiation study, and the stratification of dopaminergic monotherapy. Nonetheless, some limitations need to be addressed as well.

First, no quantitative data on treatment duration is available. Based on personal reports, the vast majority of participants have started their medication within a few weeks to months after collection of the baseline sample, resulting in almost a full year of exposure before collection of the follow-up sample. Additionally, the mean LEDD of 466mg is significantly higher than the often-used treatment initiation doses with a LEDD of 150mg or 300mg. A clinically and statistically significant decrease in MDS-UPDRSIII scores was seen as well, suggesting increased dosing and thereby longer treatment exposure to achieve adequate motor control. Nonetheless, individual participants might have started dopaminergic treatment at a later timepoint, reducing effect sizes. However, if this would have happened, the treatment effects on the microbiome would have been even larger.

Second, a follow-up period of one year was chosen deliberately, to accommodate longer medication exposure whilst minimizing the effects of disease progression on gut microbiome composition. However, the temporal stability of the gut microbiome in PD has not been assessed exhaustively. Moreover, higher LEDD scores at follow-up can be regarded a marker of faster progression. Ideally, a control group of PD subjects who were still treatment-naïve at follow-up should be included. However, the demonstrated benefit of immediate treatment initiation on quality of life of patients with disabling symptoms, makes this an unrealistic and unethical scenario.^38^

Third, sample sizes were relatively small, in particular of the groups with dopamine agonist users. Although the current study has a stronger longitudinal design, previous cross-sectional correlations between levodopa and gut microbiota required larger sample sizes to attain statistical significance and could not be replicated in a cohort three times as large as the current study.^14^ Several trends were found that are in line with previous studies, but were only statistically significant before correction for multiple testing. It can be hypothesized that larger longitudinal studies on dopaminergic treatment initiation can substantiate these trends. In addition, larger populations would allow for the inclusion of other PD medication groups. In particular COMT inhibitors, which have been associated with gut microbiome perturbations.^20,39^

Last, the current study would benefit from other data sources, providing different types of gut microbiome profiling, including metabolomics and shotgun metagenomics for a more functional readout. Also, increased taxonomic resolution as potentially provided by shotgun metagenomics could further aid clinical trial design of probiotic studies using specific species or strains.

In conclusion, the current study shows measurable and dose-dependent effects of PD medication on gut microbiota composition in PD. All previous study data on microbiome changes in non-drug naive PD patients should therefore be interpreted with caution, as several findings might be driven or accentuated by medication effects, as shown in this study. A remarkable finding is that levodopa and dopamine agonists exert differential effects on the gut microbiome. Larger studies could assess the validity of several trends revealed in the current study, that are in line with previous research. Nonetheless, PD is also characterized by microbial alterations independent from treatment initiation, including reduced levels of several SCFA producing taxa, in particular belonging to the family Lachnospiraceae. The current data on the interplay between PD medication and gut microbiota should be taken into account in future observational and interventional gut microbiota studies in PD.

## Methods

### Study population

Participants of the Dutch Parkinson Cohort of de novo PD subjects (DUPARC, NCT04180865) were included in the current study.^31^ PD was diagnosed according to the Movement Disorders Society (MDS) clinical diagnostic criteria by a movement disorder specialist.^3^ The diagnosis was further confirmed by FDOPA-PET and one-year follow-up. In seven participants no FDOPA-PET was performed, but only a clinical follow-up visit after one year was performed. For the current study, only participants who did not use PD medication at baseline, but did use PD medication at one year follow-up were included. In addition, participants with major confounders of gut microbiome composition (e.g., antibiotics usage within one month before sample collection) were excluded. For a complete overview of the exclusion criteria see Supplementary Table 1. 72 participants handed in stool samples at baseline and follow-up. Three participants had to be removed due to failed sequencing of the provided samples at one or both timepoints, five participants did not use dopaminergic medication at follow-up, and two participants had to be removed due to antibiotics usage within one month before sample collection, resulting in 62 participants in the final analysis.

The study was approved by the ethics committee of the University Medical Center Groningen, was conducted in concordance with the declaration of Helsinki and all participants provided written informed consent.

### Stool sample collection and clinical assessment

Stool sample collection and clinical assessments were performed as previously described.^17^ Briefly, stool samples were immediately frozen upon collection and kept frozen until DNA extraction. Participants were extensively characterized,^31^ including disease history, medication use, a dietary diary, a stool diary including the Bristol Stool Chart, the Montreal Cognitive Assessment (MoCA), and the Non-Motor Symptom Questionnaire (NMSQ).^40^ Motor symptomatology was assessed using the Movement Disorder Society Unified Parkinson Disease Rating Scale (MDS-UPDRS) parts II and III.^41^ MDS-UPDRS items that were not assessed were imputed using the value at the other timepoint to avoid introducing artificial differences between timepoints. If an item could not be assessed at either timepoint (e.g., due to mechanical comorbidity), the mean value of that item for all participants at both timepoints was imputed, to again avoid the introduction of artificial differences.

### Laboratory procedures

Wet lab work was performed at the University of Helsinki, Institute of Biotechnology, as previously described.^17^ DNA extraction was performed with the Qiagen Allprep DNA/RNA Mini Kit. A mixture of universal primers 515F1-4 (5’-GTGCCAGCMGCCGCGGTAA-3’) and 806R1-4 (5’-GGACTACHVGGGTWTCTAAT-3’), with partial Illumina TruSeq adapter sequences added to the 5′ ends, was used for amplification of the V4 hypervariable region of the 16S rRNA gene:

F1 ACACTCTTTCCCTACACGACGCTCTTCCGATCT,

F2 ACACTCTTTCCCTACACGACGCTCTTCCGATCTca,

F3 ACACTCTTTCCCTACACGACGCTCTTCCGATCTgca,

F4 ACACTCTTTCCCTACACGACGCTCTTCCGATCTagcaatt,

R1 GTGACTGGAGTTCAGACGTGTGCTCTTCCGATCT,

R2 GTGACTGGAGTTCAGACGTGTGCTCTTCCGATCTca,

R3 GTGACTGGAGTTCAGACGTGTGCTCTTCCGATCTatct,

R4 GTGACTGGAGTTCAGACGTGTGCTCTTCCGATCTtctact

Additional nucleotides (depicted in non-capitalized letters) were introduced for mixing in sequencing. Two-step PCR amplification was done as previously described.^27^ To assess possible contamination, a blank control was added to every DNA extraction and amplification batch. As much as practically possible, case and control samples were randomized to avoid batch effects. BARCOSEL was used to select barcodes.^42^ All samples were sequenced all together using Illumina MiSeq (v3 600 cycle kit), with 325 bases for the forward and 285 bases for the reverse reads in four repeated runs. An overview of the number the number of sequences of the blank control samples for the DNA extraction and PCR during the bio-informatics workflow, is provided in Supplementary Table 4. Very minimal to no contamination during the wet lab work was found.

### Bioinformatics

The bioinformatics pipeline was performed as previously described.^17^ Briefly, primers were removed with cutadapt (v2.10).^43^ DADA2 (v1.18) was used for further bioinformatics.^44^ Forward and reverse sequences were trimmed after 200 nucleotides and 150 nucleotides, respectively. All sequences were quality trimmed at the first instance of a (group of) nucleotide(s) with quality score 2. Sequences with ambiguous nucleotides or more than 10 expected errors were removed. Amplicon sequence variants (ASVs) were inferred using default parameters in DADA2, after which the four resulting sequence tables were merged. Subsequently, chimera were removed and identical ASVs that only differed in length were collapsed.

Taxonomic assignments are first performed up to genus level using a naïve Bayesian classifier with 50% bootstrap confidence assignments based on the SILVA (v138) reference database.^45^ Second, species are assigned in case of a 100% match with the references sequence that corresponds to the already assigned genus.

### Statistical analysis

All statistical analyses were performed with the statistical software R (v4.0.3). The threshold for statistical significance was set at (adjusted) p < 0.05 (two-sided). Clinical variables were compared using a McNemar’s Chi Square test for categorical variables, and a paired t-test or a Wilcoxon signed-rank test for continuous variables, depending on the distribution of the target variable.

A phyloseq object was created from the sequence table, taxonomy table and metadata table (phyloseq, v1.34.0).^46^ Data transformations and subsequent analysis were then performed using the R packages vegan (v2.5-7) and microbiome (v1.12.0). Alpha diversity indices Chao1, Shannon, Inverse Simpson and observed richness were calculated as measures of within-sample diversity and were tested univariably using a Wilcoxon signed-rank test.

Multivariate analyses of overall community structure were performed on Aitchinson distances (Euclidean distances calculated after centered log ratio transformation) and additionally on Bray-Curtis distances after rarefaction.^32^ The cutoff for rarefaction (26953) was based on rarefaction curves whilst keeping as many samples as possible (Supplementary Figure 1). Statistical testing was performed with a PERMANOVA with adjustment for selected variables using marginal testing.^33^ Differential abundance analyses were performed at the ASV, genus and family level with DESeq2 (v1.30.1) and ANCOM (v2.1).^34–36^ Taxa present in less than ten percent of all samples were removed before differential abundance analysis. For DESeq2, if rows representing taxa would not converge, it was tried to improve model convergence through scaling and centering of continuous variables, more stringent filtering and/or increasing the number of tries (maxit). However, more stringent filtering, as well as scaling and centering of continuous variables did not improve model convergence, and was therefore not performed in the final analyses. Taxa for which model convergence failed, were kept in the analysis to not loosen the correction for multiple testing. For ANCOM, default settings and code from github.com/FrederickHuangLin/ANCOM-Code-Archive were used. The filtering parameter zero_cut was changed from 0.9 to 1.0, since a filtering step was already performed as described above. This to prevent the additional loss of taxa when comparing very small sample sizes (eg. ropinirole users), and to have a list of taxa compatible with DESeq2 in the final analysis. Taxa were identified as differentially abundant with a false discovery rate (FDR) adjusted p-value < 0.05 in DESeq2 or with a detection threshold of 0.7 in ANCOM.

Complementary to the analysis of all participants, participants were stratified for dopaminergic monotherapy. One composite group of dopaminergic medication users was created due to the low sample sizes of those only using pramipexole or ropinirole. In addition, the effect of the medication dosage on overall community structure and differential abundance within the stratified groups were assessed using a PERMANOVA and DESeq2 respectively. For the composite group of dopamine agonist users and for all participants combined, the levodopa equivalent daily dose (LEDD) was used, calculated according to Tomlinson.^47^ Due to the low sample sizes of dopamine agonist users, statistically significant correlations between medication dosage and taxa were assessed for outlier effects through visual inspection of differential abundance dot plots. Associations were excluded if based on the presence of a taxon in less than three biological samples (Supplementary Figure 2). Otherwise, putative outliers were removed to assess if the found association could be replicated without the putative outlier(s).

All analyses using PERMANOVA, DESeq2 and ANCOM were paired through adjusting for the effect of the participant as the pairing variable. Results were plotted using ggplot2 (v3.3.3), with relative abundances calculated after adding a pseudo-count of 1 to sequence count data, in order to present data on a logarithmic scale.

### Variable selection

For the microbiome analyses concerning all participants (i.e., without stratification for dopaminergic monotherapy), all clinical and technical variables of interest were screened for their potential effect on the difference between baseline and follow-up visits using a PERMANOVA in the following model:

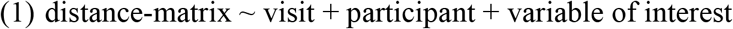

Variables that created a shift in explained variance (R^2^) of the visit variable of more than five percent compared to its explained variance without the variable (distance-matrix ∼ visit + participant), were added to the list of relevant model covariates (Supplementary Table 2). Multiple collinearity issues arose with variables that did not change between baseline and follow-up, showing collinearity with the pairing variable “participant”. Therefore, variables that resulted in a generalized variance inflation factor (GVIF) > 10 in model 1, or binary variables that did not change between baseline and follow-up (eg. sex), were excluded before variable screening. Variables with a GVIF ≥ 5 in the final model were excluded to avoid (multi)collinearity (Supplementary Table 3). This resulted in the following model with adjustment for relevant variables for the multivariate analysis of overall gut microbiome composition in all participants:

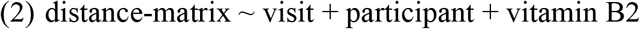

For the differential abundance analysis, variables were selected with a significant relationship (p < 0.1) in the final model of overall gut microbiome composition. However, no variables had a p-value below the selection threshold.

## Data Availability

Sequencing data of the V4 region of the 16S rRNA gene will be made available via the European Nucleotide Archive upon publication. Sample metadata can be requested from the principal investigator of the DUPARC cohort study upon reasonable request via.

## Code availability

No custom codes were used. All relevant software, packages and their versions are stated in the “Methods” section.

## Funding sources

The DUPARC study was funded by the Weston Brain Institute and personal grants for JMB to cover personnel costs.

We thank all participants in the DUPARC cohort study. For their aid in the recruitment of participants, we thank the Parkinson Platform Northern Netherlands (PPNN) Study Group collaborators.* We would like to thank Renée Speijers, Yvonne Nijman and Hanna Slomp for their help in the recruitment and logistics of the study. We thank Velma Aho for her advice concerning data analysis. We acknowledge the DNA Sequencing and Genomics Laboratory, Institute of Biotechnology, University of Helsinki, for DNA isolations and sequencing of FIN and NL cohort samples, in particular Eevakaisa Vesanen, Ursula Lönnqvist and Julia Teleni.

*PPNN Study Group:

Verwey NA^1^, Van Harten B^1^, Portman AT^2^, Langedijk MJH^2^, Oomes PG^2^, Jansen BJAM^2^, Van Wieren T^2^, Van den Bogaard SJA^3^, Van Steenbergen W^3^, Duyff R^3^, Van Amerongen JP^3^, Fransen PSS^4^, Polman SKL^4^, Zwartbol RT^4^, Van Kesteren ME^4^, Braakhekke JP^4^, Trip J^4^, Koops L^4^, De Langen CJ^4^, De Jong G^4^, Hartono JES^4^, Ybema H^4^, Bartels AL^5^, Reesink FE^5^, Postma AG^6^, Vonk GJH^7^, Oen JMTH^7^, Brinkman MJ^7^, Mondria T^7^, Holscher RS^7^, Van der Meulen AAE^8^, Rutgers AWF^8^, Boekestein WA^9^, Teune LK^9^, Orsel PJL^10^, Hoogendijk JE^10^, Van Laar T^11^.

^1^Department of Neurology, Medisch Centrum Leeuwarden, Leeuwarden, the Netherlands; ^2^Department of Neurology, Treant Zorggroep locations, Stadskanaal, Emmen, Hoogeveen, the Netherlands; ^3^Department of Neurology, Tjongerschans Ziekenhuis Heerenveen, Heerenveen, the Netherlands; ^4^Department of Neurology, Isala, Zwolle, Meppel, the Netherlands; ^5^Department of Neurology, Ommelander Ziekenhuis Groningen, Scheemda, the Netherlands; ^6^Department of Neurology, Nij Smellinghe Ziekenhuis Drachten, Drachten, the Netherlands; ^7^Department of Neurology, Antonius Zorggroep, Sneek, the Netherlands; ^8^Department of Neurology, Martini Ziekenhuis, Groningen, the Netherlands; ^9^Department of Neurology, Wilhelmina Ziekenhuis Assen, Assen, the Netherlands; ^10^Department of Neurology, Sionsberg, Dokkum, the Netherlands; ^11^Department of Neurology, University Medical Center Groningen, Groningen, the Netherlands.

## Competing interests statement

JMB received an honorarium for writing an article for the magazine “Kinetic” by Britannia Pharmaceuticals and owns exchange traded funds that might include stocks in medically-related fields, SvdZ none, PL none, FS has received grants from The Academy of Finland, The Hospital District of Helsinki and Uusimaa, OLVI-Foundation, Konung Gustaf V:s och Drottning Victorias Frimurarestiftelse, The Wilhelm and Else Stockmann Foundation, The Emil Aaltonen Foundation, The Yrjö Jahnsson Foundation, Renishaw, and honoraria from AbbVie, Orion, GE Healthcare, Merck, Teva, Bristol Myers Squibb, Sanofi, Biocodex, and Biogen. FS is founder and CEO of NeuroInnovation Oy and NeuroBiome Ltd., is a member of the scientific advisory board and has received consulting fees and stock options from Axial Biotherapeutics. TvL has received grant support from the MJFF, the UMCG, Menzis, Weston Brain Institute and the Dutch Brain Foundation. Consultancy fees were received from AbbVie, Britannia Pharm., Centrapharm and Neuroderm. Speaker fees were received from AbbVie, Britannia Pharm. and Eurocept. PABP, LP, PA, and FS have patents issued (FI127671B, US10,139,408, US11,499,971) and pending (US16/186,663, US17/116,045, US17/933,952, EP3789501, EP3149205) that are assigned to NeuroBiome Ltd. All authors declare to have no non-financial conflicts of interest.

## Author contributions

JMB contributed to the study conception, participant inclusion, data collection, data analysis, writing the first draft of the manuscript and later reviewing of the manuscript. PABP contributed to data analysis and reviewing of the manuscript. SvdZ contributed to participant inclusion, data collection and reviewing of the manuscript. PL contributed to data analysis and reviewing of the manuscript. LP contributed to the design of the laboratory work, data analysis and reviewing of the manuscript. PA contributed to the design of the laboratory procedures, data analysis and reviewing of the manuscript. FS contributed to data analysis and reviewing of the manuscript. TvL contributed to the study conception, participant inclusion, data collection and reviewing of the manuscript.

## Notes

### Clinical Protocols

https://bmcneurol.biomedcentral.com/articles/10.1186/s12883-020-01811-3

