## Supplementary Materials for "Dopaminergic Medication Accentuates Fecal Gut Microbiome Changes in Parkinson’s Disease"

### Supplementary Material

#### Contents


- Supplementary table 1: Exclusion criteria (p.2)
- Supplementary figure 1: Rarefaction curves (p.3)
- Supplementary table 2: Variable selection (p.4)
- Supplementary table 3: Collinearity screening (p.8)
- Supplementary table 4: Lost reads blank controls (p.11)
- Supplementary table 5: Differential abundances all participants at ASV, genus and family level (p.15)
- Supplementary table 6: Differential abundances levodopa users at ASV, genus and family level (p.55)
- Supplementary table 7: Differential abundances dopamine agonist users (combined, and stratified for only pramipexole and ropinirole users) at ASV, genus and family level (p.95)
- Supplementary table 8: Dose-effect relationships between dopaminergic medication and gut microbiota at ASV, genus and family level (p.193)
- Supplementary figure 2: Outlier check statistically significant associations dose-effect relationships (p.371)

#### Supplementary Table 1: exclusion criteria

All participants:

- Active or persistent primary disease of the gastrointestinal tract
- History of peritonitis, severe endometriosis, abdominal, intestinal or urogenital fistula,
- Hepatobiliar or pancreatic disease (except asymptomatic cholecystolithiasis)
- History of abdominal or anorectal surgery, except minor surgery such as uncomplicated appendectomy or cholecystectomy (>6 months ago).
- Severe gynaecological prolapse (grade III)
- Cancer and/or adjuvant treatment within the last 6 months
- Within the last three months: severe hypo- or hyperkalemia, narcosis, analgosedation, endoscopic procedure of the gastrointestinal tract, abdominal trauma
- Within the last three months: gastrointestinal tract infection, food intoxication
- Antibiotic use in the last month

PD participants:

- History of dopaminergic medication use

HC participants:

- History of neurodegenerative disease, in particular signs of parkinsonism, including probable prodromal PD.

### Supplementary Figure 1: rarefaction curves

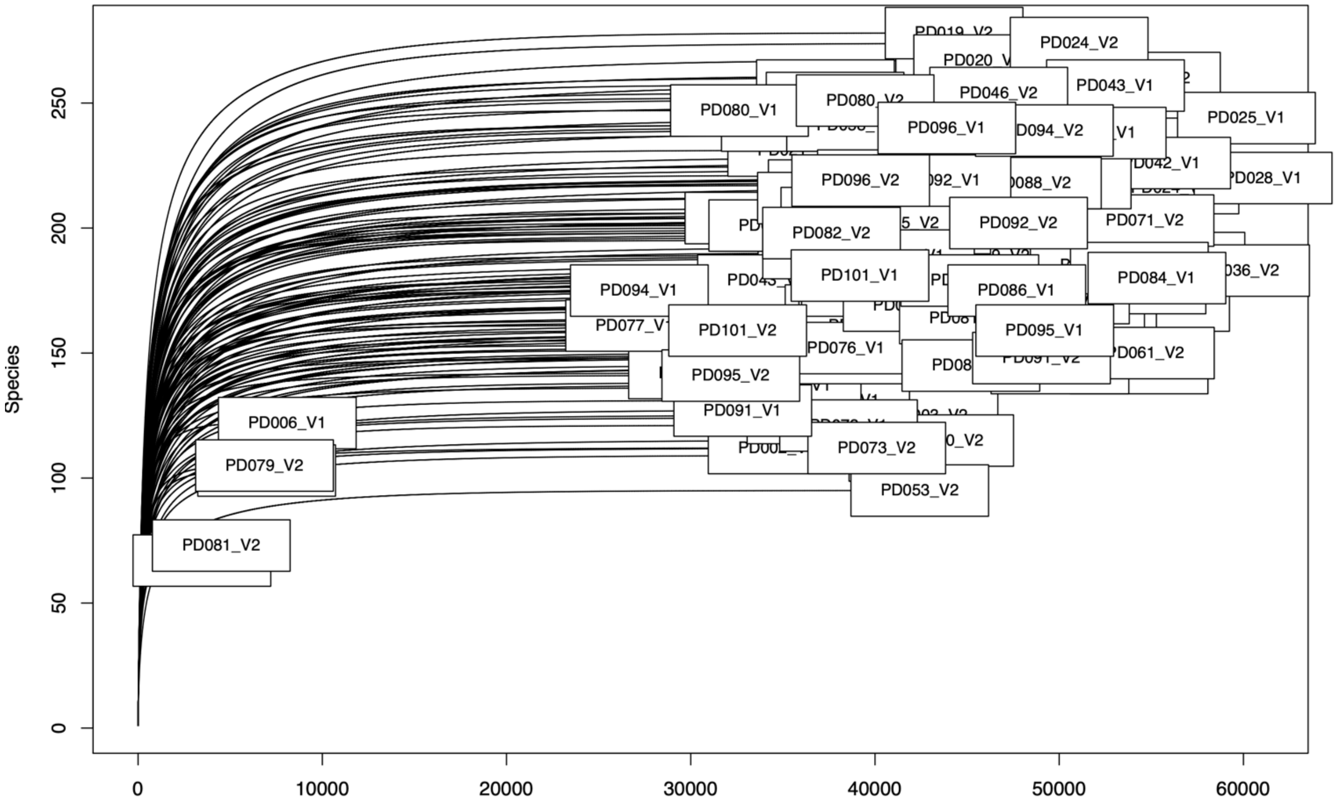

#### Supplementary Table 2: variable selection

Screening for the influence of assessed variables on the relationship between medication status and overall microbiome composition.
Variables were incorporated in a paired model with medication status (overall microbiome composition ~ medication status + participant + variable) to assess the extend to which the variable changes the explained variance (R2) of medication status on overall microbiome composition.
Variables with >5 percent shift in the R2 (%Shift_R2) of PD status, compared to the univariable R2 of PD status (calculated for each comparison separately dependent on the removal of samples due to missing data of the variable investigated) were added to the list of nuisance variables.

| variable | MEDstatus_R2 pairedNoAdjustment | MEDstatus_p pairedNoAdjustment | MEDstatus_R2 pairedAdjustedForVariable | MEDstatus_p pairedAdjustedForVariable | %Shift_R2 |
|---|---|---|---|---|---|
| vitB6_mg | 0,004649985 | 0,106589341 | 0,004284769 | 0,266473353 | -7,854139279 |
| iron_haem_mg | 0,004649985 | 0,106589341 | 0,004317416 | 0,228977102 | -7,152045758 |
| nicotinicacid_mg | 0,004649985 | 0,106589341 | 0,00433394 | 0,240575942 | -6,796698906 |
| folateequivalent_ug | 0,004649985 | 0,106589341 | 0,004346487 | 0,228277172 | -6,526866564 |
| fat_energypercentage | 0,004649985 | 0,106589341 | 0,004390173 | 0,202879712 | -5,587368878 |
| vitB2_mg | 0,004649985 | 0,106589341 | 0,004400569 | 0,202279772 | -5,363805186 |
| med_betablocker | 0,004107329 | 0,068393161 | 0,003888323 | 0,138986101 | -5,332087244 |
| med_antiepileptic | 0,004107329 | 0,068393161 | 0,003893434 | 0,125087491 | -5,207638424 |
| med_cholinesterase_inhibitor | 0,004107329 | 0,068393161 | 0,003932196 | 0,111788821 | -4,263918528 |
| cholesterol_mg | 0,004649985 | 0,106589341 | 0,004462475 | 0,175082492 | -4,0325025 |
| updrsIII_hoehnandyahr | 0,004107329 | 0,068393161 | 0,003942253 | 0,109989001 | -4,019056891 |
| med_antihistaminic | 0,004107329 | 0,068393161 | 0,00395191 | 0,111088891 | -3,783947304 |
| iron_total_mg | 0,004649985 | 0,106589341 | 0,004487078 | 0,163483652 | -3,503394615 |
| fattotal_g | 0,004649985 | 0,106589341 | 0,004489511 | 0,161683832 | -3,451068202 |
| vitK2_ug | 0,004649985 | 0,106589341 | 0,004491594 | 0,159384062 | -3,406271586 |
| folicacid_ug | 0,004649985 | 0,106589341 | 0,004492919 | 0,159984002 | -3,377785674 |
| med_phosphodiesteraseinhibitors | 0,004107329 | 0,068393161 | 0,003987225 | 0,099890011 | -2,924142061 |
| polyols_total_g | 0,004649985 | 0,106589341 | 0,004515499 | 0,150484952 | -2,892192766 |
| vitK1_ug | 0,004649985 | 0,106589341 | 0,004521844 | 0,146885311 | -2,755737273 |
| fattyacids_saturated_total_g | 0,004649985 | 0,106589341 | 0,004525436 | 0,149685031 | -2,678493656 |
| retinol_activityequivalent_ug | 0,004649985 | 0,106589341 | 0,004527121 | 0,143985601 | -2,64224337 |
| updrs_stebbins_tdpigd_subtype | 0,004300633 | 0,067193281 | 0,00441364 | 0,0449955 | 2,627694698 |

| variable | MEDstatus_R2 pairedNoAdjustment | MEDstatus_p pairedNoAdjustment | MEDstatus_R2 pairedAdjustedForVariable | MEDstatus_p pairedAdjustedForVariable | %Shift_R2 |
|---|---|---|---|---|---|
| retinol_ug | 0,004649985 | 0,106589341 | 0,004528156 | 0,143785621 | -2,619984983 |
| retinolequivalent_ug | 0,004649985 | 0,106589341 | 0,004529082 | 0,143785621 | -2,60007703 |
| med_kortisone_oldhels | 0,004107329 | 0,068393161 | 0,004001893 | 0,090790921 | -2,567029473 |
| med_tricyclicantidepressant | 0,004107329 | 0,068393161 | 0,004211472 | 0,054294571 | 2,535530346 |
| med_otherantidepressant | 0,004107329 | 0,068393161 | 0,004211472 | 0,054294571 | 2,535530346 |
| vitD3_cholecalciferol_ug | 0,004649985 | 0,106589341 | 0,004766845 | 0,081491851 | 2,513120907 |
| fattyacids_polyunsaturated_n6cis_g | 0,004649985 | 0,106589341 | 0,004544762 | 0,138386161 | -2,262880913 |
| vitK_total_ug | 0,004649985 | 0,106589341 | 0,004545414 | 0,140785921 | -2,248862321 |
| drugs_recreational | 0,004195524 | 0,089191081 | 0,004104496 | 0,117088291 | -2,169660593 |
| folate_ug | 0,004649985 | 0,106589341 | 0,004551426 | 0,139886011 | -2,119569309 |
| med_constipation | 0,004107329 | 0,068393161 | 0,004020642 | 0,084891511 | -2,110549189 |
| vitD_25hydroxy_ug | 0,004649985 | 0,106589341 | 0,004552493 | 0,138686131 | -2,096620652 |
| fattyacids_monounsaturated_cis_total_g | 0,004649985 | 0,106589341 | 0,004560076 | 0,135386461 | -1,933536204 |
| lutein_ug | 0,004649985 | 0,106589341 | 0,004563499 | 0,133186681 | -1,859934988 |
| proteinanimal_g | 0,004649985 | 0,106589341 | 0,004563847 | 0,133186681 | -1,852447029 |
| stoolconsistency_avg | 0,004123863 | 0,084091591 | 0,0041998 | 0,063493651 | 1,841390028 |
| zinc_mg | 0,004649985 | 0,106589341 | 0,004564997 | 0,132286771 | -1,827707043 |
| med_ppi | 0,004107329 | 0,068393161 | 0,004035609 | 0,085791421 | -1,746136272 |
| gamma_tocopherol_mg | 0,004649985 | 0,106589341 | 0,004730147 | 0,087991201 | 1,723912319 |
| med_corticosteroids_other | 0,004107329 | 0,068393161 | 0,00403689 | 0,078692131 | -1,714965322 |
| organicacids_total_g | 0,004649985 | 0,106589341 | 0,004729109 | 0,091890811 | 1,701600139 |
| vitB12_ug | 0,004649985 | 0,106589341 | 0,004571919 | 0,131786821 | -1,678840323 |
| iron_nonhaem_mg | 0,004649985 | 0,106589341 | 0,004574466 | 0,135786421 | -1,624076786 |
| med_calcium | 0,004107329 | 0,068393161 | 0,004170174 | 0,061393861 | 1,530064142 |
| vitB1_mg | 0,004649985 | 0,106589341 | 0,004585095 | 0,131386861 | -1,395497561 |
| med_multivit | 0,004107329 | 0,068393161 | 0,004162064 | 0,062593741 | 1,332610529 |
| ash_g | 0,004649985 | 0,106589341 | 0,004710825 | 0,091190881 | 1,30837866 |
| fattyacids_polyunsaturated_total_g | 0,004649985 | 0,106589341 | 0,004593465 | 0,122687731 | -1,215501984 |
| delta_tocopherol_mg | 0,004649985 | 0,106589341 | 0,004594056 | 0,131686831 | -1,202785074 |

| variable | MEDstatus_R2 pairedNoAdjustment | MEDstatus_p pairedNoAdjustment | MEDstatus_R2 pairedAdjustedForVariable | MEDstatus_p pairedAdjustedForVariable | %Shift_R2 |
|---|---|---|---|---|---|
| DNA_extraction_concentration | 0,004107329 | 0,068393161 | 0,004153657 | 0,059694031 | 1,127924492 |
| copper_mg | 0,004649985 | 0,106589341 | 0,004598739 | 0,126487351 | -1,102069061 |
| polysaccharides_total_g | 0,004649985 | 0,106589341 | 0,004602431 | 0,113688631 | -1,022675619 |
| vitC_mg | 0,004649985 | 0,106589341 | 0,004604619 | 0,122987701 | -0,975618186 |
| stoolfrequency_avg | 0,004123863 | 0,084091591 | 0,004085959 | 0,093790621 | -0,919138626 |
| sodium_mg | 0,004649985 | 0,106589341 | 0,004690412 | 0,097090291 | 0,869387977 |
| med_benzodiazepines | 0,004107329 | 0,068393161 | 0,004142028 | 0,061593841 | 0,844816808 |
| med_vitd | 0,004107329 | 0,068393161 | 0,00413817 | 0,065093491 | 0,750869235 |
| alpha_tocopherol_mg | 0,004649985 | 0,106589341 | 0,004684751 | 0,098090191 | 0,747662345 |
| potassium_mg | 0,004649985 | 0,106589341 | 0,004616238 | 0,118788121 | -0,725752423 |
| vitD_total_ug | 0,004649985 | 0,106589341 | 0,004619489 | 0,121087891 | -0,655826396 |
| med_antiemetic | 0,004107329 | 0,068393161 | 0,004133949 | 0,065093491 | 0,648101347 |
| magnesium_mg | 0,004649985 | 0,106589341 | 0,004620738 | 0,117488251 | -0,628985898 |
| nitrogen_total_g | 0,004649985 | 0,106589341 | 0,004622088 | 0,112788721 | -0,599943807 |
| nmsq_total | 0,003991114 | 0,211178882 | 0,003968444 | 0,219478052 | -0,56800032 |
| proteintotal_g | 0,004649985 | 0,106589341 | 0,004625672 | 0,115088491 | -0,522859638 |
| phosphor_mg | 0,004649985 | 0,106589341 | 0,004674173 | 0,102789721 | 0,520174599 |
| proteinplant_g | 0,004649985 | 0,106589341 | 0,004672607 | 0,101489851 | 0,486479602 |
| water_g | 0,004649985 | 0,106589341 | 0,004669866 | 0,102089791 | 0,427545295 |
| selenium_ug | 0,004649985 | 0,106589341 | 0,004668812 | 0,102789721 | 0,404872833 |
| nrofreads_seqtab_nolengthfilt | 0,004107329 | 0,068393161 | 0,004091268 | 0,066893311 | -0,391040051 |
| kJ | 0,004649985 | 0,106589341 | 0,004667988 | 0,101089891 | 0,387162165 |
| betacarotene_ug | 0,004649985 | 0,106589341 | 0,004667981 | 0,101689831 | 0,387005745 |
| calcium_mg | 0,004649985 | 0,106589341 | 0,004667349 | 0,102989701 | 0,373416535 |
| stoolsample_consistency | 0,004114445 | 0,137286271 | 0,00410137 | 0,136186381 | -0,317791761 |
| lycopene_ug | 0,004649985 | 0,106589341 | 0,004660042 | 0,100689931 | 0,216270779 |
| iodine_ug | 0,004649985 | 0,106589341 | 0,004640262 | 0,109389061 | -0,209112154 |
| vitE_total_mg | 0,004649985 | 0,106589341 | 0,004659003 | 0,109889011 | 0,19393833 |
| moca_total | 0,004201533 | 0,067993201 | 0,004193515 | 0,071292871 | -0,190843392 |

| variable | MEDstatus_R2 pairedNoAdjustment | MEDstatus_p pairedNoAdjustment | MEDstatus_R2 pairedAdjustedForVariable | MEDstatus_p pairedAdjustedForVariable | %Shift_R2 |
|---|---|---|---|---|---|
| zeaxanthin_ug | 0,004649985 | 0,106589341 | 0,004658803 | 0,106689331 | 0,189623902 |
| alcoholdiet_g | 0,004649985 | 0,106589341 | 0,004658548 | 0,106189381 | 0,184154321 |
| alpha_carotene_ug | 0,004649985 | 0,106589341 | 0,004658351 | 0,105089491 | 0,179903429 |
| med_corticosteroids_systemic | 0,004107329 | 0,068393161 | 0,004101942 | 0,073192681 | -0,131157204 |
| mono_disaccharides_total_g | 0,004649985 | 0,106589341 | 0,004653812 | 0,106589341 | 0,082288427 |
| fibers_total_g | 0,004649985 | 0,106589341 | 0,004647166 | 0,104089591 | -0,06062068 |
| beta_cryptoxanthin_ug | 0,004649985 | 0,106589341 | 0,004652417 | 0,104889511 | 0,052296294 |
| fattyacids_polyunsaturated_n3cis_g | 0,004649985 | 0,106589341 | 0,004648208 | 0,105089491 | -0,038211165 |
| carbstotal_g | 0,004649985 | 0,106589341 | 0,004649153 | 0,106489351 | -0,017896145 |
| beta_tocopherol_mg | 0,004649985 | 0,106589341 | 0,004650466 | 0,110188981 | 0,010329289 |
| kcal | 0,004649985 | 0,106589341 | 0,004649954 | 0,106789321 | -0,000676582 |
| med_nsaids | 0,004107329 | 0,068393161 | 0,004107329 | 0,068193181 | 1,82E-12 |
| med_statin | 0,004107329 | 0,068393161 | 0,004107329 | 0,069093091 | 1,82E-12 |
| med_systemic_hormonal_treatment | 0,004107329 | 0,068393161 | 0,004107329 | 0,060893911 | 9,08E-13 |

#### Supplementary Table 3: collinearity screening of selected variable

**Selected variables were screened for (multi-)collinearity using a generalized variance inflation factor (GVIF). Variables were excluded to obtain a maximum GVIF <5 in the final model (Supplementary Table 3a). Variables were removed based on highest GVIF with either the visit or participant variable (Supplementary Table 3b). Remaining variables were excluded based on largest GVIF until GVIF of all variables in the final model was below 5**

**Supplementary Table 3a: exclusion of variables based on GVIF**

| variable | GVIF | Df | GVIF^(1/2Df) |
|---|---|---|---|
| **visit** | **1,58754665** | **1** | **1,25997883** |
| participantID | 51538,2489 | 61 | 1,09300969 |
| vitB6_mg | 11,3554227 | 1 | 3,36978081 |
| iron_haem_mg | 5,51548061 | 1 | 2,34850604 |
| nicotinicacid_mg | 11,771431 | 1 | 3,43095191 |
| folateequivalent_ug | 8,43546621 | 1 | 2,90438741 |
| fat_energypercentage | 7,5751841 | 1 | 2,75230523 |
| vitB2_mg | 7,98766462 | 1 | 2,82624568 |
| med_betablocker | 7,95970315 | 1 | 2,82129459 |
| med_antiepileptic | 2,68144195 | 1 | 1,6375109 |

**After removal of med_betablocker and fat_energypercentage ( highest GVIF with "participantID", Supplementary Table 3b)**

| variable | GVIF | Df | GVIF^(1/2Df) |
|---|---|---|---|
| visit | 1,44744722 | 1 | 1,20309901 |
| participantID | 1118,35623 | 61 | 1,05922534 |
| vitB6_mg | 10,4969266 | 1 | 3,23989608 |
| iron_haem_mg | 5,49216867 | 1 | 2,34353764 |
| nicotinicacid_mg | 11,6086743 | 1 | 3,40715047 |
| folateequivalent_ug | 8,32939487 | 1 | 2,8860691 |
| vitB2_mg | 7,48274547 | 1 | 2,73546074 |
| med_antiepileptic | 2,10800952 | 1 | 1,45189859 |

**After removal of folateequivalent_ug and nicotinicacid_mg (highest remaining GVIF with "participantID", Supplementary Table 3b)**

| variable | GVIF | Df | GVIF^(1/2Df) |
|---|---|---|---|
| visit | 1,42316248 | 1 | 1,19296374 |
| participantID | 82,1524475 | 61 | 1,03679671 |
| vitB6_mg | 4,65308577 | 1 | 2,15710124 |
| iron_haem_mg | 5,08048117 | 1 | 2,25399227 |
| vitB2_mg | 4,69256742 | 1 | 2,16623346 |
| med_antiepileptic | 2,08108926 | 1 | 1,44259809 |

**After removal of med_antiepileptic: not the highest GVIF, but only one participant started using it, and unclear if participant really did not use the medication at baseline**

| variable | GVIF | Df | GVIF^(1/2Df) |
|---|---|---|---|
| visit | 1,40985614 | 1 | 1,18737363 |
| participantID | 39,9773983 | 61 | 1,03069371 |
| vitB6_mg | 4,63552956 | 1 | 2,153028 |
| iron_haem_mg | 5,01758188 | 1 | 2,23999596 |
| vitB2_mg | 4,56092058 | 1 | 2,13563119 |

**Removal of iron_haem_mg (highest remaining GVIF)**

| variable | GVIF | Df | GVIF^(1/2Df) |
|---|---|---|---|
| visit | 1,24722887 | 1 | 1,11679401 |
| participantID | 10,6488878 | 61 | 1,01957817 |
| vitB6_mg | 4,63397221 | 1 | 2,1526663 |
| vitB2_mg | 3,39790167 | 1 | 1,84333981 |

**Removal of vitB6_mg (highest remaining GVIF)**

| variable | GVIF | Df | GVIF^(1/2Df) |
|---|---|---|---|
| visit | 1,15685654 | 1 | 1,07557266 |
| participantID | 3,03840064 | 61 | 1,00915089 |
| vitB2_mg | 2,88397441 | 1 | 1,69822684 |

**Double check if removal of med_antiepileptic was necessary (only variable not removed due to GVIF):**
**indeed necessary, GVIF of participantID in final model would be > 5**

| **variable** | **GVIF** | **Df** | **GVIF^(1/2Df)** |
|---|---|---|---|
| visit | 1,18817646 | 1 | 1,09003507 |
| participantID | 6,15043133 | 61 | 1,01500093 |
| vitB2_mg | 2,92311853 | 1 | 1,709713 |
| med_antiepileptic | 2,04816319 | 1 | 1,43114052 |

**Supplementary Table 3b: GVIF between variables**

| | **visit** | **participant-ID** | **vitB6_mg** | **iron-haem_mg** | **nicotinic-acid_mg** | **folate-equivalent_ug** | **fat_energy-percentage** | **vitB2_mg** | **med_beta-blocker** | **med_anti-epileptic** |
|---|---|---|---|---|---|---|---|---|---|---|
| **visit** | NA | 1 | 1,06372171 | 1,07550244 | 1,04973058 | 1,03813872 | 1,01621081 | 1,02781888 | 1,01049869 | 1,00819672 |
| **participantID** | 1 | NA | 3,46439965 | 2,96512388 | 3,51279063 | 3,60747197 | 5,96011789 | 2,69949249 | 6,20967742 | 1,98387097 |
| **vitB6_mg** | 1,06372171 | 3,46439965 | NA | 1,06052202 | 1,97057245 | 1,53588905 | 1,00013706 | 1,27303485 | 1,04258193 | 1,00045651 |
| **iron_haem_mg** | 1,07550244 | 2,96512388 | 1,06052202 | NA | 1,40057418 | 1,03616133 | 1,01331483 | 1,27409059 | 1,00142935 | 1,00004522 |
| **nicotinicacid_mg** | 1,04973058 | 3,51279063 | 1,97057245 | 1,40057418 | NA | 1,12758771 | 1,03283898 | 1,2083517 | 1,00704967 | 1,00006763 |
| **folateequivalent_ug** | 1,03813872 | 3,60747197 | 1,53588905 | 1,03616133 | 1,12758771 | NA | 1,00292412 | 1,70148247 | 1,00760351 | 1,00053804 |
| **fat_energypercentage** | 1,01621081 | 5,96011789 | 1,00013706 | 1,01331483 | 1,03283898 | 1,00292412 | NA | 1,01470298 | 1,00163508 | 1,00242364 |
| **vitB2_mg** | 1,02781888 | 2,69949249 | 1,27303485 | 1,27409059 | 1,2083517 | 1,70148247 | 1,01470298 | NA | 1,0047658 | 1,00085695 |
| **med_betablocker** | 1,01049869 | 6,20967742 | 1,04258193 | 1,00142935 | 1,00704967 | 1,00760351 | 1,00163508 | 1,0047658 | NA | 1,00103581 |
| **med_antiepileptic** | 1,00819672 | 1,98387097 | 1,00045651 | 1,00004522 | 1,00006763 | 1,00053804 | 1,00242364 | 1,00085695 | 1,00103581 | NA |

**Supplementary Table 4: lost reads of blank controls for all four sequencing runs.**
**Columns represent the number of reads in the raw data,**
**the output after using cutadapt, the input used for DADA2, after quality trimming and filtering,**
**after denoising the forward reads, after denoising the reverse reads,**
**after merging and in the eventual sequence table, respectively.**
**Last column represents the percentage of lost reads at the end of the pipeline.**

**Sequencing run 1**

| sampleID | type | rawData | output-Cutadapt | input-DADA2 | filtered | denoisedF | denoisedR | merged | seqtab | percLost |
|---|---|---|---|---|---|---|---|---|---|---|
| Bl_0_ctrl_1 | PCR_control | 10 | 10 | 10 | 7 | 2 | 1 | 0 | 0 | 100 |
| Bl_0_ctrl_10 | PCR_control | 14 | 14 | 14 | 10 | 1 | 1 | 0 | 0 | 100 |
| Bl_0_ctrl_11 | PCR_control | 11 | 11 | 11 | 9 | 1 | 1 | 0 | 0 | 100 |
| Bl_0_ctrl_12 | PCR_control | 12 | 12 | 12 | 11 | 1 | 1 | 0 | 0 | 100 |
| Bl_0_ctrl_13 | PCR_control | 13 | 13 | 13 | 9 | 1 | 1 | 0 | 0 | 100 |
| Bl_0_ctrl_14 | PCR_control | 8 | 8 | 8 | 5 | 2 | 1 | 0 | 0 | 100 |
| Bl_0_ctrl_1T | PCR_control | 11 | 11 | 11 | 9 | 1 | 1 | 0 | 0 | 100 |
| Bl_0_ctrl_2 | PCR_control | 17 | 17 | 17 | 10 | 1 | 1 | 0 | 0 | 100 |
| Bl_0_ctrl_3 | PCR_control | 11 | 11 | 11 | 9 | 3 | 1 | 0 | 0 | 100 |
| Bl_0_ctrl_4 | PCR_control | 5 | 5 | 5 | 4 | 1 | 1 | 0 | 0 | 100 |
| Bl_0_ctrl_5 | PCR_control | 14 | 14 | 14 | 6 | 1 | 1 | 0 | 0 | 100 |
| Bl_0_ctrl_6 | PCR_control | 6 | 6 | 6 | 5 | 1 | 1 | 0 | 0 | 100 |
| Bl_0_ctrl_7 | PCR_control | 4 | 4 | 4 | 4 | 1 | 1 | 0 | 0 | 100 |
| Bl_0_ctrl_8 | PCR_control | 6 | 6 | 6 | 5 | 1 | 1 | 0 | 0 | 100 |
| Bl_0_ctrl_9 | PCR_control | 13 | 13 | 13 | 10 | 2 | 1 | 0 | 0 | 100 |
| kit_control_1 | DNA_extraction_control | 95 | 95 | 95 | 77 | 1 | 1 | 0 | 0 | 100 |
| kit_control_10 | DNA_extraction_control | 87 | 87 | 87 | 74 | 14 | 1 | 0 | 0 | 100 |
| kit_control_11 | DNA_extraction_control | 76 | 76 | 76 | 69 | 24 | 1 | 0 | 0 | 100 |
| kit_control_12 | DNA_extraction_control | 16 | 16 | 16 | 13 | 1 | 1 | 0 | 0 | 100 |
| kit_control_13 | DNA_extraction_control | 19 | 19 | 19 | 17 | 3 | 1 | 0 | 0 | 100 |
| kit_control_14 | DNA_extraction_control | 61 | 61 | 61 | 52 | 5 | 1 | 0 | 0 | 100 |
| kit_control_15 | DNA_extraction_control | 65 | 65 | 65 | 57 | 5 | 1 | 0 | 0 | 100 |
| kit_control_2 | DNA_extraction_control | 85 | 85 | 85 | 73 | 6 | 1 | 0 | 0 | 100 |
| kit_control_3 | DNA_extraction_control | 69 | 69 | 69 | 59 | 11 | 10 | 0 | 0 | 100 |
| kit_control_4 | DNA_extraction_control | 73 | 73 | 73 | 62 | 19 | 1 | 0 | 0 | 100 |
| kit_control_5_A | DNA_extraction_control | 68 | 68 | 68 | 54 | 8 | 1 | 0 | 0 | 100 |
| kit_control_5_B | DNA_extraction_control | 92 | 92 | 92 | 78 | 18 | 1 | 0 | 0 | 100 |
| kit_control_6 | DNA_extraction_control | 60 | 60 | 60 | 54 | 2 | 1 | 0 | 0 | 100 |
| kit_control_7 | DNA_extraction_control | 66 | 66 | 66 | 57 | 9 | 1 | 0 | 0 | 100 |
| kit_control_8 | DNA_extraction_control | 72 | 72 | 72 | 63 | 14 | 1 | 0 | 0 | 100 |
| kit_control_9 | DNA_extraction_control | 72 | 72 | 72 | 64 | 8 | 1 | 0 | 0 | 100 |

**Sequencing run 2**

| sampleID | type | rawData | output-Cutadapt | input-DADA2 | filtered | denoisedF | denoisedR | merged | seqtab | percLost |
|---|---|---|---|---|---|---|---|---|---|---|
| BI_0_ctrl_1 | PCR_control | 12 | 12 | 12 | 8 | 1 | 1 | 0 | 0 | 100 |
| BI_0_ctrl_10 | PCR_control | 6 | 6 | 6 | 6 | 1 | 1 | 0 | 0 | 100 |
| BI_0_ctrl_11 | PCR_control | 13 | 13 | 13 | 7 | 1 | 1 | 0 | 0 | 100 |
| BI_0_ctrl_12 | PCR_control | 19 | 19 | 19 | 17 | 2 | 1 | 0 | 0 | 100 |
| BI_0_ctrl_13 | PCR_control | 16 | 16 | 16 | 16 | 2 | 1 | 0 | 0 | 100 |
| BI_0_ctrl_14 | PCR_control | 7 | 7 | 7 | 7 | 2 | 1 | 0 | 0 | 100 |
| BI_0_ctrl_15 | PCR_control | 1 | 1 | 1 | 1 | 1 | 1 | 0 | 0 | 100 |
| BI_0_ctrl_16 | PCR_control | 1 | 1 | 1 | 1 | 1 | 1 | 0 | 0 | 100 |
| BI_0_ctrl_1T | PCR_control | 9 | 9 | 9 | 7 | 1 | 1 | 0 | 0 | 100 |
| BI_0_ctrl_2 | PCR_control | 24 | 24 | 24 | 13 | 3 | 1 | 0 | 0 | 100 |
| BI_0_ctrl_3 | PCR_control | 10 | 10 | 10 | 9 | 1 | 1 | 0 | 0 | 100 |
| BI_0_ctrl_4 | PCR_control | 2 | 2 | 2 | 2 | 1 | 1 | 0 | 0 | 100 |
| BI_0_ctrl_5 | PCR_control | 7 | 7 | 7 | 6 | 1 | 1 | 0 | 0 | 100 |
| BI_0_ctrl_6 | PCR_control | 5 | 5 | 5 | 4 | 1 | 1 | 0 | 0 | 100 |
| BI_0_ctrl_7 | PCR_control | 6 | 6 | 6 | 5 | 1 | 1 | 0 | 0 | 100 |
| BI_0_ctrl_8 | PCR_control | 7 | 7 | 7 | 5 | 1 | 1 | 0 | 0 | 100 |
| BI_0_ctrl_9 | PCR_control | 10 | 10 | 10 | 9 | 1 | 1 | 0 | 0 | 100 |
| kit_control_1 | DNA_extraction_control | 87 | 87 | 87 | 84 | 17 | 1 | 0 | 0 | 100 |
| kit_control_10 | DNA_extraction_control | 82 | 82 | 82 | 74 | 21 | 1 | 0 | 0 | 100 |
| kit_control_11 | DNA_extraction_control | 75 | 75 | 75 | 72 | 2 | 1 | 0 | 0 | 100 |
| kit_control_12 | DNA_extraction_control | 17 | 17 | 17 | 15 | 1 | 1 | 0 | 0 | 100 |
| kit_control_13 | DNA_extraction_control | 21 | 21 | 21 | 18 | 1 | 1 | 0 | 0 | 100 |
| kit_control_14 | DNA_extraction_control | 56 | 56 | 56 | 53 | 12 | 1 | 0 | 0 | 100 |
| kit_control_15 | DNA_extraction_control | 61 | 61 | 61 | 56 | 2 | 1 | 0 | 0 | 100 |
| kit_control_2 | DNA_extraction_control | 81 | 81 | 81 | 77 | 10 | 1 | 0 | 0 | 100 |
| kit_control_3 | DNA_extraction_control | 67 | 67 | 67 | 60 | 16 | 1 | 0 | 0 | 100 |
| kit_control_4 | DNA_extraction_control | 67 | 67 | 67 | 63 | 1 | 1 | 0 | 0 | 100 |
| kit_control_5_A | DNA_extraction_control | 79 | 79 | 79 | 69 | 16 | 1 | 0 | 0 | 100 |
| kit_control_5_B | DNA_extraction_control | 129 | 129 | 129 | 125 | 60 | 13 | 0 | 0 | 100 |
| kit_control_6 | DNA_extraction_control | 71 | 71 | 71 | 68 | 12 | 1 | 0 | 0 | 100 |
| kit_control_7 | DNA_extraction_control | 49 | 49 | 49 | 47 | 1 | 1 | 0 | 0 | 100 |
| kit_control_8 | DNA_extraction_control | 62 | 62 | 62 | 60 | 2 | 1 | 0 | 0 | 100 |
| kit_control_9 | DNA_extraction_control | 66 | 66 | 66 | 61 | 2 | 1 | 0 | 0 | 100 |

**Sequencing run 3**

| sampleID | type | rawData | output-Cutadapt | input-DADA2 | filtered | denoisedF | denoisedR | merged | seqtab | percLost |
|---|---|---|---|---|---|---|---|---|---|---|
| BI_0_ctrl_1 | PCR_control | 5 | 5 | 5 | 4 | 3 | 1 | 0 | 0 | 100 |
| BI_0_ctrl_10 | PCR_control | 10 | 10 | 10 | 7 | 1 | 1 | 0 | 0 | 100 |
| BI_0_ctrl_11 | PCR_control | 6 | 6 | 6 | 4 | 1 | 1 | 0 | 0 | 100 |
| BI_0_ctrl_12 | PCR_control | 11 | 11 | 11 | 9 | 1 | 1 | 0 | 0 | 100 |
| BI_0_ctrl_13 | PCR_control | 23 | 23 | 23 | 16 | 1 | 1 | 0 | 0 | 100 |
| BI_0_ctrl_14 | PCR_control | 7 | 7 | 7 | 5 | 1 | 1 | 0 | 0 | 100 |
| BI_0_ctrl_15 | PCR_control | 5 | 5 | 5 | 5 | 1 | 1 | 0 | 0 | 100 |
| BI_0_ctrl_16 | PCR_control | 1 | 1 | 1 | 1 | 1 | 1 | 0 | 0 | 100 |
| BI_0_ctrl_1T | PCR_control | 10 | 10 | 10 | 8 | 1 | 1 | 0 | 0 | 100 |
| BI_0_ctrl_2 | PCR_control | 30 | 30 | 30 | 20 | 1 | 1 | 0 | 0 | 100 |
| BI_0_ctrl_3 | PCR_control | 5 | 5 | 5 | 4 | 1 | 1 | 0 | 0 | 100 |
| BI_0_ctrl_4 | PCR_control | 18 | 18 | 18 | 11 | 1 | 1 | 0 | 0 | 100 |
| BI_0_ctrl_5 | PCR_control | 7 | 7 | 7 | 4 | 1 | 1 | 0 | 0 | 100 |
| BI_0_ctrl_6 | PCR_control | 7 | 7 | 7 | 6 | 2 | 2 | 1 | 1 | 85,714 |
| BI_0_ctrl_7 | PCR_control | 5 | 5 | 5 | 2 | 1 | 1 | 1 | 1 | 80 |
| BI_0_ctrl_8 | PCR_control | 5 | 5 | 5 | 3 | 1 | 1 | 0 | 0 | 100 |
| BI_0_ctrl_9 | PCR_control | 9 | 9 | 9 | 8 | 1 | 1 | 0 | 0 | 100 |
| kit_control_1 | DNA_extraction_control | 81 | 81 | 81 | 75 | 10 | 1 | 0 | 0 | 100 |
| kit_control_10 | DNA_extraction_control | 52 | 52 | 52 | 50 | 18 | 1 | 0 | 0 | 100 |
| kit_control_11 | DNA_extraction_control | 62 | 62 | 62 | 60 | 12 | 1 | 0 | 0 | 100 |
| kit_control_12 | DNA_extraction_control | 9 | 9 | 9 | 8 | 1 | 1 | 0 | 0 | 100 |
| kit_control_13 | DNA_extraction_control | 16 | 16 | 16 | 15 | 2 | 1 | 0 | 0 | 100 |
| kit_control_14 | DNA_extraction_control | 51 | 51 | 51 | 48 | 17 | 1 | 0 | 0 | 100 |
| kit_control_15 | DNA_extraction_control | 75 | 75 | 75 | 65 | 17 | 1 | 0 | 0 | 100 |
| kit_control_2 | DNA_extraction_control | 94 | 94 | 94 | 90 | 26 | 1 | 0 | 0 | 100 |
| kit_control_3 | DNA_extraction_control | 49 | 49 | 49 | 46 | 2 | 1 | 0 | 0 | 100 |
| kit_control_4 | DNA_extraction_control | 84 | 84 | 84 | 78 | 20 | 1 | 0 | 0 | 100 |
| kit_control_5_A | DNA_extraction_control | 80 | 80 | 80 | 70 | 3 | 1 | 0 | 0 | 100 |
| kit_control_5_B | DNA_extraction_control | 115 | 115 | 115 | 108 | 32 | 1 | 0 | 0 | 100 |
| kit_control_6 | DNA_extraction_control | 48 | 48 | 48 | 46 | 4 | 1 | 0 | 0 | 100 |
| kit_control_7 | DNA_extraction_control | 61 | 61 | 61 | 59 | 12 | 1 | 0 | 0 | 100 |
| kit_control_8 | DNA_extraction_control | 81 | 81 | 81 | 76 | 8 | 1 | 0 | 0 | 100 |
| kit_control_9 | DNA_extraction_control | 86 | 86 | 86 | 80 | 25 | 1 | 0 | 0 | 100 |

**Sequencing run 4**

| sampleID | type | rawData | output-Cutadapt | input-DADA2 | filtered | denoisedF | denoisedR | merged | seqtab | percLost |
|---|---|---|---|---|---|---|---|---|---|---|
| BI_0_ctrl_1 | PCR_control | 9 | 9 | 9 | 8 | 1 | 1 | 0 | 0 | 100 |
| BI_0_ctrl_10 | PCR_control | 11 | 11 | 11 | 9 | 3 | 1 | 0 | 0 | 100 |
| BI_0_ctrl_11 | PCR_control | 10 | 10 | 10 | 5 | 1 | 1 | 0 | 0 | 100 |
| BI_0_ctrl_12 | PCR_control | 6 | 6 | 6 | 6 | 1 | 1 | 0 | 0 | 100 |
| BI_0_ctrl_13 | PCR_control | 11 | 11 | 11 | 7 | 1 | 1 | 0 | 0 | 100 |
| BI_0_ctrl_14 | PCR_control | 6 | 6 | 6 | 4 | 1 | 1 | 0 | 0 | 100 |
| BI_0_ctrl_15 | PCR_control | 2 | 2 | 2 | 2 | 1 | 1 | 0 | 0 | 100 |
| BI_0_ctrl_16 | PCR_control | 1 | 1 | 1 | 1 | 1 | 1 | 0 | 0 | 100 |
| BI_0_ctrl_1T | PCR_control | 9 | 9 | 9 | 6 | 1 | 1 | 0 | 0 | 100 |
| BI_0_ctrl_2 | PCR_control | 26 | 26 | 26 | 18 | 3 | 1 | 0 | 0 | 100 |
| BI_0_ctrl_3 | PCR_control | 5 | 5 | 5 | 4 | 1 | 1 | 0 | 0 | 100 |
| BI_0_ctrl_4 | PCR_control | 5 | 5 | 5 | 4 | 1 | 1 | 0 | 0 | 100 |
| BI_0_ctrl_5 | PCR_control | 5 | 5 | 5 | 2 | 1 | 1 | 0 | 0 | 100 |
| BI_0_ctrl_6 | PCR_control | 4 | 4 | 4 | 2 | 1 | 1 | 0 | 0 | 100 |
| BI_0_ctrl_7 | PCR_control | 4 | 4 | 4 | 2 | 1 | 1 | 0 | 0 | 100 |
| BI_0_ctrl_8 | PCR_control | 9 | 9 | 9 | 6 | 1 | 1 | 0 | 0 | 100 |
| BI_0_ctrl_9 | PCR_control | 10 | 10 | 10 | 7 | 1 | 1 | 0 | 0 | 100 |
| kit_control_1 | DNA_extraction_control | 76 | 76 | 76 | 70 | 33 | 1 | 0 | 0 | 100 |
| kit_control_10 | DNA_extraction_control | 89 | 89 | 89 | 82 | 27 | 1 | 0 | 0 | 100 |
| kit_control_11 | DNA_extraction_control | 57 | 57 | 57 | 51 | 2 | 1 | 0 | 0 | 100 |
| kit_control_12 | DNA_extraction_control | 15 | 15 | 15 | 12 | 1 | 1 | 0 | 0 | 100 |
| kit_control_13 | DNA_extraction_control | 25 | 25 | 25 | 20 | 1 | 1 | 0 | 0 | 100 |
| kit_control_14 | DNA_extraction_control | 40 | 40 | 40 | 38 | 1 | 1 | 0 | 0 | 100 |
| kit_control_15 | DNA_extraction_control | 51 | 51 | 51 | 43 | 14 | 1 | 0 | 0 | 100 |
| kit_control_2 | DNA_extraction_control | 65 | 65 | 65 | 58 | 3 | 3 | 0 | 0 | 100 |
| kit_control_3 | DNA_extraction_control | 62 | 62 | 62 | 54 | 13 | 1 | 0 | 0 | 100 |
| kit_control_4 | DNA_extraction_control | 69 | 69 | 69 | 60 | 16 | 1 | 0 | 0 | 100 |
| kit_control_5_A | DNA_extraction_control | 58 | 58 | 58 | 52 | 15 | 1 | 0 | 0 | 100 |
| kit_control_5_B | DNA_extraction_control | 95 | 95 | 95 | 84 | 25 | 10 | 6 | 6 | 93,684 |
| kit_control_6 | DNA_extraction_control | 53 | 53 | 53 | 44 | 8 | 1 | 0 | 0 | 100 |
| kit_control_7 | DNA_extraction_control | 37 | 37 | 37 | 30 | 1 | 1 | 0 | 0 | 100 |
| kit_control_8 | DNA_extraction_control | 54 | 54 | 54 | 51 | 17 | 1 | 0 | 0 | 100 |
| kit_control_9 | DNA_extraction_control | 44 | 44 | 44 | 42 | 17 | 1 | 0 | 0 | 100 |

**Supplementary Table 5: differential abundance at ASV, genus and family level in all participants with DESeq2 and ANCOM**
**All columns are spread out over two pages, after which the next two pages show the next set of rows, etc.**
**A ticker is provided on both pages to easily connect the taxonomy data to the associated statistics.**

| Ticker | taxLevel | Phylum | Family | Genus | Species |
|---|---|---|---|---|---|
| 1 | ASV | Actinobacteriota | Bifidobacteriaceae | Bifidobacterium | NA |
| 2 | ASV | Verrucomicrobiota | Akkermansiaceae | Akkermansia | muciniphila |
| 3 | ASV | Firmicutes | Christensenellaceae | NA | NA |
| 4 | ASV | Firmicutes | Lachnospiraceae | Roseburia | NA |
| 5 | ASV | Firmicutes | NA | NA | NA |
| 6 | ASV | Firmicutes | NA | NA | NA |
| 7 | ASV | Firmicutes | Christensenellaceae | Christensenellaceae R-7 group | NA |
| 8 | ASV | Bacteroidota | Bacteroidaceae | Bacteroides | NA |
| 9 | ASV | Bacteroidota | Bacteroidaceae | Bacteroides | eggerthii |
| 10 | ASV | Firmicutes | UCG-010 | NA | NA |
| 11 | ASV | Firmicutes | [Eubacterium] coprostanoligenes group | NA | NA |
| 12 | ASV | Firmicutes | Ruminococcaceae | Subdoligranulum | NA |
| 13 | ASV | Firmicutes | Ruminococcaceae | Faecalibacterium | prausnitzii |
| 14 | ASV | Firmicutes | Lachnospiraceae | Lachnospira | pectinoschiza |
| 15 | ASV | Firmicutes | Oscillospiraceae | UCG-002 | NA |
| 16 | ASV | Firmicutes | Lachnospiraceae | NA | NA |
| 17 | ASV | Bacteroidota | Bacteroidaceae | Bacteroides | coprocola |
| 18 | ASV | Firmicutes | UCG-010 | NA | NA |
| 19 | ASV | Firmicutes | Ruminococcaceae | NA | NA |
| 20 | ASV | Firmicutes | Oscillospiraceae | UCG-003 | NA |
| 21 | ASV | Bacteroidota | Rikenellaceae | Alistipes | finegoldii |
| 22 | ASV | Firmicutes | NA | NA | NA |
| 23 | ASV | Firmicutes | Ruminococcaceae | Subdoligranulum | NA |
| 24 | ASV | Firmicutes | Veillonellaceae | Dialister | invisus |
| 25 | ASV | Firmicutes | Lachnospiraceae | Anaerostipes | hadrus |
| 26 | ASV | Bacteroidota | Barnesiellaceae | Barnesiella | NA |

| Ticker | DESeq2_baseMean | DESeq2_log2-FoldChange | DESeq2_lfcSE | DESeq2_stat | DESeq2_pvalue | DESeq2_padj | DESeq2 converged | ANCOM_W | ANCOM_detected_0.7 |
|---|---|---|---|---|---|---|---|---|---|
| 1 | 329,4609511 | 0,725241386 | 0,502988246 | 1,441865473 | 0,149340328 | 0,99894472 | YES | 0 | FALSE |
| 2 | 912,9538844 | -0,119399277 | 0,450207507 | -0,265209431 | 0,79084813 | 0,99894472 | YES | 0 | FALSE |
| 3 | 306,1393149 | 0,668062466 | 0,400404055 | 1,668470782 | 0,095222307 | 0,99894472 | YES | 0 | FALSE |
| 4 | 98,61254423 | 3,696327673 | 1,438183376 | 2,570136558 | 0,010165843 | 0,417816161 | YES | 0 | FALSE |
| 5 | 250,8038323 | 0,251824979 | 2,901795834 | 0,086782459 | 0,93084443 | 0,99894472 | YES | 0 | FALSE |
| 6 | 0,982444278 | 0,119608844 | 2,902589714 | 0,04120763 | 0,967130371 | 0,99894472 | YES | 0 | FALSE |
| 7 | 53,48045156 | 0,018090828 | 0,523776953 | 0,034539183 | 0,972447198 | 0,99894472 | YES | 0 | FALSE |
| 8 | 55,6836761 | 0,090562677 | 2,083117595 | 0,043474587 | 0,965323222 | 0,99894472 | YES | 0 | FALSE |
| 9 | 48,01217935 | 0,003838533 | 2,902267923 | 0,001322598 | 0,99894472 | 0,99894472 | YES | 0 | FALSE |
| 10 | 1,129599922 | 0,129679509 | 2,904750919 | 0,044643934 | 0,964391123 | 0,99894472 | YES | 0 | FALSE |
| 11 | 88,6634389 | -0,217016283 | 0,821767083 | -0,264084907 | 0,791714494 | 0,99894472 | YES | 0 | FALSE |
| 12 | 392,023144 | -0,28718425 | 0,235216042 | -1,220938193 | 0,222109422 | 0,99894472 | YES | 0 | FALSE |
| 13 | 45,96783544 | -2,873567149 | 2,904442071 | -0,989369758 | 0,322482265 | 0,99894472 | YES | 0 | FALSE |
| 14 | 72,13617156 | -1,924998545 | 0,577376246 | -3,334045276 | 0,000855927 | 0,175893048 | NO | 0 | FALSE |
| 15 | 40,77630395 | 0,267879944 | 2,903292877 | 0,092267627 | 0,926485409 | 0,99894472 | YES | 0 | FALSE |
| 16 | 146,3177638 | -0,10067464 | 2,902872575 | -0,03468104 | 0,97233408 | 0,99894472 | YES | 0 | FALSE |
| 17 | 56,2438255 | -0,90787338 | 2,903698308 | -0,312661056 | 0,754538186 | 0,99894472 | YES | 0 | FALSE |
| 18 | 2,665945516 | -0,01238474 | 2,905116901 | -0,004263078 | 0,996598566 | 0,99894472 | YES | 0 | FALSE |
| 19 | 1,533889816 | -0,063565852 | 2,904793751 | -0,021883086 | 0,982541217 | 0,99894472 | YES | 0 | FALSE |
| 20 | 138,848875 | -0,608471375 | 0,38918436 | -1,563452795 | 0,117946131 | 0,99894472 | YES | 0 | FALSE |
| 21 | 42,22019237 | 0,685760782 | 2,900809014 | 0,236403286 | 0,813119747 | 0,99894472 | YES | 3 | FALSE |
| 22 | 302,3165998 | 0,512125432 | 2,90251606 | 0,176441894 | 0,859946794 | 0,99894472 | YES | 0 | FALSE |
| 23 | 52,62298781 | -0,180888929 | 0,852657968 | -0,212147116 | 0,831992263 | 0,99894472 | YES | 0 | FALSE |
| 24 | 353,6568712 | 0,381742873 | 2,195291631 | 0,173891645 | 0,861950619 | 0,99894472 | YES | 1 | FALSE |
| 25 | 18,72093584 | 4,250487099 | 2,902497093 | 1,46442424 | 0,143078069 | 0,99894472 | YES | 0 | FALSE |
| 26 | 41,60087519 | 0,03604909 | 2,904774493 | 0,012410289 | 0,990098277 | 0,99894472 | YES | 0 | FALSE |

| Ticker | taxLevel | Phylum | Family | Genus | Species |
|---|---|---|---|---|---|
| 27 | ASV | Firmicutes | Lachnospiraceae | [Ruminococcus] torques group | NA |
| 28 | ASV | Firmicutes | NA | NA | NA |
| 29 | ASV | Firmicutes | Oscillospiraceae | UCG-002 | NA |
| 30 | ASV | Bacteroidota | Tannerellaceae | Parabacteroides | distasonis |
| 31 | ASV | Firmicutes | Lachnospiraceae | Lachnospiraceae NK4A136 group | bacterium |
| 32 | ASV | Firmicutes | Oscillospiraceae | NK4A214 group | NA |
| 33 | ASV | Firmicutes | Christensenellaceae | Christensenellaceae R-7 group | NA |
| 34 | ASV | Firmicutes | Lachnospiraceae | Fusicatenibacter | saccharivorans |
| 35 | ASV | Proteobacteria | Enterobacteriaceae | Klebsiella | NA |
| 36 | ASV | Firmicutes | Lachnospiraceae | NA | NA |
| 37 | ASV | Firmicutes | Ruminococcaceae | CAG-352 | NA |
| 38 | ASV | Actinobacteriota | Eggerthellaceae | Senegalimassilia | anaerobia |
| 39 | ASV | Actinobacteriota | Eggerthellaceae | Enterorhabdus | NA |
| 40 | ASV | Bacteroidota | Rikenellaceae | Alistipes | inops |
| 41 | ASV | Firmicutes | Ruminococcaceae | NA | NA |
| 42 | ASV | Firmicutes | Butyricicoccaceae | Butyricicoccus | faecihominis |
| 43 | ASV | Bacteroidota | Rikenellaceae | Alistipes | obesi |
| 44 | ASV | Firmicutes | Christensenellaceae | Christensenellaceae R-7 group | NA |
| 45 | ASV | Firmicutes | Lachnospiraceae | Blautia | massiliensis |
| 46 | ASV | Bacteroidota | Tannerellaceae | Parabacteroides | distasonis |
| 47 | ASV | Firmicutes | Lachnospiraceae | Lachnoclostridium | NA |
| 48 | ASV | Firmicutes | Ruminococcaceae | Faecalibacterium | prausnitzii |
| 49 | ASV | Firmicutes | Peptostreptococcaceae | Terrisporobacter | mayombei |
| 50 | ASV | Desulfobacterota | Desulfovibrionaceae | Bilophila | wadsworthia |
| 51 | ASV | Bacteroidota | Muribaculaceae | NA | NA |
| 52 | ASV | Firmicutes | Erysipelotrichaceae | Turicibacter | sanguinis |
| 53 | ASV | Firmicutes | Peptococcaceae | NA | NA |
| 54 | ASV | Firmicutes | Oscillospiraceae | Oscillibacter | NA |
| 55 | ASV | Firmicutes | NA | NA | NA |
| 56 | ASV | Firmicutes | Oscillospiraceae | UCG-002 | NA |

| Ticker | DESeq2_baseMean | DESeq2_log2-FoldChange | DESeq2_lfcSE | DESeq2_stat | DESeq2_pvalue | DESeq2_padj | DESeq2 converged | ANCOM_W | ANCOM_detected_0.7 |
|---|---|---|---|---|---|---|---|---|---|
| 27 | 121,3343881 | -1,809597497 | 0,899336567 | -2,012147135 | 0,044204429 | 0,99894472 | YES | 0 | FALSE |
| 28 | 48,88295812 | 2,345161697 | 2,90265721 | 0,807936152 | 0,419127338 | 0,99894472 | YES | 0 | FALSE |
| 29 | 223,9543426 | 0,482471647 | 0,506672887 | 0,952234981 | 0,340977824 | 0,99894472 | YES | 0 | FALSE |
| 30 | 100,1236925 | 0,34779767 | 0,588836004 | 0,59065286 | 0,554753039 | 0,99894472 | YES | 0 | FALSE |
| 31 | 76,66129975 | -0,768382373 | 1,207164555 | -0,636518335 | 0,524438634 | 0,99894472 | YES | 0 | FALSE |
| 32 | 102,520782 | 0,476383627 | 0,721073716 | 0,660658704 | 0,508831213 | 0,99894472 | YES | 0 | FALSE |
| 33 | 88,04840875 | -0,203176304 | 0,357800003 | -0,567848804 | 0,570137638 | 0,99894472 | YES | 0 | FALSE |
| 34 | 784,3099195 | -0,006458111 | 0,288116413 | -0,022414935 | 0,982116967 | 0,99894472 | YES | 0 | FALSE |
| 35 | 60,97545019 | 4,122324161 | 2,904730087 | 1,419176322 | 0,155847617 | 0,99894472 | YES | 0 | FALSE |
| 36 | 219,9122333 | 0,496962121 | 0,418255769 | 1,188177565 | 0,234763461 | 0,99894472 | YES | 0 | FALSE |
| 37 | 23,21558955 | -0,820342895 | 2,904423415 | -0,282446041 | 0,777601516 | 0,99894472 | YES | 0 | FALSE |
| 38 | 17,57410805 | 0,156101013 | 0,552273726 | 0,282651529 | 0,777443976 | 0,99894472 | YES | 0 | FALSE |
| 39 | 7,464465206 | 0,002152032 | 0,800792281 | 0,002687379 | 0,997855784 | 0,99894472 | YES | 0 | FALSE |
| 40 | 66,61369828 | 0,260977241 | 0,844551134 | 0,309012955 | 0,757311672 | 0,99894472 | YES | 0 | FALSE |
| 41 | 137,4812202 | -0,331653216 | 0,462264066 | -0,717454027 | 0,473093994 | 0,99894472 | YES | 0 | FALSE |
| 42 | 25,09711944 | 0,156370854 | 1,071912321 | 0,145880266 | 0,884015911 | 0,99894472 | YES | 0 | FALSE |
| 43 | 78,85312392 | 0,38313767 | 0,485107637 | 0,789799297 | 0,42964499 | 0,99894472 | YES | 0 | FALSE |
| 44 | 174,3923518 | 1,354111966 | 0,627796588 | 2,156927882 | 0,031011284 | 0,980433672 | YES | 0 | FALSE |
| 45 | 411,9541167 | 0,059433023 | 0,27201796 | 0,218489336 | 0,827047867 | 0,99894472 | YES | 0 | FALSE |
| 46 | 52,28557923 | -0,062450605 | 2,581563054 | -0,024191005 | 0,980700253 | 0,99894472 | YES | 0 | FALSE |
| 47 | 56,93268433 | 0,110254491 | 0,593487402 | 0,185773937 | 0,852622044 | 0,99894472 | YES | 0 | FALSE |
| 48 | 99,35817288 | -0,110512252 | 1,647013936 | -0,067098553 | 0,946503246 | 0,99894472 | YES | 0 | FALSE |
| 49 | 7,024029205 | 0,34840672 | 2,9031332 | 0,120010587 | 0,904474761 | 0,99894472 | YES | 0 | FALSE |
| 50 | 53,68000431 | 0,307888467 | 0,309608135 | 0,994445661 | 0,320005948 | 0,99894472 | YES | 0 | FALSE |
| 51 | 131,9175803 | -0,828192067 | 2,000064901 | -0,414082596 | 0,678813618 | 0,99894472 | YES | 0 | FALSE |
| 52 | 9,696741845 | 0,300595499 | 2,902138896 | 0,103577227 | 0,917504861 | 0,99894472 | YES | 0 | FALSE |
| 53 | 1,34026348 | 0,256672604 | 2,904840332 | 0,088360314 | 0,929590303 | 0,99894472 | YES | 0 | FALSE |
| 54 | 47,25557163 | 0,078209579 | 0,500698926 | 0,156200811 | 0,875874737 | 0,99894472 | YES | 0 | FALSE |
| 55 | 56,24238155 | -0,445716261 | 1,459822173 | -0,305322298 | 0,760120695 | 0,99894472 | YES | 0 | FALSE |
| 56 | 70,30307183 | -0,69007768 | 1,844526656 | -0,374121826 | 0,708313681 | 0,99894472 | YES | 0 | FALSE |

| Ticker | taxLevel | Phylum | Family | Genus | Species |
|---|---|---|---|---|---|
| 57 | ASV | Firmicutes | Christensenellaceae | Christensenellaceae R-7 group | NA |
| 58 | ASV | Proteobacteria | Sutterellaceae | Sutterella | NA |
| 59 | ASV | Firmicutes | Butyricicoccaceae | Butyricicoccus | NA |
| 60 | ASV | Firmicutes | Lachnospiraceae | Roseburia | hominis |
| 61 | ASV | Firmicutes | Streptococcaceae | Streptococcus | NA |
| 62 | ASV | Firmicutes | NA | NA | NA |
| 63 | ASV | Firmicutes | Ruminococcaceae | Ruminococcus | NA |
| 64 | ASV | Bacteroidota | Prevotellaceae | Prevotella | NA |
| 65 | ASV | Firmicutes | Oscillospiraceae | UCG-005 | NA |
| 66 | ASV | Firmicutes | Lachnospiraceae | [Eubacterium] ventriosum group | NA |
| 67 | ASV | Firmicutes | Lachnospiraceae | Roseburia | NA |
| 68 | ASV | Firmicutes | Peptostreptococcaceae | Romboutsia | NA |
| 69 | ASV | Firmicutes | Ruminococcaceae | Ruminococcus | callidus |
| 70 | ASV | Firmicutes | Oscillospiraceae | UCG-002 | NA |
| 71 | ASV | Desulfobacterota | Desulfovibrionaceae | Desulfovibrio | NA |
| 72 | ASV | Bacteroidota | Tannerellaceae | Parabacteroides | distasonis |
| 73 | ASV | Firmicutes | Oscillospiraceae | Intestinimonas | NA |
| 74 | ASV | Firmicutes | Lachnospiraceae | Lachnoclostridium | edouardi |
| 75 | ASV | Bacteroidota | Barnesiellaceae | Barnesiella | NA |
| 76 | ASV | Actinobacteriota | Coriobacteriaceae | Collinsella | aerofaciens |
| 77 | ASV | Firmicutes | Oscillospiraceae | NA | NA |
| 78 | ASV | Proteobacteria | Pasteurellaceae | Haemophilus | NA |
| 79 | ASV | Actinobacteriota | Coriobacteriales Incertae Sedis | NA | NA |
| 80 | ASV | Firmicutes | NA | NA | NA |
| 81 | ASV | Bacteroidota | Prevotellaceae | Paraprevotella | NA |
| 82 | ASV | Firmicutes | NA | NA | NA |
| 83 | ASV | Firmicutes | Lachnospiraceae | Dorea | longicatena |
| 84 | ASV | Firmicutes | Ruminococcaceae | Ruminococcus | NA |
| 85 | ASV | Firmicutes | Ruminococcaceae | Ruminococcus | NA |
| 86 | ASV | Firmicutes | NA | NA | NA |

| Ticker | DESeq2_baseMean | DESeq2_log2-FoldChange | DESeq2_lfcSE | DESeq2_stat | DESeq2_pvalue | DESeq2_padj | DESeq2 converged | ANCOM_W | ANCOM_detected_0.7 |
|---|---|---|---|---|---|---|---|---|---|
| 57 | 500,2723708 | 0,317626941 | 0,402237775 | 0,789649706 | 0,429732371 | 0,99894472 | YES | 0 | FALSE |
| 58 | 37,1261767 | -0,052354247 | 0,902207874 | -0,058029029 | 0,953725505 | 0,99894472 | YES | 0 | FALSE |
| 59 | 51,64233017 | 0,090318943 | 0,721966447 | 0,125101302 | 0,900443352 | 0,99894472 | YES | 0 | FALSE |
| 60 | 66,12943999 | -0,606315978 | 0,909750202 | -0,66646424 | 0,505114413 | 0,99894472 | YES | 0 | FALSE |
| 61 | 19,28836149 | 0,282405106 | 2,901955184 | 0,097315461 | 0,922475878 | 0,99894472 | YES | 0 | FALSE |
| 62 | 20,03721474 | 1,740703906 | 2,903715792 | 0,599474615 | 0,548856433 | 0,99894472 | YES | 0 | FALSE |
| 63 | 29,01572216 | -0,90317505 | 2,903197727 | -0,311096637 | 0,755727158 | 0,99894472 | YES | 0 | FALSE |
| 64 | 18,45072649 | 0,195382277 | 2,903310962 | 0,067296366 | 0,94634577 | 0,99894472 | YES | 0 | FALSE |
| 65 | 61,57773797 | -0,426868365 | 0,438181398 | -0,974181849 | 0,329966255 | 0,99894472 | YES | 0 | FALSE |
| 66 | 60,2889301 | -0,231465178 | 0,48702422 | -0,475264205 | 0,634598669 | 0,99894472 | YES | 0 | FALSE |
| 67 | 53,37439601 | -0,303812067 | 0,842141792 | -0,36076118 | 0,718277985 | 0,99894472 | YES | 0 | FALSE |
| 68 | 37,6955578 | 1,449355319 | 0,670180483 | 2,162634329 | 0,030569316 | 0,980433672 | YES | 0 | FALSE |
| 69 | 108,8714557 | -0,968585486 | 2,900725838 | -0,333911421 | 0,738446401 | 0,99894472 | YES | 0 | FALSE |
| 70 | 16,25948563 | 0,061183234 | 2,901169439 | 0,021089163 | 0,98317453 | 0,99894472 | YES | 0 | FALSE |
| 71 | 172,3096971 | 0,152550275 | 0,43396722 | 0,351524879 | 0,725194611 | 0,99894472 | YES | 0 | FALSE |
| 72 | 21,94634572 | 0,035425942 | 2,905171956 | 0,012194095 | 0,990270761 | 0,99894472 | YES | 0 | FALSE |
| 73 | 38,10329659 | 0,159608241 | 0,436792294 | 0,365409929 | 0,71480543 | 0,99894472 | YES | 0 | FALSE |
| 74 | 58,01814875 | -0,456126798 | 0,575178046 | -0,793018442 | 0,427767078 | 0,99894472 | YES | 0 | FALSE |
| 75 | 41,58625889 | 0,010271639 | 2,90551085 | 0,003535227 | 0,997179303 | 0,99894472 | YES | 0 | FALSE |
| 76 | 207,6161908 | -0,158750452 | 0,275265568 | -0,576717434 | 0,564130353 | 0,99894472 | YES | 0 | FALSE |
| 77 | 15,5768961 | 0,273680748 | 2,904048113 | 0,094241121 | 0,924917621 | 0,99894472 | YES | 0 | FALSE |
| 78 | 12,64520864 | -1,555283502 | 2,021121704 | -0,769515017 | 0,441587633 | 0,99894472 | YES | 0 | FALSE |
| 79 | 2,991697807 | 0,179725446 | 2,905379545 | 0,061859541 | 0,950674687 | 0,99894472 | YES | 0 | FALSE |
| 80 | 21,55097999 | 0,049332516 | 2,900948314 | 0,017005651 | 0,986432107 | 0,99894472 | YES | 0 | FALSE |
| 81 | 37,41244117 | 0,129948349 | 2,902935675 | 0,04476446 | 0,964295053 | 0,99894472 | YES | 0 | FALSE |
| 82 | 29,63800594 | 0,071421584 | 2,902649114 | 0,024605655 | 0,980369508 | 0,99894472 | YES | 0 | FALSE |
| 83 | 612,0135466 | -0,181336246 | 0,272876327 | -0,664536379 | 0,506347073 | 0,99894472 | YES | 0 | FALSE |
| 84 | 57,87216551 | -2,254656503 | 2,902604036 | -0,776770264 | 0,437294324 | 0,99894472 | YES | 0 | FALSE |
| 85 | 88,29850228 | -0,446709999 | 2,902528116 | -0,153903763 | 0,877685617 | 0,99894472 | YES | 0 | FALSE |
| 86 | 84,18084758 | 0,258772986 | 2,902145083 | 0,08916611 | 0,928949899 | 0,99894472 | YES | 0 | FALSE |

| Ticker | taxLevel | Phylum | Family | Genus | Species |
|---|---|---|---|---|---|
| 87 | ASV | Firmicutes | Lachnospiraceae | [Ruminococcus] torques group | NA |
| 88 | ASV | Firmicutes | NA | NA | NA |
| 89 | ASV | Firmicutes | Ruminococcaceae | Ruminococcus | champanellensis |
| 90 | ASV | Firmicutes | Erysipelatoclostridiaceae | Erysipelotrichaceae UCG-003 | bacterium |
| 91 | ASV | Firmicutes | Ruminococcaceae | UBA1819 | NA |
| 92 | ASV | Firmicutes | Lachnospiraceae | Lachnospiraceae NK4A136 group | NA |
| 93 | ASV | Bacteroidota | Marinifilaceae | Odoribacter | splanchnicus |
| 94 | ASV | Actinobacteriota | Eggerthellaceae | Adlercreutzia | equolifaciens |
| 95 | ASV | Bacteroidota | Bacteroidaceae | Bacteroides | intestinalis |
| 96 | ASV | Bacteroidota | Barnesiellaceae | Barnesiella | intestinihominis |
| 97 | ASV | Firmicutes | Ruminococcaceae | Ruminococcus | NA |
| 98 | ASV | Bacteroidota | Bacteroidaceae | Bacteroides | eggerthii |
| 99 | ASV | Bacteroidota | Bacteroidaceae | Bacteroides | vulgatus |
| 100 | ASV | Bacteroidota | Prevotellaceae | Prevotella | copri |
| 101 | ASV | Firmicutes | Lachnospiraceae | [Eubacterium] xylanophilum group | NA |
| 102 | ASV | Firmicutes | Oscillospiraceae | UCG-002 | NA |
| 103 | ASV | Firmicutes | Lachnospiraceae | [Eubacterium] hallii group | NA |
| 104 | ASV | Firmicutes | NA | NA | NA |
| 105 | ASV | Firmicutes | NA | NA | NA |
| 106 | ASV | Firmicutes | Oscillospiraceae | Intestinimonas | NA |
| 107 | ASV | Firmicutes | Lachnospiraceae | Blautia | NA |
| 108 | ASV | Bacteroidota | Prevotellaceae | Prevotella | NA |
| 109 | ASV | Bacteroidota | Tannerellaceae | NA | NA |
| 110 | ASV | Firmicutes | Ruminococcaceae | Faecalibacterium | prausnitzii |
| 111 | ASV | Firmicutes | Lachnospiraceae | Lachnospiraceae FCS020 group | NA |
| 112 | ASV | Firmicutes | NA | NA | NA |
| 113 | ASV | Firmicutes | Lachnospiraceae | Lachnospiraceae UCG-010 | NA |
| 114 | ASV | Firmicutes | Oscillospiraceae | UCG-002 | NA |
| 115 | ASV | Firmicutes | Lachnospiraceae | Lachnospira | pectinoschiza |
| 116 | ASV | Firmicutes | Peptococcaceae | Peptococcus | NA |

| Ticker | DESeq2_baseMean | DESeq2_log2-FoldChange | DESeq2_lfcSE | DESeq2_stat | DESeq2_pvalue | DESeq2_padj | DESeq2 converged | ANCOM_W | ANCOM_detected_0.7 |
|---|---|---|---|---|---|---|---|---|---|
| 87 | 26,05244729 | 2,082832058 | 2,904851071 | 0,717018534 | 0,47336266 | 0,99894472 | YES | 0 | FALSE |
| 88 | 27,08985775 | -0,030051993 | 2,904030426 | -0,010348374 | 0,991743339 | 0,99894472 | YES | 0 | FALSE |
| 89 | 76,71281636 | -0,662062192 | 2,905145571 | -0,227892949 | 0,819729464 | 0,99894472 | YES | 0 | FALSE |
| 90 | 171,7659937 | -0,214886554 | 0,275598389 | -0,779709034 | 0,43556216 | 0,99894472 | YES | 0 | FALSE |
| 91 | 26,24405822 | -0,032456746 | 0,794631221 | -0,040845043 | 0,96741943 | 0,99894472 | YES | 0 | FALSE |
| 92 | 19,73124026 | 0,430061975 | 2,90337117 | 0,148125041 | 0,882244087 | 0,99894472 | YES | 2 | FALSE |
| 93 | 31,05010717 | -0,021841021 | 0,257907322 | -0,08468554 | 0,932511392 | 0,99894472 | YES | 0 | FALSE |
| 94 | 8,784474931 | 0,031020996 | 1,763959792 | 0,017586 | 0,985969126 | 0,99894472 | YES | 0 | FALSE |
| 95 | 58,01666222 | 1,469260751 | 2,904772581 | 0,505809219 | 0,612990586 | 0,99894472 | YES | 2 | FALSE |
| 96 | 60,04490399 | 0,07970368 | 2,902925451 | 0,02745633 | 0,97809577 | 0,99894472 | YES | 0 | FALSE |
| 97 | 45,20814381 | 1,613658093 | 2,902943514 | 0,555869615 | 0,578299991 | 0,99894472 | YES | 0 | FALSE |
| 98 | 96,40091483 | 0,100516266 | 2,904758013 | 0,034604007 | 0,972395506 | 0,99894472 | YES | 0 | FALSE |
| 99 | 1159,525526 | 0,048054774 | 0,211090009 | 0,22765063 | 0,819917856 | 0,99894472 | YES | 0 | FALSE |
| 100 | 453,818211 | -0,517309539 | 1,409924996 | -0,366905715 | 0,713689349 | 0,99894472 | YES | 0 | FALSE |
| 101 | 29,3648638 | -0,018670075 | 1,046377953 | -0,017842573 | 0,985764442 | 0,99894472 | YES | 0 | FALSE |
| 102 | 31,71809199 | 9,739654604 | 2,903153752 | 3,354853183 | 0,000794071 | 0,175893048 | YES | 0 | FALSE |
| 103 | 14,42897349 | 0,211533681 | 2,902679758 | 0,072875308 | 0,941905343 | 0,99894472 | YES | 0 | FALSE |
| 104 | 35,48121853 | -4,529956524 | 2,902336963 | -1,560796207 | 0,118571843 | 0,99894472 | YES | 0 | FALSE |
| 105 | 47,05692284 | 0,019474869 | 2,90291298 | 0,006708733 | 0,994647245 | 0,99894472 | YES | 0 | FALSE |
| 106 | 31,19176193 | 0,29259525 | 2,572911222 | 0,113721472 | 0,909458591 | 0,99894472 | YES | 0 | FALSE |
| 107 | 46,96817061 | 0,322658699 | 2,901155363 | 0,111217311 | 0,911444024 | 0,99894472 | YES | 0 | FALSE |
| 108 | 16,73438973 | 0,178307477 | 2,905635499 | 0,061366086 | 0,951067661 | 0,99894472 | YES | 0 | FALSE |
| 109 | 55,76378301 | -0,054015135 | 2,612393108 | -0,020676496 | 0,983503719 | 0,99894472 | YES | 0 | FALSE |
| 110 | 329,5330742 | -1,26131262 | 0,442579328 | -2,84991309 | 0,004373118 | 0,224668921 | YES | 352 | TRUE |
| 111 | 40,95303873 | -0,271382582 | 0,540331591 | -0,502251926 | 0,615490321 | 0,99894472 | YES | 0 | FALSE |
| 112 | 21,48342194 | 0,553258067 | 2,902263007 | 0,190629886 | 0,848815574 | 0,99894472 | YES | 148 | FALSE |
| 113 | 37,53411887 | 0,043868548 | 2,111722289 | 0,020773825 | 0,983426078 | 0,99894472 | YES | 0 | FALSE |
| 114 | 130,7202466 | 1,394512221 | 2,902537438 | 0,480445903 | 0,630910361 | 0,99894472 | YES | 0 | FALSE |
| 115 | 28,20609117 | -0,002387499 | 1,009416676 | -0,002365226 | 0,998112824 | 0,99894472 | YES | 0 | FALSE |
| 116 | 54,36812989 | 0,235127045 | 2,903688965 | 0,080975286 | 0,935461607 | 0,99894472 | YES | 0 | FALSE |

| Ticker | taxLevel | Phylum | Family | Genus | Species |
|---|---|---|---|---|---|
| 117 | ASV | Firmicutes | Anaerovoracaceae | Family XIII AD3011 group | NA |
| 118 | ASV | Firmicutes | Lachnospiraceae | Lachnospiraceae UCG-004 | NA |
| 119 | ASV | Bacteroidota | Bacteroidaceae | Bacteroides | uniformis |
| 120 | ASV | Firmicutes | Oscillospiraceae | Intestinimonas | NA |
| 121 | ASV | Firmicutes | Oscillospiraceae | NK4A214 group | NA |
| 122 | ASV | Bacteroidota | Barnesiellaceae | Barnesiella | NA |
| 123 | ASV | Bacteroidota | Muribaculaceae | NA | NA |
| 124 | ASV | Firmicutes | Lachnospiraceae | Lachnospiraceae UCG-001 | NA |
| 125 | ASV | Firmicutes | Oscillospiraceae | Flavonifractor | plautii |
| 126 | ASV | Firmicutes | Streptococcaceae | Streptococcus | NA |
| 127 | ASV | Firmicutes | Ruminococcaceae | [Eubacterium] siraeum group | NA |
| 128 | ASV | Proteobacteria | NA | NA | NA |
| 129 | ASV | Firmicutes | Acholeplasmataceae | Anaeroplasma | NA |
| 130 | ASV | Firmicutes | [Eubacterium] coprostanoligenes group | NA | NA |
| 131 | ASV | Cyanobacteria | NA | NA | NA |
| 132 | ASV | Bacteroidota | Bacteroidaceae | Bacteroides | clarus |
| 133 | ASV | Firmicutes | Butyricicoccaceae | NA | NA |
| 134 | ASV | Bacteroidota | Bacteroidaceae | Bacteroides | uniformis |
| 135 | ASV | Bacteroidota | Prevotellaceae | Alloprevotella | NA |
| 136 | ASV | Firmicutes | [Eubacterium] coprostanoligenes group | NA | NA |
| 137 | ASV | Firmicutes | Lachnospiraceae | Tyzzerella | NA |
| 138 | ASV | Firmicutes | NA | NA | NA |
| 139 | ASV | Bacteroidota | Bacteroidaceae | Bacteroides | coprophilus |
| 140 | ASV | Euryarchaeota | Methanobacteriaceae | Methanosphaera | NA |
| 141 | ASV | Firmicutes | Ruminococcaceae | Subdoligranulum | NA |
| 142 | ASV | Proteobacteria | NA | NA | NA |
| 143 | ASV | Actinobacteriota | Bifidobacteriaceae | Bifidobacterium | NA |
| 144 | ASV | Bacteroidota | Prevotellaceae | Paraprevotella | NA |
| 145 | ASV | Firmicutes | Oscillospiraceae | NK4A214 group | NA |
| 146 | ASV | Firmicutes | Erysipelotrichaceae | Solobacterium | NA |

| Ticker | DESeq2_baseMean | DESeq2_log2-FoldChange | DESeq2_lfcSE | DESeq2_stat | DESeq2_pvalue | DESeq2_padj | DESeq2 converged | ANCOM_W | ANCOM_detected_0.7 |
|---|---|---|---|---|---|---|---|---|---|
| 117 | 8,044960964 | -0,302090811 | 1,184137073 | -0,255114731 | 0,798634487 | 0,99894472 | YES | 0 | FALSE |
| 118 | 29,34100099 | -0,450849055 | 0,786368606 | -0,573330435 | 0,566420982 | 0,99894472 | YES | 0 | FALSE |
| 119 | 381,6499484 | -0,034196257 | 0,34333081 | -0,09960148 | 0,920660719 | 0,99894472 | YES | 0 | FALSE |
| 120 | 34,09495391 | -0,168510768 | 0,441922729 | -0,381312743 | 0,702971199 | 0,99894472 | YES | 0 | FALSE |
| 121 | 52,54840517 | 0,093930715 | 0,342527603 | 0,274228162 | 0,783909287 | 0,99894472 | YES | 0 | FALSE |
| 122 | 36,79977703 | 0,190169429 | 2,90515098 | 0,065459396 | 0,947808234 | 0,99894472 | YES | 0 | FALSE |
| 123 | 74,19388671 | 0,303741265 | 2,905523433 | 0,104539259 | 0,916741415 | 0,99894472 | YES | 5 | FALSE |
| 124 | 49,5657396 | -0,60231909 | 1,025033277 | -0,587609304 | 0,556794565 | 0,99894472 | YES | 0 | FALSE |
| 125 | 12,45080414 | -0,290282229 | 2,898986524 | -0,100132314 | 0,920239281 | 0,99894472 | YES | 0 | FALSE |
| 126 | 205,3641771 | 0,838054089 | 0,42552906 | 1,969440325 | 0,048902549 | 0,99894472 | YES | 0 | FALSE |
| 127 | 25,94039767 | -0,218488007 | 2,899895148 | -0,075343416 | 0,939941479 | 0,99894472 | YES | 0 | FALSE |
| 128 | 21,90960149 | 0,230522479 | 2,903314173 | 0,07939977 | 0,936714651 | 0,99894472 | YES | 0 | FALSE |
| 129 | 32,48546035 | 0,013399819 | 2,904510816 | 0,004613451 | 0,996319011 | 0,99894472 | YES | 0 | FALSE |
| 130 | 14,90640636 | 0,103713455 | 2,905176975 | 0,035699531 | 0,971521945 | 0,99894472 | YES | 0 | FALSE |
| 131 | 9,97741108 | -6,951023858 | 2,901333754 | -2,395802913 | 0,016584005 | 0,619638727 | YES | 0 | FALSE |
| 132 | 32,82183895 | 0,201803971 | 2,903699427 | 0,069498919 | 0,944592493 | 0,99894472 | YES | 0 | FALSE |
| 133 | 41,99365617 | 0,245636975 | 1,125361533 | 0,218273833 | 0,827215761 | 0,99894472 | YES | 0 | FALSE |
| 134 | 370,4736696 | -0,819503505 | 0,496010961 | -1,652188296 | 0,098496172 | 0,99894472 | YES | 0 | FALSE |
| 135 | 104,5073726 | 0,072466053 | 2,90399985 | 0,024953876 | 0,980091754 | 0,99894472 | YES | 0 | FALSE |
| 136 | 7,762635101 | 0,023159216 | 2,903738337 | 0,007975655 | 0,993636415 | 0,99894472 | YES | 0 | FALSE |
| 137 | 59,50731398 | 0,335125429 | 2,660535998 | 0,125961622 | 0,899762304 | 0,99894472 | YES | 0 | FALSE |
| 138 | 13,06580814 | -0,202029678 | 2,902624468 | -0,069602417 | 0,944510113 | 0,99894472 | YES | 0 | FALSE |
| 139 | 55,67549779 | 0,242649998 | 2,905521046 | 0,08351342 | 0,933443307 | 0,99894472 | YES | 0 | FALSE |
| 140 | 10,35634002 | 0,178136469 | 2,904019684 | 0,061341344 | 0,951087365 | 0,99894472 | YES | 0 | FALSE |
| 141 | 13,22637554 | 0,053298886 | 2,902618704 | 0,018362345 | 0,985349792 | 0,99894472 | YES | 0 | FALSE |
| 142 | 22,37698387 | 0,343381105 | 2,90365281 | 0,118258321 | 0,905862981 | 0,99894472 | YES | 0 | FALSE |
| 143 | 314,5076205 | 0,135245399 | 0,445518932 | 0,303568241 | 0,76145685 | 0,99894472 | YES | 0 | FALSE |
| 144 | 26,68983111 | 0,119614005 | 2,903395593 | 0,04119797 | 0,967138072 | 0,99894472 | YES | 0 | FALSE |
| 145 | 15,98302489 | 0,198243894 | 2,900995732 | 0,0683365 | 0,94551777 | 0,99894472 | YES | 0 | FALSE |
| 146 | 15,98831435 | 0,108188605 | 2,904768306 | 0,037245176 | 0,970289519 | 0,99894472 | YES | 0 | FALSE |

| Ticker | taxLevel | Phylum | Family | Genus | Species |
|---|---|---|---|---|---|
| 147 | ASV | Bacteroidota | Tannerellaceae | Parabacteroides | distasonis |
| 148 | ASV | Firmicutes | Lachnospiraceae | Lachnospira | NA |
| 149 | ASV | Actinobacteriota | Coriobacteriales Incertae Sedis | NA | NA |
| 150 | ASV | Firmicutes | Lachnospiraceae | Blautia | NA |
| 151 | ASV | Bacteroidota | Bacteroidaceae | Bacteroides | ovatus |
| 152 | ASV | Firmicutes | Lachnospiraceae | Lachnospiraceae AC2044 group | NA |
| 153 | ASV | Euryarchaeota | Methanobacteriaceae | Methanobrevibacter | NA |
| 154 | ASV | Firmicutes | Lactobacillaceae | Lactobacillus | delbrueckii |
| 155 | ASV | Firmicutes | Anaerovoracaceae | Family XIII UCG-001 | NA |
| 156 | ASV | Firmicutes | Ruminococcaceae | Incertae Sedis | NA |
| 157 | ASV | Firmicutes | Oscillospiraceae | Colidextribacter | NA |
| 158 | ASV | Firmicutes | Lachnospiraceae | Lachnoclostridium | NA |
| 159 | ASV | Firmicutes | Ruminococcaceae | Incertae Sedis | NA |
| 160 | ASV | Firmicutes | Ruminococcaceae | CAG-352 | NA |
| 161 | ASV | Firmicutes | Christensenellaceae | Christensenellaceae R-7 group | NA |
| 162 | ASV | Firmicutes | Oscillospiraceae | NK4A214 group | NA |
| 163 | ASV | Firmicutes | Erysipelotrichaceae | Catenisphaera | NA |
| 164 | ASV | Firmicutes | Oscillospiraceae | NK4A214 group | NA |
| 165 | ASV | Bacteroidota | Rikenellaceae | Alistipes | ihumii |
| 166 | ASV | Firmicutes | Clostridiaceae | Clostridium sensu stricto 1 | NA |
| 167 | ASV | Actinobacteriota | Eggerthellaceae | Slackia | isoflavoniconvertens |
| 168 | ASV | Firmicutes | Oscillospiraceae | NK4A214 group | NA |
| 169 | ASV | Firmicutes | Lachnospiraceae | NA | NA |
| 170 | ASV | Firmicutes | Christensenellaceae | Christensenellaceae R-7 group | NA |
| 171 | ASV | Firmicutes | Erysipelotrichaceae | NA | NA |
| 172 | ASV | Firmicutes | Oscillospiraceae | UCG-002 | NA |
| 173 | ASV | Firmicutes | Erysipelatoclostridiaceae | Erysipelatoclostridium | NA |
| 174 | ASV | Firmicutes | [Eubacterium] coprostanoligenes group | NA | NA |
| 175 | ASV | Firmicutes | Anaerovoracaceae | Mogibacterium | NA |
| 176 | ASV | Firmicutes | Ruminococcaceae | Negativibacillus | NA |

| Ticker | DESeq2_baseMean | DESeq2_log2-FoldChange | DESeq2_lfcSE | DESeq2_stat | DESeq2_pvalue | DESeq2_padj | DESeq2 converged | ANCOM_W | ANCOM_detected_0.7 |
|---|---|---|---|---|---|---|---|---|---|
| 147 | 24,33668341 | -0,084493616 | 2,90408372 | -0,029094759 | 0,976789016 | 0,99894472 | YES | 0 | FALSE |
| 148 | 28,4383693 | -0,109996601 | 2,901145425 | -0,037914887 | 0,969755543 | 0,99894472 | YES | 0 | FALSE |
| 149 | 6,394636888 | 0,438764413 | 2,903715706 | 0,151104467 | 0,879893309 | 0,99894472 | YES | 1 | FALSE |
| 150 | 8,884013475 | 0,53604282 | 2,900307466 | 0,184822756 | 0,853368058 | 0,99894472 | YES | 0 | FALSE |
| 151 | 16,17825249 | -0,183078632 | 2,904758999 | -0,063027133 | 0,949744898 | 0,99894472 | YES | 0 | FALSE |
| 152 | 55,85242367 | 0,093828102 | 1,764726323 | 0,053168642 | 0,95759754 | 0,99894472 | YES | 0 | FALSE |
| 153 | 187,2991034 | 0,627259221 | 0,368900424 | 1,700348331 | 0,089065425 | 0,99894472 | YES | 0 | FALSE |
| 154 | 14,18298157 | -0,005278122 | 2,90522679 | -0,001816768 | 0,99855043 | 0,99894472 | YES | 0 | FALSE |
| 155 | 9,902869365 | 0,304514502 | 2,902923001 | 0,104899269 | 0,916455739 | 0,99894472 | YES | 0 | FALSE |
| 156 | 11,2416058 | 0,277267358 | 2,901999378 | 0,095543562 | 0,923883091 | 0,99894472 | YES | 0 | FALSE |
| 157 | 24,18771829 | 1,690604873 | 0,557669545 | 3,031553161 | 0,002432991 | 0,224668921 | YES | 23 | FALSE |
| 158 | 21,37708722 | -0,492274037 | 0,859521601 | -0,572730268 | 0,56682734 | 0,99894472 | YES | 0 | FALSE |
| 159 | 13,17567418 | 0,521413274 | 1,86312809 | 0,279859059 | 0,779585639 | 0,99894472 | YES | 1 | FALSE |
| 160 | 301,6970288 | -0,968286738 | 2,90029961 | -0,333857487 | 0,738487102 | 0,99894472 | YES | 0 | FALSE |
| 161 | 10,62271582 | 0,276731651 | 2,440147319 | 0,113407764 | 0,909707285 | 0,99894472 | YES | 0 | FALSE |
| 162 | 34,33950651 | 0,272156998 | 2,901789563 | 0,093789364 | 0,92527648 | 0,99894472 | YES | 0 | FALSE |
| 163 | 3,549088477 | 0,173025079 | 2,905324979 | 0,059554467 | 0,952510484 | 0,99894472 | YES | 0 | FALSE |
| 164 | 33,3035992 | 2,33356518 | 2,904435912 | 0,803448673 | 0,421715452 | 0,99894472 | YES | 0 | FALSE |
| 165 | 12,88364389 | -0,009849477 | 2,900762179 | -0,003395479 | 0,997290805 | 0,99894472 | YES | 0 | FALSE |
| 166 | 60,47003898 | 0,438991111 | 0,632429554 | 0,694134403 | 0,487597925 | 0,99894472 | YES | 0 | FALSE |
| 167 | 9,822575183 | -0,27448307 | 0,936873957 | -0,292977585 | 0,769539285 | 0,99894472 | YES | 0 | FALSE |
| 168 | 6,375368608 | -0,086490262 | 2,904304043 | -0,02978003 | 0,976242485 | 0,99894472 | YES | 0 | FALSE |
| 169 | 29,60002579 | -0,074984582 | 2,902167903 | -0,025837438 | 0,979387001 | 0,99894472 | YES | 0 | FALSE |
| 170 | 28,75714711 | -0,397846874 | 0,912940753 | -0,435786082 | 0,662991943 | 0,99894472 | YES | 17 | FALSE |
| 171 | 14,75712244 | 0,503737165 | 2,791618827 | 0,180446256 | 0,856802243 | 0,99894472 | YES | 0 | FALSE |
| 172 | 625,5412024 | 0,092207141 | 0,39937772 | 0,230877028 | 0,817410338 | 0,99894472 | YES | 0 | FALSE |
| 173 | 2,397194588 | 0,139584502 | 2,903799474 | 0,048069608 | 0,961660768 | 0,99894472 | YES | 0 | FALSE |
| 174 | 32,88496859 | 0,198811906 | 2,904772743 | 0,068443188 | 0,945432843 | 0,99894472 | YES | 0 | FALSE |
| 175 | 25,74168808 | 0,179513378 | 2,903671629 | 0,061822892 | 0,950703873 | 0,99894472 | YES | 0 | FALSE |
| 176 | 20,88080254 | 0,540413725 | 1,568000981 | 0,344651395 | 0,730356463 | 0,99894472 | YES | 0 | FALSE |

| Ticker | taxLevel | Phylum | Family | Genus | Species |
|---|---|---|---|---|---|
| 177 | ASV | Proteobacteria | Sutterellaceae | Sutterella | NA |
| 178 | ASV | Firmicutes | Streptococcaceae | Lactococcus | NA |
| 179 | ASV | Firmicutes | Ruminococcaceae | Subdoligranulum | NA |
| 180 | ASV | Proteobacteria | Enterobacteriaceae | Escherichia-Shigella | NA |
| 181 | ASV | Firmicutes | Lachnospiraceae | GCA-900066575 | NA |
| 182 | ASV | Firmicutes | Lachnospiraceae | Lachnospiraceae UCG-004 | NA |
| 183 | ASV | Firmicutes | Veillonellaceae | Veillonella | NA |
| 184 | ASV | Firmicutes | Lachnospiraceae | Lachnoclostridium | NA |
| 185 | ASV | Firmicutes | Christensenellaceae | NA | NA |
| 186 | ASV | Firmicutes | Oscillospiraceae | Colidextribacter | massiliensis |
| 187 | ASV | Bacteroidota | Rikenellaceae | Alistipes | putredinis |
| 188 | ASV | Firmicutes | Lachnospiraceae | NA | NA |
| 189 | ASV | Firmicutes | Ruminococcaceae | Subdoligranulum | NA |
| 190 | ASV | Firmicutes | Anaerovoracaceae | Family XIII UCG-001 | NA |
| 191 | ASV | Firmicutes | NA | NA | NA |
| 192 | ASV | Firmicutes | Oscillospiraceae | UCG-002 | NA |
| 193 | ASV | Firmicutes | Christensenellaceae | Christensenellaceae R-7 group | NA |
| 194 | ASV | Firmicutes | Anaerovoracaceae | Family XIII AD3011 group | NA |
| 195 | ASV | Desulfobacterota | Desulfovibrionaceae | Desulfovibrio | piger |
| 196 | ASV | Proteobacteria | NA | NA | NA |
| 197 | ASV | Proteobacteria | NA | NA | NA |
| 198 | ASV | Firmicutes | Anaerovoracaceae | [Eubacterium] brachy group | NA |
| 199 | ASV | Firmicutes | Lachnospiraceae | NA | NA |
| 200 | ASV | Firmicutes | Peptostreptococcaceae | Intestinibacter | bartlettii |
| 201 | ASV | Firmicutes | Lachnospiraceae | [Eubacterium] ventriosum group | NA |
| 202 | ASV | Bacteroidota | Barnesiellaceae | NA | NA |
| 203 | ASV | Firmicutes | Lachnospiraceae | Lachnoclostridium | NA |
| 204 | ASV | Firmicutes | Lachnospiraceae | Lachnotalea | NA |
| 205 | ASV | Firmicutes | Monoglobaceae | Monoglobus | pectinilyticus |
| 206 | ASV | Firmicutes | Lachnospiraceae | Anaerostipes | hadrus |

| Ticker | DESeq2_baseMean | DESeq2_log2-FoldChange | DESeq2_lfcSE | DESeq2_stat | DESeq2_pvalue | DESeq2_padj | DESeq2 converged | ANCOM_W | ANCOM_detected_0.7 |
|---|---|---|---|---|---|---|---|---|---|
| 177 | 16,90005335 | -0,100609273 | 2,90346772 | -0,034651418 | 0,972357701 | 0,99894472 | YES | 0 | FALSE |
| 178 | 7,317008837 | 0,550663687 | 2,90368141 | 0,189643287 | 0,849588665 | 0,99894472 | YES | 0 | FALSE |
| 179 | 488,2169575 | 0,398916829 | 0,282238447 | 1,413403569 | 0,157537097 | 0,99894472 | NO | 0 | FALSE |
| 180 | 646,9240633 | -0,490320976 | 0,50985903 | -0,961679498 | 0,336210624 | 0,99894472 | YES | 0 | FALSE |
| 181 | 8,981213672 | -0,193721351 | 1,566364497 | -0,12367578 | 0,901571989 | 0,99894472 | YES | 0 | FALSE |
| 182 | 12,3174562 | -0,454407602 | 2,899955645 | -0,156694673 | 0,875485486 | 0,99894472 | YES | 0 | FALSE |
| 183 | 18,53531193 | 0,534603826 | 2,902819598 | 0,184167086 | 0,853882379 | 0,99894472 | YES | 0 | FALSE |
| 184 | 5,512274824 | 0,374262106 | 2,903511895 | 0,128899801 | 0,897436934 | 0,99894472 | YES | 0 | FALSE |
| 185 | 11,68386395 | 0,197118637 | 2,903703493 | 0,06788525 | 0,945876981 | 0,99894472 | YES | 0 | FALSE |
| 186 | 26,08398385 | -0,726436462 | 0,592615982 | -1,225813147 | 0,220268999 | 0,99894472 | YES | 18 | FALSE |
| 187 | 322,2845272 | -0,245846333 | 0,360890042 | -0,681222267 | 0,49573086 | 0,99894472 | YES | 0 | FALSE |
| 188 | 11,38247968 | -0,431947789 | 2,90452224 | -0,148715608 | 0,881778045 | 0,99894472 | YES | 157 | FALSE |
| 189 | 23,41177155 | 0,192932493 | 2,900856204 | 0,06650881 | 0,946972744 | 0,99894472 | YES | 0 | FALSE |
| 190 | 14,13608779 | -0,151340097 | 1,413883035 | -0,107038626 | 0,914758337 | 0,99894472 | YES | 0 | FALSE |
| 191 | 10,4207477 | 0,185048931 | 2,901500932 | 0,063776968 | 0,949147818 | 0,99894472 | YES | 0 | FALSE |
| 192 | 616,792096 | -0,43925717 | 0,31259113 | -1,40521316 | 0,159957902 | 0,99894472 | YES | 0 | FALSE |
| 193 | 23,32683817 | 0,518295524 | 0,688357531 | 0,752945236 | 0,451482824 | 0,99894472 | YES | 0 | FALSE |
| 194 | 27,13864507 | -0,03315378 | 0,521108977 | -0,063621587 | 0,949271543 | 0,99894472 | YES | 0 | FALSE |
| 195 | 35,80719401 | 0,007657753 | 2,905516734 | 0,002635591 | 0,997897105 | 0,99894472 | YES | 0 | FALSE |
| 196 | 11,12584357 | 0,078702314 | 2,903149093 | 0,027109291 | 0,978372564 | 0,99894472 | YES | 0 | FALSE |
| 197 | 11,49770266 | 0,011605945 | 2,902113853 | 0,003999135 | 0,99680916 | 0,99894472 | YES | 0 | FALSE |
| 198 | 1,144020885 | 0,414152134 | 2,90528539 | 0,142551274 | 0,886644582 | 0,99894472 | YES | 5 | FALSE |
| 199 | 5,065534029 | -0,129269192 | 2,901527862 | -0,044552111 | 0,964464315 | 0,99894472 | YES | 0 | FALSE |
| 200 | 34,94531112 | 0,440071205 | 0,669243958 | 0,657564704 | 0,510817881 | 0,99894472 | YES | 0 | FALSE |
| 201 | 13,96035869 | -0,156882059 | 2,90203185 | -0,054059386 | 0,95688785 | 0,99894472 | YES | 0 | FALSE |
| 202 | 24,57831886 | 0,129484385 | 1,506353122 | 0,085958852 | 0,931499127 | 0,99894472 | YES | 0 | FALSE |
| 203 | 15,20955995 | -0,076052609 | 0,880432033 | -0,086381011 | 0,931163542 | 0,99894472 | YES | 0 | FALSE |
| 204 | 4,808606648 | -0,448859059 | 2,901820943 | -0,154681859 | 0,877072132 | 0,99894472 | YES | 5 | FALSE |
| 205 | 9,728213387 | -0,16640598 | 2,904850722 | -0,057285553 | 0,954317728 | 0,99894472 | YES | 0 | FALSE |
| 206 | 135,9333701 | -0,193643644 | 0,357242489 | -0,542050986 | 0,587783382 | 0,99894472 | NO | 0 | FALSE |

| Ticker | taxLevel | Phylum | Family | Genus | Species |
|---|---|---|---|---|---|
| 207 | ASV | Firmicutes | Christensenellaceae | Christensenellaceae R-7 group | NA |
| 208 | ASV | Firmicutes | Lachnospiraceae | NA | NA |
| 209 | ASV | Bacteroidota | Tannerellaceae | Parabacteroides | NA |
| 210 | ASV | Firmicutes | Lachnospiraceae | Agathobacter | NA |
| 211 | ASV | Firmicutes | Lachnospiraceae | Roseburia | NA |
| 212 | ASV | Firmicutes | Lachnospiraceae | [Eubacterium] xylanophilum group | NA |
| 213 | ASV | Firmicutes | Ruminococcaceae | Negativibacillus | NA |
| 214 | ASV | Firmicutes | NA | NA | NA |
| 215 | ASV | Firmicutes | Lachnospiraceae | Marvinbryantia | NA |
| 216 | ASV | Firmicutes | Lachnospiraceae | Blautia | obeum |
| 217 | ASV | Firmicutes | [Eubacterium] coprostanoligenes group | NA | NA |
| 218 | ASV | Firmicutes | Ruminococcaceae | NA | NA |
| 219 | ASV | Bacteroidota | Rikenellaceae | Alistipes | NA |
| 220 | ASV | Bacteroidota | Rikenellaceae | Alistipes | NA |
| 221 | ASV | Firmicutes | Lachnospiraceae | [Eubacterium] hallii group | NA |
| 222 | ASV | Firmicutes | UCG-010 | NA | NA |
| 223 | ASV | Firmicutes | NA | NA | NA |
| 224 | ASV | Bacteroidota | Barnesiellaceae | Coprobacter | fastidiosus |
| 225 | ASV | Firmicutes | Clostridiaceae | Clostridium sensu stricto 1 | NA |
| 226 | ASV | Firmicutes | Christensenellaceae | Christensenellaceae R-7 group | NA |
| 227 | ASV | Firmicutes | NA | NA | NA |
| 228 | ASV | Firmicutes | NA | NA | NA |
| 229 | ASV | Firmicutes | Selenomonadaceae | Mitsuokella | jalaludinii |
| 230 | ASV | Firmicutes | Ruminococcaceae | DTU089 | NA |
| 231 | ASV | Firmicutes | Lachnospiraceae | NA | NA |
| 232 | ASV | Bacteroidota | Tannerellaceae | Parabacteroides | merdae |
| 233 | ASV | Bacteroidota | Rikenellaceae | Alistipes | indistinctus |
| 234 | ASV | Firmicutes | Ruminococcaceae | Ruminococcus | bromii |
| 235 | ASV | Firmicutes | Erysipelotrichaceae | Holdemanella | NA |
| 236 | ASV | Bacteroidota | Rikenellaceae | Rikenellaceae RC9 gut group | NA |

| Ticker | DESeq2_baseMean | DESeq2_log2-FoldChange | DESeq2_lfcSE | DESeq2_stat | DESeq2_pvalue | DESeq2_padj | DESeq2 converged | ANCOM_W | ANCOM_detected_0.7 |
|---|---|---|---|---|---|---|---|---|---|
| 207 | 8,747838125 | 0,229868848 | 2,901709887 | 0,079218411 | 0,9368589 | 0,99894472 | YES | 0 | FALSE |
| 208 | 8,457088352 | 0,042010438 | 2,904189157 | 0,014465462 | 0,988458634 | 0,99894472 | YES | 0 | FALSE |
| 209 | 27,45099236 | 0,200595679 | 2,905897027 | 0,069030553 | 0,9449653 | 0,99894472 | YES | 0 | FALSE |
| 210 | 348,4599683 | -0,404276033 | 0,814521134 | -0,496335842 | 0,619657482 | 0,99894472 | YES | 0 | FALSE |
| 211 | 20,84532238 | -0,182336086 | 2,901119493 | -0,06285025 | 0,949885751 | 0,99894472 | YES | 0 | FALSE |
| 212 | 13,70410935 | 0,174944749 | 2,90052241 | 0,06031491 | 0,951904827 | 0,99894472 | YES | 0 | FALSE |
| 213 | 11,33772624 | 0,007615992 | 2,902077612 | 0,002624324 | 0,997906095 | 0,99894472 | YES | 0 | FALSE |
| 214 | 23,11357952 | -6,059498108 | 2,905313763 | -2,085660484 | 0,037009388 | 0,99894472 | YES | 0 | FALSE |
| 215 | 37,41460798 | 0,953279192 | 0,718439544 | 1,326874614 | 0,184550163 | 0,99894472 | YES | 0 | FALSE |
| 216 | 214,0964186 | -0,455205896 | 0,348360609 | -1,306708863 | 0,191311607 | 0,99894472 | YES | 0 | FALSE |
| 217 | 16,07726368 | 0,143141973 | 2,90550866 | 0,049265719 | 0,960707539 | 0,99894472 | YES | 0 | FALSE |
| 218 | 14,09617392 | -0,208859468 | 0,548846304 | -0,380542724 | 0,703542588 | 0,99894472 | YES | 0 | FALSE |
| 219 | 9,007958881 | 0,466035174 | 2,901691156 | 0,160608124 | 0,872402056 | 0,99894472 | YES | 1 | FALSE |
| 220 | 14,02234981 | 0,207353168 | 2,901941854 | 0,071453247 | 0,943037033 | 0,99894472 | YES | 0 | FALSE |
| 221 | 136,4336817 | -0,156731771 | 0,258829715 | -0,605540098 | 0,54482019 | 0,99894472 | YES | 0 | FALSE |
| 222 | 11,07180546 | -0,345039416 | 2,900710894 | -0,11894995 | 0,905315008 | 0,99894472 | YES | 0 | FALSE |
| 223 | 16,80301954 | 0,126375144 | 2,904407705 | 0,043511503 | 0,965293795 | 0,99894472 | YES | 0 | FALSE |
| 224 | 8,118634009 | -0,048215407 | 1,582026661 | -0,030476988 | 0,975686646 | 0,99894472 | YES | 0 | FALSE |
| 225 | 8,627364798 | 0,068693327 | 2,901305986 | 0,023676692 | 0,981110498 | 0,99894472 | YES | 0 | FALSE |
| 226 | 2,659951872 | 0,288819398 | 2,902109815 | 0,099520492 | 0,920725018 | 0,99894472 | YES | 0 | FALSE |
| 227 | 15,08492104 | 0,122310501 | 2,903706606 | 0,042122197 | 0,966401285 | 0,99894472 | YES | 0 | FALSE |
| 228 | 27,5064993 | 0,364832003 | 2,903338679 | 0,125659471 | 0,900001485 | 0,99894472 | YES | 0 | FALSE |
| 229 | 18,66242672 | 0,032714512 | 2,90516667 | 0,011260804 | 0,991015368 | 0,99894472 | YES | 0 | FALSE |
| 230 | 8,43598098 | 0,303272036 | 1,250189422 | 0,242580869 | 0,808330103 | 0,99894472 | YES | 0 | FALSE |
| 231 | 12,36412939 | -1,008194601 | 2,900893975 | -0,347546174 | 0,728181036 | 0,99894472 | YES | 312 | TRUE |
| 232 | 339,1420055 | -0,085316549 | 0,238359893 | -0,357931645 | 0,720394468 | 0,99894472 | YES | 0 | FALSE |
| 233 | 7,189307042 | 0,827565501 | 1,316705766 | 0,628512096 | 0,529668525 | 0,99894472 | YES | 0 | FALSE |
| 234 | 1047,396357 | 0,418236301 | 0,51825383 | 0,807010535 | 0,419660413 | 0,99894472 | YES | 0 | FALSE |
| 235 | 211,0214756 | 0,081629549 | 0,77114011 | 0,105855665 | 0,91569687 | 0,99894472 | YES | 0 | FALSE |
| 236 | 24,46505356 | 0,130693451 | 2,901968206 | 0,045036142 | 0,964078501 | 0,99894472 | YES | 0 | FALSE |

| Ticker | taxLevel | Phylum | Family | Genus | Species |
|---|---|---|---|---|---|
| 237 | ASV | Firmicutes | Oscillospiraceae | Colidextribacter | NA |
| 238 | ASV | Firmicutes | NA | NA | NA |
| 239 | ASV | Firmicutes | Ruminococcaceae | Ruminococcus | bicirculans |
| 240 | ASV | Firmicutes | Oscillospiraceae | Oscillibacter | NA |
| 241 | ASV | Firmicutes | Oscillospiraceae | UCG-003 | NA |
| 242 | ASV | Firmicutes | Oscillospiraceae | Oscillospira | NA |
| 243 | ASV | Firmicutes | UCG-010 | NA | NA |
| 244 | ASV | Actinobacteriota | Bifidobacteriaceae | Bifidobacterium | NA |
| 245 | ASV | Firmicutes | NA | NA | NA |
| 246 | ASV | Firmicutes | Lachnospiraceae | Lachnospiraceae FCS020 group | bacterium |
| 247 | ASV | Firmicutes | Christensenellaceae | Christensenellaceae R-7 group | NA |
| 248 | ASV | Firmicutes | Oscillospiraceae | NK4A214 group | NA |
| 249 | ASV | Firmicutes | Lactobacillaceae | Lactobacillus | NA |
| 250 | ASV | Firmicutes | Lachnospiraceae | Blautia | faecis |
| 251 | ASV | Bacteroidota | Marinifilaceae | Butyricimonas | virosa |
| 252 | ASV | Firmicutes | UCG-010 | NA | NA |
| 253 | ASV | Firmicutes | Christensenellaceae | Christensenellaceae R-7 group | NA |
| 254 | ASV | Firmicutes | Christensenellaceae | Christensenellaceae R-7 group | NA |
| 255 | ASV | Firmicutes | Christensenellaceae | Christensenellaceae R-7 group | NA |
| 256 | ASV | Firmicutes | UCG-010 | NA | NA |
| 257 | ASV | Bacteroidota | Bacteroidaceae | Bacteroides | NA |
| 258 | ASV | Firmicutes | [Eubacterium] coprostanoligenes group | NA | NA |
| 259 | ASV | Firmicutes | Veillonellaceae | Veillonella | NA |
| 260 | ASV | Firmicutes | Erysipelatoclostridiaceae | Coprobacillus | cateniformis |
| 261 | ASV | Firmicutes | Oscillospiraceae | UCG-005 | NA |
| 262 | ASV | Firmicutes | Christensenellaceae | Christensenellaceae R-7 group | NA |
| 263 | ASV | Actinobacteriota | Atopobiaceae | Olsenella | NA |
| 264 | ASV | Firmicutes | Lachnospiraceae | Coprococcus | comes |
| 265 | ASV | Firmicutes | NA | NA | NA |
| 266 | ASV | Firmicutes | Oscillospiraceae | UCG-005 | NA |

| Ticker | DESeq2_baseMean | DESeq2_log2-FoldChange | DESeq2_lfcSE | DESeq2_stat | DESeq2_pvalue | DESeq2_padj | DESeq2 converged | ANCOM_W | ANCOM_detected_0.7 |
|---|---|---|---|---|---|---|---|---|---|
| 237 | 9,271850602 | -0,832845372 | 1,191051507 | -0,699252188 | 0,484394441 | 0,99894472 | YES | 208 | FALSE |
| 238 | 9,058926154 | 0,274291909 | 0,596458773 | 0,459867339 | 0,645611445 | 0,99894472 | YES | 0 | FALSE |
| 239 | 196,4702635 | -0,285907992 | 0,706295679 | -0,404799294 | 0,685625048 | 0,99894472 | YES | 0 | FALSE |
| 240 | 9,133748374 | 0,474485377 | 2,904617769 | 0,163355531 | 0,870238514 | 0,99894472 | YES | 8 | FALSE |
| 241 | 3,501742822 | 0,112448653 | 2,904666268 | 0,038713106 | 0,969119124 | 0,99894472 | YES | 0 | FALSE |
| 242 | 10,18965444 | 0,496542347 | 2,90141747 | 0,17113785 | 0,864115375 | 0,99894472 | YES | 7 | FALSE |
| 243 | 16,24033686 | 0,430538397 | 2,900858392 | 0,148417585 | 0,882013223 | 0,99894472 | YES | 3 | FALSE |
| 244 | 46,7716175 | 1,178000406 | 2,904792266 | 0,405536885 | 0,685082912 | 0,99894472 | YES | 0 | FALSE |
| 245 | 19,91706631 | -0,77918988 | 2,902697315 | -0,26843649 | 0,788363355 | 0,99894472 | YES | 0 | FALSE |
| 246 | 4,351166029 | 0,205973108 | 2,904522757 | 0,070914613 | 0,943465713 | 0,99894472 | YES | 0 | FALSE |
| 247 | 7,312103265 | 0,185547992 | 2,488102804 | 0,074574086 | 0,940553593 | 0,99894472 | YES | 0 | FALSE |
| 248 | 21,25846324 | -0,166826608 | 2,902684097 | -0,057473222 | 0,954168237 | 0,99894472 | YES | 0 | FALSE |
| 249 | 8,257075044 | -0,48207339 | 2,904652747 | -0,165965928 | 0,868183763 | 0,99894472 | YES | 0 | FALSE |
| 250 | 105,7539111 | -0,389753391 | 0,450980199 | -0,864236152 | 0,387458176 | 0,99894472 | YES | 0 | FALSE |
| 251 | 8,260028247 | 0,200421436 | 2,90168106 | 0,069070801 | 0,944933263 | 0,99894472 | YES | 0 | FALSE |
| 252 | 12,92001934 | 0,175074273 | 2,901590662 | 0,060337344 | 0,95188696 | 0,99894472 | YES | 0 | FALSE |
| 253 | 10,95557651 | 0,038349905 | 1,825694103 | 0,021005658 | 0,983241143 | 0,99894472 | YES | 0 | FALSE |
| 254 | 5,736317313 | 0,495180701 | 2,899588195 | 0,170776217 | 0,864399731 | 0,99894472 | YES | 0 | FALSE |
| 255 | 4,824076391 | 0,218653256 | 2,901638629 | 0,075355095 | 0,939932186 | 0,99894472 | YES | 0 | FALSE |
| 256 | 12,24334777 | 0,318086399 | 1,714944301 | 0,185479143 | 0,852853238 | 0,99894472 | YES | 0 | FALSE |
| 257 | 60,78089087 | 0,022509288 | 2,90147066 | 0,007757889 | 0,993810162 | 0,99894472 | YES | 0 | FALSE |
| 258 | 9,85819851 | -0,073265725 | 2,139102572 | -0,034250683 | 0,972677251 | 0,99894472 | YES | 0 | FALSE |
| 259 | 7,709181677 | 0,083556202 | 2,903004704 | 0,028782661 | 0,977037929 | 0,99894472 | YES | 0 | FALSE |
| 260 | 7,089198105 | 0,259839954 | 2,903518599 | 0,089491403 | 0,928691385 | 0,99894472 | YES | 0 | FALSE |
| 261 | 14,02468503 | 0,058893583 | 2,902573183 | 0,020290128 | 0,983811931 | 0,99894472 | YES | 0 | FALSE |
| 262 | 14,5808246 | 0,071403522 | 2,903680286 | 0,024590697 | 0,98038144 | 0,99894472 | YES | 0 | FALSE |
| 263 | 4,349924288 | 0,023908061 | 2,903581135 | 0,008233991 | 0,9934303 | 0,99894472 | YES | 0 | FALSE |
| 264 | 119,6318567 | -0,403729513 | 0,299406936 | -1,348430726 | 0,177519887 | 0,99894472 | YES | 0 | FALSE |
| 265 | 3,918883104 | -0,139516806 | 2,904210991 | -0,048039487 | 0,961684773 | 0,99894472 | YES | 0 | FALSE |
| 266 | 13,08387694 | -0,202849155 | 2,904440883 | -0,069841034 | 0,944320186 | 0,99894472 | YES | 0 | FALSE |

| Ticker | taxLevel | Phylum | Family | Genus | Species |
|---|---|---|---|---|---|
| 267 | ASV | Verrucomicrobiota | Victivallaceae | Victivallis | NA |
| 268 | ASV | Actinobacteriota | Actinomycetaceae | Actinomyces | odontolyticus |
| 269 | ASV | Firmicutes | Lachnospiraceae | NA | NA |
| 270 | ASV | Bacteroidota | Marinifilaceae | Butyricimonas | paravirosa |
| 271 | ASV | Bacteroidota | Bacteroidaceae | Bacteroides | NA |
| 272 | ASV | Firmicutes | NA | NA | NA |
| 273 | ASV | Bacteroidota | Rikenellaceae | Alistipes | NA |
| 274 | ASV | Firmicutes | Christensenellaceae | Christensenellaceae R-7 group | NA |
| 275 | ASV | Firmicutes | Oscillospiraceae | Intestinimonas | massiliensis |
| 276 | ASV | Bacteroidota | Bacteroidaceae | Bacteroides | caccae |
| 277 | ASV | Firmicutes | UCG-010 | NA | NA |
| 278 | ASV | Firmicutes | Lachnospiraceae | [Eubacterium] xylanophilum group | NA |
| 279 | ASV | Firmicutes | Lachnospiraceae | NA | NA |
| 280 | ASV | Firmicutes | Lachnospiraceae | Howardella | ureilytica |
| 281 | ASV | Bacteroidota | Bacteroidaceae | Bacteroides | NA |
| 282 | ASV | Firmicutes | Anaerovoracaceae | Family XIII AD3011 group | NA |
| 283 | ASV | Firmicutes | Lachnospiraceae | Eisenbergiella | tayi |
| 284 | ASV | Firmicutes | Lachnospiraceae | Blautia | NA |
| 285 | ASV | Bacteroidota | Prevotellaceae | Prevotella | NA |
| 286 | ASV | Firmicutes | Erysipelatoclostridiaceae | Erysipelatoclostridium | NA |
| 287 | ASV | Firmicutes | Christensenellaceae | Christensenellaceae R-7 group | NA |
| 288 | ASV | Bacteroidota | Bacteroidaceae | Bacteroides | nordii |
| 289 | ASV | Firmicutes | Oscillospiraceae | Colidextribacter | NA |
| 290 | ASV | Firmicutes | Ruminococcaceae | NA | NA |
| 291 | ASV | Firmicutes | Lachnospiraceae | Marvinbryantia | NA |
| 292 | ASV | Firmicutes | Oscillospiraceae | UCG-005 | NA |
| 293 | ASV | Proteobacteria | Oxalobacteraceae | Oxalobacter | formigenes |
| 294 | ASV | Firmicutes | Lachnospiraceae | Blautia | NA |
| 295 | ASV | Firmicutes | Lachnospiraceae | Coprococcus | eutactus |
| 296 | ASV | Firmicutes | UCG-010 | NA | NA |

| Ticker | DESeq2_baseMean | DESeq2_log2-FoldChange | DESeq2_lfcSE | DESeq2_stat | DESeq2_pvalue | DESeq2_padj | DESeq2 converged | ANCOM_W | ANCOM_detected_0.7 |
|---|---|---|---|---|---|---|---|---|---|
| 267 | 5,186345754 | 0,146263978 | 2,901523935 | 0,050409365 | 0,959796174 | 0,99894472 | YES | 0 | FALSE |
| 268 | 0,977468038 | 0,208112623 | 2,904720633 | 0,071646347 | 0,942883355 | 0,99894472 | YES | 0 | FALSE |
| 269 | 6,921613119 | -0,212159825 | 2,904116556 | -0,073054859 | 0,941762463 | 0,99894472 | YES | 0 | FALSE |
| 270 | 5,716731909 | 0,244352169 | 2,904612546 | 0,084125564 | 0,932956599 | 0,99894472 | YES | 0 | FALSE |
| 271 | 124,2794829 | 0,262123314 | 0,42985839 | 0,609789922 | 0,542000979 | 0,99894472 | YES | 0 | FALSE |
| 272 | 4,244930043 | -0,062124964 | 2,904960376 | -0,021385821 | 0,982937884 | 0,99894472 | YES | 0 | FALSE |
| 273 | 180,269199 | -0,485047918 | 0,662516088 | -0,732130022 | 0,464089211 | 0,99894472 | YES | 0 | FALSE |
| 274 | 8,025712213 | 0,217575449 | 2,903523299 | 0,074934976 | 0,940266448 | 0,99894472 | YES | 0 | FALSE |
| 275 | 9,488323889 | 0,368646053 | 2,902117154 | 0,127026592 | 0,898919351 | 0,99894472 | YES | 0 | FALSE |
| 276 | 271,0007549 | 0,129797705 | 0,45570551 | 0,284828035 | 0,775775892 | 0,99894472 | YES | 0 | FALSE |
| 277 | 7,992442659 | 0,163187266 | 2,903462689 | 0,056204361 | 0,955179007 | 0,99894472 | YES | 0 | FALSE |
| 278 | 8,336489924 | -0,132373722 | 2,904068033 | -0,04558217 | 0,963643281 | 0,99894472 | YES | 0 | FALSE |
| 279 | 3,691944363 | -0,335062539 | 2,90241878 | -0,11544252 | 0,908094378 | 0,99894472 | YES | 0 | FALSE |
| 280 | 10,23380724 | -0,261106711 | 1,031632647 | -0,253100473 | 0,800190574 | 0,99894472 | YES | 0 | FALSE |
| 281 | 189,3186947 | -1,201056445 | 1,01659373 | -1,181451753 | 0,237423309 | 0,99894472 | YES | 0 | FALSE |
| 282 | 2,717937618 | 0,062593698 | 2,902941681 | 0,021562162 | 0,982797217 | 0,99894472 | YES | 0 | FALSE |
| 283 | 11,47156132 | -8,311942783 | 2,904976817 | -2,861276805 | 0,004219385 | 0,224668921 | YES | 0 | FALSE |
| 284 | 492,9887856 | 0,153807469 | 0,19670308 | 0,781927099 | 0,434257418 | 0,99894472 | YES | 0 | FALSE |
| 285 | 110,5712201 | -1,235210158 | 2,903334348 | -0,425445371 | 0,670512037 | 0,99894472 | YES | 0 | FALSE |
| 286 | 3,609034086 | -0,008679551 | 2,905598685 | -0,002987182 | 0,997616578 | 0,99894472 | YES | 0 | FALSE |
| 287 | 2,9824968 | -0,005273292 | 2,903870673 | -0,001815953 | 0,99855108 | 0,99894472 | YES | 0 | FALSE |
| 288 | 5,398458234 | 0,21218115 | 2,903817811 | 0,073069719 | 0,941750638 | 0,99894472 | YES | 0 | FALSE |
| 289 | 12,01855857 | 0,196632201 | 2,904220023 | 0,067705683 | 0,946019925 | 0,99894472 | YES | 0 | FALSE |
| 290 | 2,656381379 | -0,176719681 | 2,904367017 | -0,060846195 | 0,9514817 | 0,99894472 | YES | 0 | FALSE |
| 291 | 8,021765729 | 0,503683802 | 2,902333203 | 0,173544444 | 0,862223497 | 0,99894472 | YES | 0 | FALSE |
| 292 | 280,6266795 | -0,072089934 | 0,271504208 | -0,265520503 | 0,790608517 | 0,99894472 | YES | 0 | FALSE |
| 293 | 12,74891559 | 0,143005246 | 0,399238974 | 0,358194603 | 0,720197686 | 0,99894472 | YES | 0 | FALSE |
| 294 | 15,87663876 | 0,462852109 | 2,904819387 | 0,159339376 | 0,873401499 | 0,99894472 | YES | 34 | FALSE |
| 295 | 288,3266723 | 0,284180279 | 0,906884821 | 0,313358733 | 0,754008133 | 0,99894472 | YES | 0 | FALSE |
| 296 | 5,640723253 | 0,278348397 | 2,90420885 | 0,095843106 | 0,923645181 | 0,99894472 | YES | 0 | FALSE |

| Ticker | taxLevel | Phylum | Family | Genus | Species |
|---|---|---|---|---|---|
| 297 | ASV | Firmicutes | Lachnospiraceae | [Eubacterium] ventriosum group | NA |
| 298 | ASV | Firmicutes | UCG-010 | NA | NA |
| 299 | ASV | Firmicutes | Lachnospiraceae | NA | NA |
| 300 | ASV | Firmicutes | Lachnospiraceae | [Ruminococcus] torques group | NA |
| 301 | ASV | Firmicutes | Veillonellaceae | Allisonella | histaminiformans |
| 302 | ASV | Firmicutes | Ruminococcaceae | Negativibacillus | massiliensis |
| 303 | ASV | Firmicutes | Anaerovoracaceae | Family XIII UCG-001 | NA |
| 304 | ASV | Firmicutes | Erysipelatoclostridiaceae | Asteroleplasma | NA |
| 305 | ASV | Bacteroidota | Marinifilaceae | Butyricimonas | NA |
| 306 | ASV | Bacteroidota | Marinifilaceae | Butyricimonas | NA |
| 307 | ASV | Bacteroidota | Bacteroidaceae | Bacteroides | NA |
| 308 | ASV | Proteobacteria | Sutterellaceae | Sutterella | NA |
| 309 | ASV | Firmicutes | Lachnospiraceae | NA | NA |
| 310 | ASV | Firmicutes | Monoglobaceae | Monoglobus | NA |
| 311 | ASV | Firmicutes | Lachnospiraceae | NA | NA |
| 312 | ASV | Firmicutes | Oscillospiraceae | UCG-005 | NA |
| 313 | ASV | Bacteroidota | Bacteroidaceae | Bacteroides | massiliensis |
| 314 | ASV | Firmicutes | Lachnospiraceae | Roseburia | inulinivorans |
| 315 | ASV | Firmicutes | Erysipelatoclostridiaceae | Erysipelatoclostridium | NA |
| 316 | ASV | Bacteroidota | Bacteroidaceae | Bacteroides | NA |
| 317 | ASV | Bacteroidota | NA | NA | NA |
| 318 | ASV | Firmicutes | Anaerovoracaceae | Family XIII AD3011 group | NA |
| 319 | ASV | Firmicutes | [Eubacterium] coprostanoligenes group | NA | NA |
| 320 | ASV | Firmicutes | UCG-010 | NA | NA |
| 321 | ASV | Firmicutes | Lachnospiraceae | NA | NA |
| 322 | ASV | Firmicutes | Oscillospiraceae | Oscillibacter | NA |
| 323 | ASV | Firmicutes | Ruminococcaceae | Faecalibacterium | NA |
| 324 | ASV | Firmicutes | Ruminococcaceae | [Eubacterium] siraeum group | NA |
| 325 | ASV | Firmicutes | Lachnospiraceae | NA | NA |
| 326 | ASV | Firmicutes | Lachnospiraceae | NA | NA |

| Ticker | DESeq2_baseMean | DESeq2_log2-FoldChange | DESeq2_lfcSE | DESeq2_stat | DESeq2_pvalue | DESeq2_padj | DESeq2 converged | ANCOM_W | ANCOM_detected_0.7 |
|---|---|---|---|---|---|---|---|---|---|
| 297 | 4,140582892 | -0,115843011 | 2,904317685 | -0,039886481 | 0,968183629 | 0,99894472 | YES | 0 | FALSE |
| 298 | 5,605550656 | 0,625139955 | 2,899929365 | 0,215570752 | 0,829322356 | 0,99894472 | YES | 0 | FALSE |
| 299 | 4,63217094 | -0,167977467 | 2,904354266 | -0,057836425 | 0,953878924 | 0,99894472 | YES | 0 | FALSE |
| 300 | 297,6477888 | -0,572390506 | 0,354308668 | -1,615513698 | 0,106199496 | 0,99894472 | YES | 0 | FALSE |
| 301 | 9,225730262 | -0,074150709 | 2,901991553 | -0,025551663 | 0,979614941 | 0,99894472 | YES | 0 | FALSE |
| 302 | 9,123251396 | 0,412841104 | 2,903598673 | 0,142182564 | 0,886935804 | 0,99894472 | YES | 0 | FALSE |
| 303 | 5,816478612 | 0,16332708 | 2,904519402 | 0,05623205 | 0,955156949 | 0,99894472 | YES | 0 | FALSE |
| 304 | 2,585148566 | -0,137632518 | 2,903974494 | -0,047394534 | 0,962198785 | 0,99894472 | YES | 0 | FALSE |
| 305 | 14,23384685 | 0,029797685 | 2,902286465 | 0,010266969 | 0,991808288 | 0,99894472 | YES | 0 | FALSE |
| 306 | 7,872578089 | 0,092713889 | 2,902023042 | 0,03194802 | 0,974513504 | 0,99894472 | YES | 0 | FALSE |
| 307 | 140,0576948 | -0,024575253 | 0,730392888 | -0,033646622 | 0,973158945 | 0,99894472 | YES | 0 | FALSE |
| 308 | 14,15500388 | 0,17983592 | 2,905238314 | 0,061900574 | 0,95064201 | 0,99894472 | YES | 0 | FALSE |
| 309 | 10,19948611 | 0,170481963 | 2,901314159 | 0,058760256 | 0,953143065 | 0,99894472 | YES | 0 | FALSE |
| 310 | 79,94215441 | -0,032652499 | 0,429009849 | -0,076111305 | 0,939330547 | 0,99894472 | NO | 0 | FALSE |
| 311 | 8,514583524 | 0,287814232 | 2,904263923 | 0,099100577 | 0,921058414 | 0,99894472 | YES | 0 | FALSE |
| 312 | 25,39383483 | 5,097716442 | 2,905342736 | 1,754600715 | 0,079327629 | 0,99894472 | YES | 0 | FALSE |
| 313 | 185,8878633 | -0,383264753 | 0,534752678 | -0,716714041 | 0,47355056 | 0,99894472 | YES | 0 | FALSE |
| 314 | 164,1997714 | -1,189993708 | 0,587980541 | -2,023865801 | 0,042983962 | 0,99894472 | YES | 0 | FALSE |
| 315 | 3,091871824 | 0,173165333 | 2,905502743 | 0,059599095 | 0,95247494 | 0,99894472 | YES | 0 | FALSE |
| 316 | 142,3397159 | 0,244937553 | 0,576085473 | 0,425175716 | 0,670708586 | 0,99894472 | YES | 0 | FALSE |
| 317 | 3,018899993 | 0,186785673 | 2,905134979 | 0,064295007 | 0,948735329 | 0,99894472 | YES | 0 | FALSE |
| 318 | 6,842526689 | 0,368945238 | 2,90279874 | 0,127099834 | 0,898861383 | 0,99894472 | YES | 0 | FALSE |
| 319 | 139,6694157 | -0,223783034 | 1,037299491 | -0,215736185 | 0,829193394 | 0,99894472 | YES | 0 | FALSE |
| 320 | 6,344996583 | 0,259025369 | 2,903518458 | 0,089210857 | 0,928914337 | 0,99894472 | YES | 0 | FALSE |
| 321 | 5,084609489 | 0,490987661 | 2,904284941 | 0,169056298 | 0,865752359 | 0,99894472 | YES | 5 | FALSE |
| 322 | 8,485709209 | -0,119872912 | 2,901150712 | -0,041319092 | 0,967041513 | 0,99894472 | YES | 0 | FALSE |
| 323 | 819,0129138 | -0,256465534 | 0,268666854 | -0,954585689 | 0,339787257 | 0,99894472 | YES | 0 | FALSE |
| 324 | 157,1480003 | 0,60197138 | 0,531753845 | 1,132048947 | 0,257613857 | 0,99894472 | YES | 0 | FALSE |
| 325 | 14,15018687 | 0,34540997 | 2,904863302 | 0,118907478 | 0,905348657 | 0,99894472 | YES | 0 | FALSE |
| 326 | 6,128535647 | 0,104694503 | 2,904170311 | 0,036049712 | 0,97124272 | 0,99894472 | YES | 0 | FALSE |

| Ticker | taxLevel | Phylum | Family | Genus | Species |
|---|---|---|---|---|---|
| 327 | ASV | Firmicutes | Erysipelatoclostridiaceae | Catenibacterium | mitsuokai |
| 328 | ASV | Firmicutes | Lachnospiraceae | Lachnospiraceae UCG-010 | NA |
| 329 | ASV | Firmicutes | Oscillospiraceae | Oscillibacter | NA |
| 330 | ASV | Actinobacteriota | Atopobiaceae | NA | NA |
| 331 | ASV | Firmicutes | NA | NA | NA |
| 332 | ASV | Firmicutes | UCG-010 | NA | NA |
| 333 | ASV | Firmicutes | UCG-010 | NA | NA |
| 334 | ASV | Firmicutes | NA | NA | NA |
| 335 | ASV | Actinobacteriota | Eggerthellaceae | NA | NA |
| 336 | ASV | Firmicutes | Ruminococcaceae | Angelakisella | NA |
| 337 | ASV | Firmicutes | Oscillospiraceae | UCG-002 | NA |
| 338 | ASV | Firmicutes | Ruminococcaceae | Incertae Sedis | NA |
| 339 | ASV | Firmicutes | Lachnospiraceae | NA | NA |
| 340 | ASV | Firmicutes | Oscillospiraceae | Flavonifractor | NA |
| 341 | ASV | Firmicutes | Butyricicoccaceae | Butyricicoccus | NA |
| 342 | ASV | Firmicutes | Lachnospiraceae | NA | NA |
| 343 | ASV | Firmicutes | Erysipelotrichaceae | Holdemania | filiformis |
| 344 | ASV | Firmicutes | Erysipelotrichaceae | Holdemanella | biformis |
| 345 | ASV | Firmicutes | Christensenellaceae | Christensenellaceae R-7 group | NA |
| 346 | ASV | Bacteroidota | Prevotellaceae | NA | NA |
| 347 | ASV | Firmicutes | Ruminococcaceae | Faecalibacterium | NA |
| 348 | ASV | Firmicutes | Oscillospiraceae | NA | NA |
| 349 | ASV | Firmicutes | Ruminococcaceae | DTU089 | NA |
| 350 | ASV | Firmicutes | Christensenellaceae | NA | NA |
| 351 | ASV | Firmicutes | Ruminococcaceae | Fournierella | NA |
| 352 | ASV | Firmicutes | UCG-010 | NA | NA |
| 353 | ASV | Firmicutes | Lachnospiraceae | Agathobacter | NA |
| 354 | ASV | Firmicutes | Lachnospiraceae | Roseburia | intestinalis |
| 355 | ASV | Firmicutes | Lachnospiraceae | Lachnoclostridium | NA |
| 356 | ASV | Firmicutes | Hydrogenoanaerobacterium | NA | NA |

| Ticker | DESeq2_baseMean | DESeq2_log2-FoldChange | DESeq2_lfcSE | DESeq2_stat | DESeq2_pvalue | DESeq2_padj | DESeq2 converged | ANCOM_W | ANCOM_detected_0.7 |
|---|---|---|---|---|---|---|---|---|---|
| 327 | 64,3604178 | -0,597131615 | 2,90404315 | -0,205620779 | 0,837087145 | 0,99894472 | YES | 0 | FALSE |
| 328 | 4,081463827 | 0,112086769 | 2,905936996 | 0,038571645 | 0,96923191 | 0,99894472 | YES | 0 | FALSE |
| 329 | 1,09131054 | -0,011783933 | 2,9046787 | -0,00405688 | 0,996763087 | 0,99894472 | YES | 0 | FALSE |
| 330 | 1,405286744 | 0,246133029 | 2,905415876 | 0,084715249 | 0,932487773 | 0,99894472 | YES | 0 | FALSE |
| 331 | 168,985979 | 0,60304508 | 1,278133571 | 0,471816947 | 0,637057453 | 0,99894472 | YES | 0 | FALSE |
| 332 | 16,09675875 | 0,085323968 | 2,902221077 | 0,029399541 | 0,976545939 | 0,99894472 | YES | 0 | FALSE |
| 333 | 2,748017918 | 0,26607387 | 2,903707788 | 0,091632454 | 0,926990065 | 0,99894472 | YES | 0 | FALSE |
| 334 | 14,3385944 | 0,675847133 | 1,066505927 | 0,633702182 | 0,526275201 | 0,99894472 | YES | 10 | FALSE |
| 335 | 84,9391707 | 0,05579014 | 0,254281979 | 0,219402652 | 0,826336405 | 0,99894472 | YES | 0 | FALSE |
| 336 | 2,985334318 | 0,342410897 | 2,904255028 | 0,117899735 | 0,906147104 | 0,99894472 | YES | 0 | FALSE |
| 337 | 267,3251138 | -0,131296597 | 0,312954141 | -0,419539414 | 0,674821956 | 0,99894472 | YES | 0 | FALSE |
| 338 | 70,99439829 | 0,934131164 | 0,324618471 | 2,877627883 | 0,004006775 | 0,224668921 | YES | 25 | FALSE |
| 339 | 7,037408263 | -0,125857041 | 2,905025125 | -0,043323908 | 0,965443333 | 0,99894472 | YES | 0 | FALSE |
| 340 | 1,249713349 | -0,008240937 | 2,905099371 | -0,002836714 | 0,997736632 | 0,99894472 | YES | 0 | FALSE |
| 341 | 78,55695416 | 0,15570296 | 0,335261099 | 0,464422984 | 0,642344726 | 0,99894472 | YES | 0 | FALSE |
| 342 | 10,20531785 | 0,294332648 | 2,901644248 | 0,101436504 | 0,91920396 | 0,99894472 | YES | 0 | FALSE |
| 343 | 1,719887309 | -0,11392381 | 2,902423332 | -0,039251273 | 0,968690055 | 0,99894472 | YES | 0 | FALSE |
| 344 | 47,15928254 | -1,807964146 | 2,905159744 | -0,622328651 | 0,533725785 | 0,99894472 | YES | 0 | FALSE |
| 345 | 6,111053812 | 0,582331064 | 2,902194244 | 0,200651995 | 0,840970699 | 0,99894472 | YES | 57 | FALSE |
| 346 | 15,53629147 | 0,135888463 | 2,904498831 | 0,046785511 | 0,962684177 | 0,99894472 | YES | 0 | FALSE |
| 347 | 93,09663263 | -0,412261724 | 0,564617301 | -0,730161339 | 0,465291571 | 0,99894472 | YES | 0 | FALSE |
| 348 | 6,569822943 | -0,578054495 | 2,902931537 | -0,199127843 | 0,842162742 | 0,99894472 | YES | 130 | FALSE |
| 349 | 4,303921459 | 0,313896745 | 2,905230068 | 0,1080454 | 0,913959679 | 0,99894472 | YES | 2 | FALSE |
| 350 | 84,0393567 | 0,089108318 | 0,645858428 | 0,137968809 | 0,890265069 | 0,99894472 | YES | 0 | FALSE |
| 351 | 7,105330285 | 0,025136825 | 2,905225309 | 0,008652281 | 0,993096565 | 0,99894472 | YES | 0 | FALSE |
| 352 | 4,674932431 | -0,098635775 | 2,902664988 | -0,033981109 | 0,972892215 | 0,99894472 | YES | 0 | FALSE |
| 353 | 1686,904785 | -0,977446186 | 0,336238163 | -2,90700549 | 0,003649068 | 0,224668921 | YES | 0 | FALSE |
| 354 | 135,8032156 | -2,261498784 | 1,202284549 | -1,881001287 | 0,059971744 | 0,99894472 | YES | 0 | FALSE |
| 355 | 5,023366844 | 0,531952126 | 2,904869267 | 0,183124291 | 0,854700496 | 0,99894472 | YES | 68 | FALSE |
| 356 | 2,818052049 | -0,19483133 | 2,903856984 | -0,067093982 | 0,946506884 | 0,99894472 | YES | 0 | FALSE |

| Ticker | taxLevel | Phylum | Family | Genus | Species |
|---|---|---|---|---|---|
| 357 | ASV | Firmicutes | Lachnospiraceae | NA | NA |
| 358 | ASV | Firmicutes | [Eubacterium] coprostanoligenes group | NA | NA |
| 359 | ASV | Firmicutes | Oscillospiraceae | Oscillibacter | NA |
| 360 | ASV | Proteobacteria | Sutterellaceae | Parasutterella | excrementihominis |
| 361 | ASV | Verrucomicrobiota | vadinBE97 | NA | NA |
| 362 | ASV | Firmicutes | Ruminococcaceae | Angelakisella | NA |
| 363 | ASV | Firmicutes | Oscillospiraceae | UCG-005 | NA |
| 364 | ASV | Firmicutes | Ruminococcaceae | NA | NA |
| 365 | ASV | Firmicutes | Lachnospiraceae | Coprococcus | catus |
| 366 | ASV | Firmicutes | Lachnospiraceae | [Ruminococcus] gauvreauii group | NA |
| 367 | ASV | Verrucomicrobiota | Puniceicoccaceae | NA | NA |
| 368 | ASV | Firmicutes | Ruminococcaceae | NA | NA |
| 369 | ASV | Firmicutes | NA | NA | NA |
| 370 | ASV | Firmicutes | Lachnospiraceae | Lachnospiraceae ND3007 group | NA |
| 371 | ASV | Firmicutes | UCG-010 | NA | NA |
| 372 | ASV | Firmicutes | Lachnospiraceae | [Ruminococcus] torques group | NA |
| 373 | ASV | Firmicutes | Ruminococcaceae | Fournierella | NA |
| 374 | ASV | Firmicutes | Lachnospiraceae | [Eubacterium] eligens group | NA |
| 375 | ASV | Firmicutes | Lachnospiraceae | [Eubacterium] ruminantium group | NA |
| 376 | ASV | Firmicutes | Peptococcaceae | Peptococcus | NA |
| 377 | ASV | Firmicutes | Ruminococcaceae | Faecalibacterium | prausnitzii |
| 378 | ASV | Firmicutes | Acidaminococcaceae | Phascolarctobacterium | faecium |
| 379 | ASV | Firmicutes | Christensenellaceae | Christensenellaceae R-7 group | NA |
| 380 | ASV | Firmicutes | Peptococcaceae | NA | NA |
| 381 | ASV | Desulfobacterota | Desulfovibrionaceae | NA | NA |
| 382 | ASV | Firmicutes | Lachnospiraceae | Lachnospiraceae NK4A136 group | NA |
| 383 | ASV | Firmicutes | Ruminococcaceae | NA | NA |
| 384 | ASV | Firmicutes | Oscillospiraceae | Colidextribacter | NA |
| 385 | ASV | Firmicutes | Lachnospiraceae | Coprococcus | eutactus |
| 386 | ASV | Firmicutes | Lachnospiraceae | GCA-900066575 | NA |

| Ticker | DESeq2_baseMean | DESeq2_log2-FoldChange | DESeq2_lfcSE | DESeq2_stat | DESeq2_pvalue | DESeq2_padj | DESeq2 converged | ANCOM_W | ANCOM_detected_0.7 |
|---|---|---|---|---|---|---|---|---|---|
| 357 | 3,728044901 | 0,338628222 | 2,902146112 | 0,116682003 | 0,907112052 | 0,99894472 | YES | 0 | FALSE |
| 358 | 55,27170415 | 0,767967386 | 0,898826081 | 0,854411551 | 0,392876998 | 0,99894472 | YES | 7 | FALSE |
| 359 | 7,440181251 | 0,222456622 | 2,904555076 | 0,076588881 | 0,938950605 | 0,99894472 | YES | 0 | FALSE |
| 360 | 51,05649708 | 0,029193532 | 0,643169452 | 0,04539011 | 0,963796364 | 0,99894472 | YES | 0 | FALSE |
| 361 | 1,504875068 | 0,146663652 | 2,903267955 | 0,050516747 | 0,959710604 | 0,99894472 | YES | 0 | FALSE |
| 362 | 2,767870925 | 0,215263472 | 2,903410544 | 0,074141589 | 0,940897723 | 0,99894472 | YES | 0 | FALSE |
| 363 | 196,5919775 | 0,141471762 | 0,44613143 | 0,317107813 | 0,751161797 | 0,99894472 | YES | 0 | FALSE |
| 364 | 2,641872796 | 0,391552164 | 2,905009019 | 0,13478518 | 0,892781723 | 0,99894472 | YES | 0 | FALSE |
| 365 | 82,13050064 | -0,361247523 | 0,239930033 | -1,505636946 | 0,132160399 | 0,99894472 | YES | 19 | FALSE |
| 366 | 43,5767171 | 0,188222227 | 1,281550325 | 0,146870726 | 0,883234059 | 0,99894472 | YES | 0 | FALSE |
| 367 | 3,162923681 | 0,216716748 | 2,905420063 | 0,074590504 | 0,94054053 | 0,99894472 | YES | 0 | FALSE |
| 368 | 5,313173559 | 0,005947571 | 2,903075352 | 0,002048714 | 0,998365364 | 0,99894472 | YES | 0 | FALSE |
| 369 | 7,339116708 | -0,027440739 | 2,904895193 | -0,009446378 | 0,992462993 | 0,99894472 | YES | 0 | FALSE |
| 370 | 61,87979574 | -0,980268324 | 0,364715737 | -2,687759876 | 0,00719331 | 0,328494508 | YES | 157 | FALSE |
| 371 | 4,627302076 | 0,14844534 | 2,90328501 | 0,05113013 | 0,959221827 | 0,99894472 | YES | 0 | FALSE |
| 372 | 140,4526504 | 0,190081302 | 0,721405467 | 0,263487471 | 0,792174879 | 0,99894472 | YES | 0 | FALSE |
| 373 | 6,217452364 | 0,124719775 | 2,904467355 | 0,04294067 | 0,965748828 | 0,99894472 | YES | 0 | FALSE |
| 374 | 99,71543913 | -0,096453846 | 0,57632564 | -0,167359976 | 0,867086815 | 0,99894472 | YES | 0 | FALSE |
| 375 | 148,127431 | -0,084299997 | 1,069887379 | -0,078793338 | 0,937197003 | 0,99894472 | YES | 0 | FALSE |
| 376 | 4,027590161 | -0,265138896 | 2,903089532 | -0,091329907 | 0,927230455 | 0,99894472 | YES | 0 | FALSE |
| 377 | 972,1575633 | -0,146770082 | 0,305923312 | -0,479761027 | 0,631397328 | 0,99894472 | YES | 0 | FALSE |
| 378 | 70,47815475 | -0,004488116 | 2,904105476 | -0,001545438 | 0,998766919 | 0,99894472 | YES | 0 | FALSE |
| 379 | 1,714775994 | 0,241752496 | 2,902142927 | 0,083301375 | 0,933611907 | 0,99894472 | YES | 0 | FALSE |
| 380 | 2,729887651 | 0,059200891 | 2,904850931 | 0,02038001 | 0,98374023 | 0,99894472 | YES | 0 | FALSE |
| 381 | 1,551220695 | 0,500104232 | 2,903987285 | 0,172212955 | 0,863270113 | 0,99894472 | YES | 1 | FALSE |
| 382 | 113,9387832 | -0,862745948 | 0,780031231 | -1,106040263 | 0,268709093 | 0,99894472 | YES | 0 | FALSE |
| 383 | 1,593405793 | 0,044463561 | 2,904431825 | 0,015308867 | 0,987785768 | 0,99894472 | YES | 0 | FALSE |
| 384 | 2,421772883 | 0,103333389 | 2,904549809 | 0,035576387 | 0,971620137 | 0,99894472 | YES | 0 | FALSE |
| 385 | 92,07779332 | 0,097388573 | 2,901094521 | 0,033569597 | 0,973220367 | 0,99894472 | YES | 0 | FALSE |
| 386 | 4,748954977 | 0,478858643 | 2,90391262 | 0,164901189 | 0,869021757 | 0,99894472 | YES | 4 | FALSE |

| Ticker | taxLevel | Phylum | Family | Genus | Species |
|---|---|---|---|---|---|
| 387 | ASV | Firmicutes | Christensenellaceae | Christensenellaceae R-7 group | NA |
| 388 | ASV | Firmicutes | Lachnospiraceae | Lachnospiraceae NK4A136 group | NA |
| 389 | ASV | Verrucomicrobiota | Victivallaceae | Victivallis | vadensis |
| 390 | ASV | Bacteroidota | Bacteroidaceae | Bacteroides | NA |
| 391 | ASV | Firmicutes | Lachnospiraceae | Coprococcus | NA |
| 392 | ASV | Firmicutes | Lachnospiraceae | [Eubacterium] hallii group | NA |
| 393 | ASV | Firmicutes | NA | NA | NA |
| 394 | ASV | Proteobacteria | Sutterellaceae | Sutterella | wadsworthensis |
| 395 | ASV | Firmicutes | NA | NA | NA |
| 396 | ASV | Firmicutes | Lachnospiraceae | Dorea | formicigenerans |
| 397 | ASV | Firmicutes | Lachnospiraceae | Shuttleworthia | NA |
| 398 | ASV | Firmicutes | Lachnospiraceae | Blautia | NA |
| 399 | ASV | Desulfobacterota | Desulfovibrionaceae | NA | NA |
| 400 | ASV | Bacteroidota | Barnesiellaceae | Coprobacter | secundus |
| 401 | ASV | Bacteroidota | Bacteroidaceae | Bacteroides | NA |
| 402 | ASV | Firmicutes | Lachnospiraceae | [Eubacterium] eligens group | NA |
| 403 | ASV | Firmicutes | Erysipelotrichaceae | Turicibacter | sanguinis |
| 404 | ASV | Firmicutes | Oscillospiraceae | NK4A214 group | NA |
| 405 | ASV | Firmicutes | UCG-010 | NA | NA |
| 406 | ASV | Bacteroidota | Rikenellaceae | Alistipes | shahii |
| 407 | ASV | Bacteroidota | Prevotellaceae | Prevotella | NA |
| 408 | ASV | Firmicutes | Lachnospiraceae | CAG-56 | NA |
| 409 | ASV | Firmicutes | Lachnospiraceae | [Ruminococcus] gnavus group | NA |
| 410 | ASV | Firmicutes | Acidaminococcaceae | Phascolarctobacterium | succinatutens |
| 411 | ASV | Bacteroidota | Bacteroidaceae | Bacteroides | plebeius |
| 412 | Genus | Actinobacteriota | Bifidobacteriaceae | Bifidobacterium | NA |
| 413 | Genus | Verrucomicrobiota | Akkermansiaceae | Akkermansia | NA |
| 414 | Genus | Firmicutes | Lachnospiraceae | Lachnospira | NA |
| 415 | Genus | Firmicutes | Oscillospiraceae | UCG-003 | NA |
| 416 | Genus | Firmicutes | Veillonellaceae | Dialister | NA |

| Ticker | DESeq2_baseMean | DESeq2_log2-FoldChange | DESeq2_lfcSE | DESeq2_stat | DESeq2_pvalue | DESeq2_padj | DESeq2 converged | ANCOM_W | ANCOM_detected_0.7 |
|---|---|---|---|---|---|---|---|---|---|
| 387 | 1,275801653 | 0,18880628 | 2,904776693 | 0,064998553 | 0,948175153 | 0,99894472 | YES | 0 | FALSE |
| 388 | 55,95176432 | 0,358807687 | 1,426999312 | 0,251442088 | 0,801472331 | 0,99894472 | YES | 0 | FALSE |
| 389 | 1,209779611 | 0,193801181 | 2,904201161 | 0,066731321 | 0,946795599 | 0,99894472 | YES | 0 | FALSE |
| 390 | 42,67470703 | 0,472046354 | 2,474820869 | 0,190739605 | 0,848729608 | 0,99894472 | YES | 0 | FALSE |
| 391 | 293,4496484 | -0,351719784 | 1,021382262 | -0,344356659 | 0,730578079 | 0,99894472 | YES | 0 | FALSE |
| 392 | 58,67428243 | 0,070264676 | 0,505620091 | 0,138967333 | 0,889475963 | 0,99894472 | YES | 0 | FALSE |
| 393 | 3,347437588 | 0,176222094 | 2,900519988 | 0,060755345 | 0,951554054 | 0,99894472 | YES | 0 | FALSE |
| 394 | 36,31469707 | 0,160980091 | 1,664018334 | 0,096741777 | 0,922931462 | 0,99894472 | YES | 0 | FALSE |
| 395 | 1,709236849 | 0,48724502 | 2,90407956 | 0,167779501 | 0,866756749 | 0,99894472 | YES | 0 | FALSE |
| 396 | 61,62795417 | 0,746557693 | 0,563918862 | 1,323874308 | 0,185544792 | 0,99894472 | YES | 1 | FALSE |
| 397 | 2,990038708 | 0,058839564 | 2,904913294 | 0,020255188 | 0,983839803 | 0,99894472 | YES | 0 | FALSE |
| 398 | 64,70622657 | 1,910289055 | 0,650744621 | 2,935543365 | 0,003329641 | 0,224668921 | YES | 38 | FALSE |
| 399 | 3,275879807 | 0,133143518 | 2,905491416 | 0,045824784 | 0,963449904 | 0,99894472 | YES | 0 | FALSE |
| 400 | 3,657989246 | 0,170773156 | 2,902141618 | 0,058843839 | 0,95307649 | 0,99894472 | YES | 0 | FALSE |
| 401 | 654,2550208 | 0,054759674 | 0,400600774 | 0,13669388 | 0,891272767 | 0,99894472 | NO | 0 | FALSE |
| 402 | 72,73003574 | 0,945435895 | 0,776743753 | 1,217178627 | 0,22353626 | 0,99894472 | YES | 0 | FALSE |
| 403 | 24,93246918 | 1,439359653 | 2,900563293 | 0,496234527 | 0,619728954 | 0,99894472 | YES | 0 | FALSE |
| 404 | 97,03939266 | 0,194296011 | 0,484341252 | 0,401155199 | 0,688305863 | 0,99894472 | YES | 0 | FALSE |
| 405 | 3,458344526 | -0,254419836 | 2,903580289 | -0,0876228 | 0,93017648 | 0,99894472 | YES | 0 | FALSE |
| 406 | 93,3536042 | 1,073599705 | 0,705976917 | 1,52072919 | 0,12832781 | 0,99894472 | YES | 0 | FALSE |
| 407 | 36,03751168 | -0,167093543 | 2,90521467 | -0,057515042 | 0,954134924 | 0,99894472 | YES | 0 | FALSE |
| 408 | 107,3283829 | -0,887327361 | 0,551036442 | -1,610287985 | 0,107335001 | 0,99894472 | YES | 0 | FALSE |
| 409 | 57,72130721 | -2,520566636 | 2,904067903 | -0,867943423 | 0,385425306 | 0,99894472 | YES | 0 | FALSE |
| 410 | 200,6469766 | -0,55855006 | 0,522939777 | -1,068096337 | 0,285477058 | 0,99894472 | YES | 0 | FALSE |
| 411 | 56,73587672 | -1,188171418 | 2,904040279 | -0,409144263 | 0,682433795 | 0,99894472 | YES | 0 | FALSE |
| 412 | 721,2204328 | 0,350476123 | 0,205492301 | 1,705543817 | 0,088093047 | 0,674605374 | YES | 0 | FALSE |
| 413 | 1143,268978 | -0,051254833 | 0,406503139 | -0,126087176 | 0,899662919 | 0,999636303 | YES | 0 | FALSE |
| 414 | 123,7797096 | -1,086671986 | 0,392592986 | -2,767935303 | 0,005641265 | 0,144416392 | YES | 0 | FALSE |
| 415 | 149,7231774 | -0,650603608 | 0,33853509 | -1,921820299 | 0,054628373 | 0,528721726 | YES | 0 | FALSE |
| 416 | 564,3048754 | 0,03761961 | 0,550928218 | 0,06828405 | 0,945559521 | 0,999636303 | YES | 0 | FALSE |

| Ticker | taxLevel | Phylum | Family | Genus | Species |
|---|---|---|---|---|---|
| 417 | Genus | Desulfobacterota | Desulfovibrionaceae | Mailhella | NA |
| 418 | Genus | Firmicutes | Oscillospiraceae | NK4A214 group | NA |
| 419 | Genus | Firmicutes | Lachnospiraceae | Fusicatenibacter | NA |
| 420 | Genus | Proteobacteria | Enterobacteriaceae | Klebsiella | NA |
| 421 | Genus | Firmicutes | Ruminococcaceae | Candidatus Soleaferrea | NA |
| 422 | Genus | Actinobacteriota | Eggerthellaceae | Senegalimassilia | NA |
| 423 | Genus | Actinobacteriota | Eggerthellaceae | Enterorhabdus | NA |
| 424 | Genus | Bacteroidota | Marinifilaceae | Sanguibacteroides | NA |
| 425 | Genus | Firmicutes | Lachnospiraceae | Lachnoclostridium | NA |
| 426 | Genus | Firmicutes | Peptostreptococcaceae | Terrisporobacter | NA |
| 427 | Genus | Desulfobacterota | Desulfovibrionaceae | Bilophila | NA |
| 428 | Genus | Firmicutes | Oscillospiraceae | Oscillibacter | NA |
| 429 | Genus | Firmicutes | Christensenellaceae | Christensenellaceae R-7 group | NA |
| 430 | Genus | Firmicutes | Erysipelatoclostridiaceae | UCG-004 | NA |
| 431 | Genus | Firmicutes | Lachnospiraceae | [Eubacterium] ventriosum group | NA |
| 432 | Genus | Firmicutes | Peptostreptococcaceae | Romboutsia | NA |
| 433 | Genus | Desulfobacterota | Desulfovibrionaceae | Desulfovibrio | NA |
| 434 | Genus | Firmicutes | Ruminococcaceae | Paludicola | NA |
| 435 | Genus | Firmicutes | Oscillospiraceae | Intestinimonas | NA |
| 436 | Genus | Actinobacteriota | Coriobacteriaceae | Collinsella | NA |
| 437 | Genus | Proteobacteria | Pasteurellaceae | Haemophilus | NA |
| 438 | Genus | Firmicutes | Erysipelatoclostridiaceae | Asteroleplasma | NA |
| 439 | Genus | Firmicutes | Lachnospiraceae | Dorea | NA |
| 440 | Genus | Firmicutes | Erysipelatoclostridiaceae | Erysipelotrichaceae UCG-003 | NA |
| 441 | Genus | Firmicutes | Ruminococcaceae | UBA1819 | NA |
| 442 | Genus | Bacteroidota | Marinifilaceae | Odoribacter | NA |
| 443 | Genus | Actinobacteriota | Eggerthellaceae | Adlercreutzia | NA |
| 444 | Genus | Bacteroidota | Barnesiellaceae | Barnesiella | NA |
| 445 | Genus | Bacteroidota | Prevotellaceae | Prevotellaceae NK3B31 group | NA |
| 446 | Genus | Bacteroidota | Bacteroidaceae | Bacteroides | NA |

| Ticker | DESeq2_baseMean | DESeq2_log2-FoldChange | DESeq2_lfcSE | DESeq2_stat | DESeq2_pvalue | DESeq2_padj | DESeq2 converged | ANCOM_W | ANCOM_detected_0.7 |
|---|---|---|---|---|---|---|---|---|---|
| 417 | 5,714723446 | 0,074332497 | 2,904675348 | 0,025590638 | 0,979583853 | 0,999636303 | YES | 0 | FALSE |
| 418 | 422,4623148 | 0,38089252 | 0,183912409 | 2,071053942 | 0,038353755 | 0,490928068 | YES | 1 | FALSE |
| 419 | 768,4400643 | -0,029919198 | 0,272601769 | -0,109754234 | 0,912604288 | 0,999636303 | YES | 0 | FALSE |
| 420 | 68,07885842 | 3,931523328 | 2,90469822 | 1,353504919 | 0,17589438 | 0,913845111 | YES | 0 | FALSE |
| 421 | 6,094425354 | -0,117608389 | 2,903712132 | -0,040502772 | 0,967692297 | 0,999636303 | YES | 0 | FALSE |
| 422 | 19,54514968 | 0,150243536 | 0,510174284 | 0,294494531 | 0,768380042 | 0,999636303 | YES | 0 | FALSE |
| 423 | 8,098004857 | -0,162653704 | 0,801156581 | -0,203023613 | 0,839116576 | 0,999636303 | YES | 0 | FALSE |
| 424 | 2,94952053 | 0,02963724 | 2,903885484 | 0,010206064 | 0,991856881 | 0,999636303 | YES | 0 | FALSE |
| 425 | 182,5651877 | -0,234763998 | 0,288128404 | -0,814789499 | 0,415192813 | 0,999636303 | YES | 0 | FALSE |
| 426 | 6,591681567 | 0,279009556 | 2,902748246 | 0,096119102 | 0,92342598 | 0,999636303 | YES | 0 | FALSE |
| 427 | 64,32240255 | 0,416302329 | 0,245239676 | 1,697532531 | 0,089596026 | 0,674605374 | YES | 0 | FALSE |
| 428 | 82,17895313 | 0,097100262 | 0,286472373 | 0,338951574 | 0,734646212 | 0,999636303 | YES | 0 | FALSE |
| 429 | 1021,268723 | 0,198898135 | 0,239188347 | 0,831554452 | 0,405660481 | 0,999636303 | YES | 0 | FALSE |
| 430 | 8,366500089 | 0,212882268 | 2,90364957 | 0,073315413 | 0,941555127 | 0,999636303 | YES | 0 | FALSE |
| 431 | 101,4029537 | -0,358651864 | 0,329549631 | -1,088309106 | 0,276458672 | 0,999636303 | YES | 0 | FALSE |
| 432 | 38,6635841 | 1,232186582 | 0,622553535 | 1,97924598 | 0,047788318 | 0,528721726 | YES | 0 | FALSE |
| 433 | 265,2572369 | 0,030249509 | 0,16683235 | 0,181316806 | 0,856118916 | 0,999636303 | YES | 0 | FALSE |
| 434 | 6,197525142 | 0,055613786 | 2,900594816 | 0,019173235 | 0,984702909 | 0,999636303 | YES | 0 | FALSE |
| 435 | 131,2055575 | 0,015813406 | 0,204551161 | 0,07730783 | 0,938378662 | 0,999636303 | NO | 0 | FALSE |
| 436 | 233,933023 | -0,19285134 | 0,248152009 | -0,777150026 | 0,437070262 | 0,999636303 | YES | 0 | FALSE |
| 437 | 11,93705431 | -1,514230784 | 2,029777427 | -0,746008288 | 0,455662408 | 0,999636303 | YES | 0 | FALSE |
| 438 | 6,420258122 | -0,117107924 | 2,903418065 | -0,040334503 | 0,967826447 | 0,999636303 | YES | 0 | FALSE |
| 439 | 678,9890903 | -0,162308279 | 0,188837338 | -0,8595137 | 0,390057165 | 0,999636303 | YES | 0 | FALSE |
| 440 | 177,0676523 | -0,24123914 | 0,27623789 | -0,873302139 | 0,382498415 | 0,999636303 | YES | 0 | FALSE |
| 441 | 26,39412807 | -0,183952201 | 0,800545963 | -0,229783434 | 0,818260058 | 0,999636303 | YES | 0 | FALSE |
| 442 | 39,33887948 | 0,055089024 | 0,256718232 | 0,214589448 | 0,830087422 | 0,999636303 | YES | 0 | FALSE |
| 443 | 8,504437492 | 0,016009293 | 1,799907387 | 0,008894509 | 0,992903302 | 0,999636303 | YES | 0 | FALSE |
| 444 | 246,9722513 | -0,371143084 | 0,275853962 | -1,34543322 | 0,178485373 | 0,913845111 | YES | 0 | FALSE |
| 445 | 51,69489275 | -2,636044669 | 2,90454452 | -0,907558707 | 0,364111419 | 0,999636303 | YES | 0 | FALSE |
| 446 | 5032,644884 | -0,180654367 | 0,123711886 | -1,460283025 | 0,144212305 | 0,818156949 | YES | 0 | FALSE |

| Ticker | taxLevel | Phylum | Family | Genus | Species |
|---|---|---|---|---|---|
| 447 | Genus | Bacteroidota | Prevotellaceae | Prevotella | NA |
| 448 | Genus | Firmicutes | Lachnospiraceae | [Eubacterium] xylanophilum group | NA |
| 449 | Genus | Firmicutes | Lachnospiraceae | Lachnospiraceae FCS020 group | NA |
| 450 | Genus | Firmicutes | Lachnospiraceae | Lachnospiraceae UCG-010 | NA |
| 451 | Genus | Firmicutes | Peptococcaceae | Peptococcus | NA |
| 452 | Genus | Firmicutes | Lachnospiraceae | Lachnospiraceae UCG-004 | NA |
| 453 | Genus | Firmicutes | Lachnospiraceae | Lachnospiraceae UCG-001 | NA |
| 454 | Genus | Firmicutes | Oscillospiraceae | Flavonifractor | NA |
| 455 | Genus | Firmicutes | Streptococcaceae | Streptococcus | NA |
| 456 | Genus | Firmicutes | Acholeplasmataceae | Anaeroplasma | NA |
| 457 | Genus | Bacteroidota | Prevotellaceae | Alloprevotella | NA |
| 458 | Genus | Firmicutes | Lachnospiraceae | Tyzzerella | NA |
| 459 | Genus | Euryarchaeota | Methanobacteriaceae | Methanosphaera | NA |
| 460 | Genus | Firmicutes | Erysipelotrichaceae | Solobacterium | NA |
| 461 | Genus | Firmicutes | Lachnospiraceae | Lachnospiraceae AC2044 group | NA |
| 462 | Genus | Euryarchaeota | Methanobacteriaceae | Methanobrevibacter | NA |
| 463 | Genus | Firmicutes | Ruminococcaceae | CAG-352 | NA |
| 464 | Genus | Firmicutes | Clostridiaceae | Clostridium sensu stricto 1 | NA |
| 465 | Genus | Actinobacteriota | Eggerthellaceae | Slackia | NA |
| 466 | Genus | Firmicutes | Oscillospiraceae | UCG-002 | NA |
| 467 | Genus | Firmicutes | Erysipelotrichaceae | Catenisphaera | NA |
| 468 | Genus | Firmicutes | Anaerovoracaceae | Mogibacterium | NA |
| 469 | Genus | Firmicutes | Ruminococcaceae | Negativibacillus | NA |
| 470 | Genus | Firmicutes | Streptococcaceae | Lactococcus | NA |
| 471 | Genus | Firmicutes | Ruminococcaceae | Subdoligranulum | NA |
| 472 | Genus | Proteobacteria | Enterobacteriaceae | Escherichia-Shigella | NA |
| 473 | Genus | Firmicutes | Lachnospiraceae | GCA-900066575 | NA |
| 474 | Genus | Firmicutes | Veillonellaceae | Veillonella | NA |
| 475 | Genus | Firmicutes | Oscillospiraceae | Colidextribacter | NA |
| 476 | Genus | Bacteroidota | Rikenellaceae | Alistipes | NA |

| Ticker | DESeq2_baseMean | DESeq2_log2-FoldChange | DESeq2_lfcSE | DESeq2_stat | DESeq2_pvalue | DESeq2_padj | DESeq2 converged | ANCOM_W | ANCOM_detected_0.7 |
|---|---|---|---|---|---|---|---|---|---|
| 447 | 1133,926116 | -1,199866666 | 0,429374517 | -2,794452435 | 0,005198769 | 0,144416392 | YES | 0 | FALSE |
| 448 | 54,39281101 | 0,17712794 | 0,490929478 | 0,36080119 | 0,718248073 | 0,999636303 | YES | 0 | FALSE |
| 449 | 41,87861749 | -0,31749773 | 0,438160355 | -0,724615375 | 0,468688024 | 0,999636303 | YES | 0 | FALSE |
| 450 | 55,94278565 | -0,109385716 | 0,430279662 | -0,254220047 | 0,799325564 | 0,999636303 | YES | 0 | FALSE |
| 451 | 75,38353118 | -0,010388069 | 0,567266961 | -0,018312487 | 0,985389566 | 0,999636303 | YES | 0 | FALSE |
| 452 | 44,44180757 | -0,673253371 | 0,533499202 | -1,261957599 | 0,206964044 | 0,999636303 | YES | 0 | FALSE |
| 453 | 57,93753735 | -0,691204322 | 0,684282098 | -1,010116039 | 0,312439699 | 0,999636303 | YES | 0 | FALSE |
| 454 | 19,05663697 | -0,385044096 | 1,423918019 | -0,270411703 | 0,786843538 | 0,999636303 | YES | 0 | FALSE |
| 455 | 236,3988458 | 0,826298081 | 0,380265258 | 2,172951813 | 0,029783944 | 0,423593874 | YES | 0 | FALSE |
| 456 | 36,74837665 | 0,039188718 | 2,904065044 | 0,013494436 | 0,989233325 | 0,999636303 | YES | 0 | FALSE |
| 457 | 144,0791813 | -0,237057539 | 1,937487517 | -0,122353067 | 0,902619407 | 0,999636303 | YES | 0 | FALSE |
| 458 | 54,98143302 | 0,278326881 | 1,719757133 | 0,161840806 | 0,871431218 | 0,999636303 | YES | 0 | FALSE |
| 459 | 11,31173121 | 0,185655228 | 2,903645185 | 0,063938676 | 0,949019056 | 0,999636303 | YES | 0 | FALSE |
| 460 | 15,48373845 | 0,085845276 | 2,903964699 | 0,029561405 | 0,976416846 | 0,999636303 | YES | 0 | FALSE |
| 461 | 65,84336143 | 0,136474474 | 1,448721948 | 0,094203359 | 0,924947617 | 0,999636303 | YES | 0 | FALSE |
| 462 | 193,8582867 | 0,531176304 | 0,366286863 | 1,45016477 | 0,147012577 | 0,818156949 | YES | 0 | FALSE |
| 463 | 304,8295096 | -0,626218713 | 2,19613655 | -0,285145617 | 0,775532582 | 0,999636303 | YES | 0 | FALSE |
| 464 | 88,43123869 | 0,191892378 | 0,357822916 | 0,536277496 | 0,591766781 | 0,999636303 | YES | 0 | FALSE |
| 465 | 9,730735359 | -0,359951968 | 0,925005092 | -0,389135121 | 0,697176194 | 0,999636303 | YES | 0 | FALSE |
| 466 | 2032,576687 | 0,026751547 | 0,153669925 | 0,174084468 | 0,86179908 | 0,999636303 | YES | 0 | FALSE |
| 467 | 20,19897597 | 0,171393486 | 2,903457668 | 0,05903082 | 0,95292756 | 0,999636303 | YES | 0 | FALSE |
| 468 | 25,24876388 | 0,158649767 | 2,903612513 | 0,054638753 | 0,956426264 | 0,999636303 | YES | 0 | FALSE |
| 469 | 45,26405539 | 1,041394586 | 0,416402206 | 2,50093436 | 0,012386613 | 0,226498074 | YES | 33 | FALSE |
| 470 | 7,843484646 | 0,56280261 | 2,903734505 | 0,193820272 | 0,846316614 | 0,999636303 | YES | 0 | FALSE |
| 471 | 1006,12734 | 0,034875048 | 0,185168696 | 0,188342027 | 0,850608542 | 0,999636303 | YES | 0 | FALSE |
| 472 | 661,0305271 | -0,571961431 | 0,485395223 | -1,178341698 | 0,238660411 | 0,999636303 | YES | 0 | FALSE |
| 473 | 16,11197157 | 0,118187901 | 0,745718693 | 0,158488585 | 0,874071815 | 0,999636303 | YES | 0 | FALSE |
| 474 | 31,16545164 | -0,438459173 | 1,867863591 | -0,234738326 | 0,814411839 | 0,999636303 | YES | 0 | FALSE |
| 475 | 98,85286937 | -0,139949768 | 0,197674794 | -0,707979837 | 0,478957779 | 0,999636303 | YES | 0 | FALSE |
| 476 | 1144,772964 | -0,026988855 | 0,140362807 | -0,192279245 | 0,847523475 | 0,999636303 | YES | 0 | FALSE |

| Ticker | taxLevel | Phylum | Family | Genus | Species |
|---|---|---|---|---|---|
| 477 | Genus | Firmicutes | Anaerovoracaceae | Family XIII UCG-001 | NA |
| 478 | Genus | Firmicutes | Anaerovoracaceae | Family XIII AD3011 group | NA |
| 479 | Genus | Firmicutes | Anaerovoracaceae | [Eubacterium] brachy group | NA |
| 480 | Genus | Firmicutes | Peptostreptococcaceae | Intestinibacter | NA |
| 481 | Genus | Firmicutes | Lachnospiraceae | Lachnotalea | NA |
| 482 | Genus | Firmicutes | Lachnospiraceae | Anaerostipes | NA |
| 483 | Genus | Firmicutes | Lachnospiraceae | Marvinbryantia | NA |
| 484 | Genus | Firmicutes | Lachnospiraceae | [Eubacterium] hallii group | NA |
| 485 | Genus | Bacteroidota | Barnesiellaceae | Coprobacter | NA |
| 486 | Genus | Firmicutes | Selenomonadaceae | Mitsuokella | NA |
| 487 | Genus | Firmicutes | Ruminococcaceae | DTU089 | NA |
| 488 | Genus | Bacteroidota | Tannerellaceae | Parabacteroides | NA |
| 489 | Genus | Firmicutes | Ruminococcaceae | Ruminococcus | NA |
| 490 | Genus | Firmicutes | Erysipelotrichaceae | Holdemanella | NA |
| 491 | Genus | Bacteroidota | Prevotellaceae | Paraprevotella | NA |
| 492 | Genus | Firmicutes | Lactobacillaceae | Lactobacillus | NA |
| 493 | Genus | Firmicutes | Oscillospiraceae | Oscillospira | NA |
| 494 | Genus | Bacteroidota | Prevotellaceae | Prevotellaceae UCG-001 | NA |
| 495 | Genus | Firmicutes | Erysipelatoclostridiaceae | Coprobacillus | NA |
| 496 | Genus | Actinobacteriota | Atopobiaceae | Olsenella | NA |
| 497 | Genus | Verrucomicrobiota | Victivallaceae | Victivallis | NA |
| 498 | Genus | Actinobacteriota | Actinomycetaceae | Actinomyces | NA |
| 499 | Genus | Firmicutes | Lachnospiraceae | Howardella | NA |
| 500 | Genus | Firmicutes | Lachnospiraceae | Eisenbergiella | NA |
| 501 | Genus | Firmicutes | Lachnospiraceae | Blautia | NA |
| 502 | Genus | Firmicutes | Oscillospiraceae | UCG-005 | NA |
| 503 | Genus | Proteobacteria | Oxalobacteraceae | Oxalobacter | NA |
| 504 | Genus | Firmicutes | Lachnospiraceae | [Ruminococcus] torques group | NA |
| 505 | Genus | Firmicutes | Veillonellaceae | Allisonella | NA |
| 506 | Genus | Bacteroidota | Marinifilaceae | Butyricimonas | NA |

| Ticker | DESeq2_baseMean | DESeq2_log2-FoldChange | DESeq2_lfcSE | DESeq2_stat | DESeq2_pvalue | DESeq2_padj | DESeq2 converged | ANCOM_W | ANCOM_detected_0.7 |
|---|---|---|---|---|---|---|---|---|---|
| 477 | 28,1853619 | 0,035945154 | 0,361721435 | 0,099372473 | 0,920842538 | 0,999636303 | YES | 0 | FALSE |
| 478 | 62,06718117 | 0,103251912 | 0,231534543 | 0,445946037 | 0,655636226 | 0,999636303 | YES | 0 | FALSE |
| 479 | 1,595132946 | 0,508393433 | 2,904562273 | 0,175032719 | 0,861053924 | 0,999636303 | YES | 8 | FALSE |
| 480 | 36,62497344 | 0,404208168 | 0,666558461 | 0,606410678 | 0,54424208 | 0,999636303 | YES | 0 | FALSE |
| 481 | 4,438815253 | -0,483098607 | 2,901812935 | -0,166481651 | 0,867777922 | 0,999636303 | YES | 0 | FALSE |
| 482 | 165,7530311 | 0,097129691 | 0,289984137 | 0,33494829 | 0,737664094 | 0,999636303 | YES | 0 | FALSE |
| 483 | 63,59587409 | 0,950230648 | 0,619672389 | 1,533440355 | 0,125167392 | 0,801071308 | YES | 0 | FALSE |
| 484 | 208,3356985 | 0,172120699 | 0,173664369 | 0,991111188 | 0,32163129 | 0,999636303 | YES | 0 | FALSE |
| 485 | 12,213473 | 0,235136187 | 0,744472826 | 0,315842538 | 0,752122029 | 0,999636303 | YES | 0 | FALSE |
| 486 | 21,24036454 | 0,001324231 | 2,905120083 | 0,000455827 | 0,999636303 | 0,999636303 | YES | 0 | FALSE |
| 487 | 12,63647498 | 0,625883064 | 0,7307808 | 0,856458002 | 0,391744494 | 0,999636303 | YES | 0 | FALSE |
| 488 | 689,0294243 | -0,121921929 | 0,18105392 | -0,673401212 | 0,500692078 | 0,999636303 | YES | 0 | FALSE |
| 489 | 1802,317879 | -0,05886896 | 0,253638551 | -0,232097839 | 0,816462023 | 0,999636303 | YES | 0 | FALSE |
| 490 | 269,6096991 | -0,199547209 | 0,319526796 | -0,624508528 | 0,532293671 | 0,999636303 | YES | 0 | FALSE |
| 491 | 142,3554429 | -0,124275196 | 0,327970945 | -0,37892136 | 0,704746262 | 0,999636303 | YES | 0 | FALSE |
| 492 | 164,6392328 | 2,142796478 | 0,841662148 | 2,545910473 | 0,010899314 | 0,226498074 | YES | 0 | FALSE |
| 493 | 34,73878558 | 0,405606729 | 0,726152796 | 0,558569397 | 0,576455631 | 0,999636303 | YES | 0 | FALSE |
| 494 | 81,0783181 | 0,098380618 | 2,905122269 | 0,033864536 | 0,972985173 | 0,999636303 | YES | 0 | FALSE |
| 495 | 6,609277278 | 0,24833657 | 2,903472347 | 0,085530889 | 0,93183934 | 0,999636303 | YES | 0 | FALSE |
| 496 | 4,974908538 | 0,046528056 | 2,902234918 | 0,016031802 | 0,98720902 | 0,999636303 | YES | 0 | FALSE |
| 497 | 7,171043458 | 0,298705279 | 2,049216298 | 0,145765617 | 0,884106419 | 0,999636303 | YES | 0 | FALSE |
| 498 | 1,080148438 | 0,257552179 | 2,903726352 | 0,088697125 | 0,929322617 | 0,999636303 | YES | 0 | FALSE |
| 499 | 9,850588669 | -0,356525883 | 1,064616935 | -0,334886541 | 0,737710675 | 0,999636303 | YES | 0 | FALSE |
| 500 | 16,69629512 | -4,766242231 | 2,90307034 | -1,641793575 | 0,100632791 | 0,715610959 | YES | 0 | FALSE |
| 501 | 1358,368645 | -0,07123541 | 0,140756439 | -0,506089886 | 0,61279355 | 0,999636303 | YES | 0 | FALSE |
| 502 | 668,0635607 | -0,085967439 | 0,219711555 | -0,391274093 | 0,695594647 | 0,999636303 | YES | 0 | FALSE |
| 503 | 14,36227103 | 0,210361545 | 0,383971119 | 0,547857728 | 0,583789597 | 0,999636303 | YES | 0 | FALSE |
| 504 | 629,5657794 | -0,202945933 | 0,195562268 | -1,037756081 | 0,299383628 | 0,999636303 | YES | 0 | FALSE |
| 505 | 9,50052128 | -0,179504203 | 2,901643003 | -0,061862952 | 0,950671971 | 0,999636303 | YES | 0 | FALSE |
| 506 | 49,26473663 | 0,249884389 | 0,241592078 | 1,034323606 | 0,300984905 | 0,999636303 | YES | 0 | FALSE |

| Ticker | taxLevel | Phylum | Family | Genus | Species |
|---|---|---|---|---|---|
| 507 | Genus | Firmicutes | Monoglobaceae | Monoglobus | NA |
| 508 | Genus | Bacteroidota | Rikenellaceae | Rikenellaceae RC9 gut group | NA |
| 509 | Genus | Firmicutes | Lachnospiraceae | Roseburia | NA |
| 510 | Genus | Firmicutes | Erysipelatoclostridiaceae | Erysipelatoclostridium | NA |
| 511 | Genus | Firmicutes | Ruminococcaceae | [Eubacterium] siraeum group | NA |
| 512 | Genus | Firmicutes | Erysipelatoclostridiaceae | Catenibacterium | NA |
| 513 | Genus | Firmicutes | Ruminococcaceae | Angelakisella | NA |
| 514 | Genus | Firmicutes | Ruminococcaceae | Incertae Sedis | NA |
| 515 | Genus | Firmicutes | Butyricicoccaceae | Butyricicoccus | NA |
| 516 | Genus | Firmicutes | Erysipelotrichaceae | Holdemania | NA |
| 517 | Genus | Firmicutes | Ruminococcaceae | Fournierella | NA |
| 518 | Genus | Firmicutes | Lachnospiraceae | Agathobacter | NA |
| 519 | Genus | Firmicutes | Lachnospiraceae | Hungatella | NA |
| 520 | Genus | Proteobacteria | Sutterellaceae | Parasutterella | NA |
| 521 | Genus | Firmicutes | Ruminococcaceae | Anaerotruncus | NA |
| 522 | Genus | Firmicutes | Lachnospiraceae | [Ruminococcus] gauvreauii group | NA |
| 523 | Genus | Firmicutes | Lachnospiraceae | Lachnospiraceae ND3007 group | NA |
| 524 | Genus | Firmicutes | Defluviitaleaceae | Defluviitaleaceae UCG-011 | NA |
| 525 | Genus | Firmicutes | Lachnospiraceae | [Eubacterium] eligens group | NA |
| 526 | Genus | Firmicutes | Lachnospiraceae | [Eubacterium] ruminantium group | NA |
| 527 | Genus | Firmicutes | Ruminococcaceae | Faecalibacterium | NA |
| 528 | Genus | Firmicutes | Lachnospiraceae | Lachnospiraceae NK4A136 group | NA |
| 529 | Genus | Firmicutes | Lachnospiraceae | Coprococcus | NA |
| 530 | Genus | Actinobacteriota | Atopobiaceae | Libanicoccus | NA |
| 531 | Genus | Proteobacteria | Sutterellaceae | Sutterella | NA |
| 532 | Genus | Firmicutes | Lachnospiraceae | Shuttleworthia | NA |
| 533 | Genus | Synergistota | Synergistaceae | Cloacibacillus | NA |
| 534 | Genus | Firmicutes | Erysipelotrichaceae | Turicibacter | NA |
| 535 | Genus | Firmicutes | Lachnospiraceae | CAG-56 | NA |
| 536 | Genus | Firmicutes | Lachnospiraceae | [Ruminococcus] gnavus group | NA |

| Ticker | DESeq2_baseMean | DESeq2_log2-FoldChange | DESeq2_lfcSE | DESeq2_stat | DESeq2_pvalue | DESeq2_padj | DESeq2 converged | ANCOM_W | ANCOM_detected_0.7 |
|---|---|---|---|---|---|---|---|---|---|
| 507 | 134,5737112 | 0,528177478 | 0,274083328 | 1,927068975 | 0,053971035 | 0,528721726 | YES | 0 | FALSE |
| 508 | 207,2990445 | -0,627803579 | 0,580561434 | -1,081373204 | 0,279531136 | 0,999636303 | YES | 0 | FALSE |
| 509 | 535,2124177 | -0,484626844 | 0,255471155 | -1,896992418 | 0,057828939 | 0,528721726 | YES | 0 | FALSE |
| 510 | 15,74390741 | -0,184037256 | 1,27132393 | -0,144760318 | 0,884900114 | 0,999636303 | YES | 0 | FALSE |
| 511 | 211,2297365 | 0,029534049 | 0,40321147 | 0,073247047 | 0,941609529 | 0,999636303 | YES | 0 | FALSE |
| 512 | 76,10366348 | -0,36613825 | 2,903295365 | -0,126111264 | 0,899643851 | 0,999636303 | YES | 0 | FALSE |
| 513 | 7,049628943 | 0,753080864 | 1,661444673 | 0,453268698 | 0,65035527 | 0,999636303 | YES | 0 | FALSE |
| 514 | 120,0579137 | 0,494641018 | 0,217123706 | 2,278152978 | 0,022717465 | 0,363479436 | YES | 0 | FALSE |
| 515 | 151,7944076 | -0,136328651 | 0,198067667 | -0,688293314 | 0,491268091 | 0,999636303 | YES | 0 | FALSE |
| 516 | 2,556745844 | -0,137858054 | 1,634930482 | -0,084320438 | 0,932801663 | 0,999636303 | YES | 0 | FALSE |
| 517 | 13,76967531 | -0,068681007 | 2,90191629 | -0,023667467 | 0,981117857 | 0,999636303 | YES | 0 | FALSE |
| 518 | 1987,419 | -0,704069243 | 0,234349909 | -3,004350399 | 0,002661486 | 0,144416392 | YES | 0 | FALSE |
| 519 | 13,17537301 | -11,20360831 | 2,905764502 | -3,855649107 | 0,000115423 | 0,014774138 | YES | 0 | FALSE |
| 520 | 61,45386457 | 0,21851985 | 0,464891216 | 0,470045125 | 0,638322778 | 0,999636303 | YES | 0 | FALSE |
| 521 | 2,9255055 | 0,325342855 | 2,904048996 | 0,112030773 | 0,910799006 | 0,999636303 | YES | 0 | FALSE |
| 522 | 47,70677293 | 0,425398786 | 0,833869541 | 0,510150287 | 0,609946177 | 0,999636303 | YES | 0 | FALSE |
| 523 | 59,64700043 | -1,001607256 | 0,356258989 | -2,811458197 | 0,00493175 | 0,144416392 | YES | 51 | FALSE |
| 524 | 3,384136139 | 0,072647934 | 2,902731756 | 0,025027436 | 0,980033079 | 0,999636303 | YES | 0 | FALSE |
| 525 | 197,9799092 | -0,079729248 | 0,376285828 | -0,211884802 | 0,832196908 | 0,999636303 | NO | 0 | FALSE |
| 526 | 144,1230944 | -0,104378223 | 1,020468355 | -0,102284625 | 0,918530758 | 0,999636303 | YES | 0 | FALSE |
| 527 | 2334,575011 | -0,306025728 | 0,172751513 | -1,771479289 | 0,076481034 | 0,652638155 | YES | 0 | FALSE |
| 528 | 273,0660538 | -0,171234141 | 0,35640323 | -0,480450588 | 0,63090703 | 0,999636303 | YES | 0 | FALSE |
| 529 | 880,5389705 | -0,200296977 | 0,22879433 | -0,875445546 | 0,381331527 | 0,999636303 | YES | 0 | FALSE |
| 530 | 3,591584218 | 0,122616 | 2,904255821 | 0,042219421 | 0,96632378 | 0,999636303 | YES | 0 | FALSE |
| 531 | 157,8381505 | 0,00868578 | 0,274870288 | 0,03159956 | 0,974791394 | 0,999636303 | YES | 0 | FALSE |
| 532 | 8,078136836 | -0,143840286 | 2,900708992 | -0,049587975 | 0,960450729 | 0,999636303 | YES | 0 | FALSE |
| 533 | 12,2358954 | -0,939609804 | 2,904347223 | -0,323518413 | 0,746302665 | 0,999636303 | YES | 0 | FALSE |
| 534 | 32,84582589 | 1,285172598 | 0,860559655 | 1,493414885 | 0,135328619 | 0,818156949 | YES | 0 | FALSE |
| 535 | 119,2328775 | -0,821415656 | 0,511832865 | -1,604851333 | 0,108526526 | 0,73112607 | YES | 49 | FALSE |
| 536 | 59,7771096 | -2,868540898 | 2,904147586 | -0,987739367 | 0,323280309 | 0,999636303 | YES | 0 | FALSE |

| Ticker | taxLevel | Phylum | Family | Genus | Species |
|---|---|---|---|---|---|
| 537 | Genus | Firmicutes | Butyricicoccaceae | UCG-009 | NA |
| 538 | Genus | Firmicutes | Erysipelotrichaceae | Dielma | NA |
| 539 | Genus | Firmicutes | Acidaminococcaceae | Phascolarctobacterium | NA |
| 540 | Family | Actinobacteriota | Bifidobacteriaceae | NA | NA |
| 541 | Family | Verrucomicrobiota | Akkermansiaceae | NA | NA |
| 542 | Family | Thermoplasmatota | Methanomethylophilaceae | NA | NA |
| 543 | Family | Verrucomicrobiota | vadinBE97 | NA | NA |
| 544 | Family | Firmicutes | Veillonellaceae | NA | NA |
| 545 | Family | Bacteroidota | Muribaculaceae | NA | NA |
| 546 | Family | Firmicutes | Christensenellaceae | NA | NA |
| 547 | Family | Firmicutes | Peptostreptococcaceae | NA | NA |
| 548 | Family | Desulfobacterota | Desulfovibrionaceae | NA | NA |
| 549 | Family | Actinobacteriota | Coriobacteriaceae | NA | NA |
| 550 | Family | Proteobacteria | Pasteurellaceae | NA | NA |
| 551 | Family | Firmicutes | Erysipelatoclostridiaceae | NA | NA |
| 552 | Family | Bacteroidota | Marinifilaceae | NA | NA |
| 553 | Family | Bacteroidota | Barnesiellaceae | NA | NA |
| 554 | Family | Bacteroidota | Bacteroidaceae | NA | NA |
| 555 | Family | Bacteroidota | Prevotellaceae | NA | NA |
| 556 | Family | Firmicutes | Peptococcaceae | NA | NA |
| 557 | Family | Firmicutes | Streptococcaceae | NA | NA |
| 558 | Family | Firmicutes | Acholeplasmataceae | NA | NA |
| 559 | Family | Actinobacteriota | Coriobacteriales Incertae Sedis | NA | NA |
| 560 | Family | Euryarchaeota | Methanobacteriaceae | NA | NA |
| 561 | Family | Firmicutes | Clostridiaceae | NA | NA |
| 562 | Family | Firmicutes | Oscillospiraceae | NA | NA |
| 563 | Family | Proteobacteria | Enterobacteriaceae | NA | NA |
| 564 | Family | Bacteroidota | Rikenellaceae | NA | NA |
| 565 | Family | Firmicutes | Anaerovoracaceae | NA | NA |
| 566 | Family | Firmicutes | Selenomonadaceae | NA | NA |

| Ticker | DESeq2_baseMean | DESeq2_log2-FoldChange | DESeq2_lfcSE | DESeq2_stat | DESeq2_pvalue | DESeq2_padj | DESeq2 converged | ANCOM_W | ANCOM_detected_0.7 |
|---|---|---|---|---|---|---|---|---|---|
| 537 | 11,79079998 | 1,118191667 | 1,276583605 | 0,875925136 | 0,381070734 | 0,999636303 | YES | 0 | FALSE |
| 538 | 1,873897077 | -0,105353842 | 2,904840448 | -0,036268375 | 0,971068367 | 0,999636303 | YES | 0 | FALSE |
| 539 | 346,0890663 | -0,274787788 | 0,234685193 | -1,170878249 | 0,241647719 | 0,999636303 | YES | 0 | FALSE |
| 540 | 719,8268243 | 0,267602084 | 0,201934499 | 1,325192507 | 0,18510731 | 0,634653635 | NO | 0 | FALSE |
| 541 | 923,0790758 | -0,116451965 | 0,39599331 | -0,294075586 | 0,768700146 | 0,993663169 | YES | 0 | FALSE |
| 542 | 3,550815016 | 0,033515843 | 2,905520863 | 0,011535227 | 0,990796425 | 0,993663169 | YES | 0 | FALSE |
| 543 | 9,106987731 | 0,28914922 | 2,901557924 | 0,099653092 | 0,920619742 | 0,993663169 | YES | 0 | FALSE |
| 544 | 739,0395625 | 0,253827079 | 0,343099933 | 0,739805096 | 0,459418267 | 0,891945368 | YES | 0 | FALSE |
| 545 | 410,9691689 | 0,023633842 | 0,397185102 | 0,059503345 | 0,952551201 | 0,993663169 | YES | 0 | FALSE |
| 546 | 1338,01187 | 0,170294403 | 0,235885647 | 0,721936266 | 0,470333663 | 0,891945368 | YES | 0 | FALSE |
| 547 | 86,69618105 | 0,529011751 | 0,372647887 | 1,419602176 | 0,155723532 | 0,634653635 | YES | 0 | FALSE |
| 548 | 326,2821135 | -0,089568034 | 0,152024567 | -0,589168158 | 0,555748475 | 0,919859545 | YES | 0 | FALSE |
| 549 | 245,8784998 | -0,289499024 | 0,248508446 | -1,164946417 | 0,244040683 | 0,678087776 | YES | 0 | FALSE |
| 550 | 13,90091425 | -1,707213985 | 1,449626133 | -1,177692611 | 0,238919174 | 0,678087776 | YES | 0 | FALSE |
| 551 | 295,1800348 | -0,300690098 | 0,171845789 | -1,749767043 | 0,08015852 | 0,427512106 | YES | 0 | FALSE |
| 552 | 85,69374475 | -0,061119458 | 0,172579262 | -0,354152968 | 0,723224248 | 0,993663169 | YES | 0 | FALSE |
| 553 | 310,119534 | -0,48759986 | 0,239233045 | -2,038179385 | 0,041531996 | 0,305730961 | YES | 0 | FALSE |
| 554 | 5171,570003 | -0,251178527 | 0,119015277 | -2,11047298 | 0,034817636 | 0,305730961 | YES | 0 | FALSE |
| 555 | 1582,448966 | -0,619438205 | 0,299095561 | -2,071037775 | 0,038355266 | 0,305730961 | YES | 0 | FALSE |
| 556 | 87,25824975 | -0,225669446 | 0,318378271 | -0,708809195 | 0,478442891 | 0,891945368 | YES | 0 | FALSE |
| 557 | 291,678827 | 0,85982129 | 0,369861952 | 2,324708681 | 0,020087543 | 0,305730961 | YES | 0 | FALSE |
| 558 | 27,48862548 | -0,023064377 | 2,904056581 | -0,007942124 | 0,993663169 | 0,993663169 | YES | 0 | FALSE |
| 559 | 24,05154352 | 0,37511567 | 0,581627446 | 0,644941488 | 0,518965104 | 0,891945368 | YES | 0 | FALSE |
| 560 | 172,2411344 | 0,469356038 | 0,351454485 | 1,335467489 | 0,181723389 | 0,634653635 | YES | 0 | FALSE |
| 561 | 100,5839013 | 0,081091529 | 0,360897424 | 0,224694119 | 0,82221724 | 0,993663169 | YES | 0 | FALSE |
| 562 | 3604,729134 | -0,027556664 | 0,100294631 | -0,274757117 | 0,783502846 | 0,993663169 | YES | 0 | FALSE |
| 563 | 816,1672056 | -0,308589124 | 0,444845048 | -0,693700257 | 0,487870205 | 0,891945368 | YES | 0 | FALSE |
| 564 | 1311,209727 | -0,263084463 | 0,130982827 | -2,008541646 | 0,044585765 | 0,305730961 | YES | 0 | FALSE |
| 565 | 117,7214411 | 0,184139434 | 0,162966207 | 1,129924033 | 0,258508236 | 0,678087776 | YES | 0 | FALSE |
| 566 | 18,38767109 | -0,087745347 | 2,905102547 | -0,030203873 | 0,97590446 | 0,993663169 | YES | 0 | FALSE |

| Ticker | taxLevel | Phylum | Family | Genus | Species |
|---|---|---|---|---|---|
| 567 | Family | Bacteroidota | Tannerellaceae | NA | NA |
| 568 | Family | Firmicutes | Ruminococcaceae | NA | NA |
| 569 | Family | Firmicutes | Erysipelotrichaceae | NA | NA |
| 570 | Family | Firmicutes | Lactobacillaceae | NA | NA |
| 571 | Family | Firmicutes | UCG-010 | NA | NA |
| 572 | Family | Actinobacteriota | Atopobiaceae | NA | NA |
| 573 | Family | Verrucomicrobiota | Victivallaceae | NA | NA |
| 574 | Family | Actinobacteriota | Actinomycetaceae | NA | NA |
| 575 | Family | Proteobacteria | Oxalobacteraceae | NA | NA |
| 576 | Family | Firmicutes | Monoglobaceae | NA | NA |
| 577 | Family | Firmicutes | [Eubacterium] coprostanoligenes group | NA | NA |
| 578 | Family | Actinobacteriota | Eggerthellaceae | NA | NA |
| 579 | Family | Firmicutes | Butyricicoccaceae | NA | NA |
| 580 | Family | Firmicutes | Lachnospiraceae | NA | NA |
| 581 | Family | Firmicutes | Hydrogenoanaerobacterium | NA | NA |
| 582 | Family | Proteobacteria | Sutterellaceae | NA | NA |
| 583 | Family | Verrucomicrobiota | Puniceicoccaceae | NA | NA |
| 584 | Family | Firmicutes | Defluviitaleaceae | NA | NA |
| 585 | Family | Firmicutes | Ruminiclostridium | NA | NA |
| 586 | Family | Synergistota | Synergistaceae | NA | NA |
| 587 | Family | Firmicutes | Acidaminococcaceae | NA | NA |

| Ticker | DESeq2_baseMean | DESeq2_log2-FoldChange | DESeq2_lfcSE | DESeq2_stat | DESeq2_pvalue | DESeq2_padj | DESeq2 converged | ANCOM_W | ANCOM_detected_0.7 |
|---|---|---|---|---|---|---|---|---|---|
| 567 | 715,5677576 | -0,192933562 | 0,174327238 | -1,106732165 | 0,268409745 | 0,678087776 | YES | 0 | FALSE |
| 568 | 6452,345527 | -0,160043891 | 0,088849043 | -1,801301236 | 0,071655413 | 0,427512106 | YES | 0 | FALSE |
| 569 | 363,4789923 | -0,283347425 | 0,206131802 | -1,374593447 | 0,169257521 | 0,634653635 | YES | 0 | FALSE |
| 570 | 197,0007947 | 2,100616551 | 0,820258495 | 2,560920202 | 0,010439533 | 0,250548796 | YES | 0 | FALSE |
| 571 | 163,9145478 | 0,008891399 | 0,269861214 | 0,032948044 | 0,97371602 | 0,993663169 | YES | 0 | FALSE |
| 572 | 14,18352558 | -0,071117811 | 0,874906811 | -0,081286156 | 0,935214384 | 0,993663169 | YES | 0 | FALSE |
| 573 | 9,01369094 | -0,079758259 | 1,156429136 | -0,06896943 | 0,945013953 | 0,993663169 | YES | 0 | FALSE |
| 574 | 1,250716078 | 0,159910217 | 2,902813443 | 0,05508801 | 0,956068348 | 0,993663169 | YES | 0 | FALSE |
| 575 | 13,04688119 | 0,202180886 | 0,369409825 | 0,547307821 | 0,584167272 | 0,934667634 | YES | 0 | FALSE |
| 576 | 145,7697528 | 0,448407852 | 0,278400044 | 1,610660135 | 0,107253819 | 0,514818332 | YES | 0 | FALSE |
| 577 | 428,7437323 | 0,160461242 | 0,132342781 | 1,212466902 | 0,225333705 | 0,678087776 | YES | 0 | FALSE |
| 578 | 141,9186143 | -0,180823578 | 0,176710592 | -1,023275262 | 0,306177717 | 0,73482652 | YES | 0 | FALSE |
| 579 | 220,0873183 | -0,117508934 | 0,182784956 | -0,642880775 | 0,520301465 | 0,891945368 | YES | 0 | FALSE |
| 580 | 10920,74407 | -0,293540896 | 0,109802611 | -2,673350788 | 0,007509768 | 0,250548796 | YES | 0 | FALSE |
| 581 | 2,712422674 | -0,311560441 | 2,903788099 | -0,107294482 | 0,914555362 | 0,993663169 | YES | 0 | FALSE |
| 582 | 202,7427138 | -0,155045108 | 0,191725857 | -0,80868126 | 0,41869851 | 0,891945368 | YES | 0 | FALSE |
| 583 | 5,435988665 | -0,103215862 | 2,901063965 | -0,035578623 | 0,971618354 | 0,993663169 | YES | 0 | FALSE |
| 584 | 2,457580465 | 0,026873526 | 2,902787278 | 0,009257835 | 0,992613422 | 0,993663169 | YES | 0 | FALSE |
| 585 | 4,25990335 | 0,262069779 | 2,904483195 | 0,090229401 | 0,928104921 | 0,993663169 | YES | 0 | FALSE |
| 586 | 9,128634694 | 0,027368302 | 2,90331952 | 0,009426555 | 0,992478808 | 0,993663169 | YES | 0 | FALSE |
| 587 | 393,6239555 | -0,214693962 | 0,233280548 | -0,920325179 | 0,357402855 | 0,816920811 | YES | 0 | FALSE |

#### Supplementary Table 6: differential abundance at ASV, genus and family level in levodopa users with DESeq2 and ANCOM

**All columns are spread out over two pages, after which the next two pages show the next set of rows, etc.**

**A ticker is provided on both pages to easily connect the taxonomy data to the associated statistics.**

| Ticker | taxLevel | Phylum | Family | Genus | Species |
|---|---|---|---|---|---|
| 1 | ASV | Actinobacteriota | Bifidobacteriaceae | Bifidobacterium | NA |
| 2 | ASV | Verrucomicrobiota | Akkermansiaceae | Akkermansia | muciniphila |
| 3 | ASV | Firmicutes | Christensenellaceae | NA | NA |
| 4 | ASV | Firmicutes | Lachnospiraceae | Roseburia | NA |
| 5 | ASV | Firmicutes | NA | NA | NA |
| 6 | ASV | Firmicutes | NA | NA | NA |
| 7 | ASV | Firmicutes | Christensenellaceae | Christensenellaceae R-7 group | NA |
| 8 | ASV | Bacteroidota | Bacteroidaceae | Bacteroides | NA |
| 9 | ASV | Bacteroidota | Bacteroidaceae | Bacteroides | eggerthii |
| 10 | ASV | Firmicutes | UCG-010 | NA | NA |
| 11 | ASV | Firmicutes | [Eubacterium] coprostanoligenes group | NA | NA |
| 12 | ASV | Firmicutes | Ruminococcaceae | Subdoligranulum | NA |
| 13 | ASV | Firmicutes | Ruminococcaceae | Faecalibacterium | prausnitzii |
| 14 | ASV | Firmicutes | Lachnospiraceae | Lachnospira | pectinoschiza |
| 15 | ASV | Firmicutes | Oscillospiraceae | UCG-002 | NA |
| 16 | ASV | Firmicutes | Lachnospiraceae | NA | NA |
| 17 | ASV | Bacteroidota | Bacteroidaceae | Bacteroides | coprocola |
| 18 | ASV | Firmicutes | UCG-010 | NA | NA |
| 19 | ASV | Firmicutes | Ruminococcaceae | NA | NA |
| 20 | ASV | Firmicutes | Oscillospiraceae | UCG-003 | NA |
| 21 | ASV | Bacteroidota | Rikenellaceae | Alistipes | finegoldii |
| 22 | ASV | Firmicutes | NA | NA | NA |
| 23 | ASV | Firmicutes | Ruminococcaceae | Subdoligranulum | NA |
| 24 | ASV | Firmicutes | Veillonellaceae | Dialister | invisus |
| 25 | ASV | Firmicutes | Lachnospiraceae | Anaerostipes | hadrus |
| 26 | ASV | Bacteroidota | Barnesiellaceae | Barnesiella | NA |

| Ticker | DESeq2_baseMean | DESeq2_log2-FoldChange | DESeq2_lfcSE | DESeq2_stat | DESeq2_pvalue | DESeq2_padj | DESeq2 converged | ANCOM_W | ANCOM_detected_0.7 |
|---|---|---|---|---|---|---|---|---|---|
| 1 | 346,6327265 | -0,021273579 | 0,596309589 | -0,035675393 | 0,971541192 | 0,999299353 | YES | 0 | FALSE |
| 2 | 1016,441608 | 0,003849834 | 0,501323403 | 0,007679342 | 0,993872832 | 0,999299353 | YES | 0 | FALSE |
| 3 | 357,1733653 | 0,818310952 | 0,405112748 | 2,019958532 | 0,043387689 | 0,999299353 | YES | 0 | FALSE |
| 4 | 88,91672255 | 8,799021579 | 2,290605236 | 3,841352251 | 0,000122358 | 0,045125672 | YES | 0 | FALSE |
| 5 | 256,2142209 | -0,425917915 | 2,907957486 | -0,146466349 | 0,883553253 | 0,999299353 | YES | 0 | FALSE |
| 6 | 1,13833772 | 0,068342717 | 2,909405015 | 0,023490273 | 0,981259198 | 0,999299353 | YES | 0 | FALSE |
| 7 | 58,46974618 | -0,046882794 | 0,531189671 | -0,088259988 | 0,929670039 | 0,999299353 | YES | 0 | FALSE |
| 8 | 46,37553392 | -0,018375607 | 2,747895859 | -0,006687156 | 0,994664462 | 0,999299353 | YES | 0 | FALSE |
| 9 | 30,40443022 | -0,124550638 | 2,908826252 | -0,042818177 | 0,965846474 | 0,999299353 | YES | 0 | FALSE |
| 10 | 1,26539883 | 0,314305989 | 2,91249499 | 0,107916405 | 0,914062004 | 0,999299353 | YES | 0 | FALSE |
| 11 | 100,9978767 | -0,459154851 | 0,84379593 | -0,544153906 | 0,586335568 | 0,999299353 | YES | 0 | FALSE |
| 12 | 374,3666094 | -0,39798066 | 0,296629049 | -1,341677969 | 0,179700433 | 0,999299353 | YES | 0 | FALSE |
| 13 | 50,51980185 | -1,227524404 | 2,911537672 | -0,421606911 | 0,673311961 | 0,999299353 | YES | 0 | FALSE |
| 14 | 77,76089022 | -2,095141889 | 0,700661321 | -2,990234832 | 0,00278763 | 0,336998991 | YES | 0 | FALSE |
| 15 | 28,85511555 | 0,102347873 | 2,911554615 | 0,035152311 | 0,971958289 | 0,999299353 | YES | 0 | FALSE |
| 16 | 197,448331 | -0,118255739 | 2,907206043 | -0,040676766 | 0,967553585 | 0,999299353 | YES | 0 | FALSE |
| 17 | 54,90178431 | -0,355534773 | 2,912282933 | -0,122081124 | 0,902834771 | 0,999299353 | YES | 0 | FALSE |
| 18 | 3,02499994 | -0,115065239 | 2,911991606 | -0,039514276 | 0,968480372 | 0,999299353 | YES | 0 | FALSE |
| 19 | 1,638703655 | -0,177071148 | 2,91207667 | -0,060805799 | 0,951513872 | 0,999299353 | YES | 0 | FALSE |
| 20 | 130,9450931 | -0,564484397 | 0,468424698 | -1,205069671 | 0,228176407 | 0,999299353 | YES | 0 | FALSE |
| 21 | 33,60063308 | 0,571793819 | 2,907392896 | 0,196668919 | 0,844086634 | 0,999299353 | YES | 0 | FALSE |
| 22 | 398,3639327 | 0,488953756 | 2,907226889 | 0,16818562 | 0,866437254 | 0,999299353 | YES | 0 | FALSE |
| 23 | 59,90597499 | -0,314903267 | 0,824661407 | -0,38185765 | 0,702566954 | 0,999299353 | YES | 0 | FALSE |
| 24 | 276,5548356 | 0,176227905 | 2,179097097 | 0,080871984 | 0,935543761 | 0,999299353 | YES | 0 | FALSE |
| 25 | 9,332851223 | 6,059517425 | 2,910745659 | 2,081774959 | 0,03736303 | 0,999299353 | YES | 0 | FALSE |
| 26 | 37,59825428 | 0,090255361 | 2,912969289 | 0,030983973 | 0,975282322 | 0,999299353 | YES | 0 | FALSE |

| Ticker | taxLevel | Phylum | Family | Genus | Species |
|---|---|---|---|---|---|
| 27 | ASV | Firmicutes | Lachnospiraceae | [Ruminococcus] torques group | NA |
| 28 | ASV | Firmicutes | NA | NA | NA |
| 29 | ASV | Firmicutes | Oscillospiraceae | UCG-002 | NA |
| 30 | ASV | Bacteroidota | Tannerellaceae | Parabacteroides | distasonis |
| 31 | ASV | Firmicutes | Lachnospiraceae | Lachnospiraceae NK4A136 group | bacterium |
| 32 | ASV | Firmicutes | Oscillospiraceae | NK4A214 group | NA |
| 33 | ASV | Firmicutes | Christensenellaceae | Christensenellaceae R-7 group | NA |
| 34 | ASV | Firmicutes | Lachnospiraceae | Fusicatenibacter | saccharivorans |
| 35 | ASV | Proteobacteria | Enterobacteriaceae | Klebsiella | NA |
| 36 | ASV | Firmicutes | Lachnospiraceae | NA | NA |
| 37 | ASV | Firmicutes | Ruminococcaceae | CAG-352 | NA |
| 38 | ASV | Actinobacteriota | Eggerthellaceae | Senegalimassilia | anaerobia |
| 39 | ASV | Actinobacteriota | Eggerthellaceae | Enterorhabdus | NA |
| 40 | ASV | Bacteroidota | Rikenellaceae | Alistipes | inops |
| 41 | ASV | Firmicutes | Ruminococcaceae | NA | NA |
| 42 | ASV | Firmicutes | Butyricicoccaceae | Butyricicoccus | faecihominis |
| 43 | ASV | Bacteroidota | Rikenellaceae | Alistipes | obesi |
| 44 | ASV | Firmicutes | Christensenellaceae | Christensenellaceae R-7 group | NA |
| 45 | ASV | Firmicutes | Lachnospiraceae | Blautia | massiliensis |
| 46 | ASV | Bacteroidota | Tannerellaceae | Parabacteroides | distasonis |
| 47 | ASV | Firmicutes | Lachnospiraceae | Lachnoclostridium | NA |
| 48 | ASV | Firmicutes | Ruminococcaceae | Faecalibacterium | prausnitzii |
| 49 | ASV | Firmicutes | Peptostreptococcaceae | Terrisporobacter | mayombei |
| 50 | ASV | Desulfobacterota | Desulfovibrionaceae | Bilophila | wadsworthia |
| 51 | ASV | Bacteroidota | Muribaculaceae | NA | NA |
| 52 | ASV | Firmicutes | Erysipelotrichaceae | Turicibacter | sanguinis |
| 53 | ASV | Firmicutes | Peptococcaceae | NA | NA |
| 54 | ASV | Firmicutes | Oscillospiraceae | Oscillibacter | NA |
| 55 | ASV | Firmicutes | NA | NA | NA |
| 56 | ASV | Firmicutes | Oscillospiraceae | UCG-002 | NA |

| Ticker | DESeq2_baseMean | DESeq2_log2-FoldChange | DESeq2_lfcSE | DESeq2_stat | DESeq2_pvalue | DESeq2_padj | DESeq2 converged | ANCOM_W | ANCOM_detected_0.7 |
|---|---|---|---|---|---|---|---|---|---|
| 27 | 123,9750089 | -1,604191695 | 0,967597487 | -1,657912217 | 0,097335186 | 0,999299353 | YES | 0 | FALSE |
| 28 | 36,79873362 | 6,431904187 | 2,911296599 | 2,209291966 | 0,02715434 | 0,999299353 | YES | 0 | FALSE |
| 29 | 248,9458341 | 0,366541884 | 0,48055759 | 0,762742888 | 0,445616748 | 0,999299353 | YES | 0 | FALSE |
| 30 | 79,46445507 | -0,078544255 | 0,651066357 | -0,1206394 | 0,90397666 | 0,999299353 | YES | 0 | FALSE |
| 31 | 52,11686268 | -0,876855013 | 1,462108565 | -0,599719497 | 0,548693192 | 0,999299353 | YES | 0 | FALSE |
| 32 | 107,8757466 | 0,472625879 | 0,78955812 | 0,598595427 | 0,549442707 | 0,999299353 | YES | 0 | FALSE |
| 33 | 85,01391949 | -0,010396484 | 0,364833795 | -0,028496495 | 0,977266163 | 0,999299353 | YES | 0 | FALSE |
| 34 | 737,9910079 | -0,29593472 | 0,288184425 | -1,026893526 | 0,304470603 | 0,999299353 | YES | 0 | FALSE |
| 35 | 62,02797969 | 2,706184393 | 2,911288856 | 0,92954857 | 0,352604866 | 0,999299353 | YES | 0 | FALSE |
| 36 | 215,6612668 | 0,341182777 | 0,520138452 | 0,655946078 | 0,511858819 | 0,999299353 | YES | 0 | FALSE |
| 37 | 29,67807042 | -10,76312124 | 2,912617198 | -3,695343572 | 0,00021959 | 0,045125672 | YES | 0 | FALSE |
| 38 | 19,55149837 | 0,343595503 | 0,601131613 | 0,571581157 | 0,567605764 | 0,999299353 | YES | 0 | FALSE |
| 39 | 7,454505117 | 0,161841455 | 0,861704862 | 0,187815414 | 0,851021352 | 0,999299353 | YES | 0 | FALSE |
| 40 | 64,03685612 | 0,234403984 | 1,015835019 | 0,230750052 | 0,817508987 | 0,999299353 | YES | 0 | FALSE |
| 41 | 136,9658738 | 0,015736896 | 0,446759778 | 0,035224514 | 0,971900715 | 0,999299353 | YES | 0 | FALSE |
| 42 | 23,4462083 | 0,05270481 | 1,143401571 | 0,04609475 | 0,96323473 | 0,999299353 | YES | 0 | FALSE |
| 43 | 76,68153906 | -0,072874553 | 0,574925064 | -0,126754872 | 0,899134415 | 0,999299353 | YES | 0 | FALSE |
| 44 | 227,287366 | 1,46078687 | 0,571894593 | 2,554293901 | 0,01064034 | 0,600681553 | YES | 0 | FALSE |
| 45 | 367,1702874 | -0,056089148 | 0,33203886 | -0,168923445 | 0,865856858 | 0,999299353 | YES | 0 | FALSE |
| 46 | 45,6144019 | 0,054767513 | 1,940396577 | 0,028224907 | 0,977482772 | 0,999299353 | YES | 0 | FALSE |
| 47 | 51,77060175 | -0,290577652 | 0,727085558 | -0,399647124 | 0,689416442 | 0,999299353 | YES | 0 | FALSE |
| 48 | 114,1341006 | -0,032728616 | 1,799686748 | -0,018185729 | 0,985490687 | 0,999299353 | YES | 0 | FALSE |
| 49 | 7,13841624 | 0,458628097 | 2,909619322 | 0,157624777 | 0,874752479 | 0,999299353 | YES | 0 | FALSE |
| 50 | 50,9190978 | 0,48238532 | 0,333890928 | 1,444739225 | 0,148531148 | 0,999299353 | YES | 0 | FALSE |
| 51 | 87,98895088 | -0,027381046 | 2,908599806 | -0,009413824 | 0,992488966 | 0,999299353 | YES | 0 | FALSE |
| 52 | 11,42916362 | 0,397308187 | 2,907545455 | 0,136647283 | 0,8913096 | 0,999299353 | YES | 0 | FALSE |
| 53 | 1,555478949 | 0,274215278 | 2,910947443 | 0,094201384 | 0,924949185 | 0,999299353 | YES | 0 | FALSE |
| 54 | 37,25355638 | -0,088469571 | 0,592175352 | -0,149397591 | 0,881239912 | 0,999299353 | YES | 0 | FALSE |
| 55 | 57,36799493 | -1,239308339 | 1,310336261 | -0,945794127 | 0,344253602 | 0,999299353 | YES | 0 | FALSE |
| 56 | 86,06796294 | -0,895785221 | 1,654893092 | -0,541294918 | 0,588304323 | 0,999299353 | YES | 0 | FALSE |

| Ticker | taxLevel | Phylum | Family | Genus | Species |
|---|---|---|---|---|---|
| 57 | ASV | Firmicutes | Christensenellaceae | Christensenellaceae R-7 group | NA |
| 58 | ASV | Proteobacteria | Sutterellaceae | Sutterella | NA |
| 59 | ASV | Firmicutes | Butyricicoccaceae | Butyricicoccus | NA |
| 60 | ASV | Firmicutes | Lachnospiraceae | Roseburia | hominis |
| 61 | ASV | Firmicutes | Streptococcaceae | Streptococcus | NA |
| 62 | ASV | Firmicutes | NA | NA | NA |
| 63 | ASV | Firmicutes | Ruminococcaceae | Ruminococcus | NA |
| 64 | ASV | Bacteroidota | Prevotellaceae | Prevotella | NA |
| 65 | ASV | Firmicutes | Oscillospiraceae | UCG-005 | NA |
| 66 | ASV | Firmicutes | Lachnospiraceae | [Eubacterium] ventriosum group | NA |
| 67 | ASV | Firmicutes | Lachnospiraceae | Roseburia | NA |
| 68 | ASV | Firmicutes | Peptostreptococcaceae | Romboutsia | NA |
| 69 | ASV | Firmicutes | Ruminococcaceae | Ruminococcus | callidus |
| 70 | ASV | Firmicutes | Oscillospiraceae | UCG-002 | NA |
| 71 | ASV | Desulfobacterota | Desulfovibrionaceae | Desulfovibrio | NA |
| 72 | ASV | Bacteroidota | Tannerellaceae | Parabacteroides | distasonis |
| 73 | ASV | Firmicutes | Oscillospiraceae | Intestinimonas | NA |
| 74 | ASV | Firmicutes | Lachnospiraceae | Lachnoclostridium | edouardi |
| 75 | ASV | Bacteroidota | Barnesiellaceae | Barnesiella | NA |
| 76 | ASV | Actinobacteriota | Coriobacteriaceae | Collinsella | aerofaciens |
| 77 | ASV | Firmicutes | Oscillospiraceae | NA | NA |
| 78 | ASV | Proteobacteria | Pasteurellaceae | Haemophilus | NA |
| 79 | ASV | Actinobacteriota | Coriobacteriales Incertae Sedis | NA | NA |
| 80 | ASV | Firmicutes | NA | NA | NA |
| 81 | ASV | Bacteroidota | Prevotellaceae | Paraprevotella | NA |
| 82 | ASV | Firmicutes | NA | NA | NA |
| 83 | ASV | Firmicutes | Lachnospiraceae | Dorea | longicatena |
| 84 | ASV | Firmicutes | Ruminococcaceae | Ruminococcus | NA |
| 85 | ASV | Firmicutes | Ruminococcaceae | Ruminococcus | NA |
| 86 | ASV | Firmicutes | NA | NA | NA |

| Ticker | DESeq2_baseMean | DESeq2_log2-FoldChange | DESeq2_lfcSE | DESeq2_stat | DESeq2_pvalue | DESeq2_padj | DESeq2 converged | ANCOM_W | ANCOM_detected_0.7 |
|---|---|---|---|---|---|---|---|---|---|
| 57 | 637,6403702 | 0,446456579 | 0,333705236 | 1,337877057 | 0,180936517 | 0,999299353 | YES | 0 | FALSE |
| 58 | 23,06248383 | -0,168975883 | 1,36157093 | -0,124103621 | 0,901233231 | 0,999299353 | YES | 0 | FALSE |
| 59 | 52,62220893 | 0,092015134 | 0,83519721 | 0,110171745 | 0,91227317 | 0,999299353 | YES | 0 | FALSE |
| 60 | 55,16735837 | 0,296824061 | 1,056435181 | 0,280967603 | 0,778735248 | 0,999299353 | YES | 0 | FALSE |
| 61 | 15,96534437 | 0,266098118 | 2,909575414 | 0,091455996 | 0,92713027 | 0,999299353 | YES | 0 | FALSE |
| 62 | 22,58783515 | -0,467662348 | 2,910833249 | -0,160662707 | 0,872359064 | 0,999299353 | YES | 0 | FALSE |
| 63 | 25,12366646 | -0,074461662 | 2,909647082 | -0,025591304 | 0,979583322 | 0,999299353 | YES | 0 | FALSE |
| 64 | 24,01537456 | 0,240047801 | 2,908737753 | 0,08252645 | 0,934228086 | 0,999299353 | YES | 0 | FALSE |
| 65 | 67,23477775 | -0,480648771 | 0,511935484 | -0,938885438 | 0,347789567 | 0,999299353 | YES | 0 | FALSE |
| 66 | 62,51801145 | -0,099502345 | 0,572997248 | -0,173652397 | 0,862138651 | 0,999299353 | YES | 0 | FALSE |
| 67 | 53,00377714 | -0,443731486 | 0,945846107 | -0,469137085 | 0,638971652 | 0,999299353 | YES | 0 | FALSE |
| 68 | 38,42899062 | 0,448740458 | 0,861000087 | 0,521185148 | 0,602237799 | 0,999299353 | YES | 0 | FALSE |
| 69 | 115,3116721 | -1,382222261 | 2,816940628 | -0,490682071 | 0,623651329 | 0,999299353 | YES | 0 | FALSE |
| 70 | 14,2452233 | 0,391523966 | 2,908814961 | 0,134599131 | 0,892928829 | 0,999299353 | YES | 0 | FALSE |
| 71 | 164,5704062 | 0,137560679 | 0,590458505 | 0,232972644 | 0,815782648 | 0,999299353 | YES | 0 | FALSE |
| 72 | 27,56044501 | 0,072444954 | 2,912266139 | 0,024875801 | 0,980154029 | 0,999299353 | YES | 0 | FALSE |
| 73 | 36,744968 | 0,284383428 | 0,567582372 | 0,501043447 | 0,616340546 | 0,999299353 | YES | 0 | FALSE |
| 74 | 49,92685806 | -0,661926885 | 0,770246222 | -0,859370506 | 0,390136137 | 0,999299353 | YES | 0 | FALSE |
| 75 | 50,95215862 | -0,049660392 | 2,91221008 | -0,017052476 | 0,986394752 | 0,999299353 | YES | 0 | FALSE |
| 76 | 195,593335 | -0,287421042 | 0,340138042 | -0,845012926 | 0,398103622 | 0,999299353 | YES | 0 | FALSE |
| 77 | 15,72556168 | 0,214584536 | 2,911534899 | 0,073701516 | 0,941247892 | 0,999299353 | YES | 0 | FALSE |
| 78 | 8,543927258 | -0,001984263 | 2,259642652 | -0,000878131 | 0,999299353 | 0,999299353 | YES | 0 | FALSE |
| 79 | 3,275222135 | 0,213896874 | 2,912614171 | 0,073438108 | 0,941457494 | 0,999299353 | YES | 0 | FALSE |
| 80 | 25,94509867 | 0,152636706 | 2,907044996 | 0,052505794 | 0,958125679 | 0,999299353 | YES | 0 | FALSE |
| 81 | 40,13409453 | 0,264330054 | 2,908796171 | 0,090872663 | 0,927593772 | 0,999299353 | YES | 0 | FALSE |
| 82 | 37,65647991 | 0,245098396 | 2,90821946 | 0,08427782 | 0,932835547 | 0,999299353 | YES | 0 | FALSE |
| 83 | 526,8625731 | -0,101963876 | 0,313475176 | -0,32526938 | 0,744977203 | 0,999299353 | YES | 0 | FALSE |
| 84 | 68,35691256 | -2,06612958 | 2,908772351 | -0,710309825 | 0,477512028 | 0,999299353 | YES | 0 | FALSE |
| 85 | 66,43328135 | -0,542802294 | 2,910110089 | -0,186522941 | 0,85203469 | 0,999299353 | YES | 0 | FALSE |
| 86 | 68,78971624 | 0,129834891 | 2,908666787 | 0,044637251 | 0,96439645 | 0,999299353 | YES | 0 | FALSE |

| Ticker | taxLevel | Phylum | Family | Genus | Species |
|---|---|---|---|---|---|
| 87 | ASV | Firmicutes | Lachnospiraceae | [Ruminococcus] torques group | NA |
| 88 | ASV | Firmicutes | NA | NA | NA |
| 89 | ASV | Firmicutes | Ruminococcaceae | Ruminococcus | champanellensis |
| 90 | ASV | Firmicutes | Erysipelatoclostridiaceae | Erysipelotrichaceae UCG-003 | bacterium |
| 91 | ASV | Firmicutes | Ruminococcaceae | UBA1819 | NA |
| 92 | ASV | Firmicutes | Lachnospiraceae | Lachnospiraceae NK4A136 group | NA |
| 93 | ASV | Bacteroidota | Marinifilaceae | Odoribacter | splanchnicus |
| 94 | ASV | Actinobacteriota | Eggerthellaceae | Adlercreutzia | equolifaciens |
| 95 | ASV | Bacteroidota | Bacteroidaceae | Bacteroides | intestinalis |
| 96 | ASV | Bacteroidota | Barnesiellaceae | Barnesiella | intestinihominis |
| 97 | ASV | Firmicutes | Ruminococcaceae | Ruminococcus | NA |
| 98 | ASV | Bacteroidota | Bacteroidaceae | Bacteroides | eggerthii |
| 99 | ASV | Bacteroidota | Bacteroidaceae | Bacteroides | vulgatus |
| 100 | ASV | Bacteroidota | Prevotellaceae | Prevotella | copri |
| 101 | ASV | Firmicutes | Lachnospiraceae | [Eubacterium] xylanophilum group | NA |
| 102 | ASV | Firmicutes | Oscillospiraceae | UCG-002 | NA |
| 103 | ASV | Firmicutes | Lachnospiraceae | [Eubacterium] hallii group | NA |
| 104 | ASV | Firmicutes | NA | NA | NA |
| 105 | ASV | Firmicutes | NA | NA | NA |
| 106 | ASV | Firmicutes | Oscillospiraceae | Intestinimonas | NA |
| 107 | ASV | Firmicutes | Lachnospiraceae | Blautia | NA |
| 108 | ASV | Bacteroidota | Prevotellaceae | Prevotella | NA |
| 109 | ASV | Bacteroidota | Tannerellaceae | NA | NA |
| 110 | ASV | Firmicutes | Ruminococcaceae | Faecalibacterium | prausnitzii |
| 111 | ASV | Firmicutes | Lachnospiraceae | Lachnospiraceae FCS020 group | NA |
| 112 | ASV | Firmicutes | NA | NA | NA |
| 113 | ASV | Firmicutes | Lachnospiraceae | Lachnospiraceae UCG-010 | NA |
| 114 | ASV | Firmicutes | Oscillospiraceae | UCG-002 | NA |
| 115 | ASV | Firmicutes | Lachnospiraceae | Lachnospira | pectinoschiza |
| 116 | ASV | Firmicutes | Peptococcaceae | Peptococcus | NA |

| Ticker | DESeq2_baseMean | DESeq2_log2-FoldChange | DESeq2_lfcSE | DESeq2_stat | DESeq2_pvalue | DESeq2_padj | DESeq2 converged | ANCOM_W | ANCOM_detected_0.7 |
|---|---|---|---|---|---|---|---|---|---|
| 87 | 27,80198025 | 3,128193828 | 2,911042406 | 1,07459576 | 0,282555751 | 0,999299353 | YES | 0 | FALSE |
| 88 | 30,59069723 | -0,031155212 | 2,910823482 | -0,01070323 | 0,991460221 | 0,999299353 | YES | 0 | FALSE |
| 89 | 53,85886049 | 0,576354803 | 2,913652994 | 0,197811752 | 0,843192351 | 0,999299353 | YES | 0 | FALSE |
| 90 | 139,7331795 | -0,297975105 | 0,236721322 | -1,258759047 | 0,208117377 | 0,999299353 | YES | 0 | FALSE |
| 91 | 26,66292816 | -0,17656714 | 1,086669391 | -0,162484691 | 0,870924183 | 0,999299353 | YES | 0 | FALSE |
| 92 | 9,32575453 | 0,269265114 | 2,910967097 | 0,092500226 | 0,926300612 | 0,999299353 | YES | 0 | FALSE |
| 93 | 33,17019841 | 0,128506982 | 0,290192463 | 0,44283363 | 0,657886076 | 0,999299353 | YES | 0 | FALSE |
| 94 | 9,054189399 | -0,099209612 | 1,888194102 | -0,052542062 | 0,958096781 | 0,999299353 | YES | 0 | FALSE |
| 95 | 73,19466546 | 0,370080218 | 2,910797235 | 0,127140501 | 0,898829197 | 0,999299353 | YES | 0 | FALSE |
| 96 | 56,00017742 | -0,091375093 | 2,908726358 | -0,031414125 | 0,974939277 | 0,999299353 | YES | 0 | FALSE |
| 97 | 47,69546804 | 3,253940453 | 2,908159175 | 1,118900396 | 0,263182634 | 0,999299353 | YES | 0 | FALSE |
| 98 | 101,360087 | 0,050378809 | 2,912203399 | 0,017299207 | 0,986197918 | 0,999299353 | YES | 0 | FALSE |
| 99 | 1030,976376 | 0,005422985 | 0,227913883 | 0,023794009 | 0,981016919 | 0,999299353 | YES | 0 | FALSE |
| 100 | 292,2343386 | -0,968695746 | 1,808476941 | -0,535641746 | 0,592206171 | 0,999299353 | YES | 0 | FALSE |
| 101 | 23,50210386 | 0,012908893 | 1,960433646 | 0,006584713 | 0,994746197 | 0,999299353 | YES | 0 | FALSE |
| 102 | 29,39239845 | 0,506860306 | 2,906688422 | 0,17437724 | 0,861569 | 0,999299353 | YES | 0 | FALSE |
| 103 | 12,27098395 | -0,046929702 | 2,909589083 | -0,016129323 | 0,98713122 | 0,999299353 | YES | 0 | FALSE |
| 104 | 34,35032394 | -1,802520069 | 2,908767322 | -0,619685203 | 0,535465059 | 0,999299353 | YES | 0 | FALSE |
| 105 | 36,01031628 | -0,071983309 | 2,910755784 | -0,024730109 | 0,980270239 | 0,999299353 | YES | 0 | FALSE |
| 106 | 29,59105329 | 0,247915246 | 2,908004397 | 0,085252707 | 0,932060489 | 0,999299353 | YES | 0 | FALSE |
| 107 | 62,35934014 | 0,330655472 | 1,698362207 | 0,194690786 | 0,845635026 | 0,999299353 | YES | 0 | FALSE |
| 108 | 7,239848411 | 0,237956952 | 2,913188483 | 0,081682649 | 0,934899076 | 0,999299353 | YES | 0 | FALSE |
| 109 | 63,88848232 | 0,054239539 | 2,908671433 | 0,01864753 | 0,985122286 | 0,999299353 | YES | 0 | FALSE |
| 110 | 347,8353987 | -1,312225307 | 0,520455132 | -2,521303423 | 0,011692098 | 0,600681553 | YES | 237 | FALSE |
| 111 | 43,96681586 | -0,547744539 | 0,659001612 | -0,831173292 | 0,40587574 | 0,999299353 | YES | 0 | FALSE |
| 112 | 9,600944567 | 0,427483229 | 2,910280739 | 0,146887282 | 0,88322099 | 0,999299353 | YES | 0 | FALSE |
| 113 | 25,70293356 | 0,067868549 | 2,906064398 | 0,023354111 | 0,981367809 | 0,999299353 | YES | 0 | FALSE |
| 114 | 165,5478588 | 1,264795329 | 2,90797906 | 0,434939627 | 0,66360625 | 0,999299353 | YES | 0 | FALSE |
| 115 | 23,3952391 | -0,089833783 | 1,469768947 | -0,061121024 | 0,951262826 | 0,999299353 | YES | 0 | FALSE |
| 116 | 44,56053753 | 0,261982052 | 2,910181421 | 0,090022584 | 0,928269268 | 0,999299353 | YES | 0 | FALSE |

| Ticker | taxLevel | Phylum | Family | Genus | Species |
|---|---|---|---|---|---|
| 117 | ASV | Firmicutes | Anaerovoracaceae | Family XIII AD3011 group | NA |
| 118 | ASV | Firmicutes | Lachnospiraceae | Lachnospiraceae UCG-004 | NA |
| 119 | ASV | Bacteroidota | Bacteroidaceae | Bacteroides | uniformis |
| 120 | ASV | Firmicutes | Oscillospiraceae | Intestinimonas | NA |
| 121 | ASV | Firmicutes | Oscillospiraceae | NK4A214 group | NA |
| 122 | ASV | Bacteroidota | Barnesiellaceae | Barnesiella | NA |
| 123 | ASV | Bacteroidota | Muribaculaceae | NA | NA |
| 124 | ASV | Firmicutes | Lachnospiraceae | Lachnospiraceae UCG-001 | NA |
| 125 | ASV | Firmicutes | Oscillospiraceae | Flavonifractor | plautii |
| 126 | ASV | Firmicutes | Streptococcaceae | Streptococcus | NA |
| 127 | ASV | Firmicutes | Ruminococcaceae | [Eubacterium] siraeum group | NA |
| 128 | ASV | Proteobacteria | NA | NA | NA |
| 129 | ASV | Firmicutes | Acholeplasmataceae | Anaeroplasma | NA |
| 130 | ASV | Firmicutes | [Eubacterium] coprostanoligenes group | NA | NA |
| 131 | ASV | Cyanobacteria | NA | NA | NA |
| 132 | ASV | Bacteroidota | Bacteroidaceae | Bacteroides | clarus |
| 133 | ASV | Firmicutes | Butyricicoccaceae | NA | NA |
| 134 | ASV | Bacteroidota | Bacteroidaceae | Bacteroides | uniformis |
| 135 | ASV | Bacteroidota | Prevotellaceae | Alloprevotella | NA |
| 136 | ASV | Firmicutes | [Eubacterium] coprostanoligenes group | NA | NA |
| 137 | ASV | Firmicutes | Lachnospiraceae | Tyzzerella | NA |
| 138 | ASV | Firmicutes | NA | NA | NA |
| 139 | ASV | Bacteroidota | Bacteroidaceae | Bacteroides | coprophilus |
| 140 | ASV | Euryarchaeota | Methanobacteriaceae | Methanosphaera | NA |
| 141 | ASV | Firmicutes | Ruminococcaceae | Subdoligranulum | NA |
| 142 | ASV | Proteobacteria | NA | NA | NA |
| 143 | ASV | Actinobacteriota | Bifidobacteriaceae | Bifidobacterium | NA |
| 144 | ASV | Bacteroidota | Prevotellaceae | Paraprevotella | NA |
| 145 | ASV | Firmicutes | Oscillospiraceae | NK4A214 group | NA |
| 146 | ASV | Firmicutes | Erysipelotrichaceae | Solobacterium | NA |

| Ticker | DESeq2_baseMean | DESeq2_log2-FoldChange | DESeq2_lfcSE | DESeq2_stat | DESeq2_pvalue | DESeq2_padj | DESeq2 converged | ANCOM_W | ANCOM_detected_0.7 |
|---|---|---|---|---|---|---|---|---|---|
| 117 | 7,697372797 | -0,508251865 | 1,819877617 | -0,279278046 | 0,780031453 | 0,999299353 | YES | 0 | FALSE |
| 118 | 27,64704295 | -0,623136704 | 0,994737006 | -0,62643362 | 0,531030564 | 0,999299353 | YES | 0 | FALSE |
| 119 | 339,2459582 | 0,085447253 | 0,488986477 | 0,174743592 | 0,861281113 | 0,999299353 | YES | 0 | FALSE |
| 120 | 31,52528109 | -0,039449608 | 0,491998952 | -0,080182302 | 0,936092266 | 0,999299353 | YES | 0 | FALSE |
| 121 | 48,41260651 | 0,286322969 | 0,381974553 | 0,749586502 | 0,453503783 | 0,999299353 | YES | 0 | FALSE |
| 122 | 45,82330375 | 0,229763164 | 2,911546292 | 0,078914481 | 0,937100645 | 0,999299353 | YES | 0 | FALSE |
| 123 | 67,31078843 | 0,271601457 | 2,912950831 | 0,093239286 | 0,925713464 | 0,999299353 | YES | 0 | FALSE |
| 124 | 50,61471211 | -0,814793264 | 1,311503477 | -0,621266568 | 0,534424247 | 0,999299353 | YES | 0 | FALSE |
| 125 | 5,424195007 | -0,22488739 | 2,907267562 | -0,077353524 | 0,938342313 | 0,999299353 | YES | 0 | FALSE |
| 126 | 160,1730331 | 0,561387241 | 0,553459735 | 1,014323546 | 0,310428392 | 0,999299353 | YES | 0 | FALSE |
| 127 | 32,01737841 | -0,074800334 | 2,481405919 | -0,030144336 | 0,975951942 | 0,999299353 | YES | 0 | FALSE |
| 128 | 18,57610302 | 0,322565422 | 2,910139339 | 0,110841917 | 0,911741705 | 0,999299353 | YES | 0 | FALSE |
| 129 | 34,67554047 | 0,021931616 | 2,910837202 | 0,00753447 | 0,993988419 | 0,999299353 | YES | 0 | FALSE |
| 130 | 16,72179855 | 0,275086146 | 2,913681517 | 0,094411879 | 0,92478198 | 0,999299353 | YES | 0 | FALSE |
| 131 | 10,56994452 | -0,271044529 | 2,905145479 | -0,093298092 | 0,925666748 | 0,999299353 | YES | 0 | FALSE |
| 132 | 27,19033597 | 0,24175565 | 2,910208611 | 0,083071588 | 0,933794617 | 0,999299353 | YES | 0 | FALSE |
| 133 | 52,03263594 | 0,253082681 | 1,041530825 | 0,242991061 | 0,808012322 | 0,999299353 | YES | 0 | FALSE |
| 134 | 412,0140052 | -0,602657752 | 0,523583379 | -1,151025367 | 0,2497218 | 0,999299353 | YES | 0 | FALSE |
| 135 | 110,1361143 | 0,111828359 | 2,911491057 | 0,038409309 | 0,969361339 | 0,999299353 | YES | 0 | FALSE |
| 136 | 9,596872634 | 0,072973235 | 2,910159378 | 0,02507534 | 0,97999487 | 0,999299353 | YES | 0 | FALSE |
| 137 | 72,61139852 | 0,241138085 | 2,332094897 | 0,103399774 | 0,917645692 | 0,999299353 | YES | 0 | FALSE |
| 138 | 16,60882207 | -0,196104471 | 2,907988349 | -0,067436471 | 0,946234236 | 0,999299353 | YES | 0 | FALSE |
| 139 | 21,11198514 | 0,260163241 | 2,912960649 | 0,089312309 | 0,928833712 | 0,999299353 | YES | 0 | FALSE |
| 140 | 10,34906432 | 0,186838023 | 2,911487931 | 0,064172694 | 0,94883272 | 0,999299353 | YES | 0 | FALSE |
| 141 | 12,30986784 | 0,096050097 | 2,908092572 | 0,033028556 | 0,973651816 | 0,999299353 | YES | 0 | FALSE |
| 142 | 24,01745867 | 0,348607515 | 2,911748977 | 0,11972444 | 0,904701439 | 0,999299353 | YES | 0 | FALSE |
| 143 | 287,2858951 | 0,20362739 | 0,537410267 | 0,378904913 | 0,704758476 | 0,999299353 | YES | 0 | FALSE |
| 144 | 25,80915858 | 0,045180555 | 2,91161391 | 0,015517358 | 0,987619437 | 0,999299353 | YES | 0 | FALSE |
| 145 | 15,84550348 | 0,056158223 | 2,906427294 | 0,019322081 | 0,984584169 | 0,999299353 | YES | 0 | FALSE |
| 146 | 18,9083293 | 0,130609556 | 2,911520733 | 0,044859566 | 0,964219246 | 0,999299353 | YES | 0 | FALSE |

| Ticker | taxLevel | Phylum | Family | Genus | Species |
|---|---|---|---|---|---|
| 147 | ASV | Bacteroidota | Tannerellaceae | Parabacteroides | distasonis |
| 148 | ASV | Firmicutes | Lachnospiraceae | Lachnospira | NA |
| 149 | ASV | Actinobacteriota | Coriobacteriales Incertae Sedis | NA | NA |
| 150 | ASV | Firmicutes | Lachnospiraceae | Blautia | NA |
| 151 | ASV | Bacteroidota | Bacteroidaceae | Bacteroides | ovatus |
| 152 | ASV | Firmicutes | Lachnospiraceae | Lachnospiraceae AC2044 group | NA |
| 153 | ASV | Euryarchaeota | Methanobacteriaceae | Methanobrevibacter | NA |
| 154 | ASV | Firmicutes | Lactobacillaceae | Lactobacillus | delbrueckii |
| 155 | ASV | Firmicutes | Anaerovoracaceae | Family XIII UCG-001 | NA |
| 156 | ASV | Firmicutes | Ruminococcaceae | Incertae Sedis | NA |
| 157 | ASV | Firmicutes | Oscillospiraceae | Colidextribacter | NA |
| 158 | ASV | Firmicutes | Lachnospiraceae | Lachnoclostridium | NA |
| 159 | ASV | Firmicutes | Ruminococcaceae | Incertae Sedis | NA |
| 160 | ASV | Firmicutes | Ruminococcaceae | CAG-352 | NA |
| 161 | ASV | Firmicutes | Christensenellaceae | Christensenellaceae R-7 group | NA |
| 162 | ASV | Firmicutes | Oscillospiraceae | NK4A214 group | NA |
| 163 | ASV | Firmicutes | Erysipelotrichaceae | Catenisphaera | NA |
| 164 | ASV | Firmicutes | Oscillospiraceae | NK4A214 group | NA |
| 165 | ASV | Bacteroidota | Rikenellaceae | Alistipes | ihumii |
| 166 | ASV | Firmicutes | Clostridiaceae | Clostridium sensu stricto 1 | NA |
| 167 | ASV | Actinobacteriota | Eggerthellaceae | Slackia | isoflavoniconvertens |
| 168 | ASV | Firmicutes | Oscillospiraceae | NK4A214 group | NA |
| 169 | ASV | Firmicutes | Lachnospiraceae | NA | NA |
| 170 | ASV | Firmicutes | Christensenellaceae | Christensenellaceae R-7 group | NA |
| 171 | ASV | Firmicutes | Erysipelotrichaceae | NA | NA |
| 172 | ASV | Firmicutes | Oscillospiraceae | UCG-002 | NA |
| 173 | ASV | Firmicutes | Erysipelatoclostridiaceae | Erysipelatoclostridium | NA |
| 174 | ASV | Firmicutes | [Eubacterium] coprostanoligenes group | NA | NA |
| 175 | ASV | Firmicutes | Anaerovoracaceae | Mogibacterium | NA |
| 176 | ASV | Firmicutes | Ruminococcaceae | Negativibacillus | NA |

| Ticker | DESeq2_baseMean | DESeq2_log2-FoldChange | DESeq2_lfcSE | DESeq2_stat | DESeq2_pvalue | DESeq2_padj | DESeq2 converged | ANCOM_W | ANCOM_detected_0.7 |
|---|---|---|---|---|---|---|---|---|---|
| 147 | 30,01116406 | 0,090139125 | 2,910869786 | 0,030966389 | 0,975296345 | 0,999299353 | YES | 0 | FALSE |
| 148 | 34,19366839 | -0,045077675 | 2,90532844 | -0,015515518 | 0,987620905 | 0,999299353 | YES | 0 | FALSE |
| 149 | 5,952421825 | 0,363349242 | 2,910760564 | 0,124829657 | 0,900658408 | 0,999299353 | YES | 0 | FALSE |
| 150 | 9,246885654 | 0,411319643 | 2,905641161 | 0,141558995 | 0,887428358 | 0,999299353 | YES | 0 | FALSE |
| 151 | 12,87202367 | -0,015819157 | 2,912240276 | -0,005431954 | 0,995665949 | 0,999299353 | YES | 0 | FALSE |
| 152 | 74,80869052 | 0,14803533 | 1,248286554 | 0,118590823 | 0,905599536 | 0,999299353 | YES | 0 | FALSE |
| 153 | 218,7222979 | 0,86514281 | 0,449579892 | 1,924336086 | 0,05431247 | 0,999299353 | YES | 0 | FALSE |
| 154 | 8,552884092 | -0,05802592 | 2,912393323 | -0,019923792 | 0,984104165 | 0,999299353 | YES | 0 | FALSE |
| 155 | 8,730686227 | 0,383173905 | 2,910663099 | 0,131644884 | 0,895265182 | 0,999299353 | YES | 0 | FALSE |
| 156 | 7,341634343 | 0,212530856 | 2,908358135 | 0,073075889 | 0,941745728 | 0,999299353 | YES | 0 | FALSE |
| 157 | 26,3743812 | 1,728214807 | 0,587784233 | 2,940219745 | 0,003279796 | 0,336998991 | YES | 20 | FALSE |
| 158 | 23,88138539 | -0,808551563 | 0,839360984 | -0,963294195 | 0,335399906 | 0,999299353 | YES | 0 | FALSE |
| 159 | 12,64849219 | 0,51604484 | 1,828164151 | 0,282274893 | 0,777732736 | 0,999299353 | YES | 0 | FALSE |
| 160 | 348,6736511 | -1,011402487 | 2,904440471 | -0,348226275 | 0,727670256 | 0,999299353 | YES | 0 | FALSE |
| 161 | 7,869461939 | 0,238379373 | 2,908288851 | 0,081965508 | 0,934674142 | 0,999299353 | YES | 0 | FALSE |
| 162 | 42,70925267 | 0,351108996 | 2,014253587 | 0,174312211 | 0,861620103 | 0,999299353 | YES | 0 | FALSE |
| 163 | 2,3850677 | 0,132196971 | 2,913252005 | 0,045377801 | 0,963806175 | 0,999299353 | YES | 0 | FALSE |
| 164 | 27,83267671 | 2,665104494 | 2,911377011 | 0,915410297 | 0,359976275 | 0,999299353 | YES | 0 | FALSE |
| 165 | 14,3535372 | -0,08109143 | 2,905893783 | -0,027905848 | 0,977737244 | 0,999299353 | YES | 0 | FALSE |
| 166 | 67,08498444 | 0,916286423 | 0,704626773 | 1,300385479 | 0,193468884 | 0,999299353 | YES | 21 | FALSE |
| 167 | 10,45285503 | -0,362127704 | 1,007791059 | -0,359328157 | 0,719349613 | 0,999299353 | YES | 0 | FALSE |
| 168 | 8,575826931 | -0,071336844 | 2,911184237 | -0,024504407 | 0,980450268 | 0,999299353 | YES | 0 | FALSE |
| 169 | 27,71648061 | -0,107941392 | 2,909330414 | -0,037101799 | 0,970403838 | 0,999299353 | YES | 0 | FALSE |
| 170 | 36,29230641 | -0,328858783 | 0,880389345 | -0,373537896 | 0,708748146 | 0,999299353 | YES | 0 | FALSE |
| 171 | 16,83887458 | 0,393515571 | 2,905408026 | 0,135442446 | 0,892262066 | 0,999299353 | YES | 0 | FALSE |
| 172 | 631,2624844 | 0,137849641 | 0,45594053 | 0,302341274 | 0,762391917 | 0,999299353 | YES | 0 | FALSE |
| 173 | 1,948383656 | 0,056380677 | 2,912069906 | 0,019361031 | 0,984553097 | 0,999299353 | YES | 0 | FALSE |
| 174 | 42,35700937 | 0,245453616 | 2,910828176 | 0,084324323 | 0,932798574 | 0,999299353 | YES | 0 | FALSE |
| 175 | 24,16835401 | 0,140483794 | 2,90942519 | 0,048285756 | 0,961488507 | 0,999299353 | YES | 0 | FALSE |
| 176 | 13,81514126 | 0,312579376 | 2,905620964 | 0,107577478 | 0,914330863 | 0,999299353 | YES | 0 | FALSE |

| Ticker | taxLevel | Phylum | Family | Genus | Species |
|---|---|---|---|---|---|
| 177 | ASV | Proteobacteria | Sutterellaceae | Sutterella | NA |
| 178 | ASV | Firmicutes | Streptococcaceae | Lactococcus | NA |
| 179 | ASV | Firmicutes | Ruminococcaceae | Subdoligranulum | NA |
| 180 | ASV | Proteobacteria | Enterobacteriaceae | Escherichia-Shigella | NA |
| 181 | ASV | Firmicutes | Lachnospiraceae | GCA-900066575 | NA |
| 182 | ASV | Firmicutes | Lachnospiraceae | Lachnospiraceae UCG-004 | NA |
| 183 | ASV | Firmicutes | Veillonellaceae | Veillonella | NA |
| 184 | ASV | Firmicutes | Lachnospiraceae | Lachnoclostridium | NA |
| 185 | ASV | Firmicutes | Christensenellaceae | NA | NA |
| 186 | ASV | Firmicutes | Oscillospiraceae | Colidextribacter | massiliensis |
| 187 | ASV | Bacteroidota | Rikenellaceae | Alistipes | putredinis |
| 188 | ASV | Firmicutes | Lachnospiraceae | NA | NA |
| 189 | ASV | Firmicutes | Ruminococcaceae | Subdoligranulum | NA |
| 190 | ASV | Firmicutes | Anaerovoracaceae | Family XIII UCG-001 | NA |
| 191 | ASV | Firmicutes | NA | NA | NA |
| 192 | ASV | Firmicutes | Oscillospiraceae | UCG-002 | NA |
| 193 | ASV | Firmicutes | Christensenellaceae | Christensenellaceae R-7 group | NA |
| 194 | ASV | Firmicutes | Anaerovoracaceae | Family XIII AD3011 group | NA |
| 195 | ASV | Desulfobacterota | Desulfovibrionaceae | Desulfovibrio | piger |
| 196 | ASV | Proteobacteria | NA | NA | NA |
| 197 | ASV | Proteobacteria | NA | NA | NA |
| 198 | ASV | Firmicutes | Anaerovoracaceae | [Eubacterium] brachy group | NA |
| 199 | ASV | Firmicutes | Lachnospiraceae | NA | NA |
| 200 | ASV | Firmicutes | Peptostreptococcaceae | Intestinibacter | bartlettii |
| 201 | ASV | Firmicutes | Lachnospiraceae | [Eubacterium] ventriosum group | NA |
| 202 | ASV | Bacteroidota | Barnesiellaceae | NA | NA |
| 203 | ASV | Firmicutes | Lachnospiraceae | Lachnoclostridium | NA |
| 204 | ASV | Firmicutes | Lachnospiraceae | Lachnotalea | NA |
| 205 | ASV | Firmicutes | Monoglobaceae | Monoglobus | pectinilyticus |
| 206 | ASV | Firmicutes | Lachnospiraceae | Anaerostipes | hadrus |

| Ticker | DESeq2_baseMean | DESeq2_log2-FoldChange | DESeq2_lfcSE | DESeq2_stat | DESeq2_pvalue | DESeq2_padj | DESeq2 converged | ANCOM_W | ANCOM_detected_0.7 |
|---|---|---|---|---|---|---|---|---|---|
| 177 | 19,24472386 | -0,016669143 | 2,910912054 | -0,005726433 | 0,995430992 | 0,999299353 | YES | 0 | FALSE |
| 178 | 8,470308428 | 0,486492275 | 2,910530013 | 0,167149032 | 0,867252786 | 0,999299353 | YES | 0 | FALSE |
| 179 | 531,6351586 | 0,388108298 | 0,320753641 | 1,209988752 | 0,226283209 | 0,999299353 | YES | 0 | FALSE |
| 180 | 723,4835665 | -0,471507938 | 0,600690154 | -0,784943678 | 0,432486592 | 0,999299353 | YES | 0 | FALSE |
| 181 | 7,912403662 | -0,140226784 | 2,240777958 | -0,062579509 | 0,950101347 | 0,999299353 | YES | 0 | FALSE |
| 182 | 9,88773165 | -0,523307316 | 2,907017662 | -0,180015183 | 0,857140649 | 0,999299353 | YES | 0 | FALSE |
| 183 | 10,79563595 | 0,109602844 | 2,909809769 | 0,03766667 | 0,969953451 | 0,999299353 | YES | 0 | FALSE |
| 184 | 4,134367288 | 0,358513363 | 2,911199607 | 0,123149702 | 0,901988555 | 0,999299353 | YES | 0 | FALSE |
| 185 | 12,76608642 | 0,355070914 | 2,90946088 | 0,122040106 | 0,902867256 | 0,999299353 | YES | 0 | FALSE |
| 186 | 26,1628603 | -0,944650004 | 0,714023591 | -1,322995508 | 0,185836872 | 0,999299353 | YES | 0 | FALSE |
| 187 | 273,8873852 | -0,474428246 | 0,427720774 | -1,109200852 | 0,267343545 | 0,999299353 | YES | 0 | FALSE |
| 188 | 8,651357003 | -0,596917238 | 2,911059474 | -0,205051543 | 0,837531855 | 0,999299353 | YES | 167 | FALSE |
| 189 | 16,743465 | 0,352788686 | 2,906865832 | 0,121363939 | 0,903402779 | 0,999299353 | YES | 0 | FALSE |
| 190 | 13,36751846 | -0,123847973 | 1,680391769 | -0,073701845 | 0,941247631 | 0,999299353 | YES | 0 | FALSE |
| 191 | 13,39252942 | 0,3341766 | 2,905875584 | 0,115000312 | 0,908444874 | 0,999299353 | YES | 0 | FALSE |
| 192 | 541,6644648 | -0,617181674 | 0,409390557 | -1,507562067 | 0,131666647 | 0,999299353 | YES | 0 | FALSE |
| 193 | 28,60355617 | 0,435632467 | 0,613876145 | 0,709642279 | 0,477925994 | 0,999299353 | YES | 0 | FALSE |
| 194 | 30,22859143 | 0,027318075 | 0,589411662 | 0,046348039 | 0,963032851 | 0,999299353 | YES | 0 | FALSE |
| 195 | 42,22204903 | 0,053180608 | 2,912242188 | 0,018261053 | 0,985430598 | 0,999299353 | YES | 0 | FALSE |
| 196 | 13,18180794 | 0,071690103 | 2,909017823 | 0,024644092 | 0,980338849 | 0,999299353 | YES | 0 | FALSE |
| 197 | 14,90153933 | -0,129933893 | 2,906367636 | -0,044706627 | 0,964341152 | 0,999299353 | YES | 0 | FALSE |
| 198 | 0,554566966 | 0,307942189 | 2,914884652 | 0,105644726 | 0,915864237 | 0,999299353 | YES | 0 | FALSE |
| 199 | 3,227624179 | -0,10634843 | 2,909242774 | -0,036555364 | 0,970839534 | 0,999299353 | YES | 0 | FALSE |
| 200 | 35,22493639 | 0,647451009 | 0,834735792 | 0,775635853 | 0,437964025 | 0,999299353 | YES | 0 | FALSE |
| 201 | 12,47942199 | -0,079233872 | 2,907607312 | -0,027250541 | 0,978259905 | 0,999299353 | YES | 0 | FALSE |
| 202 | 21,92437397 | 0,005414265 | 1,81062047 | 0,002990281 | 0,997614104 | 0,999299353 | YES | 0 | FALSE |
| 203 | 14,93406139 | -0,010184227 | 1,117444935 | -0,009113851 | 0,9927283 | 0,999299353 | YES | 0 | FALSE |
| 204 | 3,92166846 | -0,606833964 | 2,907822408 | -0,208690174 | 0,834690111 | 0,999299353 | YES | 0 | FALSE |
| 205 | 9,771837182 | -1,758915548 | 2,911685932 | -0,604088349 | 0,545784905 | 0,999299353 | YES | 0 | FALSE |
| 206 | 132,8820657 | -0,404735218 | 0,446687661 | -0,906081033 | 0,364892963 | 0,999299353 | YES | 0 | FALSE |

| Ticker | taxLevel | Phylum | Family | Genus | Species |
|---|---|---|---|---|---|
| 207 | ASV | Firmicutes | Christensenellaceae | Christensenellaceae R-7 group | NA |
| 208 | ASV | Firmicutes | Lachnospiraceae | NA | NA |
| 209 | ASV | Bacteroidota | Tannerellaceae | Parabacteroides | NA |
| 210 | ASV | Firmicutes | Lachnospiraceae | Agathobacter | NA |
| 211 | ASV | Firmicutes | Lachnospiraceae | Roseburia | NA |
| 212 | ASV | Firmicutes | Lachnospiraceae | [Eubacterium] xylanophilum group | NA |
| 213 | ASV | Firmicutes | Ruminococcaceae | Negativibacillus | NA |
| 214 | ASV | Firmicutes | NA | NA | NA |
| 215 | ASV | Firmicutes | Lachnospiraceae | Marvinbryantia | NA |
| 216 | ASV | Firmicutes | Lachnospiraceae | Blautia | obeum |
| 217 | ASV | Firmicutes | [Eubacterium] coprostanoligenes group | NA | NA |
| 218 | ASV | Firmicutes | Ruminococcaceae | NA | NA |
| 219 | ASV | Bacteroidota | Rikenellaceae | Alistipes | NA |
| 220 | ASV | Bacteroidota | Rikenellaceae | Alistipes | NA |
| 221 | ASV | Firmicutes | Lachnospiraceae | [Eubacterium] hallii group | NA |
| 222 | ASV | Firmicutes | UCG-010 | NA | NA |
| 223 | ASV | Firmicutes | NA | NA | NA |
| 224 | ASV | Bacteroidota | Barnesiellaceae | Coprobacter | fastidiosus |
| 225 | ASV | Firmicutes | Clostridiaceae | Clostridium sensu stricto 1 | NA |
| 226 | ASV | Firmicutes | Christensenellaceae | Christensenellaceae R-7 group | NA |
| 227 | ASV | Firmicutes | NA | NA | NA |
| 228 | ASV | Firmicutes | NA | NA | NA |
| 229 | ASV | Firmicutes | Selenomonadaceae | Mitsuokella | jalaludinii |
| 230 | ASV | Firmicutes | Ruminococcaceae | DTU089 | NA |
| 231 | ASV | Firmicutes | Lachnospiraceae | NA | NA |
| 232 | ASV | Bacteroidota | Tannerellaceae | Parabacteroides | merdae |
| 233 | ASV | Bacteroidota | Rikenellaceae | Alistipes | indistinctus |
| 234 | ASV | Firmicutes | Ruminococcaceae | Ruminococcus | bromii |
| 235 | ASV | Firmicutes | Erysipelotrichaceae | Holdemanella | NA |
| 236 | ASV | Bacteroidota | Rikenellaceae | Rikenellaceae RC9 gut group | NA |

| Ticker | DESeq2_baseMean | DESeq2_log2-FoldChange | DESeq2_lfcSE | DESeq2_stat | DESeq2_pvalue | DESeq2_padj | DESeq2 converged | ANCOM_W | ANCOM_detected_0.7 |
|---|---|---|---|---|---|---|---|---|---|
| 207 | 8,220221102 | 0,220315652 | 2,909648011 | 0,075719005 | 0,939642656 | 0,999299353 | YES | 0 | FALSE |
| 208 | 2,705995867 | 0,20823481 | 2,913141278 | 0,071481192 | 0,943014793 | 0,999299353 | YES | 0 | FALSE |
| 209 | 32,52636146 | 0,182945616 | 2,913649945 | 0,062789154 | 0,949934402 | 0,999299353 | YES | 0 | FALSE |
| 210 | 320,0109162 | -0,81502665 | 0,948281866 | -0,859477208 | 0,390077289 | 0,999299353 | YES | 0 | FALSE |
| 211 | 20,64397058 | -0,250201165 | 2,905284139 | -0,086119344 | 0,931371546 | 0,999299353 | YES | 0 | FALSE |
| 212 | 9,948554992 | 0,072136678 | 2,905486257 | 0,024827747 | 0,980192359 | 0,999299353 | YES | 0 | FALSE |
| 213 | 11,61358952 | -0,076173238 | 2,907097232 | -0,026202508 | 0,979095816 | 0,999299353 | YES | 0 | FALSE |
| 214 | 18,11799983 | -0,116942604 | 2,912187556 | -0,040156275 | 0,967968537 | 0,999299353 | YES | 0 | FALSE |
| 215 | 35,49744459 | 0,569999301 | 0,854594556 | 0,666982135 | 0,504783545 | 0,999299353 | YES | 0 | FALSE |
| 216 | 195,4902888 | -0,419385771 | 0,415531537 | -1,009275433 | 0,312842559 | 0,999299353 | YES | 0 | FALSE |
| 217 | 22,52490053 | 0,17276685 | 2,911509304 | 0,059339275 | 0,95268188 | 0,999299353 | YES | 0 | FALSE |
| 218 | 11,23850092 | -0,306397364 | 0,947950853 | -0,323220728 | 0,746528084 | 0,999299353 | YES | 0 | FALSE |
| 219 | 8,9381144 | 0,558622049 | 2,909071582 | 0,192027605 | 0,847720582 | 0,999299353 | YES | 0 | FALSE |
| 220 | 16,41997627 | 0,04336229 | 2,909469279 | 0,014903849 | 0,988108889 | 0,999299353 | YES | 0 | FALSE |
| 221 | 143,4480555 | -0,276659389 | 0,299629434 | -0,923338491 | 0,355830835 | 0,999299353 | YES | 0 | FALSE |
| 222 | 12,43737457 | -0,528533252 | 1,721852764 | -0,306956125 | 0,758876768 | 0,999299353 | YES | 0 | FALSE |
| 223 | 17,72672335 | 0,285830766 | 2,910867362 | 0,098194363 | 0,921777959 | 0,999299353 | YES | 0 | FALSE |
| 224 | 7,537075577 | -0,182524908 | 2,182885688 | -0,083616338 | 0,933361477 | 0,999299353 | YES | 0 | FALSE |
| 225 | 9,162474875 | -0,031700435 | 2,907525865 | -0,01090289 | 0,991300925 | 0,999299353 | YES | 0 | FALSE |
| 226 | 3,42743513 | 0,172649728 | 2,906290035 | 0,05940554 | 0,952629101 | 0,999299353 | YES | 0 | FALSE |
| 227 | 20,82591292 | 0,064169865 | 2,908763984 | 0,02206087 | 0,9823994 | 0,999299353 | YES | 0 | FALSE |
| 228 | 29,78158179 | 0,364167997 | 2,909481732 | 0,125165934 | 0,900392186 | 0,999299353 | YES | 0 | FALSE |
| 229 | 19,59731936 | -0,013696886 | 2,912268403 | -0,004703167 | 0,996247429 | 0,999299353 | YES | 0 | FALSE |
| 230 | 7,625424248 | 0,33411902 | 1,766316073 | 0,189161512 | 0,849966232 | 0,999299353 | YES | 0 | FALSE |
| 231 | 12,27709941 | -1,156155191 | 2,905987916 | -0,397852718 | 0,690738755 | 0,999299353 | YES | 274 | FALSE |
| 232 | 331,2558741 | 0,049661748 | 0,235954337 | 0,210471858 | 0,833299414 | 0,999299353 | YES | 0 | FALSE |
| 233 | 6,162876947 | 0,600041214 | 1,965344663 | 0,305310934 | 0,760129349 | 0,999299353 | YES | 0 | FALSE |
| 234 | 1141,850194 | 0,583909737 | 0,596521477 | 0,978857861 | 0,327650215 | 0,999299353 | YES | 0 | FALSE |
| 235 | 223,2780932 | 0,08724833 | 0,759112841 | 0,114934599 | 0,908496961 | 0,999299353 | YES | 0 | FALSE |
| 236 | 25,91588437 | 0,224005845 | 2,907486932 | 0,077044489 | 0,938588153 | 0,999299353 | YES | 0 | FALSE |

| Ticker | taxLevel | Phylum | Family | Genus | Species |
|---|---|---|---|---|---|
| 237 | ASV | Firmicutes | Oscillospiraceae | Colidextribacter | NA |
| 238 | ASV | Firmicutes | NA | NA | NA |
| 239 | ASV | Firmicutes | Ruminococcaceae | Ruminococcus | bicirculans |
| 240 | ASV | Firmicutes | Oscillospiraceae | Oscillibacter | NA |
| 241 | ASV | Firmicutes | Oscillospiraceae | UCG-003 | NA |
| 242 | ASV | Firmicutes | Oscillospiraceae | Oscillospira | NA |
| 243 | ASV | Firmicutes | UCG-010 | NA | NA |
| 244 | ASV | Actinobacteriota | Bifidobacteriaceae | Bifidobacterium | NA |
| 245 | ASV | Firmicutes | NA | NA | NA |
| 246 | ASV | Firmicutes | Lachnospiraceae | Lachnospiraceae FCS020 group | bacterium |
| 247 | ASV | Firmicutes | Christensenellaceae | Christensenellaceae R-7 group | NA |
| 248 | ASV | Firmicutes | Oscillospiraceae | NK4A214 group | NA |
| 249 | ASV | Firmicutes | Lactobacillaceae | Lactobacillus | NA |
| 250 | ASV | Firmicutes | Lachnospiraceae | Blautia | faecis |
| 251 | ASV | Bacteroidota | Marinifilaceae | Butyricimonas | virosa |
| 252 | ASV | Firmicutes | UCG-010 | NA | NA |
| 253 | ASV | Firmicutes | Christensenellaceae | Christensenellaceae R-7 group | NA |
| 254 | ASV | Firmicutes | Christensenellaceae | Christensenellaceae R-7 group | NA |
| 255 | ASV | Firmicutes | Christensenellaceae | Christensenellaceae R-7 group | NA |
| 256 | ASV | Firmicutes | UCG-010 | NA | NA |
| 257 | ASV | Bacteroidota | Bacteroidaceae | Bacteroides | NA |
| 258 | ASV | Firmicutes | [Eubacterium] coprostanoligenes group | NA | NA |
| 259 | ASV | Firmicutes | Veillonellaceae | Veillonella | NA |
| 260 | ASV | Firmicutes | Erysipelatoclostridiaceae | Coprobacillus | cateniformis |
| 261 | ASV | Firmicutes | Oscillospiraceae | UCG-005 | NA |
| 262 | ASV | Firmicutes | Christensenellaceae | Christensenellaceae R-7 group | NA |
| 263 | ASV | Actinobacteriota | Atopobiaceae | Olsenella | NA |
| 264 | ASV | Firmicutes | Lachnospiraceae | Coprococcus | comes |
| 265 | ASV | Firmicutes | NA | NA | NA |
| 266 | ASV | Firmicutes | Oscillospiraceae | UCG-005 | NA |

| Ticker | DESeq2_baseMean | DESeq2_log2-FoldChange | DESeq2_lfcSE | DESeq2_stat | DESeq2_pvalue | DESeq2_padj | DESeq2 converged | ANCOM_W | ANCOM_detected_0.7 |
|---|---|---|---|---|---|---|---|---|---|
| 237 | 9,989651428 | -0,862039131 | 1,086801683 | -0,793188992 | 0,42766772 | 0,999299353 | YES | 87 | FALSE |
| 238 | 8,77073113 | -0,132223419 | 0,710508845 | -0,186096796 | 0,852368855 | 0,999299353 | YES | 0 | FALSE |
| 239 | 199,656055 | -0,3173419 | 0,833842695 | -0,380577658 | 0,703516662 | 0,999299353 | YES | 0 | FALSE |
| 240 | 2,121682783 | 0,395724185 | 2,914598623 | 0,135773133 | 0,892000631 | 0,999299353 | YES | 0 | FALSE |
| 241 | 4,265605972 | 0,11417967 | 2,911170267 | 0,039221227 | 0,96871401 | 0,999299353 | YES | 0 | FALSE |
| 242 | 9,415592399 | 0,557542872 | 2,909156619 | 0,191651033 | 0,848015565 | 0,999299353 | YES | 0 | FALSE |
| 243 | 20,03444959 | 0,440462191 | 2,3302386 | 0,189020211 | 0,850076977 | 0,999299353 | YES | 6 | FALSE |
| 244 | 45,15141618 | -0,513440523 | 2,911553378 | -0,176345908 | 0,860022198 | 0,999299353 | YES | 0 | FALSE |
| 245 | 26,44148839 | -1,446411398 | 2,906929593 | -0,497573592 | 0,618784621 | 0,999299353 | YES | 0 | FALSE |
| 246 | 5,488811908 | 0,287044721 | 2,9103492 | 0,098628962 | 0,921432874 | 0,999299353 | YES | 0 | FALSE |
| 247 | 4,933840203 | -0,032635956 | 2,906666438 | -0,011227967 | 0,991041566 | 0,999299353 | YES | 0 | FALSE |
| 248 | 22,75111542 | -0,05018256 | 2,908775844 | -0,017252123 | 0,98623548 | 0,999299353 | YES | 0 | FALSE |
| 249 | 9,990301438 | -4,27227283 | 2,91273093 | -1,466758493 | 0,142441765 | 0,999299353 | YES | 0 | FALSE |
| 250 | 78,55330611 | -0,896209195 | 0,562346065 | -1,593696928 | 0,111003933 | 0,999299353 | YES | 0 | FALSE |
| 251 | 7,24003739 | 0,051119337 | 2,909667984 | 0,017568787 | 0,985982857 | 0,999299353 | YES | 0 | FALSE |
| 252 | 15,37727118 | 0,168727834 | 2,907438332 | 0,05803316 | 0,953722215 | 0,999299353 | YES | 0 | FALSE |
| 253 | 12,68864099 | 0,305828542 | 1,625416396 | 0,188153966 | 0,850755957 | 0,999299353 | YES | 0 | FALSE |
| 254 | 5,638992417 | 0,582573756 | 2,665219477 | 0,218583783 | 0,826974287 | 0,999299353 | YES | 0 | FALSE |
| 255 | 5,804203834 | 0,331782974 | 2,906662756 | 0,114145672 | 0,909122318 | 0,999299353 | YES | 0 | FALSE |
| 256 | 10,03455377 | 0,208751324 | 2,906282798 | 0,071827602 | 0,942739106 | 0,999299353 | YES | 0 | FALSE |
| 257 | 40,25734808 | -0,435745969 | 2,908773379 | -0,149804028 | 0,880919231 | 0,999299353 | YES | 0 | FALSE |
| 258 | 11,04850758 | 0,312944348 | 1,539502151 | 0,20327633 | 0,838919055 | 0,999299353 | YES | 0 | FALSE |
| 259 | 9,382984409 | 0,209376467 | 2,909687885 | 0,071958394 | 0,942635019 | 0,999299353 | YES | 0 | FALSE |
| 260 | 8,923422661 | -1,149487022 | 2,909786637 | -0,395041687 | 0,692812123 | 0,999299353 | YES | 0 | FALSE |
| 261 | 13,76928275 | -0,083893743 | 2,909418167 | -0,02883523 | 0,976996003 | 0,999299353 | YES | 0 | FALSE |
| 262 | 18,63459123 | 0,091490429 | 2,90874857 | 0,031453536 | 0,974907846 | 0,999299353 | YES | 0 | FALSE |
| 263 | 4,677784239 | 0,01578562 | 2,910453542 | 0,005423766 | 0,995672482 | 0,999299353 | YES | 0 | FALSE |
| 264 | 121,089678 | -0,572760781 | 0,315452212 | -1,815681615 | 0,069419226 | 0,999299353 | YES | 57 | FALSE |
| 265 | 5,002586655 | -0,049589348 | 2,9102279 | -0,017039679 | 0,986404961 | 0,999299353 | YES | 0 | FALSE |
| 266 | 12,31047961 | -0,425390363 | 2,911589882 | -0,146102432 | 0,883840526 | 0,999299353 | YES | 101 | FALSE |

| Ticker | taxLevel | Phylum | Family | Genus | Species |
|---|---|---|---|---|---|
| 267 | ASV | Verrucomicrobiota | Victivallaceae | Victivallis | NA |
| 268 | ASV | Actinobacteriota | Actinomycetaceae | Actinomyces | odontolyticus |
| 269 | ASV | Firmicutes | Lachnospiraceae | NA | NA |
| 270 | ASV | Bacteroidota | Marinifilaceae | Butyricimonas | paravirosa |
| 271 | ASV | Bacteroidota | Bacteroidaceae | Bacteroides | NA |
| 272 | ASV | Firmicutes | NA | NA | NA |
| 273 | ASV | Bacteroidota | Rikenellaceae | Alistipes | NA |
| 274 | ASV | Firmicutes | Christensenellaceae | Christensenellaceae R-7 group | NA |
| 275 | ASV | Firmicutes | Oscillospiraceae | Intestinimonas | massiliensis |
| 276 | ASV | Bacteroidota | Bacteroidaceae | Bacteroides | caccae |
| 277 | ASV | Firmicutes | UCG-010 | NA | NA |
| 278 | ASV | Firmicutes | Lachnospiraceae | [Eubacterium] xylanophilum group | NA |
| 279 | ASV | Firmicutes | Lachnospiraceae | NA | NA |
| 280 | ASV | Firmicutes | Lachnospiraceae | Howardella | ureilytica |
| 281 | ASV | Bacteroidota | Bacteroidaceae | Bacteroides | NA |
| 282 | ASV | Firmicutes | Anaerovoracaceae | Family XIII AD3011 group | NA |
| 283 | ASV | Firmicutes | Lachnospiraceae | Eisenbergiella | tayi |
| 284 | ASV | Firmicutes | Lachnospiraceae | Blautia | NA |
| 285 | ASV | Bacteroidota | Prevotellaceae | Prevotella | NA |
| 286 | ASV | Firmicutes | Erysipelatoclostridiaceae | Erysipelatoclostridium | NA |
| 287 | ASV | Firmicutes | Christensenellaceae | Christensenellaceae R-7 group | NA |
| 288 | ASV | Bacteroidota | Bacteroidaceae | Bacteroides | nordii |
| 289 | ASV | Firmicutes | Oscillospiraceae | Colidextribacter | NA |
| 290 | ASV | Firmicutes | Ruminococcaceae | NA | NA |
| 291 | ASV | Firmicutes | Lachnospiraceae | Marvinbryantia | NA |
| 292 | ASV | Firmicutes | Oscillospiraceae | UCG-005 | NA |
| 293 | ASV | Proteobacteria | Oxalobacteraceae | Oxalobacter | formigenes |
| 294 | ASV | Firmicutes | Lachnospiraceae | Blautia | NA |
| 295 | ASV | Firmicutes | Lachnospiraceae | Coprococcus | eutactus |
| 296 | ASV | Firmicutes | UCG-010 | NA | NA |

| Ticker | DESeq2_baseMean | DESeq2_log2-FoldChange | DESeq2_lfcSE | DESeq2_stat | DESeq2_pvalue | DESeq2_padj | DESeq2 converged | ANCOM_W | ANCOM_detected_0.7 |
|---|---|---|---|---|---|---|---|---|---|
| 267 | 5,775539756 | -0,015409653 | 2,907666177 | -0,005299664 | 0,9957715 | 0,999299353 | YES | 0 | FALSE |
| 268 | 0,88183281 | 0,239113483 | 2,913083801 | 0,082082597 | 0,934581032 | 0,999299353 | YES | 0 | FALSE |
| 269 | 4,728764316 | -0,254053398 | 2,911644753 | -0,087254256 | 0,930469413 | 0,999299353 | YES | 0 | FALSE |
| 270 | 6,96757681 | 0,332410404 | 2,910519131 | 0,114210005 | 0,909071321 | 0,999299353 | YES | 0 | FALSE |
| 271 | 123,4362494 | 0,320576927 | 0,539026481 | 0,594733169 | 0,552021846 | 0,999299353 | YES | 0 | FALSE |
| 272 | 5,371350046 | -0,141588165 | 2,911142008 | -0,04863664 | 0,96120887 | 0,999299353 | YES | 0 | FALSE |
| 273 | 199,5822721 | -0,486205256 | 0,795768377 | -0,610988412 | 0,541207252 | 0,999299353 | YES | 0 | FALSE |
| 274 | 5,565362966 | 0,475567909 | 2,91130501 | 0,163352142 | 0,870241182 | 0,999299353 | YES | 1 | FALSE |
| 275 | 8,649431688 | 0,369884507 | 2,909084678 | 0,127148072 | 0,898823205 | 0,999299353 | YES | 0 | FALSE |
| 276 | 261,9482242 | 0,335208402 | 0,558967803 | 0,599691789 | 0,548711661 | 0,999299353 | YES | 0 | FALSE |
| 277 | 9,327913783 | 0,317278231 | 2,909692678 | 0,109041836 | 0,913169308 | 0,999299353 | YES | 0 | FALSE |
| 278 | 8,325782108 | 0,108819817 | 2,912234509 | 0,037366433 | 0,970192836 | 0,999299353 | YES | 0 | FALSE |
| 279 | 4,052568483 | -0,448702652 | 2,906974951 | -0,154353808 | 0,877330773 | 0,999299353 | YES | 0 | FALSE |
| 280 | 9,254855614 | -0,370992199 | 1,803633267 | -0,205691592 | 0,837031826 | 0,999299353 | YES | 6 | FALSE |
| 281 | 138,5637744 | -1,102279492 | 1,352832906 | -0,814793525 | 0,415190508 | 0,999299353 | YES | 0 | FALSE |
| 282 | 2,806398495 | 0,070239377 | 2,909308367 | 0,024142981 | 0,980738559 | 0,999299353 | YES | 0 | FALSE |
| 283 | 7,024069658 | -0,927120728 | 2,911750561 | -0,318406645 | 0,750176498 | 0,999299353 | YES | 0 | FALSE |
| 284 | 415,0190944 | 0,232887296 | 0,219439377 | 1,061283071 | 0,288561277 | 0,999299353 | YES | 0 | FALSE |
| 285 | 45,54055903 | -4,606448371 | 2,911725576 | -1,582033832 | 0,113641847 | 0,999299353 | YES | 0 | FALSE |
| 286 | 4,75739345 | 0,048838134 | 2,912306177 | 0,016769574 | 0,986620443 | 0,999299353 | YES | 0 | FALSE |
| 287 | 3,183859222 | -0,059164853 | 2,909749933 | -0,020333312 | 0,983777482 | 0,999299353 | YES | 0 | FALSE |
| 288 | 5,386847909 | 0,096427266 | 2,911662898 | 0,033117593 | 0,973580813 | 0,999299353 | YES | 0 | FALSE |
| 289 | 10,98952418 | 0,1491334 | 2,910398407 | 0,051241576 | 0,959133023 | 0,999299353 | YES | 0 | FALSE |
| 290 | 2,805795971 | -0,28499859 | 2,911928196 | -0,097872808 | 0,922033292 | 0,999299353 | YES | 0 | FALSE |
| 291 | 7,050817322 | 0,385971534 | 2,908913976 | 0,132685785 | 0,894441886 | 0,999299353 | YES | 0 | FALSE |
| 292 | 302,0586381 | -0,012560091 | 0,312774877 | -0,040156969 | 0,967967984 | 0,999299353 | YES | 0 | FALSE |
| 293 | 13,73103523 | -0,135032077 | 0,497891337 | -0,271207926 | 0,786231117 | 0,999299353 | YES | 0 | FALSE |
| 294 | 14,3654402 | 0,501430647 | 2,912304775 | 0,17217657 | 0,863298717 | 0,999299353 | YES | 11 | FALSE |
| 295 | 291,6553856 | 0,029055452 | 0,850228465 | 0,0341737 | 0,972738639 | 0,999299353 | YES | 0 | FALSE |
| 296 | 7,768918053 | 0,366173292 | 2,909036575 | 0,12587442 | 0,899831331 | 0,999299353 | YES | 0 | FALSE |

| Ticker | taxLevel | Phylum | Family | Genus | Species |
|---|---|---|---|---|---|
| 297 | ASV | Firmicutes | Lachnospiraceae | [Eubacterium] ventriosum group | NA |
| 298 | ASV | Firmicutes | UCG-010 | NA | NA |
| 299 | ASV | Firmicutes | Lachnospiraceae | NA | NA |
| 300 | ASV | Firmicutes | Lachnospiraceae | [Ruminococcus] torques group | NA |
| 301 | ASV | Firmicutes | Veillonellaceae | Allisonella | histaminiformans |
| 302 | ASV | Firmicutes | Ruminococcaceae | Negativibacillus | massiliensis |
| 303 | ASV | Firmicutes | Anaerovoracaceae | Family XIII UCG-001 | NA |
| 304 | ASV | Firmicutes | Erysipelatoclostridiaceae | Asteroleplasma | NA |
| 305 | ASV | Bacteroidota | Marinifilaceae | Butyricimonas | NA |
| 306 | ASV | Bacteroidota | Marinifilaceae | Butyricimonas | NA |
| 307 | ASV | Bacteroidota | Bacteroidaceae | Bacteroides | NA |
| 308 | ASV | Proteobacteria | Sutterellaceae | Sutterella | NA |
| 309 | ASV | Firmicutes | Lachnospiraceae | NA | NA |
| 310 | ASV | Firmicutes | Monoglobaceae | Monoglobus | NA |
| 311 | ASV | Firmicutes | Lachnospiraceae | NA | NA |
| 312 | ASV | Firmicutes | Oscillospiraceae | UCG-005 | NA |
| 313 | ASV | Bacteroidota | Bacteroidaceae | Bacteroides | massiliensis |
| 314 | ASV | Firmicutes | Lachnospiraceae | Roseburia | inulinivorans |
| 315 | ASV | Firmicutes | Erysipelatoclostridiaceae | Erysipelatoclostridium | NA |
| 316 | ASV | Bacteroidota | Bacteroidaceae | Bacteroides | NA |
| 317 | ASV | Bacteroidota | NA | NA | NA |
| 318 | ASV | Firmicutes | Anaerovoracaceae | Family XIII AD3011 group | NA |
| 319 | ASV | Firmicutes | [Eubacterium] coprostanoligenes group | NA | NA |
| 320 | ASV | Firmicutes | UCG-010 | NA | NA |
| 321 | ASV | Firmicutes | Lachnospiraceae | NA | NA |
| 322 | ASV | Firmicutes | Oscillospiraceae | Oscillibacter | NA |
| 323 | ASV | Firmicutes | Ruminococcaceae | Faecalibacterium | NA |
| 324 | ASV | Firmicutes | Ruminococcaceae | [Eubacterium] siraeum group | NA |
| 325 | ASV | Firmicutes | Lachnospiraceae | NA | NA |
| 326 | ASV | Firmicutes | Lachnospiraceae | NA | NA |

| Ticker | DESeq2_baseMean | DESeq2_log2-FoldChange | DESeq2_lfcSE | DESeq2_stat | DESeq2_pvalue | DESeq2_padj | DESeq2 converged | ANCOM_W | ANCOM_detected_0.7 |
|---|---|---|---|---|---|---|---|---|---|
| 297 | 2,86496118 | 0,088319391 | 2,911346768 | 0,030336266 | 0,975798874 | 0,999299353 | YES | 0 | FALSE |
| 298 | 6,437604221 | 0,458256292 | 2,904039934 | 0,157799584 | 0,874614727 | 0,999299353 | YES | 0 | FALSE |
| 299 | 5,034264887 | -0,122829393 | 2,910564766 | -0,042201223 | 0,966338288 | 0,999299353 | YES | 0 | FALSE |
| 300 | 249,2766116 | -0,624914757 | 0,44654718 | -1,399437249 | 0,161681904 | 0,999299353 | YES | 0 | FALSE |
| 301 | 8,770524285 | -0,086018639 | 2,9076228 | -0,029583837 | 0,976398956 | 0,999299353 | YES | 0 | FALSE |
| 302 | 9,056758508 | 0,473189353 | 2,910604834 | 0,162574235 | 0,870853675 | 0,999299353 | YES | 0 | FALSE |
| 303 | 7,625175334 | 0,111812904 | 2,910300961 | 0,038419705 | 0,96935305 | 0,999299353 | YES | 0 | FALSE |
| 304 | 3,166917521 | -0,164722933 | 2,910885557 | -0,056588598 | 0,954872917 | 0,999299353 | YES | 0 | FALSE |
| 305 | 12,31516585 | 0,072449891 | 2,907428452 | 0,02491889 | 0,98011966 | 0,999299353 | YES | 0 | FALSE |
| 306 | 6,374459177 | 0,263281971 | 2,908420649 | 0,090524034 | 0,927870796 | 0,999299353 | YES | 0 | FALSE |
| 307 | 144,2814565 | 0,02667813 | 0,853503393 | 0,031257205 | 0,975064419 | 0,999299353 | YES | 0 | FALSE |
| 308 | 2,523890074 | 0,120821962 | 2,913822018 | 0,041465114 | 0,966925104 | 0,999299353 | YES | 0 | FALSE |
| 309 | 11,48671386 | 0,130087559 | 2,906893676 | 0,044751399 | 0,964305464 | 0,999299353 | YES | 0 | FALSE |
| 310 | 78,31427244 | 0,554767254 | 0,533476839 | 1,039908789 | 0,298382279 | 0,999299353 | YES | 0 | FALSE |
| 311 | 11,45333846 | 0,284081256 | 2,909802673 | 0,097629045 | 0,92222686 | 0,999299353 | YES | 0 | FALSE |
| 312 | 30,91172048 | 1,605568818 | 2,912388262 | 0,551289414 | 0,581435294 | 0,999299353 | YES | 0 | FALSE |
| 313 | 164,1406568 | -0,326266792 | 0,559514453 | -0,583124869 | 0,559809243 | 0,999299353 | YES | 0 | FALSE |
| 314 | 153,9674257 | -1,111816058 | 0,709794642 | -1,566391167 | 0,11725707 | 0,999299353 | YES | 0 | FALSE |
| 315 | 0,388289001 | 0,16497248 | 2,914850406 | 0,056597237 | 0,954866035 | 0,999299353 | YES | 0 | FALSE |
| 316 | 151,8729563 | 0,115527662 | 0,647576373 | 0,178400057 | 0,858408804 | 0,999299353 | YES | 0 | FALSE |
| 317 | 3,547781421 | 0,246047637 | 2,911515626 | 0,084508438 | 0,932652195 | 0,999299353 | YES | 0 | FALSE |
| 318 | 7,920860219 | 0,643296327 | 2,90911845 | 0,221131019 | 0,824990418 | 0,999299353 | YES | 0 | FALSE |
| 319 | 164,9695729 | -0,925754272 | 0,934454455 | -0,990689559 | 0,321837191 | 0,999299353 | YES | 0 | FALSE |
| 320 | 7,495680267 | 0,33359262 | 2,908437658 | 0,114698219 | 0,908684325 | 0,999299353 | YES | 0 | FALSE |
| 321 | 5,872426435 | 0,477285617 | 2,910534599 | 0,16398555 | 0,869742519 | 0,999299353 | YES | 0 | FALSE |
| 322 | 10,26691315 | -0,16236776 | 2,490594139 | -0,06519238 | 0,948020828 | 0,999299353 | YES | 0 | FALSE |
| 323 | 779,6942869 | -0,587498783 | 0,306417827 | -1,917312675 | 0,055198221 | 0,999299353 | YES | 0 | FALSE |
| 324 | 115,647046 | 0,54036327 | 0,709131835 | 0,762006785 | 0,446055954 | 0,999299353 | YES | 0 | FALSE |
| 325 | 17,93930574 | 0,436889416 | 2,911668188 | 0,150047803 | 0,880726901 | 0,999299353 | YES | 0 | FALSE |
| 326 | 4,914105369 | 0,10079652 | 2,913011115 | 0,034602175 | 0,972396967 | 0,999299353 | YES | 0 | FALSE |

| Ticker | taxLevel | Phylum | Family | Genus | Species |
|---|---|---|---|---|---|
| 327 | ASV | Firmicutes | Erysipelatoclostridiaceae | Catenibacterium | mitsuokai |
| 328 | ASV | Firmicutes | Lachnospiraceae | Lachnospiraceae UCG-010 | NA |
| 329 | ASV | Firmicutes | Oscillospiraceae | Oscillibacter | NA |
| 330 | ASV | Actinobacteriota | Atopobiaceae | NA | NA |
| 331 | ASV | Firmicutes | NA | NA | NA |
| 332 | ASV | Firmicutes | UCG-010 | NA | NA |
| 333 | ASV | Firmicutes | UCG-010 | NA | NA |
| 334 | ASV | Firmicutes | NA | NA | NA |
| 335 | ASV | Actinobacteriota | Eggerthellaceae | NA | NA |
| 336 | ASV | Firmicutes | Ruminococcaceae | Angelakisella | NA |
| 337 | ASV | Firmicutes | Oscillospiraceae | UCG-002 | NA |
| 338 | ASV | Firmicutes | Ruminococcaceae | Incertae Sedis | NA |
| 339 | ASV | Firmicutes | Lachnospiraceae | NA | NA |
| 340 | ASV | Firmicutes | Oscillospiraceae | Flavonifractor | NA |
| 341 | ASV | Firmicutes | Butyricicoccaceae | Butyricicoccus | NA |
| 342 | ASV | Firmicutes | Lachnospiraceae | NA | NA |
| 343 | ASV | Firmicutes | Erysipelotrichaceae | Holdemania | filiformis |
| 344 | ASV | Firmicutes | Erysipelotrichaceae | Holdemanella | biformis |
| 345 | ASV | Firmicutes | Christensenellaceae | Christensenellaceae R-7 group | NA |
| 346 | ASV | Bacteroidota | Prevotellaceae | NA | NA |
| 347 | ASV | Firmicutes | Ruminococcaceae | Faecalibacterium | NA |
| 348 | ASV | Firmicutes | Oscillospiraceae | NA | NA |
| 349 | ASV | Firmicutes | Ruminococcaceae | DTU089 | NA |
| 350 | ASV | Firmicutes | Christensenellaceae | NA | NA |
| 351 | ASV | Firmicutes | Ruminococcaceae | Fournierella | NA |
| 352 | ASV | Firmicutes | UCG-010 | NA | NA |
| 353 | ASV | Firmicutes | Lachnospiraceae | Agathobacter | NA |
| 354 | ASV | Firmicutes | Lachnospiraceae | Roseburia | intestinalis |
| 355 | ASV | Firmicutes | Lachnospiraceae | Lachnoclostridium | NA |
| 356 | ASV | Firmicutes | Hydrogenoanaerobacterium | NA | NA |

| Ticker | DESeq2_baseMean | DESeq2_log2-FoldChange | DESeq2_lfcSE | DESeq2_stat | DESeq2_pvalue | DESeq2_padj | DESeq2 converged | ANCOM_W | ANCOM_detected_0.7 |
|---|---|---|---|---|---|---|---|---|---|
| 327 | 67,01747497 | -1,077925318 | 2,912270446 | -0,370132286 | 0,711283927 | 0,999299353 | YES | 0 | FALSE |
| 328 | 3,055112192 | 0,153194079 | 2,91374665 | 0,052576321 | 0,958069484 | 0,999299353 | YES | 0 | FALSE |
| 329 | 0,505963203 | -0,146729132 | 2,913482167 | -0,050362118 | 0,959833824 | 0,999299353 | YES | 0 | FALSE |
| 330 | 1,81793053 | 0,294501734 | 2,912253003 | 0,101125051 | 0,919451192 | 0,999299353 | YES | 0 | FALSE |
| 331 | 184,8812144 | 0,717309943 | 1,13321501 | 0,632986624 | 0,526742378 | 0,999299353 | YES | 0 | FALSE |
| 332 | 19,61685753 | 0,260489549 | 2,908770649 | 0,089553141 | 0,928642322 | 0,999299353 | YES | 0 | FALSE |
| 333 | 3,211174268 | 0,337061533 | 2,909463607 | 0,11585006 | 0,907771375 | 0,999299353 | YES | 0 | FALSE |
| 334 | 15,32174136 | 0,876829244 | 0,933125026 | 0,939669625 | 0,34738705 | 0,999299353 | YES | 23 | FALSE |
| 335 | 83,76738392 | 0,021165718 | 0,304886814 | 0,069421558 | 0,944654069 | 0,999299353 | YES | 0 | FALSE |
| 336 | 1,756793572 | 0,170990005 | 2,912532964 | 0,05870835 | 0,953184408 | 0,999299353 | YES | 0 | FALSE |
| 337 | 297,2102856 | -0,102120175 | 0,278071128 | -0,367244798 | 0,713436426 | 0,999299353 | YES | 0 | FALSE |
| 338 | 74,33547493 | 0,96513267 | 0,378720446 | 2,548403923 | 0,010821709 | 0,600681553 | YES | 0 | FALSE |
| 339 | 7,072394613 | -0,17277927 | 2,911936899 | -0,059334826 | 0,952685423 | 0,999299353 | YES | 0 | FALSE |
| 340 | 0,930872367 | 0,029193684 | 2,91344172 | 0,010020343 | 0,992005057 | 0,999299353 | YES | 0 | FALSE |
| 341 | 72,12331995 | 0,076015014 | 0,389218187 | 0,195301804 | 0,845156685 | 0,999299353 | YES | 0 | FALSE |
| 342 | 9,362214561 | 0,314738681 | 2,906974736 | 0,108270181 | 0,913781376 | 0,999299353 | YES | 0 | FALSE |
| 343 | 1,219516932 | -0,487049956 | 2,909755126 | -0,167385204 | 0,867066966 | 0,999299353 | YES | 0 | FALSE |
| 344 | 49,83996399 | -1,758153569 | 2,911541464 | -0,603856614 | 0,545938976 | 0,999299353 | YES | 0 | FALSE |
| 345 | 7,198649845 | 0,74348241 | 2,906556493 | 0,255794928 | 0,79810919 | 0,999299353 | YES | 51 | FALSE |
| 346 | 5,961837949 | 0,151126416 | 2,912379518 | 0,051891045 | 0,95861551 | 0,999299353 | YES | 0 | FALSE |
| 347 | 96,46549968 | -0,485893677 | 0,592983658 | -0,819404835 | 0,412555477 | 0,999299353 | YES | 0 | FALSE |
| 348 | 8,130540982 | -0,728913878 | 2,907993094 | -0,250658738 | 0,802077965 | 0,999299353 | YES | 83 | FALSE |
| 349 | 3,018838706 | 0,189501475 | 2,913698306 | 0,065038125 | 0,948143645 | 0,999299353 | YES | 0 | FALSE |
| 350 | 88,23778836 | 0,185172376 | 0,679963864 | 0,27232679 | 0,785370759 | 0,999299353 | YES | 0 | FALSE |
| 351 | 8,946274281 | 0,069619873 | 2,913000167 | 0,023899715 | 0,980932602 | 0,999299353 | YES | 0 | FALSE |
| 352 | 4,254865218 | -0,088593047 | 2,908812967 | -0,03045677 | 0,97570277 | 0,999299353 | YES | 0 | FALSE |
| 353 | 1602,321472 | -1,064418515 | 0,391695392 | -2,717464991 | 0,006578411 | 0,540745421 | YES | 0 | FALSE |
| 354 | 135,8232871 | -0,587926608 | 1,31698031 | -0,446420196 | 0,655293747 | 0,999299353 | YES | 0 | FALSE |
| 355 | 2,559200671 | 0,472782217 | 2,913784161 | 0,162257117 | 0,871103383 | 0,999299353 | YES | 0 | FALSE |
| 356 | 2,721956617 | -0,181501152 | 2,911109915 | -0,06234775 | 0,950285904 | 0,999299353 | YES | 0 | FALSE |

| Ticker | taxLevel | Phylum | Family | Genus | Species |
|---|---|---|---|---|---|
| 357 | ASV | Firmicutes | Lachnospiraceae | NA | NA |
| 358 | ASV | Firmicutes | [Eubacterium] coprostanoligenes group | NA | NA |
| 359 | ASV | Firmicutes | Oscillospiraceae | Oscillibacter | NA |
| 360 | ASV | Proteobacteria | Sutterellaceae | Parasutterella | excrementihominis |
| 361 | ASV | Verrucomicrobiota | vadinBE97 | NA | NA |
| 362 | ASV | Firmicutes | Ruminococcaceae | Angelakisella | NA |
| 363 | ASV | Firmicutes | Oscillospiraceae | UCG-005 | NA |
| 364 | ASV | Firmicutes | Ruminococcaceae | NA | NA |
| 365 | ASV | Firmicutes | Lachnospiraceae | Coprococcus | catus |
| 366 | ASV | Firmicutes | Lachnospiraceae | [Ruminococcus] gauvreauii group | NA |
| 367 | ASV | Verrucomicrobiota | Puniceicoccaceae | NA | NA |
| 368 | ASV | Firmicutes | Ruminococcaceae | NA | NA |
| 369 | ASV | Firmicutes | NA | NA | NA |
| 370 | ASV | Firmicutes | Lachnospiraceae | Lachnospiraceae ND3007 group | NA |
| 371 | ASV | Firmicutes | UCG-010 | NA | NA |
| 372 | ASV | Firmicutes | Lachnospiraceae | [Ruminococcus] torques group | NA |
| 373 | ASV | Firmicutes | Ruminococcaceae | Fournierella | NA |
| 374 | ASV | Firmicutes | Lachnospiraceae | [Eubacterium] eligens group | NA |
| 375 | ASV | Firmicutes | Lachnospiraceae | [Eubacterium] ruminantium group | NA |
| 376 | ASV | Firmicutes | Peptococcaceae | Peptococcus | NA |
| 377 | ASV | Firmicutes | Ruminococcaceae | Faecalibacterium | prausnitzii |
| 378 | ASV | Firmicutes | Acidaminococcaceae | Phascolarctobacterium | faecium |
| 379 | ASV | Firmicutes | Christensenellaceae | Christensenellaceae R-7 group | NA |
| 380 | ASV | Firmicutes | Peptococcaceae | NA | NA |
| 381 | ASV | Desulfobacterota | Desulfovibrionaceae | NA | NA |
| 382 | ASV | Firmicutes | Lachnospiraceae | Lachnospiraceae NK4A136 group | NA |
| 383 | ASV | Firmicutes | Ruminococcaceae | NA | NA |
| 384 | ASV | Firmicutes | Oscillospiraceae | Colidextribacter | NA |
| 385 | ASV | Firmicutes | Lachnospiraceae | Coprococcus | eutactus |
| 386 | ASV | Firmicutes | Lachnospiraceae | GCA-900066575 | NA |

| Ticker | DESeq2_baseMean | DESeq2_log2-FoldChange | DESeq2_lfcSE | DESeq2_stat | DESeq2_pvalue | DESeq2_padj | DESeq2 converged | ANCOM_W | ANCOM_detected_0.7 |
|---|---|---|---|---|---|---|---|---|---|
| 357 | 3,696366896 | 0,225106169 | 2,908330992 | 0,077400464 | 0,938304971 | 0,999299353 | YES | 0 | FALSE |
| 358 | 45,26333147 | 0,859607749 | 1,14109459 | 0,753318574 | 0,4512585 | 0,999299353 | YES | 0 | FALSE |
| 359 | 5,558682084 | 0,300100546 | 2,913101705 | 0,103017531 | 0,917949057 | 0,999299353 | YES | 0 | FALSE |
| 360 | 52,42523433 | 0,11898548 | 0,715175983 | 0,16637231 | 0,867863964 | 0,999299353 | YES | 0 | FALSE |
| 361 | 1,801031007 | 0,136310344 | 2,910008376 | 0,046841908 | 0,962639228 | 0,999299353 | YES | 0 | FALSE |
| 362 | 2,641995519 | 0,225022281 | 2,910133142 | 0,077323707 | 0,938366032 | 0,999299353 | YES | 0 | FALSE |
| 363 | 225,4569035 | 0,142527139 | 0,4460371 | 0,319540997 | 0,749316309 | 0,999299353 | YES | 0 | FALSE |
| 364 | 2,152719414 | 0,359118574 | 2,913246877 | 0,123270903 | 0,901892581 | 0,999299353 | YES | 0 | FALSE |
| 365 | 87,92888506 | -0,127295354 | 0,232923456 | -0,54651153 | 0,584714364 | 0,999299353 | YES | 0 | FALSE |
| 366 | 34,87997267 | 0,034614094 | 1,747114786 | 0,019812147 | 0,984193228 | 0,999299353 | YES | 0 | FALSE |
| 367 | 4,171374726 | 0,184318992 | 2,912044988 | 0,063295379 | 0,949531295 | 0,999299353 | YES | 0 | FALSE |
| 368 | 5,988697938 | 0,109814069 | 2,908994488 | 0,037749837 | 0,96988714 | 0,999299353 | YES | 0 | FALSE |
| 369 | 8,554994248 | 0,01137668 | 2,912412157 | 0,003906274 | 0,996883252 | 0,999299353 | YES | 0 | FALSE |
| 370 | 72,59997327 | -0,964252483 | 0,413896902 | -2,329692437 | 0,019822413 | 0,905223508 | YES | 0 | FALSE |
| 371 | 6,197101774 | 0,250762265 | 2,907951168 | 0,086233313 | 0,931280949 | 0,999299353 | YES | 0 | FALSE |
| 372 | 125,0317041 | 0,311593006 | 0,84433738 | 0,369038506 | 0,712099023 | 0,999299353 | YES | 0 | FALSE |
| 373 | 7,879044875 | 0,286481359 | 2,910944973 | 0,098415244 | 0,921602571 | 0,999299353 | YES | 0 | FALSE |
| 374 | 101,7224426 | -0,087805983 | 0,685123422 | -0,128160824 | 0,898021702 | 0,999299353 | YES | 0 | FALSE |
| 375 | 181,6773233 | -0,50209039 | 0,99812917 | -0,503031476 | 0,614942143 | 0,999299353 | YES | 0 | FALSE |
| 376 | 4,546450581 | -0,056020016 | 2,909478739 | -0,019254313 | 0,98463823 | 0,999299353 | YES | 0 | FALSE |
| 377 | 1045,211502 | -0,202775072 | 0,371722045 | -0,545501874 | 0,585408392 | 0,999299353 | YES | 0 | FALSE |
| 378 | 91,33627835 | 0,032371048 | 2,910781678 | 0,011121084 | 0,991126842 | 0,999299353 | YES | 0 | FALSE |
| 379 | 2,293529572 | 0,299775901 | 2,906673497 | 0,103133669 | 0,917856884 | 0,999299353 | YES | 0 | FALSE |
| 380 | 1,253800201 | 0,238691334 | 2,914451193 | 0,081899239 | 0,93472684 | 0,999299353 | YES | 0 | FALSE |
| 381 | 1,725099267 | 0,355842761 | 2,911205019 | 0,12223212 | 0,902715189 | 0,999299353 | YES | 0 | FALSE |
| 382 | 100,0852058 | -1,107313347 | 0,879089376 | -1,259614069 | 0,207808618 | 0,999299353 | YES | 0 | FALSE |
| 383 | 1,295754344 | 0,006690863 | 2,912018025 | 0,002297672 | 0,998166724 | 0,999299353 | YES | 0 | FALSE |
| 384 | 1,608697539 | 0,067324028 | 2,913099343 | 0,02311079 | 0,981561898 | 0,999299353 | YES | 0 | FALSE |
| 385 | 119,8655212 | 0,642053928 | 2,175091047 | 0,295184852 | 0,767852673 | 0,999299353 | YES | 0 | FALSE |
| 386 | 3,236851239 | 0,431248779 | 2,911883063 | 0,148099621 | 0,882264148 | 0,999299353 | YES | 0 | FALSE |

| Ticker | taxLevel | Phylum | Family | Genus | Species |
|---|---|---|---|---|---|
| 387 | ASV | Firmicutes | Christensenellaceae | Christensenellaceae R-7 group | NA |
| 388 | ASV | Firmicutes | Lachnospiraceae | Lachnospiraceae NK4A136 group | NA |
| 389 | ASV | Verrucomicrobiota | Victivallaceae | Victivallis | vadensis |
| 390 | ASV | Bacteroidota | Bacteroidaceae | Bacteroides | NA |
| 391 | ASV | Firmicutes | Lachnospiraceae | Coprococcus | NA |
| 392 | ASV | Firmicutes | Lachnospiraceae | [Eubacterium] hallii group | NA |
| 393 | ASV | Firmicutes | NA | NA | NA |
| 394 | ASV | Proteobacteria | Sutterellaceae | Sutterella | wadsworthensis |
| 395 | ASV | Firmicutes | NA | NA | NA |
| 396 | ASV | Firmicutes | Lachnospiraceae | Dorea | formicigenerans |
| 397 | ASV | Firmicutes | Lachnospiraceae | Shuttleworthia | NA |
| 398 | ASV | Firmicutes | Lachnospiraceae | Blautia | NA |
| 399 | ASV | Desulfobacterota | Desulfovibrionaceae | NA | NA |
| 400 | ASV | Bacteroidota | Barnesiellaceae | Coprobacter | secundus |
| 401 | ASV | Bacteroidota | Bacteroidaceae | Bacteroides | NA |
| 402 | ASV | Firmicutes | Lachnospiraceae | [Eubacterium] eligens group | NA |
| 403 | ASV | Firmicutes | Erysipelotrichaceae | Turicibacter | sanguinis |
| 404 | ASV | Firmicutes | Oscillospiraceae | NK4A214 group | NA |
| 405 | ASV | Firmicutes | UCG-010 | NA | NA |
| 406 | ASV | Bacteroidota | Rikenellaceae | Alistipes | shahii |
| 407 | ASV | Bacteroidota | Prevotellaceae | Prevotella | NA |
| 408 | ASV | Firmicutes | Lachnospiraceae | CAG-56 | NA |
| 409 | ASV | Firmicutes | Lachnospiraceae | [Ruminococcus] gnavus group | NA |
| 410 | ASV | Firmicutes | Acidaminococcaceae | Phascolarctobacterium | succinatutens |
| 411 | ASV | Bacteroidota | Bacteroidaceae | Bacteroides | plebeius |
| 412 | Genus | Actinobacteriota | Bifidobacteriaceae | Bifidobacterium | NA |
| 413 | Genus | Verrucomicrobiota | Akkermansiaceae | Akkermansia | NA |
| 414 | Genus | Firmicutes | Lachnospiraceae | Lachnospira | NA |
| 415 | Genus | Firmicutes | Oscillospiraceae | UCG-003 | NA |
| 416 | Genus | Firmicutes | Veillonellaceae | Dialister | NA |

| Ticker | DESeq2_baseMean | DESeq2_log2-FoldChange | DESeq2_lfcSE | DESeq2_stat | DESeq2_pvalue | DESeq2_padj | DESeq2 converged | ANCOM_W | ANCOM_detected_0.7 |
|---|---|---|---|---|---|---|---|---|---|
| 387 | 1,033120643 | 0,189543808 | 2,912704565 | 0,065074849 | 0,948114406 | 0,999299353 | YES | 0 | FALSE |
| 388 | 52,16535846 | 0,273974106 | 1,835082416 | 0,149297985 | 0,881318506 | 0,999299353 | YES | 0 | FALSE |
| 389 | 1,493976754 | 0,474587872 | 2,910752425 | 0,163046458 | 0,870481856 | 0,999299353 | YES | 0 | FALSE |
| 390 | 38,42992757 | 0,600600717 | 2,904624298 | 0,206773977 | 0,836186381 | 0,999299353 | YES | 0 | FALSE |
| 391 | 235,0164518 | -2,07064404 | 1,39099958 | -1,488601485 | 0,13659234 | 0,999299353 | YES | 0 | FALSE |
| 392 | 56,64768015 | -0,217000095 | 0,580216238 | -0,373998659 | 0,708405314 | 0,999299353 | YES | 0 | FALSE |
| 393 | 4,357255709 | 0,429226154 | 1,729477495 | 0,248182561 | 0,803993159 | 0,999299353 | YES | 0 | FALSE |
| 394 | 41,68723249 | 0,034936504 | 2,012245912 | 0,017361946 | 0,986147868 | 0,999299353 | YES | 0 | FALSE |
| 395 | 1,922600258 | 0,299799684 | 2,911111014 | 0,102984628 | 0,917975171 | 0,999299353 | YES | 0 | FALSE |
| 396 | 42,28933085 | 0,913106737 | 0,731003588 | 1,249113892 | 0,21162342 | 0,999299353 | YES | 0 | FALSE |
| 397 | 2,894780216 | 0,070591764 | 2,912479715 | 0,024237684 | 0,980663019 | 0,999299353 | YES | 0 | FALSE |
| 398 | 71,7924502 | 1,425563359 | 0,701090683 | 2,033350884 | 0,042017086 | 0,999299353 | YES | 0 | FALSE |
| 399 | 1,374876307 | 0,183411463 | 2,91347344 | 0,062952852 | 0,949804048 | 0,999299353 | YES | 0 | FALSE |
| 400 | 3,875087249 | -0,071307515 | 2,908774885 | -0,024514621 | 0,980442121 | 0,999299353 | YES | 0 | FALSE |
| 401 | 661,8643954 | -0,045558802 | 0,411814215 | -0,110629504 | 0,91191015 | 0,999299353 | YES | 0 | FALSE |
| 402 | 79,09621748 | 1,028563287 | 0,910089594 | 1,130178055 | 0,258401205 | 0,999299353 | YES | 0 | FALSE |
| 403 | 30,82796885 | 0,85224907 | 2,905999682 | 0,293272252 | 0,769314061 | 0,999299353 | YES | 0 | FALSE |
| 404 | 104,516405 | -0,039997272 | 0,5801044 | -0,068948402 | 0,945030691 | 0,999299353 | YES | 0 | FALSE |
| 405 | 4,561937719 | -0,148177476 | 2,909611521 | -0,050926894 | 0,959383775 | 0,999299353 | YES | 0 | FALSE |
| 406 | 92,30897336 | 1,546384264 | 0,78067354 | 1,980833453 | 0,047609954 | 0,999299353 | YES | 0 | FALSE |
| 407 | 18,93040995 | -0,132668988 | 2,912325338 | -0,045554316 | 0,963665482 | 0,999299353 | YES | 0 | FALSE |
| 408 | 117,0625667 | -0,576044495 | 0,62005055 | -0,929028277 | 0,352874432 | 0,999299353 | YES | 0 | FALSE |
| 409 | 55,700982 | 0,034699345 | 2,910239556 | 0,011923192 | 0,990486895 | 0,999299353 | YES | 0 | FALSE |
| 410 | 185,5716817 | -0,416535146 | 0,656917269 | -0,6340755 | 0,526031551 | 0,999299353 | YES | 0 | FALSE |
| 411 | 40,62062215 | 0,077014468 | 2,912202688 | 0,026445435 | 0,978902055 | 0,999299353 | YES | 0 | FALSE |
| 412 | 716,9067968 | 0,18355832 | 0,241890852 | 0,758847713 | 0,44794366 | 0,999339594 | YES | 0 | FALSE |
| 413 | 1059,636449 | -0,00943369 | 0,452913155 | -0,020828917 | 0,98338213 | 0,999339594 | YES | 0 | FALSE |
| 414 | 130,5204589 | -0,946668912 | 0,469230108 | -2,017493965 | 0,04364399 | 0,698303843 | YES | 0 | FALSE |
| 415 | 141,0241001 | -0,626869316 | 0,401626459 | -1,560826738 | 0,118564637 | 0,872829116 | YES | 0 | FALSE |
| 416 | 486,5707601 | -0,073954556 | 0,44471186 | -0,166297692 | 0,867922682 | 0,999339594 | YES | 0 | FALSE |

| Ticker | taxLevel | Phylum | Family | Genus | Species |
|---|---|---|---|---|---|
| 417 | Genus | Desulfobacterota | Desulfovibrionaceae | Mailhella | NA |
| 418 | Genus | Firmicutes | Oscillospiraceae | NK4A214 group | NA |
| 419 | Genus | Firmicutes | Lachnospiraceae | Fusicatenibacter | NA |
| 420 | Genus | Proteobacteria | Enterobacteriaceae | Klebsiella | NA |
| 421 | Genus | Firmicutes | Ruminococcaceae | Candidatus Soleaferrea | NA |
| 422 | Genus | Actinobacteriota | Eggerthellaceae | Senegalimassilia | NA |
| 423 | Genus | Actinobacteriota | Eggerthellaceae | Enterorhabdus | NA |
| 424 | Genus | Bacteroidota | Marinifilaceae | Sanguibacteroides | NA |
| 425 | Genus | Firmicutes | Lachnospiraceae | Lachnoclostridium | NA |
| 426 | Genus | Firmicutes | Peptostreptococcaceae | Terrisporobacter | NA |
| 427 | Genus | Desulfobacterota | Desulfovibrionaceae | Bilophila | NA |
| 428 | Genus | Firmicutes | Oscillospiraceae | Oscillibacter | NA |
| 429 | Genus | Firmicutes | Christensenellaceae | Christensenellaceae R-7 group | NA |
| 430 | Genus | Firmicutes | Erysipelatoclostridiaceae | UCG-004 | NA |
| 431 | Genus | Firmicutes | Lachnospiraceae | [Eubacterium] ventriosum group | NA |
| 432 | Genus | Firmicutes | Peptostreptococcaceae | Romboutsia | NA |
| 433 | Genus | Desulfobacterota | Desulfovibrionaceae | Desulfovibrio | NA |
| 434 | Genus | Firmicutes | Ruminococcaceae | Paludicola | NA |
| 435 | Genus | Firmicutes | Oscillospiraceae | Intestinimonas | NA |
| 436 | Genus | Actinobacteriota | Coriobacteriaceae | Collinsella | NA |
| 437 | Genus | Proteobacteria | Pasteurellaceae | Haemophilus | NA |
| 438 | Genus | Firmicutes | Erysipelatoclostridiaceae | Asteroleplasma | NA |
| 439 | Genus | Firmicutes | Lachnospiraceae | Dorea | NA |
| 440 | Genus | Firmicutes | Erysipelatoclostridiaceae | Erysipelotrichaceae UCG-003 | NA |
| 441 | Genus | Firmicutes | Ruminococcaceae | UBA1819 | NA |
| 442 | Genus | Bacteroidota | Marinifilaceae | Odoribacter | NA |
| 443 | Genus | Actinobacteriota | Eggerthellaceae | Adlercreutzia | NA |
| 444 | Genus | Bacteroidota | Barnesiellaceae | Barnesiella | NA |
| 445 | Genus | Bacteroidota | Prevotellaceae | Prevotellaceae NK3B31 group | NA |
| 446 | Genus | Bacteroidota | Bacteroidaceae | Bacteroides | NA |

| Ticker | DESeq2_baseMean | DESeq2_log2-FoldChange | DESeq2_lfcSE | DESeq2_stat | DESeq2_pvalue | DESeq2_padj | DESeq2 converged | ANCOM_W | ANCOM_detected_0.7 |
|---|---|---|---|---|---|---|---|---|---|
| 417 | 5,408764909 | 0,105139874 | 2,912001987 | 0,036105701 | 0,971198076 | 0,999339594 | YES | 0 | FALSE |
| 418 | 444,1717823 | 0,453058263 | 0,21079271 | 2,149307069 | 0,031610066 | 0,578012644 | YES | 0 | FALSE |
| 419 | 735,5686131 | -0,256802368 | 0,274494315 | -0,935547128 | 0,349506408 | 0,999339594 | YES | 0 | FALSE |
| 420 | 72,08342991 | 2,173142375 | 2,911213196 | 0,746473112 | 0,455381668 | 0,999339594 | YES | 0 | FALSE |
| 421 | 6,99789109 | 0,017845729 | 2,911176785 | 0,006130074 | 0,995108939 | 0,999339594 | YES | 0 | FALSE |
| 422 | 22,02411631 | 0,438265878 | 0,558943596 | 0,784096787 | 0,432983321 | 0,999339594 | YES | 0 | FALSE |
| 423 | 8,411446589 | -0,054142993 | 0,88245561 | -0,06135492 | 0,951076553 | 0,999339594 | YES | 0 | FALSE |
| 424 | 2,994673597 | 0,009267151 | 2,91164185 | 0,003182792 | 0,997460503 | 0,999339594 | YES | 0 | FALSE |
| 425 | 168,1303521 | -0,257367701 | 0,370212646 | -0,695188842 | 0,486936964 | 0,999339594 | YES | 0 | FALSE |
| 426 | 6,236371299 | 0,371721391 | 2,908899072 | 0,127787655 | 0,89831702 | 0,999339594 | YES | 0 | FALSE |
| 427 | 59,72897528 | 0,476215837 | 0,246284269 | 1,93360233 | 0,053162041 | 0,702814088 | YES | 0 | FALSE |
| 428 | 59,15597452 | -0,116608447 | 0,33416369 | -0,348956068 | 0,727122292 | 0,999339594 | YES | 0 | FALSE |
| 429 | 1241,920264 | 0,405274964 | 0,24822709 | 1,63267822 | 0,102536691 | 0,872829116 | YES | 0 | FALSE |
| 430 | 11,24689325 | 0,169067616 | 2,909970387 | 0,058099428 | 0,95366943 | 0,999339594 | YES | 0 | FALSE |
| 431 | 78,28506593 | 0,088415711 | 0,412264131 | 0,214463748 | 0,830185434 | 0,999339594 | YES | 0 | FALSE |
| 432 | 39,21052986 | 0,33355643 | 0,78119886 | 0,42698018 | 0,669393764 | 0,999339594 | YES | 0 | FALSE |
| 433 | 267,8382523 | 0,020560328 | 0,187232769 | 0,10981159 | 0,912558799 | 0,999339594 | YES | 0 | FALSE |
| 434 | 6,081551098 | 0,032506632 | 2,90733428 | 0,011180906 | 0,991079113 | 0,999339594 | YES | 0 | FALSE |
| 435 | 126,1457446 | 0,001544473 | 0,25973599 | 0,005946319 | 0,995255552 | 0,999339594 | YES | 0 | FALSE |
| 436 | 205,2048867 | -0,222764096 | 0,303434569 | -0,734142115 | 0,462862128 | 0,999339594 | YES | 0 | FALSE |
| 437 | 7,985578301 | -0,01394645 | 2,388691104 | -0,005838532 | 0,995341552 | 0,999339594 | YES | 0 | FALSE |
| 438 | 4,33460539 | -0,101585305 | 2,909959966 | -0,03490952 | 0,97215189 | 0,999339594 | YES | 0 | FALSE |
| 439 | 563,3530868 | -0,086546043 | 0,231423065 | -0,373973284 | 0,708424193 | 0,999339594 | YES | 0 | FALSE |
| 440 | 142,8586301 | -0,283878122 | 0,256751451 | -1,105653426 | 0,268876556 | 0,999339594 | YES | 0 | FALSE |
| 441 | 25,33044464 | -0,249940065 | 1,092994629 | -0,228674559 | 0,81912187 | 0,999339594 | YES | 0 | FALSE |
| 442 | 44,49373392 | 0,243296961 | 0,299349976 | 0,812750897 | 0,416360893 | 0,999339594 | YES | 0 | FALSE |
| 443 | 9,224301864 | -0,099331792 | 1,97958935 | -0,050177979 | 0,95998056 | 0,999339594 | YES | 0 | FALSE |
| 444 | 256,6041676 | -0,396184323 | 0,342936778 | -1,155269275 | 0,247980174 | 0,999339594 | NO | 0 | FALSE |
| 445 | 40,16815987 | -0,278884624 | 2,911281261 | -0,095794463 | 0,923683815 | 0,999339594 | YES | 0 | FALSE |
| 446 | 4450,191083 | -0,075138246 | 0,138118426 | -0,544013193 | 0,586432394 | 0,999339594 | YES | 0 | FALSE |

| Ticker | taxLevel | Phylum | Family | Genus | Species |
|---|---|---|---|---|---|
| 447 | Genus | Bacteroidota | Prevotellaceae | Prevotella | NA |
| 448 | Genus | Firmicutes | Lachnospiraceae | [Eubacterium] xylanophilum group | NA |
| 449 | Genus | Firmicutes | Lachnospiraceae | Lachnospiraceae FCS020 group | NA |
| 450 | Genus | Firmicutes | Lachnospiraceae | Lachnospiraceae UCG-010 | NA |
| 451 | Genus | Firmicutes | Peptococcaceae | Peptococcus | NA |
| 452 | Genus | Firmicutes | Lachnospiraceae | Lachnospiraceae UCG-004 | NA |
| 453 | Genus | Firmicutes | Lachnospiraceae | Lachnospiraceae UCG-001 | NA |
| 454 | Genus | Firmicutes | Oscillospiraceae | Flavonifractor | NA |
| 455 | Genus | Firmicutes | Streptococcaceae | Streptococcus | NA |
| 456 | Genus | Firmicutes | Acholeplasmataceae | Anaeroplasma | NA |
| 457 | Genus | Bacteroidota | Prevotellaceae | Alloprevotella | NA |
| 458 | Genus | Firmicutes | Lachnospiraceae | Tyzzerella | NA |
| 459 | Genus | Euryarchaeota | Methanobacteriaceae | Methanosphaera | NA |
| 460 | Genus | Firmicutes | Erysipelotrichaceae | Solobacterium | NA |
| 461 | Genus | Firmicutes | Lachnospiraceae | Lachnospiraceae AC2044 group | NA |
| 462 | Genus | Euryarchaeota | Methanobacteriaceae | Methanobrevibacter | NA |
| 463 | Genus | Firmicutes | Ruminococcaceae | CAG-352 | NA |
| 464 | Genus | Firmicutes | Clostridiaceae | Clostridium sensu stricto 1 | NA |
| 465 | Genus | Actinobacteriota | Eggerthellaceae | Slackia | NA |
| 466 | Genus | Firmicutes | Oscillospiraceae | UCG-002 | NA |
| 467 | Genus | Firmicutes | Erysipelotrichaceae | Catenisphaera | NA |
| 468 | Genus | Firmicutes | Anaerovoracaceae | Mogibacterium | NA |
| 469 | Genus | Firmicutes | Ruminococcaceae | Negativibacillus | NA |
| 470 | Genus | Firmicutes | Streptococcaceae | Lactococcus | NA |
| 471 | Genus | Firmicutes | Ruminococcaceae | Subdoligranulum | NA |
| 472 | Genus | Proteobacteria | Enterobacteriaceae | Escherichia-Shigella | NA |
| 473 | Genus | Firmicutes | Lachnospiraceae | GCA-900066575 | NA |
| 474 | Genus | Firmicutes | Veillonellaceae | Veillonella | NA |
| 475 | Genus | Firmicutes | Oscillospiraceae | Colidextribacter | NA |
| 476 | Genus | Bacteroidota | Rikenellaceae | Alistipes | NA |

| Ticker | DESeq2_baseMean | DESeq2_log2-FoldChange | DESeq2_lfcSE | DESeq2_stat | DESeq2_pvalue | DESeq2_padj | DESeq2 converged | ANCOM_W | ANCOM_detected_0.7 |
|---|---|---|---|---|---|---|---|---|---|
| 447 | 726,241955 | -1,22040181 | 0,536730445 | -2,273770423 | 0,022979793 | 0,490235594 | NO | 0 | FALSE |
| 448 | 43,51055513 | 0,331048568 | 0,653501847 | 0,506576331 | 0,612452119 | 0,999339594 | YES | 0 | FALSE |
| 449 | 46,25841108 | -0,571486126 | 0,529294718 | -1,079712504 | 0,280270224 | 0,999339594 | YES | 0 | FALSE |
| 450 | 42,29156391 | -0,155713374 | 0,569224259 | -0,27355365 | 0,784427658 | 0,999339594 | YES | 0 | FALSE |
| 451 | 74,43544401 | 0,411556638 | 0,612732035 | 0,671674754 | 0,501790777 | 0,999339594 | YES | 0 | FALSE |
| 452 | 39,18375991 | -0,901353627 | 0,680818083 | -1,323927271 | 0,1855272 | 0,999339594 | YES | 0 | FALSE |
| 453 | 61,35643696 | -0,608464113 | 0,855323811 | -0,711384513 | 0,476845993 | 0,999339594 | YES | 0 | FALSE |
| 454 | 7,524681296 | -0,675880121 | 2,902611725 | -0,232852405 | 0,815876018 | 0,999339594 | YES | 0 | FALSE |
| 455 | 183,201132 | 0,601465278 | 0,472504514 | 1,272930227 | 0,203042802 | 0,999339594 | YES | 0 | FALSE |
| 456 | 38,90305883 | 0,099333137 | 2,909972305 | 0,034135424 | 0,972769161 | 0,999339594 | YES | 0 | FALSE |
| 457 | 125,4365284 | 0,475947641 | 2,908657411 | 0,163631385 | 0,870021336 | 0,999339594 | YES | 0 | FALSE |
| 458 | 68,38846969 | 0,219383784 | 1,504646356 | 0,145804217 | 0,884075947 | 0,999339594 | YES | 0 | FALSE |
| 459 | 10,94265096 | 0,234918627 | 2,910726675 | 0,080707897 | 0,935674257 | 0,999339594 | YES | 0 | FALSE |
| 460 | 16,7696842 | 0,184863679 | 2,910681902 | 0,063512155 | 0,949358681 | 0,999339594 | YES | 0 | FALSE |
| 461 | 89,88255606 | 0,255102721 | 1,06137139 | 0,240351986 | 0,810057397 | 0,999339594 | YES | 0 | FALSE |
| 462 | 227,7305299 | 0,844874941 | 0,440128742 | 1,91960865 | 0,054907351 | 0,702814088 | YES | 0 | FALSE |
| 463 | 344,5054983 | -1,436041944 | 1,66842368 | -0,860717791 | 0,389393492 | 0,999339594 | YES | 0 | FALSE |
| 464 | 97,80632938 | 0,671092025 | 0,434823173 | 1,543367665 | 0,122741594 | 0,872829116 | YES | 0 | FALSE |
| 465 | 10,71585446 | -0,482576995 | 1,019160763 | -0,473504292 | 0,63585344 | 0,999339594 | YES | 0 | FALSE |
| 466 | 2038,191126 | 0,081397418 | 0,173620291 | 0,468824337 | 0,639195203 | 0,999339594 | YES | 0 | FALSE |
| 467 | 23,74637616 | 0,080984676 | 2,910935276 | 0,027820844 | 0,977805041 | 0,999339594 | YES | 0 | FALSE |
| 468 | 24,31824105 | 0,151269106 | 2,909268743 | 0,051995577 | 0,958532218 | 0,999339594 | YES | 0 | FALSE |
| 469 | 38,51324307 | 0,735815666 | 0,557209702 | 1,32053635 | 0,186656008 | 0,999339594 | YES | 0 | FALSE |
| 470 | 8,931084815 | 0,550318221 | 2,910658975 | 0,189069975 | 0,850037973 | 0,999339594 | YES | 0 | FALSE |
| 471 | 1006,420478 | -0,020267531 | 0,234184764 | -0,086545045 | 0,93103315 | 0,999339594 | YES | 0 | FALSE |
| 472 | 726,1129814 | -0,400048794 | 0,570386875 | -0,701363953 | 0,483075912 | 0,999339594 | YES | 0 | FALSE |
| 473 | 14,47144605 | 0,070255156 | 1,133604861 | 0,061974995 | 0,950582745 | 0,999339594 | YES | 0 | FALSE |
| 474 | 24,55111242 | -1,954654013 | 2,33977716 | -0,835401784 | 0,403491526 | 0,999339594 | YES | 0 | FALSE |
| 475 | 92,9260607 | -0,340817385 | 0,23666391 | -1,440090232 | 0,149841872 | 0,999339594 | YES | 0 | FALSE |
| 476 | 1172,576683 | 0,008189579 | 0,145929361 | 0,056120162 | 0,955246082 | 0,999339594 | YES | 0 | FALSE |

| Ticker | taxLevel | Phylum | Family | Genus | Species |
|---|---|---|---|---|---|
| 477 | Genus | Firmicutes | Anaerovoracaceae | Family XIII UCG-001 | NA |
| 478 | Genus | Firmicutes | Anaerovoracaceae | Family XIII AD3011 group | NA |
| 479 | Genus | Firmicutes | Anaerovoracaceae | [Eubacterium] brachy group | NA |
| 480 | Genus | Firmicutes | Peptostreptococcaceae | Intestinibacter | NA |
| 481 | Genus | Firmicutes | Lachnospiraceae | Lachnotalea | NA |
| 482 | Genus | Firmicutes | Lachnospiraceae | Anaerostipes | NA |
| 483 | Genus | Firmicutes | Lachnospiraceae | Marvinbryantia | NA |
| 484 | Genus | Firmicutes | Lachnospiraceae | [Eubacterium] hallii group | NA |
| 485 | Genus | Bacteroidota | Barnesiellaceae | Coprobacter | NA |
| 486 | Genus | Firmicutes | Selenomonadaceae | Mitsuokella | NA |
| 487 | Genus | Firmicutes | Ruminococcaceae | DTU089 | NA |
| 488 | Genus | Bacteroidota | Tannerellaceae | Parabacteroides | NA |
| 489 | Genus | Firmicutes | Ruminococcaceae | Ruminococcus | NA |
| 490 | Genus | Firmicutes | Erysipelotrichaceae | Holdemanella | NA |
| 491 | Genus | Bacteroidota | Prevotellaceae | Paraprevotella | NA |
| 492 | Genus | Firmicutes | Lactobacillaceae | Lactobacillus | NA |
| 493 | Genus | Firmicutes | Oscillospiraceae | Oscillospira | NA |
| 494 | Genus | Bacteroidota | Prevotellaceae | Prevotellaceae UCG-001 | NA |
| 495 | Genus | Firmicutes | Erysipelatoclostridiaceae | Coprobacillus | NA |
| 496 | Genus | Actinobacteriota | Atopobiaceae | Olsenella | NA |
| 497 | Genus | Verrucomicrobiota | Victivallaceae | Victivallis | NA |
| 498 | Genus | Actinobacteriota | Actinomycetaceae | Actinomyces | NA |
| 499 | Genus | Firmicutes | Lachnospiraceae | Howardella | NA |
| 500 | Genus | Firmicutes | Lachnospiraceae | Eisenbergiella | NA |
| 501 | Genus | Firmicutes | Lachnospiraceae | Blautia | NA |
| 502 | Genus | Firmicutes | Oscillospiraceae | UCG-005 | NA |
| 503 | Genus | Proteobacteria | Oxalobacteraceae | Oxalobacter | NA |
| 504 | Genus | Firmicutes | Lachnospiraceae | [Ruminococcus] torques group | NA |
| 505 | Genus | Firmicutes | Veillonellaceae | Allisonella | NA |
| 506 | Genus | Bacteroidota | Marinifilaceae | Butyricimonas | NA |

| Ticker | DESeq2_baseMean | DESeq2_log2-FoldChange | DESeq2_lfcSE | DESeq2_stat | DESeq2_pvalue | DESeq2_padj | DESeq2 converged | ANCOM_W | ANCOM_detected_0.7 |
|---|---|---|---|---|---|---|---|---|---|
| 477 | 28,65779471 | 0,073070448 | 0,454631474 | 0,16072457 | 0,872310337 | 0,999339594 | YES | 0 | FALSE |
| 478 | 65,48340884 | 0,038996995 | 0,296491972 | 0,131527997 | 0,895357641 | 0,999339594 | YES | 0 | FALSE |
| 479 | 0,876784564 | 0,492728201 | 2,913608577 | 0,169112696 | 0,865707999 | 0,999339594 | YES | 0 | FALSE |
| 480 | 34,28616114 | 0,650481631 | 0,827334904 | 0,786237384 | 0,431728428 | 0,999339594 | YES | 0 | FALSE |
| 481 | 3,602630922 | -0,598845779 | 2,907626803 | -0,205956892 | 0,836824585 | 0,999339594 | YES | 0 | FALSE |
| 482 | 151,3924744 | -0,057955171 | 0,382074858 | -0,151685383 | 0,879435086 | 0,999339594 | NO | 0 | FALSE |
| 483 | 68,44937291 | 0,280076502 | 0,715505658 | 0,391438556 | 0,695473098 | 0,999339594 | YES | 0 | FALSE |
| 484 | 214,7423804 | -0,021364752 | 0,225754388 | -0,094637151 | 0,92460304 | 0,999339594 | YES | 0 | FALSE |
| 485 | 12,23923207 | 0,0113031 | 0,884221631 | 0,012783107 | 0,989800834 | 0,999339594 | YES | 0 | FALSE |
| 486 | 25,868923 | 0,002410423 | 2,912205846 | 0,000827697 | 0,999339594 | 0,999339594 | YES | 0 | FALSE |
| 487 | 10,14568357 | 0,375982382 | 1,141713633 | 0,329314086 | 0,741918298 | 0,999339594 | YES | 0 | FALSE |
| 488 | 632,4957271 | -0,128689005 | 0,154298334 | -0,834027184 | 0,404265665 | 0,999339594 | YES | 0 | FALSE |
| 489 | 1932,526695 | 0,051697632 | 0,256718541 | 0,201378644 | 0,840402513 | 0,999339594 | YES | 0 | FALSE |
| 490 | 263,2560422 | -0,073020267 | 0,284988889 | -0,256221452 | 0,797779845 | 0,999339594 | YES | 0 | FALSE |
| 491 | 171,8329806 | -0,064985958 | 0,38274493 | -0,169789206 | 0,865175916 | 0,999339594 | YES | 0 | FALSE |
| 492 | 146,5234805 | 0,753386293 | 0,985872532 | 0,764182253 | 0,444758641 | 0,999339594 | YES | 0 | FALSE |
| 493 | 29,82142639 | 0,597732827 | 1,106968319 | 0,539972841 | 0,589215763 | 0,999339594 | YES | 0 | FALSE |
| 494 | 105,4778736 | 0,116228704 | 2,912822151 | 0,039902437 | 0,968170908 | 0,999339594 | YES | 0 | FALSE |
| 495 | 8,161865312 | -0,947855215 | 2,909792631 | -0,325746655 | 0,744616042 | 0,999339594 | YES | 0 | FALSE |
| 496 | 5,319040721 | -0,070593213 | 2,909524119 | -0,024262804 | 0,980642983 | 0,999339594 | YES | 0 | FALSE |
| 497 | 8,206624497 | 0,49527236 | 2,664323825 | 0,185890452 | 0,85253067 | 0,999339594 | YES | 0 | FALSE |
| 498 | 0,9372743 | 0,362677486 | 2,911294615 | 0,124576017 | 0,900859216 | 0,999339594 | YES | 0 | FALSE |
| 499 | 9,07038158 | -0,441230292 | 1,919182283 | -0,229905359 | 0,818165312 | 0,999339594 | YES | 0 | FALSE |
| 500 | 8,023429284 | -7,600684696 | 2,911649238 | -2,610439677 | 0,009042592 | 0,385817252 | YES | 0 | FALSE |
| 501 | 1223,163228 | -0,095386938 | 0,158107043 | -0,603306066 | 0,546305098 | 0,999339594 | YES | 0 | FALSE |
| 502 | 722,6710778 | -0,142561651 | 0,229612583 | -0,620879086 | 0,534679183 | 0,999339594 | YES | 0 | FALSE |
| 503 | 16,05310098 | -0,002905897 | 0,465936031 | -0,006236687 | 0,995023876 | 0,999339594 | YES | 0 | FALSE |
| 504 | 559,9342945 | -0,002368141 | 0,236092839 | -0,01003055 | 0,991996913 | 0,999339594 | YES | 0 | FALSE |
| 505 | 9,952250596 | -0,178569051 | 2,906948773 | -0,061428345 | 0,951018079 | 0,999339594 | YES | 0 | FALSE |
| 506 | 48,10819097 | 0,542914023 | 0,300652505 | 1,805785797 | 0,070951805 | 0,814074526 | YES | 0 | FALSE |

| Ticker | taxLevel | Phylum | Family | Genus | Species |
|---|---|---|---|---|---|
| 507 | Genus | Firmicutes | Monoglobaceae | Monoglobus | NA |
| 508 | Genus | Bacteroidota | Rikenellaceae | Rikenellaceae RC9 gut group | NA |
| 509 | Genus | Firmicutes | Lachnospiraceae | Roseburia | NA |
| 510 | Genus | Firmicutes | Erysipelatoclostridiaceae | Erysipelatoclostridium | NA |
| 511 | Genus | Firmicutes | Ruminococcaceae | [Eubacterium] siraeum group | NA |
| 512 | Genus | Firmicutes | Erysipelatoclostridiaceae | Catenibacterium | NA |
| 513 | Genus | Firmicutes | Ruminococcaceae | Angelakisella | NA |
| 514 | Genus | Firmicutes | Ruminococcaceae | Incertae Sedis | NA |
| 515 | Genus | Firmicutes | Butyricicoccaceae | Butyricicoccus | NA |
| 516 | Genus | Firmicutes | Erysipelotrichaceae | Holdemania | NA |
| 517 | Genus | Firmicutes | Ruminococcaceae | Fournierella | NA |
| 518 | Genus | Firmicutes | Lachnospiraceae | Agathobacter | NA |
| 519 | Genus | Firmicutes | Lachnospiraceae | Hungatella | NA |
| 520 | Genus | Proteobacteria | Sutterellaceae | Parasutterella | NA |
| 521 | Genus | Firmicutes | Ruminococcaceae | Anaerotruncus | NA |
| 522 | Genus | Firmicutes | Lachnospiraceae | [Ruminococcus] gauvreauii group | NA |
| 523 | Genus | Firmicutes | Lachnospiraceae | Lachnospiraceae ND3007 group | NA |
| 524 | Genus | Firmicutes | Defluviitaleaceae | Defluviitaleaceae UCG-011 | NA |
| 525 | Genus | Firmicutes | Lachnospiraceae | [Eubacterium] eligens group | NA |
| 526 | Genus | Firmicutes | Lachnospiraceae | [Eubacterium] ruminantium group | NA |
| 527 | Genus | Firmicutes | Ruminococcaceae | Faecalibacterium | NA |
| 528 | Genus | Firmicutes | Lachnospiraceae | Lachnospiraceae NK4A136 group | NA |
| 529 | Genus | Firmicutes | Lachnospiraceae | Coprococcus | NA |
| 530 | Genus | Actinobacteriota | Atopobiaceae | Libanicoccus | NA |
| 531 | Genus | Proteobacteria | Sutterellaceae | Sutterella | NA |
| 532 | Genus | Firmicutes | Lachnospiraceae | Shuttleworthia | NA |
| 533 | Genus | Synergistota | Synergistaceae | Cloacibacillus | NA |
| 534 | Genus | Firmicutes | Erysipelotrichaceae | Turicibacter | NA |
| 535 | Genus | Firmicutes | Lachnospiraceae | CAG-56 | NA |
| 536 | Genus | Firmicutes | Lachnospiraceae | [Ruminococcus] gnavus group | NA |

| Ticker | DESeq2_baseMean | DESeq2_log2-FoldChange | DESeq2_lfcSE | DESeq2_stat | DESeq2_pvalue | DESeq2_padj | DESeq2 converged | ANCOM_W | ANCOM_detected_0.7 |
|---|---|---|---|---|---|---|---|---|---|
| 507 | 127,7908033 | 0,791647442 | 0,319640007 | 2,476684474 | 0,013260908 | 0,417919648 | YES | 23 | FALSE |
| 508 | 187,5269105 | -0,135632516 | 0,411705361 | -0,329440734 | 0,741822583 | 0,999339594 | YES | 0 | FALSE |
| 509 | 502,1298847 | -0,461092591 | 0,287171973 | -1,605632282 | 0,108354728 | 0,872829116 | YES | 0 | FALSE |
| 510 | 8,972415919 | -0,257453416 | 2,051030765 | -0,125523917 | 0,900108792 | 0,999339594 | YES | 0 | FALSE |
| 511 | 174,2386528 | 0,04461628 | 0,500770898 | 0,089095193 | 0,929006257 | 0,999339594 | YES | 0 | FALSE |
| 512 | 77,10698596 | -0,758253492 | 2,911555111 | -0,260429037 | 0,794532848 | 0,999339594 | YES | 0 | FALSE |
| 513 | 5,275101543 | 0,238098402 | 2,902028883 | 0,082045497 | 0,934610534 | 0,999339594 | YES | 0 | FALSE |
| 514 | 116,9962084 | 0,421315287 | 0,26699322 | 1,577999947 | 0,114565622 | 0,872829116 | YES | 0 | FALSE |
| 515 | 145,9429848 | -0,217401887 | 0,264955329 | -0,820522793 | 0,411918141 | 0,999339594 | YES | 0 | FALSE |
| 516 | 1,872431237 | -0,426413155 | 2,905156436 | -0,146778036 | 0,883307222 | 0,999339594 | YES | 0 | FALSE |
| 517 | 16,90710766 | 0,114852205 | 2,908571059 | 0,039487502 | 0,968501718 | 0,999339594 | YES | 0 | FALSE |
| 518 | 1903,075798 | -0,868011759 | 0,278200688 | -3,120092068 | 0,001807945 | 0,115708494 | YES | 7 | FALSE |
| 519 | 16,87569309 | -9,741488141 | 2,912341369 | -3,344899141 | 0,000823125 | 0,105359997 | YES | 0 | FALSE |
| 520 | 61,85286103 | 0,207744919 | 0,538053821 | 0,386104346 | 0,699419391 | 0,999339594 | YES | 0 | FALSE |
| 521 | 3,394205931 | 0,408740537 | 2,911443486 | 0,140391026 | 0,888351048 | 0,999339594 | YES | 0 | FALSE |
| 522 | 42,6394857 | 0,254642532 | 1,048527736 | 0,242857221 | 0,808116007 | 0,999339594 | YES | 0 | FALSE |
| 523 | 70,42894352 | -0,97069059 | 0,404190386 | -2,401567739 | 0,016324986 | 0,417919648 | YES | 1 | FALSE |
| 524 | 4,440207473 | -0,050849078 | 2,907766766 | -0,01748733 | 0,98604784 | 0,999339594 | YES | 0 | FALSE |
| 525 | 191,6833352 | 0,079287023 | 0,422459031 | 0,187679792 | 0,851127672 | 0,999339594 | YES | 0 | FALSE |
| 526 | 172,3895039 | -0,417744135 | 0,941580733 | -0,443662578 | 0,657286555 | 0,999339594 | YES | 0 | FALSE |
| 527 | 2421,900823 | -0,391629256 | 0,220953325 | -1,77245242 | 0,076319487 | 0,814074526 | YES | 0 | FALSE |
| 528 | 207,5213105 | -0,204970759 | 0,454421842 | -0,451058333 | 0,651947507 | 0,999339594 | YES | 0 | FALSE |
| 529 | 878,2528472 | -0,447522034 | 0,262008498 | -1,708043961 | 0,087628186 | 0,862800598 | YES | 0 | FALSE |
| 530 | 3,839444221 | 0,134825682 | 2,911538554 | 0,046307366 | 0,963065268 | 0,999339594 | YES | 0 | FALSE |
| 531 | 128,2324088 | -0,185470289 | 0,360154858 | -0,514973726 | 0,60657138 | 0,999339594 | YES | 0 | FALSE |
| 532 | 7,490118204 | 0,063169558 | 2,906479303 | 0,021734047 | 0,982660105 | 0,999339594 | YES | 0 | FALSE |
| 533 | 13,57382377 | 0,561986928 | 2,91052467 | 0,193087842 | 0,846890174 | 0,999339594 | YES | 0 | FALSE |
| 534 | 39,53899495 | 1,04794823 | 0,966663836 | 1,08408755 | 0,278325982 | 0,999339594 | YES | 0 | FALSE |
| 535 | 117,8471172 | -0,561989891 | 0,580225071 | -0,96857223 | 0,332758665 | 0,999339594 | YES | 0 | FALSE |
| 536 | 57,51803175 | 0,057951636 | 2,910242102 | 0,019912995 | 0,984112779 | 0,999339594 | YES | 0 | FALSE |

| Ticker | taxLevel | Phylum | Family | Genus | Species |
|---|---|---|---|---|---|
| 537 | Genus | Firmicutes | Butyricicoccaceae | UCG-009 | NA |
| 538 | Genus | Firmicutes | Erysipelotrichaceae | Dielma | NA |
| 539 | Genus | Firmicutes | Acidaminococcaceae | Phascolarctobacterium | NA |
| 540 | Family | Actinobacteriota | Bifidobacteriaceae | NA | NA |
| 541 | Family | Verrucomicrobiota | Akkermansiaceae | NA | NA |
| 542 | Family | Thermoplasmatota | Methanomethylophilaceae | NA | NA |
| 543 | Family | Verrucomicrobiota | vadinBE97 | NA | NA |
| 544 | Family | Firmicutes | Veillonellaceae | NA | NA |
| 545 | Family | Bacteroidota | Muribaculaceae | NA | NA |
| 546 | Family | Firmicutes | Christensenellaceae | NA | NA |
| 547 | Family | Firmicutes | Peptostreptococcaceae | NA | NA |
| 548 | Family | Desulfobacterota | Desulfovibrionaceae | NA | NA |
| 549 | Family | Actinobacteriota | Coriobacteriaceae | NA | NA |
| 550 | Family | Proteobacteria | Pasteurellaceae | NA | NA |
| 551 | Family | Firmicutes | Erysipelatoclostridiaceae | NA | NA |
| 552 | Family | Bacteroidota | Marinifilaceae | NA | NA |
| 553 | Family | Bacteroidota | Barnesiellaceae | NA | NA |
| 554 | Family | Bacteroidota | Bacteroidaceae | NA | NA |
| 555 | Family | Bacteroidota | Prevotellaceae | NA | NA |
| 556 | Family | Firmicutes | Peptococcaceae | NA | NA |
| 557 | Family | Firmicutes | Streptococcaceae | NA | NA |
| 558 | Family | Firmicutes | Acholeplasmataceae | NA | NA |
| 559 | Family | Actinobacteriota | Coriobacteriales Incertae Sedis | NA | NA |
| 560 | Family | Euryarchaeota | Methanobacteriaceae | NA | NA |
| 561 | Family | Firmicutes | Clostridiaceae | NA | NA |
| 562 | Family | Firmicutes | Oscillospiraceae | NA | NA |
| 563 | Family | Proteobacteria | Enterobacteriaceae | NA | NA |
| 564 | Family | Bacteroidota | Rikenellaceae | NA | NA |
| 565 | Family | Firmicutes | Anaerovoracaceae | NA | NA |
| 566 | Family | Firmicutes | Selenomonadaceae | NA | NA |

| Ticker | DESeq2_baseMean | DESeq2_log2-FoldChange | DESeq2_lfcSE | DESeq2_stat | DESeq2_pvalue | DESeq2_padj | DESeq2 converged | ANCOM_W | ANCOM_detected_0.7 |
|---|---|---|---|---|---|---|---|---|---|
| 537 | 13,40761928 | 0,983012713 | 1,830964091 | 0,536882574 | 0,591348729 | 0,999339594 | YES | 0 | FALSE |
| 538 | 2,467284572 | -0,10160205 | 2,91134735 | -0,034898636 | 0,972160569 | 0,999339594 | YES | 0 | FALSE |
| 539 | 310,2423739 | -0,258917913 | 0,263008562 | -0,984446708 | 0,324895922 | 0,999339594 | YES | 0 | FALSE |
| 540 | 696,4177771 | 0,076073944 | 0,231363549 | 0,328806955 | 0,742301606 | 0,999467513 | NO | 0 | FALSE |
| 541 | 911,7268497 | -0,108773051 | 0,43760337 | -0,248565386 | 0,803696986 | 0,999467513 | YES | 0 | FALSE |
| 542 | 4,633680537 | -0,021740304 | 2,912864226 | -0,007463549 | 0,994045005 | 0,999467513 | YES | 0 | FALSE |
| 543 | 8,370674172 | 0,296732355 | 2,906930207 | 0,102077564 | 0,918695109 | 0,999467513 | YES | 0 | FALSE |
| 544 | 642,4361392 | -0,101359529 | 0,415740764 | -0,243804644 | 0,807382123 | 0,999467513 | YES | 0 | FALSE |
| 545 | 382,535211 | -0,072061634 | 0,473607805 | -0,152154659 | 0,879064954 | 0,999467513 | YES | 0 | FALSE |
| 546 | 1661,879145 | 0,219687986 | 0,248485617 | 0,884107452 | 0,376638211 | 0,951507059 | YES | 0 | FALSE |
| 547 | 87,91799439 | 0,206141179 | 0,449272077 | 0,458833721 | 0,646353577 | 0,999467513 | YES | 0 | FALSE |
| 548 | 324,543189 | -0,060885017 | 0,168019774 | -0,362368164 | 0,717076927 | 0,999467513 | YES | 0 | FALSE |
| 549 | 198,2351386 | -0,378168933 | 0,300443583 | -1,258701982 | 0,208137995 | 0,624413986 | YES | 0 | FALSE |
| 550 | 9,306920299 | -2,200341946 | 1,682712729 | -1,307615915 | 0,191003616 | 0,611211571 | YES | 0 | FALSE |
| 551 | 261,6799201 | -0,414448187 | 0,195992434 | -2,11461319 | 0,034462929 | 0,4200768 | YES | 0 | FALSE |
| 552 | 93,33142425 | 0,118861793 | 0,206439599 | 0,575770312 | 0,564770443 | 0,999467513 | YES | 0 | FALSE |
| 553 | 328,3399431 | -0,615320229 | 0,294644595 | -2,088347247 | 0,036766522 | 0,4200768 | YES | 0 | FALSE |
| 554 | 4686,872673 | -0,199736352 | 0,14273258 | -1,399374641 | 0,161700667 | 0,567822391 | YES | 0 | FALSE |
| 555 | 1322,65234 | -0,563861623 | 0,370369537 | -1,522429808 | 0,127901421 | 0,567822391 | YES | 0 | FALSE |
| 556 | 86,5675905 | 0,177295434 | 0,347885977 | 0,509636622 | 0,610306062 | 0,999467513 | YES | 0 | FALSE |
| 557 | 224,4978492 | 0,676046079 | 0,453521549 | 1,490659221 | 0,136050988 | 0,567822391 | YES | 0 | FALSE |
| 558 | 32,18864131 | 0,001942055 | 2,909995084 | 0,000667374 | 0,999467513 | 0,999467513 | YES | 0 | FALSE |
| 559 | 24,93678287 | 0,285213805 | 0,821661005 | 0,347118584 | 0,728502232 | 0,999467513 | YES | 0 | FALSE |
| 560 | 205,3982853 | 0,694902665 | 0,422771256 | 1,643684747 | 0,100241338 | 0,567822391 | YES | 0 | FALSE |
| 561 | 118,888961 | 0,509078808 | 0,436429956 | 1,166461654 | 0,243427844 | 0,687325676 | YES | 0 | FALSE |
| 562 | 3614,639811 | -0,040589626 | 0,120729512 | -0,336203013 | 0,736717781 | 0,999467513 | YES | 0 | FALSE |
| 563 | 964,6541405 | -0,406096286 | 0,518214152 | -0,783645688 | 0,433248039 | 0,999467513 | YES | 0 | FALSE |
| 564 | 1368,812 | -0,233288085 | 0,140726568 | -1,657740175 | 0,097369921 | 0,567822391 | YES | 0 | FALSE |
| 565 | 121,6766104 | 0,106145549 | 0,204819262 | 0,518240071 | 0,60429078 | 0,999467513 | YES | 0 | FALSE |
| 566 | 19,43253785 | -0,108111539 | 2,912158792 | -0,037124191 | 0,970385984 | 0,999467513 | YES | 0 | FALSE |

| Ticker | taxLevel | Phylum | Family | Genus | Species |
|---|---|---|---|---|---|
| 567 | Family | Bacteroidota | Tannerellaceae | NA | NA |
| 568 | Family | Firmicutes | Ruminococcaceae | NA | NA |
| 569 | Family | Firmicutes | Erysipelotrichaceae | NA | NA |
| 570 | Family | Firmicutes | Lactobacillaceae | NA | NA |
| 571 | Family | Firmicutes | UCG-010 | NA | NA |
| 572 | Family | Actinobacteriota | Atopobiaceae | NA | NA |
| 573 | Family | Verrucomicrobiota | Victivallaceae | NA | NA |
| 574 | Family | Actinobacteriota | Actinomycetaceae | NA | NA |
| 575 | Family | Proteobacteria | Oxalobacteraceae | NA | NA |
| 576 | Family | Firmicutes | Monoglobaceae | NA | NA |
| 577 | Family | Firmicutes | [Eubacterium] coprostanoligenes group | NA | NA |
| 578 | Family | Actinobacteriota | Eggerthellaceae | NA | NA |
| 579 | Family | Firmicutes | Butyricicoccaceae | NA | NA |
| 580 | Family | Firmicutes | Lachnospiraceae | NA | NA |
| 581 | Family | Firmicutes | Hydrogenoanaerobacterium | NA | NA |
| 582 | Family | Proteobacteria | Sutterellaceae | NA | NA |
| 583 | Family | Verrucomicrobiota | Puniceicoccaceae | NA | NA |
| 584 | Family | Firmicutes | Defluviitaleaceae | NA | NA |
| 585 | Family | Firmicutes | Ruminiclostridium | NA | NA |
| 586 | Family | Synergistota | Synergistaceae | NA | NA |
| 587 | Family | Firmicutes | Acidaminococcaceae | NA | NA |

| Ticker | DESeq2_baseMean | DESeq2_log2-FoldChange | DESeq2_lfcSE | DESeq2_stat | DESeq2_pvalue | DESeq2_padj | DESeq2 converged | ANCOM_W | ANCOM_detected_0.7 |
|---|---|---|---|---|---|---|---|---|---|
| 567 | 702,7423541 | -0,230909683 | 0,153578179 | -1,503531848 | 0,132701951 | 0,567822391 | YES | 0 | FALSE |
| 568 | 6758,921304 | -0,224905058 | 0,10951638 | -2,053620267 | 0,040012459 | 0,4200768 | YES | 0 | FALSE |
| 569 | 380,6336525 | -0,349249325 | 0,251905025 | -1,386432547 | 0,165614864 | 0,567822391 | YES | 0 | FALSE |
| 570 | 185,0695562 | 0,667341809 | 0,928329112 | 0,718863386 | 0,472225098 | 0,999467513 | YES | 0 | FALSE |
| 571 | 194,0721117 | -0,030826575 | 0,277920648 | -0,110918621 | 0,91168088 | 0,999467513 | YES | 0 | FALSE |
| 572 | 17,34544845 | -0,209457002 | 0,987045474 | -0,212206031 | 0,831946302 | 0,999467513 | YES | 0 | FALSE |
| 573 | 11,35129158 | 0,06004212 | 1,341397885 | 0,044760858 | 0,964297925 | 0,999467513 | YES | 0 | FALSE |
| 574 | 1,22960968 | 0,19979114 | 2,909599716 | 0,068666195 | 0,945255327 | 0,999467513 | YES | 0 | FALSE |
| 575 | 15,30891264 | 0,001742285 | 0,451198152 | 0,003861463 | 0,996919006 | 0,999467513 | YES | 0 | FALSE |
| 576 | 135,7952013 | 0,663699213 | 0,329150313 | 2,016401584 | 0,043758 | 0,4200768 | YES | 10 | FALSE |
| 577 | 470,7983445 | 0,091749496 | 0,158083447 | 0,580386481 | 0,56165402 | 0,999467513 | YES | 0 | FALSE |
| 578 | 144,578283 | -0,285012523 | 0,191451088 | -1,488696282 | 0,136567364 | 0,567822391 | YES | 0 | FALSE |
| 579 | 230,3853905 | -0,236322509 | 0,233290995 | -1,012994561 | 0,311062759 | 0,829500691 | YES | 0 | FALSE |
| 580 | 10353,20266 | -0,38151699 | 0,129662027 | -2,942395709 | 0,003256835 | 0,156328064 | YES | 1 | FALSE |
| 581 | 2,829129513 | -0,284645616 | 2,910940574 | -0,097784757 | 0,922103212 | 0,999467513 | YES | 0 | FALSE |
| 582 | 194,2738504 | -0,325446759 | 0,226022629 | -1,439885735 | 0,149899729 | 0,567822391 | YES | 0 | FALSE |
| 583 | 6,964085125 | 0,169080604 | 2,906981304 | 0,058163637 | 0,953618286 | 0,999467513 | YES | 0 | FALSE |
| 584 | 3,371600749 | -0,094446325 | 2,907948444 | -0,032478679 | 0,974090319 | 0,999467513 | YES | 0 | FALSE |
| 585 | 4,886888113 | 0,185004328 | 2,911145074 | 0,063550364 | 0,949328256 | 0,999467513 | YES | 0 | FALSE |
| 586 | 11,69017375 | 0,234808109 | 2,909069974 | 0,080715868 | 0,935667917 | 0,999467513 | YES | 0 | FALSE |
| 587 | 360,9905604 | -0,202363616 | 0,261646586 | -0,773423491 | 0,439271792 | 0,999467513 | NO | 0 | FALSE |

**Supplementary Table 7: differential abundance at ASV, genus and family level in dopamine agonist (a. any dopamine agonist, b. pramipexole, c. ropinirole) users with DESeq2 and ANCOM**

**All columns are spread out over two pages, after which the next two pages show the next set of rows, etc.**
**A ticker is provided on both pages to easily connect the taxonomy data to the associated statistics.**

**Supplementary Table 7a: users of any dopamine agonist without concurrent levodopa therapy**

| Medication | Ticker | taxLevel | Phylum | Family | Genus | Species |
|---|---|---|---|---|---|---|
| Dopamine agonists | 1 | ASV1 | Actinobacteriota | Bifidobacteriaceae | Bifidobacterium | NA |
| Dopamine agonists | 2 | ASV | Verrucomicrobiota | Akkermansiaceae | Akkermansia | muciniphila |
| Dopamine agonists | 3 | ASV | Firmicutes | Christensenellaceae | NA | NA |
| Dopamine agonists | 4 | ASV | Firmicutes | Lachnospiraceae | Roseburia | NA |
| Dopamine agonists | 5 | ASV | Firmicutes | NA | NA | NA |
| Dopamine agonists | 6 | ASV | Firmicutes | Christensenellaceae | Christensenellaceae R-7 group | NA |
| Dopamine agonists | 7 | ASV | Bacteroidota | Bacteroidaceae | Bacteroides | NA |
| Dopamine agonists | 8 | ASV | Bacteroidota | Bacteroidaceae | Bacteroides | eggerthii |
| Dopamine agonists | 9 | ASV | Firmicutes | UCG-010 | NA | NA |
| Dopamine agonists | 10 | ASV | Firmicutes | [Eubacterium] coprostanoligenes group | NA | NA |
| Dopamine agonists | 11 | ASV | Firmicutes | Ruminococcaceae | Subdoligranulum | NA |
| Dopamine agonists | 12 | ASV | Firmicutes | Ruminococcaceae | Faecalibacterium | prausnitzii |
| Dopamine agonists | 13 | ASV | Firmicutes | Lachnospiraceae | Lachnospira | pectinoschiza |
| Dopamine agonists | 14 | ASV | Bacteroidota | Bacteroidaceae | Bacteroides | coprocola |
| Dopamine agonists | 15 | ASV | Firmicutes | Ruminococcaceae | NA | NA |
| Dopamine agonists | 16 | ASV | Firmicutes | Oscillospiraceae | UCG-003 | NA |
| Dopamine agonists | 17 | ASV | Bacteroidota | Rikenellaceae | Alistipes | finegoldii |
| Dopamine agonists | 18 | ASV | Firmicutes | NA | NA | NA |
| Dopamine agonists | 19 | ASV | Firmicutes | Ruminococcaceae | Subdoligranulum | NA |
| Dopamine agonists | 20 | ASV | Firmicutes | Veillonellaceae | Dialister | invisus |
| Dopamine agonists | 21 | ASV | Firmicutes | Lachnospiraceae | Anaerostipes | hadrus |
| Dopamine agonists | 22 | ASV | Firmicutes | Lachnospiraceae | [Ruminococcus] torques group | NA |
| Dopamine agonists | 23 | ASV | Firmicutes | NA | NA | NA |
| Dopamine agonists | 24 | ASV | Firmicutes | Oscillospiraceae | UCG-002 | NA |

| Ticker | DESeq2_baseMean | DESeq2_log2-FoldChange | DESeq2_lfcSE | DESeq2_stat | DESeq2_pvalue | DESeq2_padj | DESeq2 converged | ANCOM_W | ANCOM_detected_0.7 |
|---|---|---|---|---|---|---|---|---|---|
| 1 | 400,8590305 | 2,367754236 | 1,745278402 | 1,356662772 | 0,174888386 | 0,998956236 | YES | 0 | FALSE |
| 2 | 391,9776858 | -1,50714991 | 2,352262596 | -0,640723494 | 0,521702347 | 0,998956236 | YES | 0 | FALSE |
| 3 | 145,4880729 | 0,539526538 | 2,949779672 | 0,182904012 | 0,854873335 | 0,998956236 | YES | 0 | FALSE |
| 4 | 53,73004373 | 2,233947447 | 2,971325352 | 0,751835354 | 0,452150078 | 0,998956236 | YES | 0 | FALSE |
| 5 | 1,655977854 | 2,119017011 | 2,941765051 | 0,720321635 | 0,471326987 | 0,998956236 | YES | 0 | FALSE |
| 6 | 42,31758336 | -0,299173763 | 3,013957826 | -0,099262757 | 0,920929648 | 0,998956236 | YES | 0 | FALSE |
| 7 | 50,52826394 | 0,088443609 | 2,991439905 | 0,029565564 | 0,976413529 | 0,998956236 | YES | 0 | FALSE |
| 8 | 134,4484627 | 0,031291742 | 3,01371755 | 0,010383104 | 0,991715631 | 0,998956236 | YES | 0 | FALSE |
| 9 | 0,184537664 | 0,575653944 | 3,044083828 | 0,189105812 | 0,850009887 | 0,998956236 | YES | 0 | FALSE |
| 10 | 56,50189389 | -0,481221245 | 2,970708898 | -0,161988691 | 0,87131476 | 0,998956236 | YES | 0 | FALSE |
| 11 | 312,1109785 | 0,166115997 | 1,354350884 | 0,12265359 | 0,902381416 | 0,998956236 | YES | 0 | FALSE |
| 12 | 27,57702312 | -0,36418004 | 3,036809092 | -0,119921941 | 0,904544984 | 0,998956236 | YES | 0 | FALSE |
| 13 | 57,02858643 | -0,696376317 | 2,049269025 | -0,339816934 | 0,733994395 | 0,998956236 | YES | 0 | FALSE |
| 14 | 7,967951352 | -0,363825783 | 3,03695335 | -0,119799596 | 0,904641902 | 0,998956236 | YES | 0 | FALSE |
| 15 | 2,083762464 | 0,412884929 | 3,037608468 | 0,135924341 | 0,891881093 | 0,998956236 | YES | 0 | FALSE |
| 16 | 127,1856264 | -0,369390746 | 2,211639432 | -0,167021234 | 0,867353342 | 0,998956236 | YES | 0 | FALSE |
| 17 | 28,32546676 | 0,94179992 | 2,992214795 | 0,314750105 | 0,752951398 | 0,998956236 | YES | 0 | FALSE |
| 18 | 63,10985334 | 1,876915025 | 3,014051501 | 0,622721617 | 0,533467475 | 0,998956236 | YES | 0 | FALSE |
| 19 | 10,53852819 | -1,238142715 | 3,019441852 | -0,410056817 | 0,681764269 | 0,998956236 | YES | 0 | FALSE |
| 20 | 1017,182378 | 0,865672696 | 2,491347884 | 0,347471624 | 0,728237033 | 0,998956236 | YES | 0 | FALSE |
| 21 | 74,08795427 | -0,37735732 | 2,974814356 | -0,126850712 | 0,899058557 | 0,998956236 | YES | 0 | FALSE |
| 22 | 18,34931008 | -0,011450114 | 3,01397297 | -0,00379901 | 0,996968836 | 0,998956236 | YES | 0 | FALSE |
| 23 | 168,5745109 | 0,77730252 | 3,013899072 | 0,257905955 | 0,796479489 | 0,998956236 | YES | 0 | FALSE |
| 24 | 17,2853955 | 0,175490463 | 3,014285124 | 0,058219596 | 0,953573712 | 0,998956236 | YES | 0 | FALSE |

| Medication | Ticker | taxLevel | Phylum | Family | Genus | Species |
|---|---|---|---|---|---|---|
| Dopamine agonists | 25 | ASV | Bacteroidota | Tannerellaceae | Parabacteroides | distasonis |
| Dopamine agonists | 26 | ASV | Firmicutes | Lachnospiraceae | Lachnospiraceae NK4A136 group | bacterium |
| Dopamine agonists | 27 | ASV | Firmicutes | Oscillospiraceae | NK4A214 group | NA |
| Dopamine agonists | 28 | ASV | Firmicutes | Christensenellaceae | Christensenellaceae R-7 group | NA |
| Dopamine agonists | 29 | ASV | Firmicutes | Lachnospiraceae | Fusicatenibacter | saccharivorans |
| Dopamine agonists | 30 | ASV | Firmicutes | Lachnospiraceae | NA | NA |
| Dopamine agonists | 31 | ASV | Firmicutes | Ruminococcaceae | CAG-352 | NA |
| Dopamine agonists | 32 | ASV | Actinobacteriota | Eggerthellaceae | Senegalimassilia | anaerobia |
| Dopamine agonists | 33 | ASV | Actinobacteriota | Eggerthellaceae | Enterorhabdus | NA |
| Dopamine agonists | 34 | ASV | Bacteroidota | Rikenellaceae | Alistipes | inops |
| Dopamine agonists | 35 | ASV | Firmicutes | Ruminococcaceae | NA | NA |
| Dopamine agonists | 36 | ASV | Firmicutes | Butyricicoccaceae | Butyricicoccus | faecihominis |
| Dopamine agonists | 37 | ASV | Bacteroidota | Rikenellaceae | Alistipes | obesi |
| Dopamine agonists | 38 | ASV | Firmicutes | Christensenellaceae | Christensenellaceae R-7 group | NA |
| Dopamine agonists | 39 | ASV | Firmicutes | Lachnospiraceae | Blautia | massiliensis |
| Dopamine agonists | 40 | ASV | Bacteroidota | Tannerellaceae | Parabacteroides | distasonis |
| Dopamine agonists | 41 | ASV | Firmicutes | Lachnospiraceae | Lachnoclostridium | NA |
| Dopamine agonists | 42 | ASV | Firmicutes | Ruminococcaceae | Faecalibacterium | prausnitzii |
| Dopamine agonists | 43 | ASV | Firmicutes | Peptostreptococcaceae | Terrisporobacter | mayombei |
| Dopamine agonists | 44 | ASV | Desulfobacterota | Desulfovibrionaceae | Bilophila | wadsworthia |
| Dopamine agonists | 45 | ASV | Bacteroidota | Muribaculaceae | NA | NA |
| Dopamine agonists | 46 | ASV | Firmicutes | Erysipelotrichaceae | Turicibacter | sanguinis |
| Dopamine agonists | 47 | ASV | Firmicutes | Peptococcaceae | NA | NA |
| Dopamine agonists | 48 | ASV | Firmicutes | Oscillospiraceae | Oscillibacter | NA |
| Dopamine agonists | 49 | ASV | Firmicutes | NA | NA | NA |
| Dopamine agonists | 50 | ASV | Firmicutes | Oscillospiraceae | UCG-002 | NA |
| Dopamine agonists | 51 | ASV | Firmicutes | Christensenellaceae | Christensenellaceae R-7 group | NA |
| Dopamine agonists | 52 | ASV | Proteobacteria | Sutterellaceae | Sutterella | NA |
| Dopamine agonists | 53 | ASV | Firmicutes | Butyricicoccaceae | Butyricicoccus | NA |
| Dopamine agonists | 54 | ASV | Firmicutes | Lachnospiraceae | Roseburia | hominis |

| Ticker | DESeq2_baseMean | DESeq2_log2-FoldChange | DESeq2_lfcSE | DESeq2_stat | DESeq2_pvalue | DESeq2_padj | DESeq2 converged | ANCOM_W | ANCOM_detected_0.7 |
|---|---|---|---|---|---|---|---|---|---|
| 25 | 111,0222979 | 0,578865383 | 2,507743292 | 0,230831196 | 0,817445945 | 0,998956236 | YES | 0 | FALSE |
| 26 | 223,0800973 | -1,059171201 | 2,29578743 | -0,461354212 | 0,644544497 | 0,998956236 | YES | 0 | FALSE |
| 27 | 78,55255695 | 0,38505527 | 2,971500026 | 0,129582792 | 0,896896519 | 0,998956236 | YES | 0 | FALSE |
| 28 | 29,06788126 | -0,470677731 | 3,014490801 | -0,156138387 | 0,875923941 | 0,998956236 | YES | 0 | FALSE |
| 29 | 717,8497493 | 1,437296715 | 1,82587695 | 0,787181587 | 0,431175574 | 0,998956236 | YES | 0 | FALSE |
| 30 | 205,1227669 | 0,652938243 | 2,018669312 | 0,323449829 | 0,746354598 | 0,998956236 | YES | 0 | FALSE |
| 31 | 4,576534068 | 0,595710688 | 3,037061265 | 0,196147076 | 0,84449505 | 0,998956236 | YES | 0 | FALSE |
| 32 | 6,284731383 | 1,047711302 | 2,979836623 | 0,35160025 | 0,725138078 | 0,998956236 | YES | 0 | FALSE |
| 33 | 10,16755101 | -1,194660519 | 2,293003671 | -0,521002445 | 0,602365068 | 0,998956236 | YES | 1 | FALSE |
| 34 | 141,1948633 | 0,865746764 | 2,608392794 | 0,331908126 | 0,739958634 | 0,998956236 | YES | 0 | FALSE |
| 35 | 64,64348299 | -0,120305132 | 2,97056733 | -0,040499042 | 0,96769527 | 0,998956236 | YES | 0 | FALSE |
| 36 | 11,06828922 | 1,18489137 | 2,993322816 | 0,395844833 | 0,692219501 | 0,998956236 | YES | 0 | FALSE |
| 37 | 80,9778734 | 2,874273705 | 1,928529017 | 1,490396919 | 0,136119902 | 0,998956236 | YES | 0 | FALSE |
| 38 | 10,7821158 | 0,175474364 | 3,014303796 | 0,058213895 | 0,953578253 | 0,998956236 | YES | 0 | FALSE |
| 39 | 451,7473085 | 0,581965058 | 1,388282533 | 0,419197854 | 0,675071541 | 0,998956236 | YES | 0 | FALSE |
| 40 | 36,88112669 | -0,175788268 | 3,03687257 | -0,057884638 | 0,95384052 | 0,998956236 | YES | 0 | FALSE |
| 41 | 10,93357082 | 0,546043838 | 3,014486851 | 0,181139897 | 0,856257769 | 0,998956236 | YES | 0 | FALSE |
| 42 | 60,19932348 | -0,605030713 | 3,014001281 | -0,200740032 | 0,840901856 | 0,998956236 | YES | 0 | FALSE |
| 43 | 7,678192803 | -1,236590746 | 3,019774577 | -0,409497701 | 0,682174454 | 0,998956236 | YES | 0 | FALSE |
| 44 | 54,48100851 | 0,017172571 | 2,011822089 | 0,00853583 | 0,993189476 | 0,998956236 | YES | 0 | FALSE |
| 45 | 304,0847259 | -0,181486224 | 2,991170598 | -0,06067398 | 0,951618855 | 0,998956236 | YES | 0 | FALSE |
| 46 | 5,227121934 | 0,505455289 | 3,002354943 | 0,168352942 | 0,866305626 | 0,998956236 | YES | 0 | FALSE |
| 47 | 1,545222137 | 0,36113028 | 3,037657604 | 0,118884459 | 0,905366895 | 0,998956236 | YES | 0 | FALSE |
| 48 | 147,7543168 | 0,334916589 | 1,906642487 | 0,175657781 | 0,860562805 | 0,998956236 | YES | 0 | FALSE |
| 49 | 11,98018217 | -0,363990976 | 3,036885435 | -0,11985667 | 0,904596689 | 0,998956236 | YES | 0 | FALSE |
| 50 | 46,70146365 | 0,479829887 | 2,993336271 | 0,16029936 | 0,872645263 | 0,998956236 | YES | 0 | FALSE |
| 51 | 67,81235193 | -25,09372524 | 3,049197925 | -8,229615086 | 1,88E-16 | 6,86E-14 | YES | 0 | FALSE |
| 52 | 8,128584484 | 0,123949971 | 3,015113606 | 0,041109552 | 0,967208559 | 0,998956236 | YES | 0 | FALSE |
| 53 | 19,94635241 | -0,300899008 | 3,014254842 | -0,099825338 | 0,920482992 | 0,998956236 | YES | 0 | FALSE |
| 54 | 87,97309925 | -0,205606441 | 2,969562448 | -0,069237958 | 0,944800209 | 0,998956236 | YES | 0 | FALSE |

| Medication | Ticker | taxLevel | Phylum | Family | Genus | Species |
|---|---|---|---|---|---|---|
| Dopamine agonists | 55 | ASV | Firmicutes | Streptococcaceae | Streptococcus | NA |
| Dopamine agonists | 56 | ASV | Firmicutes | NA | NA | NA |
| Dopamine agonists | 57 | ASV | Firmicutes | Ruminococcaceae | Ruminococcus | NA |
| Dopamine agonists | 58 | ASV | Firmicutes | Oscillospiraceae | UCG-005 | NA |
| Dopamine agonists | 59 | ASV | Firmicutes | Lachnospiraceae | [Eubacterium] ventriosum group | NA |
| Dopamine agonists | 60 | ASV | Firmicutes | Lachnospiraceae | Roseburia | NA |
| Dopamine agonists | 61 | ASV | Firmicutes | Peptostreptococcaceae | Romboutsia | NA |
| Dopamine agonists | 62 | ASV | Firmicutes | Ruminococcaceae | Ruminococcus | callidus |
| Dopamine agonists | 63 | ASV | Firmicutes | Oscillospiraceae | UCG-002 | NA |
| Dopamine agonists | 64 | ASV | Desulfobacterota | Desulfovibrionaceae | Desulfovibrio | NA |
| Dopamine agonists | 65 | ASV | Bacteroidota | Tannerellaceae | Parabacteroides | distasonis |
| Dopamine agonists | 66 | ASV | Firmicutes | Oscillospiraceae | Intestinimonas | NA |
| Dopamine agonists | 67 | ASV | Firmicutes | Lachnospiraceae | Lachnoclostridium | edouardi |
| Dopamine agonists | 68 | ASV | Actinobacteriota | Coriobacteriaceae | Collinsella | aerofaciens |
| Dopamine agonists | 69 | ASV | Firmicutes | Oscillospiraceae | NA | NA |
| Dopamine agonists | 70 | ASV | Proteobacteria | Pasteurellaceae | Haemophilus | NA |
| Dopamine agonists | 71 | ASV | Actinobacteriota | Coriobacteriales Incertae Sedis | NA | NA |
| Dopamine agonists | 72 | ASV | Firmicutes | NA | NA | NA |
| Dopamine agonists | 73 | ASV | Bacteroidota | Prevotellaceae | Paraprevotella | NA |
| Dopamine agonists | 74 | ASV | Firmicutes | NA | NA | NA |
| Dopamine agonists | 75 | ASV | Firmicutes | Lachnospiraceae | Dorea | longicatena |
| Dopamine agonists | 76 | ASV | Firmicutes | Ruminococcaceae | Ruminococcus | NA |
| Dopamine agonists | 77 | ASV | Firmicutes | Ruminococcaceae | Ruminococcus | NA |
| Dopamine agonists | 78 | ASV | Firmicutes | Lachnospiraceae | [Ruminococcus] torques group | NA |
| Dopamine agonists | 79 | ASV | Firmicutes | NA | NA | NA |
| Dopamine agonists | 80 | ASV | Firmicutes | Ruminococcaceae | Ruminococcus | champanellensis |
| Dopamine agonists | 81 | ASV | Firmicutes | Erysipelatoclostridiaceae | Erysipelotrichaceae UCG-003 | bacterium |
| Dopamine agonists | 82 | ASV | Firmicutes | Ruminococcaceae | UBA1819 | NA |
| Dopamine agonists | 83 | ASV | Firmicutes | Lachnospiraceae | Lachnospiraceae NK4A136 group | NA |
| Dopamine agonists | 84 | ASV | Bacteroidota | Marinifilaceae | Odoribacter | splanchnicus |

| Ticker | DESeq2_baseMean | DESeq2_log2-FoldChange | DESeq2_lfcSE | DESeq2_stat | DESeq2_pvalue | DESeq2_padj | DESeq2 converged | ANCOM_W | ANCOM_detected_0.7 |
|---|---|---|---|---|---|---|---|---|---|
| 55 | 50,75517682 | -0,534594369 | 2,992217867 | -0,178661579 | 0,858203439 | 0,998956236 | YES | 0 | FALSE |
| 56 | 47,83749254 | 5,657969646 | 2,97858407 | 1,899550093 | 0,057492187 | 0,998956236 | YES | 0 | FALSE |
| 57 | 27,20871536 | 0,160212155 | 3,013910272 | 0,053157573 | 0,95760636 | 0,998956236 | YES | 0 | FALSE |
| 58 | 17,46368906 | -0,036398486 | 2,633747502 | -0,013820036 | 0,988973558 | 0,998956236 | YES | 0 | FALSE |
| 59 | 83,97160382 | 0,031908471 | 2,241734348 | 0,014233833 | 0,988643428 | 0,998956236 | YES | 0 | FALSE |
| 60 | 73,58395473 | -0,0401245 | 2,313845087 | -0,017341049 | 0,986164538 | 0,998956236 | YES | 0 | FALSE |
| 61 | 44,10389525 | 2,360467132 | 1,936737805 | 1,218785075 | 0,222925778 | 0,998956236 | YES | 0 | FALSE |
| 62 | 28,8143168 | 0,294064648 | 3,036908869 | 0,096830251 | 0,9228612 | 0,998956236 | YES | 0 | FALSE |
| 63 | 18,62244929 | -1,957554369 | 2,960966652 | -0,661120032 | 0,508535339 | 0,998956236 | YES | 0 | FALSE |
| 64 | 33,66228937 | 0,225612311 | 3,036789806 | 0,074293028 | 0,940777224 | 0,998956236 | YES | 0 | FALSE |
| 65 | 6,199235025 | 0,131884378 | 3,036959194 | 0,043426457 | 0,965361588 | 0,998956236 | YES | 0 | FALSE |
| 66 | 12,56394146 | -0,627143659 | 2,993860668 | -0,209476568 | 0,834076225 | 0,998956236 | YES | 0 | FALSE |
| 67 | 22,34143269 | -0,219678896 | 2,60505864 | -0,084327812 | 0,9327958 | 0,998956236 | YES | 0 | FALSE |
| 68 | 502,9478624 | 0,133811581 | 1,387287365 | 0,096455561 | 0,923158767 | 0,998956236 | YES | 0 | FALSE |
| 69 | 38,81620558 | 0,763057707 | 2,992318103 | 0,255005544 | 0,798718817 | 0,998956236 | YES | 0 | FALSE |
| 70 | 37,11322796 | -0,370484239 | 2,986779817 | -0,124041363 | 0,901282525 | 0,998956236 | YES | 0 | FALSE |
| 71 | 7,097758825 | 0,036930944 | 3,036964635 | 0,012160479 | 0,990297581 | 0,998956236 | YES | 0 | FALSE |
| 72 | 3,986068578 | -0,181436108 | 3,019471674 | -0,060088693 | 0,952084995 | 0,998956236 | YES | 0 | FALSE |
| 73 | 15,47677946 | -0,194187647 | 3,036825026 | -0,063944299 | 0,949014579 | 0,998956236 | YES | 0 | FALSE |
| 74 | 7,515869006 | -0,363796496 | 3,036965542 | -0,119789471 | 0,904649923 | 0,998956236 | YES | 0 | FALSE |
| 75 | 577,6307374 | -0,275042609 | 1,572886808 | -0,174864846 | 0,861185833 | 0,998956236 | YES | 0 | FALSE |
| 76 | 4,520823462 | -0,363453224 | 3,037107784 | -0,119670835 | 0,904743905 | 0,998956236 | YES | 0 | FALSE |
| 77 | 187,8251628 | 6,324370184 | 3,014879283 | 2,097719209 | 0,035929957 | 0,998956236 | YES | 0 | FALSE |
| 78 | 63,31194315 | 0,077856477 | 3,03677698 | 0,025637865 | 0,979546184 | 0,998956236 | YES | 0 | FALSE |
| 79 | 3,088965164 | -0,319702228 | 3,037209662 | -0,105261824 | 0,916168055 | 0,998956236 | YES | 0 | FALSE |
| 80 | 267,4217133 | -1,13960007 | 2,969396826 | -0,383781669 | 0,701140276 | 0,998956236 | YES | 0 | FALSE |
| 81 | 143,7463084 | 0,166518416 | 2,211117657 | 0,075309613 | 0,939968373 | 0,998956236 | YES | 0 | FALSE |
| 82 | 28,72913582 | 0,10442026 | 2,440198837 | 0,042791701 | 0,965867579 | 0,998956236 | YES | 0 | FALSE |
| 83 | 76,22456119 | 1,581514784 | 2,970174878 | 0,532465208 | 0,594403833 | 0,998956236 | YES | 0 | FALSE |
| 84 | 22,93412291 | -0,812457663 | 1,567214225 | -0,518408811 | 0,604173069 | 0,998956236 | YES | 0 | FALSE |

| Medication | Ticker | taxLevel | Phylum | Family | Genus | Species |
|---|---|---|---|---|---|---|
| Dopamine agonists | 85 | ASV | Actinobacteriota | Eggerthellaceae | Adlercreutzia | equolifaciens |
| Dopamine agonists | 86 | ASV | Bacteroidota | Bacteroidaceae | Bacteroides | intestinalis |
| Dopamine agonists | 87 | ASV | Bacteroidota | Barnesiellaceae | Barnesiella | intestinihominis |
| Dopamine agonists | 88 | ASV | Firmicutes | Ruminococcaceae | Ruminococcus | NA |
| Dopamine agonists | 89 | ASV | Bacteroidota | Bacteroidaceae | Bacteroides | eggerthii |
| Dopamine agonists | 90 | ASV | Bacteroidota | Bacteroidaceae | Bacteroides | vulgatus |
| Dopamine agonists | 91 | ASV | Bacteroidota | Prevotellaceae | Prevotella | copri |
| Dopamine agonists | 92 | ASV | Firmicutes | Lachnospiraceae | [Eubacterium] xylanophilum group | NA |
| Dopamine agonists | 93 | ASV | Firmicutes | Oscillospiraceae | UCG-002 | NA |
| Dopamine agonists | 94 | ASV | Firmicutes | Lachnospiraceae | [Eubacterium] hallii group | NA |
| Dopamine agonists | 95 | ASV | Firmicutes | NA | NA | NA |
| Dopamine agonists | 96 | ASV | Firmicutes | NA | NA | NA |
| Dopamine agonists | 97 | ASV | Firmicutes | Oscillospiraceae | Intestinimonas | NA |
| Dopamine agonists | 98 | ASV | Bacteroidota | Prevotellaceae | Prevotella | NA |
| Dopamine agonists | 99 | ASV | Bacteroidota | Tannerellaceae | NA | NA |
| Dopamine agonists | 100 | ASV | Firmicutes | Ruminococcaceae | Faecalibacterium | prausnitzii |
| Dopamine agonists | 101 | ASV | Firmicutes | Lachnospiraceae | Lachnospiraceae FCS020 group | NA |
| Dopamine agonists | 102 | ASV | Firmicutes | NA | NA | NA |
| Dopamine agonists | 103 | ASV | Firmicutes | Lachnospiraceae | Lachnospiraceae UCG-010 | NA |
| Dopamine agonists | 104 | ASV | Firmicutes | Lachnospiraceae | Lachnospira | pectinoschiza |
| Dopamine agonists | 105 | ASV | Firmicutes | Peptococcaceae | Peptococcus | NA |
| Dopamine agonists | 106 | ASV | Firmicutes | Anaerovoracaceae | Family XIII AD3011 group | NA |
| Dopamine agonists | 107 | ASV | Firmicutes | Lachnospiraceae | Lachnospiraceae UCG-004 | NA |
| Dopamine agonists | 108 | ASV | Bacteroidota | Bacteroidaceae | Bacteroides | uniformis |
| Dopamine agonists | 109 | ASV | Firmicutes | Oscillospiraceae | Intestinimonas | NA |
| Dopamine agonists | 110 | ASV | Firmicutes | Oscillospiraceae | NK4A214 group | NA |
| Dopamine agonists | 111 | ASV | Bacteroidota | Barnesiellaceae | Barnesiella | NA |
| Dopamine agonists | 112 | ASV | Firmicutes | Lachnospiraceae | Lachnospiraceae UCG-001 | NA |
| Dopamine agonists | 113 | ASV | Firmicutes | Oscillospiraceae | Flavonifractor | plautii |
| Dopamine agonists | 114 | ASV | Firmicutes | Streptococcaceae | Streptococcus | NA |

| Ticker | DESeq2_baseMean | DESeq2_log2-FoldChange | DESeq2_lfcSE | DESeq2_stat | DESeq2_pvalue | DESeq2_padj | DESeq2 converged | ANCOM_W | ANCOM_detected_0.7 |
|---|---|---|---|---|---|---|---|---|---|
| 85 | 9,805297733 | 1,026779142 | 2,878913534 | 0,356655082 | 0,721350036 | 0,998956236 | YES | 0 | FALSE |
| 86 | 14,23534299 | 0,237199425 | 3,036830936 | 0,07810755 | 0,937742501 | 0,998956236 | YES | 0 | FALSE |
| 87 | 137,8979738 | 0,445218 | 3,0137046 | 0,147731135 | 0,882554959 | 0,998956236 | YES | 0 | FALSE |
| 88 | 35,77101564 | -0,364213917 | 3,036795652 | -0,119933627 | 0,904535726 | 0,998956236 | YES | 0 | FALSE |
| 89 | 214,0966625 | 0,003942395 | 3,013683334 | 0,001308165 | 0,998956236 | 0,998956236 | YES | 0 | FALSE |
| 90 | 1053,515881 | 0,194626494 | 2,1963282 | 0,088614486 | 0,929388295 | 0,998956236 | YES | 0 | FALSE |
| 91 | 781,5005243 | 0,434413898 | 3,017125505 | 0,143982707 | 0,885514125 | 0,998956236 | YES | 0 | FALSE |
| 92 | 48,2493914 | -1,57134968 | 2,244734767 | -0,700015745 | 0,483917472 | 0,998956236 | YES | 0 | FALSE |
| 93 | 108,2671864 | 0,363432996 | 2,992988999 | 0,12142811 | 0,903351953 | 0,998956236 | YES | 0 | FALSE |
| 94 | 0,553612992 | 0,589285482 | 3,039279844 | 0,19388984 | 0,846262139 | 0,998956236 | YES | 0 | FALSE |
| 95 | 0,295260262 | 0,583165467 | 3,041421392 | 0,191741095 | 0,847945014 | 0,998956236 | YES | 0 | FALSE |
| 96 | 3,27759701 | -0,36312842 | 3,037242968 | -0,119558568 | 0,904832842 | 0,998956236 | YES | 0 | FALSE |
| 97 | 65,44470241 | 0,557143388 | 2,59211663 | 0,214937623 | 0,829815951 | 0,998956236 | YES | 0 | FALSE |
| 98 | 18,95485928 | 0,114796882 | 3,036809093 | 0,03780181 | 0,969845701 | 0,998956236 | YES | 0 | FALSE |
| 99 | 2,091095852 | 0,152199889 | 3,037284751 | 0,05011051 | 0,960034324 | 0,998956236 | YES | 0 | FALSE |
| 100 | 460,9638656 | -1,516862267 | 1,853170312 | -0,818522862 | 0,413058693 | 0,998956236 | YES | 0 | FALSE |
| 101 | 27,74552205 | 0,392401807 | 1,844497877 | 0,212741805 | 0,831528358 | 0,998956236 | YES | 0 | FALSE |
| 102 | 40,74832547 | 0,667811205 | 3,013912032 | 0,22157621 | 0,824643804 | 0,998956236 | YES | 0 | FALSE |
| 103 | 31,64309664 | -0,371901589 | 2,999252956 | -0,123998074 | 0,9013168 | 0,998956236 | YES | 0 | FALSE |
| 104 | 68,96140702 | 1,187824945 | 2,192530238 | 0,541759892 | 0,587983925 | 0,998956236 | YES | 0 | FALSE |
| 105 | 58,56256001 | 0,178100316 | 3,036772744 | 0,058647891 | 0,953232565 | 0,998956236 | YES | 0 | FALSE |
| 106 | 6,803883342 | 1,285750429 | 3,000002919 | 0,428583059 | 0,668226676 | 0,998956236 | YES | 0 | FALSE |
| 107 | 30,95444734 | -0,068861679 | 2,991803509 | -0,023016779 | 0,981636889 | 0,998956236 | YES | 0 | FALSE |
| 108 | 729,6551081 | 0,135808234 | 1,482768782 | 0,091590972 | 0,927023025 | 0,998956236 | YES | 0 | FALSE |
| 109 | 44,37254422 | 0,991601527 | 1,905022428 | 0,520519608 | 0,602701465 | 0,998956236 | YES | 0 | FALSE |
| 110 | 41,9580284 | -0,276875259 | 2,99137943 | -0,09255772 | 0,926254934 | 0,998956236 | YES | 0 | FALSE |
| 111 | 13,68705374 | 0,085649053 | 3,036831394 | 0,028203427 | 0,977499904 | 0,998956236 | YES | 0 | FALSE |
| 112 | 24,13944265 | -1,073362205 | 2,971141525 | -0,361262564 | 0,717903176 | 0,998956236 | YES | 0 | FALSE |
| 113 | 73,16987435 | -1,432532625 | 2,088617225 | -0,685876094 | 0,492791246 | 0,998956236 | YES | 0 | FALSE |
| 114 | 655,5226161 | 0,930255324 | 1,451587611 | 0,640853722 | 0,521617725 | 0,998956236 | YES | 0 | FALSE |

| Medication | Ticker | taxLevel | Phylum | Family | Genus | Species |
|---|---|---|---|---|---|---|
| Dopamine agonists | 115 | ASV | Firmicutes | Ruminococcaceae | [Eubacterium] siraeum group | NA |
| Dopamine agonists | 116 | ASV | Proteobacteria | NA | NA | NA |
| Dopamine agonists | 117 | ASV | Firmicutes | Acholeplasmataceae | Anaeroplasma | NA |
| Dopamine agonists | 118 | ASV | Firmicutes | [Eubacterium] coprostanoligenes group | NA | NA |
| Dopamine agonists | 119 | ASV | Cyanobacteria | NA | NA | NA |
| Dopamine agonists | 120 | ASV | Bacteroidota | Bacteroidaceae | Bacteroides | clarus |
| Dopamine agonists | 121 | ASV | Firmicutes | Butyricicoccaceae | NA | NA |
| Dopamine agonists | 122 | ASV | Bacteroidota | Bacteroidaceae | Bacteroides | uniformis |
| Dopamine agonists | 123 | ASV | Bacteroidota | Prevotellaceae | Alloprevotella | NA |
| Dopamine agonists | 124 | ASV | Firmicutes | [Eubacterium] coprostanoligenes group | NA | NA |
| Dopamine agonists | 125 | ASV | Firmicutes | Lachnospiraceae | Tyzzerella | NA |
| Dopamine agonists | 126 | ASV | Firmicutes | NA | NA | NA |
| Dopamine agonists | 127 | ASV | Euryarchaeota | Methanobacteriaceae | Methanosphaera | NA |
| Dopamine agonists | 128 | ASV | Proteobacteria | NA | NA | NA |
| Dopamine agonists | 129 | ASV | Actinobacteriota | Bifidobacteriaceae | Bifidobacterium | NA |
| Dopamine agonists | 130 | ASV | Bacteroidota | Prevotellaceae | Paraprevotella | NA |
| Dopamine agonists | 131 | ASV | Firmicutes | Oscillospiraceae | NK4A214 group | NA |
| Dopamine agonists | 132 | ASV | Firmicutes | Erysipelotrichaceae | Solobacterium | NA |
| Dopamine agonists | 133 | ASV | Bacteroidota | Tannerellaceae | Parabacteroides | distasonis |
| Dopamine agonists | 134 | ASV | Firmicutes | Lachnospiraceae | Lachnospira | NA |
| Dopamine agonists | 135 | ASV | Actinobacteriota | Coriobacteriales Incertae Sedis | NA | NA |
| Dopamine agonists | 136 | ASV | Firmicutes | Lachnospiraceae | Blautia | NA |
| Dopamine agonists | 137 | ASV | Bacteroidota | Bacteroidaceae | Bacteroides | ovatus |
| Dopamine agonists | 138 | ASV | Firmicutes | Lachnospiraceae | Lachnospiraceae AC2044 group | NA |
| Dopamine agonists | 139 | ASV | Euryarchaeota | Methanobacteriaceae | Methanobrevibacter | NA |
| Dopamine agonists | 140 | ASV | Firmicutes | Lactobacillaceae | Lactobacillus | delbrueckii |
| Dopamine agonists | 141 | ASV | Firmicutes | Anaerovoracaceae | Family XIII UCG-001 | NA |
| Dopamine agonists | 142 | ASV | Firmicutes | Ruminococcaceae | Incertae Sedis | NA |
| Dopamine agonists | 143 | ASV | Firmicutes | Oscillospiraceae | Colidextribacter | NA |
| Dopamine agonists | 144 | ASV | Firmicutes | Lachnospiraceae | Lachnoclostridium | NA |

| Ticker | DESeq2_baseMean | DESeq2_log2-FoldChange | DESeq2_lfcSE | DESeq2_stat | DESeq2_pvalue | DESeq2_padj | DESeq2 converged | ANCOM_W | ANCOM_detected_0.7 |
|---|---|---|---|---|---|---|---|---|---|
| 115 | 20,2441453 | -2,079459716 | 2,969506322 | -0,700271187 | 0,483757962 | 0,998956236 | YES | 0 | FALSE |
| 116 | 4,980552695 | 0,205155028 | 3,015226201 | 0,068039681 | 0,945754047 | 0,998956236 | YES | 0 | FALSE |
| 117 | 58,26968806 | 0,300021255 | 3,036776318 | 0,098795968 | 0,92130027 | 0,998956236 | YES | 0 | FALSE |
| 118 | 1,988620903 | -0,329530502 | 3,037368111 | -0,108492119 | 0,913605332 | 0,998956236 | YES | 0 | FALSE |
| 119 | 10,34967123 | -0,343968632 | 2,971117627 | -0,115770789 | 0,907834202 | 0,998956236 | YES | 0 | FALSE |
| 120 | 110,0502031 | -0,080347087 | 3,013761015 | -0,026660073 | 0,978730859 | 0,998956236 | YES | 0 | FALSE |
| 121 | 6,050103979 | 0,67932252 | 3,015744264 | 0,225258663 | 0,821778056 | 0,998956236 | YES | 0 | FALSE |
| 122 | 208,0200067 | -0,074870666 | 2,222826187 | -0,033682646 | 0,973130218 | 0,998956236 | YES | 0 | FALSE |
| 123 | 118,5908202 | -0,24540152 | 3,014035617 | -0,081419582 | 0,935108277 | 0,998956236 | YES | 0 | FALSE |
| 124 | 0,452082346 | -0,355951537 | 3,040261861 | -0,117079236 | 0,906797264 | 0,998956236 | YES | 0 | FALSE |
| 125 | 14,16800105 | 1,105065337 | 3,014488717 | 0,366584665 | 0,713928848 | 0,998956236 | YES | 0 | FALSE |
| 126 | 2,634519437 | -0,62546276 | 3,039049317 | -0,20580869 | 0,836940353 | 0,998956236 | YES | 0 | FALSE |
| 127 | 5,878889909 | 0,41474847 | 3,015250766 | 0,137550241 | 0,890595884 | 0,998956236 | YES | 0 | FALSE |
| 128 | 34,79510218 | 0,866231951 | 3,010331199 | 0,287753039 | 0,773535787 | 0,998956236 | YES | 0 | FALSE |
| 129 | 597,1037513 | 0,921780082 | 1,355591995 | 0,679983421 | 0,496514959 | 0,998956236 | YES | 0 | FALSE |
| 130 | 42,08744841 | 0,096983506 | 2,606817213 | 0,0372038 | 0,970322509 | 0,998956236 | YES | 0 | FALSE |
| 131 | 13,88456112 | 0,016026603 | 3,014633264 | 0,00531627 | 0,995758251 | 0,998956236 | YES | 0 | FALSE |
| 132 | 4,407802875 | -0,363431293 | 3,037116929 | -0,119663253 | 0,904749911 | 0,998956236 | YES | 0 | FALSE |
| 133 | 27,91939325 | -0,827378683 | 3,015838201 | -0,27434452 | 0,783819875 | 0,998956236 | YES | 0 | FALSE |
| 134 | 1,299736745 | -0,361349021 | 3,037987369 | -0,118943556 | 0,905320074 | 0,998956236 | YES | 0 | FALSE |
| 135 | 14,05825884 | 0,702903935 | 3,014908642 | 0,233142698 | 0,8156506 | 0,998956236 | YES | 0 | FALSE |
| 136 | 2,208170287 | 0,604586766 | 3,037503165 | 0,199040703 | 0,842230905 | 0,998956236 | YES | 0 | FALSE |
| 137 | 50,21470141 | -1,082424138 | 3,01404853 | -0,359126314 | 0,719500597 | 0,998956236 | YES | 0 | FALSE |
| 138 | 5,116219409 | 0,1769436 | 3,015106578 | 0,058685687 | 0,95320246 | 0,998956236 | YES | 0 | FALSE |
| 139 | 78,94475221 | 0,25638986 | 1,659318688 | 0,15451514 | 0,877203574 | 0,998956236 | YES | 0 | FALSE |
| 140 | 48,50443725 | 0,167032232 | 3,013771514 | 0,055422991 | 0,95580148 | 0,998956236 | YES | 0 | FALSE |
| 141 | 18,70344525 | -0,513049667 | 2,427754253 | -0,211326853 | 0,83263223 | 0,998956236 | YES | 0 | FALSE |
| 142 | 74,61123849 | 0,648943596 | 2,45535688 | 0,264297057 | 0,791551028 | 0,998956236 | YES | 0 | FALSE |
| 143 | 20,01693115 | 0,100947687 | 2,974382469 | 0,033939041 | 0,972925761 | 0,998956236 | YES | 0 | FALSE |
| 144 | 5,935439758 | -0,612719104 | 3,015439389 | -0,203193971 | 0,838983425 | 0,998956236 | YES | 0 | FALSE |

| Medication | Ticker | taxLevel | Phylum | Family | Genus | Species |
|---|---|---|---|---|---|---|
| Dopamine agonists | 145 | ASV | Firmicutes | Ruminococcaceae | Incertae Sedis | NA |
| Dopamine agonists | 146 | ASV | Firmicutes | Ruminococcaceae | CAG-352 | NA |
| Dopamine agonists | 147 | ASV | Firmicutes | Christensenellaceae | Christensenellaceae R-7 group | NA |
| Dopamine agonists | 148 | ASV | Firmicutes | Erysipelotrichaceae | Catenisphaera | NA |
| Dopamine agonists | 149 | ASV | Firmicutes | Oscillospiraceae | NK4A214 group | NA |
| Dopamine agonists | 150 | ASV | Bacteroidota | Rikenellaceae | Alistipes | ihumii |
| Dopamine agonists | 151 | ASV | Firmicutes | Clostridiaceae | Clostridium sensu stricto 1 | NA |
| Dopamine agonists | 152 | ASV | Actinobacteriota | Eggerthellaceae | Slackia | isoflavoniconvertens |
| Dopamine agonists | 153 | ASV | Firmicutes | Oscillospiraceae | NK4A214 group | NA |
| Dopamine agonists | 154 | ASV | Firmicutes | Lachnospiraceae | NA | NA |
| Dopamine agonists | 155 | ASV | Firmicutes | Christensenellaceae | Christensenellaceae R-7 group | NA |
| Dopamine agonists | 156 | ASV | Firmicutes | Erysipelotrichaceae | NA | NA |
| Dopamine agonists | 157 | ASV | Firmicutes | Oscillospiraceae | UCG-002 | NA |
| Dopamine agonists | 158 | ASV | Firmicutes | Erysipelatoclostridiaceae | Erysipelatoclostridium | NA |
| Dopamine agonists | 159 | ASV | Firmicutes | Anaerovoracaceae | Mogibacterium | NA |
| Dopamine agonists | 160 | ASV | Firmicutes | Ruminococcaceae | Negativibacillus | NA |
| Dopamine agonists | 161 | ASV | Proteobacteria | Sutterellaceae | Sutterella | NA |
| Dopamine agonists | 162 | ASV | Firmicutes | Streptococcaceae | Lactococcus | NA |
| Dopamine agonists | 163 | ASV | Firmicutes | Ruminococcaceae | Subdoligranulum | NA |
| Dopamine agonists | 164 | ASV | Proteobacteria | Enterobacteriaceae | Escherichia-Shigella | NA |
| Dopamine agonists | 165 | ASV | Firmicutes | Lachnospiraceae | GCA-900066575 | NA |
| Dopamine agonists | 166 | ASV | Firmicutes | Lachnospiraceae | Lachnospiraceae UCG-004 | NA |
| Dopamine agonists | 167 | ASV | Firmicutes | Veillonellaceae | Veillonella | NA |
| Dopamine agonists | 168 | ASV | Firmicutes | Lachnospiraceae | Lachnoclostridium | NA |
| Dopamine agonists | 169 | ASV | Firmicutes | Christensenellaceae | NA | NA |
| Dopamine agonists | 170 | ASV | Firmicutes | Oscillospiraceae | Colidextribacter | massiliensis |
| Dopamine agonists | 171 | ASV | Bacteroidota | Rikenellaceae | Alistipes | putredinis |
| Dopamine agonists | 172 | ASV | Firmicutes | Lachnospiraceae | NA | NA |
| Dopamine agonists | 173 | ASV | Firmicutes | Ruminococcaceae | Subdoligranulum | NA |
| Dopamine agonists | 174 | ASV | Firmicutes | NA | NA | NA |

| Ticker | DESeq2_baseMean | DESeq2_log2-FoldChange | DESeq2_lfcSE | DESeq2_stat | DESeq2_pvalue | DESeq2_padj | DESeq2 converged | ANCOM_W | ANCOM_detected_0.7 |
|---|---|---|---|---|---|---|---|---|---|
| 145 | 29,97171447 | 1,432325983 | 2,546455459 | 0,562478318 | 0,573790172 | 0,998956236 | YES | 0 | FALSE |
| 146 | 194,2826734 | 2,321416452 | 3,014076432 | 0,770191634 | 0,441186226 | 0,998956236 | YES | 0 | FALSE |
| 147 | 24,43761099 | -0,078094819 | 1,756182935 | -0,044468499 | 0,964530962 | 0,998956236 | YES | 0 | FALSE |
| 148 | 6,715675586 | 0,109446922 | 3,036988068 | 0,036037982 | 0,971252073 | 0,998956236 | YES | 0 | FALSE |
| 149 | 141,6776611 | 0,196303518 | 3,01377367 | 0,065135455 | 0,948066152 | 0,998956236 | YES | 0 | FALSE |
| 150 | 3,374096179 | 0,359372513 | 3,01902911 | 0,119035789 | 0,905247002 | 0,998956236 | YES | 0 | FALSE |
| 151 | 41,82297384 | -0,289424041 | 2,465095247 | -0,117408867 | 0,906536058 | 0,998956236 | YES | 0 | FALSE |
| 152 | 4,32930549 | 0,535481086 | 2,997012997 | 0,178671593 | 0,858195576 | 0,998956236 | YES | 0 | FALSE |
| 153 | 3,415144542 | -1,023420205 | 3,017690111 | -0,339140259 | 0,734504072 | 0,998956236 | YES | 0 | FALSE |
| 154 | 5,191965365 | -0,191439735 | 3,015223956 | -0,06349105 | 0,949375486 | 0,998956236 | YES | 0 | FALSE |
| 155 | 7,240776119 | -0,096463867 | 3,014392475 | -0,032001097 | 0,974471176 | 0,998956236 | YES | 0 | FALSE |
| 156 | 15,26354989 | 0,840702686 | 2,434074135 | 0,345389105 | 0,729801867 | 0,998956236 | YES | 0 | FALSE |
| 157 | 411,8775246 | 0,415125739 | 2,159585842 | 0,192224699 | 0,847566199 | 0,998956236 | YES | 0 | FALSE |
| 158 | 11,29170044 | 0,695021695 | 2,977565204 | 0,233419471 | 0,815435695 | 0,998956236 | YES | 0 | FALSE |
| 159 | 6,908737034 | 0,328076498 | 3,036971616 | 0,108027516 | 0,913973866 | 0,998956236 | YES | 0 | FALSE |
| 160 | 71,07282003 | 1,357248537 | 1,852605542 | 0,732616041 | 0,463792645 | 0,998956236 | YES | 0 | FALSE |
| 161 | 11,4823769 | 0,266164653 | 3,036895411 | 0,087643668 | 0,930159894 | 0,998956236 | YES | 0 | FALSE |
| 162 | 11,46938317 | -0,047024763 | 3,036897587 | -0,015484474 | 0,987645671 | 0,998956236 | YES | 0 | FALSE |
| 163 | 515,8006642 | 1,69574736 | 2,179790899 | 0,77794038 | 0,436604162 | 0,998956236 | YES | 0 | FALSE |
| 164 | 133,001607 | -0,056361678 | 2,141176312 | -0,026322763 | 0,978999899 | 0,998956236 | YES | 0 | FALSE |
| 165 | 4,522088092 | -0,773603632 | 2,996152454 | -0,258199022 | 0,796253313 | 0,998956236 | YES | 0 | FALSE |
| 166 | 15,76402688 | -0,344205523 | 2,991875492 | -0,11504674 | 0,908408075 | 0,998956236 | YES | 0 | FALSE |
| 167 | 56,75108944 | 4,993181199 | 2,993798818 | 1,667841262 | 0,095347242 | 0,998956236 | YES | 0 | FALSE |
| 168 | 10,58221923 | 0,160754514 | 3,014329115 | 0,053330113 | 0,957468887 | 0,998956236 | YES | 0 | FALSE |
| 169 | 25,82786753 | -0,024108222 | 3,03681561 | -0,007938652 | 0,993665939 | 0,998956236 | YES | 0 | FALSE |
| 170 | 15,72089964 | -0,372978319 | 2,971599471 | -0,125514331 | 0,900116381 | 0,998956236 | YES | 0 | FALSE |
| 171 | 438,7820828 | 0,3606665 | 1,665414938 | 0,216562547 | 0,828549277 | 0,998956236 | YES | 0 | FALSE |
| 172 | 27,29458452 | -0,341442778 | 3,013892757 | -0,113289624 | 0,909800943 | 0,998956236 | YES | 0 | FALSE |
| 173 | 11,10269959 | 0,81686395 | 3,016553685 | 0,270793772 | 0,78654965 | 0,998956236 | YES | 0 | FALSE |
| 174 | 3,538269188 | -0,319826295 | 3,037151457 | -0,105304691 | 0,916134041 | 0,998956236 | YES | 0 | FALSE |

| Medication | Ticker | taxLevel | Phylum | Family | Genus | Species |
|---|---|---|---|---|---|---|
| Dopamine agonists | 175 | ASV | Firmicutes | Oscillospiraceae | UCG-002 | NA |
| Dopamine agonists | 176 | ASV | Firmicutes | Christensenellaceae | Christensenellaceae R-7 group | NA |
| Dopamine agonists | 177 | ASV | Firmicutes | Anaerovoracaceae | Family XIII AD3011 group | NA |
| Dopamine agonists | 178 | ASV | Proteobacteria | NA | NA | NA |
| Dopamine agonists | 179 | ASV | Proteobacteria | NA | NA | NA |
| Dopamine agonists | 180 | ASV | Firmicutes | Anaerovoracaceae | [Eubacterium] brachy group | NA |
| Dopamine agonists | 181 | ASV | Firmicutes | Lachnospiraceae | NA | NA |
| Dopamine agonists | 182 | ASV | Firmicutes | Peptostreptococcaceae | Intestinibacter | bartlettii |
| Dopamine agonists | 183 | ASV | Firmicutes | Lachnospiraceae | [Eubacterium] ventriosum group | NA |
| Dopamine agonists | 184 | ASV | Bacteroidota | Barnesiellaceae | NA | NA |
| Dopamine agonists | 185 | ASV | Firmicutes | Lachnospiraceae | Lachnoclostridium | NA |
| Dopamine agonists | 186 | ASV | Firmicutes | Lachnospiraceae | Lachnotalea | NA |
| Dopamine agonists | 187 | ASV | Firmicutes | Monoglobaceae | Monoglobus | pectinilyticus |
| Dopamine agonists | 188 | ASV | Firmicutes | Lachnospiraceae | Anaerostipes | hadrus |
| Dopamine agonists | 189 | ASV | Firmicutes | Christensenellaceae | Christensenellaceae R-7 group | NA |
| Dopamine agonists | 190 | ASV | Firmicutes | Lachnospiraceae | NA | NA |
| Dopamine agonists | 191 | ASV | Bacteroidota | Tannerellaceae | Parabacteroides | NA |
| Dopamine agonists | 192 | ASV | Firmicutes | Lachnospiraceae | Agathobacter | NA |
| Dopamine agonists | 193 | ASV | Firmicutes | Lachnospiraceae | Roseburia | NA |
| Dopamine agonists | 194 | ASV | Firmicutes | Lachnospiraceae | [Eubacterium] xylanophilum group | NA |
| Dopamine agonists | 195 | ASV | Firmicutes | NA | NA | NA |
| Dopamine agonists | 196 | ASV | Firmicutes | Lachnospiraceae | Marvinbryantia | NA |
| Dopamine agonists | 197 | ASV | Firmicutes | Lachnospiraceae | Blautia | obeum |
| Dopamine agonists | 198 | ASV | Firmicutes | Ruminococcaceae | NA | NA |
| Dopamine agonists | 199 | ASV | Bacteroidota | Rikenellaceae | Alistipes | NA |
| Dopamine agonists | 200 | ASV | Bacteroidota | Rikenellaceae | Alistipes | NA |
| Dopamine agonists | 201 | ASV | Firmicutes | Lachnospiraceae | [Eubacterium] hallii group | NA |
| Dopamine agonists | 202 | ASV | Firmicutes | UCG-010 | NA | NA |
| Dopamine agonists | 203 | ASV | Firmicutes | NA | NA | NA |
| Dopamine agonists | 204 | ASV | Bacteroidota | Barnesiellaceae | Coprobacter | fastidiosus |

| Ticker | DESeq2_baseMean | DESeq2_log2-FoldChange | DESeq2_lfcSE | DESeq2_stat | DESeq2_pvalue | DESeq2_padj | DESeq2 converged | ANCOM_W | ANCOM_detected_0.7 |
|---|---|---|---|---|---|---|---|---|---|
| 175 | 425,2658718 | 0,145765897 | 1,864591099 | 0,078175798 | 0,937688214 | 0,998956236 | YES | 0 | FALSE |
| 176 | 2,842283495 | -0,722481415 | 3,006267043 | -0,240325096 | 0,810078241 | 0,998956236 | YES | 0 | FALSE |
| 177 | 12,38549144 | -0,076257718 | 2,976822791 | -0,025617151 | 0,979562706 | 0,998956236 | YES | 0 | FALSE |
| 178 | 3,371579368 | 0,5195434 | 3,019660374 | 0,172053588 | 0,8633954 | 0,998956236 | YES | 0 | FALSE |
| 179 | 1,43990671 | -0,017796004 | 3,037851672 | -0,005858089 | 0,995325948 | 0,998956236 | YES | 0 | FALSE |
| 180 | 9,922552433 | 3,349569814 | 2,513752264 | 1,33249798 | 0,182696605 | 0,998956236 | YES | 2 | FALSE |
| 181 | 15,01808272 | -1,316797419 | 2,960794439 | -0,444744627 | 0,656504315 | 0,998956236 | YES | 0 | FALSE |
| 182 | 47,48497571 | -0,611244561 | 2,101967226 | -0,290796428 | 0,771207017 | 0,998956236 | YES | 0 | FALSE |
| 183 | 31,85861736 | -0,04753537 | 3,013833344 | -0,015772395 | 0,987415971 | 0,998956236 | YES | 0 | FALSE |
| 184 | 52,8323071 | 0,47480413 | 2,99250015 | 0,158664697 | 0,873933053 | 0,998956236 | YES | 0 | FALSE |
| 185 | 22,39241141 | -1,200682899 | 2,976554776 | -0,403380078 | 0,68666864 | 0,998956236 | YES | 0 | FALSE |
| 186 | 6,454170329 | -0,04367961 | 3,036922533 | -0,014382853 | 0,98852454 | 0,998956236 | YES | 0 | FALSE |
| 187 | 27,54735273 | 0,446669584 | 3,013920903 | 0,148202159 | 0,882183228 | 0,998956236 | YES | 0 | FALSE |
| 188 | 104,897279 | 0,738626635 | 1,688314836 | 0,437493422 | 0,661753545 | 0,998956236 | YES | 0 | FALSE |
| 189 | 25,10210339 | 0,698307393 | 2,053195995 | 0,340107518 | 0,733775561 | 0,998956236 | YES | 0 | FALSE |
| 190 | 11,21380822 | -0,471612529 | 3,014747127 | -0,156435187 | 0,875690004 | 0,998956236 | YES | 0 | FALSE |
| 191 | 2,322330941 | 0,458239627 | 3,037391504 | 0,150866171 | 0,880081286 | 0,998956236 | YES | 0 | FALSE |
| 192 | 512,9968805 | 1,749857104 | 2,326751978 | 0,752060005 | 0,452014974 | 0,998956236 | YES | 0 | FALSE |
| 193 | 19,79554004 | 0,516132042 | 3,036818022 | 0,169958173 | 0,865043031 | 0,998956236 | YES | 0 | FALSE |
| 194 | 22,80175238 | 0,235787379 | 3,036800664 | 0,077643351 | 0,938111757 | 0,998956236 | YES | 0 | FALSE |
| 195 | 15,00686138 | 0,85094459 | 3,014361067 | 0,282296835 | 0,777715912 | 0,998956236 | YES | 0 | FALSE |
| 196 | 24,70018565 | 1,200116707 | 2,992084485 | 0,401097199 | 0,688348563 | 0,998956236 | YES | 0 | FALSE |
| 197 | 134,6497628 | -0,546808091 | 1,940722841 | -0,281754859 | 0,778131486 | 0,998956236 | YES | 0 | FALSE |
| 198 | 27,99546171 | 0,200332576 | 1,737619591 | 0,115291389 | 0,908214164 | 0,998956236 | YES | 0 | FALSE |
| 199 | 19,71155851 | 0,879261188 | 2,972078228 | 0,295840527 | 0,767351872 | 0,998956236 | YES | 0 | FALSE |
| 200 | 3,48410036 | 1,257200448 | 3,016958018 | 0,416711283 | 0,6768896 | 0,998956236 | YES | 0 | FALSE |
| 201 | 61,88242008 | 0,097403744 | 1,825801339 | 0,05334849 | 0,957454246 | 0,998956236 | YES | 0 | FALSE |
| 202 | 1,469267625 | -0,361687232 | 3,037845509 | -0,119060443 | 0,90522747 | 0,998956236 | YES | 0 | FALSE |
| 203 | 8,193992525 | -0,363839197 | 3,036947757 | -0,119804233 | 0,904638229 | 0,998956236 | YES | 0 | FALSE |
| 204 | 4,848200515 | 0,436238225 | 3,016557385 | 0,144614595 | 0,885015173 | 0,998956236 | YES | 0 | FALSE |

| Medication | Ticker | taxLevel | Phylum | Family | Genus | Species |
|---|---|---|---|---|---|---|
| Dopamine agonists | 205 | ASV | Firmicutes | Clostridiaceae | Clostridium sensu stricto 1 | NA |
| Dopamine agonists | 206 | ASV | Firmicutes | NA | NA | NA |
| Dopamine agonists | 207 | ASV | Firmicutes | NA | NA | NA |
| Dopamine agonists | 208 | ASV | Firmicutes | Ruminococcaceae | DTU089 | NA |
| Dopamine agonists | 209 | ASV | Firmicutes | Lachnospiraceae | NA | NA |
| Dopamine agonists | 210 | ASV | Bacteroidota | Tannerellaceae | Parabacteroides | merdae |
| Dopamine agonists | 211 | ASV | Bacteroidota | Rikenellaceae | Alistipes | indistinctus |
| Dopamine agonists | 212 | ASV | Firmicutes | Ruminococcaceae | Ruminococcus | bromii |
| Dopamine agonists | 213 | ASV | Firmicutes | Erysipelotrichaceae | Holdemanella | NA |
| Dopamine agonists | 214 | ASV | Firmicutes | Oscillospiraceae | Colidextribacter | NA |
| Dopamine agonists | 215 | ASV | Firmicutes | NA | NA | NA |
| Dopamine agonists | 216 | ASV | Firmicutes | Ruminococcaceae | Ruminococcus | bicirculans |
| Dopamine agonists | 217 | ASV | Firmicutes | Oscillospiraceae | Oscillibacter | NA |
| Dopamine agonists | 218 | ASV | Firmicutes | Oscillospiraceae | UCG-003 | NA |
| Dopamine agonists | 219 | ASV | Firmicutes | Oscillospiraceae | Oscillospira | NA |
| Dopamine agonists | 220 | ASV | Firmicutes | UCG-010 | NA | NA |
| Dopamine agonists | 221 | ASV | Actinobacteriota | Bifidobacteriaceae | Bifidobacterium | NA |
| Dopamine agonists | 222 | ASV | Firmicutes | NA | NA | NA |
| Dopamine agonists | 223 | ASV | Firmicutes | Lachnospiraceae | Lachnospiraceae FCS020 group | bacterium |
| Dopamine agonists | 224 | ASV | Firmicutes | Christensenellaceae | Christensenellaceae R-7 group | NA |
| Dopamine agonists | 225 | ASV | Firmicutes | Oscillospiraceae | NK4A214 group | NA |
| Dopamine agonists | 226 | ASV | Firmicutes | Lactobacillaceae | Lactobacillus | NA |
| Dopamine agonists | 227 | ASV | Firmicutes | Lachnospiraceae | Blautia | faecis |
| Dopamine agonists | 228 | ASV | Bacteroidota | Marinifilaceae | Butyricimonas | virosa |
| Dopamine agonists | 229 | ASV | Firmicutes | UCG-010 | NA | NA |
| Dopamine agonists | 230 | ASV | Firmicutes | Christensenellaceae | Christensenellaceae R-7 group | NA |
| Dopamine agonists | 231 | ASV | Firmicutes | Christensenellaceae | Christensenellaceae R-7 group | NA |
| Dopamine agonists | 232 | ASV | Firmicutes | Christensenellaceae | Christensenellaceae R-7 group | NA |
| Dopamine agonists | 233 | ASV | Firmicutes | UCG-010 | NA | NA |
| Dopamine agonists | 234 | ASV | Bacteroidota | Bacteroidaceae | Bacteroides | NA |

| Ticker | DESeq2_baseMean | DESeq2_log2-FoldChange | DESeq2_lfcSE | DESeq2_stat | DESeq2_pvalue | DESeq2_padj | DESeq2 converged | ANCOM_W | ANCOM_detected_0.7 |
|---|---|---|---|---|---|---|---|---|---|
| 205 | 7,461734711 | 0,268684273 | 2,993201736 | 0,089764839 | 0,928474089 | 0,998956236 | YES | 0 | FALSE |
| 206 | 1,291763648 | 0,59342684 | 3,037845582 | 0,195344636 | 0,845123156 | 0,998956236 | YES | 0 | FALSE |
| 207 | 1,550116378 | 0,593952961 | 3,037664234 | 0,195529497 | 0,844978448 | 0,998956236 | YES | 0 | FALSE |
| 208 | 22,20999818 | 0,403123675 | 2,50310492 | 0,161049452 | 0,872054452 | 0,998956236 | YES | 0 | FALSE |
| 209 | 4,360669351 | -0,280853645 | 3,01921838 | -0,093021971 | 0,925886106 | 0,998956236 | YES | 0 | FALSE |
| 210 | 311,7696625 | -0,267460872 | 1,813828279 | -0,147456557 | 0,882771668 | 0,998956236 | YES | 0 | FALSE |
| 211 | 15,81276091 | 0,238232661 | 2,337592896 | 0,101913666 | 0,918825203 | 0,998956236 | YES | 0 | FALSE |
| 212 | 753,7235241 | -0,587934417 | 2,198361923 | -0,267442049 | 0,789128828 | 0,998956236 | YES | 0 | FALSE |
| 213 | 120,8395767 | -0,405369254 | 2,991338434 | -0,13551434 | 0,892205227 | 0,998956236 | YES | 0 | FALSE |
| 214 | 6,75389286 | -0,370897542 | 2,993296358 | -0,123909395 | 0,901387014 | 0,998956236 | YES | 0 | FALSE |
| 215 | 5,98221351 | 0,583579613 | 2,394275813 | 0,24373951 | 0,807432572 | 0,998956236 | YES | 0 | FALSE |
| 216 | 200,0878474 | 0,365111393 | 2,373917098 | 0,15380124 | 0,877766456 | 0,998956236 | YES | 0 | FALSE |
| 217 | 74,22539422 | 1,516920344 | 2,461186325 | 0,61633706 | 0,537672084 | 0,998956236 | YES | 0 | FALSE |
| 218 | 0,621613226 | -0,358186004 | 3,039317675 | -0,117850795 | 0,906185882 | 0,998956236 | YES | 0 | FALSE |
| 219 | 29,91155672 | -0,098789467 | 2,187971941 | -0,045151158 | 0,963986825 | 0,998956236 | YES | 0 | FALSE |
| 220 | 0,627428058 | 0,590128087 | 3,038986921 | 0,194185794 | 0,846030406 | 0,998956236 | YES | 0 | FALSE |
| 221 | 60,7130086 | 20,77874406 | 3,049198742 | 6,81449319 | 9,46E-12 | 1,73E-09 | YES | 0 | FALSE |
| 222 | 4,982516929 | 0,595784467 | 3,037035988 | 0,196173002 | 0,844474759 | 0,998956236 | YES | 0 | FALSE |
| 223 | 4,417389267 | -0,147221153 | 3,037104888 | -0,048474175 | 0,961338346 | 0,998956236 | YES | 0 | FALSE |
| 224 | 22,84262922 | 2,333194828 | 2,375350212 | 0,982252982 | 0,325975232 | 0,998956236 | YES | 0 | FALSE |
| 225 | 20,81801385 | -0,32261927 | 3,014300438 | -0,107029567 | 0,914765524 | 0,998956236 | YES | 0 | FALSE |
| 226 | 0,58311163 | 0,431189733 | 3,03966985 | 0,141854134 | 0,887195224 | 0,998956236 | YES | 0 | FALSE |
| 227 | 257,355953 | 0,336087043 | 1,561126491 | 0,215284953 | 0,82954516 | 0,998956236 | YES | 0 | FALSE |
| 228 | 13,37726803 | 0,95600481 | 2,315339184 | 0,412900544 | 0,679679482 | 0,998956236 | YES | 0 | FALSE |
| 229 | 8,287123424 | -0,322162266 | 3,014327394 | -0,106876999 | 0,914886561 | 0,998956236 | YES | 0 | FALSE |
| 230 | 1,354895711 | -0,617917811 | 3,041131661 | -0,2031868 | 0,83898903 | 0,998956236 | YES | 0 | FALSE |
| 231 | 1,154331945 | -0,110677989 | 3,023377631 | -0,036607398 | 0,970798045 | 0,998956236 | YES | 0 | FALSE |
| 232 | 2,316922024 | -0,36264087 | 3,037446409 | -0,119390047 | 0,904966346 | 0,998956236 | YES | 0 | FALSE |
| 233 | 21,27090817 | 0,295750806 | 3,013930558 | 0,098127943 | 0,921830699 | 0,998956236 | YES | 0 | FALSE |
| 234 | 246,6626698 | -1,016226252 | 2,391449851 | -0,424941486 | 0,670879331 | 0,998956236 | YES | 0 | FALSE |

| Medication | Ticker | taxLevel | Phylum | Family | Genus | Species |
| --- | --- | --- | --- | --- | --- | --- |
| Dopamine agonists | 235 | ASV | Firmicutes | [Eubacterium] coprostanoligenes group | NA | NA |
| Dopamine agonists | 236 | ASV | Firmicutes | Veillonellaceae | Veillonella | NA |
| Dopamine agonists | 237 | ASV | Firmicutes | Erysipelatoclostridiaceae | Coprobacillus | cateniformis |
| Dopamine agonists | 238 | ASV | Firmicutes | Oscillospiraceae | UCG-005 | NA |
| Dopamine agonists | 239 | ASV | Actinobacteriota | Atopobiaceae | Olsenella | NA |
| Dopamine agonists | 240 | ASV | Firmicutes | Lachnospiraceae | Coprococcus | comes |
| Dopamine agonists | 241 | ASV | Firmicutes | Oscillospiraceae | UCG-005 | NA |
| Dopamine agonists | 242 | ASV | Verrucomicrobiota | Victivallaceae | Victivallis | NA |
| Dopamine agonists | 243 | ASV | Actinobacteriota | Actinomycetaceae | Actinomyces | odontolyticus |
| Dopamine agonists | 244 | ASV | Firmicutes | Lachnospiraceae | NA | NA |
| Dopamine agonists | 245 | ASV | Bacteroidota | Bacteroidaceae | Bacteroides | NA |
| Dopamine agonists | 246 | ASV | Bacteroidota | Rikenellaceae | Alistipes | NA |
| Dopamine agonists | 247 | ASV | Firmicutes | Christensenellaceae | Christensenellaceae R-7 group | NA |
| Dopamine agonists | 248 | ASV | Firmicutes | Oscillospiraceae | Intestinimonas | massiliensis |
| Dopamine agonists | 249 | ASV | Bacteroidota | Bacteroidaceae | Bacteroides | caccae |
| Dopamine agonists | 250 | ASV | Firmicutes | UCG-010 | NA | NA |
| Dopamine agonists | 251 | ASV | Firmicutes | Lachnospiraceae | [Eubacterium] xylanophilum group | NA |
| Dopamine agonists | 252 | ASV | Firmicutes | Lachnospiraceae | Howardella | ureilytica |
| Dopamine agonists | 253 | ASV | Bacteroidota | Bacteroidaceae | Bacteroides | NA |
| Dopamine agonists | 254 | ASV | Firmicutes | Anaerovoracaceae | Family XIII AD3011 group | NA |
| Dopamine agonists | 255 | ASV | Firmicutes | Lachnospiraceae | Eisenbergiella | tayi |
| Dopamine agonists | 256 | ASV | Firmicutes | Lachnospiraceae | Blautia | NA |
| Dopamine agonists | 257 | ASV | Bacteroidota | Prevotellaceae | Prevotella | NA |
| Dopamine agonists | 258 | ASV | Firmicutes | Erysipelatoclostridiaceae | Erysipelatoclostridium | NA |
| Dopamine agonists | 259 | ASV | Bacteroidota | Bacteroidaceae | Bacteroides | nordii |
| Dopamine agonists | 260 | ASV | Firmicutes | Oscillospiraceae | Colidextribacter | NA |
| Dopamine agonists | 261 | ASV | Firmicutes | Ruminococcaceae | NA | NA |
| Dopamine agonists | 262 | ASV | Firmicutes | Lachnospiraceae | Marvinbryantia | NA |
| Dopamine agonists | 263 | ASV | Firmicutes | Oscillospiraceae | UCG-005 | NA |
| Dopamine agonists | 264 | ASV | Proteobacteria | Oxalobacteraceae | Oxalobacter | formigenes |

| Ticker | DESeq2_baseMean | DESeq2_log2-FoldChange | DESeq2_lfcSE | DESeq2_stat | DESeq2_pvalue | DESeq2_padj | DESeq2 converged | ANCOM_W | ANCOM_detected_0.7 |
|---|---|---|---|---|---|---|---|---|---|
| 235 | 9,544460345 | -0,597609835 | 3,015427297 | -0,19818413 | 0,842901004 | 0,998956236 | YES | 0 | FALSE |
| 236 | 5,086397915 | -1,201758062 | 3,01997716 | -0,397936143 | 0,690677257 | 0,998956236 | YES | 0 | FALSE |
| 237 | 2,123983293 | 0,560925595 | 3,037582909 | 0,184661822 | 0,853494293 | 0,998956236 | YES | 0 | FALSE |
| 238 | 3,470899527 | 0,561802867 | 3,037260927 | 0,184970235 | 0,853252382 | 0,998956236 | YES | 0 | FALSE |
| 239 | 1,551705206 | 0,204870684 | 3,022143627 | 0,067789857 | 0,945952918 | 0,998956236 | YES | 0 | FALSE |
| 240 | 71,6824924 | -0,310338484 | 2,352627189 | -0,131911459 | 0,895054325 | 0,998956236 | YES | 0 | FALSE |
| 241 | 28,07579217 | 0,611501516 | 2,991775008 | 0,204394219 | 0,838045447 | 0,998956236 | YES | 0 | FALSE |
| 242 | 0,439374009 | 0,220655035 | 3,024290543 | 0,072960925 | 0,941837212 | 0,998956236 | YES | 0 | FALSE |
| 243 | 1,467775284 | 0,117733461 | 3,017750181 | 0,039013654 | 0,968879503 | 0,998956236 | YES | 0 | FALSE |
| 244 | 2,447124746 | 0,608318824 | 3,037593565 | 0,200263403 | 0,841274583 | 0,998956236 | YES | 0 | FALSE |
| 245 | 159,5331272 | -0,372560067 | 1,626369572 | -0,229074667 | 0,818810883 | 0,998956236 | YES | 0 | FALSE |
| 246 | 232,3846727 | 0,509395076 | 2,028461042 | 0,251123914 | 0,801718308 | 0,998956236 | YES | 0 | FALSE |
| 247 | 14,74533947 | -0,633569829 | 2,992959361 | -0,211686746 | 0,832351429 | 0,998956236 | YES | 0 | FALSE |
| 248 | 21,72295123 | 0,226914683 | 3,01430726 | 0,075279215 | 0,939992559 | 0,998956236 | YES | 0 | FALSE |
| 249 | 446,0773252 | -0,429017463 | 1,648246574 | -0,260287186 | 0,794642256 | 0,998956236 | YES | 0 | FALSE |
| 250 | 1,921349971 | -0,36230004 | 3,037589036 | -0,119272237 | 0,905059678 | 0,998956236 | YES | 0 | FALSE |
| 251 | 10,75637349 | -0,761948777 | 2,993381804 | -0,254544467 | 0,799074955 | 0,998956236 | YES | 0 | FALSE |
| 252 | 5,909439436 | -0,21479177 | 2,997221263 | -0,071663635 | 0,942869597 | 0,998956236 | YES | 0 | FALSE |
| 253 | 782,0433039 | -0,134282372 | 2,532882551 | -0,053015633 | 0,957719452 | 0,998956236 | YES | 0 | FALSE |
| 254 | 1,404075075 | -0,318534431 | 3,037758101 | -0,104858392 | 0,916488175 | 0,998956236 | YES | 0 | FALSE |
| 255 | 3,931410209 | -0,319912492 | 3,037111422 | -0,10533446 | 0,91611042 | 0,998956236 | YES | 0 | FALSE |
| 256 | 886,1689657 | -0,610946956 | 1,199666394 | -0,509264041 | 0,610567159 | 0,998956236 | YES | 0 | FALSE |
| 257 | 0,777067058 | 0,557074686 | 3,039004796 | 0,183308262 | 0,854556152 | 0,998956236 | YES | 0 | FALSE |
| 258 | 0,473939308 | -0,478554398 | 3,040664373 | -0,157384814 | 0,874941581 | 0,998956236 | YES | 0 | FALSE |
| 259 | 3,435030008 | 1,190847298 | 2,998280977 | 0,397176685 | 0,691237174 | 0,998956236 | YES | 0 | FALSE |
| 260 | 42,29824705 | 0,136828508 | 3,036790481 | 0,045056947 | 0,964061918 | 0,998956236 | YES | 0 | FALSE |
| 261 | 4,01270662 | 0,907033304 | 2,998578033 | 0,302487811 | 0,762280224 | 0,998956236 | YES | 0 | FALSE |
| 262 | 5,305152038 | -0,117541834 | 3,015624197 | -0,038977614 | 0,968908237 | 0,998956236 | YES | 0 | FALSE |
| 263 | 112,3228646 | 0,119639024 | 2,176745114 | 0,054962349 | 0,95616846 | 0,998956236 | YES | 0 | FALSE |
| 264 | 9,924570874 | 1,479976301 | 2,285112186 | 0,647660237 | 0,517204727 | 0,998956236 | YES | 0 | FALSE |

| Medication | Ticker | taxLevel | Phylum | Family | Genus | Species |
|---|---|---|---|---|---|---|
| Dopamine agonists | 265 | ASV | Firmicutes | Lachnospiraceae | Blautia | NA |
| Dopamine agonists | 266 | ASV | Firmicutes | Lachnospiraceae | Coprococcus | eutactus |
| Dopamine agonists | 267 | ASV | Firmicutes | Lachnospiraceae | [Eubacterium] ventriosum group | NA |
| Dopamine agonists | 268 | ASV | Firmicutes | UCG-010 | NA | NA |
| Dopamine agonists | 269 | ASV | Firmicutes | Lachnospiraceae | NA | NA |
| Dopamine agonists | 270 | ASV | Firmicutes | Lachnospiraceae | [Ruminococcus] torques group | NA |
| Dopamine agonists | 271 | ASV | Firmicutes | Veillonellaceae | Allisonella | histaminiformans |
| Dopamine agonists | 272 | ASV | Firmicutes | Ruminococcaceae | Negativibacillus | massiliensis |
| Dopamine agonists | 273 | ASV | Firmicutes | Anaerovoracaceae | Family XIII UCG-001 | NA |
| Dopamine agonists | 274 | ASV | Firmicutes | Erysipelatoclostridiaceae | Asteroleplasma | NA |
| Dopamine agonists | 275 | ASV | Bacteroidota | Marinifilaceae | Butyricimonas | NA |
| Dopamine agonists | 276 | ASV | Bacteroidota | Marinifilaceae | Butyricimonas | NA |
| Dopamine agonists | 277 | ASV | Bacteroidota | Bacteroidaceae | Bacteroides | NA |
| Dopamine agonists | 278 | ASV | Proteobacteria | Sutterellaceae | Sutterella | NA |
| Dopamine agonists | 279 | ASV | Firmicutes | Lachnospiraceae | NA | NA |
| Dopamine agonists | 280 | ASV | Firmicutes | Monoglobaceae | Monoglobus | NA |
| Dopamine agonists | 281 | ASV | Firmicutes | Lachnospiraceae | NA | NA |
| Dopamine agonists | 282 | ASV | Firmicutes | Oscillospiraceae | UCG-005 | NA |
| Dopamine agonists | 283 | ASV | Bacteroidota | Bacteroidaceae | Bacteroides | massiliensis |
| Dopamine agonists | 284 | ASV | Firmicutes | Lachnospiraceae | Roseburia | inulinivorans |
| Dopamine agonists | 285 | ASV | Firmicutes | Erysipelatoclostridiaceae | Erysipelatoclostridium | NA |
| Dopamine agonists | 286 | ASV | Bacteroidota | Bacteroidaceae | Bacteroides | NA |
| Dopamine agonists | 287 | ASV | Firmicutes | Anaerovoracaceae | Family XIII AD3011 group | NA |
| Dopamine agonists | 288 | ASV | Firmicutes | [Eubacterium] coprostanoligenes group | NA | NA |
| Dopamine agonists | 289 | ASV | Firmicutes | Oscillospiraceae | Oscillibacter | NA |
| Dopamine agonists | 290 | ASV | Firmicutes | Ruminococcaceae | Faecalibacterium | NA |
| Dopamine agonists | 291 | ASV | Firmicutes | Ruminococcaceae | [Eubacterium] siraeum group | NA |
| Dopamine agonists | 292 | ASV | Firmicutes | Lachnospiraceae | NA | NA |
| Dopamine agonists | 293 | ASV | Firmicutes | Lachnospiraceae | NA | NA |
| Dopamine agonists | 294 | ASV | Firmicutes | Erysipelatoclostridiaceae | Catenibacterium | mitsuokai |

| Ticker | DESeq2_baseMean | DESeq2_log2-FoldChange | DESeq2_lfcSE | DESeq2_stat | DESeq2_pvalue | DESeq2_padj | DESeq2 converged | ANCOM_W | ANCOM_detected_0.7 |
|---|---|---|---|---|---|---|---|---|---|
| 265 | 1,886407998 | 0,299754965 | 3,037371933 | 0,098688923 | 0,921385264 | 0,998956236 | YES | 0 | FALSE |
| 266 | 11,21988997 | 0,596252379 | 3,036877372 | 0,196337325 | 0,844346149 | 0,998956236 | YES | 0 | FALSE |
| 267 | 10,30099633 | -1,231456247 | 3,015751238 | -0,408341455 | 0,683023009 | 0,998956236 | YES | 0 | FALSE |
| 268 | 4,105164263 | 1,019851862 | 2,99966336 | 0,339988772 | 0,733864984 | 0,998956236 | YES | 0 | FALSE |
| 269 | 9,484818942 | -0,714112128 | 3,018930783 | -0,236544717 | 0,813010012 | 0,998956236 | YES | 0 | FALSE |
| 270 | 403,5003596 | 0,063780107 | 1,560508502 | 0,040871362 | 0,967398448 | 0,998956236 | YES | 0 | FALSE |
| 271 | 7,070697629 | -0,274909222 | 2,992675791 | -0,091860676 | 0,926808735 | 0,998956236 | YES | 0 | FALSE |
| 272 | 23,55148509 | -0,136173653 | 2,992548003 | -0,04550425 | 0,963705387 | 0,998956236 | YES | 0 | FALSE |
| 273 | 2,020374352 | 0,560810196 | 3,037625357 | 0,184621252 | 0,853526116 | 0,998956236 | YES | 0 | FALSE |
| 274 | 0,751458363 | 0,542392051 | 3,020904495 | 0,179546243 | 0,857508811 | 0,998956236 | YES | 0 | FALSE |
| 275 | 1,235586066 | -0,318244357 | 3,03789476 | -0,10475819 | 0,916567687 | 0,998956236 | YES | 0 | FALSE |
| 276 | 10,16390106 | -0,38859302 | 3,015139876 | -0,128880595 | 0,897452132 | 0,998956236 | YES | 0 | FALSE |
| 277 | 286,1460699 | 0,087792808 | 2,182361313 | 0,040228356 | 0,967911071 | 0,998956236 | YES | 0 | FALSE |
| 278 | 53,05744211 | 0,75143586 | 3,013832666 | 0,249328992 | 0,803106307 | 0,998956236 | YES | 0 | FALSE |
| 279 | 2,599473491 | -0,36282109 | 3,037371018 | -0,119452345 | 0,904916993 | 0,998956236 | YES | 0 | FALSE |
| 280 | 117,3142266 | -0,994842613 | 1,570169331 | -0,63358938 | 0,526348833 | 0,998956236 | YES | 0 | FALSE |
| 281 | 0,775058189 | 0,591341204 | 3,038566349 | 0,194611911 | 0,845696778 | 0,998956236 | YES | 0 | FALSE |
| 282 | 8,378009947 | 0,59612535 | 3,036920387 | 0,196292716 | 0,844381063 | 0,998956236 | YES | 0 | FALSE |
| 283 | 143,7464759 | -0,57697544 | 2,991606629 | -0,192864742 | 0,847064899 | 0,998956236 | YES | 0 | FALSE |
| 284 | 165,9657635 | -1,589177919 | 2,213972762 | -0,717794702 | 0,47288388 | 0,998956236 | YES | 0 | FALSE |
| 285 | 35,23305081 | 0,361354751 | 2,422515639 | 0,149165085 | 0,88142337 | 0,998956236 | YES | 0 | FALSE |
| 286 | 151,3403235 | 0,039389012 | 2,596359054 | 0,015170865 | 0,987895866 | 0,998956236 | YES | 0 | FALSE |
| 287 | 0,520829284 | -0,325775661 | 3,039088743 | -0,107195179 | 0,91463414 | 0,998956236 | YES | 0 | FALSE |
| 288 | 49,13549751 | 0,492259901 | 3,013984853 | 0,163325274 | 0,870262335 | 0,998956236 | YES | 0 | FALSE |
| 289 | 2,316922024 | -0,36264087 | 3,037446409 | -0,119390047 | 0,904966346 | 0,998956236 | YES | 0 | FALSE |
| 290 | 977,2739625 | 1,610213496 | 1,569457286 | 1,025968346 | 0,304906505 | 0,998956236 | YES | 0 | FALSE |
| 291 | 356,4491153 | 1,838189332 | 1,656530203 | 1,10966243 | 0,267144518 | 0,998956236 | YES | 0 | FALSE |
| 292 | 1,791534384 | 0,604072113 | 3,037677047 | 0,198859886 | 0,842372349 | 0,998956236 | YES | 0 | FALSE |
| 293 | 7,442933781 | -0,099041049 | 3,037009408 | -0,032611374 | 0,9739845 | 0,998956236 | YES | 0 | FALSE |
| 294 | 75,00824213 | -0,389201892 | 3,014122797 | -0,12912609 | 0,897257878 | 0,998956236 | YES | 0 | FALSE |

| Medication | Ticker | taxLevel | Phylum | Family | Genus | Species |
|---|---|---|---|---|---|---|
| Dopamine agonists | 295 | ASV | Firmicutes | Lachnospiraceae | Lachnospiraceae UCG-010 | NA |
| Dopamine agonists | 296 | ASV | Firmicutes | Oscillospiraceae | Oscillibacter | NA |
| Dopamine agonists | 297 | ASV | Firmicutes | NA | NA | NA |
| Dopamine agonists | 298 | ASV | Firmicutes | NA | NA | NA |
| Dopamine agonists | 299 | ASV | Actinobacteriota | Eggerthellaceae | NA | NA |
| Dopamine agonists | 300 | ASV | Firmicutes | Ruminococcaceae | Angelakisella | NA |
| Dopamine agonists | 301 | ASV | Firmicutes | Oscillospiraceae | UCG-002 | NA |
| Dopamine agonists | 302 | ASV | Firmicutes | Ruminococcaceae | Incertae Sedis | NA |
| Dopamine agonists | 303 | ASV | Firmicutes | Oscillospiraceae | Flavonifractor | NA |
| Dopamine agonists | 304 | ASV | Firmicutes | Butyricicoccaceae | Butyricicoccus | NA |
| Dopamine agonists | 305 | ASV | Firmicutes | Lachnospiraceae | NA | NA |
| Dopamine agonists | 306 | ASV | Firmicutes | Erysipelotrichaceae | Holdemania | filiformis |
| Dopamine agonists | 307 | ASV | Bacteroidota | Prevotellaceae | NA | NA |
| Dopamine agonists | 308 | ASV | Firmicutes | Ruminococcaceae | Faecalibacterium | NA |
| Dopamine agonists | 309 | ASV | Firmicutes | Ruminococcaceae | DTU089 | NA |
| Dopamine agonists | 310 | ASV | Firmicutes | Christensenellaceae | NA | NA |
| Dopamine agonists | 311 | ASV | Firmicutes | Ruminococcaceae | Fournierella | NA |
| Dopamine agonists | 312 | ASV | Firmicutes | UCG-010 | NA | NA |
| Dopamine agonists | 313 | ASV | Firmicutes | Lachnospiraceae | Agathobacter | NA |
| Dopamine agonists | 314 | ASV | Firmicutes | Lachnospiraceae | Roseburia | intestinalis |
| Dopamine agonists | 315 | ASV | Firmicutes | Lachnospiraceae | Lachnoclostridium | NA |
| Dopamine agonists | 316 | ASV | Firmicutes | Hydrogenoanaerobacterium | NA | NA |
| Dopamine agonists | 317 | ASV | Firmicutes | Lachnospiraceae | NA | NA |
| Dopamine agonists | 318 | ASV | Firmicutes | [Eubacterium] coprostanoligenes group | NA | NA |
| Dopamine agonists | 319 | ASV | Firmicutes | Oscillospiraceae | Oscillibacter | NA |
| Dopamine agonists | 320 | ASV | Proteobacteria | Sutterellaceae | Parasutterella | excrementihominis |
| Dopamine agonists | 321 | ASV | Verrucomicrobiota | vadinBE97 | NA | NA |
| Dopamine agonists | 322 | ASV | Firmicutes | Ruminococcaceae | Angelakisella | NA |
| Dopamine agonists | 323 | ASV | Firmicutes | Oscillospiraceae | UCG-005 | NA |
| Dopamine agonists | 324 | ASV | Firmicutes | Ruminococcaceae | NA | NA |

| Ticker | DESeq2_baseMean | DESeq2_log2-FoldChange | DESeq2_lfcSE | DESeq2_stat | DESeq2_pvalue | DESeq2_padj | DESeq2 converged | ANCOM_W | ANCOM_detected_0.7 |
|---|---|---|---|---|---|---|---|---|---|
| 295 | 19,70269215 | -0,072878272 | 3,014029973 | -0,024179677 | 0,980709289 | 0,998956236 | YES | 0 | FALSE |
| 296 | 4,695124639 | -0,315880225 | 3,03712358 | -0,104006379 | 0,917164287 | 0,998956236 | YES | 0 | FALSE |
| 297 | 15,65657266 | -0,632539819 | 3,037144516 | -0,208267936 | 0,835019766 | 0,998956236 | YES | 0 | FALSE |
| 298 | 8,33456202 | -0,215402165 | 3,014653817 | -0,071451708 | 0,943038257 | 0,998956236 | YES | 0 | FALSE |
| 299 | 128,7041458 | 0,732593045 | 1,831265408 | 0,400047444 | 0,689121573 | 0,998956236 | YES | 0 | FALSE |
| 300 | 19,16140334 | 1,438859106 | 2,971901088 | 0,48415444 | 0,628276268 | 0,998956236 | YES | 0 | FALSE |
| 301 | 145,5328527 | 0,22801143 | 2,429457233 | 0,093852827 | 0,925226067 | 0,998956236 | YES | 0 | FALSE |
| 302 | 57,80571486 | 0,475083346 | 2,473480654 | 0,192070775 | 0,847686767 | 0,998956236 | YES | 0 | FALSE |
| 303 | 5,053294801 | -0,46860166 | 2,994488677 | -0,156488039 | 0,875648347 | 0,998956236 | YES | 0 | FALSE |
| 304 | 124,0117155 | 0,405537503 | 1,995203176 | 0,203256244 | 0,838934754 | 0,998956236 | YES | 0 | FALSE |
| 305 | 2,039270621 | 0,607820314 | 3,037760949 | 0,200088264 | 0,841411552 | 0,998956236 | YES | 0 | FALSE |
| 306 | 5,005656773 | 0,381752641 | 2,670394 | 0,142957422 | 0,886323808 | 0,998956236 | YES | 0 | FALSE |
| 307 | 50,04995218 | 0,408502901 | 3,036775284 | 0,134518647 | 0,892992467 | 0,998956236 | YES | 0 | FALSE |
| 308 | 37,65228701 | -0,873218178 | 2,991994398 | -0,291851542 | 0,770400135 | 0,998956236 | YES | 0 | FALSE |
| 309 | 18,05574172 | 1,129694345 | 2,994998565 | 0,377193618 | 0,706029723 | 0,998956236 | YES | 0 | FALSE |
| 310 | 105,3457468 | -0,318355543 | 3,014176296 | -0,105619417 | 0,915884319 | 0,998956236 | YES | 0 | FALSE |
| 311 | 0,332167795 | 0,584599546 | 3,040917306 | 0,192244473 | 0,847550711 | 0,998956236 | YES | 0 | FALSE |
| 312 | 1,529963724 | 0,290559706 | 3,039714591 | 0,095587825 | 0,923847935 | 0,998956236 | YES | 0 | FALSE |
| 313 | 2342,640596 | 0,310488251 | 1,669278006 | 0,186001523 | 0,852443567 | 0,998956236 | YES | 0 | FALSE |
| 314 | 50,19902288 | -1,12700161 | 2,991482566 | -0,376736814 | 0,706369202 | 0,998956236 | YES | 0 | FALSE |
| 315 | 37,13206487 | 1,131624176 | 2,970957329 | 0,380895466 | 0,703280817 | 0,998956236 | YES | 0 | FALSE |
| 316 | 3,395585132 | -0,86679226 | 3,016173943 | -0,28738139 | 0,773820309 | 0,998956236 | YES | 0 | FALSE |
| 317 | 0,777067058 | 0,557074686 | 3,039004796 | 0,183308262 | 0,854556152 | 0,998956236 | YES | 0 | FALSE |
| 318 | 48,60197185 | 0,181260307 | 2,992791523 | 0,060565631 | 0,951705146 | 0,998956236 | YES | 0 | FALSE |
| 319 | 1,958188745 | 0,604304023 | 3,03759867 | 0,198941364 | 0,842308612 | 0,998956236 | YES | 0 | FALSE |
| 320 | 101,9339438 | -0,103425258 | 2,604623787 | -0,039708329 | 0,968325661 | 0,998956236 | YES | 0 | FALSE |
| 321 | 0,291645132 | 0,590714805 | 3,042255894 | 0,194169993 | 0,846042778 | 0,998956236 | YES | 0 | FALSE |
| 322 | 6,234247042 | -0,156896008 | 3,037027961 | -0,051661035 | 0,958798785 | 0,998956236 | YES | 0 | FALSE |
| 323 | 131,7532683 | 0,162836017 | 2,970338306 | 0,054820697 | 0,956281311 | 0,998956236 | YES | 0 | FALSE |
| 324 | 2,795814763 | 1,221856511 | 3,018034047 | 0,404851798 | 0,685586451 | 0,998956236 | YES | 0 | FALSE |

| Medication | Ticker | taxLevel | Phylum | Family | Genus | Species |
|---|---|---|---|---|---|---|
| Dopamine agonists | 325 | ASV | Firmicutes | Lachnospiraceae | Coprococcus | catus |
| Dopamine agonists | 326 | ASV | Firmicutes | Lachnospiraceae | [Ruminococcus] gauvreauii group | NA |
| Dopamine agonists | 327 | ASV | Verrucomicrobiota | Puniceicoccaceae | NA | NA |
| Dopamine agonists | 328 | ASV | Firmicutes | Ruminococcaceae | NA | NA |
| Dopamine agonists | 329 | ASV | Firmicutes | NA | NA | NA |
| Dopamine agonists | 330 | ASV | Firmicutes | Lachnospiraceae | Lachnospiraceae ND3007 group | NA |
| Dopamine agonists | 331 | ASV | Firmicutes | Lachnospiraceae | [Ruminococcus] torques group | NA |
| Dopamine agonists | 332 | ASV | Firmicutes | Lachnospiraceae | [Eubacterium] eligens group | NA |
| Dopamine agonists | 333 | ASV | Firmicutes | Lachnospiraceae | [Eubacterium] ruminantium group | NA |
| Dopamine agonists | 334 | ASV | Firmicutes | Ruminococcaceae | Faecalibacterium | prausnitzii |
| Dopamine agonists | 335 | ASV | Firmicutes | Acidaminococcaceae | Phascolarctobacterium | faecium |
| Dopamine agonists | 336 | ASV | Firmicutes | Peptococcaceae | NA | NA |
| Dopamine agonists | 337 | ASV | Desulfobacterota | Desulfovibrionaceae | NA | NA |
| Dopamine agonists | 338 | ASV | Firmicutes | Lachnospiraceae | Lachnospiraceae NK4A136 group | NA |
| Dopamine agonists | 339 | ASV | Firmicutes | Ruminococcaceae | NA | NA |
| Dopamine agonists | 340 | ASV | Firmicutes | Oscillospiraceae | Colidextribacter | NA |
| Dopamine agonists | 341 | ASV | Firmicutes | Lachnospiraceae | GCA-900066575 | NA |
| Dopamine agonists | 342 | ASV | Firmicutes | Christensenellaceae | Christensenellaceae R-7 group | NA |
| Dopamine agonists | 343 | ASV | Firmicutes | Lachnospiraceae | Lachnospiraceae NK4A136 group | NA |
| Dopamine agonists | 344 | ASV | Verrucomicrobiota | Victivallaceae | Victivallis | vadensis |
| Dopamine agonists | 345 | ASV | Bacteroidota | Bacteroidaceae | Bacteroides | NA |
| Dopamine agonists | 346 | ASV | Firmicutes | Lachnospiraceae | Coprococcus | NA |
| Dopamine agonists | 347 | ASV | Firmicutes | Lachnospiraceae | [Eubacterium] hallii group | NA |
| Dopamine agonists | 348 | ASV | Firmicutes | NA | NA | NA |
| Dopamine agonists | 349 | ASV | Proteobacteria | Sutterellaceae | Sutterella | wadsworthensis |
| Dopamine agonists | 350 | ASV | Firmicutes | NA | NA | NA |
| Dopamine agonists | 351 | ASV | Firmicutes | Lachnospiraceae | Dorea | formicigenerans |
| Dopamine agonists | 352 | ASV | Firmicutes | Lachnospiraceae | Shuttleworthia | NA |
| Dopamine agonists | 353 | ASV | Firmicutes | Lachnospiraceae | Blautia | NA |
| Dopamine agonists | 354 | ASV | Desulfobacterota | Desulfovibrionaceae | NA | NA |

| Ticker | DESeq2_baseMean | DESeq2_log2-FoldChange | DESeq2_lfcSE | DESeq2_stat | DESeq2_pvalue | DESeq2_padj | DESeq2 converged | ANCOM_W | ANCOM_detected_0.7 |
|---|---|---|---|---|---|---|---|---|---|
| 325 | 28,27143501 | -0,749260149 | 2,184935932 | -0,342920878 | 0,731657985 | 0,998956236 | YES | 0 | FALSE |
| 326 | 23,15202526 | 0,503165289 | 2,972174455 | 0,169291977 | 0,865566986 | 0,998956236 | YES | 0 | FALSE |
| 327 | 1,666543613 | 0,603867336 | 3,037746043 | 0,198787959 | 0,842428615 | 0,998956236 | YES | 0 | FALSE |
| 328 | 2,316922024 | -0,36264087 | 3,037446409 | -0,119390047 | 0,904966346 | 0,998956236 | YES | 0 | FALSE |
| 329 | 2,995045544 | -0,363017488 | 3,037289296 | -0,119520221 | 0,904863221 | 0,998956236 | YES | 0 | FALSE |
| 330 | 27,0844115 | -1,219329825 | 2,163552681 | -0,563577599 | 0,573041636 | 0,998956236 | YES | 0 | FALSE |
| 331 | 250,8500642 | 0,353377382 | 2,465964779 | 0,143301877 | 0,886051773 | 0,998956236 | YES | 0 | FALSE |
| 332 | 89,34026653 | -0,296611406 | 2,356637645 | -0,125862118 | 0,899841069 | 0,998956236 | YES | 0 | FALSE |
| 333 | 13,80818368 | 0,475596531 | 3,036871129 | 0,156607413 | 0,87555426 | 0,998956236 | YES | 0 | FALSE |
| 334 | 605,778642 | -0,071665452 | 1,848487982 | -0,038769769 | 0,969073948 | 0,998956236 | YES | 0 | FALSE |
| 335 | 4,627415686 | -0,30060631 | 3,019849806 | -0,099543464 | 0,92070678 | 0,998956236 | YES | 0 | FALSE |
| 336 | 17,07699902 | -0,500642641 | 2,202816417 | -0,227273883 | 0,82021078 | 0,998956236 | YES | 0 | FALSE |
| 337 | 0,751315944 | 1,236215654 | 3,027566951 | 0,40831984 | 0,683038875 | 0,998956236 | YES | 0 | FALSE |
| 338 | 132,7178957 | 0,257976264 | 2,969298295 | 0,086881222 | 0,930765926 | 0,998956236 | YES | 0 | FALSE |
| 339 | 4,403870367 | 0,440303402 | 3,016353073 | 0,145972103 | 0,883943411 | 0,998956236 | YES | 0 | FALSE |
| 340 | 7,484426045 | -0,010708584 | 2,992845893 | -0,003578061 | 0,997145127 | 0,998956236 | YES | 0 | FALSE |
| 341 | 28,66619448 | 1,444019265 | 2,261778609 | 0,638444125 | 0,523184615 | 0,998956236 | YES | 0 | FALSE |
| 342 | 0,33906176 | -0,353260815 | 3,041402619 | -0,116150625 | 0,907533167 | 0,998956236 | YES | 0 | FALSE |
| 343 | 82,44934701 | 1,154873711 | 2,594609136 | 0,445105081 | 0,656243817 | 0,998956236 | YES | 0 | FALSE |
| 344 | 0,516255622 | -0,870656745 | 3,030816075 | -0,28726809 | 0,773907054 | 0,998956236 | YES | 0 | FALSE |
| 345 | 110,7148008 | -0,643126073 | 2,969662505 | -0,216565375 | 0,828547072 | 0,998956236 | YES | 0 | FALSE |
| 346 | 333,6328387 | 0,599371175 | 2,991205587 | 0,200377793 | 0,841185126 | 0,998956236 | YES | 0 | FALSE |
| 347 | 166,9880699 | 0,750718824 | 1,788444245 | 0,419760821 | 0,674660189 | 0,998956236 | YES | 0 | FALSE |
| 348 | 1,116911288 | -1,02239481 | 3,026900435 | -0,337769554 | 0,735536855 | 0,998956236 | YES | 0 | FALSE |
| 349 | 25,69401646 | 0,923665921 | 2,438481821 | 0,37878729 | 0,704845826 | 0,998956236 | YES | 0 | FALSE |
| 350 | 1,473218583 | 1,254714885 | 3,023899247 | 0,414932768 | 0,678191121 | 0,998956236 | YES | 0 | FALSE |
| 351 | 89,42595153 | 0,381291456 | 2,23537084 | 0,170571902 | 0,864560393 | 0,998956236 | YES | 0 | FALSE |
| 352 | 9,595750395 | -0,00430364 | 2,65133722 | -0,001623196 | 0,998704878 | 0,998956236 | YES | 0 | FALSE |
| 353 | 30,38632518 | 7,939342651 | 3,015718778 | 2,632653518 | 0,008472073 | 0,998956236 | YES | 0 | FALSE |
| 354 | 1,932859215 | 0,463053824 | 3,037522714 | 0,152444563 | 0,878836311 | 0,998956236 | YES | 0 | FALSE |

| Medication | Ticker | taxLevel | Phylum | Family | Genus | Species |
|---|---|---|---|---|---|---|
| Dopamine agonists | 355 | ASV | Bacteroidota | Barnesiellaceae | Coprobacter | secundus |
| Dopamine agonists | 356 | ASV | Bacteroidota | Bacteroidaceae | Bacteroides | NA |
| Dopamine agonists | 357 | ASV | Firmicutes | Lachnospiraceae | [Eubacterium] eligens group | NA |
| Dopamine agonists | 358 | ASV | Firmicutes | Erysipelotrichaceae | Turicibacter | sanguinis |
| Dopamine agonists | 359 | ASV | Firmicutes | Oscillospiraceae | NK4A214 group | NA |
| Dopamine agonists | 360 | ASV | Firmicutes | UCG-010 | NA | NA |
| Dopamine agonists | 361 | ASV | Bacteroidota | Rikenellaceae | Alistipes | shahii |
| Dopamine agonists | 362 | ASV | Firmicutes | Lachnospiraceae | CAG-56 | NA |
| Dopamine agonists | 363 | ASV | Firmicutes | Lachnospiraceae | [Ruminococcus] gnavus group | NA |
| Dopamine agonists | 364 | ASV | Firmicutes | Acidaminococcaceae | Phascolarctobacterium | succinatutens |
| Dopamine agonists | 365 | ASV | Bacteroidota | Bacteroidaceae | Bacteroides | plebeius |
| Dopamine agonists | 366 | Genus | Actinobacteriota | Bifidobacteriaceae | Bifidobacterium | NA |
| Dopamine agonists | 367 | Genus | Verrucomicrobiota | Akkermansiaceae | Akkermansia | NA |
| Dopamine agonists | 368 | Genus | Firmicutes | Lachnospiraceae | Lachnospira | NA |
| Dopamine agonists | 369 | Genus | Firmicutes | Oscillospiraceae | UCG-003 | NA |
| Dopamine agonists | 370 | Genus | Firmicutes | Veillonellaceae | Dialister | NA |
| Dopamine agonists | 371 | Genus | Firmicutes | Oscillospiraceae | NK4A214 group | NA |
| Dopamine agonists | 372 | Genus | Firmicutes | Lachnospiraceae | Fusicatenibacter | NA |
| Dopamine agonists | 373 | Genus | Firmicutes | Ruminococcaceae | Candidatus Soleaferrea | NA |
| Dopamine agonists | 374 | Genus | Actinobacteriota | Eggerthellaceae | Senegalimassilia | NA |
| Dopamine agonists | 375 | Genus | Actinobacteriota | Eggerthellaceae | Enterorhabdus | NA |
| Dopamine agonists | 376 | Genus | Firmicutes | Lachnospiraceae | Lachnoclostridium | NA |
| Dopamine agonists | 377 | Genus | Firmicutes | Peptostreptococcaceae | Terrisporobacter | NA |
| Dopamine agonists | 378 | Genus | Desulfobacterota | Desulfovibrionaceae | Bilophila | NA |
| Dopamine agonists | 379 | Genus | Firmicutes | Oscillospiraceae | Oscillibacter | NA |
| Dopamine agonists | 380 | Genus | Firmicutes | Christensenellaceae | Christensenellaceae R-7 group | NA |
| Dopamine agonists | 381 | Genus | Firmicutes | Erysipelatoclostridiaceae | UCG-004 | NA |
| Dopamine agonists | 382 | Genus | Firmicutes | Lachnospiraceae | [Eubacterium] ventriosum group | NA |
| Dopamine agonists | 383 | Genus | Firmicutes | Peptostreptococcaceae | Romboutsia | NA |
| Dopamine agonists | 384 | Genus | Desulfobacterota | Desulfovibrionaceae | Desulfovibrio | NA |

| Ticker | DESeq2_baseMean | DESeq2_log2-FoldChange | DESeq2_lfcSE | DESeq2_stat | DESeq2_pvalue | DESeq2_padj | DESeq2 converged | ANCOM_W | ANCOM_detected_0.7 |
|---|---|---|---|---|---|---|---|---|---|
| 355 | 4,352557783 | 1,246854368 | 2,997700795 | 0,415936897 | 0,677456179 | 0,998956236 | YES | 0 | FALSE |
| 356 | 1397,971376 | -0,908328279 | 1,621284588 | -0,560252213 | 0,575307417 | 0,998956236 | YES | 0 | FALSE |
| 357 | 53,93957092 | -1,588741214 | 2,570871281 | -0,617977736 | 0,536590015 | 0,998956236 | YES | 0 | FALSE |
| 358 | 7,09932009 | 0,722030004 | 2,959136086 | 0,24400027 | 0,807230611 | 0,998956236 | YES | 0 | FALSE |
| 359 | 55,39903495 | 0,679900371 | 2,431490076 | 0,279622926 | 0,779766816 | 0,998956236 | YES | 0 | FALSE |
| 360 | 0,282551466 | -0,351136301 | 3,042304969 | -0,115417851 | 0,908113931 | 0,998956236 | YES | 0 | FALSE |
| 361 | 82,21928101 | 0,1444773 | 2,479584097 | 0,058266747 | 0,953536154 | 0,998956236 | YES | 0 | FALSE |
| 362 | 79,77909046 | -2,56531161 | 2,974400497 | -0,862463415 | 0,388432557 | 0,998956236 | YES | 0 | FALSE |
| 363 | 121,7579298 | -1,058772632 | 2,996645974 | -0,353319225 | 0,723849135 | 0,998956236 | YES | 0 | FALSE |
| 364 | 39,74120698 | -0,184732255 | 3,013802559 | -0,061295407 | 0,951123948 | 0,998956236 | YES | 0 | FALSE |
| 365 | 81,97677146 | -2,834053266 | 3,014363162 | -0,940183088 | 0,347123655 | 0,998956236 | YES | 0 | FALSE |
| 366 | 1054,994047 | 1,200398916 | 0,569836244 | 2,106568209 | 0,035155025 | 0,850751601 | YES | 0 | FALSE |
| 367 | 861,3548571 | -0,388914813 | 1,263309424 | -0,307853964 | 0,758193457 | 0,998881056 | YES | 0 | FALSE |
| 368 | 133,9734081 | -0,922240808 | 1,061351414 | -0,868930682 | 0,384885046 | 0,998881056 | YES | 0 | FALSE |
| 369 | 183,3027975 | -0,406996253 | 1,371181005 | -0,296821683 | 0,766602651 | 0,998881056 | YES | 0 | FALSE |
| 370 | 1196,04593 | 0,883512973 | 1,979677205 | 0,446291431 | 0,655386745 | 0,998881056 | YES | 0 | FALSE |
| 371 | 384,9475 | 0,244622007 | 0,841904466 | 0,290557916 | 0,77138945 | 0,998881056 | YES | 0 | FALSE |
| 372 | 711,149347 | 1,446176965 | 1,298208785 | 1,113978723 | 0,265288315 | 0,998881056 | YES | 0 | FALSE |
| 373 | 1,072416593 | -0,621915154 | 3,008435118 | -0,206723805 | 0,836225565 | 0,998881056 | YES | 0 | FALSE |
| 374 | 6,290512682 | 1,032320575 | 2,981942702 | 0,346190614 | 0,729199468 | 0,998881056 | YES | 0 | FALSE |
| 375 | 9,34864251 | -1,317784784 | 1,790909486 | -0,735818752 | 0,461841008 | 0,998881056 | YES | 0 | FALSE |
| 376 | 215,256091 | -0,597897219 | 0,510389178 | -1,171453559 | 0,241416515 | 0,998881056 | YES | 0 | FALSE |
| 377 | 6,873461442 | -1,326610154 | 3,019266679 | -0,439381577 | 0,660385074 | 0,998881056 | YES | 0 | FALSE |
| 378 | 65,01195307 | -0,066665474 | 1,11193935 | -0,059954235 | 0,952192084 | 0,998881056 | YES | 0 | FALSE |
| 379 | 267,8988818 | 0,539299051 | 0,687018296 | 0,784984992 | 0,432462368 | 0,998881056 | YES | 0 | FALSE |
| 380 | 306,551642 | 0,003418093 | 1,117501822 | 0,003058691 | 0,997559521 | 0,998881056 | YES | 0 | FALSE |
| 381 | 0,407412491 | 0,12512018 | 3,03526886 | 0,041222108 | 0,967118829 | 0,998881056 | YES | 0 | FALSE |
| 382 | 251,7753773 | -1,330897512 | 0,915711729 | -1,453402278 | 0,146112095 | 0,998881056 | YES | 0 | FALSE |
| 383 | 45,56525839 | 2,510647686 | 1,368158412 | 1,83505628 | 0,066497351 | 0,998881056 | YES | 0 | FALSE |
| 384 | 72,4324127 | 0,365429371 | 2,035950079 | 0,179488375 | 0,857554244 | 0,998881056 | YES | 0 | FALSE |

| Medication | Ticker | taxLevel | Phylum | Family | Genus | Species |
|---|---|---|---|---|---|---|
| Dopamine agonists | 385 | Genus | Firmicutes | Ruminococcaceae | Paludicola | NA |
| Dopamine agonists | 386 | Genus | Firmicutes | Oscillospiraceae | Intestinimonas | NA |
| Dopamine agonists | 387 | Genus | Actinobacteriota | Coriobacteriaceae | Collinsella | NA |
| Dopamine agonists | 388 | Genus | Proteobacteria | Pasteurellaceae | Haemophilus | NA |
| Dopamine agonists | 389 | Genus | Firmicutes | Erysipelatoclostridiaceae | Asteroleplasma | NA |
| Dopamine agonists | 390 | Genus | Firmicutes | Lachnospiraceae | Dorea | NA |
| Dopamine agonists | 391 | Genus | Firmicutes | Erysipelatoclostridiaceae | Erysipelotrichaceae UCG-003 | NA |
| Dopamine agonists | 392 | Genus | Firmicutes | Ruminococcaceae | UBA1819 | NA |
| Dopamine agonists | 393 | Genus | Bacteroidota | Marinifilaceae | Odoribacter | NA |
| Dopamine agonists | 394 | Genus | Actinobacteriota | Eggerthellaceae | Adlercreutzia | NA |
| Dopamine agonists | 395 | Genus | Bacteroidota | Barnesiellaceae | Barnesiella | NA |
| Dopamine agonists | 396 | Genus | Bacteroidota | Prevotellaceae | Prevotellaceae NK3B31 group | NA |
| Dopamine agonists | 397 | Genus | Bacteroidota | Bacteroidaceae | Bacteroides | NA |
| Dopamine agonists | 398 | Genus | Bacteroidota | Prevotellaceae | Prevotella | NA |
| Dopamine agonists | 399 | Genus | Firmicutes | Lachnospiraceae | [Eubacterium] xylanophilum group | NA |
| Dopamine agonists | 400 | Genus | Firmicutes | Lachnospiraceae | Lachnospiraceae FCS020 group | NA |
| Dopamine agonists | 401 | Genus | Firmicutes | Lachnospiraceae | Lachnospiraceae UCG-010 | NA |
| Dopamine agonists | 402 | Genus | Firmicutes | Peptococcaceae | Peptococcus | NA |
| Dopamine agonists | 403 | Genus | Firmicutes | Lachnospiraceae | Lachnospiraceae UCG-004 | NA |
| Dopamine agonists | 404 | Genus | Firmicutes | Lachnospiraceae | Lachnospiraceae UCG-001 | NA |
| Dopamine agonists | 405 | Genus | Firmicutes | Oscillospiraceae | Flavonifractor | NA |
| Dopamine agonists | 406 | Genus | Firmicutes | Streptococcaceae | Streptococcus | NA |
| Dopamine agonists | 407 | Genus | Firmicutes | Acholeplasmataceae | Anaeroplasma | NA |
| Dopamine agonists | 408 | Genus | Bacteroidota | Prevotellaceae | Alloprevotella | NA |
| Dopamine agonists | 409 | Genus | Firmicutes | Lachnospiraceae | Tyzzerella | NA |
| Dopamine agonists | 410 | Genus | Euryarchaeota | Methanobacteriaceae | Methanosphaera | NA |
| Dopamine agonists | 411 | Genus | Firmicutes | Erysipelotrichaceae | Solobacterium | NA |
| Dopamine agonists | 412 | Genus | Firmicutes | Lachnospiraceae | Lachnospiraceae AC2044 group | NA |
| Dopamine agonists | 413 | Genus | Euryarchaeota | Methanobacteriaceae | Methanobrevibacter | NA |
| Dopamine agonists | 414 | Genus | Firmicutes | Ruminococcaceae | CAG-352 | NA |

| Ticker | DESeq2_baseMean | DESeq2_log2-FoldChange | DESeq2_lfcSE | DESeq2_stat | DESeq2_pvalue | DESeq2_padj | DESeq2 converged | ANCOM_W | ANCOM_detected_0.7 |
|---|---|---|---|---|---|---|---|---|---|
| 385 | 4,924808752 | -0,033093868 | 2,975634166 | -0,011121618 | 0,991126415 | 0,998881056 | YES | 0 | FALSE |
| 386 | 177,2590394 | -0,011872391 | 0,657579716 | -0,01805468 | 0,985595232 | 0,998881056 | YES | 0 | FALSE |
| 387 | 438,0900912 | 0,051951997 | 0,737162771 | 0,070475611 | 0,943815112 | 0,998881056 | YES | 0 | FALSE |
| 388 | 40,05658099 | -0,460576816 | 2,781066562 | -0,165611576 | 0,868462637 | 0,998881056 | YES | 0 | FALSE |
| 389 | 26,50145184 | 0,405436061 | 3,016899348 | 0,134388329 | 0,89309551 | 0,998881056 | YES | 0 | FALSE |
| 390 | 758,8763265 | -0,285780975 | 0,464501781 | -0,615241935 | 0,538394957 | 0,998881056 | YES | 0 | FALSE |
| 391 | 133,7131249 | 0,171179839 | 1,672131468 | 0,102372237 | 0,91846122 | 0,998881056 | YES | 0 | FALSE |
| 392 | 24,55110178 | -0,06914917 | 1,918682328 | -0,036039927 | 0,971250523 | 0,998881056 | YES | 0 | FALSE |
| 393 | 22,25984237 | -0,822744843 | 0,852458513 | -0,965143559 | 0,334472912 | 0,998881056 | YES | 0 | FALSE |
| 394 | 8,913992309 | 0,702452417 | 2,733641076 | 0,256965855 | 0,797205132 | 0,998881056 | YES | 0 | FALSE |
| 395 | 227,6580989 | 0,393216023 | 0,685829927 | 0,573343343 | 0,566412244 | 0,998881056 | YES | 0 | FALSE |
| 396 | 127,0218344 | -0,056565826 | 3,036761356 | -0,018627024 | 0,985138645 | 0,998881056 | YES | 0 | FALSE |
| 397 | 7247,818069 | -0,351714224 | 0,384050522 | -0,91580197 | 0,35977077 | 0,998881056 | YES | 0 | FALSE |
| 398 | 1202,055699 | -1,107770608 | 1,344940355 | -0,823657795 | 0,410134027 | 0,998881056 | YES | 0 | FALSE |
| 399 | 100,5747759 | -1,402873141 | 0,899979526 | -1,558783394 | 0,119047656 | 0,998881056 | YES | 0 | FALSE |
| 400 | 32,84736195 | 0,520519533 | 0,659680458 | 0,789047979 | 0,430083965 | 0,998881056 | YES | 0 | FALSE |
| 401 | 83,90158914 | 0,263848059 | 0,994583583 | 0,265284953 | 0,790789955 | 0,998881056 | YES | 0 | FALSE |
| 402 | 63,6478755 | -0,30455504 | 1,852300749 | -0,164419865 | 0,869400626 | 0,998881056 | YES | 0 | FALSE |
| 403 | 67,12358874 | 0,544404032 | 1,798660911 | 0,302671854 | 0,76213995 | 0,998881056 | YES | 0 | FALSE |
| 404 | 35,98088275 | -1,416372534 | 2,693534133 | -0,525841688 | 0,598998207 | 0,998881056 | YES | 0 | FALSE |
| 405 | 108,9075689 | 0,607169483 | 1,197004705 | 0,507240682 | 0,611985954 | 0,998881056 | YES | 0 | FALSE |
| 406 | 723,5098522 | 0,715407334 | 0,876218088 | 0,816471771 | 0,414230363 | 0,998881056 | YES | 0 | FALSE |
| 407 | 59,87875326 | 0,213830991 | 3,036779062 | 0,070413747 | 0,94386435 | 0,998881056 | YES | 0 | FALSE |
| 408 | 190,3388284 | -0,33056552 | 3,014025163 | -0,109675767 | 0,91266652 | 0,998881056 | YES | 0 | FALSE |
| 409 | 11,54643079 | 1,049462505 | 3,014591405 | 0,348127611 | 0,727744349 | 0,998881056 | YES | 0 | FALSE |
| 410 | 6,87489613 | 0,319741022 | 3,01525615 | 0,106041081 | 0,915549758 | 0,998881056 | YES | 0 | FALSE |
| 411 | 13,59437992 | -0,746989898 | 3,014107796 | -0,247831182 | 0,804265028 | 0,998881056 | YES | 0 | FALSE |
| 412 | 7,831182968 | 0,16640074 | 3,01494573 | 0,055191952 | 0,95598554 | 0,998881056 | YES | 0 | FALSE |
| 413 | 84,42452394 | -0,106700456 | 1,077161075 | -0,099057104 | 0,921092931 | 0,998881056 | YES | 0 | FALSE |
| 414 | 183,1689752 | 12,70769294 | 3,015397296 | 4,2142682 | 2,51E-05 | 0,003032131 | YES | 0 | FALSE |

| Medication | Ticker | taxLevel | Phylum | Family | Genus | Species |
|---|---|---|---|---|---|---|
| Dopamine agonists | 415 | Genus | Firmicutes | Clostridiaceae | Clostridium sensu stricto 1 | NA |
| Dopamine agonists | 416 | Genus | Actinobacteriota | Eggerthellaceae | Slackia | NA |
| Dopamine agonists | 417 | Genus | Firmicutes | Oscillospiraceae | UCG-002 | NA |
| Dopamine agonists | 418 | Genus | Firmicutes | Erysipelotrichaceae | Catenisphaera | NA |
| Dopamine agonists | 419 | Genus | Firmicutes | Anaerovoracaceae | Mogibacterium | NA |
| Dopamine agonists | 420 | Genus | Firmicutes | Ruminococcaceae | Negativibacillus | NA |
| Dopamine agonists | 421 | Genus | Firmicutes | Streptococcaceae | Lactococcus | NA |
| Dopamine agonists | 422 | Genus | Firmicutes | Ruminococcaceae | Subdoligranulum | NA |
| Dopamine agonists | 423 | Genus | Proteobacteria | Enterobacteriaceae | Escherichia-Shigella | NA |
| Dopamine agonists | 424 | Genus | Firmicutes | Lachnospiraceae | GCA-900066575 | NA |
| Dopamine agonists | 425 | Genus | Firmicutes | Veillonellaceae | Veillonella | NA |
| Dopamine agonists | 426 | Genus | Firmicutes | Oscillospiraceae | Colidextribacter | NA |
| Dopamine agonists | 427 | Genus | Bacteroidota | Rikenellaceae | Alistipes | NA |
| Dopamine agonists | 428 | Genus | Firmicutes | Anaerovoracaceae | Family XIII UCG-001 | NA |
| Dopamine agonists | 429 | Genus | Firmicutes | Anaerovoracaceae | Family XIII AD3011 group | NA |
| Dopamine agonists | 430 | Genus | Firmicutes | Anaerovoracaceae | [Eubacterium] brachy group | NA |
| Dopamine agonists | 431 | Genus | Firmicutes | Peptostreptococcaceae | Intestinibacter | NA |
| Dopamine agonists | 432 | Genus | Firmicutes | Lachnospiraceae | Lachnotalea | NA |
| Dopamine agonists | 433 | Genus | Firmicutes | Lachnospiraceae | Anaerostipes | NA |
| Dopamine agonists | 434 | Genus | Firmicutes | Lachnospiraceae | Marvinbryantia | NA |
| Dopamine agonists | 435 | Genus | Firmicutes | Lachnospiraceae | [Eubacterium] hallii group | NA |
| Dopamine agonists | 436 | Genus | Bacteroidota | Barnesiellaceae | Coprobacter | NA |
| Dopamine agonists | 437 | Genus | Firmicutes | Ruminococcaceae | DTU089 | NA |
| Dopamine agonists | 438 | Genus | Bacteroidota | Tannerellaceae | Parabacteroides | NA |
| Dopamine agonists | 439 | Genus | Firmicutes | Ruminococcaceae | Ruminococcus | NA |
| Dopamine agonists | 440 | Genus | Firmicutes | Erysipelotrichaceae | Holdemanella | NA |
| Dopamine agonists | 441 | Genus | Bacteroidota | Prevotellaceae | Paraprevotella | NA |
| Dopamine agonists | 442 | Genus | Firmicutes | Lactobacillaceae | Lactobacillus | NA |
| Dopamine agonists | 443 | Genus | Firmicutes | Oscillospiraceae | Oscillospira | NA |
| Dopamine agonists | 444 | Genus | Firmicutes | Erysipelatoclostridiaceae | Coprobacillus | NA |

| Ticker | DESeq2_baseMean | DESeq2_log2-FoldChange | DESeq2_lfcSE | DESeq2_stat | DESeq2_pvalue | DESeq2_padj | DESeq2 converged | ANCOM_W | ANCOM_detected_0.7 |
|---|---|---|---|---|---|---|---|---|---|
| 415 | 56,51096338 | -0,823840161 | 1,035771526 | -0,795387921 | 0,426387884 | 0,998881056 | YES | 0 | FALSE |
| 416 | 3,66075168 | 0,504888441 | 2,998349405 | 0,168388794 | 0,866277423 | 0,998881056 | YES | 0 | FALSE |
| 417 | 1408,342623 | 0,15689328 | 0,816924077 | 0,192053686 | 0,847700153 | 0,998881056 | YES | 0 | FALSE |
| 418 | 8,934247857 | 0,580158002 | 3,014507977 | 0,192455288 | 0,847385588 | 0,998881056 | YES | 0 | FALSE |
| 419 | 7,003045608 | 0,245488307 | 3,036998369 | 0,080832545 | 0,935575126 | 0,998881056 | YES | 0 | FALSE |
| 420 | 92,12266094 | 2,350263171 | 0,830334804 | 2,830500613 | 0,004647522 | 0,281175094 | YES | 1 | FALSE |
| 421 | 9,575547846 | -0,134559543 | 3,036897234 | -0,044308231 | 0,964658711 | 0,998881056 | YES | 0 | FALSE |
| 422 | 1065,856619 | 0,648951886 | 0,531065623 | 1,221980595 | 0,221714965 | 0,998881056 | YES | 0 | FALSE |
| 423 | 116,2531639 | -0,112897163 | 1,675357904 | -0,067386893 | 0,946273704 | 0,998881056 | YES | 0 | FALSE |
| 424 | 29,27683927 | 0,384147092 | 1,280326301 | 0,300038429 | 0,764147843 | 0,998881056 | YES | 0 | FALSE |
| 425 | 67,34805537 | 0,222636202 | 2,992180018 | 0,074406019 | 0,94068732 | 0,998881056 | YES | 0 | FALSE |
| 426 | 207,9260896 | 0,732536115 | 0,556478817 | 1,316377359 | 0,188047418 | 0,998881056 | YES | 0 | FALSE |
| 427 | 1042,659174 | 0,59816083 | 0,631188003 | 0,947674587 | 0,343295142 | 0,998881056 | YES | 0 | FALSE |
| 428 | 19,59074966 | 0,002492819 | 1,777551508 | 0,001402389 | 0,998881056 | 0,998881056 | YES | 0 | FALSE |
| 429 | 53,55976405 | 0,968659241 | 0,651318176 | 1,487228942 | 0,136954355 | 0,998881056 | YES | 0 | FALSE |
| 430 | 9,443006777 | 2,12562051 | 1,867682262 | 1,138106066 | 0,255076196 | 0,998881056 | YES | 0 | FALSE |
| 431 | 46,54954651 | -0,773966021 | 1,407987023 | -0,549696843 | 0,582527323 | 0,998881056 | YES | 0 | FALSE |
| 432 | 10,50194988 | -0,131369756 | 3,036922889 | -0,043257521 | 0,965496253 | 0,998881056 | YES | 0 | FALSE |
| 433 | 166,5056402 | 0,467517658 | 0,659326023 | 0,709084189 | 0,478272234 | 0,998881056 | YES | 0 | FALSE |
| 434 | 33,45446218 | 5,188519683 | 2,073120203 | 2,502758728 | 0,012322952 | 0,440428816 | YES | 0 | FALSE |
| 435 | 236,0654733 | 0,422324523 | 0,239524896 | 1,763175892 | 0,077870829 | 0,998881056 | YES | 0 | FALSE |
| 436 | 7,703336528 | 1,213364459 | 2,997990208 | 0,404725958 | 0,685678959 | 0,998881056 | YES | 0 | FALSE |
| 437 | 34,94914423 | 1,495250273 | 0,813095072 | 1,838961181 | 0,065920896 | 0,998881056 | YES | 0 | FALSE |
| 438 | 662,4121459 | 0,381256852 | 0,909003145 | 0,419423029 | 0,674906997 | 0,998881056 | YES | 0 | FALSE |
| 439 | 1506,084107 | -0,161013239 | 0,856080555 | -0,188081878 | 0,850812467 | 0,998881056 | YES | 0 | FALSE |
| 440 | 147,8981584 | -0,383675029 | 2,99128592 | -0,128264245 | 0,89793986 | 0,998881056 | YES | 0 | FALSE |
| 441 | 76,89042138 | -0,616187654 | 1,191686746 | -0,517071836 | 0,605106013 | 0,998881056 | YES | 0 | FALSE |
| 442 | 92,84598104 | 2,772431542 | 1,366264975 | 2,029204871 | 0,042437428 | 0,855821459 | YES | 0 | FALSE |
| 443 | 33,72458445 | -0,298256339 | 0,736681131 | -0,404864909 | 0,685576813 | 0,998881056 | YES | 0 | FALSE |
| 444 | 1,947881622 | 0,542650238 | 3,037825089 | 0,178631166 | 0,858227321 | 0,998881056 | YES | 0 | FALSE |

| Medication | Ticker | taxLevel | Phylum | Family | Genus | Species |
|---|---|---|---|---|---|---|
| Dopamine agonists | 445 | Genus | Actinobacteriota | Atopobiaceae | Olsenella | NA |
| Dopamine agonists | 446 | Genus | Verrucomicrobiota | Victivallaceae | Victivallis | NA |
| Dopamine agonists | 447 | Genus | Actinobacteriota | Actinomycetaceae | Actinomyces | NA |
| Dopamine agonists | 448 | Genus | Firmicutes | Lachnospiraceae | Howardella | NA |
| Dopamine agonists | 449 | Genus | Firmicutes | Lachnospiraceae | Eisenbergiella | NA |
| Dopamine agonists | 450 | Genus | Firmicutes | Lachnospiraceae | Blautia | NA |
| Dopamine agonists | 451 | Genus | Firmicutes | Oscillospiraceae | UCG-005 | NA |
| Dopamine agonists | 452 | Genus | Proteobacteria | Oxalobacteraceae | Oxalobacter | NA |
| Dopamine agonists | 453 | Genus | Firmicutes | Lachnospiraceae | [Ruminococcus] torques group | NA |
| Dopamine agonists | 454 | Genus | Firmicutes | Veillonellaceae | Allisonella | NA |
| Dopamine agonists | 455 | Genus | Bacteroidota | Marinifilaceae | Butyricimonas | NA |
| Dopamine agonists | 456 | Genus | Firmicutes | Monoglobaceae | Monoglobus | NA |
| Dopamine agonists | 457 | Genus | Bacteroidota | Rikenellaceae | Rikenellaceae RC9 gut group | NA |
| Dopamine agonists | 458 | Genus | Firmicutes | Lachnospiraceae | Roseburia | NA |
| Dopamine agonists | 459 | Genus | Firmicutes | Erysipelatoclostridiaceae | Erysipelatoclostridium | NA |
| Dopamine agonists | 460 | Genus | Firmicutes | Ruminococcaceae | [Eubacterium] siraeum group | NA |
| Dopamine agonists | 461 | Genus | Firmicutes | Erysipelatoclostridiaceae | Catenibacterium | NA |
| Dopamine agonists | 462 | Genus | Firmicutes | Ruminococcaceae | Angelakisella | NA |
| Dopamine agonists | 463 | Genus | Firmicutes | Ruminococcaceae | Incertae Sedis | NA |
| Dopamine agonists | 464 | Genus | Firmicutes | Butyricicoccaceae | Butyricicoccus | NA |
| Dopamine agonists | 465 | Genus | Firmicutes | Erysipelotrichaceae | Holdemania | NA |
| Dopamine agonists | 466 | Genus | Firmicutes | Ruminococcaceae | Fournierella | NA |
| Dopamine agonists | 467 | Genus | Firmicutes | Lachnospiraceae | Agathobacter | NA |
| Dopamine agonists | 468 | Genus | Proteobacteria | Sutterellaceae | Parasutterella | NA |
| Dopamine agonists | 469 | Genus | Firmicutes | Ruminococcaceae | Anaerotruncus | NA |
| Dopamine agonists | 470 | Genus | Firmicutes | Lachnospiraceae | [Ruminococcus] gauvreauii group | NA |
| Dopamine agonists | 471 | Genus | Firmicutes | Lachnospiraceae | Lachnospiraceae ND3007 group | NA |
| Dopamine agonists | 472 | Genus | Firmicutes | Defluviitaleaceae | Defluviitaleaceae UCG-011 | NA |
| Dopamine agonists | 473 | Genus | Firmicutes | Lachnospiraceae | [Eubacterium] eligens group | NA |
| Dopamine agonists | 474 | Genus | Firmicutes | Lachnospiraceae | [Eubacterium] ruminantium group | NA |

| Ticker | DESeq2_baseMean | DESeq2_log2-FoldChange | DESeq2_lfcSE | DESeq2_stat | DESeq2_pvalue | DESeq2_padj | DESeq2 converged | ANCOM_W | ANCOM_detected_0.7 |
|---|---|---|---|---|---|---|---|---|---|
| 445 | 1,366993497 | 0,085376546 | 3,02203937 | 0,028251302 | 0,977461721 | 0,998881056 | YES | 0 | FALSE |
| 446 | 1,446560666 | -1,209363477 | 2,989316234 | -0,404561907 | 0,685799564 | 0,998881056 | YES | 0 | FALSE |
| 447 | 1,21009094 | 0,03126212 | 3,017759656 | 0,01035938 | 0,991734558 | 0,998881056 | YES | 0 | FALSE |
| 448 | 5,48928889 | -0,232819475 | 2,996845946 | -0,077688169 | 0,938076105 | 0,998881056 | YES | 0 | FALSE |
| 449 | 26,77826566 | -0,25043012 | 2,991653957 | -0,083709588 | 0,933287334 | 0,998881056 | YES | 0 | FALSE |
| 450 | 1879,779624 | -0,490519641 | 0,549381567 | -0,892857843 | 0,371933305 | 0,998881056 | YES | 0 | FALSE |
| 451 | 408,1887532 | 1,346825933 | 1,121369966 | 1,201054045 | 0,229730238 | 0,998881056 | YES | 0 | FALSE |
| 452 | 10,23479096 | 1,37828668 | 1,77614419 | 0,775999318 | 0,437749389 | 0,998881056 | YES | 0 | FALSE |
| 453 | 675,4032774 | -0,763304601 | 0,57416103 | -1,329426 | 0,183707465 | 0,998881056 | YES | 0 | FALSE |
| 454 | 9,31522785 | -0,301346357 | 2,992561185 | -0,100698478 | 0,91978982 | 0,998881056 | YES | 0 | FALSE |
| 455 | 26,30575282 | -0,404879954 | 0,572127439 | -0,707674422 | 0,479147465 | 0,998881056 | YES | 0 | FALSE |
| 456 | 225,7522721 | 0,301498949 | 0,632539685 | 0,476648273 | 0,633612603 | 0,998881056 | YES | 0 | FALSE |
| 457 | 102,0607186 | -2,898973619 | 3,013846987 | -0,961884804 | 0,336107473 | 0,998881056 | YES | 0 | FALSE |
| 458 | 546,2324732 | -0,20541556 | 1,118265184 | -0,183691277 | 0,854255652 | 0,998881056 | YES | 0 | FALSE |
| 459 | 75,75037649 | -0,398804576 | 0,942216261 | -0,423262252 | 0,672103932 | 0,998881056 | YES | 0 | FALSE |
| 460 | 486,2965926 | 1,016544963 | 0,871100483 | 1,166966364 | 0,243223954 | 0,998881056 | YES | 0 | FALSE |
| 461 | 64,06289774 | -0,4049521 | 3,014044021 | -0,134355072 | 0,893121807 | 0,998881056 | YES | 0 | FALSE |
| 462 | 24,46009323 | 2,541807552 | 1,700931891 | 1,494361746 | 0,135081094 | 0,998881056 | YES | 0 | FALSE |
| 463 | 206,4588675 | 0,605746574 | 0,557641433 | 1,086265363 | 0,277361607 | 0,998881056 | YES | 0 | FALSE |
| 464 | 193,8532185 | 0,10303907 | 0,359089583 | 0,286945306 | 0,774154199 | 0,998881056 | YES | 0 | FALSE |
| 465 | 5,55987889 | 0,223569081 | 1,141016615 | 0,195938497 | 0,844658305 | 0,998881056 | YES | 0 | FALSE |
| 466 | 1,672806929 | 0,447940329 | 3,004510885 | 0,149089268 | 0,881483194 | 0,998881056 | YES | 0 | FALSE |
| 467 | 2890,414456 | 0,007432346 | 0,444043974 | 0,01673786 | 0,986645743 | 0,998881056 | YES | 0 | FALSE |
| 468 | 117,353991 | 0,301537698 | 1,135533534 | 0,265547154 | 0,79058799 | 0,998881056 | YES | 0 | FALSE |
| 469 | 2,34582764 | -0,095690175 | 3,016210831 | -0,031725294 | 0,974691124 | 0,998881056 | YES | 0 | FALSE |
| 470 | 27,28353221 | 0,516072458 | 2,951723481 | 0,174837671 | 0,861207187 | 0,998881056 | YES | 0 | FALSE |
| 471 | 26,88123588 | -1,359122777 | 1,634365337 | -0,831590555 | 0,405640095 | 0,998881056 | YES | 0 | FALSE |
| 472 | 0,534033279 | 0,141026526 | 3,024914465 | 0,046621658 | 0,962814771 | 0,998881056 | YES | 0 | FALSE |
| 473 | 133,936392 | -0,712511253 | 1,511915625 | -0,4712639 | 0,637452292 | 0,998881056 | YES | 0 | FALSE |
| 474 | 13,09952607 | 0,4246652 | 3,036897217 | 0,139835223 | 0,888790183 | 0,998881056 | YES | 0 | FALSE |

| Medication | Ticker | taxLevel | Phylum | Family | Genus | Species |
|---|---|---|---|---|---|---|
| Dopamine agonists | 475 | Genus | Firmicutes | Ruminococcaceae | Faecalibacterium | NA |
| Dopamine agonists | 476 | Genus | Firmicutes | Lachnospiraceae | Lachnospiraceae NK4A136 group | NA |
| Dopamine agonists | 477 | Genus | Firmicutes | Lachnospiraceae | Coprococcus | NA |
| Dopamine agonists | 478 | Genus | Actinobacteriota | Atopobiaceae | Libanicoccus | NA |
| Dopamine agonists | 479 | Genus | Proteobacteria | Sutterellaceae | Sutterella | NA |
| Dopamine agonists | 480 | Genus | Firmicutes | Lachnospiraceae | Shuttleworthia | NA |
| Dopamine agonists | 481 | Genus | Firmicutes | Erysipelotrichaceae | Turicibacter | NA |
| Dopamine agonists | 482 | Genus | Firmicutes | Lachnospiraceae | CAG-56 | NA |
| Dopamine agonists | 483 | Genus | Firmicutes | Lachnospiraceae | [Ruminococcus] gnavus group | NA |
| Dopamine agonists | 484 | Genus | Firmicutes | Butyricicoccaceae | UCG-009 | NA |
| Dopamine agonists | 485 | Genus | Firmicutes | Erysipelotrichaceae | Dielma | NA |
| Dopamine agonists | 486 | Genus | Firmicutes | Acidaminococcaceae | Phascolarctobacterium | NA |
| Dopamine agonists | 487 | Family | Actinobacteriota | Bifidobacteriaceae | NA | NA |
| Dopamine agonists | 488 | Family | Verrucomicrobiota | Akkermansiaceae | NA | NA |
| Dopamine agonists | 489 | Family | Thermoplasmatota | Methanomethylophilaceae | NA | NA |
| Dopamine agonists | 490 | Family | Verrucomicrobiota | vadinBE97 | NA | NA |
| Dopamine agonists | 491 | Family | Firmicutes | Veillonellaceae | NA | NA |
| Dopamine agonists | 492 | Family | Bacteroidota | Muribaculaceae | NA | NA |
| Dopamine agonists | 493 | Family | Firmicutes | Christensenellaceae | NA | NA |
| Dopamine agonists | 494 | Family | Firmicutes | Peptostreptococcaceae | NA | NA |
| Dopamine agonists | 495 | Family | Desulfobacterota | Desulfovibrionaceae | NA | NA |
| Dopamine agonists | 496 | Family | Actinobacteriota | Coriobacteriaceae | NA | NA |
| Dopamine agonists | 497 | Family | Proteobacteria | Pasteurellaceae | NA | NA |
| Dopamine agonists | 498 | Family | Firmicutes | Erysipelatoclostridiaceae | NA | NA |
| Dopamine agonists | 499 | Family | Bacteroidota | Marinifilaceae | NA | NA |
| Dopamine agonists | 500 | Family | Bacteroidota | Barnesiellaceae | NA | NA |
| Dopamine agonists | 501 | Family | Bacteroidota | Bacteroidaceae | NA | NA |
| Dopamine agonists | 502 | Family | Bacteroidota | Prevotellaceae | NA | NA |
| Dopamine agonists | 503 | Family | Firmicutes | Peptococcaceae | NA | NA |
| Dopamine agonists | 504 | Family | Firmicutes | Streptococcaceae | NA | NA |

| Ticker | DESeq2_baseMean | DESeq2_log2-FoldChange | DESeq2_lfcSE | DESeq2_stat | DESeq2_pvalue | DESeq2_padj | DESeq2 converged | ANCOM_W | ANCOM_detected_0.7 |
|---|---|---|---|---|---|---|---|---|---|
| 475 | 2320,25665 | -0,058773396 | 0,320910014 | -0,183146033 | 0,854683438 | 0,998881056 | YES | 0 | FALSE |
| 476 | 642,5453446 | 0,463703431 | 0,862077501 | 0,537890654 | 0,590652541 | 0,998881056 | YES | 0 | FALSE |
| 477 | 495,238952 | 0,682981203 | 0,883632845 | 0,772924192 | 0,439567247 | 0,998881056 | YES | 0 | FALSE |
| 478 | 4,806469958 | -0,243178381 | 3,036982419 | -0,08007237 | 0,936179697 | 0,998881056 | YES | 0 | FALSE |
| 479 | 123,5047879 | 0,84337033 | 0,857777354 | 0,983204238 | 0,325506929 | 0,998881056 | YES | 0 | FALSE |
| 480 | 19,52623002 | -0,204464115 | 1,303227503 | -0,156890577 | 0,875331086 | 0,998881056 | YES | 0 | FALSE |
| 481 | 12,0624508 | 1,51982716 | 1,792333007 | 0,847960258 | 0,396460104 | 0,998881056 | YES | 0 | FALSE |
| 482 | 178,255087 | -4,57908921 | 1,874255243 | -2,443151341 | 0,01455963 | 0,440428816 | YES | 0 | FALSE |
| 483 | 132,4461467 | -1,14957802 | 2,996344824 | -0,383660122 | 0,701230373 | 0,998881056 | YES | 1 | FALSE |
| 484 | 13,40497275 | 1,429169613 | 1,692687843 | 0,844319653 | 0,398490807 | 0,998881056 | YES | 0 | FALSE |
| 485 | 0,502482894 | 0,14439477 | 3,039536654 | 0,04750552 | 0,962110331 | 0,998881056 | YES | 0 | FALSE |
| 486 | 347,7782958 | -0,357042903 | 1,221156439 | -0,292380969 | 0,769995357 | 0,998881056 | YES | 0 | FALSE |
| 487 | 1051,128543 | 1,24965854 | 0,597652397 | 2,090945417 | 0,036532957 | 0,56017201 | YES | 0 | FALSE |
| 488 | 761,6767525 | -0,473920478 | 1,155086875 | -0,4102899 | 0,6815933 | 0,986521117 | YES | 0 | FALSE |
| 489 | 2,948528745 | 0,195532868 | 3,037183032 | 0,064379679 | 0,948667911 | 0,986521117 | YES | 0 | FALSE |
| 490 | 0,147824868 | 0,614561365 | 3,043740044 | 0,201909938 | 0,839987132 | 0,986521117 | YES | 0 | FALSE |
| 491 | 1608,066586 | 1,979870494 | 0,639247101 | 3,097191199 | 0,001953638 | 0,089867358 | YES | 2 | FALSE |
| 492 | 517,5474959 | -0,283829436 | 2,637510061 | -0,107612646 | 0,914302965 | 0,986521117 | YES | 0 | FALSE |
| 493 | 502,0114562 | 0,630080832 | 0,953588718 | 0,660746944 | 0,508774613 | 0,986521117 | YES | 0 | FALSE |
| 494 | 101,3902113 | 0,618695225 | 0,862630272 | 0,717219469 | 0,473238688 | 0,986521117 | YES | 0 | FALSE |
| 495 | 137,0403272 | -0,149684914 | 0,843756685 | -0,177402937 | 0,8591919 | 0,986521117 | YES | 0 | FALSE |
| 496 | 521,0394889 | 0,196621388 | 0,755043267 | 0,260410756 | 0,794546947 | 0,986521117 | YES | 0 | FALSE |
| 497 | 44,08973922 | -0,238790269 | 2,185548327 | -0,109258746 | 0,912997267 | 0,986521117 | YES | 0 | FALSE |
| 498 | 281,8983195 | -0,221529326 | 0,709739218 | -0,31212778 | 0,754943415 | 0,986521117 | YES | 0 | FALSE |
| 499 | 45,35172578 | -0,72895974 | 0,50338611 | -1,448112543 | 0,147585578 | 0,986521117 | YES | 0 | FALSE |
| 500 | 252,5257347 | 0,623050186 | 0,714580352 | 0,871910603 | 0,383257149 | 0,986521117 | YES | 0 | FALSE |
| 501 | 7180,909544 | -0,299757096 | 0,343869113 | -0,871718584 | 0,38336192 | 0,986521117 | YES | 0 | FALSE |
| 502 | 1620,824833 | -0,873860009 | 0,6347497 | -1,376700152 | 0,168604972 | 0,986521117 | YES | 0 | FALSE |
| 503 | 72,25285468 | -0,59932715 | 0,699486895 | -0,856809689 | 0,39155007 | 0,986521117 | YES | 1 | FALSE |
| 504 | 781,8006256 | 0,700148613 | 0,88702856 | 0,789319132 | 0,429925507 | 0,986521117 | YES | 0 | FALSE |

| Medication | Ticker | taxLevel | Phylum | Family | Genus | Species |
|---|---|---|---|---|---|---|
| Dopamine agonists | 505 | Family | Firmicutes | Acholeplasmataceae | NA | NA |
| Dopamine agonists | 506 | Family | Actinobacteriota | Coriobacteriales Incertae Sedis | NA | NA |
| Dopamine agonists | 507 | Family | Euryarchaeota | Methanobacteriaceae | NA | NA |
| Dopamine agonists | 508 | Family | Firmicutes | Clostridiaceae | NA | NA |
| Dopamine agonists | 509 | Family | Firmicutes | Oscillospiraceae | NA | NA |
| Dopamine agonists | 510 | Family | Proteobacteria | Enterobacteriaceae | NA | NA |
| Dopamine agonists | 511 | Family | Bacteroidota | Rikenellaceae | NA | NA |
| Dopamine agonists | 512 | Family | Firmicutes | Anaerovoracaceae | NA | NA |
| Dopamine agonists | 513 | Family | Bacteroidota | Tannerellaceae | NA | NA |
| Dopamine agonists | 514 | Family | Firmicutes | Ruminococcaceae | NA | NA |
| Dopamine agonists | 515 | Family | Firmicutes | Erysipelotrichaceae | NA | NA |
| Dopamine agonists | 516 | Family | Firmicutes | Lactobacillaceae | NA | NA |
| Dopamine agonists | 517 | Family | Firmicutes | UCG-010 | NA | NA |
| Dopamine agonists | 518 | Family | Actinobacteriota | Atopobiaceae | NA | NA |
| Dopamine agonists | 519 | Family | Verrucomicrobiota | Victivallaceae | NA | NA |
| Dopamine agonists | 520 | Family | Actinobacteriota | Actinomycetaceae | NA | NA |
| Dopamine agonists | 521 | Family | Proteobacteria | Oxalobacteraceae | NA | NA |
| Dopamine agonists | 522 | Family | Firmicutes | Monoglobaceae | NA | NA |
| Dopamine agonists | 523 | Family | Firmicutes | [Eubacterium] coprostanoligenes group | NA | NA |
| Dopamine agonists | 524 | Family | Actinobacteriota | Eggerthellaceae | NA | NA |
| Dopamine agonists | 525 | Family | Firmicutes | Butyricicoccaceae | NA | NA |
| Dopamine agonists | 526 | Family | Firmicutes | Lachnospiraceae | NA | NA |
| Dopamine agonists | 527 | Family | Firmicutes | Hydrogenoanaerobacterium | NA | NA |
| Dopamine agonists | 528 | Family | Proteobacteria | Sutterellaceae | NA | NA |
| Dopamine agonists | 529 | Family | Verrucomicrobiota | Puniceicoccaceae | NA | NA |
| Dopamine agonists | 530 | Family | Firmicutes | Defluviitaleaceae | NA | NA |
| Dopamine agonists | 531 | Family | Firmicutes | Ruminiclostridium | NA | NA |
| Dopamine agonists | 532 | Family | Firmicutes | Acidaminococcaceae | NA | NA |

| Ticker | DESeq2_baseMean | DESeq2_log2-FoldChange | DESeq2_lfcSE | DESeq2_stat | DESeq2_pvalue | DESeq2_padj | DESeq2 converged | ANCOM_W | ANCOM_detected_0.7 |
|---|---|---|---|---|---|---|---|---|---|
| 505 | 43,66029827 | 0,264951388 | 3,036780438 | 0,087247463 | 0,930474813 | 0,986521117 | YES | 0 | FALSE |
| 506 | 40,1387451 | 0,581398813 | 1,28594465 | 0,45211807 | 0,651183927 | 0,986521117 | YES | 0 | FALSE |
| 507 | 79,20511498 | -0,064296768 | 1,041353758 | -0,061743445 | 0,950767142 | 0,986521117 | YES | 0 | FALSE |
| 508 | 56,99955518 | -0,839650108 | 1,055680694 | -0,795363705 | 0,426401966 | 0,986521117 | YES | 0 | FALSE |
| 509 | 3327,623644 | 0,320762647 | 0,422427931 | 0,759331056 | 0,447654545 | 0,986521117 | YES | 0 | FALSE |
| 510 | 133,7261228 | 0,288483596 | 1,541729712 | 0,187116843 | 0,851569023 | 0,986521117 | YES | 0 | FALSE |
| 511 | 1099,133854 | 0,345116936 | 0,570473233 | 0,604966046 | 0,545201558 | 0,986521117 | YES | 0 | FALSE |
| 512 | 97,51950525 | 0,765979991 | 0,527377106 | 1,452433147 | 0,146381205 | 0,986521117 | YES | 0 | FALSE |
| 513 | 611,6429564 | 0,443530042 | 0,820598614 | 0,540495724 | 0,588855209 | 0,986521117 | YES | 0 | FALSE |
| 514 | 6687,853491 | 0,122650483 | 0,358368004 | 0,342247303 | 0,732164789 | 0,986521117 | YES | 0 | FALSE |
| 515 | 171,3521872 | -0,466751943 | 0,623035952 | -0,74915732 | 0,45376239 | 0,986521117 | YES | 0 | FALSE |
| 516 | 105,0044887 | 2,909282736 | 1,231757097 | 2,361896468 | 0,018181719 | 0,418179538 | YES | 0 | FALSE |
| 517 | 77,65154318 | 0,261747628 | 1,335860098 | 0,195939401 | 0,844657597 | 0,986521117 | YES | 0 | FALSE |
| 518 | 6,066628147 | 0,495403979 | 2,983723818 | 0,166035468 | 0,868129038 | 0,986521117 | YES | 0 | FALSE |
| 519 | 2,060516529 | -1,382509214 | 2,980750322 | -0,463812485 | 0,642782098 | 0,986521117 | YES | 0 | FALSE |
| 520 | 0,969820909 | 0,079788448 | 3,017775861 | 0,026439488 | 0,978906798 | 0,986521117 | YES | 0 | FALSE |
| 521 | 8,282395154 | 1,399022383 | 2,0007177 | 0,699260262 | 0,484389396 | 0,986521117 | YES | 0 | FALSE |
| 522 | 244,6838494 | 0,405937825 | 0,662149211 | 0,613060952 | 0,539836038 | 0,986521117 | YES | 0 | FALSE |
| 523 | 328,8566557 | 0,632607326 | 0,510849331 | 1,23834424 | 0,215588446 | 0,986521117 | YES | 0 | FALSE |
| 524 | 154,3967646 | 0,734058833 | 0,711133456 | 1,032237798 | 0,301960732 | 0,986521117 | YES | 0 | FALSE |
| 525 | 213,8474286 | 0,311437272 | 0,332395756 | 0,936947199 | 0,348785719 | 0,986521117 | YES | 0 | FALSE |
| 526 | 11942,20647 | -0,06786816 | 0,355050383 | -0,191150786 | 0,84840746 | 0,986521117 | YES | 0 | FALSE |
| 527 | 2,490877974 | -0,823142606 | 3,014502425 | -0,273060854 | 0,784806438 | 0,986521117 | YES | 0 | FALSE |
| 528 | 223,3608361 | 0,460601018 | 0,681507173 | 0,67585645 | 0,49913179 | 0,986521117 | YES | 0 | FALSE |
| 529 | 1,494746013 | -0,110118475 | 3,001644669 | -0,036686046 | 0,970735335 | 0,986521117 | YES | 0 | FALSE |
| 530 | 0,432160341 | 0,25687168 | 3,025577849 | 0,08490004 | 0,93234086 | 0,986521117 | YES | 0 | FALSE |
| 531 | 4,965768924 | -0,05130584 | 3,036912586 | -0,016894079 | 0,986521117 | 0,986521117 | YES | 0 | FALSE |
| 532 | 383,4330824 | -0,246005201 | 1,161032699 | -0,211884817 | 0,832196896 | 0,986521117 | YES | 0 | FALSE |

**Supplementary Table 7b: users of only pramipexole**

| Medication | Ticker | taxLevel | Phylum | Family | Genus | Species |
|---|---|---|---|---|---|---|
| Pramipexole | 1 | ASV | Actinobacteriota | Bifidobacteriaceae | Bifidobacterium | NA |
| Pramipexole | 2 | ASV | Verrucomicrobiota | Akkermansiaceae | Akkermansia | muciniphila |
| Pramipexole | 3 | ASV | Firmicutes | Christensenellaceae | NA | NA |
| Pramipexole | 4 | ASV | Firmicutes | Lachnospiraceae | Roseburia | NA |
| Pramipexole | 5 | ASV | Firmicutes | NA | NA | NA |
| Pramipexole | 6 | ASV | Firmicutes | Christensenellaceae | Christensenellaceae R-7 group | NA |
| Pramipexole | 7 | ASV | Bacteroidota | Bacteroidaceae | Bacteroides | NA |
| Pramipexole | 8 | ASV | Bacteroidota | Bacteroidaceae | Bacteroides | eggerthii |
| Pramipexole | 9 | ASV | Firmicutes | [Eubacterium] coprostanoligenes group | NA | NA |
| Pramipexole | 10 | ASV | Firmicutes | Ruminococcaceae | Subdoligranulum | NA |
| Pramipexole | 11 | ASV | Firmicutes | Ruminococcaceae | Faecalibacterium | prausnitzii |
| Pramipexole | 12 | ASV | Firmicutes | Lachnospiraceae | Lachnospira | pectinoschiza |
| Pramipexole | 13 | ASV | Bacteroidota | Bacteroidaceae | Bacteroides | coprocola |
| Pramipexole | 14 | ASV | Firmicutes | Ruminococcaceae | NA | NA |
| Pramipexole | 15 | ASV | Firmicutes | Oscillospiraceae | UCG-003 | NA |
| Pramipexole | 16 | ASV | Bacteroidota | Rikenellaceae | Alistipes | finegoldii |
| Pramipexole | 17 | ASV | Firmicutes | NA | NA | NA |
| Pramipexole | 18 | ASV | Firmicutes | Ruminococcaceae | Subdoligranulum | NA |
| Pramipexole | 19 | ASV | Firmicutes | Veillonellaceae | Dialister | invisus |
| Pramipexole | 20 | ASV | Firmicutes | Lachnospiraceae | Anaerostipes | hadrus |
| Pramipexole | 21 | ASV | Firmicutes | Lachnospiraceae | [Ruminococcus] torques group | NA |
| Pramipexole | 22 | ASV | Firmicutes | NA | NA | NA |
| Pramipexole | 23 | ASV | Firmicutes | Oscillospiraceae | UCG-002 | NA |
| Pramipexole | 24 | ASV | Bacteroidota | Tannerellaceae | Parabacteroides | distasonis |
| Pramipexole | 25 | ASV | Firmicutes | Lachnospiraceae | Lachnospiraceae NK4A136 group | bacterium |
| Pramipexole | 26 | ASV | Firmicutes | Oscillospiraceae | NK4A214 group | NA |
| Pramipexole | 27 | ASV | Firmicutes | Christensenellaceae | Christensenellaceae R-7 group | NA |
| Pramipexole | 28 | ASV | Firmicutes | Lachnospiraceae | Fusicatenibacter | saccharivorans |
| Pramipexole | 29 | ASV | Firmicutes | Lachnospiraceae | NA | NA |

| Ticker | DESeq2_baseMean | DESeq2_log2-FoldChange | DESeq2_lfcSE | DESeq2_stat | DESeq2_pvalue | DESeq2_padj | DESeq2 converged | ANCOM_W | ANCOM_detected_0.7 |
|---|---|---|---|---|---|---|---|---|---|
| 1 | 538,6681246 | 3,634967203 | 1,756735556 | 2,069160148 | 0,038531061 | 0,999805755 | YES | 142 | FALSE |
| 2 | 435,6769742 | -1,890503189 | 2,640364449 | -0,71600085 | 0,473990824 | 0,999805755 | YES | 0 | FALSE |
| 3 | 137,3160995 | 1,645146732 | 2,988227783 | 0,550542613 | 0,581947257 | 0,999805755 | YES | 0 | FALSE |
| 4 | 75,89357853 | 2,399860894 | 2,988821712 | 0,802945483 | 0,422006245 | 0,999805755 | YES | 0 | FALSE |
| 5 | 2,316101286 | 3,251950459 | 3,017008612 | 1,077872448 | 0,281090683 | 0,999805755 | YES | 0 | FALSE |
| 6 | 10,88069728 | 0,27839282 | 3,099829023 | 0,089809089 | 0,928438925 | 0,999805755 | YES | 0 | FALSE |
| 7 | 79,02373633 | 0,08762014 | 2,660618247 | 0,032932248 | 0,973728616 | 0,999805755 | YES | 0 | FALSE |
| 8 | 194,604676 | -0,730894512 | 3,0994269 | -0,235816019 | 0,813575438 | 0,999805755 | YES | 0 | FALSE |
| 9 | 72,43561471 | -0,475622468 | 3,044067076 | -0,156245725 | 0,875839336 | 0,999805755 | YES | 0 | FALSE |
| 10 | 334,2045594 | -0,020151313 | 1,870176468 | -0,010775086 | 0,991402891 | 0,999805755 | YES | 0 | FALSE |
| 11 | 33,15660909 | -0,734769636 | 3,099565435 | -0,237055694 | 0,812613584 | 0,999805755 | YES | 0 | FALSE |
| 12 | 72,2808885 | -0,261514129 | 2,475701311 | -0,105632343 | 0,915874063 | 0,999805755 | YES | 0 | FALSE |
| 13 | 9,580085824 | -0,733296869 | 3,100047634 | -0,236543742 | 0,813010769 | 0,999805755 | YES | 0 | FALSE |
| 14 | 3,999290986 | 0,674672403 | 3,101650128 | 0,217520473 | 0,827802754 | 0,999805755 | YES | 0 | FALSE |
| 15 | 64,248212 | -0,538283092 | 3,04185702 | -0,176958709 | 0,859540823 | 0,999805755 | YES | 0 | FALSE |
| 16 | 7,009449063 | 0,992616769 | 3,100405831 | 0,320157045 | 0,748849284 | 0,999805755 | YES | 0 | FALSE |
| 17 | 3,193361941 | -0,729221878 | 3,101393195 | -0,235127193 | 0,814110013 | 0,999805755 | YES | 0 | FALSE |
| 18 | 0,543550969 | -0,701397484 | 3,110790556 | -0,225472423 | 0,821611776 | 0,999805755 | YES | 0 | FALSE |
| 19 | 1313,509776 | 1,170981571 | 2,65406994 | 0,441202228 | 0,6590666 | 0,999805755 | YES | 0 | FALSE |
| 20 | 130,5332795 | 0,608702245 | 2,563118738 | 0,237484997 | 0,812280557 | 0,999805755 | YES | 0 | FALSE |
| 21 | 31,3318211 | -0,164012106 | 3,042275663 | -0,053910994 | 0,957006077 | 0,999805755 | YES | 0 | FALSE |
| 22 | 346,05527 | 1,33415145 | 3,042140719 | 0,438556784 | 0,660982717 | 0,999805755 | YES | 0 | FALSE |
| 23 | 18,20895745 | -0,734275945 | 3,099726504 | -0,236884107 | 0,8127467 | 0,999805755 | YES | 0 | FALSE |
| 24 | 162,320947 | 1,283141063 | 2,65516904 | 0,483261534 | 0,628910049 | 0,999805755 | YES | 0 | FALSE |
| 25 | 89,10619009 | -0,000620186 | 2,547491854 | -0,00024345 | 0,999805755 | 0,999805755 | YES | 0 | FALSE |
| 26 | 43,08507005 | 0,210509636 | 3,048569383 | 0,069051942 | 0,944948274 | 0,999805755 | YES | 0 | FALSE |
| 27 | 2,51392323 | -0,727591411 | 3,101934274 | -0,23456055 | 0,814549832 | 0,999805755 | YES | 0 | FALSE |
| 28 | 902,9077777 | 3,157972608 | 2,218954686 | 1,423180305 | 0,154683893 | 0,999805755 | YES | 0 | FALSE |
| 29 | 187,7566819 | 2,324599677 | 2,56517024 | 0,906216531 | 0,364821255 | 0,999805755 | YES | 0 | FALSE |

| Medication | Ticker | taxLevel | Phylum | Family | Genus | Species |
|---|---|---|---|---|---|---|
| Pramipexole | 30 | ASV | Actinobacteriota | Eggerthellaceae | Senegalimassilia | anaerobia |
| Pramipexole | 31 | ASV | Actinobacteriota | Eggerthellaceae | Enterorhabdus | NA |
| Pramipexole | 32 | ASV | Bacteroidota | Rikenellaceae | Alistipes | inops |
| Pramipexole | 33 | ASV | Firmicutes | Ruminococcaceae | NA | NA |
| Pramipexole | 34 | ASV | Firmicutes | Butyricicoccaceae | Butyricicoccus | faecihominis |
| Pramipexole | 35 | ASV | Bacteroidota | Rikenellaceae | Alistipes | obesi |
| Pramipexole | 36 | ASV | Firmicutes | Christensenellaceae | Christensenellaceae R-7 group | NA |
| Pramipexole | 37 | ASV | Firmicutes | Lachnospiraceae | Blautia | massiliensis |
| Pramipexole | 38 | ASV | Firmicutes | Lachnospiraceae | Lachnoclostridium | NA |
| Pramipexole | 39 | ASV | Firmicutes | Ruminococcaceae | Faecalibacterium | prausnitzii |
| Pramipexole | 40 | ASV | Firmicutes | Peptostreptococcaceae | Terrisporobacter | mayombei |
| Pramipexole | 41 | ASV | Desulfobacterota | Desulfovibrionaceae | Bilophila | wadsworthia |
| Pramipexole | 42 | ASV | Bacteroidota | Muribaculaceae | NA | NA |
| Pramipexole | 43 | ASV | Firmicutes | Erysipelotrichaceae | Turicibacter | sanguinis |
| Pramipexole | 44 | ASV | Firmicutes | Oscillospiraceae | Oscillibacter | NA |
| Pramipexole | 45 | ASV | Firmicutes | NA | NA | NA |
| Pramipexole | 46 | ASV | Firmicutes | Christensenellaceae | Christensenellaceae R-7 group | NA |
| Pramipexole | 47 | ASV | Firmicutes | Butyricicoccaceae | Butyricicoccus | NA |
| Pramipexole | 48 | ASV | Firmicutes | Lachnospiraceae | Roseburia | hominis |
| Pramipexole | 49 | ASV | Firmicutes | Streptococcaceae | Streptococcus | NA |
| Pramipexole | 50 | ASV | Firmicutes | NA | NA | NA |
| Pramipexole | 51 | ASV | Firmicutes | Ruminococcaceae | Ruminococcus | NA |
| Pramipexole | 52 | ASV | Firmicutes | Oscillospiraceae | UCG-005 | NA |
| Pramipexole | 53 | ASV | Firmicutes | Lachnospiraceae | [Eubacterium] ventriosum group | NA |
| Pramipexole | 54 | ASV | Firmicutes | Lachnospiraceae | Roseburia | NA |
| Pramipexole | 55 | ASV | Firmicutes | Peptostreptococcaceae | Romboutsia | NA |
| Pramipexole | 56 | ASV | Firmicutes | Oscillospiraceae | UCG-002 | NA |
| Pramipexole | 57 | ASV | Firmicutes | Oscillospiraceae | Intestinimonas | NA |
| Pramipexole | 58 | ASV | Firmicutes | Lachnospiraceae | Lachnoclostridium | edouardi |
| Pramipexole | 59 | ASV | Actinobacteriota | Coriobacteriaceae | Collinsella | aerofaciens |

| Ticker | DESeq2_baseMean | DESeq2_log2-FoldChange | DESeq2_lfcSE | DESeq2_stat | DESeq2_pvalue | DESeq2_padj | DESeq2 converged | ANCOM_W | ANCOM_detected_0.7 |
|---|---|---|---|---|---|---|---|---|---|
| 30 | 0,412320533 | 0,944951362 | 3,115784583 | 0,303278785 | 0,761677413 | 0,999805755 | YES | 0 | FALSE |
| 31 | 10,07662933 | 0,045945081 | 2,582428394 | 0,017791425 | 0,985805246 | 0,999805755 | YES | 0 | FALSE |
| 32 | 236,5477575 | 0,811529674 | 3,041662824 | 0,266804613 | 0,789619602 | 0,999805755 | YES | 0 | FALSE |
| 33 | 9,648029695 | -0,733311356 | 3,100042868 | -0,236548779 | 0,813006861 | 0,999805755 | YES | 0 | FALSE |
| 34 | 17,18883172 | 2,334880391 | 2,99490527 | 0,779617444 | 0,435616085 | 0,999805755 | YES | 0 | FALSE |
| 35 | 73,45126475 | 1,930612786 | 2,171891408 | 0,888908524 | 0,374052244 | 0,999805755 | YES | 0 | FALSE |
| 36 | 10,12363679 | -0,733407345 | 3,100011294 | -0,236582153 | 0,812980967 | 0,999805755 | YES | 0 | FALSE |
| 37 | 654,9178766 | 1,486182317 | 1,877466438 | 0,791589286 | 0,428600196 | 0,999805755 | YES | 0 | FALSE |
| 38 | 5,575019215 | 1,02451633 | 3,101447574 | 0,330334886 | 0,741146936 | 0,999805755 | YES | 0 | FALSE |
| 39 | 13,66365008 | -0,138334769 | 3,100013859 | -0,04462392 | 0,964407077 | 0,999805755 | YES | 0 | FALSE |
| 40 | 0,543550969 | -0,701397484 | 3,110790556 | -0,225472423 | 0,821611776 | 0,999805755 | YES | 0 | FALSE |
| 41 | 73,02845104 | 0,675767654 | 2,353129395 | 0,287178281 | 0,773975816 | 0,999805755 | YES | 0 | FALSE |
| 42 | 60,60593301 | -0,735044894 | 3,099476471 | -0,237151306 | 0,812539411 | 0,999805755 | YES | 0 | FALSE |
| 43 | 0,274880355 | 0,923060493 | 3,123178622 | 0,295551617 | 0,767572527 | 0,999805755 | YES | 0 | FALSE |
| 44 | 226,320627 | 1,314713827 | 1,88225175 | 0,698479269 | 0,484877518 | 0,999805755 | YES | 0 | FALSE |
| 45 | 14,40410067 | -0,733987847 | 3,099820775 | -0,236783963 | 0,812824394 | 0,999805755 | YES | 0 | FALSE |
| 46 | 81,53264531 | -0,73513145 | 3,099448875 | -0,237181344 | 0,812516109 | 0,999805755 | YES | 0 | FALSE |
| 47 | 3,465137426 | -0,729697665 | 3,101235704 | -0,235292553 | 0,813981675 | 0,999805755 | YES | 0 | FALSE |
| 48 | 34,40256681 | -0,643101544 | 3,0427724 | -0,211353811 | 0,832611196 | 0,999805755 | YES | 0 | FALSE |
| 49 | 2,125665448 | 0,546955581 | 3,101948439 | 0,176326458 | 0,860037477 | 0,999805755 | YES | 0 | FALSE |
| 50 | 66,5504132 | 2,641832776 | 2,99043532 | 0,883427492 | 0,377005342 | 0,999805755 | YES | 0 | FALSE |
| 51 | 45,98798335 | 0,238894681 | 3,042098044 | 0,07852958 | 0,937406802 | 0,999805755 | YES | 0 | FALSE |
| 52 | 6,191430665 | -0,456307993 | 3,100263173 | -0,147183632 | 0,88298708 | 0,999805755 | YES | 0 | FALSE |
| 53 | 65,37327323 | 1,168315248 | 3,042048618 | 0,384055416 | 0,700937375 | 0,999805755 | YES | 0 | FALSE |
| 54 | 63,25682592 | 1,069762194 | 2,597181053 | 0,411893577 | 0,680417429 | 0,999805755 | YES | 0 | FALSE |
| 55 | 35,47408363 | 3,305504623 | 2,462598039 | 1,342283463 | 0,179504103 | 0,999805755 | YES | 0 | FALSE |
| 56 | 17,09603952 | -1,323358962 | 2,995446677 | -0,441790192 | 0,658641037 | 0,999805755 | YES | 0 | FALSE |
| 57 | 1,087101937 | -0,717772049 | 3,105217782 | -0,231150309 | 0,817198033 | 0,999805755 | YES | 0 | FALSE |
| 58 | 13,39462051 | 0,117610366 | 3,0997338 | 0,037942086 | 0,969733857 | 0,999805755 | YES | 0 | FALSE |
| 59 | 835,9405041 | 1,398881079 | 1,602609227 | 0,872877215 | 0,382730007 | 0,999805755 | YES | 0 | FALSE |

| Medication | Ticker | taxLevel | Phylum | Family | Genus | Species |
|---|---|---|---|---|---|---|
| Pramipexole | 60 | ASV | Firmicutes | Oscillospiraceae | NA | NA |
| Pramipexole | 61 | ASV | Proteobacteria | Pasteurellaceae | Haemophilus | NA |
| Pramipexole | 62 | ASV | Actinobacteriota | Coriobacteriales Incertae Sedis | NA | NA |
| Pramipexole | 63 | ASV | Firmicutes | NA | NA | NA |
| Pramipexole | 64 | ASV | Firmicutes | NA | NA | NA |
| Pramipexole | 65 | ASV | Firmicutes | Lachnospiraceae | Dorea | longicatena |
| Pramipexole | 66 | ASV | Firmicutes | Ruminococcaceae | Ruminococcus | NA |
| Pramipexole | 67 | ASV | Firmicutes | Ruminococcaceae | Ruminococcus | NA |
| Pramipexole | 68 | ASV | Firmicutes | Lachnospiraceae | [Ruminococcus] torques group | NA |
| Pramipexole | 69 | ASV | Firmicutes | Ruminococcaceae | Ruminococcus | champanellensis |
| Pramipexole | 70 | ASV | Firmicutes | Erysipelatoclostridiaceae | Erysipelotrichaceae UCG-003 | bacterium |
| Pramipexole | 71 | ASV | Firmicutes | Ruminococcaceae | UBA1819 | NA |
| Pramipexole | 72 | ASV | Firmicutes | Lachnospiraceae | Lachnospiraceae NK4A136 group | NA |
| Pramipexole | 73 | ASV | Bacteroidota | Marinifilaceae | Odoribacter | splanchnicus |
| Pramipexole | 74 | ASV | Actinobacteriota | Eggerthellaceae | Adlercreutzia | equolifaciens |
| Pramipexole | 75 | ASV | Bacteroidota | Barnesiellaceae | Barnesiella | intestinihominis |
| Pramipexole | 76 | ASV | Firmicutes | Ruminococcaceae | Ruminococcus | NA |
| Pramipexole | 77 | ASV | Bacteroidota | Bacteroidaceae | Bacteroides | eggerthii |
| Pramipexole | 78 | ASV | Bacteroidota | Bacteroidaceae | Bacteroides | vulgatus |
| Pramipexole | 79 | ASV | Bacteroidota | Prevotellaceae | Prevotella | copri |
| Pramipexole | 80 | ASV | Firmicutes | Lachnospiraceae | [Eubacterium] xylanophilum group | NA |
| Pramipexole | 81 | ASV | Firmicutes | Oscillospiraceae | UCG-002 | NA |
| Pramipexole | 82 | ASV | Firmicutes | NA | NA | NA |
| Pramipexole | 83 | ASV | Firmicutes | Oscillospiraceae | Intestinimonas | NA |
| Pramipexole | 84 | ASV | Firmicutes | Ruminococcaceae | Faecalibacterium | prausnitzii |
| Pramipexole | 85 | ASV | Firmicutes | Lachnospiraceae | Lachnospiraceae FCS020 group | NA |
| Pramipexole | 86 | ASV | Firmicutes | NA | NA | NA |
| Pramipexole | 87 | ASV | Firmicutes | Lachnospiraceae | Lachnospiraceae UCG-010 | NA |
| Pramipexole | 88 | ASV | Firmicutes | Lachnospiraceae | Lachnospira | pectinoschiza |
| Pramipexole | 89 | ASV | Firmicutes | Anaerovoracaceae | Family XIII AD3011 group | NA |

| Ticker | DESeq2_baseMean | DESeq2_log2-FoldChange | DESeq2_lfcSE | DESeq2_stat | DESeq2_pvalue | DESeq2_padj | DESeq2 converged | ANCOM_W | ANCOM_detected_0.7 |
|---|---|---|---|---|---|---|---|---|---|
| 60 | 65,60773752 | 1,314262899 | 2,554842199 | 0,514420382 | 0,606958112 | 0,999805755 | YES | 0 | FALSE |
| 61 | 13,88186595 | -1,036444726 | 3,029269114 | -0,342143496 | 0,732242905 | 0,999805755 | YES | 0 | FALSE |
| 62 | 14,09708202 | 0,065593968 | 3,099956646 | 0,02115964 | 0,983118309 | 0,999805755 | YES | 0 | FALSE |
| 63 | 3,872800652 | -0,730286968 | 3,101040614 | -0,235497389 | 0,813822705 | 0,999805755 | YES | 0 | FALSE |
| 64 | 9,036534855 | -0,733173173 | 3,100088331 | -0,236500736 | 0,813044136 | 0,999805755 | YES | 0 | FALSE |
| 65 | 512,566191 | -0,497983064 | 2,261384871 | -0,220211549 | 0,825706403 | 0,999805755 | YES | 1 | FALSE |
| 66 | 5,435509687 | -0,731734117 | 3,100562478 | -0,236000443 | 0,813432327 | 0,999805755 | YES | 0 | FALSE |
| 67 | 324,1512848 | 6,067619739 | 3,043659069 | 1,993528053 | 0,046203655 | 0,999805755 | YES | 0 | FALSE |
| 68 | 103,5982391 | 0,110429427 | 3,099441308 | 0,035628817 | 0,97157833 | 0,999805755 | YES | 0 | FALSE |
| 69 | 62,3688652 | -0,699422895 | 3,099526641 | -0,225654745 | 0,821469958 | 0,999805755 | YES | 0 | FALSE |
| 70 | 227,8322627 | 0,403572043 | 2,244027252 | 0,179842755 | 0,857276017 | 0,999805755 | YES | 0 | FALSE |
| 71 | 12,78580628 | 0,647269575 | 2,559235125 | 0,252915243 | 0,80033371 | 0,999805755 | YES | 0 | FALSE |
| 72 | 23,57099048 | 0,994957756 | 3,099678331 | 0,320987422 | 0,748219923 | 0,999805755 | YES | 0 | FALSE |
| 73 | 22,98590963 | -0,176488888 | 1,932590752 | -0,091322432 | 0,927236394 | 0,999805755 | YES | 0 | FALSE |
| 74 | 12,52817079 | 2,307801484 | 3,003475075 | 0,768377105 | 0,442263178 | 0,999805755 | YES | 0 | FALSE |
| 75 | 202,7115034 | 0,732315854 | 3,041560273 | 0,240769798 | 0,809733539 | 0,999805755 | YES | 0 | FALSE |
| 76 | 43,0084704 | -0,734909036 | 3,099520449 | -0,23710411 | 0,812576024 | 0,999805755 | YES | 0 | FALSE |
| 77 | 223,3965113 | -0,732935778 | 3,099419502 | -0,236475178 | 0,813063966 | 0,999805755 | YES | 0 | FALSE |
| 78 | 1305,812412 | 0,910642088 | 3,041457152 | 0,299409803 | 0,764627383 | 0,999805755 | YES | 0 | FALSE |
| 79 | 1275,5993 | 0,730204342 | 3,051051551 | 0,239328746 | 0,810850677 | 0,999805755 | YES | 0 | FALSE |
| 80 | 51,36103563 | -0,891403598 | 2,521513332 | -0,353519288 | 0,723699171 | 0,999805755 | YES | 0 | FALSE |
| 81 | 156,8534097 | 1,192067582 | 3,047653356 | 0,391142772 | 0,695691707 | 0,999805755 | YES | 0 | FALSE |
| 82 | 3,940744523 | -0,73037346 | 3,101011996 | -0,235527454 | 0,813799373 | 0,999805755 | YES | 0 | FALSE |
| 83 | 69,02539648 | 0,618064606 | 3,041948445 | 0,2031805 | 0,838993954 | 0,999805755 | YES | 0 | FALSE |
| 84 | 688,2219105 | -1,327931812 | 2,351815253 | -0,564641211 | 0,572317829 | 0,999805755 | YES | 0 | FALSE |
| 85 | 35,15341408 | 0,648733621 | 2,264423535 | 0,286489524 | 0,774503216 | 0,999805755 | YES | 0 | FALSE |
| 86 | 78,20423814 | 1,15252951 | 3,042151383 | 0,378853438 | 0,704796702 | 0,999805755 | YES | 0 | FALSE |
| 87 | 9,483372262 | 0,993481988 | 3,100136305 | 0,32046397 | 0,748616639 | 0,999805755 | YES | 0 | FALSE |
| 88 | 83,48713658 | -1,926051269 | 2,558023331 | -0,752945153 | 0,451482873 | 0,999805755 | YES | 0 | FALSE |
| 89 | 11,8953349 | 2,164039083 | 2,529607819 | 0,855484027 | 0,392283243 | 0,999805755 | YES | 0 | FALSE |

| Medication | Ticker | taxLevel | Phylum | Family | Genus | Species |
|---|---|---|---|---|---|---|
| Pramipexole | 90 | ASV | Firmicutes | Lachnospiraceae | Lachnospiraceae UCG-004 | NA |
| Pramipexole | 91 | ASV | Bacteroidota | Bacteroidaceae | Bacteroides | uniformis |
| Pramipexole | 92 | ASV | Firmicutes | Oscillospiraceae | Intestinimonas | NA |
| Pramipexole | 93 | ASV | Firmicutes | Oscillospiraceae | NK4A214 group | NA |
| Pramipexole | 94 | ASV | Firmicutes | Lachnospiraceae | Lachnospiraceae UCG-001 | NA |
| Pramipexole | 95 | ASV | Firmicutes | Oscillospiraceae | Flavonifractor | plautii |
| Pramipexole | 96 | ASV | Firmicutes | Streptococcaceae | Streptococcus | NA |
| Pramipexole | 97 | ASV | Firmicutes | Ruminococcaceae | [Eubacterium] siraeum group | NA |
| Pramipexole | 98 | ASV | Proteobacteria | NA | NA | NA |
| Pramipexole | 99 | ASV | Firmicutes | Acholeplasmataceae | Anaeroplasma | NA |
| Pramipexole | 100 | ASV | Cyanobacteria | NA | NA | NA |
| Pramipexole | 101 | ASV | Bacteroidota | Bacteroidaceae | Bacteroides | clarus |
| Pramipexole | 102 | ASV | Firmicutes | Butyricicoccaceae | NA | NA |
| Pramipexole | 103 | ASV | Bacteroidota | Bacteroidaceae | Bacteroides | uniformis |
| Pramipexole | 104 | ASV | Bacteroidota | Prevotellaceae | Alloprevotella | NA |
| Pramipexole | 105 | ASV | Firmicutes | [Eubacterium] coprostanoligenes group | NA | NA |
| Pramipexole | 106 | ASV | Firmicutes | Lachnospiraceae | Tyzzerella | NA |
| Pramipexole | 107 | ASV | Euryarchaeota | Methanobacteriaceae | Methanosphaera | NA |
| Pramipexole | 108 | ASV | Proteobacteria | NA | NA | NA |
| Pramipexole | 109 | ASV | Actinobacteriota | Bifidobacteriaceae | Bifidobacterium | NA |
| Pramipexole | 110 | ASV | Bacteroidota | Prevotellaceae | Paraprevotella | NA |
| Pramipexole | 111 | ASV | Firmicutes | Erysipelotrichaceae | Solobacterium | NA |
| Pramipexole | 112 | ASV | Bacteroidota | Tannerellaceae | Parabacteroides | distasonis |
| Pramipexole | 113 | ASV | Firmicutes | Lachnospiraceae | Lachnospira | NA |
| Pramipexole | 114 | ASV | Actinobacteriota | Coriobacteriales Incertae Sedis | NA | NA |
| Pramipexole | 115 | ASV | Bacteroidota | Bacteroidaceae | Bacteroides | ovatus |
| Pramipexole | 116 | ASV | Firmicutes | Lachnospiraceae | Lachnospiraceae AC2044 group | NA |
| Pramipexole | 117 | ASV | Euryarchaeota | Methanobacteriaceae | Methanobrevibacter | NA |
| Pramipexole | 118 | ASV | Firmicutes | Anaerovoracaceae | Family XIII UCG-001 | NA |
| Pramipexole | 119 | ASV | Firmicutes | Ruminococcaceae | Incertae Sedis | NA |

| Ticker | DESeq2_baseMean | DESeq2_log2-FoldChange | DESeq2_lfcSE | DESeq2_stat | DESeq2_pvalue | DESeq2_padj | DESeq2 converged | ANCOM_W | ANCOM_detected_0.7 |
|---|---|---|---|---|---|---|---|---|---|
| 90 | 41,25216796 | -0,042463005 | 3,042044083 | -0,013958708 | 0,988862924 | 0,999805755 | YES | 0 | FALSE |
| 91 | 542,3847711 | 0,031100777 | 2,018475751 | 0,015408051 | 0,98770664 | 0,999805755 | YES | 0 | FALSE |
| 92 | 40,29552417 | -1,976014809 | 2,387301047 | -0,827719157 | 0,407829568 | 0,999805755 | YES | 0 | FALSE |
| 93 | 11,55045809 | -0,733648718 | 3,099932136 | -0,236666058 | 0,812915869 | 0,999805755 | YES | 0 | FALSE |
| 94 | 11,8970432 | 0,185573106 | 3,044286007 | 0,060957842 | 0,951392784 | 0,999805755 | YES | 0 | FALSE |
| 95 | 118,9162928 | -0,022243029 | 2,107834443 | -0,01055255 | 0,991580439 | 0,999805755 | YES | 0 | FALSE |
| 96 | 546,0398678 | 2,392253836 | 1,900099562 | 1,259014993 | 0,208024917 | 0,999805755 | YES | 0 | FALSE |
| 97 | 20,52132053 | -2,368490088 | 3,001370164 | -0,789136281 | 0,430032359 | 0,999805755 | YES | 0 | FALSE |
| 98 | 6,388883913 | -0,648457685 | 3,100832569 | -0,209123734 | 0,834351647 | 0,999805755 | YES | 0 | FALSE |
| 99 | 75,53470726 | 0,492270593 | 3,099440151 | 0,158825649 | 0,873806241 | 0,999805755 | YES | 0 | FALSE |
| 100 | 2,183770884 | -1,200726505 | 3,030099196 | -0,396266402 | 0,69190851 | 0,999805755 | YES | 0 | FALSE |
| 101 | 80,22179945 | 0,014408067 | 3,099462245 | 0,00464857 | 0,996290991 | 0,999805755 | YES | 0 | FALSE |
| 102 | 8,684758312 | 1,168646233 | 3,047362074 | 0,383494381 | 0,701353236 | 0,999805755 | YES | 0 | FALSE |
| 103 | 167,3961647 | -0,193542014 | 2,345629446 | -0,08251176 | 0,934239767 | 0,999805755 | YES | 0 | FALSE |
| 104 | 7,307610916 | -0,116017985 | 3,100417099 | -0,037420122 | 0,970150029 | 0,999805755 | YES | 0 | FALSE |
| 105 | 0,543550969 | -0,701397484 | 3,110790556 | -0,225472423 | 0,821611776 | 0,999805755 | YES | 0 | FALSE |
| 106 | 7,906390886 | 1,026537039 | 3,100839072 | 0,331051375 | 0,74060568 | 0,999805755 | YES | 0 | FALSE |
| 107 | 6,36457686 | 0,285923008 | 3,100156298 | 0,092228578 | 0,926516433 | 0,999805755 | YES | 0 | FALSE |
| 108 | 56,81768963 | 0,240546444 | 3,075593347 | 0,078211394 | 0,937659898 | 0,999805755 | YES | 0 | FALSE |
| 109 | 594,797018 | 1,651082686 | 1,583020359 | 1,042995232 | 0,296950501 | 0,999805755 | YES | 0 | FALSE |
| 110 | 53,74541723 | -0,215705374 | 3,041985752 | -0,070909397 | 0,943469865 | 0,999805755 | YES | 0 | FALSE |
| 111 | 5,299621945 | -0,731641332 | 3,10059292 | -0,2359682 | 0,813457346 | 0,999805755 | YES | 0 | FALSE |
| 112 | 41,34844229 | -0,190859063 | 3,099557426 | -0,061576231 | 0,950900306 | 0,999805755 | YES | 0 | FALSE |
| 113 | 1,562709035 | -0,722987167 | 3,103468502 | -0,232961013 | 0,81579168 | 0,999805755 | YES | 0 | FALSE |
| 114 | 22,99427955 | 0,701738218 | 3,099831496 | 0,226379472 | 0,820906292 | 0,999805755 | YES | 0 | FALSE |
| 115 | 25,44965612 | -0,52661077 | 3,099769167 | -0,169887092 | 0,865098933 | 0,999805755 | YES | 0 | FALSE |
| 116 | 1,358877422 | -0,721183536 | 3,104071184 | -0,232334728 | 0,816278041 | 0,999805755 | YES | 0 | FALSE |
| 117 | 57,00131746 | 1,442904943 | 2,03394812 | 0,709410889 | 0,478069533 | 0,999805755 | YES | 0 | FALSE |
| 118 | 17,45796301 | -0,104300261 | 2,563491922 | -0,040686791 | 0,967545592 | 0,999805755 | YES | 0 | FALSE |
| 119 | 123,2049149 | 1,154051835 | 2,587088316 | 0,446081345 | 0,655538488 | 0,999805755 | YES | 0 | FALSE |

| Medication | Ticker | taxLevel | Phylum | Family | Genus | Species |
|---|---|---|---|---|---|---|
| Pramipexole | 120 | ASV | Firmicutes | Oscillospiraceae | Colidextribacter | NA |
| Pramipexole | 121 | ASV | Firmicutes | Lachnospiraceae | Lachnoclostridium | NA |
| Pramipexole | 122 | ASV | Firmicutes | Ruminococcaceae | Incertae Sedis | NA |
| Pramipexole | 123 | ASV | Firmicutes | Ruminococcaceae | CAG-352 | NA |
| Pramipexole | 124 | ASV | Firmicutes | Christensenellaceae | Christensenellaceae R-7 group | NA |
| Pramipexole | 125 | ASV | Firmicutes | Erysipelotrichaceae | Catenisphaera | NA |
| Pramipexole | 126 | ASV | Firmicutes | Oscillospiraceae | NK4A214 group | NA |
| Pramipexole | 127 | ASV | Bacteroidota | Rikenellaceae | Alistipes | ihumii |
| Pramipexole | 128 | ASV | Firmicutes | Clostridiaceae | Clostridium sensu stricto 1 | NA |
| Pramipexole | 129 | ASV | Actinobacteriota | Eggerthellaceae | Slackia | isoflavoniconvertens |
| Pramipexole | 130 | ASV | Firmicutes | Oscillospiraceae | NK4A214 group | NA |
| Pramipexole | 131 | ASV | Firmicutes | Lachnospiraceae | NA | NA |
| Pramipexole | 132 | ASV | Firmicutes | Christensenellaceae | Christensenellaceae R-7 group | NA |
| Pramipexole | 133 | ASV | Firmicutes | Erysipelotrichaceae | NA | NA |
| Pramipexole | 134 | ASV | Firmicutes | Oscillospiraceae | UCG-002 | NA |
| Pramipexole | 135 | ASV | Firmicutes | Erysipelatoclostridiaceae | Erysipelatoclostridium | NA |
| Pramipexole | 136 | ASV | Firmicutes | Anaerovoracaceae | Mogibacterium | NA |
| Pramipexole | 137 | ASV | Firmicutes | Ruminococcaceae | Negativibacillus | NA |
| Pramipexole | 138 | ASV | Proteobacteria | Sutterellaceae | Sutterella | NA |
| Pramipexole | 139 | ASV | Firmicutes | Streptococcaceae | Lactococcus | NA |
| Pramipexole | 140 | ASV | Firmicutes | Ruminococcaceae | Subdoligranulum | NA |
| Pramipexole | 141 | ASV | Proteobacteria | Enterobacteriaceae | Escherichia-Shigella | NA |
| Pramipexole | 142 | ASV | Firmicutes | Lachnospiraceae | GCA-900066575 | NA |
| Pramipexole | 143 | ASV | Firmicutes | Lachnospiraceae | Lachnospiraceae UCG-004 | NA |
| Pramipexole | 144 | ASV | Firmicutes | Veillonellaceae | Veillonella | NA |
| Pramipexole | 145 | ASV | Firmicutes | Lachnospiraceae | Lachnoclostridium | NA |
| Pramipexole | 146 | ASV | Firmicutes | Christensenellaceae | NA | NA |
| Pramipexole | 147 | ASV | Firmicutes | Oscillospiraceae | Colidextribacter | massiliensis |
| Pramipexole | 148 | ASV | Bacteroidota | Rikenellaceae | Alistipes | putredinis |
| Pramipexole | 149 | ASV | Firmicutes | Lachnospiraceae | NA | NA |

| Ticker | DESeq2_baseMean | DESeq2_log2-FoldChange | DESeq2_lfcSE | DESeq2_stat | DESeq2_pvalue | DESeq2_padj | DESeq2 converged | ANCOM_W | ANCOM_detected_0.7 |
|---|---|---|---|---|---|---|---|---|---|
| 120 | 33,00466302 | -0,57097658 | 3,049915919 | -0,187210597 | 0,851495516 | 0,999805755 | YES | 0 | FALSE |
| 121 | 1,358877422 | -0,721183536 | 3,104071184 | -0,232334728 | 0,816278041 | 0,999805755 | YES | 0 | FALSE |
| 122 | 44,02602912 | 4,065334249 | 2,373701438 | 1,712656101 | 0,086775826 | 0,999805755 | YES | 0 | FALSE |
| 123 | 411,9532093 | 3,732936417 | 3,042487486 | 1,226935668 | 0,219846771 | 0,999805755 | YES | 0 | FALSE |
| 124 | 21,80343709 | 0,969749354 | 2,121093078 | 0,45719321 | 0,647532178 | 0,999805755 | YES | 0 | FALSE |
| 125 | 11,91508582 | 0,175477811 | 3,100008606 | 0,056605588 | 0,954859383 | 0,999805755 | YES | 0 | FALSE |
| 126 | 211,4863622 | 0,119129508 | 3,099404403 | 0,038436258 | 0,969339852 | 0,999805755 | YES | 0 | FALSE |
| 127 | 5,115312645 | -0,280914364 | 3,100907935 | -0,090591004 | 0,92781758 | 0,999805755 | YES | 0 | FALSE |
| 128 | 11,63613081 | 0,555849494 | 3,043815923 | 0,182616002 | 0,855099328 | 0,999805755 | YES | 0 | FALSE |
| 129 | 5,521707361 | 2,445753294 | 3,055174384 | 0,800528214 | 0,423404824 | 0,999805755 | YES | 0 | FALSE |
| 130 | 5,966074393 | -2,296515759 | 3,06446285 | -0,749402382 | 0,453614716 | 0,999805755 | YES | 0 | FALSE |
| 131 | 1,358877422 | -0,721183536 | 3,104071184 | -0,232334728 | 0,816278041 | 0,999805755 | YES | 0 | FALSE |
| 132 | 4,552239363 | -0,731036796 | 3,100792639 | -0,23575804 | 0,813620429 | 0,999805755 | YES | 0 | FALSE |
| 133 | 8,386122009 | 0,853558667 | 2,583543034 | 0,330382988 | 0,741110594 | 0,999805755 | YES | 0 | FALSE |
| 134 | 118,4631776 | 0,147466072 | 3,099410308 | 0,047578751 | 0,962051967 | 0,999805755 | YES | 0 | FALSE |
| 135 | 18,36670979 | 1,337484871 | 2,427444455 | 0,550984748 | 0,58164413 | 0,999805755 | YES | 0 | FALSE |
| 136 | 8,982149978 | 0,540556695 | 3,099980319 | 0,174374235 | 8,62E-01 | 1,00E+00 | YES | 0 | FALSE |
| 137 | 95,31389306 | 1,85290007 | 1,918711641 | 0,965700124 | 0,334194257 | 0,999805755 | YES | 0 | FALSE |
| 138 | 21,27744894 | 0,433663512 | 3,099752215 | 0,139902638 | 0,888736917 | 0,999805755 | YES | 0 | FALSE |
| 139 | 18,91393827 | -0,097202509 | 3,099768485 | -0,03135799 | 0,974984044 | 0,999805755 | YES | 0 | FALSE |
| 140 | 872,8219076 | 2,641752925 | 2,98688284 | 0,884451472 | 0,376452548 | 0,999805755 | YES | 0 | FALSE |
| 141 | 148,7567609 | 2,383465259 | 2,305656509 | 1,033746896 | 0,301254503 | 0,999805755 | YES | 0 | FALSE |
| 142 | 1,834484519 | -0,724778535 | 3,102870195 | -0,23358326 | 0,815308525 | 0,999805755 | YES | 0 | FALSE |
| 143 | 15,4038418 | -0,16327411 | 3,04306585 | -0,053654478 | 0,957210452 | 0,999805755 | YES | 0 | FALSE |
| 144 | 14,6533267 | 0,130637358 | 3,045330761 | 0,042897592 | 0,965783168 | 0,999805755 | YES | 0 | FALSE |
| 145 | 18,22978649 | 0,249654855 | 3,043147745 | 0,082038362 | 0,934616208 | 0,999805755 | YES | 0 | FALSE |
| 146 | 42,53257762 | -0,058892009 | 3,099545825 | -0,019000206 | 0,984840941 | 0,999805755 | YES | 0 | FALSE |
| 147 | 17,18629522 | -1,923549342 | 3,000936405 | -0,640983041 | 0,521533702 | 0,999805755 | YES | 0 | FALSE |
| 148 | 290,3412161 | 0,968202643 | 2,362514715 | 0,409818672 | 0,681938968 | 0,999805755 | YES | 0 | FALSE |
| 149 | 10,48431403 | -0,360134653 | 3,09987546 | -0,116177136 | 0,907512157 | 0,999805755 | YES | 0 | FALSE |

| Medication | Ticker | taxLevel | Phylum | Family | Genus | Species |
|---|---|---|---|---|---|---|
| Pramipexole | 150 | ASV | Firmicutes | Ruminococcaceae | Subdoligranulum | NA |
| Pramipexole | 151 | ASV | Firmicutes | Oscillospiraceae | UCG-002 | NA |
| Pramipexole | 152 | ASV | Firmicutes | Christensenellaceae | Christensenellaceae R-7 group | NA |
| Pramipexole | 153 | ASV | Firmicutes | Anaerovoracaceae | Family XIII AD3011 group | NA |
| Pramipexole | 154 | ASV | Proteobacteria | NA | NA | NA |
| Pramipexole | 155 | ASV | Proteobacteria | NA | NA | NA |
| Pramipexole | 156 | ASV | Firmicutes | Anaerovoracaceae | [Eubacterium] brachy group | NA |
| Pramipexole | 157 | ASV | Firmicutes | Lachnospiraceae | NA | NA |
| Pramipexole | 158 | ASV | Firmicutes | Peptostreptococcaceae | Intestinibacter | bartlettii |
| Pramipexole | 159 | ASV | Firmicutes | Lachnospiraceae | [Eubacterium] ventriosum group | NA |
| Pramipexole | 160 | ASV | Bacteroidota | Barnesiellaceae | NA | NA |
| Pramipexole | 161 | ASV | Firmicutes | Lachnospiraceae | Lachnoclostridium | NA |
| Pramipexole | 162 | ASV | Firmicutes | Monoglobaceae | Monoglobus | pectinilyticus |
| Pramipexole | 163 | ASV | Firmicutes | Lachnospiraceae | Anaerostipes | hadrus |
| Pramipexole | 164 | ASV | Firmicutes | Christensenellaceae | Christensenellaceae R-7 group | NA |
| Pramipexole | 165 | ASV | Firmicutes | Lachnospiraceae | Agathobacter | NA |
| Pramipexole | 166 | ASV | Firmicutes | NA | NA | NA |
| Pramipexole | 167 | ASV | Firmicutes | Lachnospiraceae | Marvinbryantia | NA |
| Pramipexole | 168 | ASV | Firmicutes | Lachnospiraceae | Blautia | obeum |
| Pramipexole | 169 | ASV | Firmicutes | Ruminococcaceae | NA | NA |
| Pramipexole | 170 | ASV | Bacteroidota | Rikenellaceae | Alistipes | NA |
| Pramipexole | 171 | ASV | Bacteroidota | Rikenellaceae | Alistipes | NA |
| Pramipexole | 172 | ASV | Firmicutes | Lachnospiraceae | [Eubacterium] hallii group | NA |
| Pramipexole | 173 | ASV | Firmicutes | UCG-010 | NA | NA |
| Pramipexole | 174 | ASV | Firmicutes | NA | NA | NA |
| Pramipexole | 175 | ASV | Bacteroidota | Barnesiellaceae | Coprobacter | fastidiosus |
| Pramipexole | 176 | ASV | Firmicutes | Clostridiaceae | Clostridium sensu stricto 1 | NA |
| Pramipexole | 177 | ASV | Firmicutes | Ruminococcaceae | DTU089 | NA |
| Pramipexole | 178 | ASV | Bacteroidota | Tannerellaceae | Parabacteroides | merdae |
| Pramipexole | 179 | ASV | Bacteroidota | Rikenellaceae | Alistipes | indistinctus |

| Ticker | DESeq2_baseMean | DESeq2_log2-FoldChange | DESeq2_lfcSE | DESeq2_stat | DESeq2_pvalue | DESeq2_padj | DESeq2 converged | ANCOM_W | ANCOM_detected_0.7 |
|---|---|---|---|---|---|---|---|---|---|
| 150 | 13,63571058 | 0,446608864 | 3,099755923 | 0,144078719 | 0,885438309 | 0,999805755 | YES | 0 | FALSE |
| 151 | 90,69184796 | 0,389256096 | 3,046390167 | 0,127776179 | 0,898326102 | 0,999805755 | YES | 0 | FALSE |
| 152 | 0,339719355 | -0,683103079 | 3,117155922 | -0,219143057 | 0,826538611 | 0,999805755 | YES | 0 | FALSE |
| 153 | 6,284213504 | 0,060751041 | 3,100144203 | 0,019596198 | 0,984365496 | 0,999805755 | YES | 0 | FALSE |
| 154 | 0,560098632 | 1,013393343 | 3,118901106 | 0,324919999 | 0,745241621 | 0,999805755 | YES | 0 | FALSE |
| 155 | 2,460439192 | -0,043818953 | 3,102437923 | -0,014124039 | 0,988731022 | 0,999805755 | YES | 0 | FALSE |
| 156 | 17,56213136 | 4,558612912 | 2,54445087 | 1,79159007 | 0,073198655 | 0,999805755 | YES | 37 | FALSE |
| 157 | 21,61991235 | -1,716778455 | 2,996251298 | -0,572975456 | 0,566661312 | 0,999805755 | YES | 0 | FALSE |
| 158 | 42,4840573 | -0,627536876 | 2,275248883 | -0,275810212 | 0,78269384 | 0,999805755 | YES | 0 | FALSE |
| 159 | 42,81919195 | -0,003091729 | 3,099563078 | -0,000997473 | 0,999204132 | 0,999805755 | YES | 0 | FALSE |
| 160 | 2,947201643 | 0,483025516 | 3,101180101 | 0,15575539 | 0,876225835 | 0,999805755 | YES | 0 | FALSE |
| 161 | 38,6579463 | -0,619831036 | 2,581808947 | -0,24007626 | 0,810271138 | 0,999805755 | YES | 0 | FALSE |
| 162 | 52,2071447 | 0,740984324 | 3,042127073 | 0,243574415 | 0,807560445 | 0,999805755 | YES | 0 | FALSE |
| 163 | 167,2686241 | -1,153093708 | 1,890705645 | -0,60987479 | 0,541944754 | 0,999805755 | YES | 0 | FALSE |
| 164 | 40,84893866 | 1,27980295 | 2,135952943 | 0,599171885 | 0,549058269 | 0,999805755 | YES | 0 | FALSE |
| 165 | 487,2873051 | 10,31243599 | 2,990991384 | 3,447832061 | 0,000565105 | 0,171792007 | YES | 0 | FALSE |
| 166 | 5,733651328 | 0,46577509 | 3,100792276 | 0,150211639 | 0,880597643 | 0,999805755 | YES | 0 | FALSE |
| 167 | 36,87430144 | 0,95213481 | 3,042465263 | 0,312948457 | 0,754319822 | 0,999805755 | YES | 0 | FALSE |
| 168 | 154,2226588 | 0,14685083 | 2,580719443 | 0,056903059 | 0,954622418 | 0,999805755 | YES | 0 | FALSE |
| 169 | 28,99652836 | 1,036666456 | 2,166134737 | 0,478578935 | 0,632238207 | 0,999805755 | YES | 0 | FALSE |
| 170 | 24,16993243 | 2,619975497 | 3,001392609 | 0,872919953 | 0,38270671 | 0,999805755 | YES | 0 | FALSE |
| 171 | 1,305681688 | 0,978611848 | 3,10482166 | 0,315191001 | 0,752616638 | 0,999805755 | YES | 0 | FALSE |
| 172 | 75,61948249 | 0,286637064 | 2,720397854 | 0,105365862 | 0,916085504 | 0,999805755 | YES | 0 | FALSE |
| 173 | 1,766540648 | -0,724381209 | 3,103002777 | -0,233445234 | 0,815415692 | 0,999805755 | YES | 0 | FALSE |
| 174 | 9,851861308 | -0,733353624 | 3,100028964 | -0,236563475 | 0,812995459 | 0,999805755 | YES | 0 | FALSE |
| 175 | 0,824641066 | 0,969067109 | 3,107880647 | 0,311809628 | 0,755185206 | 0,999805755 | YES | 0 | FALSE |
| 176 | 6,150366803 | 0,276716506 | 3,046506871 | 0,090830751 | 0,927627076 | 0,999805755 | YES | 0 | FALSE |
| 177 | 37,23194595 | 2,08284184 | 2,362510908 | 0,881622105 | 0,377981197 | 0,999805755 | YES | 0 | FALSE |
| 178 | 401,239006 | 0,082507455 | 2,261133779 | 0,036489418 | 0,970892116 | 0,999805755 | YES | 0 | FALSE |
| 179 | 21,25255595 | -0,90593196 | 2,568818306 | -0,352664865 | 0,724339703 | 0,999805755 | YES | 0 | FALSE |

| Medication | Ticker | taxLevel | Phylum | Family | Genus | Species |
|---|---|---|---|---|---|---|
| Pramipexole | 180 | ASV | Firmicutes | Ruminococcaceae | Ruminococcus | bromii |
| Pramipexole | 181 | ASV | Firmicutes | Erysipelotrichaceae | Holdemanella | NA |
| Pramipexole | 182 | ASV | Firmicutes | Oscillospiraceae | Colidextribacter | NA |
| Pramipexole | 183 | ASV | Firmicutes | NA | NA | NA |
| Pramipexole | 184 | ASV | Firmicutes | Ruminococcaceae | Ruminococcus | bicirculans |
| Pramipexole | 185 | ASV | Firmicutes | Oscillospiraceae | Oscillibacter | NA |
| Pramipexole | 186 | ASV | Firmicutes | Oscillospiraceae | UCG-003 | NA |
| Pramipexole | 187 | ASV | Firmicutes | Oscillospiraceae | Oscillospira | NA |
| Pramipexole | 188 | ASV | Actinobacteriota | Bifidobacteriaceae | Bifidobacterium | NA |
| Pramipexole | 189 | ASV | Firmicutes | Lachnospiraceae | Lachnospiraceae FCS020 group | bacterium |
| Pramipexole | 190 | ASV | Firmicutes | Christensenellaceae | Christensenellaceae R-7 group | NA |
| Pramipexole | 191 | ASV | Firmicutes | Lactobacillaceae | Lactobacillus | NA |
| Pramipexole | 192 | ASV | Firmicutes | Lachnospiraceae | Blautia | faecis |
| Pramipexole | 193 | ASV | Bacteroidota | Marinifilaceae | Butyricimonas | virosa |
| Pramipexole | 194 | ASV | Firmicutes | UCG-010 | NA | NA |
| Pramipexole | 195 | ASV | Firmicutes | Christensenellaceae | Christensenellaceae R-7 group | NA |
| Pramipexole | 196 | ASV | Bacteroidota | Bacteroidaceae | Bacteroides | NA |
| Pramipexole | 197 | ASV | Firmicutes | Veillonellaceae | Veillonella | NA |
| Pramipexole | 198 | ASV | Firmicutes | Erysipelatoclostridiaceae | Coprobacillus | cateniformis |
| Pramipexole | 199 | ASV | Firmicutes | Oscillospiraceae | UCG-005 | NA |
| Pramipexole | 200 | ASV | Firmicutes | Lachnospiraceae | Coprococcus | comes |
| Pramipexole | 201 | ASV | Actinobacteriota | Actinomycetaceae | Actinomyces | odontolyticus |
| Pramipexole | 202 | ASV | Firmicutes | Lachnospiraceae | NA | NA |
| Pramipexole | 203 | ASV | Bacteroidota | Bacteroidaceae | Bacteroides | NA |
| Pramipexole | 204 | ASV | Bacteroidota | Rikenellaceae | Alistipes | NA |
| Pramipexole | 205 | ASV | Firmicutes | Christensenellaceae | Christensenellaceae R-7 group | NA |
| Pramipexole | 206 | ASV | Firmicutes | Oscillospiraceae | Intestinimonas | massiliensis |
| Pramipexole | 207 | ASV | Bacteroidota | Bacteroidaceae | Bacteroides | caccae |
| Pramipexole | 208 | ASV | Firmicutes | UCG-010 | NA | NA |
| Pramipexole | 209 | ASV | Firmicutes | Lachnospiraceae | [Eubacterium] xylanophilum group | NA |

| Ticker | DESeq2_baseMean | DESeq2_log2-FoldChange | DESeq2_lfcSE | DESeq2_stat | DESeq2_pvalue | DESeq2_padj | DESeq2 converged | ANCOM_W | ANCOM_detected_0.7 |
|---|---|---|---|---|---|---|---|---|---|
| 180 | 829,5920807 | -0,818770408 | 2,986773951 | -0,274132031 | 0,783983159 | 0,999805755 | YES | 0 | FALSE |
| 181 | 74,37586975 | -1,078241678 | 3,042305571 | -0,354415969 | 0,723027169 | 0,999805755 | YES | 0 | FALSE |
| 182 | 7,860171156 | -2,13534238 | 3,046473431 | -0,700922699 | 0,483351258 | 0,999805755 | YES | 0 | FALSE |
| 183 | 10,63970657 | -0,926565388 | 2,317799467 | -0,399760808 | 0,6893327 | 0,999805755 | YES | 0 | FALSE |
| 184 | 224,0627518 | 1,205804957 | 2,645201112 | 0,455846231 | 0,648500557 | 0,999805755 | YES | 0 | FALSE |
| 185 | 128,8129072 | 1,582672789 | 2,6020148 | 0,608248957 | 5,43E-01 | 1,00E+00 | YES | 0 | FALSE |
| 186 | 0,747382582 | -0,710191596 | 3,107783117 | -0,228520321 | 0,81924176 | 0,999805755 | YES | 0 | FALSE |
| 187 | 52,05245337 | -0,141527881 | 1,913408841 | -0,073966357 | 0,941037155 | 0,999805755 | YES | 0 | FALSE |
| 188 | 96,71036381 | 1,078473896 | 3,099494097 | 0,347951589 | 0,727876541 | 0,999805755 | YES | 0 | FALSE |
| 189 | 8,430134655 | -0,243056384 | 3,100400136 | -0,078395166 | 0,937513719 | 0,999805755 | YES | 0 | FALSE |
| 190 | 40,54368899 | 0,885053874 | 2,490051817 | 0,355435926 | 0,722263038 | 0,999805755 | YES | 0 | FALSE |
| 191 | 1,242401646 | 0,730297613 | 3,107390266 | 0,235019599 | 0,814193522 | 0,999805755 | YES | 0 | FALSE |
| 192 | 268,157392 | 0,325698928 | 1,603612052 | 0,203103318 | 0,839054279 | 0,999805755 | YES | 0 | FALSE |
| 193 | 13,44926126 | 0,241541256 | 3,043170659 | 0,079371578 | 0,936737075 | 0,999805755 | YES | 0 | FALSE |
| 194 | 6,658499367 | -0,73239587 | 3,100344178 | -0,236230505 | 0,81325381 | 0,999805755 | YES | 0 | FALSE |
| 195 | 2,785698715 | -0,728337637 | 3,101686482 | -0,234819877 | 0,81434854 | 0,999805755 | YES | 0 | FALSE |
| 196 | 143,4182641 | -1,132349846 | 2,562326206 | -0,441922595 | 0,65854522 | 0,999805755 | YES | 0 | FALSE |
| 197 | 0,986165204 | -0,687202071 | 3,110438554 | -0,220934141 | 0,825143713 | 0,999805755 | YES | 0 | FALSE |
| 198 | 2,817523643 | 0,987750041 | 3,10193016 | 0,31843078 | 0,750158193 | 0,999805755 | YES | 0 | FALSE |
| 199 | 4,604245953 | 0,990895591 | 3,100943516 | 0,319546482 | 0,74931215 | 0,999805755 | YES | 0 | FALSE |
| 200 | 48,39972217 | -1,038731841 | 3,042699065 | -0,341385007 | 0,732813761 | 0,999805755 | YES | 0 | FALSE |
| 201 | 1,024744735 | 0,226031468 | 3,106671096 | 0,072756806 | 0,941999643 | 0,999805755 | YES | 0 | FALSE |
| 202 | 4,865471315 | 1,023523402 | 3,101747174 | 0,329982859 | 0,741412914 | 0,999805755 | YES | 0 | FALSE |
| 203 | 80,22945882 | 0,891258688 | 2,2579764 | 0,394715679 | 0,69305273 | 0,999805755 | YES | 0 | FALSE |
| 204 | 331,7576695 | 2,88522749 | 2,53489459 | 1,138204129 | 0,255035255 | 0,999805755 | YES | 0 | FALSE |
| 205 | 7,038863537 | -1,214659059 | 3,047799759 | -0,398536372 | 0,690234853 | 0,999805755 | YES | 0 | FALSE |
| 206 | 5,74610761 | 0,744015682 | 3,101133547 | 0,23991733 | 0,810394345 | 0,999805755 | YES | 0 | FALSE |
| 207 | 536,4100119 | 0,763389677 | 1,956118266 | 0,390257425 | 0,696346202 | 0,999805755 | YES | 0 | FALSE |
| 208 | 2,310091617 | -0,726918549 | 3,102157927 | -0,234326739 | 0,814731329 | 0,999805755 | YES | 0 | FALSE |
| 209 | 3,252522719 | -0,644704295 | 3,102229211 | -0,207819684 | 0,835369762 | 0,999805755 | YES | 0 | FALSE |

| Medication | Ticker | taxLevel | Phylum | Family | Genus | Species |
|---|---|---|---|---|---|---|
| Pramipexole | 210 | ASV | Firmicutes | Lachnospiraceae | Howardella | ureilytica |
| Pramipexole | 211 | ASV | Bacteroidota | Bacteroidaceae | Bacteroides | NA |
| Pramipexole | 212 | ASV | Firmicutes | Lachnospiraceae | Blautia | NA |
| Pramipexole | 213 | ASV | Bacteroidota | Prevotellaceae | Prevotella | NA |
| Pramipexole | 214 | ASV | Firmicutes | Erysipelatoclostridiaceae | Erysipelatoclostridium | NA |
| Pramipexole | 215 | ASV | Bacteroidota | Bacteroidaceae | Bacteroides | nordii |
| Pramipexole | 216 | ASV | Firmicutes | Oscillospiraceae | Colidextribacter | NA |
| Pramipexole | 217 | ASV | Firmicutes | Ruminococcaceae | NA | NA |
| Pramipexole | 218 | ASV | Firmicutes | Oscillospiraceae | UCG-005 | NA |
| Pramipexole | 219 | ASV | Proteobacteria | Oxalobacteraceae | Oxalobacter | formigenes |
| Pramipexole | 220 | ASV | Firmicutes | Lachnospiraceae | [Eubacterium] ventriosum group | NA |
| Pramipexole | 221 | ASV | Firmicutes | UCG-010 | NA | NA |
| Pramipexole | 222 | ASV | Firmicutes | Lachnospiraceae | NA | NA |
| Pramipexole | 223 | ASV | Firmicutes | Lachnospiraceae | [Ruminococcus] torques group | NA |
| Pramipexole | 224 | ASV | Firmicutes | Veillonellaceae | Allisonella | histaminiformans |
| Pramipexole | 225 | ASV | Firmicutes | Ruminococcaceae | Negativibacillus | massiliensis |
| Pramipexole | 226 | ASV | Firmicutes | Anaerovoracaceae | Family XIII UCG-001 | NA |
| Pramipexole | 227 | ASV | Firmicutes | Erysipelatoclostridiaceae | Asteroleplasma | NA |
| Pramipexole | 228 | ASV | Bacteroidota | Bacteroidaceae | Bacteroides | NA |
| Pramipexole | 229 | ASV | Proteobacteria | Sutterellaceae | Sutterella | NA |
| Pramipexole | 230 | ASV | Firmicutes | Lachnospiraceae | NA | NA |
| Pramipexole | 231 | ASV | Firmicutes | Monoglobaceae | Monoglobus | NA |
| Pramipexole | 232 | ASV | Bacteroidota | Bacteroidaceae | Bacteroides | massiliensis |
| Pramipexole | 233 | ASV | Firmicutes | Lachnospiraceae | Roseburia | inulinivorans |
| Pramipexole | 234 | ASV | Firmicutes | Erysipelatoclostridiaceae | Erysipelatoclostridium | NA |
| Pramipexole | 235 | ASV | Bacteroidota | Bacteroidaceae | Bacteroides | NA |
| Pramipexole | 236 | ASV | Firmicutes | [Eubacterium] coprostanoligenes group | NA | NA |
| Pramipexole | 237 | ASV | Firmicutes | Oscillospiraceae | Oscillibacter | NA |
| Pramipexole | 238 | ASV | Firmicutes | Ruminococcaceae | Faecalibacterium | NA |
| Pramipexole | 239 | ASV | Firmicutes | Ruminococcaceae | [Eubacterium] siraeum group | NA |

| Ticker | DESeq2_baseMean | DESeq2_log2-FoldChange | DESeq2_lfcSE | DESeq2_stat | DESeq2_pvalue | DESeq2_padj | DESeq2 converged | ANCOM_W | ANCOM_detected_0.7 |
|---|---|---|---|---|---|---|---|---|---|
| 210 | 0,747382582 | -0,710191596 | 3,107783117 | -0,228520321 | 0,81924176 | 0,999805755 | YES | 0 | FALSE |
| 211 | 1273,945506 | -0,075486758 | 3,04148545 | -0,024819043 | 0,980199302 | 0,999805755 | YES | 0 | FALSE |
| 212 | 1308,754837 | -0,965767576 | 1,591367952 | -0,606878865 | 0,543931305 | 0,999805755 | YES | 0 | FALSE |
| 213 | 1,030801333 | 0,974200691 | 3,106230612 | 0,313627935 | 0,753803641 | 0,999805755 | YES | 0 | FALSE |
| 214 | 0,848613936 | -0,928506305 | 3,114183403 | -0,298154021 | 0,765585612 | 0,999805755 | YES | 0 | FALSE |
| 215 | 1,931225187 | -0,05024095 | 3,103229632 | -0,016189891 | 0,9870829 | 0,999805755 | YES | 0 | FALSE |
| 216 | 68,95655279 | 0,208553839 | 3,099478933 | 0,067286742 | 0,946353431 | 0,999805755 | YES | 0 | FALSE |
| 217 | 6,352552955 | 2,035538615 | 3,012437089 | 0,675711577 | 0,499223784 | 0,999805755 | YES | 0 | FALSE |
| 218 | 21,20583703 | -0,151127592 | 3,099603905 | -0,048757066 | 0,961112898 | 0,999805755 | YES | 0 | FALSE |
| 219 | 12,25359827 | 0,728220359 | 2,60494763 | 0,279552783 | 0,779820637 | 0,999805755 | YES | 0 | FALSE |
| 220 | 4,646461028 | -0,646990924 | 3,101377404 | -0,208614057 | 0,834749536 | 0,999805755 | YES | 0 | FALSE |
| 221 | 4,763700512 | 0,584350546 | 3,057428876 | 0,191124821 | 0,848427802 | 0,999805755 | YES | 0 | FALSE |
| 222 | 13,63473783 | -0,056093687 | 3,099920141 | -0,018095204 | 0,985562904 | 0,999805755 | YES | 0 | FALSE |
| 223 | 603,4481863 | 0,903653072 | 1,903300454 | 0,474782145 | 0,634942261 | 0,999805755 | YES | 0 | FALSE |
| 224 | 3,329249684 | -0,729469095 | 3,101311261 | -0,235213119 | 0,814043324 | 0,999805755 | YES | 0 | FALSE |
| 225 | 30,8501947 | -0,675292771 | 3,043765984 | -0,221860936 | 0,824422141 | 0,999805755 | YES | 0 | FALSE |
| 226 | 2,680083465 | 0,987337659 | 3,10205987 | 0,318284527 | 0,75026912 | 0,999805755 | YES | 0 | FALSE |
| 227 | 0,549760711 | 0,956702015 | 3,111901783 | 0,30743323 | 0,758513638 | 0,999805755 | YES | 0 | FALSE |
| 228 | 415,0059752 | -0,355461933 | 3,041504572 | -0,116870425 | 0,906962735 | 0,999805755 | YES | 0 | FALSE |
| 229 | 51,7567803 | 0,331844005 | 3,099467069 | 0,107064859 | 0,914737525 | 0,999805755 | YES | 0 | FALSE |
| 230 | 3,12541807 | -0,729090313 | 3,10143681 | -0,235081466 | 0,814145503 | 0,999805755 | YES | 0 | FALSE |
| 231 | 132,3023306 | 0,164462452 | 1,990899376 | 0,082607114 | 0,934163944 | 0,999805755 | YES | 0 | FALSE |
| 232 | 207,8403903 | -0,509399166 | 3,099417525 | -0,164353193 | 0,869453109 | 0,999805755 | YES | 0 | FALSE |
| 233 | 127,5742219 | -2,097494968 | 2,654643677 | -0,790122978 | 0,429455952 | 0,999805755 | YES | 0 | FALSE |
| 234 | 58,45840405 | 0,887718241 | 2,528057331 | 0,351146404 | 0,725478516 | 0,999805755 | YES | 0 | FALSE |
| 235 | 174,7509689 | 0,234357604 | 3,041657007 | 0,07704932 | 0,93858431 | 0,999805755 | YES | 0 | FALSE |
| 236 | 59,57505941 | 0,40361216 | 3,099504381 | 0,13021829 | 0,896393725 | 0,999805755 | YES | 0 | FALSE |
| 237 | 2,785698715 | -0,728337637 | 3,101686482 | -0,234819877 | 0,81434854 | 0,999805755 | YES | 0 | FALSE |
| 238 | 1287,39372 | 3,048797391 | 2,158301443 | 1,412591092 | 0,15777599 | 0,999805755 | YES | 0 | FALSE |
| 239 | 277,2992052 | 0,357321498 | 2,280088202 | 0,156713893 | 0,875470337 | 0,999805755 | YES | 0 | FALSE |

| Medication | Ticker | taxLevel | Phylum | Family | Genus | Species |
|---|---|---|---|---|---|---|
| Pramipexole | 240 | ASV | Firmicutes | Lachnospiraceae | NA | NA |
| Pramipexole | 241 | ASV | Firmicutes | Erysipelatoclostridiaceae | Catenibacterium | mitsuokai |
| Pramipexole | 242 | ASV | Firmicutes | Lachnospiraceae | Lachnospiraceae UCG-010 | NA |
| Pramipexole | 243 | ASV | Firmicutes | Oscillospiraceae | Oscillibacter | NA |
| Pramipexole | 244 | ASV | Firmicutes | NA | NA | NA |
| Pramipexole | 245 | ASV | Actinobacteriota | Eggerthellaceae | NA | NA |
| Pramipexole | 246 | ASV | Firmicutes | Ruminococcaceae | Angelakisella | NA |
| Pramipexole | 247 | ASV | Firmicutes | Oscillospiraceae | UCG-002 | NA |
| Pramipexole | 248 | ASV | Firmicutes | Ruminococcaceae | Incertae Sedis | NA |
| Pramipexole | 249 | ASV | Firmicutes | Oscillospiraceae | Flavonifractor | NA |
| Pramipexole | 250 | ASV | Firmicutes | Butyricicoccaceae | Butyricicoccus | NA |
| Pramipexole | 251 | ASV | Firmicutes | Lachnospiraceae | NA | NA |
| Pramipexole | 252 | ASV | Firmicutes | Erysipelotrichaceae | Holdemania | filiformis |
| Pramipexole | 253 | ASV | Firmicutes | Ruminococcaceae | Faecalibacterium | NA |
| Pramipexole | 254 | ASV | Firmicutes | Ruminococcaceae | DTU089 | NA |
| Pramipexole | 255 | ASV | Firmicutes | Christensenellaceae | NA | NA |
| Pramipexole | 256 | ASV | Firmicutes | Lachnospiraceae | Agathobacter | NA |
| Pramipexole | 257 | ASV | Firmicutes | Lachnospiraceae | Roseburia | intestinalis |
| Pramipexole | 258 | ASV | Firmicutes | Lachnospiraceae | Lachnoclostridium | NA |
| Pramipexole | 259 | ASV | Firmicutes | Hydrogenoanaerobacterium | NA | NA |
| Pramipexole | 260 | ASV | Firmicutes | Lachnospiraceae | NA | NA |
| Pramipexole | 261 | ASV | Firmicutes | [Eubacterium] coprostanoligenes group | NA | NA |
| Pramipexole | 262 | ASV | Proteobacteria | Sutterellaceae | Parasutterella | excrementihominis |
| Pramipexole | 263 | ASV | Firmicutes | Ruminococcaceae | Angelakisella | NA |
| Pramipexole | 264 | ASV | Firmicutes | Oscillospiraceae | UCG-005 | NA |
| Pramipexole | 265 | ASV | Firmicutes | Ruminococcaceae | NA | NA |
| Pramipexole | 266 | ASV | Firmicutes | Lachnospiraceae | Coprococcus | catus |
| Pramipexole | 267 | ASV | Firmicutes | Lachnospiraceae | [Ruminococcus] gauvreauii group | NA |
| Pramipexole | 268 | ASV | Firmicutes | Ruminococcaceae | NA | NA |
| Pramipexole | 269 | ASV | Firmicutes | NA | NA | NA |

| Ticker | DESeq2_baseMean | DESeq2_log2-FoldChange | DESeq2_lfcSE | DESeq2_stat | DESeq2_pvalue | DESeq2_padj | DESeq2 converged | ANCOM_W | ANCOM_detected_0.7 |
|---|---|---|---|---|---|---|---|---|---|
| 240 | 11,96956381 | -0,188718793 | 3,10011313 | -0,060874808 | 0,951458912 | 0,999805755 | YES | 0 | FALSE |
| 241 | 4,280463879 | -0,730765144 | 3,100882444 | -0,235663608 | 0,813693711 | 0,999805755 | YES | 0 | FALSE |
| 242 | 32,94686015 | -0,130538151 | 3,042409371 | -0,042906176 | 0,965776326 | 0,999805755 | YES | 0 | FALSE |
| 243 | 7,40906732 | -0,696042595 | 3,100690338 | -0,22447988 | 0,822383921 | 0,999805755 | YES | 0 | FALSE |
| 244 | 3,625895907 | -0,969499478 | 3,103074896 | -0,312431865 | 0,754712336 | 0,999805755 | YES | 0 | FALSE |
| 245 | 177,8807931 | 1,109276943 | 2,250774469 | 0,492842334 | 0,622123996 | 0,999805755 | YES | 0 | FALSE |
| 246 | 35,12840033 | 3,145097772 | 2,402962567 | 1,308841767 | 0,190587956 | 0,999805755 | YES | 0 | FALSE |
| 247 | 82,02825286 | 0,006694823 | 3,041765069 | 0,002200966 | 0,998243884 | 0,999805755 | YES | 0 | FALSE |
| 248 | 14,33377851 | 0,578160532 | 3,099759283 | 0,186517881 | 0,852038658 | 0,999805755 | YES | 0 | FALSE |
| 249 | 5,603909884 | -1,428552923 | 3,052515881 | -0,467991971 | 0,639790332 | 0,999805755 | YES | 0 | FALSE |
| 250 | 132,1223433 | 0,077290326 | 2,990670414 | 0,025843813 | 0,979381916 | 0,999805755 | YES | 0 | FALSE |
| 251 | 4,054559429 | 1,021972781 | 3,102216202 | 0,329433126 | 0,741828333 | 0,999805755 | YES | 0 | FALSE |
| 252 | 8,314560698 | 0,607654602 | 2,304351542 | 0,263698742 | 0,792012065 | 0,999805755 | YES | 0 | FALSE |
| 253 | 6,047004527 | -0,732098201 | 3,100442341 | -0,236127017 | 0,81333411 | 0,999805755 | YES | 0 | FALSE |
| 254 | 20,82580503 | 0,667213581 | 3,09980704 | 0,215243585 | 0,829577411 | 0,999805755 | YES | 0 | FALSE |
| 255 | 3,545500781 | -0,156352123 | 3,100772108 | -0,05042361 | 0,959784822 | 0,999805755 | YES | 0 | FALSE |
| 256 | 3345,312936 | 1,128712752 | 1,954013466 | 0,577638164 | 0,563508435 | 0,999805755 | YES | 0 | FALSE |
| 257 | 39,39869301 | -1,079315672 | 3,04244571 | -0,354752648 | 0,722774905 | 0,999805755 | YES | 0 | FALSE |
| 258 | 61,12933745 | 0,745170289 | 2,568647217 | 0,290102231 | 0,771738029 | 0,999805755 | YES | 0 | FALSE |
| 259 | 2,207068988 | -0,641137047 | 3,103563492 | -0,206580935 | 0,836337151 | 0,999805755 | YES | 0 | FALSE |
| 260 | 1,030801333 | 0,974200691 | 3,106230612 | 0,313627935 | 0,753803641 | 0,999805755 | YES | 0 | FALSE |
| 261 | 12,14282493 | 0,204173907 | 3,045820433 | 0,067034125 | 0,946554537 | 0,999805755 | YES | 0 | FALSE |
| 262 | 162,2978453 | -0,224186922 | 2,273915357 | -0,098590707 | 0,921463249 | 0,999805755 | YES | 0 | FALSE |
| 263 | 10,34678682 | -0,285128263 | 3,100132103 | -0,09197294 | 0,926719539 | 0,999805755 | YES | 0 | FALSE |
| 264 | 153,5235801 | -1,058387413 | 3,042064909 | -0,347917433 | 0,727902193 | 0,999805755 | YES | 0 | FALSE |
| 265 | 3,942889868 | 2,461041696 | 3,063648885 | 0,803304095 | 0,421798991 | 0,999805755 | YES | 0 | FALSE |
| 266 | 15,47441742 | -0,754150774 | 3,043353002 | -0,247802596 | 0,804287146 | 0,999805755 | YES | 0 | FALSE |
| 267 | 6,794697969 | 0,23227117 | 3,04958108 | 0,076164943 | 0,939287873 | 0,999805755 | YES | 0 | FALSE |
| 268 | 2,785698715 | -0,728337637 | 3,101686482 | -0,234819877 | 0,81434854 | 0,999805755 | YES | 0 | FALSE |
| 269 | 3,601025168 | -0,729908749 | 3,101165804 | -0,235365922 | 0,813924733 | 0,999805755 | YES | 0 | FALSE |

| Medication | Ticker | taxLevel | Phylum | Family | Genus | Species |
|---|---|---|---|---|---|---|
| Pramipexole | 270 | ASV | Firmicutes | Lachnospiraceae | Lachnospiraceae ND3007 group | NA |
| Pramipexole | 271 | ASV | Firmicutes | Lachnospiraceae | [Ruminococcus] torques group | NA |
| Pramipexole | 272 | ASV | Firmicutes | Lachnospiraceae | [Eubacterium] eligens group | NA |
| Pramipexole | 273 | ASV | Firmicutes | Lachnospiraceae | [Eubacterium] ruminantium group | NA |
| Pramipexole | 274 | ASV | Firmicutes | Ruminococcaceae | Faecalibacterium | prausnitzii |
| Pramipexole | 275 | ASV | Firmicutes | Acidaminococcaceae | Phascolarctobacterium | faecium |
| Pramipexole | 276 | ASV | Firmicutes | Peptococcaceae | NA | NA |
| Pramipexole | 277 | ASV | Desulfobacterota | Desulfovibrionaceae | NA | NA |
| Pramipexole | 278 | ASV | Firmicutes | Lachnospiraceae | Lachnospiraceae NK4A136 group | NA |
| Pramipexole | 279 | ASV | Firmicutes | Ruminococcaceae | NA | NA |
| Pramipexole | 280 | ASV | Firmicutes | Oscillospiraceae | Colidextribacter | NA |
| Pramipexole | 281 | ASV | Firmicutes | Lachnospiraceae | GCA-900066575 | NA |
| Pramipexole | 282 | ASV | Firmicutes | Christensenellaceae | Christensenellaceae R-7 group | NA |
| Pramipexole | 283 | ASV | Firmicutes | Lachnospiraceae | Lachnospiraceae NK4A136 group | NA |
| Pramipexole | 284 | ASV | Verrucomicrobiota | Victivallaceae | Victivallis | vadensis |
| Pramipexole | 285 | ASV | Bacteroidota | Bacteroidaceae | Bacteroides | NA |
| Pramipexole | 286 | ASV | Firmicutes | Lachnospiraceae | Coprococcus | NA |
| Pramipexole | 287 | ASV | Firmicutes | Lachnospiraceae | [Eubacterium] hallii group | NA |
| Pramipexole | 288 | ASV | Firmicutes | NA | NA | NA |
| Pramipexole | 289 | ASV | Proteobacteria | Sutterellaceae | Sutterella | wadsworthensis |
| Pramipexole | 290 | ASV | Firmicutes | NA | NA | NA |
| Pramipexole | 291 | ASV | Firmicutes | Lachnospiraceae | Dorea | formicigenerans |
| Pramipexole | 292 | ASV | Firmicutes | Lachnospiraceae | Shuttleworthia | NA |
| Pramipexole | 293 | ASV | Firmicutes | Lachnospiraceae | Blautia | NA |
| Pramipexole | 294 | ASV | Bacteroidota | Barnesiellaceae | Coprobacter | secundus |
| Pramipexole | 295 | ASV | Bacteroidota | Bacteroidaceae | Bacteroides | NA |
| Pramipexole | 296 | ASV | Firmicutes | Lachnospiraceae | [Eubacterium] eligens group | NA |
| Pramipexole | 297 | ASV | Firmicutes | Erysipelotrichaceae | Turicibacter | sanguinis |
| Pramipexole | 298 | ASV | Firmicutes | Oscillospiraceae | NK4A214 group | NA |
| Pramipexole | 299 | ASV | Firmicutes | UCG-010 | NA | NA |

| Ticker | DESeq2_baseMean | DESeq2_log2-FoldChange | DESeq2_lfcSE | DESeq2_stat | DESeq2_pvalue | DESeq2_padj | DESeq2 converged | ANCOM_W | ANCOM_detected_0.7 |
|---|---|---|---|---|---|---|---|---|---|
| 270 | 14,46116544 | -0,453557741 | 2,991959722 | -0,151592195 | 0,87950859 | 0,999805755 | YES | 0 | FALSE |
| 271 | 302,2008219 | 0,794563163 | 2,607264017 | 0,30474979 | 0,760556725 | 0,999805755 | YES | 0 | FALSE |
| 272 | 140,623728 | -0,484292473 | 2,520688372 | -0,192127071 | 0,847642671 | 0,999805755 | YES | 0 | FALSE |
| 273 | 18,19762785 | 0,811136875 | 3,099721016 | 0,261680606 | 0,793567694 | 0,999805755 | YES | 0 | FALSE |
| 274 | 462,9451701 | 0,355597321 | 2,714978831 | 0,130976093 | 0,895794219 | 0,999805755 | YES | 0 | FALSE |
| 275 | 6,109153174 | 0,618337987 | 3,10080675 | 0,199411971 | 0,841940497 | 0,999805755 | YES | 0 | FALSE |
| 276 | 26,27030935 | -0,735917426 | 2,276518981 | -0,323264349 | 0,746495051 | 0,999805755 | YES | 0 | FALSE |
| 277 | 0,46674886 | 1,002277977 | 3,122393598 | 0,32099668 | 0,748212907 | 0,999805755 | YES | 0 | FALSE |
| 278 | 14,8117639 | -0,734025659 | 3,099808362 | -0,236797109 | 0,812814194 | 0,999805755 | YES | 0 | FALSE |
| 279 | 1,306896808 | 1,048492719 | 3,108215345 | 0,337329497 | 0,735868526 | 0,999805755 | YES | 0 | FALSE |
| 280 | 8,789093928 | 0,698997977 | 3,044471922 | 0,229595803 | 0,818405868 | 0,999805755 | YES | 0 | FALSE |
| 281 | 44,77586329 | 2,60299378 | 2,36535497 | 1,100466447 | 0,271128941 | 0,999805755 | YES | 0 | FALSE |
| 282 | 0,407663227 | -0,691064414 | 3,114367629 | -0,221895581 | 0,824395171 | 0,999805755 | YES | 0 | FALSE |
| 283 | 41,34021869 | 1,47912421 | 3,042729943 | 0,486117479 | 0,626883875 | 0,999805755 | YES | 0 | FALSE |
| 284 | 0,784639242 | -1,706525376 | 3,104731936 | -0,549653049 | 0,582557365 | 0,999805755 | YES | 0 | FALSE |
| 285 | 55,29048009 | -0,505296012 | 3,042738891 | -0,166066176 | 0,868104872 | 0,999805755 | YES | 0 | FALSE |
| 286 | 507,9557155 | 0,560796704 | 3,041491496 | 0,184382138 | 0,853713682 | 0,999805755 | YES | 0 | FALSE |
| 287 | 278,2596982 | 1,691605112 | 1,848755172 | 0,914996825 | 0,360193297 | 0,999805755 | YES | 0 | FALSE |
| 288 | 1,895833441 | -1,900862452 | 3,087724421 | -0,615619205 | 0,538145873 | 0,999805755 | YES | 0 | FALSE |
| 289 | 36,89237627 | 1,099934358 | 3,045943436 | 0,361114506 | 0,71801385 | 0,999805755 | YES | 0 | FALSE |
| 290 | 2,790106318 | 2,382843612 | 3,087206877 | 0,771844488 | 0,440206539 | 0,999805755 | YES | 0 | FALSE |
| 291 | 111,3930899 | 2,034421817 | 2,464278377 | 0,825564935 | 0,409050933 | 0,999805755 | YES | 0 | FALSE |
| 292 | 9,167173499 | -0,5429823 | 3,045111207 | -0,178312798 | 0,858477328 | 0,999805755 | YES | 0 | FALSE |
| 293 | 1,099521422 | 0,975501641 | 3,105814215 | 0,314088859 | 0,753453551 | 0,999805755 | YES | 0 | FALSE |
| 294 | 2,93437005 | 2,382137456 | 3,066999662 | 0,776699615 | 0,437336014 | 0,999805755 | YES | 0 | FALSE |
| 295 | 1524,367925 | 0,255670058 | 2,223502488 | 0,114985281 | 0,908456789 | 0,999805755 | YES | 0 | FALSE |
| 296 | 70,83354964 | -1,114845414 | 2,992310769 | -0,372570064 | 0,709468457 | 0,999805755 | YES | 0 | FALSE |
| 297 | 7,956798089 | 1,467149455 | 3,009047494 | 0,487579361 | 0,625847817 | 0,999805755 | YES | 0 | FALSE |
| 298 | 42,16748708 | 0,308900845 | 3,042028316 | 0,101544369 | 0,919118338 | 0,999805755 | YES | 0 | FALSE |
| 299 | 0,339719355 | -0,683103079 | 3,117155922 | -0,219143057 | 0,826538611 | 0,999805755 | YES | 0 | FALSE |

| Medication | Ticker | taxLevel | Phylum | Family | Genus | Species |
|---|---|---|---|---|---|---|
| Pramipexole | 300 | ASV | Bacteroidota | Rikenellaceae | Alistipes | shahii |
| Pramipexole | 301 | ASV | Firmicutes | Lachnospiraceae | CAG-56 | NA |
| Pramipexole | 302 | ASV | Firmicutes | Lachnospiraceae | [Ruminococcus] gnavus group | NA |
| Pramipexole | 303 | ASV | Firmicutes | Acidaminococcaceae | Phascolarctobacterium | succinatutens |
| Pramipexole | 304 | ASV | Bacteroidota | Bacteroidaceae | Bacteroides | plebeius |
| Pramipexole | 305 | Genus | Actinobacteriota | Bifidobacteriaceae | Bifidobacterium | NA |
| Pramipexole | 306 | Genus | Verrucomicrobiota | Akkermansiaceae | Akkermansia | NA |
| Pramipexole | 307 | Genus | Firmicutes | Lachnospiraceae | Lachnospira | NA |
| Pramipexole | 308 | Genus | Firmicutes | Oscillospiraceae | UCG-003 | NA |
| Pramipexole | 309 | Genus | Firmicutes | Veillonellaceae | Dialister | NA |
| Pramipexole | 310 | Genus | Firmicutes | Oscillospiraceae | NK4A214 group | NA |
| Pramipexole | 311 | Genus | Firmicutes | Lachnospiraceae | Fusicatenibacter | NA |
| Pramipexole | 312 | Genus | Firmicutes | Ruminococcaceae | Candidatus Soleaferrea | NA |
| Pramipexole | 313 | Genus | Actinobacteriota | Eggerthellaceae | Senegalimassilia | NA |
| Pramipexole | 314 | Genus | Actinobacteriota | Eggerthellaceae | Enterorhabdus | NA |
| Pramipexole | 315 | Genus | Firmicutes | Lachnospiraceae | Lachnoclostridium | NA |
| Pramipexole | 316 | Genus | Firmicutes | Peptostreptococcaceae | Terrisporobacter | NA |
| Pramipexole | 317 | Genus | Desulfobacterota | Desulfovibrionaceae | Bilophila | NA |
| Pramipexole | 318 | Genus | Firmicutes | Oscillospiraceae | Oscillibacter | NA |
| Pramipexole | 319 | Genus | Firmicutes | Christensenellaceae | Christensenellaceae R-7 group | NA |
| Pramipexole | 320 | Genus | Firmicutes | Erysipelatoclostridiaceae | UCG-004 | NA |
| Pramipexole | 321 | Genus | Firmicutes | Lachnospiraceae | [Eubacterium] ventriosum group | NA |
| Pramipexole | 322 | Genus | Firmicutes | Peptostreptococcaceae | Romboutsia | NA |
| Pramipexole | 323 | Genus | Desulfobacterota | Desulfovibrionaceae | Desulfovibrio | NA |
| Pramipexole | 324 | Genus | Firmicutes | Ruminococcaceae | Paludicola | NA |
| Pramipexole | 325 | Genus | Firmicutes | Oscillospiraceae | Intestinimonas | NA |
| Pramipexole | 326 | Genus | Actinobacteriota | Coriobacteriaceae | Collinsella | NA |
| Pramipexole | 327 | Genus | Proteobacteria | Pasteurellaceae | Haemophilus | NA |
| Pramipexole | 328 | Genus | Firmicutes | Erysipelatoclostridiaceae | Asteroleplasma | NA |
| Pramipexole | 329 | Genus | Firmicutes | Lachnospiraceae | Dorea | NA |

| Ticker | DESeq2_baseMean | DESeq2_log2-FoldChange | DESeq2_lfcSE | DESeq2_stat | DESeq2_pvalue | DESeq2_padj | DESeq2 converged | ANCOM_W | ANCOM_detected_0.7 |
|---|---|---|---|---|---|---|---|---|---|
| 300 | 65,95549915 | -0,195805869 | 2,631231231 | -0,074416063 | 0,940679328 | 0,999805755 | YES | 0 | FALSE |
| 301 | 97,32724809 | -1,329019478 | 3,042284094 | -0,436849234 | 0,66222069 | 0,999805755 | YES | 0 | FALSE |
| 302 | 2,792784839 | -2,24599434 | 3,073100969 | -0,730856019 | 0,464867102 | 0,999805755 | YES | 0 | FALSE |
| 303 | 30,98240522 | -0,734727472 | 3,099579213 | -0,237041037 | 0,812624954 | 0,999805755 | YES | 0 | FALSE |
| 304 | 99,52452745 | -2,810175313 | 3,043251107 | -0,923412237 | 0,355792417 | 0,999805755 | YES | 0 | FALSE |
| 305 | 1344,355805 | 2,075906804 | 0,634969733 | 3,269300404 | 0,001078138 | 0,127220251 | YES | 0 | FALSE |
| 306 | 1364,830344 | -1,853052843 | 1,774211955 | -1,044437131 | 0,296283192 | 0,998567161 | YES | 0 | FALSE |
| 307 | 177,5815805 | -1,855418696 | 1,179368311 | -1,573230922 | 0,115665358 | 0,998567161 | YES | 0 | FALSE |
| 308 | 77,4927846 | -0,798937652 | 2,374922701 | -0,33640575 | 0,736564914 | 0,998567161 | YES | 0 | FALSE |
| 309 | 1236,026384 | 0,975524526 | 2,276874509 | 0,428448964 | 0,668324283 | 0,998567161 | YES | 0 | FALSE |
| 310 | 307,3640933 | 0,510846692 | 1,316950445 | 0,387901226 | 0,698089131 | 0,998567161 | YES | 0 | FALSE |
| 311 | 899,3581077 | 2,879365121 | 1,793806475 | 1,605170436 | 0,108456302 | 0,998567161 | YES | 0 | FALSE |
| 312 | 1,731469796 | -1,618920159 | 3,052877151 | -0,530293254 | 0,595908623 | 0,998567161 | YES | 0 | FALSE |
| 313 | 0,423565529 | 0,724649898 | 3,117075581 | 0,232477487 | 0,816167171 | 0,998567161 | YES | 0 | FALSE |
| 314 | 9,98438168 | -0,183100816 | 2,297123699 | -0,079708731 | 0,936468915 | 0,998567161 | YES | 0 | FALSE |
| 315 | 308,8576131 | -0,612370924 | 0,729632679 | -0,839286591 | 0,401308506 | 0,998567161 | YES | 0 | FALSE |
| 316 | 0,735861495 | -0,919432729 | 3,110000803 | -0,295637457 | 0,767506965 | 0,998567161 | YES | 0 | FALSE |
| 317 | 98,93991685 | 0,53399798 | 1,223804083 | 0,436342702 | 0,662588104 | 0,998567161 | YES | 0 | FALSE |
| 318 | 409,7871718 | 1,139142712 | 0,580013995 | 1,963991769 | 0,049531046 | 0,998567161 | YES | 0 | FALSE |
| 319 | 286,0527492 | 0,089815544 | 1,331752252 | 0,067441631 | 0,946230128 | 0,998567161 | YES | 0 | FALSE |
| 320 | 0,14118851 | 0,640569765 | 3,145757306 | 0,203629747 | 0,838642845 | 0,998567161 | YES | 0 | FALSE |
| 321 | 111,1692141 | -0,295201379 | 1,371541069 | -0,21523335 | 0,829585391 | 0,998567161 | YES | 0 | FALSE |
| 322 | 33,54813725 | 3,262462818 | 2,019700085 | 1,615320434 | 0,106241321 | 0,998567161 | YES | 0 | FALSE |
| 323 | 91,9738297 | 0,274393657 | 2,078220048 | 0,132033014 | 0,894958179 | 0,998567161 | YES | 0 | FALSE |
| 324 | 6,285305421 | -0,052137283 | 3,002824442 | -0,017362748 | 0,986147228 | 0,998567161 | YES | 0 | FALSE |
| 325 | 170,7079586 | -0,429383514 | 0,970784172 | -0,442305846 | 0,658267901 | 0,998567161 | YES | 0 | FALSE |
| 326 | 807,8694594 | 1,151350074 | 0,701813903 | 1,640534718 | 0,100894036 | 0,998567161 | YES | 0 | FALSE |
| 327 | 15,46153561 | -1,167107902 | 3,029406578 | -0,385259579 | 0,700045106 | 0,998567161 | YES | 0 | FALSE |
| 328 | 37,71216728 | 0,560347731 | 3,099556357 | 0,180783205 | 0,856537747 | 0,998567161 | YES | 0 | FALSE |
| 329 | 677,4623792 | -0,70296082 | 0,737196746 | -0,953559311 | 0,34030676 | 0,998567161 | YES | 0 | FALSE |

| Medication | Ticker | taxLevel | Phylum | Family | Genus | Species |
|---|---|---|---|---|---|---|
| Pramipexole | 330 | Genus | Firmicutes | Erysipelatoclostridiaceae | Erysipelotrichaceae UCG-003 | NA |
| Pramipexole | 331 | Genus | Firmicutes | Ruminococcaceae | UBA1819 | NA |
| Pramipexole | 332 | Genus | Bacteroidota | Marinifilaceae | Odoribacter | NA |
| Pramipexole | 333 | Genus | Actinobacteriota | Eggerthellaceae | Adlercreutzia | NA |
| Pramipexole | 334 | Genus | Bacteroidota | Barnesiellaceae | Barnesiella | NA |
| Pramipexole | 335 | Genus | Bacteroidota | Prevotellaceae | Prevotellaceae NK3B31 group | NA |
| Pramipexole | 336 | Genus | Bacteroidota | Bacteroidaceae | Bacteroides | NA |
| Pramipexole | 337 | Genus | Bacteroidota | Prevotellaceae | Prevotella | NA |
| Pramipexole | 338 | Genus | Firmicutes | Lachnospiraceae | [Eubacterium] xylanophilum group | NA |
| Pramipexole | 339 | Genus | Firmicutes | Lachnospiraceae | Lachnospiraceae FCS020 group | NA |
| Pramipexole | 340 | Genus | Firmicutes | Lachnospiraceae | Lachnospiraceae UCG-010 | NA |
| Pramipexole | 341 | Genus | Firmicutes | Peptococcaceae | Peptococcus | NA |
| Pramipexole | 342 | Genus | Firmicutes | Lachnospiraceae | Lachnospiraceae UCG-004 | NA |
| Pramipexole | 343 | Genus | Firmicutes | Lachnospiraceae | Lachnospiraceae UCG-001 | NA |
| Pramipexole | 344 | Genus | Firmicutes | Oscillospiraceae | Flavonifractor | NA |
| Pramipexole | 345 | Genus | Firmicutes | Streptococcaceae | Streptococcus | NA |
| Pramipexole | 346 | Genus | Firmicutes | Acholeplasmataceae | Anaeroplasma | NA |
| Pramipexole | 347 | Genus | Bacteroidota | Prevotellaceae | Alloprevotella | NA |
| Pramipexole | 348 | Genus | Firmicutes | Lachnospiraceae | Tyzzerella | NA |
| Pramipexole | 349 | Genus | Euryarchaeota | Methanobacteriaceae | Methanosphaera | NA |
| Pramipexole | 350 | Genus | Firmicutes | Erysipelotrichaceae | Solobacterium | NA |
| Pramipexole | 351 | Genus | Firmicutes | Lachnospiraceae | Lachnospiraceae AC2044 group | NA |
| Pramipexole | 352 | Genus | Euryarchaeota | Methanobacteriaceae | Methanobrevibacter | NA |
| Pramipexole | 353 | Genus | Firmicutes | Ruminococcaceae | CAG-352 | NA |
| Pramipexole | 354 | Genus | Firmicutes | Clostridiaceae | Clostridium sensu stricto 1 | NA |
| Pramipexole | 355 | Genus | Actinobacteriota | Eggerthellaceae | Slackia | NA |
| Pramipexole | 356 | Genus | Firmicutes | Oscillospiraceae | UCG-002 | NA |
| Pramipexole | 357 | Genus | Firmicutes | Erysipelotrichaceae | Catenisphaera | NA |
| Pramipexole | 358 | Genus | Firmicutes | Anaerovoracaceae | Mogibacterium | NA |
| Pramipexole | 359 | Genus | Firmicutes | Ruminococcaceae | Negativibacillus | NA |

| Ticker | DESeq2_baseMean | DESeq2_log2-FoldChange | DESeq2_lfcSE | DESeq2_stat | DESeq2_pvalue | DESeq2_padj | DESeq2 converged | ANCOM_W | ANCOM_detected_0.7 |
|---|---|---|---|---|---|---|---|---|---|
| 330 | 241,6263661 | 0,302265405 | 1,677697708 | 0,180166786 | 0,857021632 | 0,998567161 | YES | 0 | FALSE |
| 331 | 12,83772834 | 0,279226645 | 2,147958876 | 0,129996271 | 0,896569377 | 0,998567161 | YES | 0 | FALSE |
| 332 | 24,07193252 | -0,408919821 | 1,038047904 | -0,393931552 | 0,693631575 | 0,998567161 | YES | 0 | FALSE |
| 333 | 11,47978911 | 2,169928891 | 2,990644031 | 0,725572441 | 0,468100921 | 0,998567161 | YES | 0 | FALSE |
| 334 | 259,4575144 | 1,023639988 | 1,204817107 | 0,84962272 | 0,395534876 | 0,998567161 | YES | 0 | FALSE |
| 335 | 231,8491692 | -0,201645977 | 3,099399092 | -0,065059701 | 0,948126467 | 0,998567161 | YES | 0 | FALSE |
| 336 | 7941,910738 | -0,055341247 | 0,536821693 | -0,103090557 | 0,9178911 | 0,998567161 | YES | 0 | FALSE |
| 337 | 1982,176061 | -1,766460292 | 1,435067154 | -1,230925178 | 0,21835085 | 0,998567161 | YES | 0 | FALSE |
| 338 | 57,28847948 | -2,387598392 | 1,745546444 | -1,367822896 | 0,171367519 | 0,998567161 | YES | 0 | FALSE |
| 339 | 43,80668629 | 0,047654211 | 0,83220938 | 0,057262285 | 0,954336263 | 0,998567161 | YES | 0 | FALSE |
| 340 | 71,03005293 | 0,725710934 | 0,970474508 | 0,747789796 | 0,454586958 | 0,998567161 | YES | 0 | FALSE |
| 341 | 10,76653394 | -0,59562815 | 3,047544832 | -0,195445246 | 0,845044398 | 0,998567161 | YES | 0 | FALSE |
| 342 | 89,92750812 | 0,246492895 | 2,240015248 | 0,110040722 | 0,91237708 | 0,998567161 | YES | 0 | FALSE |
| 343 | 29,79850509 | -0,629661812 | 3,042615949 | -0,206947516 | 0,836050847 | 0,998567161 | YES | 0 | FALSE |
| 344 | 161,6784889 | 0,435500559 | 1,035944641 | 0,420389799 | 0,674200719 | 0,998567161 | YES | 0 | FALSE |
| 345 | 531,702743 | 1,932785493 | 1,224594377 | 1,578306686 | 0,114495171 | 0,998567161 | YES | 0 | FALSE |
| 346 | 83,16336531 | 0,240933849 | 3,099442303 | 0,077734581 | 0,938039186 | 0,998567161 | YES | 0 | FALSE |
| 347 | 7,419443565 | -0,366024919 | 3,100424947 | -0,118056371 | 0,906022992 | 0,998567161 | YES | 0 | FALSE |
| 348 | 7,349290314 | 0,758838987 | 3,100781888 | 0,244725045 | 0,806669335 | 0,998567161 | YES | 0 | FALSE |
| 349 | 7,292472392 | 0,030094563 | 3,100166983 | 0,009707401 | 0,992254736 | 0,998567161 | YES | 0 | FALSE |
| 350 | 7,174649577 | -0,946341438 | 3,100501822 | -0,305222023 | 0,76019706 | 0,998567161 | YES | 0 | FALSE |
| 351 | 1,839653738 | -0,937090657 | 3,103729985 | -0,301924027 | 0,762709979 | 0,998567161 | YES | 0 | FALSE |
| 352 | 80,54366585 | 1,019536748 | 1,332545562 | 0,765104607 | 0,444209257 | 0,998567161 | YES | 0 | FALSE |
| 353 | 312,4986088 | 4,066695598 | 3,042512882 | 1,336623954 | 0,181345416 | 0,998567161 | YES | 0 | FALSE |
| 354 | 34,84101479 | -0,830901423 | 1,59003712 | -0,522567312 | 0,601275393 | 0,998567161 | YES | 0 | FALSE |
| 355 | 5,414722143 | 2,219107307 | 3,055487107 | 0,72626957 | 0,46767353 | 0,998567161 | YES | 0 | FALSE |
| 356 | 526,5545115 | 0,163093119 | 1,274496814 | 0,127966674 | 0,898175346 | 0,998567161 | YES | 0 | FALSE |
| 357 | 16,85469065 | 0,825832305 | 3,043602899 | 0,271333789 | 0,786134321 | 0,998567161 | YES | 0 | FALSE |
| 358 | 9,803946621 | 0,290969269 | 3,100001379 | 0,093861013 | 0,925219564 | 0,998567161 | YES | 0 | FALSE |
| 359 | 117,8970404 | 1,595678491 | 0,788503911 | 2,023678601 | 0,043003232 | 0,998567161 | YES | 0 | FALSE |

| Medication | Ticker | taxLevel | Phylum | Family | Genus | Species |
|---|---|---|---|---|---|---|
| Pramipexole | 360 | Genus | Firmicutes | Streptococcaceae | Lactococcus | NA |
| Pramipexole | 361 | Genus | Firmicutes | Ruminococcaceae | Subdoligranulum | NA |
| Pramipexole | 362 | Genus | Proteobacteria | Enterobacteriaceae | Escherichia-Shigella | NA |
| Pramipexole | 363 | Genus | Firmicutes | Lachnospiraceae | GCA-900066575 | NA |
| Pramipexole | 364 | Genus | Firmicutes | Veillonellaceae | Veillonella | NA |
| Pramipexole | 365 | Genus | Firmicutes | Oscillospiraceae | Colidextribacter | NA |
| Pramipexole | 366 | Genus | Bacteroidota | Rikenellaceae | Alistipes | NA |
| Pramipexole | 367 | Genus | Firmicutes | Anaerovoracaceae | Family XIII UCG-001 | NA |
| Pramipexole | 368 | Genus | Firmicutes | Anaerovoracaceae | Family XIII AD3011 group | NA |
| Pramipexole | 369 | Genus | Firmicutes | Anaerovoracaceae | [Eubacterium] brachy group | NA |
| Pramipexole | 370 | Genus | Firmicutes | Peptostreptococcaceae | Intestinibacter | NA |
| Pramipexole | 371 | Genus | Firmicutes | Lachnospiraceae | Anaerostipes | NA |
| Pramipexole | 372 | Genus | Firmicutes | Lachnospiraceae | Marvinbryantia | NA |
| Pramipexole | 373 | Genus | Firmicutes | Lachnospiraceae | [Eubacterium] hallii group | NA |
| Pramipexole | 374 | Genus | Bacteroidota | Barnesiellaceae | Coprobacter | NA |
| Pramipexole | 375 | Genus | Firmicutes | Ruminococcaceae | DTU089 | NA |
| Pramipexole | 376 | Genus | Bacteroidota | Tannerellaceae | Parabacteroides | NA |
| Pramipexole | 377 | Genus | Firmicutes | Ruminococcaceae | Ruminococcus | NA |
| Pramipexole | 378 | Genus | Firmicutes | Erysipelotrichaceae | Holdemanella | NA |
| Pramipexole | 379 | Genus | Bacteroidota | Prevotellaceae | Paraprevotella | NA |
| Pramipexole | 380 | Genus | Firmicutes | Lactobacillaceae | Lactobacillus | NA |
| Pramipexole | 381 | Genus | Firmicutes | Oscillospiraceae | Oscillospira | NA |
| Pramipexole | 382 | Genus | Firmicutes | Erysipelatoclostridiaceae | Coprobacillus | NA |
| Pramipexole | 383 | Genus | Verrucomicrobiota | Victivallaceae | Victivallis | NA |
| Pramipexole | 384 | Genus | Actinobacteriota | Actinomycetaceae | Actinomyces | NA |
| Pramipexole | 385 | Genus | Firmicutes | Lachnospiraceae | Howardella | NA |
| Pramipexole | 386 | Genus | Firmicutes | Lachnospiraceae | Eisenbergiella | NA |
| Pramipexole | 387 | Genus | Firmicutes | Lachnospiraceae | Blautia | NA |
| Pramipexole | 388 | Genus | Firmicutes | Oscillospiraceae | UCG-005 | NA |
| Pramipexole | 389 | Genus | Proteobacteria | Oxalobacteraceae | Oxalobacter | NA |

| Ticker | DESeq2_baseMean | DESeq2_log2-FoldChange | DESeq2_lfcSE | DESeq2_stat | DESeq2_pvalue | DESeq2_padj | DESeq2 converged | ANCOM_W | ANCOM_detected_0.7 |
|---|---|---|---|---|---|---|---|---|---|
| 360 | 17,17654912 | -0,339204244 | 3,099787475 | -0,109428226 | 0,912862847 | 0,998567161 | YES | 0 | FALSE |
| 361 | 1353,359098 | 0,413380057 | 0,69971633 | 0,590782349 | 0,554666263 | 0,998567161 | YES | 0 | FALSE |
| 362 | 130,0618911 | 2,373947079 | 1,816504274 | 1,306876682 | 0,191254597 | 0,998567161 | YES | 0 | FALSE |
| 363 | 44,19644707 | 0,697056185 | 1,458911323 | 0,477792018 | 0,632798243 | 0,998567161 | YES | 0 | FALSE |
| 364 | 17,04352115 | 0,005639676 | 3,044013356 | 0,001852711 | 0,998521752 | 0,998567161 | YES | 0 | FALSE |
| 365 | 331,4686411 | 0,181096768 | 0,636398794 | 0,284564914 | 0,775977495 | 0,998567161 | YES | 0 | FALSE |
| 366 | 1102,079131 | 1,15214913 | 0,930236122 | 1,23855557 | 0,215510131 | 0,998567161 | YES | 0 | FALSE |
| 367 | 19,80434916 | 0,694238291 | 1,976863707 | 0,351181667 | 0,725452063 | 0,998567161 | YES | 0 | FALSE |
| 368 | 66,47623354 | 0,890024108 | 0,824649543 | 1,079275572 | 0,280464899 | 0,998567161 | YES | 0 | FALSE |
| 369 | 15,82505315 | 3,08468982 | 1,647008507 | 1,872904606 | 0,061081559 | 0,998567161 | YES | 0 | FALSE |
| 370 | 42,5719079 | -1,008499654 | 1,317856347 | -0,765257652 | 0,444118136 | 0,998567161 | YES | 0 | FALSE |
| 371 | 281,498817 | -0,049459835 | 0,644067874 | -0,076792892 | 0,938788305 | 0,998567161 | YES | 0 | FALSE |
| 372 | 41,68597386 | 1,970488781 | 2,020579413 | 0,975209768 | 0,329456218 | 0,998567161 | YES | 0 | FALSE |
| 373 | 384,917426 | 0,673797274 | 0,468136934 | 1,43931663 | 0,150060833 | 0,998567161 | YES | 0 | FALSE |
| 374 | 3,742647479 | 2,169321493 | 3,064759155 | 0,707827723 | 0,479052248 | 0,998567161 | YES | 0 | FALSE |
| 375 | 52,03740731 | 2,101219057 | 1,117217969 | 1,880760169 | 0,060004551 | 0,998567161 | YES | 0 | FALSE |
| 376 | 834,0789902 | 0,828408859 | 1,406958545 | 0,588794077 | 0,555999419 | 0,998567161 | YES | 0 | FALSE |
| 377 | 1502,742616 | -0,860725616 | 1,13799685 | -0,756351493 | 0,449438483 | 0,998567161 | YES | 0 | FALSE |
| 378 | 87,88670941 | -1,409420653 | 3,042270878 | -0,463279145 | 0,643164292 | 0,998567161 | YES | 0 | FALSE |
| 379 | 73,12775143 | -0,521537948 | 2,123783592 | -0,245570194 | 0,806014965 | 0,998567161 | YES | 0 | FALSE |
| 380 | 4,952380139 | 4,362167394 | 3,041269419 | 1,434324551 | 0,151479667 | 0,998567161 | YES | 0 | FALSE |
| 381 | 55,07116979 | -0,161021223 | 0,909255105 | -0,17709136 | 0,859436629 | 0,998567161 | YES | 0 | FALSE |
| 382 | 2,89436445 | 0,772408325 | 3,102156134 | 0,248990796 | 0,803367901 | 0,998567161 | YES | 0 | FALSE |
| 383 | 0,778211941 | -2,047919181 | 3,111660383 | -0,658143541 | 0,5104459 | 0,998567161 | YES | 0 | FALSE |
| 384 | 0,96555808 | -0,010116455 | 3,106445969 | -0,003256601 | 0,997401613 | 0,998567161 | YES | 0 | FALSE |
| 385 | 1,011809556 | -0,927303819 | 3,107188687 | -0,298438206 | 0,765368732 | 0,998567161 | YES | 0 | FALSE |
| 386 | 42,40951613 | 0,236995904 | 2,375908694 | 0,099749584 | 0,920543134 | 0,998567161 | YES | 0 | FALSE |
| 387 | 2676,734993 | -0,350942123 | 0,668877178 | -0,52467349 | 0,599810189 | 0,998567161 | YES | 0 | FALSE |
| 388 | 207,6260688 | 1,638865436 | 2,031638891 | 0,80667162 | 0,419855699 | 0,998567161 | YES | 0 | FALSE |
| 389 | 12,48260945 | 0,534782305 | 2,28425114 | 0,234117123 | 8,15E-01 | 0,998567161 | YES | 0 | FALSE |

| Medication | Ticker | taxLevel | Phylum | Family | Genus | Species |
|---|---|---|---|---|---|---|
| Pramipexole | 390 | Genus | Firmicutes | Lachnospiraceae | [Ruminococcus] torques group | NA |
| Pramipexole | 391 | Genus | Firmicutes | Veillonellaceae | Allisonella | NA |
| Pramipexole | 392 | Genus | Bacteroidota | Marinifilaceae | Butyricimonas | NA |
| Pramipexole | 393 | Genus | Firmicutes | Monoglobaceae | Monoglobus | NA |
| Pramipexole | 394 | Genus | Bacteroidota | Rikenellaceae | Rikenellaceae RC9 gut group | NA |
| Pramipexole | 395 | Genus | Firmicutes | Lachnospiraceae | Roseburia | NA |
| Pramipexole | 396 | Genus | Firmicutes | Erysipelatoclostridiaceae | Erysipelatoclostridium | NA |
| Pramipexole | 397 | Genus | Firmicutes | Ruminococcaceae | [Eubacterium] siraeum group | NA |
| Pramipexole | 398 | Genus | Firmicutes | Erysipelatoclostridiaceae | Catenibacterium | NA |
| Pramipexole | 399 | Genus | Firmicutes | Ruminococcaceae | Angelakisella | NA |
| Pramipexole | 400 | Genus | Firmicutes | Ruminococcaceae | Incertae Sedis | NA |
| Pramipexole | 401 | Genus | Firmicutes | Butyricicoccaceae | Butyricicoccus | NA |
| Pramipexole | 402 | Genus | Firmicutes | Erysipelotrichaceae | Holdemania | NA |
| Pramipexole | 403 | Genus | Firmicutes | Ruminococcaceae | Fournierella | NA |
| Pramipexole | 404 | Genus | Firmicutes | Lachnospiraceae | Agathobacter | NA |
| Pramipexole | 405 | Genus | Proteobacteria | Sutterellaceae | Parasutterella | NA |
| Pramipexole | 406 | Genus | Firmicutes | Ruminococcaceae | Anaerotruncus | NA |
| Pramipexole | 407 | Genus | Firmicutes | Lachnospiraceae | [Ruminococcus] gauvreauii group | NA |
| Pramipexole | 408 | Genus | Firmicutes | Lachnospiraceae | Lachnospiraceae ND3007 group | NA |
| Pramipexole | 409 | Genus | Firmicutes | Defluviitaleaceae | Defluviitaleaceae UCG-011 | NA |
| Pramipexole | 410 | Genus | Firmicutes | Lachnospiraceae | [Eubacterium] eligens group | NA |
| Pramipexole | 411 | Genus | Firmicutes | Lachnospiraceae | [Eubacterium] ruminantium group | NA |
| Pramipexole | 412 | Genus | Firmicutes | Ruminococcaceae | Faecalibacterium | NA |
| Pramipexole | 413 | Genus | Firmicutes | Lachnospiraceae | Lachnospiraceae NK4A136 group | NA |
| Pramipexole | 414 | Genus | Firmicutes | Lachnospiraceae | Coprococcus | NA |
| Pramipexole | 415 | Genus | Proteobacteria | Sutterellaceae | Sutterella | NA |
| Pramipexole | 416 | Genus | Firmicutes | Lachnospiraceae | Shuttleworthia | NA |
| Pramipexole | 417 | Genus | Firmicutes | Erysipelotrichaceae | Turicibacter | NA |
| Pramipexole | 418 | Genus | Firmicutes | Lachnospiraceae | CAG-56 | NA |
| Pramipexole | 419 | Genus | Firmicutes | Lachnospiraceae | [Ruminococcus] gnavus group | NA |

| Ticker | DESeq2_baseMean | DESeq2_log2-FoldChange | DESeq2_lfcSE | DESeq2_stat | DESeq2_pvalue | DESeq2_padj | DESeq2 converged | ANCOM_W | ANCOM_detected_0.7 |
|---|---|---|---|---|---|---|---|---|---|
| 390 | 1058,1215 | -0,311373394 | 0,645254484 | -0,482559055 | 0,629408858 | 0,998567161 | YES | 0 | FALSE |
| 391 | 4,507151657 | -0,94442347 | 3,101167459 | -0,304538043 | 0,760718014 | 0,998567161 | YES | 0 | FALSE |
| 392 | 18,7865398 | 0,107045018 | 1,315213963 | 0,081389813 | 0,935131951 | 0,998567161 | YES | 0 | FALSE |
| 393 | 218,6629912 | 0,854179592 | 0,970664993 | 0,879994229 | 0,378862436 | 0,998567161 | YES | 0 | FALSE |
| 394 | 116,3580989 | -0,949431821 | 3,099439075 | -0,306323757 | 0,759358153 | 0,998567161 | YES | 0 | FALSE |
| 395 | 371,9546907 | -0,801230726 | 1,444931672 | -0,554511152 | 0,579229076 | 0,998567161 | YES | 0 | FALSE |
| 396 | 110,1501044 | 0,255279921 | 1,385042787 | 0,184311939 | 0,853768749 | 0,998567161 | YES | 0 | FALSE |
| 397 | 349,2160858 | -0,478337344 | 0,761869094 | -0,627847156 | 0,530104071 | 0,998567161 | YES | 0 | FALSE |
| 398 | 5,794909274 | -0,94556809 | 3,100770037 | -0,304946216 | 0,760407116 | 0,998567161 | YES | 0 | FALSE |
| 399 | 41,88704901 | 2,467219179 | 1,551716002 | 1,58999403 | 0,111836151 | 0,998567161 | YES | 0 | FALSE |
| 400 | 281,134366 | 1,197715847 | 0,737222602 | 1,624632564 | 0,104240874 | 0,998567161 | YES | 0 | FALSE |
| 401 | 196,1325531 | -0,104463565 | 0,534948757 | -0,195277703 | 0,845175551 | 0,998567161 | YES | 0 | FALSE |
| 402 | 9,418012638 | 0,949120128 | 0,969198817 | 0,979283209 | 0,327440065 | 0,998567161 | YES | 0 | FALSE |
| 403 | 1,790211743 | 0,74503356 | 3,105051451 | 0,23994242 | 0,810374895 | 0,998567161 | YES | 0 | FALSE |
| 404 | 4103,809375 | 0,470273483 | 0,670916875 | 0,700941504 | 0,483339522 | 0,998567161 | YES | 0 | FALSE |
| 405 | 218,5495706 | -0,287296612 | 0,925678028 | -0,310363435 | 0,756284597 | 0,998567161 | YES | 0 | FALSE |
| 406 | 4,452740537 | -0,491783149 | 3,049316549 | -0,161276516 | 0,871875619 | 0,998567161 | YES | 0 | FALSE |
| 407 | 6,575366094 | 0,005475778 | 3,049215651 | 0,001795799 | 0,998567161 | 0,998567161 | YES | 0 | FALSE |
| 408 | 16,3345729 | -0,627010956 | 2,992319191 | -0,209540131 | 0,834026611 | 0,998567161 | YES | 0 | FALSE |
| 409 | 0,855235155 | -0,088385824 | 3,069112425 | -0,028798497 | 0,9770253 | 0,998567161 | YES | 0 | FALSE |
| 410 | 184,1701242 | -0,865868355 | 2,245404775 | -0,385617936 | 0,699779648 | 0,998567161 | YES | 0 | FALSE |
| 411 | 19,07108089 | 0,577247649 | 3,099743124 | 0,18622435 | 0,852268829 | 0,998567161 | YES | 0 | FALSE |
| 412 | 2498,489811 | 0,207527229 | 0,513473587 | 0,404163396 | 0,686092568 | 0,998567161 | YES | 0 | FALSE |
| 413 | 180,0333742 | 0,570502771 | 1,513815991 | 0,376864014 | 0,706274666 | 0,998567161 | YES | 0 | FALSE |
| 414 | 574,4731389 | -0,377469855 | 0,983022702 | -0,38398895 | 0,700986637 | 0,998567161 | YES | 0 | FALSE |
| 415 | 145,7481738 | 0,709057917 | 1,051273675 | 0,6744751 | 0,500009311 | 0,998567161 | YES | 0 | FALSE |
| 416 | 8,044064653 | -0,810062913 | 2,791055478 | -0,290235332 | 0,771636208 | 0,998567161 | YES | 0 | FALSE |
| 417 | 7,336055277 | 1,973853729 | 3,012001481 | 0,655329601 | 0,512255566 | 0,998567161 | YES | 0 | FALSE |
| 418 | 343,2845611 | -3,448042823 | 2,067135371 | -1,668029521 | 0,095309867 | 0,998567161 | YES | 0 | FALSE |
| 419 | 2,812958118 | -2,489480242 | 3,072504988 | -0,810244491 | 0,417799671 | 0,998567161 | YES | 0 | FALSE |

| Medication | Ticker | taxLevel | Phylum | Family | Genus | Species |
|---|---|---|---|---|---|---|
| Pramipexole | 420 | Genus | Firmicutes | Butyricicoccaceae | UCG-009 | NA |
| Pramipexole | 421 | Genus | Firmicutes | Erysipelotrichaceae | Dielma | NA |
| Pramipexole | 422 | Genus | Firmicutes | Acidaminococcaceae | Phascolarctobacterium | NA |
| Pramipexole | 423 | Family | Actinobacteriota | Bifidobacteriaceae | NA | NA |
| Pramipexole | 424 | Family | Verrucomicrobiota | Akkermansiaceae | NA | NA |
| Pramipexole | 425 | Family | Thermoplasmatota | Methanomethylophilaceae | NA | NA |
| Pramipexole | 426 | Family | Firmicutes | Veillonellaceae | NA | NA |
| Pramipexole | 427 | Family | Bacteroidota | Muribaculaceae | NA | NA |
| Pramipexole | 428 | Family | Firmicutes | Christensenellaceae | NA | NA |
| Pramipexole | 429 | Family | Firmicutes | Peptostreptococcaceae | NA | NA |
| Pramipexole | 430 | Family | Desulfobacterota | Desulfovibrionaceae | NA | NA |
| Pramipexole | 431 | Family | Actinobacteriota | Coriobacteriaceae | NA | NA |
| Pramipexole | 432 | Family | Proteobacteria | Pasteurellaceae | NA | NA |
| Pramipexole | 433 | Family | Firmicutes | Erysipelatoclostridiaceae | NA | NA |
| Pramipexole | 434 | Family | Bacteroidota | Marinifilaceae | NA | NA |
| Pramipexole | 435 | Family | Bacteroidota | Barnesiellaceae | NA | NA |
| Pramipexole | 436 | Family | Bacteroidota | Bacteroidaceae | NA | NA |
| Pramipexole | 437 | Family | Bacteroidota | Prevotellaceae | NA | NA |
| Pramipexole | 438 | Family | Firmicutes | Peptococcaceae | NA | NA |
| Pramipexole | 439 | Family | Firmicutes | Streptococcaceae | NA | NA |
| Pramipexole | 440 | Family | Firmicutes | Acholeplasmataceae | NA | NA |
| Pramipexole | 441 | Family | Actinobacteriota | Coriobacteriales Incertae Sedis | NA | NA |
| Pramipexole | 442 | Family | Euryarchaeota | Methanobacteriaceae | NA | NA |
| Pramipexole | 443 | Family | Firmicutes | Clostridiaceae | NA | NA |
| Pramipexole | 444 | Family | Firmicutes | Oscillospiraceae | NA | NA |
| Pramipexole | 445 | Family | Proteobacteria | Enterobacteriaceae | NA | NA |
| Pramipexole | 446 | Family | Bacteroidota | Rikenellaceae | NA | NA |
| Pramipexole | 447 | Family | Firmicutes | Anaerovoracaceae | NA | NA |
| Pramipexole | 448 | Family | Bacteroidota | Tannerellaceae | NA | NA |
| Pramipexole | 449 | Family | Firmicutes | Ruminococcaceae | NA | NA |

| Ticker | DESeq2_baseMean | DESeq2_log2-FoldChange | DESeq2_lfcSE | DESeq2_stat | DESeq2_pvalue | DESeq2_padj | DESeq2 converged | ANCOM_W | ANCOM_detected_0.7 |
|---|---|---|---|---|---|---|---|---|---|
| 420 | 12,89337023 | 1,016089183 | 2,06725263 | 0,491516697 | 0,623061046 | 0,998567161 | YES | 0 | FALSE |
| 421 | 0,959063134 | 0,126992231 | 3,106621985 | 0,040877915 | 0,967393224 | 0,998567161 | YES | 0 | FALSE |
| 422 | 587,9129411 | -1,099522474 | 1,691177272 | -0,650152112 | 0,515593971 | 0,998567161 | YES | 0 | FALSE |
| 423 | 1452,852193 | 2,045519186 | 0,542507586 | 3,770489554 | 0,000162928 | 0,007168815 | YES | 0 | FALSE |
| 424 | 1064,304049 | -1,76586698 | 1,610276374 | -1,096623542 | 0,272805992 | 0,887421601 | YES | 0 | FALSE |
| 425 | 5,867307413 | 0,087713395 | 3,100476575 | 0,028290294 | 0,977430621 | 0,999074427 | YES | 0 | FALSE |
| 426 | 1819,828224 | 1,424396479 | 0,946304586 | 1,505219883 | 0,132267555 | 0,714067636 | YES | 0 | FALSE |
| 427 | 127,7645415 | -1,014278764 | 2,854683215 | -0,355303439 | 0,722362279 | 0,999074427 | YES | 0 | FALSE |
| 428 | 468,487708 | 1,039196222 | 1,268796677 | 0,819040782 | 0,412763146 | 0,887421601 | YES | 0 | FALSE |
| 429 | 89,80318698 | 0,631068119 | 0,705457591 | 0,894551462 | 0,371026908 | 0,887421601 | YES | 0 | FALSE |
| 430 | 174,7569869 | 0,731347809 | 1,099623715 | 0,665089156 | 0,50599347 | 0,887421601 | YES | 0 | FALSE |
| 431 | 874,8211621 | 1,146285388 | 0,654378182 | 1,751717004 | 0,079822482 | 0,584118713 | YES | 0 | FALSE |
| 432 | 15,2039086 | -1,293053208 | 2,916583782 | -0,443345127 | 0,657516119 | 0,982897865 | YES | 0 | FALSE |
| 433 | 361,249562 | -0,443041206 | 0,945322499 | -0,468666732 | 0,63930787 | 0,982897865 | YES | 0 | FALSE |
| 434 | 41,71305716 | -0,565520886 | 0,690927453 | -0,818495319 | 0,413074414 | 0,887421601 | YES | 0 | FALSE |
| 435 | 232,0328804 | 1,07836753 | 1,080315868 | 0,998196511 | 0,318184078 | 0,887421601 | YES | 0 | FALSE |
| 436 | 8015,177685 | -0,094825674 | 0,479885511 | -0,197600618 | 0,843357551 | 0,999074427 | YES | 0 | FALSE |
| 437 | 2183,684499 | -1,412019444 | 0,759848658 | -1,858290371 | 0,063127791 | 0,584118713 | YES | 0 | FALSE |
| 438 | 49,90636662 | -1,153610221 | 0,993717487 | -1,160903613 | 0,245681099 | 0,887421601 | YES | 0 | FALSE |
| 439 | 477,6137919 | 1,645745265 | 1,167448872 | 1,409693653 | 0,15863016 | 0,714067636 | YES | 0 | FALSE |
| 440 | 88,39021296 | 0,199238074 | 3,0994447 | 0,064281861 | 0,948745796 | 0,999074427 | YES | 0 | FALSE |
| 441 | 41,06695359 | 1,353946056 | 1,646897662 | 0,822119119 | 0,411009105 | 0,887421601 | YES | 0 | FALSE |
| 442 | 70,49045809 | 0,912824582 | 1,249789952 | 0,730382398 | 0,465156475 | 0,887421601 | YES | 0 | FALSE |
| 443 | 36,31313935 | -0,804346767 | 1,502449267 | -0,535357023 | 0,592403002 | 0,965397485 | YES | 0 | FALSE |
| 444 | 2443,524326 | 0,261599171 | 0,614181072 | 0,425931673 | 0,670157635 | 0,982897865 | YES | 0 | FALSE |
| 445 | 143,5553921 | 2,842728113 | 1,689195877 | 1,68288838 | 0,092396701 | 0,584118713 | YES | 0 | FALSE |
| 446 | 1198,635133 | 0,540191575 | 0,848557431 | 0,636599899 | 0,524385491 | 0,887421601 | YES | 0 | FALSE |
| 447 | 117,5343183 | 1,162882423 | 0,692129788 | 1,68015081 | 0,092927977 | 0,584118713 | YES | 0 | FALSE |
| 448 | 741,2548036 | 0,835684825 | 1,267475886 | 0,659329959 | 0,509683908 | 0,887421601 | YES | 0 | FALSE |
| 449 | 7957,067181 | 0,05900857 | 0,468895449 | 0,125845901 | 0,899853907 | 0,999074427 | YES | 0 | FALSE |

| Medication | Ticker | taxLevel | Phylum | Family | Genus | Species |
|---|---|---|---|---|---|---|
| Pramipexole | 450 | Family | Firmicutes | Erysipelotrichaceae | NA | NA |
| Pramipexole | 451 | Family | Firmicutes | Lactobacillaceae | NA | NA |
| Pramipexole | 452 | Family | Firmicutes | UCG-010 | NA | NA |
| Pramipexole | 453 | Family | Actinobacteriota | Atopobiaceae | NA | NA |
| Pramipexole | 454 | Family | Verrucomicrobiota | Victivallaceae | NA | NA |
| Pramipexole | 455 | Family | Actinobacteriota | Actinomycetaceae | NA | NA |
| Pramipexole | 456 | Family | Proteobacteria | Oxalobacteraceae | NA | NA |
| Pramipexole | 457 | Family | Firmicutes | Monoglobaceae | NA | NA |
| Pramipexole | 458 | Family | Firmicutes | [Eubacterium] coprostanoligenes group | NA | NA |
| Pramipexole | 459 | Family | Actinobacteriota | Eggerthellaceae | NA | NA |
| Pramipexole | 460 | Family | Firmicutes | Butyricicoccaceae | NA | NA |
| Pramipexole | 461 | Family | Firmicutes | Lachnospiraceae | NA | NA |
| Pramipexole | 462 | Family | Firmicutes | Hydrogenoanaerobacterium | NA | NA |
| Pramipexole | 463 | Family | Proteobacteria | Sutterellaceae | NA | NA |
| Pramipexole | 464 | Family | Verrucomicrobiota | Puniceicoccaceae | NA | NA |
| Pramipexole | 465 | Family | Firmicutes | Defluviitaleaceae | NA | NA |
| Pramipexole | 466 | Family | Firmicutes | Acidaminococcaceae | NA | NA |

| Ticker | DESeq2_baseMean | DESeq2_log2-FoldChange | DESeq2_lfcSE | DESeq2_stat | DESeq2_pvalue | DESeq2_padj | DESeq2 converged | ANCOM_W | ANCOM_detected_0.7 |
|---|---|---|---|---|---|---|---|---|---|
| 450 | 137,2007408 | -0,597182953 | 0,772215752 | -0,773336923 | 0,43932301 | 0,887421601 | YES | 0 | FALSE |
| 451 | 4,047915222 | 4,260552203 | 3,048875947 | 1,397417369 | 0,162288099 | 0,714067636 | YES | 0 | FALSE |
| 452 | 32,48961552 | -0,212568206 | 2,725685933 | -0,07798705 | 0,937838354 | 0,999074427 | YES | 0 | FALSE |
| 453 | 1,36035841 | -0,205100965 | 3,061329334 | -0,066997354 | 0,94658381 | 0,999074427 | YES | 0 | FALSE |
| 454 | 1,73190797 | -2,099862113 | 3,076694611 | -0,682505864 | 0,494919136 | 0,887421601 | YES | 0 | FALSE |
| 455 | 0,556230961 | -0,059616229 | 3,10659155 | -0,019190237 | 0,984689346 | 0,999074427 | YES | 0 | FALSE |
| 456 | 12,30161991 | 0,41854002 | 2,219345543 | 0,188587136 | 0,850416416 | 0,999074427 | YES | 0 | FALSE |
| 457 | 243,3811586 | 0,904186732 | 0,951371579 | 0,950403347 | 0,341907344 | 0,887421601 | YES | 0 | FALSE |
| 458 | 395,6115501 | 1,225674274 | 0,642916684 | 1,906427853 | 0,056594718 | 0,584118713 | YES | 0 | FALSE |
| 459 | 184,6983295 | 1,690073345 | 0,886048653 | 1,907427249 | 0,056465284 | 0,584118713 | YES | 0 | FALSE |
| 460 | 222,8593832 | 0,116079937 | 0,427108834 | 0,271780697 | 0,785790648 | 0,999074427 | YES | 0 | FALSE |
| 461 | 14073,51074 | -0,036978599 | 0,461229178 | -0,080174024 | 0,93609885 | 0,999074427 | YES | 0 | FALSE |
| 462 | 1,256263016 | -0,971379696 | 3,103906599 | -0,312953907 | 0,754315682 | 0,999074427 | YES | 0 | FALSE |
| 463 | 364,3547965 | 0,056811161 | 0,698581336 | 0,081323618 | 0,935184592 | 0,999074427 | YES | 0 | FALSE |
| 464 | 1,060703435 | -2,018310566 | 3,095244348 | -0,65206825 | 0,514357145 | 0,887421601 | YES | 0 | FALSE |
| 465 | 0,690960416 | 0,0035603 | 3,069132959 | 0,001160034 | 0,999074427 | 0,999074427 | YES | 0 | FALSE |
| 466 | 571,4730027 | -1,130911266 | 1,605895547 | -0,704224673 | 0,481292868 | 0,887421601 | YES | 0 | FALSE |

**Supplementary Table 7c: users of only ropinirole**

| Medication | Ticker | taxLevel | Phylum | Family | Genus | Species |
|---|---|---|---|---|---|---|
| Ropinirole | 1 | ASV | Actinobacteriota | Bifidobacteriaceae | Bifidobacterium | NA |
| Ropinirole | 2 | ASV | Verrucomicrobiota | Akkermansiaceae | Akkermansia | muciniphila |
| Ropinirole | 3 | ASV | Firmicutes | Christensenellaceae | NA | NA |
| Ropinirole | 4 | ASV | Firmicutes | Lachnospiraceae | Roseburia | NA |
| Ropinirole | 5 | ASV | Firmicutes | NA | NA | NA |
| Ropinirole | 6 | ASV | Firmicutes | Christensenellaceae | Christensenellaceae R-7 group | NA |
| Ropinirole | 7 | ASV | Bacteroidota | Bacteroidaceae | Bacteroides | eggerthii |
| Ropinirole | 8 | ASV | Firmicutes | UCG-010 | NA | NA |
| Ropinirole | 9 | ASV | Firmicutes | [Eubacterium] coprostanoligenes group | NA | NA |
| Ropinirole | 10 | ASV | Firmicutes | Ruminococcaceae | Subdoligranulum | NA |
| Ropinirole | 11 | ASV | Firmicutes | Lachnospiraceae | Lachnospira | pectinoschiza |
| Ropinirole | 12 | ASV | Firmicutes | Oscillospiraceae | UCG-003 | NA |
| Ropinirole | 13 | ASV | Bacteroidota | Rikenellaceae | Alistipes | finegoldii |
| Ropinirole | 14 | ASV | Firmicutes | NA | NA | NA |
| Ropinirole | 15 | ASV | Firmicutes | Ruminococcaceae | Subdoligranulum | NA |
| Ropinirole | 16 | ASV | Firmicutes | Veillonellaceae | Dialister | invisus |
| Ropinirole | 17 | ASV | Firmicutes | Lachnospiraceae | Anaerostipes | hadrus |
| Ropinirole | 18 | ASV | Firmicutes | Oscillospiraceae | UCG-002 | NA |
| Ropinirole | 19 | ASV | Bacteroidota | Tannerellaceae | Parabacteroides | distasonis |
| Ropinirole | 20 | ASV | Firmicutes | Lachnospiraceae | Lachnospiraceae NK4A136 group | bacterium |
| Ropinirole | 21 | ASV | Firmicutes | Oscillospiraceae | NK4A214 group | NA |
| Ropinirole | 22 | ASV | Firmicutes | Christensenellaceae | Christensenellaceae R-7 group | NA |
| Ropinirole | 23 | ASV | Firmicutes | Lachnospiraceae | Fusicatenibacter | saccharivorans |
| Ropinirole | 24 | ASV | Firmicutes | Lachnospiraceae | NA | NA |
| Ropinirole | 25 | ASV | Firmicutes | Ruminococcaceae | CAG-352 | NA |
| Ropinirole | 26 | ASV | Actinobacteriota | Eggerthellaceae | Senegalimassilia | anaerobia |
| Ropinirole | 27 | ASV | Actinobacteriota | Eggerthellaceae | Enterorhabdus | NA |
| Ropinirole | 28 | ASV | Bacteroidota | Rikenellaceae | Alistipes | inops |
| Ropinirole | 29 | ASV | Firmicutes | Ruminococcaceae | NA | NA |

| Ticker | DESeq2_baseMean | DESeq2_log2-FoldChange | DESeq2_lfcSE | DESeq2_stat | DESeq2_pvalue | DESeq2_padj | DESeq2 converged | ANCOM_W | ANCOM_detected_0.7 |
|---|---|---|---|---|---|---|---|---|---|
| 1 | 229,8495876 | 0,170147007 | 3,362767449 | 0,050597316 | 0,959646402 | 0,996273765 | YES | 0 | FALSE |
| 2 | 216,0470614 | 1,832579338 | 3,091957122 | 0,592692352 | 0,553387061 | 0,996273765 | YES | 0 | FALSE |
| 3 | 166,1995977 | -0,174811454 | 2,744208014 | -0,063701969 | 0,949207537 | 0,996273765 | YES | 0 | FALSE |
| 4 | 5,788917359 | 2,508378563 | 3,947091374 | 0,635500505 | 0,525102037 | 0,996273765 | YES | 0 | FALSE |
| 5 | 0,819166674 | 0,032785404 | 3,954555955 | 0,00829054 | 0,993385182 | 0,996273765 | YES | 0 | FALSE |
| 6 | 71,77510376 | -1,078296685 | 3,389522776 | -0,318126402 | 0,750389057 | 0,996273765 | YES | 0 | FALSE |
| 7 | 60,78666764 | 1,611774543 | 3,405899396 | 0,473230227 | 0,636048935 | 0,996273765 | YES | 0 | FALSE |
| 8 | 0,480409336 | 1,956000933 | 4,030767633 | 0,485267599 | 0,627486536 | 0,996273765 | YES | 1 | FALSE |
| 9 | 29,90846617 | 0,070682619 | 3,278899675 | 0,021556811 | 0,982801486 | 0,996273765 | YES | 0 | FALSE |
| 10 | 245,2198831 | 0,750598616 | 2,232511533 | 0,336212649 | 0,736710515 | 0,996273765 | YES | 0 | FALSE |
| 11 | 28,63969775 | 0,064630673 | 2,908966074 | 0,022217747 | 0,982274261 | 0,996273765 | YES | 0 | FALSE |
| 12 | 254,1192167 | 0,110443141 | 2,733551513 | 0,040402802 | 0,967771996 | 0,996273765 | YES | 0 | FALSE |
| 13 | 73,38945966 | 1,100549654 | 2,778784836 | 0,39605429 | 0,692064978 | 0,996273765 | YES | 0 | FALSE |
| 14 | 133,3288744 | 2,578919115 | 3,908984049 | 0,659741529 | 0,509419711 | 0,996273765 | YES | 0 | FALSE |
| 15 | 20,17219767 | -2,405545867 | 3,894225359 | -0,617721279 | 0,536759083 | 0,996273765 | YES | 0 | FALSE |
| 16 | 860,0452301 | 0,661299499 | 3,353000031 | 0,197226213 | 0,843650518 | 0,996273765 | YES | 0 | FALSE |
| 17 | 1,957004252 | -1,962197257 | 4,024283681 | -0,487589199 | 0,625840848 | 0,996273765 | YES | 0 | FALSE |
| 18 | 5,572748292 | 2,551537227 | 3,959741438 | 0,644369656 | 0,519335757 | 0,996273765 | YES | 0 | FALSE |
| 19 | 48,51559059 | -0,247308796 | 3,416955101 | -0,072376952 | 0,942301926 | 0,996273765 | YES | 0 | FALSE |
| 20 | 450,6191727 | -2,255058792 | 3,162192781 | -0,713131345 | 0,475764485 | 0,996273765 | YES | 0 | FALSE |
| 21 | 139,2105032 | 0,514988325 | 2,725513518 | 0,188950934 | 0,850131273 | 0,996273765 | YES | 0 | FALSE |
| 22 | 62,64125852 | -0,077040485 | 3,395871024 | -0,022686517 | 0,981900331 | 0,996273765 | YES | 0 | FALSE |
| 23 | 436,2633886 | 0,077023425 | 2,727569521 | 0,028238849 | 0,977471652 | 0,996273765 | YES | 0 | FALSE |
| 24 | 242,7791617 | -1,590389782 | 2,548101663 | -0,624146911 | 0,532531108 | 0,996273765 | YES | 0 | FALSE |
| 25 | 11,91415152 | 2,600912196 | 3,950840986 | 0,658318622 | 0,510333414 | 0,996273765 | YES | 0 | FALSE |
| 26 | 14,24401045 | 1,891857754 | 2,682207512 | 0,705336088 | 0,480601107 | 0,996273765 | YES | 0 | FALSE |
| 27 | 9,290547146 | -3,166088926 | 3,295976772 | -0,960592002 | 0,336757352 | 0,996273765 | YES | 0 | FALSE |
| 28 | 24,70129956 | 1,18516015 | 3,459439592 | 0,342587323 | 0,73190894 | 0,996273765 | YES | 0 | FALSE |
| 29 | 133,4255727 | 1,74656473 | 2,688759154 | 0,649580208 | 0,515963421 | 0,996273765 | YES | 0 | FALSE |

| Medication | Ticker | taxLevel | Phylum | Family | Genus | Species |
|---|---|---|---|---|---|---|
| Ropinirole | 30 | ASV | Bacteroidota | Rikenellaceae | Alistipes | obesi |
| Ropinirole | 31 | ASV | Firmicutes | Christensenellaceae | Christensenellaceae R-7 group | NA |
| Ropinirole | 32 | ASV | Firmicutes | Lachnospiraceae | Blautia | massiliensis |
| Ropinirole | 33 | ASV | Bacteroidota | Tannerellaceae | Parabacteroides | distasonis |
| Ropinirole | 34 | ASV | Firmicutes | Lachnospiraceae | Lachnoclostridium | NA |
| Ropinirole | 35 | ASV | Firmicutes | Ruminococcaceae | Faecalibacterium | prausnitzii |
| Ropinirole | 36 | ASV | Firmicutes | Peptostreptococcaceae | Terrisporobacter | mayombei |
| Ropinirole | 37 | ASV | Desulfobacterota | Desulfovibrionaceae | Bilophila | wadsworthia |
| Ropinirole | 38 | ASV | Bacteroidota | Muribaculaceae | NA | NA |
| Ropinirole | 39 | ASV | Firmicutes | Erysipelotrichaceae | Turicibacter | sanguinis |
| Ropinirole | 40 | ASV | Firmicutes | Peptococcaceae | NA | NA |
| Ropinirole | 41 | ASV | Firmicutes | Oscillospiraceae | Oscillibacter | NA |
| Ropinirole | 42 | ASV | Firmicutes | Oscillospiraceae | UCG-002 | NA |
| Ropinirole | 43 | ASV | Proteobacteria | Sutterellaceae | Sutterella | NA |
| Ropinirole | 44 | ASV | Firmicutes | Butyricicoccaceae | Butyricicoccus | NA |
| Ropinirole | 45 | ASV | Firmicutes | Lachnospiraceae | Roseburia | hominis |
| Ropinirole | 46 | ASV | Firmicutes | Streptococcaceae | Streptococcus | NA |
| Ropinirole | 47 | ASV | Firmicutes | NA | NA | NA |
| Ropinirole | 48 | ASV | Firmicutes | Oscillospiraceae | UCG-005 | NA |
| Ropinirole | 49 | ASV | Firmicutes | Lachnospiraceae | [Eubacterium] ventriosum group | NA |
| Ropinirole | 50 | ASV | Firmicutes | Lachnospiraceae | Roseburia | NA |
| Ropinirole | 51 | ASV | Firmicutes | Peptostreptococcaceae | Romboutsia | NA |
| Ropinirole | 52 | ASV | Firmicutes | Ruminococcaceae | Ruminococcus | callidus |
| Ropinirole | 53 | ASV | Firmicutes | Oscillospiraceae | UCG-002 | NA |
| Ropinirole | 54 | ASV | Desulfobacterota | Desulfovibrionaceae | Desulfovibrio | NA |
| Ropinirole | 55 | ASV | Bacteroidota | Tannerellaceae | Parabacteroides | distasonis |
| Ropinirole | 56 | ASV | Firmicutes | Oscillospiraceae | Intestinimonas | NA |
| Ropinirole | 57 | ASV | Firmicutes | Lachnospiraceae | Lachnoclostridium | edouardi |
| Ropinirole | 58 | ASV | Actinobacteriota | Coriobacteriaceae | Collinsella | aerofaciens |
| Ropinirole | 59 | ASV | Proteobacteria | Pasteurellaceae | Haemophilus | NA |

| Ticker | DESeq2_baseMean | DESeq2_log2-FoldChange | DESeq2_lfcSE | DESeq2_stat | DESeq2_pvalue | DESeq2_padj | DESeq2 converged | ANCOM_W | ANCOM_detected_0.7 |
|---|---|---|---|---|---|---|---|---|---|
| 30 | 78,51327234 | 8,12829037 | 2,997438765 | 2,711745262 | 0,006693001 | 0,996273765 | YES | 0 | FALSE |
| 31 | 6,149239495 | 2,559884289 | 3,958227804 | 0,646724851 | 0,517810036 | 0,996273765 | YES | 0 | FALSE |
| 32 | 216,6989455 | -0,822473359 | 2,224265861 | -0,369772954 | 0,711551669 | 0,996273765 | YES | 0 | FALSE |
| 33 | 84,50901957 | -0,371843901 | 3,382866283 | -0,109919775 | 0,912472999 | 0,996273765 | YES | 0 | FALSE |
| 34 | 27,26601311 | 0,031199618 | 3,451135089 | 0,009040393 | 0,992786908 | 0,996273765 | YES | 0 | FALSE |
| 35 | 108,3279621 | -1,264048361 | 3,378094308 | -0,374189779 | 0,708263128 | 0,996273765 | YES | 0 | FALSE |
| 36 | 14,4517237 | -2,379184328 | 3,944038698 | -0,603235544 | 0,546352005 | 0,996273765 | YES | 0 | FALSE |
| 37 | 26,14121155 | -1,97253263 | 2,969077915 | -0,664358662 | 0,506460784 | 0,996273765 | YES | 0 | FALSE |
| 38 | 797,6950536 | 2,412448213 | 2,842472543 | 0,848714693 | 0,396040068 | 0,996273765 | YES | 0 | FALSE |
| 39 | 10,87009923 | -0,583689788 | 3,763030079 | -0,155111646 | 0,876733301 | 0,996273765 | YES | 0 | FALSE |
| 40 | 3,465278207 | 1,032733926 | 3,95714369 | 0,260979638 | 0,79410821 | 0,996273765 | YES | 0 | FALSE |
| 41 | 34,97471262 | -7,156423615 | 3,480753034 | -2,055998672 | 0,039782644 | 0,996273765 | YES | 0 | FALSE |
| 42 | 111,5309248 | 1,960513536 | 2,541512906 | 0,771396255 | 0,440472094 | 0,996273765 | YES | 0 | FALSE |
| 43 | 19,36741831 | 0,496574261 | 2,912100018 | 0,170521019 | 0,864600406 | 0,996273765 | YES | 0 | FALSE |
| 44 | 54,57508989 | 0,300638662 | 3,400285573 | 0,08841571 | 0,929546276 | 0,996273765 | YES | 0 | FALSE |
| 45 | 154,8795821 | 1,295314753 | 3,20911409 | 0,403636242 | 0,68648023 | 0,996273765 | YES | 0 | FALSE |
| 46 | 107,4417965 | -2,713122781 | 3,146342198 | -0,86231014 | 0,388516874 | 0,996273765 | YES | 0 | FALSE |
| 47 | 29,0539862 | -2,427446261 | 3,85884288 | -0,629060663 | 0,529309342 | 0,996273765 | YES | 0 | FALSE |
| 48 | 38,52111495 | 0,727707706 | 2,794727494 | 0,260385926 | 0,794566098 | 0,996273765 | YES | 0 | FALSE |
| 49 | 97,5878392 | -1,482708905 | 2,753345286 | -0,538511792 | 0,590223766 | 0,996273765 | YES | 0 | FALSE |
| 50 | 106,2770123 | -2,064785841 | 3,084767818 | -0,669348866 | 0,503272962 | 0,996273765 | YES | 0 | FALSE |
| 51 | 49,26327331 | 1,185676473 | 2,616236269 | 0,45319931 | 0,650405229 | 0,996273765 | YES | 0 | FALSE |
| 52 | 75,75607392 | 1,010625525 | 3,931026154 | 0,257089494 | 0,797109688 | 0,996273765 | YES | 0 | FALSE |
| 53 | 27,96510718 | -2,557541976 | 3,113279303 | -0,821494549 | 0,411364628 | 0,996273765 | YES | 0 | FALSE |
| 54 | 76,38188359 | 0,628954729 | 3,384754676 | 0,185819886 | 0,852586009 | 0,996273765 | YES | 0 | FALSE |
| 55 | 14,18830209 | 0,366741763 | 3,533244423 | 0,10379745 | 0,91733009 | 0,996273765 | YES | 0 | FALSE |
| 56 | 29,42991897 | -0,387436561 | 2,7880547 | -0,138963042 | 0,889479354 | 0,996273765 | YES | 0 | FALSE |
| 57 | 34,48027654 | -0,549657641 | 2,79046164 | -0,196977315 | 0,84384529 | 0,996273765 | YES | 0 | FALSE |
| 58 | 68,43117881 | -3,30189231 | 2,57079956 | -1,284383412 | 0,199007834 | 0,996273765 | YES | 0 | FALSE |
| 59 | 69,915044 | 0,757996421 | 3,419978458 | 0,221637776 | 8,25E-01 | 9,96E-01 | YES | 0 | FALSE |

| Medication | Ticker | taxLevel | Phylum | Family | Genus | Species |
|---|---|---|---|---|---|---|
| Ropinirole | 60 | ASV | Firmicutes | NA | NA | NA |
| Ropinirole | 61 | ASV | Bacteroidota | Prevotellaceae | Paraprevotella | NA |
| Ropinirole | 62 | ASV | Firmicutes | Lachnospiraceae | Dorea | longicatena |
| Ropinirole | 63 | ASV | Firmicutes | NA | NA | NA |
| Ropinirole | 64 | ASV | Firmicutes | Ruminococcaceae | Ruminococcus | champanellensis |
| Ropinirole | 65 | ASV | Firmicutes | Erysipelatoclostridiaceae | Erysipelotrichaceae UCG-003 | bacterium |
| Ropinirole | 66 | ASV | Firmicutes | Ruminococcaceae | UBA1819 | NA |
| Ropinirole | 67 | ASV | Firmicutes | Lachnospiraceae | Lachnospiraceae NK4A136 group | NA |
| Ropinirole | 68 | ASV | Bacteroidota | Marinifilaceae | Odoribacter | splanchnicus |
| Ropinirole | 69 | ASV | Actinobacteriota | Eggerthellaceae | Adlercreutzia | equolifaciens |
| Ropinirole | 70 | ASV | Bacteroidota | Bacteroidaceae | Bacteroides | intestinalis |
| Ropinirole | 71 | ASV | Bacteroidota | Bacteroidaceae | Bacteroides | eggerthii |
| Ropinirole | 72 | ASV | Bacteroidota | Bacteroidaceae | Bacteroides | vulgatus |
| Ropinirole | 73 | ASV | Firmicutes | Lachnospiraceae | [Eubacterium] xylanophilum group | NA |
| Ropinirole | 74 | ASV | Firmicutes | Oscillospiraceae | UCG-002 | NA |
| Ropinirole | 75 | ASV | Firmicutes | Lachnospiraceae | [Eubacterium] hallii group | NA |
| Ropinirole | 76 | ASV | Firmicutes | NA | NA | NA |
| Ropinirole | 77 | ASV | Firmicutes | Oscillospiraceae | Intestinimonas | NA |
| Ropinirole | 78 | ASV | Bacteroidota | Prevotellaceae | Prevotella | NA |
| Ropinirole | 79 | ASV | Bacteroidota | Tannerellaceae | NA | NA |
| Ropinirole | 80 | ASV | Firmicutes | Ruminococcaceae | Faecalibacterium | prausnitzii |
| Ropinirole | 81 | ASV | Firmicutes | Lachnospiraceae | Lachnospiraceae FCS020 group | NA |
| Ropinirole | 82 | ASV | Firmicutes | Lachnospiraceae | Lachnospiraceae UCG-010 | NA |
| Ropinirole | 83 | ASV | Firmicutes | Lachnospiraceae | Lachnospira | pectinoschiza |
| Ropinirole | 84 | ASV | Firmicutes | Peptococcaceae | Peptococcus | NA |
| Ropinirole | 85 | ASV | Firmicutes | Lachnospiraceae | Lachnospiraceae UCG-004 | NA |
| Ropinirole | 86 | ASV | Bacteroidota | Bacteroidaceae | Bacteroides | uniformis |
| Ropinirole | 87 | ASV | Firmicutes | Oscillospiraceae | Intestinimonas | NA |
| Ropinirole | 88 | ASV | Firmicutes | Oscillospiraceae | NK4A214 group | NA |
| Ropinirole | 89 | ASV | Bacteroidota | Barnesiellaceae | Barnesiella | NA |

| Ticker | DESeq2_baseMean | DESeq2_log2-FoldChange | DESeq2_lfcSE | DESeq2_stat | DESeq2_pvalue | DESeq2_padj | DESeq2 converged | ANCOM_W | ANCOM_detected_0.7 |
|---|---|---|---|---|---|---|---|---|---|
| 60 | 2,011223201 | 0,958516758 | 3,98581304 | 0,240482117 | 0,809956525 | 0,996273765 | YES | 0 | FALSE |
| 61 | 56,48736206 | -0,548131333 | 3,431804576 | -0,159721022 | 0,873100841 | 0,996273765 | YES | 0 | FALSE |
| 62 | 700,6026635 | 0,242563189 | 2,200281079 | 0,11024191 | 0,912217525 | 0,996273765 | YES | 0 | FALSE |
| 63 | 7,338269311 | -1,320396129 | 3,938743813 | -0,335232803 | 0,737449479 | 0,996273765 | YES | 0 | FALSE |
| 64 | 564,9063736 | -2,36540801 | 2,663493143 | -0,888084888 | 0,374495091 | 0,996273765 | YES | 0 | FALSE |
| 65 | 7,958833976 | -0,025121483 | 3,812942428 | -0,006588477 | 0,994743194 | 0,996273765 | YES | 0 | FALSE |
| 66 | 50,89218706 | -0,702612344 | 3,438268165 | -0,204350653 | 0,83807949 | 0,996273765 | YES | 0 | FALSE |
| 67 | 163,0389885 | 3,166588836 | 2,563959788 | 1,235038416 | 0,216816207 | 0,996273765 | YES | 0 | FALSE |
| 68 | 19,62201318 | -1,864875192 | 2,686766302 | -0,694096539 | 0,487621669 | 0,996273765 | YES | 0 | FALSE |
| 69 | 7,048892715 | -4,108338705 | 3,541984478 | -1,159897433 | 0,246090568 | 0,996273765 | YES | 0 | FALSE |
| 70 | 43,04718327 | 0,629751323 | 3,411884275 | 0,184575816 | 0,853561756 | 0,996273765 | YES | 0 | FALSE |
| 71 | 202,8205626 | 1,46587351 | 3,369375706 | 0,435057897 | 0,663520403 | 0,996273765 | YES | 0 | FALSE |
| 72 | 629,0234291 | -1,004577385 | 2,725864017 | -0,3685354 | 0,712474055 | 0,996273765 | YES | 0 | FALSE |
| 73 | 36,92033723 | -2,823587938 | 3,113564796 | -0,906866606 | 0,364477342 | 0,996273765 | YES | 0 | FALSE |
| 74 | 45,25323134 | -1,031059687 | 3,94294484 | -0,261494829 | 0,793710937 | 0,996273765 | YES | 0 | FALSE |
| 75 | 1,441228007 | 2,354235735 | 3,998173471 | 0,588827812 | 0,555976787 | 0,996273765 | YES | 0 | FALSE |
| 76 | 0,768654937 | 2,231325997 | 4,027383669 | 0,554038597 | 0,579552431 | 0,996273765 | YES | 0 | FALSE |
| 77 | 66,19255201 | 0,697245973 | 3,390154299 | 0,205667917 | 0,837050321 | 0,996273765 | YES | 0 | FALSE |
| 78 | 60,75139152 | 0,288180153 | 3,395458148 | 0,084872244 | 0,932362958 | 0,996273765 | YES | 0 | FALSE |
| 79 | 6,584373047 | 0,401768287 | 3,732607113 | 0,107637443 | 0,914283294 | 0,996273765 | YES | 0 | FALSE |
| 80 | 90,15691841 | -2,358586979 | 2,585021877 | -0,912405036 | 0,361555539 | 0,996273765 | YES | 0 | FALSE |
| 81 | 15,84385141 | 0,151556927 | 2,882402291 | 0,052580075 | 0,958066493 | 0,996273765 | YES | 0 | FALSE |
| 82 | 85,24482502 | -2,105027828 | 3,058506953 | -0,688253406 | 0,491293218 | 0,996273765 | YES | 0 | FALSE |
| 83 | 34,31389825 | 6,060773556 | 2,935561637 | 2,064604429 | 0,038960444 | 0,996273765 | YES | 1 | FALSE |
| 84 | 133,4585594 | 0,493097893 | 3,369921517 | 0,146323257 | 0,883666207 | 0,996273765 | YES | 0 | FALSE |
| 85 | 7,234308225 | -0,04662076 | 3,704197953 | -0,012585926 | 0,989958149 | 0,996273765 | YES | 0 | FALSE |
| 86 | 912,4786994 | 0,435052277 | 2,26530855 | 0,192049899 | 0,847703119 | 0,996273765 | YES | 0 | FALSE |
| 87 | 46,64306583 | 2,773751588 | 2,605523151 | 1,064566088 | 0,287072333 | 0,996273765 | YES | 0 | FALSE |
| 88 | 84,44170634 | 0,425402239 | 2,764489803 | 0,153880922 | 0,877703627 | 0,996273765 | YES | 0 | FALSE |
| 89 | 44,47130369 | 0,211323031 | 3,411998498 | 0,061935265 | 0,950614384 | 0,996273765 | YES | 0 | FALSE |

| Medication | Ticker | taxLevel | Phylum | Family | Genus | Species |
|---|---|---|---|---|---|---|
| Ropinirole | 90 | ASV | Firmicutes | Lachnospiraceae | Lachnospiraceae UCG-001 | NA |
| Ropinirole | 91 | ASV | Firmicutes | Oscillospiraceae | Flavonifractor | plautii |
| Ropinirole | 92 | ASV | Firmicutes | Streptococcaceae | Streptococcus | NA |
| Ropinirole | 93 | ASV | Firmicutes | Ruminococcaceae | [Eubacterium] siraeum group | NA |
| Ropinirole | 94 | ASV | Proteobacteria | NA | NA | NA |
| Ropinirole | 95 | ASV | Firmicutes | [Eubacterium] coprostanoligenes group | NA | NA |
| Ropinirole | 96 | ASV | Cyanobacteria | NA | NA | NA |
| Ropinirole | 97 | ASV | Bacteroidota | Bacteroidaceae | Bacteroides | clarus |
| Ropinirole | 98 | ASV | Bacteroidota | Bacteroidaceae | Bacteroides | uniformis |
| Ropinirole | 99 | ASV | Bacteroidota | Prevotellaceae | Alloprevotella | NA |
| Ropinirole | 100 | ASV | Firmicutes | Lachnospiraceae | Tyzzerella | NA |
| Ropinirole | 101 | ASV | Firmicutes | NA | NA | NA |
| Ropinirole | 102 | ASV | Euryarchaeota | Methanobacteriaceae | Methanosphaera | NA |
| Ropinirole | 103 | ASV | Proteobacteria | NA | NA | NA |
| Ropinirole | 104 | ASV | Actinobacteriota | Bifidobacteriaceae | Bifidobacterium | NA |
| Ropinirole | 105 | ASV | Bacteroidota | Prevotellaceae | Paraprevotella | NA |
| Ropinirole | 106 | ASV | Firmicutes | Oscillospiraceae | NK4A214 group | NA |
| Ropinirole | 107 | ASV | Bacteroidota | Tannerellaceae | Parabacteroides | distasonis |
| Ropinirole | 108 | ASV | Actinobacteriota | Coriobacteriales Incertae Sedis | NA | NA |
| Ropinirole | 109 | ASV | Firmicutes | Lachnospiraceae | Blautia | NA |
| Ropinirole | 110 | ASV | Bacteroidota | Bacteroidaceae | Bacteroides | ovatus |
| Ropinirole | 111 | ASV | Firmicutes | Lachnospiraceae | Lachnospiraceae AC2044 group | NA |
| Ropinirole | 112 | ASV | Euryarchaeota | Methanobacteriaceae | Methanobrevibacter | NA |
| Ropinirole | 113 | ASV | Firmicutes | Lactobacillaceae | Lactobacillus | delbrueckii |
| Ropinirole | 114 | ASV | Firmicutes | Anaerovoracaceae | Family XIII UCG-001 | NA |
| Ropinirole | 115 | ASV | Firmicutes | Ruminococcaceae | Incertae Sedis | NA |
| Ropinirole | 116 | ASV | Firmicutes | Oscillospiraceae | Colidextribacter | NA |
| Ropinirole | 117 | ASV | Firmicutes | Lachnospiraceae | Lachnoclostridium | NA |
| Ropinirole | 118 | ASV | Firmicutes | Ruminococcaceae | Incertae Sedis | NA |
| Ropinirole | 119 | ASV | Firmicutes | Christensenellaceae | Christensenellaceae R-7 group | NA |

| Ticker | DESeq2_baseMean | DESeq2_log2-FoldChange | DESeq2_lfcSE | DESeq2_stat | DESeq2_pvalue | DESeq2_padj | DESeq2 converged | ANCOM_W | ANCOM_detected_0.7 |
|---|---|---|---|---|---|---|---|---|---|
| 90 | 37,99489292 | -2,618606477 | 3,164440269 | -0,827510161 | 0,407947965 | 0,996273765 | YES | 0 | FALSE |
| 91 | 8,325923641 | -5,479976727 | 3,732870724 | -1,468032818 | 0,142095309 | 0,996273765 | YES | 0 | FALSE |
| 92 | 813,8455384 | -0,797526702 | 2,32025891 | -0,34372315 | 0,731054499 | 0,996273765 | YES | 0 | FALSE |
| 93 | 33,71455881 | -1,030228276 | 3,943176282 | -0,261268633 | 0,793885354 | 0,996273765 | YES | 0 | FALSE |
| 94 | 2,57285216 | 2,432351636 | 3,960537312 | 0,614146881 | 0,539118271 | 0,996273765 | YES | 0 | FALSE |
| 95 | 7,57225385 | -1,01927963 | 3,946279916 | -0,25828873 | 0,796184083 | 0,996273765 | YES | 0 | FALSE |
| 96 | 19,46971616 | 2,157893129 | 3,709919066 | 0,581655042 | 0,56079906 | 0,996273765 | YES | 0 | FALSE |
| 97 | 147,2683699 | 0,026050217 | 3,372519873 | 0,007724259 | 0,993836994 | 0,996273765 | YES | 0 | FALSE |
| 98 | 332,30271 | 0,103874624 | 3,361429043 | 0,030901924 | 0,975347756 | 0,996273765 | YES | 0 | FALSE |
| 99 | 404,511955 | -0,319885016 | 3,372857118 | -0,094840963 | 0,92444115 | 0,996273765 | YES | 0 | FALSE |
| 100 | 22,65841788 | 1,492607851 | 3,473824594 | 0,429672774 | 0,667433691 | 0,996273765 | YES | 0 | FALSE |
| 101 | 5,268857601 | -2,273171334 | 3,994305458 | -0,569103029 | 0,569286223 | 0,996273765 | YES | 0 | FALSE |
| 102 | 2,6309324 | 0,857334784 | 3,960133252 | 0,216491398 | 0,82860473 | 0,996273765 | YES | 0 | FALSE |
| 103 | 1,837751543 | 2,387655902 | 3,969232044 | 0,601541022 | 0,547479697 | 0,996273765 | YES | 0 | FALSE |
| 104 | 578,8214631 | -0,138517002 | 2,719729047 | -0,050930442 | 0,959380948 | 0,996273765 | YES | 0 | FALSE |
| 105 | 6,320062301 | 0,80719197 | 3,747134544 | 0,215415796 | 0,829443156 | 0,996273765 | YES | 0 | FALSE |
| 106 | 46,98216282 | -0,013023212 | 2,788601268 | -0,004670159 | 0,996273765 | 0,996273765 | YES | 0 | FALSE |
| 107 | 6,322629121 | -2,297342849 | 3,988143064 | -0,576043239 | 0,564585956 | 0,996273765 | YES | 0 | FALSE |
| 108 | 3,097727899 | 0,955089645 | 3,957603151 | 0,241330323 | 0,809299111 | 0,996273765 | YES | 0 | FALSE |
| 109 | 4,870041588 | 2,494735492 | 3,949265968 | 0,631695994 | 0,527585546 | 0,996273765 | YES | 0 | FALSE |
| 110 | 67,59191608 | -2,456068076 | 3,828747898 | -0,641480751 | 0,521210383 | 0,996273765 | YES | 0 | FALSE |
| 111 | 10,37684165 | 2,594148399 | 3,952045512 | 0,656406509 | 0,511562603 | 0,996273765 | YES | 0 | FALSE |
| 112 | 101,9192515 | -2,2163841 | 2,583329657 | -0,857956357 | 0,390916563 | 0,996273765 | YES | 0 | FALSE |
| 113 | 135,3358851 | 0,354713555 | 2,731417949 | 0,129864254 | 0,896673826 | 0,996273765 | YES | 0 | FALSE |
| 114 | 19,19879679 | -0,790549847 | 3,490969605 | -0,226455666 | 0,820847037 | 0,996273765 | YES | 0 | FALSE |
| 115 | 20,4025655 | 0,143208448 | 3,47980991 | 0,041154101 | 0,967173045 | 0,996273765 | YES | 0 | FALSE |
| 116 | 8,079533426 | 1,734927533 | 3,372190214 | 0,514480923 | 0,606915795 | 0,996273765 | YES | 0 | FALSE |
| 117 | 17,18597086 | -0,374082209 | 3,523571148 | -0,106165646 | 0,915450927 | 0,996273765 | YES | 0 | FALSE |
| 118 | 6,172090332 | -2,294299548 | 3,988911814 | -0,575169283 | 0,565176815 | 0,996273765 | YES | 0 | FALSE |
| 119 | 26,57182091 | -1,440527751 | 2,800229272 | -0,514432074 | 0,60694994 | 0,996273765 | YES | 0 | FALSE |

| Medication | Ticker | taxLevel | Phylum | Family | Genus | Species |
|---|---|---|---|---|---|---|
| Ropinirole | 120 | ASV | Firmicutes | Oscillospiraceae | NK4A214 group | NA |
| Ropinirole | 121 | ASV | Bacteroidota | Rikenellaceae | Alistipes | ihumii |
| Ropinirole | 122 | ASV | Firmicutes | Clostridiaceae | Clostridium sensu stricto 1 | NA |
| Ropinirole | 123 | ASV | Actinobacteriota | Eggerthellaceae | Slackia | isoflavoniconvertens |
| Ropinirole | 124 | ASV | Firmicutes | Lachnospiraceae | NA | NA |
| Ropinirole | 125 | ASV | Firmicutes | Christensenellaceae | Christensenellaceae R-7 group | NA |
| Ropinirole | 126 | ASV | Firmicutes | Erysipelotrichaceae | NA | NA |
| Ropinirole | 127 | ASV | Firmicutes | Oscillospiraceae | UCG-002 | NA |
| Ropinirole | 128 | ASV | Firmicutes | Ruminococcaceae | Negativibacillus | NA |
| Ropinirole | 129 | ASV | Firmicutes | Ruminococcaceae | Subdoligranulum | NA |
| Ropinirole | 130 | ASV | Proteobacteria | Enterobacteriaceae | Escherichia-Shigella | NA |
| Ropinirole | 131 | ASV | Firmicutes | Lachnospiraceae | GCA-900066575 | NA |
| Ropinirole | 132 | ASV | Firmicutes | Lachnospiraceae | Lachnospiraceae UCG-004 | NA |
| Ropinirole | 133 | ASV | Firmicutes | Veillonellaceae | Veillonella | NA |
| Ropinirole | 134 | ASV | Firmicutes | Oscillospiraceae | Colidextribacter | massiliensis |
| Ropinirole | 135 | ASV | Bacteroidota | Rikenellaceae | Alistipes | putredinis |
| Ropinirole | 136 | ASV | Firmicutes | Lachnospiraceae | NA | NA |
| Ropinirole | 137 | ASV | Firmicutes | Ruminococcaceae | Subdoligranulum | NA |
| Ropinirole | 138 | ASV | Firmicutes | NA | NA | NA |
| Ropinirole | 139 | ASV | Firmicutes | Oscillospiraceae | UCG-002 | NA |
| Ropinirole | 140 | ASV | Firmicutes | Christensenellaceae | Christensenellaceae R-7 group | NA |
| Ropinirole | 141 | ASV | Firmicutes | Anaerovoracaceae | Family XIII AD3011 group | NA |
| Ropinirole | 142 | ASV | Proteobacteria | NA | NA | NA |
| Ropinirole | 143 | ASV | Firmicutes | Anaerovoracaceae | [Eubacterium] brachy group | NA |
| Ropinirole | 144 | ASV | Firmicutes | Lachnospiraceae | NA | NA |
| Ropinirole | 145 | ASV | Firmicutes | Peptostreptococcaceae | Intestinibacter | bartlettii |
| Ropinirole | 146 | ASV | Firmicutes | Lachnospiraceae | [Eubacterium] ventriosum group | NA |
| Ropinirole | 147 | ASV | Bacteroidota | Barnesiellaceae | NA | NA |
| Ropinirole | 148 | ASV | Firmicutes | Lachnospiraceae | Lachnoclostridium | NA |
| Ropinirole | 149 | ASV | Firmicutes | Lachnospiraceae | Lachnotalea | NA |

| Ticker | DESeq2_baseMean | DESeq2_log2-FoldChange | DESeq2_lfcSE | DESeq2_stat | DESeq2_pvalue | DESeq2_padj | DESeq2 converged | ANCOM_W | ANCOM_detected_0.7 |
|---|---|---|---|---|---|---|---|---|---|
| 120 | 28,21300956 | 0,516839694 | 3,443580645 | 0,150087873 | 0,880695288 | 0,996273765 | YES | 0 | FALSE |
| 121 | 0,64321304 | 2,165563135 | 4,01460611 | 0,539421073 | 0,589596344 | 0,996273765 | YES | 0 | FALSE |
| 122 | 74,57984974 | -1,154229623 | 2,73995879 | -0,421258023 | 0,673566679 | 0,996273765 | YES | 0 | FALSE |
| 123 | 1,655926674 | -1,865267468 | 4,024893343 | -0,463432769 | 0,643054194 | 0,996273765 | YES | 0 | FALSE |
| 124 | 12,3989851 | 0,575399344 | 3,559325962 | 0,161659637 | 0,871573891 | 0,996273765 | YES | 0 | FALSE |
| 125 | 7,78144696 | 0,872961488 | 3,676653939 | 0,237433684 | 0,812320361 | 0,996273765 | YES | 0 | FALSE |
| 126 | 24,06435572 | 1,011031997 | 3,460705241 | 0,292146232 | 0,770174819 | 0,996273765 | YES | 0 | FALSE |
| 127 | 832,7630492 | 0,9798141 | 2,208501339 | 0,443655652 | 0,657291564 | 0,996273765 | YES | 0 | FALSE |
| 128 | 40,62664904 | 0,85912312 | 3,415665981 | 0,251524337 | 0,801408749 | 0,996273765 | YES | 0 | FALSE |
| 129 | 156,836539 | 0,339685732 | 2,74847443 | 0,123590647 | 0,901639399 | 0,996273765 | YES | 0 | FALSE |
| 130 | 105,8925823 | -3,332440196 | 3,093166791 | -1,077355481 | 0,281321485 | 0,996273765 | YES | 0 | FALSE |
| 131 | 6,437801 | -0,45167356 | 3,856447889 | -0,117121655 | 0,90676365 | 0,996273765 | YES | 0 | FALSE |
| 132 | 23,32776262 | -0,473978337 | 3,491710015 | -0,135743901 | 0,892023741 | 0,996273765 | YES | 0 | FALSE |
| 133 | 120,6733921 | 1,010419929 | 3,935208407 | 0,25676402 | 0,797360947 | 0,996273765 | YES | 0 | FALSE |
| 134 | 13,9719109 | 0,782736619 | 3,536642722 | 0,221321937 | 0,82484177 | 0,996273765 | YES | 0 | FALSE |
| 135 | 596,6671214 | -0,337808664 | 2,237490508 | -0,15097658 | 0,879994191 | 0,996273765 | YES | 0 | FALSE |
| 136 | 65,09911417 | -0,195984918 | 3,399934901 | -0,057643727 | 0,954032419 | 0,996273765 | YES | 0 | FALSE |
| 137 | 1,441228007 | 2,354235735 | 3,998173471 | 0,588827812 | 0,555976787 | 0,996273765 | YES | 0 | FALSE |
| 138 | 8,405653939 | -1,322900186 | 3,938231844 | -0,335912216 | 0,736937065 | 0,996273765 | YES | 0 | FALSE |
| 139 | 1048,595926 | -0,022986607 | 2,285588497 | -0,010057194 | 0,991975655 | 0,996273765 | YES | 0 | FALSE |
| 140 | 6,25502432 | -0,215705288 | 3,87891287 | -0,055609728 | 0,955652715 | 0,996273765 | YES | 0 | FALSE |
| 141 | 22,60629182 | -0,382594313 | 3,001884614 | -0,127451372 | 0,898583159 | 0,996273765 | YES | 0 | FALSE |
| 142 | 6,975813746 | 0,10175665 | 3,712030464 | 0,027412666 | 0,978130596 | 0,996273765 | YES | 0 | FALSE |
| 143 | 2,391027292 | -0,620240022 | 3,893535932 | -0,159299935 | 0,873432572 | 0,996273765 | YES | 0 | FALSE |
| 144 | 3,034240331 | -0,590536503 | 3,885181276 | -0,151997156 | 0,879189179 | 0,996273765 | YES | 0 | FALSE |
| 145 | 45,61025965 | -0,175676095 | 3,216904803 | -0,054610287 | 0,956448943 | 0,996273765 | YES | 0 | FALSE |
| 146 | 33,71868837 | 0,059368349 | 3,432479665 | 0,017296053 | 0,986200435 | 0,996273765 | YES | 0 | FALSE |
| 147 | 114,1415579 | 1,063079481 | 2,884788308 | 0,368512129 | 0,712491403 | 0,996273765 | YES | 0 | FALSE |
| 148 | 2,709698195 | -2,152753392 | 4,023297925 | -0,535071832 | 0,592600187 | 0,996273765 | YES | 0 | FALSE |
| 149 | 22,220056 | -0,123982924 | 3,475322921 | -0,035675224 | 0,971541326 | 0,996273765 | YES | 0 | FALSE |

| Medication | Ticker | taxLevel | Phylum | Family | Genus | Species |
|---|---|---|---|---|---|---|
| Ropinirole | 150 | ASV | Firmicutes | Lachnospiraceae | Anaerostipes | hadrus |
| Ropinirole | 151 | ASV | Firmicutes | Christensenellaceae | Christensenellaceae R-7 group | NA |
| Ropinirole | 152 | ASV | Firmicutes | Lachnospiraceae | NA | NA |
| Ropinirole | 153 | ASV | Bacteroidota | Tannerellaceae | Parabacteroides | NA |
| Ropinirole | 154 | ASV | Firmicutes | Lachnospiraceae | Agathobacter | NA |
| Ropinirole | 155 | ASV | Firmicutes | Lachnospiraceae | Roseburia | NA |
| Ropinirole | 156 | ASV | Firmicutes | Lachnospiraceae | [Eubacterium] xylanophilum group | NA |
| Ropinirole | 157 | ASV | Firmicutes | NA | NA | NA |
| Ropinirole | 158 | ASV | Firmicutes | Lachnospiraceae | Marvinbryantia | NA |
| Ropinirole | 159 | ASV | Firmicutes | Lachnospiraceae | Blautia | obeum |
| Ropinirole | 160 | ASV | Firmicutes | Ruminococcaceae | NA | NA |
| Ropinirole | 161 | ASV | Bacteroidota | Rikenellaceae | Alistipes | NA |
| Ropinirole | 162 | ASV | Bacteroidota | Rikenellaceae | Alistipes | NA |
| Ropinirole | 163 | ASV | Firmicutes | Lachnospiraceae | [Eubacterium] hallii group | NA |
| Ropinirole | 164 | ASV | Bacteroidota | Barnesiellaceae | Coprobacter | fastidiosus |
| Ropinirole | 165 | ASV | Firmicutes | Clostridiaceae | Clostridium sensu stricto 1 | NA |
| Ropinirole | 166 | ASV | Firmicutes | NA | NA | NA |
| Ropinirole | 167 | ASV | Firmicutes | NA | NA | NA |
| Ropinirole | 168 | ASV | Firmicutes | Ruminococcaceae | DTU089 | NA |
| Ropinirole | 169 | ASV | Firmicutes | Lachnospiraceae | NA | NA |
| Ropinirole | 170 | ASV | Bacteroidota | Tannerellaceae | Parabacteroides | merdae |
| Ropinirole | 171 | ASV | Bacteroidota | Rikenellaceae | Alistipes | indistinctus |
| Ropinirole | 172 | ASV | Firmicutes | Ruminococcaceae | Ruminococcus | bromii |
| Ropinirole | 173 | ASV | Firmicutes | Erysipelotrichaceae | Holdemanella | NA |
| Ropinirole | 174 | ASV | Firmicutes | Oscillospiraceae | Colidextribacter | NA |
| Ropinirole | 175 | ASV | Firmicutes | NA | NA | NA |
| Ropinirole | 176 | ASV | Firmicutes | Ruminococcaceae | Ruminococcus | bicirculans |
| Ropinirole | 177 | ASV | Firmicutes | Oscillospiraceae | Oscillibacter | NA |
| Ropinirole | 178 | ASV | Firmicutes | UCG-010 | NA | NA |
| Ropinirole | 179 | ASV | Firmicutes | NA | NA | NA |

| Ticker | DESeq2_baseMean | DESeq2_log2-FoldChange | DESeq2_lfcSE | DESeq2_stat | DESeq2_pvalue | DESeq2_padj | DESeq2 converged | ANCOM_W | ANCOM_detected_0.7 |
|---|---|---|---|---|---|---|---|---|---|
| 150 | 29,58458004 | 4,962858365 | 3,001809478 | 1,653288925 | 0,098272076 | 0,996273765 | YES | 0 | FALSE |
| 151 | 8,318861105 | 0,03992674 | 3,657187665 | 0,010917334 | 0,991289401 | 0,996273765 | YES | 0 | FALSE |
| 152 | 27,80701959 | -1,855149546 | 3,166213647 | -0,58592052 | 0,557928927 | 0,996273765 | YES | 0 | FALSE |
| 153 | 5,169606671 | 1,38868077 | 3,836221648 | 0,361991797 | 0,71735816 | 0,996273765 | YES | 0 | FALSE |
| 154 | 532,6939736 | -0,585402745 | 2,711440899 | -0,215900979 | 0,829064934 | 0,996273765 | YES | 0 | FALSE |
| 155 | 53,01676487 | 1,746123788 | 3,409399065 | 0,512150017 | 0,608546022 | 0,996273765 | YES | 0 | FALSE |
| 156 | 68,99813587 | 0,625176099 | 3,388754705 | 0,184485498 | 0,853632603 | 0,996273765 | YES | 0 | FALSE |
| 157 | 25,26908371 | 2,562214794 | 3,937486134 | 0,650723509 | 0,515224985 | 0,996273765 | YES | 0 | FALSE |
| 158 | 8,359122438 | 2,582732243 | 3,954232929 | 0,653156324 | 0,513655504 | 0,996273765 | YES | 0 | FALSE |
| 159 | 98,89352133 | -1,522408171 | 2,516919928 | -0,604869529 | 0,545265691 | 0,996273765 | YES | 0 | FALSE |
| 160 | 25,97374053 | -1,52006809 | 2,644574513 | -0,574787393 | 0,565435094 | 0,996273765 | YES | 0 | FALSE |
| 161 | 13,10899267 | -0,534177536 | 3,55316433 | -0,15033854 | 0,880497528 | 0,996273765 | YES | 0 | FALSE |
| 162 | 5,513254628 | 2,50507324 | 3,947664156 | 0,634571012 | 0,525708239 | 0,996273765 | YES | 0 | FALSE |
| 163 | 40,35163701 | 0,010933051 | 2,255033801 | 0,004848287 | 0,996131642 | 0,996273765 | YES | 0 | FALSE |
| 164 | 9,828050468 | -0,107939364 | 3,614327718 | -0,029864299 | 0,976175278 | 0,996273765 | YES | 0 | FALSE |
| 165 | 8,914269441 | 0,464949815 | 3,63666798 | 0,127850499 | 0,898267286 | 0,996273765 | YES | 0 | FALSE |
| 166 | 3,362865349 | 2,497300463 | 3,969777174 | 0,629078246 | 0,529297831 | 0,996273765 | YES | 0 | FALSE |
| 167 | 4,035438419 | 2,519215279 | 3,965674954 | 0,63525511 | 0,525262046 | 0,996273765 | YES | 0 | FALSE |
| 168 | 4,666702446 | -2,255572862 | 3,998869791 | -0,56405259 | 0,572718342 | 0,996273765 | YES | 0 | FALSE |
| 169 | 11,54856209 | -0,970981523 | 3,276245116 | -0,296370231 | 0,766947357 | 0,996273765 | YES | 0 | FALSE |
| 170 | 147,9798323 | -0,604915835 | 2,677661578 | -0,225911982 | 0,821269878 | 0,996273765 | YES | 0 | FALSE |
| 171 | 6,750517999 | 2,942399921 | 3,426610594 | 0,858691071 | 0,390510978 | 0,996273765 | YES | 0 | FALSE |
| 172 | 668,7963802 | -0,245698203 | 2,794185707 | -0,087931952 | 0,929930761 | 0,996273765 | YES | 0 | FALSE |
| 173 | 167,5637378 | 0,556983934 | 3,365770555 | 0,16548482 | 0,868562397 | 0,996273765 | YES | 0 | FALSE |
| 174 | 2,498128545 | 2,455267573 | 3,977835462 | 0,617237087 | 0,537078357 | 0,996273765 | YES | 0 | FALSE |
| 175 | 0,275662731 | 1,694135804 | 4,03746169 | 0,41960418 | 0,674774634 | 0,996273765 | YES | 0 | FALSE |
| 176 | 167,9029641 | -1,214827634 | 3,151762185 | -0,38544394 | 0,699908533 | 0,996273765 | YES | 0 | FALSE |
| 177 | 18,01623432 | 1,737243562 | 3,517668037 | 0,493862281 | 0,621403444 | 0,996273765 | YES | 0 | FALSE |
| 178 | 1,633391741 | 2,38002415 | 3,992848996 | 0,596071665 | 0,551127352 | 0,996273765 | YES | 0 | FALSE |
| 179 | 12,97105206 | 2,604665331 | 3,950172566 | 0,659380138 | 0,509651693 | 0,996273765 | YES | 0 | FALSE |

| Medication | Ticker | taxLevel | Phylum | Family | Genus | Species |
|---|---|---|---|---|---|---|
| Ropinirole | 180 | ASV | Firmicutes | Christensenellaceae | Christensenellaceae R-7 group | NA |
| Ropinirole | 181 | ASV | Firmicutes | Oscillospiraceae | NK4A214 group | NA |
| Ropinirole | 182 | ASV | Firmicutes | Lachnospiraceae | Blautia | faecis |
| Ropinirole | 183 | ASV | Bacteroidota | Marinifilaceae | Butyricimonas | virosa |
| Ropinirole | 184 | ASV | Firmicutes | UCG-010 | NA | NA |
| Ropinirole | 185 | ASV | Firmicutes | Christensenellaceae | Christensenellaceae R-7 group | NA |
| Ropinirole | 186 | ASV | Firmicutes | Christensenellaceae | Christensenellaceae R-7 group | NA |
| Ropinirole | 187 | ASV | Firmicutes | UCG-010 | NA | NA |
| Ropinirole | 188 | ASV | Bacteroidota | Bacteroidaceae | Bacteroides | NA |
| Ropinirole | 189 | ASV | Firmicutes | [Eubacterium] coprostanoligenes group | NA | NA |
| Ropinirole | 190 | ASV | Firmicutes | Veillonellaceae | Veillonella | NA |
| Ropinirole | 191 | ASV | Actinobacteriota | Atopobiaceae | Olsenella | NA |
| Ropinirole | 192 | ASV | Firmicutes | Lachnospiraceae | Coprococcus | comes |
| Ropinirole | 193 | ASV | Firmicutes | Oscillospiraceae | UCG-005 | NA |
| Ropinirole | 194 | ASV | Verrucomicrobiota | Victivallaceae | Victivallis | NA |
| Ropinirole | 195 | ASV | Actinobacteriota | Actinomycetaceae | Actinomyces | odontolyticus |
| Ropinirole | 196 | ASV | Bacteroidota | Bacteroidaceae | Bacteroides | NA |
| Ropinirole | 197 | ASV | Bacteroidota | Rikenellaceae | Alistipes | NA |
| Ropinirole | 198 | ASV | Firmicutes | Christensenellaceae | Christensenellaceae R-7 group | NA |
| Ropinirole | 199 | ASV | Firmicutes | Oscillospiraceae | Intestinimonas | massiliensis |
| Ropinirole | 200 | ASV | Bacteroidota | Bacteroidaceae | Bacteroides | caccae |
| Ropinirole | 201 | ASV | Firmicutes | Lachnospiraceae | [Eubacterium] xylanophilum group | NA |
| Ropinirole | 202 | ASV | Firmicutes | Lachnospiraceae | Howardella | ureilytica |
| Ropinirole | 203 | ASV | Bacteroidota | Bacteroidaceae | Bacteroides | NA |
| Ropinirole | 204 | ASV | Firmicutes | Anaerovoracaceae | Family XIII AD3011 group | NA |
| Ropinirole | 205 | ASV | Firmicutes | Lachnospiraceae | Eisenbergiella | tayi |
| Ropinirole | 206 | ASV | Firmicutes | Lachnospiraceae | Blautia | NA |
| Ropinirole | 207 | ASV | Bacteroidota | Bacteroidaceae | Bacteroides | nordii |
| Ropinirole | 208 | ASV | Firmicutes | Lachnospiraceae | Marvinbryantia | NA |
| Ropinirole | 209 | ASV | Firmicutes | Oscillospiraceae | UCG-005 | NA |

| Ticker | DESeq2_baseMean | DESeq2_log2-FoldChange | DESeq2_lfcSE | DESeq2_stat | DESeq2_pvalue | DESeq2_padj | DESeq2 converged | ANCOM_W | ANCOM_detected_0.7 |
|---|---|---|---|---|---|---|---|---|---|
| 180 | 1,411867977 | 3,870572725 | 3,97250939 | 0,974339478 | 0,329888009 | 0,996273765 | YES | 0 | FALSE |
| 181 | 61,49209071 | -0,80222396 | 2,754475675 | -0,291243799 | 0,770864867 | 0,996273765 | YES | 0 | FALSE |
| 182 | 222,9423771 | 1,136072168 | 3,015095007 | 0,376794816 | 0,706326094 | 0,996273765 | YES | 0 | FALSE |
| 183 | 12,10162028 | 5,386518279 | 3,134895035 | 1,718245179 | 0,085751904 | 0,996273765 | YES | 0 | FALSE |
| 184 | 8,867699966 | 0,264078326 | 3,639502416 | 0,072558909 | 0,942157126 | 0,996273765 | YES | 0 | FALSE |
| 185 | 2,709698195 | -2,152753392 | 4,023297925 | -0,535071832 | 0,592600187 | 0,996273765 | YES | 0 | FALSE |
| 186 | 2,687163261 | -0,184823503 | 3,896323429 | -0,04743536 | 0,962166248 | 0,996273765 | YES | 0 | FALSE |
| 187 | 58,50497648 | 0,71173631 | 2,751185931 | 0,258701639 | 0,795865455 | 0,996273765 | YES | 0 | FALSE |
| 188 | 350,1142862 | -0,979213343 | 3,358166726 | -0,29159164 | 0,770598868 | 0,996273765 | YES | 0 | FALSE |
| 189 | 26,38571486 | -2,176436769 | 3,143940452 | -0,692263992 | 0,488771558 | 0,996273765 | YES | 0 | FALSE |
| 190 | 8,881788527 | -2,335525476 | 3,978653942 | -0,587013978 | 0,557194319 | 0,996273765 | YES | 0 | FALSE |
| 191 | 3,719904428 | 1,356641148 | 3,912616009 | 0,34673506 | 0,728790368 | 0,996273765 | YES | 0 | FALSE |
| 192 | 91,7422339 | 1,488568694 | 2,599112227 | 0,572721977 | 0,566832954 | 0,996273765 | YES | 0 | FALSE |
| 193 | 65,99196383 | 2,419397094 | 2,597256961 | 0,931520112 | 0,351584586 | 0,996273765 | YES | 0 | FALSE |
| 194 | 1,272492496 | 1,726521729 | 3,911591514 | 0,441385999 | 0,658933576 | 0,996273765 | YES | 0 | FALSE |
| 195 | 1,935908323 | 0,136177636 | 3,959440255 | 0,034393153 | 0,972563643 | 0,996273765 | YES | 0 | FALSE |
| 196 | 271,7800089 | -1,617256092 | 2,211946397 | -0,73114615 | 0,464689888 | 0,996273765 | YES | 0 | FALSE |
| 197 | 76,84794037 | -2,511715086 | 2,607641838 | -0,963213218 | 0,335440533 | 0,996273765 | YES | 0 | FALSE |
| 198 | 24,64193245 | 0,158317577 | 3,458014906 | 0,045782792 | 0,963483375 | 0,996273765 | YES | 0 | FALSE |
| 199 | 44,56095926 | -0,376894124 | 3,422443711 | -0,110124273 | 0,912310818 | 0,996273765 | YES | 0 | FALSE |
| 200 | 203,6058767 | -1,757143628 | 2,720585007 | -0,645869776 | 0,518363695 | 0,996273765 | YES | 0 | FALSE |
| 201 | 28,75569958 | -0,517206876 | 3,755916376 | -0,137704577 | 0,890473902 | 0,996273765 | YES | 0 | FALSE |
| 202 | 12,3594249 | 0,420275629 | 2,910638615 | 0,144392927 | 0,885190202 | 0,996273765 | YES | 0 | FALSE |
| 203 | 156,0403863 | 0,150521599 | 3,299569328 | 0,045618559 | 0,963614276 | 0,996273765 | YES | 0 | FALSE |
| 204 | 3,33557696 | -1,293790695 | 3,943592862 | -0,328074104 | 0,742855633 | 0,996273765 | YES | 0 | FALSE |
| 205 | 9,339615487 | -1,324636168 | 3,937878673 | -0,336383184 | 0,736581928 | 0,996273765 | YES | 0 | FALSE |
| 206 | 298,3218128 | 0,216889836 | 2,221611275 | 0,097627266 | 0,922228273 | 0,996273765 | YES | 0 | FALSE |
| 207 | 5,722969127 | 5,032180531 | 3,80933406 | 1,321013188 | 0,186496967 | 0,996273765 | YES | 0 | FALSE |
| 208 | 10,92677978 | -0,814085589 | 3,755072955 | -0,216796211 | 0,828367165 | 0,996273765 | YES | 0 | FALSE |
| 209 | 223,8674951 | 0,742835432 | 2,21124064 | 0,335936043 | 0,736919096 | 0,996273765 | YES | 0 | FALSE |

| Medication | Ticker | taxLevel | Phylum | Family | Genus | Species |
|---|---|---|---|---|---|---|
| Ropinirole | 210 | ASV | Proteobacteria | Oxalobacteraceae | Oxalobacter | formigenes |
| Ropinirole | 211 | ASV | Firmicutes | Lachnospiraceae | Blautia | NA |
| Ropinirole | 212 | ASV | Firmicutes | Lachnospiraceae | Coprococcus | eutactus |
| Ropinirole | 213 | ASV | Firmicutes | Lachnospiraceae | [Eubacterium] ventriosum group | NA |
| Ropinirole | 214 | ASV | Firmicutes | UCG-010 | NA | NA |
| Ropinirole | 215 | ASV | Firmicutes | Lachnospiraceae | NA | NA |
| Ropinirole | 216 | ASV | Firmicutes | Lachnospiraceae | [Ruminococcus] torques group | NA |
| Ropinirole | 217 | ASV | Firmicutes | Veillonellaceae | Allisonella | histaminiformans |
| Ropinirole | 218 | ASV | Firmicutes | Ruminococcaceae | Negativibacillus | massiliensis |
| Ropinirole | 219 | ASV | Firmicutes | Erysipelatoclostridiaceae | Asteroleplasma | NA |
| Ropinirole | 220 | ASV | Bacteroidota | Marinifilaceae | Butyricimonas | NA |
| Ropinirole | 221 | ASV | Bacteroidota | Marinifilaceae | Butyricimonas | NA |
| Ropinirole | 222 | ASV | Bacteroidota | Bacteroidaceae | Bacteroides | NA |
| Ropinirole | 223 | ASV | Proteobacteria | Sutterellaceae | Sutterella | NA |
| Ropinirole | 224 | ASV | Firmicutes | Monoglobaceae | Monoglobus | NA |
| Ropinirole | 225 | ASV | Firmicutes | Lachnospiraceae | NA | NA |
| Ropinirole | 226 | ASV | Firmicutes | Oscillospiraceae | UCG-005 | NA |
| Ropinirole | 227 | ASV | Bacteroidota | Bacteroidaceae | Bacteroides | massiliensis |
| Ropinirole | 228 | ASV | Firmicutes | Lachnospiraceae | Roseburia | inulinivorans |
| Ropinirole | 229 | ASV | Firmicutes | Erysipelatoclostridiaceae | Erysipelatoclostridium | NA |
| Ropinirole | 230 | ASV | Bacteroidota | Bacteroidaceae | Bacteroides | NA |
| Ropinirole | 231 | ASV | Firmicutes | Anaerovoracaceae | Family XIII AD3011 group | NA |
| Ropinirole | 232 | ASV | Firmicutes | [Eubacterium] coprostanoligenes group | NA | NA |
| Ropinirole | 233 | ASV | Firmicutes | Ruminococcaceae | Faecalibacterium | NA |
| Ropinirole | 234 | ASV | Firmicutes | Ruminococcaceae | [Eubacterium] siraeum group | NA |
| Ropinirole | 235 | ASV | Firmicutes | Lachnospiraceae | NA | NA |
| Ropinirole | 236 | ASV | Firmicutes | Erysipelatoclostridiaceae | Catenibacterium | mitsuokai |
| Ropinirole | 237 | ASV | Firmicutes | NA | NA | NA |
| Ropinirole | 238 | ASV | Firmicutes | NA | NA | NA |
| Ropinirole | 239 | ASV | Actinobacteriota | Eggerthellaceae | NA | NA |

| Ticker | DESeq2_baseMean | DESeq2_log2-FoldChange | DESeq2_lfcSE | DESeq2_stat | DESeq2_pvalue | DESeq2_padj | DESeq2 converged | ANCOM_W | ANCOM_detected_0.7 |
|---|---|---|---|---|---|---|---|---|---|
| 210 | 4,166803908 | 2,438688611 | 3,558452136 | 0,685322865 | 0,493140207 | 0,996273765 | YES | 0 | FALSE |
| 211 | 5,538238425 | 0,821065863 | 3,800140359 | 0,216061983 | 0,828939433 | 0,996273765 | YES | 0 | FALSE |
| 212 | 29,2088876 | 2,636368639 | 3,915291716 | 0,673351778 | 0,500723519 | 0,996273765 | YES | 0 | FALSE |
| 213 | 15,05387886 | -2,382752391 | 3,936920612 | -0,605232522 | 0,54502451 | 0,996273765 | YES | 0 | FALSE |
| 214 | 3,362865349 | 2,497300463 | 3,969777174 | 0,629078246 | 0,529297831 | 0,996273765 | YES | 0 | FALSE |
| 215 | 2,408620617 | -2,083529226 | 4,023626536 | -0,517823711 | 0,604581273 | 0,996273765 | YES | 0 | FALSE |
| 216 | 124,8641022 | -1,255249494 | 2,567044216 | -0,488986316 | 0,624851385 | 0,996273765 | YES | 0 | FALSE |
| 217 | 12,95684549 | 1,068191494 | 3,01022229 | 0,354854689 | 0,722698455 | 0,996273765 | YES | 0 | FALSE |
| 218 | 12,73774694 | 1,000304037 | 3,956542312 | 0,252822783 | 0,800405162 | 0,996273765 | YES | 0 | FALSE |
| 219 | 1,105494502 | 0,192006663 | 3,969156824 | 0,048374673 | 0,961417644 | 0,996273765 | YES | 0 | FALSE |
| 220 | 2,935307725 | -1,288905772 | 3,944739677 | -0,326740388 | 0,74386425 | 0,996273765 | YES | 0 | FALSE |
| 221 | 23,2449133 | -0,951318118 | 2,815521282 | -0,337883476 | 0,735451 | 0,996273765 | YES | 0 | FALSE |
| 222 | 112,8103918 | 0,863542165 | 2,727215267 | 0,316638798 | 0,751517693 | 0,996273765 | YES | 0 | FALSE |
| 223 | 28,02571102 | 2,56451672 | 3,930407419 | 0,652481142 | 0,514090835 | 0,996273765 | YES | 0 | FALSE |
| 224 | 98,03847048 | -1,963447455 | 2,526828172 | -0,777040353 | 0,437134963 | 0,996273765 | YES | 2 | FALSE |
| 225 | 2,017719209 | 2,419778618 | 3,984821939 | 0,607248869 | 0,543685765 | 0,996273765 | YES | 0 | FALSE |
| 226 | 21,81058383 | 2,624325295 | 3,938258288 | 0,666366983 | 0,505176561 | 0,996273765 | YES | 0 | FALSE |
| 227 | 31,16965165 | -0,583730449 | 2,877064848 | -0,20289096 | 0,83922026 | 0,996273765 | YES | 0 | FALSE |
| 228 | 173,2381547 | -0,537957619 | 2,881871781 | -0,186669519 | 0,851919756 | 0,996273765 | YES | 0 | FALSE |
| 229 | 3,362026684 | -0,172100856 | 3,955946469 | -0,043504344 | 0,965299502 | 0,996273765 | YES | 0 | FALSE |
| 230 | 184,355587 | -0,090669988 | 3,3708216 | -0,026898483 | 0,978540703 | 0,996273765 | YES | 0 | FALSE |
| 231 | 1,983209342 | -0,981751936 | 3,957215839 | -0,248091582 | 0,804063549 | 0,996273765 | YES | 0 | FALSE |
| 232 | 44,24691043 | 1,008760138 | 3,936756377 | 0,256241444 | 0,797764409 | 0,996273765 | YES | 0 | FALSE |
| 233 | 629,0680022 | 0,348074578 | 2,202102299 | 0,158064672 | 0,874405838 | 0,996273765 | YES | 0 | FALSE |
| 234 | 450,5266071 | 4,186859843 | 2,222664227 | 1,883712255 | 0,059603908 | 0,996273765 | YES | 2 | FALSE |
| 235 | 3,951165816 | 2,479597967 | 3,952275999 | 0,62738482 | 0,530407016 | 0,996273765 | YES | 0 | FALSE |
| 236 | 164,8782222 | 0,115009683 | 3,367731975 | 0,034150486 | 0,97275715 | 0,996273765 | YES | 0 | FALSE |
| 237 | 31,31206803 | -2,431137717 | 3,85352688 | -0,630886404 | 0,528114802 | 0,996273765 | YES | 0 | FALSE |
| 238 | 14,20221341 | 0,901827702 | 3,533915662 | 0,255192197 | 0,798574658 | 0,996273765 | YES | 0 | FALSE |
| 239 | 56,86391424 | 0,27239746 | 2,746327877 | 0,099186067 | 0,920990537 | 0,996273765 | YES | 0 | FALSE |

| Medication | Ticker | taxLevel | Phylum | Family | Genus | Species |
|---|---|---|---|---|---|---|
| Ropinirole | 240 | ASV | Firmicutes | Oscillospiraceae | UCG-002 | NA |
| Ropinirole | 241 | ASV | Firmicutes | Ruminococcaceae | Incertae Sedis | NA |
| Ropinirole | 242 | ASV | Firmicutes | Oscillospiraceae | Flavonifractor | NA |
| Ropinirole | 243 | ASV | Firmicutes | Butyricicoccaceae | Butyricicoccus | NA |
| Ropinirole | 244 | ASV | Bacteroidota | Prevotellaceae | NA | NA |
| Ropinirole | 245 | ASV | Firmicutes | Ruminococcaceae | Faecalibacterium | NA |
| Ropinirole | 246 | ASV | Firmicutes | Ruminococcaceae | DTU089 | NA |
| Ropinirole | 247 | ASV | Firmicutes | Christensenellaceae | NA | NA |
| Ropinirole | 248 | ASV | Firmicutes | Ruminococcaceae | Fournierella | NA |
| Ropinirole | 249 | ASV | Firmicutes | UCG-010 | NA | NA |
| Ropinirole | 250 | ASV | Firmicutes | Lachnospiraceae | Agathobacter | NA |
| Ropinirole | 251 | ASV | Firmicutes | Lachnospiraceae | Roseburia | intestinalis |
| Ropinirole | 252 | ASV | Firmicutes | Lachnospiraceae | Lachnoclostridium | NA |
| Ropinirole | 253 | ASV | Firmicutes | Hydrogenoanaerobacterium | NA | NA |
| Ropinirole | 254 | ASV | Firmicutes | [Eubacterium] coprostanoligenes group | NA | NA |
| Ropinirole | 255 | ASV | Firmicutes | Oscillospiraceae | Oscillibacter | NA |
| Ropinirole | 256 | ASV | Verrucomicrobiota | vadinBE97 | NA | NA |
| Ropinirole | 257 | ASV | Firmicutes | Oscillospiraceae | UCG-005 | NA |
| Ropinirole | 258 | ASV | Firmicutes | Lachnospiraceae | Coprococcus | catus |
| Ropinirole | 259 | ASV | Firmicutes | Lachnospiraceae | [Ruminococcus] gauvreauii group | NA |
| Ropinirole | 260 | ASV | Verrucomicrobiota | Puniceicoccaceae | NA | NA |
| Ropinirole | 261 | ASV | Firmicutes | Lachnospiraceae | Lachnospiraceae ND3007 group | NA |
| Ropinirole | 262 | ASV | Firmicutes | Lachnospiraceae | [Ruminococcus] torques group | NA |
| Ropinirole | 263 | ASV | Firmicutes | Lachnospiraceae | [Eubacterium] eligens group | NA |
| Ropinirole | 264 | ASV | Firmicutes | Ruminococcaceae | Faecalibacterium | prausnitzii |
| Ropinirole | 265 | ASV | Firmicutes | Acidaminococcaceae | Phascolarctobacterium | faecium |
| Ropinirole | 266 | ASV | Firmicutes | Peptococcaceae | NA | NA |
| Ropinirole | 267 | ASV | Desulfobacterota | Desulfovibrionaceae | NA | NA |
| Ropinirole | 268 | ASV | Firmicutes | Lachnospiraceae | Lachnospiraceae NK4A136 group | NA |
| Ropinirole | 269 | ASV | Firmicutes | Ruminococcaceae | NA | NA |

| Ticker | DESeq2_baseMean | DESeq2_log2-FoldChange | DESeq2_lfcSE | DESeq2_stat | DESeq2_pvalue | DESeq2_padj | DESeq2 converged | ANCOM_W | ANCOM_detected_0.7 |
|---|---|---|---|---|---|---|---|---|---|
| 240 | 261,6859641 | 0,674808049 | 2,72418022 | 0,247710502 | 0,804358406 | 0,996273765 | YES | 0 | FALSE |
| 241 | 111,8646858 | 0,354296079 | 2,687233079 | 0,131844194 | 0,89510753 | 0,996273765 | YES | 0 | FALSE |
| 242 | 4,333801902 | 1,080437108 | 3,915652738 | 0,275927714 | 0,782603587 | 0,996273765 | YES | 0 | FALSE |
| 243 | 113,1693386 | 0,790194975 | 2,215831807 | 0,356613247 | 7,21E-01 | 9,96E-01 | YES | 0 | FALSE |
| 244 | 139,9907637 | 1,206385585 | 3,370196513 | 0,357957045 | 0,72037546 | 0,996273765 | YES | 0 | FALSE |
| 245 | 98,9610722 | -0,977389212 | 2,738765086 | -0,356872233 | 0,721187458 | 0,996273765 | YES | 0 | FALSE |
| 246 | 16,64121602 | 2,779664523 | 3,194985685 | 0,870008444 | 0,38429579 | 0,996273765 | YES | 1 | FALSE |
| 247 | 240,6254946 | -0,483811512 | 3,38007628 | -0,14313627 | 0,88618256 | 0,996273765 | YES | 0 | FALSE |
| 248 | 0,864736804 | 2,267221516 | 4,022389027 | 0,563650483 | 0,572992023 | 0,996273765 | YES | 0 | FALSE |
| 249 | 4,022446403 | 0,982923246 | 3,968589379 | 0,247675724 | 0,804385317 | 0,996273765 | YES | 0 | FALSE |
| 250 | 649,4738201 | -1,225113589 | 2,791165583 | -0,438925443 | 0,660715561 | 0,996273765 | YES | 0 | FALSE |
| 251 | 87,08091928 | -1,032243843 | 3,942618972 | -0,26181679 | 0,793462694 | 0,996273765 | YES | 0 | FALSE |
| 252 | 7,902331633 | 2,524739698 | 3,944014985 | 0,64014455 | 0,522078628 | 0,996273765 | YES | 0 | FALSE |
| 253 | 4,9366539 | -1,305716238 | 3,940830999 | -0,331330179 | 0,740395098 | 0,996273765 | YES | 0 | FALSE |
| 254 | 93,82993685 | 0,182478923 | 3,379493596 | 0,053995937 | 0,956938401 | 0,996273765 | YES | 0 | FALSE |
| 255 | 4,318716125 | 2,485224363 | 3,950953797 | 0,629018837 | 0,529336724 | 0,996273765 | YES | 0 | FALSE |
| 256 | 0,64321304 | 2,165563135 | 4,01460611 | 0,539421073 | 0,589596344 | 0,996273765 | YES | 0 | FALSE |
| 257 | 126,6456263 | 4,39876736 | 3,067930946 | 1,43378956 | 0,151632325 | 0,996273765 | YES | 0 | FALSE |
| 258 | 41,35040522 | -0,507680892 | 2,766247493 | -0,183526924 | 0,854384596 | 0,996273765 | YES | 0 | FALSE |
| 259 | 47,46343103 | 2,109814973 | 3,745377689 | 0,563311673 | 0,573222671 | 0,996273765 | YES | 0 | FALSE |
| 260 | 3,675503085 | 2,470936726 | 3,953509691 | 0,624998272 | 0,531972192 | 0,996273765 | YES | 0 | FALSE |
| 261 | 40,87751298 | -1,966718612 | 2,586950856 | -0,760245834 | 0,447107652 | 0,996273765 | YES | 0 | FALSE |
| 262 | 157,5810915 | -0,138444895 | 3,371133753 | -0,041067755 | 0,967241881 | 0,996273765 | YES | 0 | FALSE |
| 263 | 20,78455532 | -0,123974305 | 3,481322393 | -0,035611268 | 0,971592323 | 0,996273765 | YES | 0 | FALSE |
| 264 | 767,4170769 | -0,568626169 | 2,265765893 | -0,250964219 | 0,801841773 | 0,996273765 | YES | 0 | FALSE |
| 265 | 1,655926674 | -1,865267468 | 4,024893343 | -0,463432769 | 0,643054194 | 0,996273765 | YES | 0 | FALSE |
| 266 | 6,198242023 | 0,120828951 | 3,766686643 | 0,032078312 | 0,974409599 | 0,996273765 | YES | 0 | FALSE |
| 267 | 1,010763348 | 2,27509798 | 3,99118638 | 0,570030503 | 0,56865701 | 0,996273765 | YES | 0 | FALSE |
| 268 | 334,3969195 | 1,577716121 | 2,735427621 | 0,576771291 | 0,564093966 | 0,996273765 | YES | 0 | FALSE |
| 269 | 8,342763457 | -0,144758502 | 3,6588692 | -0,039563727 | 0,968440947 | 0,996273765 | YES | 0 | FALSE |

| Medication | Ticker | taxLevel | Phylum | Family | Genus | Species |
|---|---|---|---|---|---|---|
| Ropinirole | 270 | ASV | Firmicutes | Oscillospiraceae | Colidextribacter | NA |
| Ropinirole | 271 | ASV | Firmicutes | Lachnospiraceae | GCA-900066575 | NA |
| Ropinirole | 272 | ASV | Firmicutes | Lachnospiraceae | Lachnospiraceae NK4A136 group | NA |
| Ropinirole | 273 | ASV | Bacteroidota | Bacteroidaceae | Bacteroides | NA |
| Ropinirole | 274 | ASV | Firmicutes | Lachnospiraceae | Coprococcus | NA |
| Ropinirole | 275 | ASV | Firmicutes | Lachnospiraceae | [Eubacterium] hallii group | NA |
| Ropinirole | 276 | ASV | Proteobacteria | Sutterellaceae | Sutterella | wadsworthensis |
| Ropinirole | 277 | ASV | Firmicutes | Lachnospiraceae | Dorea | formicigenerans |
| Ropinirole | 278 | ASV | Firmicutes | Lachnospiraceae | Shuttleworthia | NA |
| Ropinirole | 279 | ASV | Firmicutes | Lachnospiraceae | Blautia | NA |
| Ropinirole | 280 | ASV | Desulfobacterota | Desulfovibrionaceae | NA | NA |
| Ropinirole | 281 | ASV | Bacteroidota | Barnesiellaceae | Coprobacter | secundus |
| Ropinirole | 282 | ASV | Bacteroidota | Bacteroidaceae | Bacteroides | NA |
| Ropinirole | 283 | ASV | Firmicutes | Lachnospiraceae | [Eubacterium] eligens group | NA |
| Ropinirole | 284 | ASV | Firmicutes | Erysipelotrichaceae | Turicibacter | sanguinis |
| Ropinirole | 285 | ASV | Firmicutes | Oscillospiraceae | NK4A214 group | NA |
| Ropinirole | 286 | ASV | Bacteroidota | Rikenellaceae | Alistipes | shahii |
| Ropinirole | 287 | ASV | Firmicutes | Lachnospiraceae | CAG-56 | NA |
| Ropinirole | 288 | ASV | Firmicutes | Lachnospiraceae | [Ruminococcus] gnavus group | NA |
| Ropinirole | 289 | ASV | Firmicutes | Acidaminococcaceae | Phascolarctobacterium | succinatutens |
| Ropinirole | 290 | Genus | Actinobacteriota | Bifidobacteriaceae | Bifidobacterium | NA |
| Ropinirole | 291 | Genus | Verrucomicrobiota | Akkermansiaceae | Akkermansia | NA |
| Ropinirole | 292 | Genus | Firmicutes | Lachnospiraceae | Lachnospira | NA |
| Ropinirole | 293 | Genus | Firmicutes | Oscillospiraceae | UCG-003 | NA |
| Ropinirole | 294 | Genus | Firmicutes | Veillonellaceae | Dialister | NA |
| Ropinirole | 295 | Genus | Firmicutes | Oscillospiraceae | NK4A214 group | NA |
| Ropinirole | 296 | Genus | Firmicutes | Lachnospiraceae | Fusicatenibacter | NA |
| Ropinirole | 297 | Genus | Actinobacteriota | Eggerthellaceae | Senegalimassilia | NA |
| Ropinirole | 298 | Genus | Actinobacteriota | Eggerthellaceae | Enterorhabdus | NA |
| Ropinirole | 299 | Genus | Firmicutes | Lachnospiraceae | Lachnoclostridium | NA |

| Ticker | DESeq2_baseMean | DESeq2_log2-FoldChange | DESeq2_lfcSE | DESeq2_stat | DESeq2_pvalue | DESeq2_padj | DESeq2 converged | ANCOM_W | ANCOM_detected_0.7 |
|---|---|---|---|---|---|---|---|---|---|
| 270 | 4,002692352 | -1,299896114 | 3,942178861 | -0,329740522 | 0,741596033 | 0,996273765 | YES | 0 | FALSE |
| 271 | 10,98360084 | 0,389232444 | 3,58468091 | 0,108582173 | 0,913533901 | 0,996273765 | YES | 0 | FALSE |
| 272 | 126,9372717 | 0,606962223 | 2,998922666 | 0,202393423 | 0,83960917 | 0,996273765 | YES | 0 | FALSE |
| 273 | 168,508339 | -1,160346293 | 3,249971123 | -0,357032801 | 0,72106725 | 0,996273765 | YES | 0 | FALSE |
| 274 | 122,0142076 | 1,010401266 | 3,935379823 | 0,256748093 | 0,797373243 | 0,996273765 | YES | 0 | FALSE |
| 275 | 19,6223459 | -0,492787023 | 3,48715051 | -0,1413151 | 0,887621021 | 0,996273765 | YES | 0 | FALSE |
| 276 | 18,95763385 | 0,820331823 | 2,905279266 | 0,282359026 | 0,77766823 | 0,996273765 | YES | 0 | FALSE |
| 277 | 42,0828179 | -1,933908619 | 3,161443172 | -0,611717027 | 0,540724994 | 0,996273765 | YES | 0 | FALSE |
| 278 | 10,13780504 | 1,19370599 | 3,606898834 | 0,330950782 | 0,740681663 | 0,996273765 | YES | 0 | FALSE |
| 279 | 59,54103669 | -0,835727698 | 3,651881235 | -0,228848542 | 0,818986636 | 0,996273765 | YES | 0 | FALSE |
| 280 | 4,301082976 | 1,404196928 | 3,925772705 | 0,357686762 | 0,720577741 | 0,996273765 | YES | 0 | FALSE |
| 281 | 5,632766388 | 0,190235214 | 3,791831106 | 0,050169749 | 0,959987118 | 0,996273765 | YES | 0 | FALSE |
| 282 | 1396,578398 | -1,831573654 | 2,195036905 | -0,834415882 | 0,404046671 | 0,996273765 | YES | 0 | FALSE |
| 283 | 39,02358076 | -2,358904822 | 3,111170593 | -0,758204911 | 0,448328322 | 0,996273765 | YES | 0 | FALSE |
| 284 | 7,231184968 | -0,857061227 | 3,853248932 | -0,222425606 | 0,823982581 | 0,996273765 | YES | 0 | FALSE |
| 285 | 69,29626908 | 1,923657044 | 3,10445158 | 0,619644724 | 0,535491715 | 0,996273765 | YES | 0 | FALSE |
| 286 | 85,09983947 | 0,676067737 | 3,381210008 | 0,199948461 | 0,841520889 | 0,996273765 | YES | 0 | FALSE |
| 287 | 32,54258343 | -5,969174594 | 3,468511919 | -1,720961246 | 0,085257857 | 0,996273765 | YES | 0 | FALSE |
| 288 | 300,8338418 | 0,398772826 | 3,370897493 | 0,11829871 | 0,905830979 | 0,996273765 | YES | 0 | FALSE |
| 289 | 31,70473749 | 0,628713467 | 3,433380543 | 0,183117909 | 0,854705504 | 0,996273765 | YES | 0 | FALSE |
| 290 | 775,0952342 | -0,497761622 | 1,98050441 | -0,251330731 | 0,801558417 | 0,998915527 | YES | 0 | FALSE |
| 291 | 220,900072 | 1,790254039 | 1,913346822 | 0,935666246 | 0,349445054 | 0,998915527 | YES | 0 | FALSE |
| 292 | 66,9975868 | 1,497482466 | 2,232000982 | 0,670914788 | 0,502274815 | 0,998915527 | YES | 0 | FALSE |
| 293 | 292,1278078 | 0,373792178 | 2,024598028 | 0,184625379 | 0,853522879 | 0,998915527 | YES | 0 | FALSE |
| 294 | 1025,394869 | 0,823185216 | 2,592727082 | 0,317497828 | 0,750865888 | 0,998915527 | YES | 0 | FALSE |
| 295 | 477,7791718 | 0,14898011 | 1,553230979 | 0,095916262 | 0,923587079 | 0,998915527 | YES | 0 | FALSE |
| 296 | 443,5395504 | 0,306792019 | 2,013274315 | 0,152384609 | 0,878883596 | 0,998915527 | YES | 0 | FALSE |
| 297 | 14,42728858 | 1,973947467 | 2,377776956 | 0,83016511 | 0,406445439 | 0,998915527 | YES | 0 | FALSE |
| 298 | 8,715536085 | -3,017833863 | 3,22478562 | -0,935824646 | 0,34936348 | 0,998915527 | YES | 0 | FALSE |
| 299 | 110,8206978 | -0,308346225 | 1,660959288 | -0,185643457 | 0,852724372 | 0,998915527 | YES | 0 | FALSE |

| Medication | Ticker | taxLevel | Phylum | Family | Genus | Species |
|---|---|---|---|---|---|---|
| Ropinirole | 300 | Genus | Firmicutes | Peptostreptococcaceae | Terrisporobacter | NA |
| Ropinirole | 301 | Genus | Desulfobacterota | Desulfovibrionaceae | Bilophila | NA |
| Ropinirole | 302 | Genus | Firmicutes | Oscillospiraceae | Oscillibacter | NA |
| Ropinirole | 303 | Genus | Firmicutes | Christensenellaceae | Christensenellaceae R-7 group | NA |
| Ropinirole | 304 | Genus | Firmicutes | Erysipelatoclostridiaceae | UCG-004 | NA |
| Ropinirole | 305 | Genus | Firmicutes | Lachnospiraceae | [Eubacterium] ventriosum group | NA |
| Ropinirole | 306 | Genus | Firmicutes | Peptostreptococcaceae | Romboutsia | NA |
| Ropinirole | 307 | Genus | Desulfobacterota | Desulfovibrionaceae | Desulfovibrio | NA |
| Ropinirole | 308 | Genus | Firmicutes | Ruminococcaceae | Paludicola | NA |
| Ropinirole | 309 | Genus | Firmicutes | Oscillospiraceae | Intestinimonas | NA |
| Ropinirole | 310 | Genus | Actinobacteriota | Coriobacteriaceae | Collinsella | NA |
| Ropinirole | 311 | Genus | Proteobacteria | Pasteurellaceae | Haemophilus | NA |
| Ropinirole | 312 | Genus | Firmicutes | Erysipelatoclostridiaceae | Asteroleplasma | NA |
| Ropinirole | 313 | Genus | Firmicutes | Lachnospiraceae | Dorea | NA |
| Ropinirole | 314 | Genus | Firmicutes | Erysipelatoclostridiaceae | Erysipelotrichaceae UCG-003 | NA |
| Ropinirole | 315 | Genus | Firmicutes | Ruminococcaceae | UBA1819 | NA |
| Ropinirole | 316 | Genus | Bacteroidota | Marinifilaceae | Odoribacter | NA |
| Ropinirole | 317 | Genus | Actinobacteriota | Eggerthellaceae | Adlercreutzia | NA |
| Ropinirole | 318 | Genus | Bacteroidota | Barnesiellaceae | Barnesiella | NA |
| Ropinirole | 319 | Genus | Bacteroidota | Bacteroidaceae | Bacteroides | NA |
| Ropinirole | 320 | Genus | Bacteroidota | Prevotellaceae | Prevotella | NA |
| Ropinirole | 321 | Genus | Firmicutes | Lachnospiraceae | [Eubacterium] xylanophilum group | NA |
| Ropinirole | 322 | Genus | Firmicutes | Lachnospiraceae | Lachnospiraceae FCS020 group | NA |
| Ropinirole | 323 | Genus | Firmicutes | Lachnospiraceae | Lachnospiraceae UCG-010 | NA |
| Ropinirole | 324 | Genus | Firmicutes | Peptococcaceae | Peptococcus | NA |
| Ropinirole | 325 | Genus | Firmicutes | Lachnospiraceae | Lachnospiraceae UCG-004 | NA |
| Ropinirole | 326 | Genus | Firmicutes | Lachnospiraceae | Lachnospiraceae UCG-001 | NA |
| Ropinirole | 327 | Genus | Firmicutes | Oscillospiraceae | Flavonifractor | NA |
| Ropinirole | 328 | Genus | Firmicutes | Streptococcaceae | Streptococcus | NA |
| Ropinirole | 329 | Genus | Bacteroidota | Prevotellaceae | Alloprevotella | NA |

| Ticker | DESeq2_baseMean | DESeq2_log2-FoldChange | DESeq2_lfcSE | DESeq2_stat | DESeq2_pvalue | DESeq2_padj | DESeq2 converged | ANCOM_W | ANCOM_detected_0.7 |
|---|---|---|---|---|---|---|---|---|---|
| 300 | 12,69970671 | -2,271430813 | 3,912072486 | -0,58062084 | 0,561496024 | 0,998915527 | YES | 0 | FALSE |
| 301 | 26,56160531 | -1,534039168 | 2,543869928 | -0,60303365 | 0,546486304 | 0,998915527 | YES | 0 | FALSE |
| 302 | 88,48221631 | -1,131235729 | 2,016925317 | -0,560871402 | 0,574885207 | 0,998915527 | YES | 0 | FALSE |
| 303 | 262,6169075 | 0,225057025 | 2,113905895 | 0,106465016 | 0,915213411 | 0,998915527 | YES | 0 | FALSE |
| 304 | 0,728481251 | -0,961475044 | 3,980123303 | -0,24156916 | 0,809114021 | 0,998915527 | YES | 0 | FALSE |
| 305 | 391,2471127 | -2,475101527 | 1,594272387 | -1,552496014 | 0,120543586 | 0,998915527 | YES | 0 | FALSE |
| 306 | 54,040719 | 1,533787011 | 2,095522661 | 0,731935302 | 0,464208058 | 0,998915527 | YES | 0 | FALSE |
| 307 | 70,36519532 | 0,744865154 | 2,744045233 | 0,271447841 | 0,786046611 | 0,998915527 | YES | 0 | FALSE |
| 308 | 4,983906894 | 0,087098254 | 3,953985403 | 0,022027965 | 0,982425648 | 0,998915527 | YES | 0 | FALSE |
| 309 | 185,3406644 | 0,509215271 | 1,568920022 | 0,324564199 | 0,745510926 | 0,998915527 | YES | 0 | FALSE |
| 310 | 69,23958923 | -3,132145097 | 2,03083401 | -1,542294979 | 0,123001928 | 0,998915527 | YES | 0 | FALSE |
| 311 | 70,26092568 | 1,037446805 | 2,803615231 | 0,370038939 | 0,711353477 | 0,998915527 | YES | 0 | FALSE |
| 312 | 1,238547867 | 0,335866846 | 3,969134697 | 0,084619664 | 0,932563765 | 0,998915527 | YES | 0 | FALSE |
| 313 | 789,0232489 | 0,328455223 | 1,540886042 | 0,21315997 | 0,83120219 | 0,998915527 | YES | 0 | FALSE |
| 314 | 10,2251921 | -0,084175897 | 3,753243791 | -0,022427506 | 0,982106939 | 0,998915527 | YES | 0 | FALSE |
| 315 | 49,07126391 | -0,658927075 | 2,877050964 | -0,229028642 | 8,19E-01 | 0,998915527 | YES | 0 | FALSE |
| 316 | 18,06683066 | -1,719803058 | 2,319752883 | -0,741373389 | 0,458467075 | 0,998915527 | YES | 0 | FALSE |
| 317 | 7,152143617 | -3,978910852 | 3,647472119 | -1,090868065 | 0,275330941 | 0,998915527 | YES | 0 | FALSE |
| 318 | 167,7390849 | -0,253375235 | 1,620266721 | -0,156378719 | 0,87573451 | 0,998915527 | YES | 0 | FALSE |
| 319 | 5783,119515 | -0,687613865 | 1,542657481 | -0,445733336 | 0,655789882 | 0,998915527 | YES | 0 | FALSE |
| 320 | 153,4665116 | -0,013032925 | 2,665326821 | -0,004889804 | 9,96E-01 | 9,99E-01 | YES | 0 | FALSE |
| 321 | 145,1226248 | -0,55471427 | 1,602792949 | -0,346092283 | 0,729273363 | 0,998915527 | YES | 0 | FALSE |
| 322 | 16,85277489 | 2,109239936 | 2,256945205 | 0,934555226 | 0,350017563 | 0,998915527 | YES | 0 | FALSE |
| 323 | 100,7158558 | -1,172930518 | 2,430948805 | -0,482499062 | 0,629451465 | 0,998915527 | YES | 0 | FALSE |
| 324 | 137,8624935 | -0,00284528 | 2,093371426 | -0,001359185 | 0,998915527 | 0,998915527 | YES | 0 | FALSE |
| 325 | 33,30020942 | 2,057293672 | 2,720656679 | 0,756175407 | 0,449544036 | 0,998915527 | YES | 0 | FALSE |
| 326 | 35,57741409 | -2,658990322 | 2,67472191 | -0,994118421 | 0,320165218 | 0,998915527 | YES | 0 | FALSE |
| 327 | 42,70432203 | 1,376686652 | 2,853612875 | 0,482436375 | 0,629495986 | 0,998915527 | YES | 0 | FALSE |
| 328 | 994,4670852 | -0,758953073 | 1,748399022 | -0,43408459 | 0,664227015 | 0,998915527 | YES | 0 | FALSE |
| 329 | 429,5712851 | -0,311615215 | 2,632391542 | -0,118377228 | 0,905768768 | 0,998915527 | YES | 0 | FALSE |

| Medication | Ticker | taxLevel | Phylum | Family | Genus | Species |
|---|---|---|---|---|---|---|
| Ropinirole | 330 | Genus | Firmicutes | Lachnospiraceae | Tyzzerella | NA |
| Ropinirole | 331 | Genus | Euryarchaeota | Methanobacteriaceae | Methanosphaera | NA |
| Ropinirole | 332 | Genus | Firmicutes | Erysipelotrichaceae | Solobacterium | NA |
| Ropinirole | 333 | Genus | Firmicutes | Lachnospiraceae | Lachnospiraceae AC2044 group | NA |
| Ropinirole | 334 | Genus | Euryarchaeota | Methanobacteriaceae | Methanobrevibacter | NA |
| Ropinirole | 335 | Genus | Firmicutes | Ruminococcaceae | CAG-352 | NA |
| Ropinirole | 336 | Genus | Firmicutes | Clostridiaceae | Clostridium sensu stricto 1 | NA |
| Ropinirole | 337 | Genus | Actinobacteriota | Eggerthellaceae | Slackia | NA |
| Ropinirole | 338 | Genus | Firmicutes | Oscillospiraceae | UCG-002 | NA |
| Ropinirole | 339 | Genus | Firmicutes | Ruminococcaceae | Negativibacillus | NA |
| Ropinirole | 340 | Genus | Firmicutes | Ruminococcaceae | Subdoligranulum | NA |
| Ropinirole | 341 | Genus | Proteobacteria | Enterobacteriaceae | Escherichia-Shigella | NA |
| Ropinirole | 342 | Genus | Firmicutes | Lachnospiraceae | GCA-900066575 | NA |
| Ropinirole | 343 | Genus | Firmicutes | Veillonellaceae | Veillonella | NA |
| Ropinirole | 344 | Genus | Firmicutes | Oscillospiraceae | Colidextribacter | NA |
| Ropinirole | 345 | Genus | Bacteroidota | Rikenellaceae | Alistipes | NA |
| Ropinirole | 346 | Genus | Firmicutes | Anaerovoracaceae | Family XIII UCG-001 | NA |
| Ropinirole | 347 | Genus | Firmicutes | Anaerovoracaceae | Family XIII AD3011 group | NA |
| Ropinirole | 348 | Genus | Firmicutes | Anaerovoracaceae | [Eubacterium] brachy group | NA |
| Ropinirole | 349 | Genus | Firmicutes | Peptostreptococcaceae | Intestinibacter | NA |
| Ropinirole | 350 | Genus | Firmicutes | Lachnospiraceae | Lachnotalea | NA |
| Ropinirole | 351 | Genus | Firmicutes | Lachnospiraceae | Anaerostipes | NA |
| Ropinirole | 352 | Genus | Firmicutes | Lachnospiraceae | Marvinbryantia | NA |
| Ropinirole | 353 | Genus | Firmicutes | Lachnospiraceae | [Eubacterium] hallii group | NA |
| Ropinirole | 354 | Genus | Bacteroidota | Barnesiellaceae | Coprobacter | NA |
| Ropinirole | 355 | Genus | Firmicutes | Ruminococcaceae | DTU089 | NA |
| Ropinirole | 356 | Genus | Bacteroidota | Tannerellaceae | Parabacteroides | NA |
| Ropinirole | 357 | Genus | Firmicutes | Ruminococcaceae | Ruminococcus | NA |
| Ropinirole | 358 | Genus | Firmicutes | Erysipelotrichaceae | Holdemanella | NA |
| Ropinirole | 359 | Genus | Bacteroidota | Prevotellaceae | Paraprevotella | NA |

| Ticker | DESeq2_baseMean | DESeq2_log2-FoldChange | DESeq2_lfcSE | DESeq2_stat | DESeq2_pvalue | DESeq2_padj | DESeq2 converged | ANCOM_W | ANCOM_detected_0.7 |
|---|---|---|---|---|---|---|---|---|---|
| 330 | 19,77158324 | 1,655667402 | 3,119150804 | 0,530807103 | 0,595552457 | 0,998915527 | YES | 0 | FALSE |
| 331 | 3,205901334 | 1,010981555 | 3,959694357 | 0,255318079 | 0,798477438 | 0,998915527 | YES | 0 | FALSE |
| 332 | 17,50218372 | -0,578299233 | 3,227764182 | -0,179164028 | 0,857808908 | 0,998915527 | YES | 0 | FALSE |
| 333 | 13,77179862 | 2,735901278 | 3,919929027 | 0,697946636 | 0,485210568 | 0,998915527 | YES | 0 | FALSE |
| 334 | 98,19313528 | -2,145138839 | 2,036322229 | -1,053437815 | 0,292140381 | 0,998915527 | YES | 0 | FALSE |
| 335 | 15,81206508 | 2,758532069 | 3,831464946 | 0,719967978 | 0,471544712 | 0,998915527 | YES | 0 | FALSE |
| 336 | 77,0983509 | -0,780453119 | 2,083606126 | -0,374568451 | 0,70798144 | 0,998915527 | YES | 0 | FALSE |
| 337 | 1,455174728 | -1,719547356 | 4,024826115 | -0,427235191 | 0,669208032 | 0,998915527 | YES | 0 | FALSE |
| 338 | 2408,052753 | 0,210728901 | 1,538324701 | 0,13698597 | 0,891041885 | 0,998915527 | YES | 0 | FALSE |
| 339 | 63,70001016 | 5,524419427 | 2,343275925 | 2,357562491 | 0,018395359 | 0,674496492 | YES | 0 | FALSE |
| 340 | 623,727588 | 1,26308849 | 1,578115413 | 0,800377767 | 0,423491958 | 0,998915527 | YES | 0 | FALSE |
| 341 | 95,82908542 | -3,178590307 | 2,492250046 | -1,275389808 | 0,202171304 | 0,998915527 | YES | 0 | FALSE |
| 342 | 16,59031394 | -0,056966043 | 2,679520481 | -0,02125979 | 0,983038419 | 0,998915527 | YES | 0 | FALSE |
| 343 | 130,0522936 | 1,022407812 | 2,72596113 | 0,375063239 | 0,707613436 | 0,998915527 | YES | 0 | FALSE |
| 344 | 62,92704697 | 1,953416454 | 1,685214584 | 1,159149982 | 0,246395056 | 0,998915527 | YES | 0 | FALSE |
| 345 | 940,0463924 | 0,011574484 | 1,566758272 | 0,007387537 | 0,994105652 | 0,998915527 | YES | 0 | FALSE |
| 346 | 17,77146951 | -0,646960272 | 3,168781201 | -0,204166912 | 0,838223067 | 0,998915527 | YES | 0 | FALSE |
| 347 | 38,56080584 | 1,618085511 | 2,08768576 | 0,775061814 | 0,438303135 | 0,998915527 | YES | 0 | FALSE |
| 348 | 2,087173488 | -0,286057268 | 3,893840288 | -0,073464047 | 0,941436853 | 0,998915527 | YES | 0 | FALSE |
| 349 | 40,5772326 | 0,163845505 | 2,732014276 | 0,059972419 | 0,952177602 | 0,998915527 | YES | 0 | FALSE |
| 350 | 24,05981237 | -0,027221206 | 3,03362859 | -0,00897315 | 0,992840558 | 0,998915527 | YES | 0 | FALSE |
| 351 | 36,53435864 | 2,177027459 | 2,069627655 | 1,051893298 | 0,292848507 | 0,998915527 | YES | 0 | FALSE |
| 352 | 22,76436955 | 7,665368083 | 3,095814955 | 2,476042075 | 0,013284792 | 0,674496492 | YES | 0 | FALSE |
| 353 | 61,15688542 | 0,161874032 | 1,63419075 | 0,099054551 | 0,921094957 | 0,998915527 | YES | 0 | FALSE |
| 354 | 14,48752516 | 0,110967829 | 3,283095654 | 0,033799755 | 0,973036831 | 0,998915527 | YES | 0 | FALSE |
| 355 | 19,23929244 | 0,70369753 | 2,424471343 | 0,290247823 | 0,771626652 | 0,998915527 | YES | 0 | FALSE |
| 356 | 398,1486777 | -0,065284458 | 1,583706236 | -0,041222581 | 0,967118452 | 0,998915527 | YES | 0 | FALSE |
| 357 | 1513,493292 | 1,37383825 | 1,670682938 | 0,822321351 | 0,410894028 | 0,998915527 | YES | 0 | FALSE |
| 358 | 225,0410883 | 0,973856074 | 2,633453264 | 0,36980192 | 0,711530084 | 0,998915527 | YES | 0 | FALSE |
| 359 | 65,6993896 | -0,293366686 | 2,18333516 | -0,134366309 | 0,893112922 | 0,998915527 | YES | 0 | FALSE |

| Medication | Ticker | taxLevel | Phylum | Family | Genus | Species |
|---|---|---|---|---|---|---|
| Ropinirole | 360 | Genus | Firmicutes | Lactobacillaceae | Lactobacillus | NA |
| Ropinirole | 361 | Genus | Firmicutes | Oscillospiraceae | Oscillospira | NA |
| Ropinirole | 362 | Genus | Actinobacteriota | Atopobiaceae | Olsenella | NA |
| Ropinirole | 363 | Genus | Verrucomicrobiota | Victivallaceae | Victivallis | NA |
| Ropinirole | 364 | Genus | Actinobacteriota | Actinomycetaceae | Actinomyces | NA |
| Ropinirole | 365 | Genus | Firmicutes | Lachnospiraceae | Howardella | NA |
| Ropinirole | 366 | Genus | Firmicutes | Lachnospiraceae | Eisenbergiella | NA |
| Ropinirole | 367 | Genus | Firmicutes | Lachnospiraceae | Blautia | NA |
| Ropinirole | 368 | Genus | Firmicutes | Oscillospiraceae | UCG-005 | NA |
| Ropinirole | 369 | Genus | Proteobacteria | Oxalobacteraceae | Oxalobacter | NA |
| Ropinirole | 370 | Genus | Firmicutes | Lachnospiraceae | [Ruminococcus] torques group | NA |
| Ropinirole | 371 | Genus | Firmicutes | Veillonellaceae | Allisonella | NA |
| Ropinirole | 372 | Genus | Bacteroidota | Marinifilaceae | Butyricimonas | NA |
| Ropinirole | 373 | Genus | Firmicutes | Monoglobaceae | Monoglobus | NA |
| Ropinirole | 374 | Genus | Bacteroidota | Rikenellaceae | Rikenellaceae RC9 gut group | NA |
| Ropinirole | 375 | Genus | Firmicutes | Lachnospiraceae | Roseburia | NA |
| Ropinirole | 376 | Genus | Firmicutes | Erysipelatoclostridiaceae | Erysipelatoclostridium | NA |
| Ropinirole | 377 | Genus | Firmicutes | Ruminococcaceae | [Eubacterium] siraeum group | NA |
| Ropinirole | 378 | Genus | Firmicutes | Erysipelatoclostridiaceae | Catenibacterium | NA |
| Ropinirole | 379 | Genus | Firmicutes | Ruminococcaceae | Angelakisella | NA |
| Ropinirole | 380 | Genus | Firmicutes | Ruminococcaceae | Incertae Sedis | NA |
| Ropinirole | 381 | Genus | Firmicutes | Butyricicoccaceae | Butyricicoccus | NA |
| Ropinirole | 382 | Genus | Firmicutes | Erysipelotrichaceae | Holdemania | NA |
| Ropinirole | 383 | Genus | Firmicutes | Ruminococcaceae | Fournierella | NA |
| Ropinirole | 384 | Genus | Firmicutes | Lachnospiraceae | Agathobacter | NA |
| Ropinirole | 385 | Genus | Proteobacteria | Sutterellaceae | Parasutterella | NA |
| Ropinirole | 386 | Genus | Firmicutes | Lachnospiraceae | [Ruminococcus] gauvreauii group | NA |
| Ropinirole | 387 | Genus | Firmicutes | Lachnospiraceae | Lachnospiraceae ND3007 group | NA |
| Ropinirole | 388 | Genus | Firmicutes | Lachnospiraceae | [Eubacterium] eligens group | NA |
| Ropinirole | 389 | Genus | Firmicutes | Ruminococcaceae | Faecalibacterium | NA |

| Ticker | DESeq2_baseMean | DESeq2_log2-FoldChange | DESeq2_lfcSE | DESeq2_stat | DESeq2_pvalue | DESeq2_padj | DESeq2 converged | ANCOM_W | ANCOM_detected_0.7 |
|---|---|---|---|---|---|---|---|---|---|
| 360 | 208,3182409 | 1,203653798 | 1,732481347 | 0,694757147 | 0,487207508 | 0,998915527 | YES | 0 | FALSE |
| 361 | 13,572396 | 0,093375293 | 3,32365976 | 0,028094119 | 0,977587084 | 0,998915527 | YES | 0 | FALSE |
| 362 | 3,760878694 | 1,669012447 | 3,912634862 | 0,426569947 | 0,669692591 | 0,998915527 | YES | 0 | FALSE |
| 363 | 2,113081886 | 1,837956491 | 3,909714825 | 0,470099885 | 0,638283656 | 0,998915527 | YES | 0 | FALSE |
| 364 | 1,802291109 | 0,269515165 | 3,95939707 | 0,068069749 | 0,945730111 | 0,998915527 | YES | 0 | FALSE |
| 365 | 11,54581472 | 0,51872356 | 2,642010926 | 0,196336645 | 0,844346681 | 0,998915527 | YES | 0 | FALSE |
| 366 | 9,102273821 | -0,964024027 | 3,941077891 | -0,24460923 | 0,806759017 | 0,998915527 | YES | 0 | FALSE |
| 367 | 861,989844 | -0,665632078 | 1,630832038 | -0,408154894 | 0,683159961 | 0,998915527 | YES | 0 | FALSE |
| 368 | 614,1585699 | 1,164319243 | 1,55416677 | 0,749159785 | 0,453760905 | 0,998915527 | YES | 0 | FALSE |
| 369 | 3,869289193 | 2,59699978 | 3,91434086 | 0,66345775 | 0,50703743 | 0,998915527 | YES | 0 | FALSE |
| 370 | 276,9588331 | -1,825276468 | 1,920229081 | -0,950551414 | 0,341832142 | 0,998915527 | YES | 0 | FALSE |
| 371 | 12,69300113 | 1,103314502 | 2,738092186 | 0,402950093 | 0,686984939 | 0,998915527 | YES | 0 | FALSE |
| 372 | 36,72177525 | -0,760932167 | 1,748977459 | -0,435072598 | 0,663509732 | 0,998915527 | YES | 0 | FALSE |
| 373 | 231,8641466 | -0,296677817 | 1,557904079 | -0,190433944 | 0,848969101 | 0,998915527 | YES | 0 | FALSE |
| 374 | 8,926982486 | 2,76659746 | 3,947672619 | 0,700817349 | 0,48341701 | 0,998915527 | YES | 0 | FALSE |
| 375 | 708,3171437 | 1,127057633 | 2,005187623 | 0,56207091 | 0,574067707 | 0,998915527 | YES | 0 | FALSE |
| 376 | 40,97346121 | -1,12124496 | 2,196059323 | -0,510571344 | 0,609651246 | 0,998915527 | YES | 0 | FALSE |
| 377 | 587,3382827 | 3,998092723 | 1,593817649 | 2,508500722 | 0,012124472 | 0,674496492 | YES | 0 | FALSE |
| 378 | 153,459711 | 0,169465146 | 2,663490267 | 0,063625217 | 0,949268652 | 0,998915527 | YES | 0 | FALSE |
| 379 | 5,450989306 | 2,734777258 | 3,95309843 | 0,691806012 | 0,489059161 | 0,998915527 | YES | 0 | FALSE |
| 380 | 132,880875 | -0,166764243 | 1,668180279 | -0,099967759 | 0,920369922 | 0,998915527 | YES | 0 | FALSE |
| 381 | 178,694455 | 0,573880796 | 1,560539487 | 0,36774513 | 0,713063286 | 0,998915527 | YES | 0 | FALSE |
| 382 | 1,058308893 | -1,537185596 | 4,026198783 | -0,381795753 | 0,702612869 | 0,998915527 | YES | 0 | FALSE |
| 383 | 2,073670166 | -0,302862954 | 3,911688966 | -0,077425112 | 0,938285365 | 0,998915527 | YES | 0 | FALSE |
| 384 | 1254,197603 | -0,625833539 | 1,558494935 | -0,401562767 | 0,688005836 | 0,998915527 | YES | 0 | FALSE |
| 385 | 0,710998605 | 2,400344032 | 4,019341669 | 0,597198305 | 0,550374991 | 0,998915527 | YES | 0 | FALSE |
| 386 | 62,18513984 | 1,869244696 | 2,698587414 | 0,692675244 | 0,488513378 | 0,998915527 | YES | 0 | FALSE |
| 387 | 38,49542381 | -1,721447027 | 2,03481885 | -0,845995223 | 0,397555409 | 0,998915527 | YES | 0 | FALSE |
| 388 | 61,52353179 | -0,441752021 | 2,116078884 | -0,208759713 | 0,834635822 | 0,998915527 | YES | 0 | FALSE |
| 389 | 1830,650072 | -0,321526259 | 1,545952372 | -0,207979408 | 0,835245046 | 0,998915527 | YES | 0 | FALSE |

| Medication | Ticker | taxLevel | Phylum | Family | Genus | Species |
|---|---|---|---|---|---|---|
| Ropinirole | 390 | Genus | Firmicutes | Lachnospiraceae | Lachnospiraceae NK4A136 group | NA |
| Ropinirole | 391 | Genus | Firmicutes | Lachnospiraceae | Coprococcus | NA |
| Ropinirole | 392 | Genus | Actinobacteriota | Atopobiaceae | Libanicoccus | NA |
| Ropinirole | 393 | Genus | Proteobacteria | Sutterellaceae | Sutterella | NA |
| Ropinirole | 394 | Genus | Firmicutes | Lachnospiraceae | Shuttleworthia | NA |
| Ropinirole | 395 | Genus | Firmicutes | Erysipelotrichaceae | Turicibacter | NA |
| Ropinirole | 396 | Genus | Firmicutes | Lachnospiraceae | CAG-56 | NA |
| Ropinirole | 397 | Genus | Firmicutes | Lachnospiraceae | [Ruminococcus] gnavus group | NA |
| Ropinirole | 398 | Genus | Firmicutes | Butyricicoccaceae | UCG-009 | NA |
| Ropinirole | 399 | Genus | Firmicutes | Acidaminococcaceae | Phascolarctobacterium | NA |
| Ropinirole | 400 | Family | Actinobacteriota | Bifidobacteriaceae | NA | NA |
| Ropinirole | 401 | Family | Verrucomicrobiota | Akkermansiaceae | NA | NA |
| Ropinirole | 402 | Family | Verrucomicrobiota | vadinBE97 | NA | NA |
| Ropinirole | 403 | Family | Firmicutes | Veillonellaceae | NA | NA |
| Ropinirole | 404 | Family | Bacteroidota | Muribaculaceae | NA | NA |
| Ropinirole | 405 | Family | Firmicutes | Christensenellaceae | NA | NA |
| Ropinirole | 406 | Family | Firmicutes | Peptostreptococcaceae | NA | NA |
| Ropinirole | 407 | Family | Desulfobacterota | Desulfovibrionaceae | NA | NA |
| Ropinirole | 408 | Family | Actinobacteriota | Coriobacteriaceae | NA | NA |
| Ropinirole | 409 | Family | Proteobacteria | Pasteurellaceae | NA | NA |
| Ropinirole | 410 | Family | Firmicutes | Erysipelatoclostridiaceae | NA | NA |
| Ropinirole | 411 | Family | Bacteroidota | Marinifilaceae | NA | NA |
| Ropinirole | 412 | Family | Bacteroidota | Barnesiellaceae | NA | NA |
| Ropinirole | 413 | Family | Bacteroidota | Bacteroidaceae | NA | NA |
| Ropinirole | 414 | Family | Bacteroidota | Prevotellaceae | NA | NA |
| Ropinirole | 415 | Family | Firmicutes | Peptococcaceae | NA | NA |
| Ropinirole | 416 | Family | Firmicutes | Streptococcaceae | NA | NA |
| Ropinirole | 417 | Family | Actinobacteriota | Coriobacteriales Incertae Sedis | NA | NA |
| Ropinirole | 418 | Family | Euryarchaeota | Methanobacteriaceae | NA | NA |
| Ropinirole | 419 | Family | Firmicutes | Clostridiaceae | NA | NA |

| Ticker | DESeq2_baseMean | DESeq2_log2-FoldChange | DESeq2_lfcSE | DESeq2_stat | DESeq2_pvalue | DESeq2_padj | DESeq2 converged | ANCOM_W | ANCOM_detected_0.7 |
|---|---|---|---|---|---|---|---|---|---|
| 390 | 1188,883235 | 0,485306304 | 1,523781978 | 0,318488019 | 0,750114781 | 0,998915527 | YES | 0 | FALSE |
| 391 | 297,4399249 | 2,826501483 | 1,890951852 | 1,494750636 | 0,134979534 | 0,998915527 | YES | 0 | FALSE |
| 392 | 13,20882232 | -0,297342905 | 3,370083415 | -0,088230132 | 0,929693769 | 0,998915527 | YES | 0 | FALSE |
| 393 | 62,12576903 | 1,665081819 | 1,886186807 | 0,882776729 | 0,377356916 | 0,998915527 | YES | 0 | FALSE |
| 394 | 34,44927347 | 0,909148618 | 2,231284681 | 0,407455233 | 0,683673668 | 0,998915527 | YES | 0 | FALSE |
| 395 | 16,63640348 | 1,26433671 | 2,452895685 | 0,515446587 | 0,606240985 | 0,998915527 | YES | 0 | FALSE |
| 396 | 28,8579116 | -5,672306604 | 3,023655721 | -1,875976344 | 0,060658528 | 0,998915527 | YES | 0 | FALSE |
| 397 | 298,3901702 | 0,669334803 | 2,645887835 | 0,252971723 | 0,800290065 | 0,998915527 | YES | 0 | FALSE |
| 398 | 16,13403659 | 2,825808336 | 2,934847972 | 0,962846581 | 0,335624521 | 0,998915527 | YES | 0 | FALSE |
| 399 | 73,55906196 | 0,867602771 | 2,108476756 | 0,411483204 | 0,680718255 | 0,998915527 | YES | 0 | FALSE |
| 400 | 626,50076 | -0,423855252 | 1,006951253 | -0,420929266 | 0,673806734 | 0,992508719 | YES | 0 | FALSE |
| 401 | 178,1127135 | 1,603981515 | 1,301310726 | 1,232589176 | 0,217729077 | 0,992508719 | YES | 0 | FALSE |
| 402 | 0,444887609 | 2,44999426 | 4,028164558 | 0,608216031 | 0,543044204 | 0,992508719 | YES | 0 | FALSE |
| 403 | 1398,278448 | 2,774351619 | 0,890444467 | 3,115693031 | 0,001835132 | 0,078910675 | YES | 0 | FALSE |
| 404 | 1021,270773 | 2,909031224 | 1,417276359 | 2,052550447 | 0,040116198 | 0,86249825 | YES | 0 | FALSE |
| 405 | 556,4461087 | -0,121316526 | 1,231057528 | -0,098546593 | 0,921498276 | 0,992508719 | YES | 0 | FALSE |
| 406 | 114,2054329 | 0,987013143 | 1,62290414 | 0,608177106 | 0,543070017 | 0,992508719 | YES | 0 | FALSE |
| 407 | 87,17576814 | -1,494516775 | 1,406353214 | -1,062689487 | 0,28792279 | 0,992508719 | YES | 0 | FALSE |
| 408 | 62,97320948 | -2,864476972 | 1,661841884 | -1,723676001 | 0,08476635 | 0,911238261 | YES | 0 | FALSE |
| 409 | 77,66308689 | 1,529233623 | 2,190136782 | 0,698236583 | 0,485029252 | 0,992508719 | YES | 0 | FALSE |
| 410 | 179,2542659 | 0,295085501 | 1,142932994 | 0,258182678 | 0,796265926 | 0,992508719 | YES | 0 | FALSE |
| 411 | 51,53716572 | -0,880525901 | 1,278590071 | -0,688669434 | 0,491031315 | 0,992508719 | YES | 0 | FALSE |
| 412 | 274,8745053 | 0,084028278 | 1,021202767 | 0,082283637 | 0,934421166 | 0,992508719 | YES | 0 | FALSE |
| 413 | 6003,305767 | -0,522559378 | 0,738072302 | -0,708005675 | 0,478941733 | 0,992508719 | YES | 0 | FALSE |
| 414 | 922,5244736 | 0,138522111 | 1,023429361 | 0,135350925 | 0,892334423 | 0,992508719 | YES | 0 | FALSE |
| 415 | 123,4033056 | 0,267037061 | 1,144520122 | 0,233317926 | 0,81551454 | 0,992508719 | YES | 0 | FALSE |
| 416 | 1041,929649 | -0,548497518 | 1,14833611 | -0,47764545 | 0,632902576 | 0,992508719 | YES | 0 | FALSE |
| 417 | 36,4048039 | -0,488277792 | 2,027663912 | -0,24080805 | 0,80970389 | 0,992508719 | YES | 0 | FALSE |
| 418 | 86,15911518 | -1,93757654 | 1,601851096 | -1,209585925 | 0,226437822 | 0,992508719 | YES | 0 | FALSE |
| 419 | 79,13344444 | -0,704484828 | 1,468086309 | -0,479866084 | 0,631322619 | 0,992508719 | YES | 0 | FALSE |

| Medication | Ticker | taxLevel | Phylum | Family | Genus | Species |
|---|---|---|---|---|---|---|
| Ropinirole | 420 | Family | Firmicutes | Oscillospiraceae | NA | NA |
| Ropinirole | 421 | Family | Proteobacteria | Enterobacteriaceae | NA | NA |
| Ropinirole | 422 | Family | Bacteroidota | Rikenellaceae | NA | NA |
| Ropinirole | 423 | Family | Firmicutes | Anaerovoracaceae | NA | NA |
| Ropinirole | 424 | Family | Bacteroidota | Tannerellaceae | NA | NA |
| Ropinirole | 425 | Family | Firmicutes | Ruminococcaceae | NA | NA |
| Ropinirole | 426 | Family | Firmicutes | Erysipelotrichaceae | NA | NA |
| Ropinirole | 427 | Family | Firmicutes | Lactobacillaceae | NA | NA |
| Ropinirole | 428 | Family | Firmicutes | UCG-010 | NA | NA |
| Ropinirole | 429 | Family | Actinobacteriota | Atopobiaceae | NA | NA |
| Ropinirole | 430 | Family | Verrucomicrobiota | Victivallaceae | NA | NA |
| Ropinirole | 431 | Family | Actinobacteriota | Actinomycetaceae | NA | NA |
| Ropinirole | 432 | Family | Proteobacteria | Oxalobacteraceae | NA | NA |
| Ropinirole | 433 | Family | Firmicutes | Monoglobaceae | NA | NA |
| Ropinirole | 434 | Family | Firmicutes | [Eubacterium] coprostanoligenes group | NA | NA |
| Ropinirole | 435 | Family | Actinobacteriota | Eggerthellaceae | NA | NA |
| Ropinirole | 436 | Family | Firmicutes | Butyricicoccaceae | NA | NA |
| Ropinirole | 437 | Family | Firmicutes | Lachnospiraceae | NA | NA |
| Ropinirole | 438 | Family | Firmicutes | Hydrogenoanaerobacterium | NA | NA |
| Ropinirole | 439 | Family | Proteobacteria | Sutterellaceae | NA | NA |
| Ropinirole | 440 | Family | Verrucomicrobiota | Puniceicoccaceae | NA | NA |
| Ropinirole | 441 | Family | Firmicutes | Ruminiclostridium | NA | NA |
| Ropinirole | 442 | Family | Firmicutes | Acidaminococcaceae | NA | NA |

| Ticker | DESeq2_baseMean | DESeq2_log2-FoldChange | DESeq2_lfcSE | DESeq2_stat | DESeq2_pvalue | DESeq2_padj | DESeq2 converged | ANCOM_W | ANCOM_detected_0.7 |
|---|---|---|---|---|---|---|---|---|---|
| 420 | 4447,37526 | 0,531400308 | 0,67082108 | 0,792163996 | 0,428265059 | 0,992508719 | YES | 0 | FALSE |
| 421 | 103,8794616 | -3,258583358 | 1,865030222 | -1,74720137 | 0,080602414 | 0,911238261 | YES | 0 | FALSE |
| 422 | 978,8666317 | 0,210450068 | 0,835358857 | 0,251927739 | 0,801096919 | 0,992508719 | YES | 0 | FALSE |
| 423 | 67,10174575 | 0,295466365 | 1,1625417 | 0,254155498 | 0,799375429 | 0,992508719 | YES | 0 | FALSE |
| 424 | 421,0129823 | 0,112807397 | 0,848073056 | 0,133016131 | 0,894180623 | 0,992508719 | YES | 0 | FALSE |
| 425 | 5266,685092 | 0,371492781 | 0,706830667 | 0,525575358 | 0,599183282 | 0,992508719 | YES | 0 | FALSE |
| 426 | 230,3135919 | -0,073891375 | 1,114939874 | -0,066273865 | 0,94715979 | 0,992508719 | YES | 0 | FALSE |
| 427 | 233,2572654 | 1,431542301 | 1,171461452 | 1,222014005 | 0,221702331 | 0,992508719 | YES | 0 | FALSE |
| 428 | 135,7797637 | 0,845237472 | 1,040205259 | 0,812567966 | 0,416465804 | 0,992508719 | YES | 0 | FALSE |
| 429 | 13,32550939 | 1,879861605 | 3,369314973 | 0,557935848 | 0,576888192 | 0,992508719 | YES | 0 | FALSE |
| 430 | 2,509140312 | 0,016896384 | 3,859825142 | 0,0043775 | 0,996507272 | 0,996507272 | YES | 0 | FALSE |
| 431 | 1,402128322 | 0,431175739 | 3,959386034 | 0,108899646 | 0,913282087 | 0,992508719 | YES | 0 | FALSE |
| 432 | 3,344068027 | 2,77914123 | 3,915101255 | 0,709851687 | 0,477796113 | 0,992508719 | YES | 0 | FALSE |
| 433 | 246,1453133 | -0,137628337 | 0,876032813 | -0,157104089 | 0,875162815 | 0,992508719 | YES | 0 | FALSE |
| 434 | 242,0207145 | -0,250592886 | 0,896955421 | -0,27938165 | 0,779951952 | 0,992508719 | YES | 0 | FALSE |
| 435 | 89,52129243 | -0,637517303 | 1,143301178 | -0,557610991 | 0,577110049 | 0,992508719 | YES | 0 | FALSE |
| 436 | 211,8057314 | 0,768302913 | 0,852185392 | 0,901567805 | 0,3672865 | 0,992508719 | YES | 0 | FALSE |
| 437 | 8883,985597 | 0,02710928 | 0,707319102 | 0,038326803 | 0,969427121 | 0,992508719 | YES | 0 | FALSE |
| 438 | 3,674039732 | -0,631638384 | 3,951743764 | -0,15983789 | 0,873008776 | 0,992508719 | YES | 0 | FALSE |
| 439 | 56,93635731 | 1,75286069 | 1,5426645 | 1,136255284 | 0,255849739 | 0,992508719 | YES | 0 | FALSE |
| 440 | 2,542214907 | 2,773390695 | 3,971629418 | 0,698300471 | 0,484989305 | 0,992508719 | YES | 0 | FALSE |
| 441 | 12,9845155 | 0,263555766 | 3,840931733 | 0,068617665 | 0,945293958 | 0,992508719 | YES | 0 | FALSE |
| 442 | 72,1926898 | 1,080686728 | 1,542694522 | 0,700518938 | 0,483603283 | 0,992508719 | YES | 0 | FALSE |

**Supplementary Table 8: dose-effect relationships at ASV, genus and family level of dopaminergic medication (a. ledd, b. levodopa, c. dopamine agonists, d. pramipexol, e. ropinirole) with DESeq2 and ANCOM**
**All columns are spread out over two pages, after which the next two pages show the next set of rows, etc.**
**A ticker is provided on both pages to easily connect the taxonomy data to the associated statistics.**

**Supplementary Table 8a: LEDD**

| Medication | Ticker | ASVnr | taxLevel | Phylum | Family | Genus | Species |
|---|---|---|---|---|---|---|---|
| LEDD | 1 | ASV76 | ASV | Firmicutes | Lachnospiraceae | Lachnospiraceae ND3007 group | NA |
| LEDD | 2 | ASV74 | ASV | Firmicutes | Lachnospiraceae | Coprococcus | catus |
| LEDD | 3 | ASV53 | ASV | Firmicutes | Lachnospiraceae | [Ruminococcus] torques group | NA |
| LEDD | 4 | ASV309 | ASV | Firmicutes | Oscillospiraceae | Colidextribacter | massiliensis |
| LEDD | 5 | ASV45 | ASV | Firmicutes | Lachnospiraceae | Coprococcus | comes |
| LEDD | 6 | ASV11 | ASV | Firmicutes | Ruminococcaceae | Subdoligranulum | NA |
| LEDD | 7 | ASV36 | ASV | Firmicutes | Lachnospiraceae | Blautia | obeum |
| LEDD | 8 | ASV157 | ASV | Firmicutes | Oscillospiraceae | UCG-005 | NA |
| LEDD | 9 | ASV30 | ASV | Proteobacteria | Enterobacteriaceae | Escherichia-Shigella | NA |
| LEDD | 10 | ASV51 | ASV | Firmicutes | Oscillospiraceae | UCG-005 | NA |
| LEDD | 11 | ASV55 | ASV | Firmicutes | Monoglobaceae | Monoglobus | NA |
| LEDD | 12 | ASV37 | ASV | Firmicutes | Lachnospiraceae | [Eubacterium] hallii group | NA |
| LEDD | 13 | ASV21 | ASV | Firmicutes | Ruminococcaceae | Faecalibacterium | prausnitzii |
| LEDD | 14 | ASV65 | ASV | Firmicutes | Ruminococcaceae | Incertae Sedis | NA |
| LEDD | 15 | ASV131 | ASV | Firmicutes | Lachnospiraceae | NA | NA |
| LEDD | 16 | ASV211 | ASV | Firmicutes | Lachnospiraceae | Lachnospiraceae FCS020 group | NA |
| LEDD | 17 | ASV19 | ASV | Firmicutes | Erysipelatoclostridiaceae | Erysipelotrichaceae UCG-003 | bacterium |
| LEDD | 18 | ASV96 | ASV | Firmicutes | Lachnospiraceae | CAG-56 | NA |
| LEDD | 19 | ASV88 | ASV | Firmicutes | Lachnospiraceae | Dorea | formicigenerans |
| LEDD | 20 | ASV111 | ASV | Firmicutes | Lachnospiraceae | Lachnospira | pectinoschiza |
| LEDD | 21 | ASV14 | ASV | Firmicutes | Lachnospiraceae | Blautia | massiliensis |
| LEDD | 22 | ASV144 | ASV | Desulfobacterota | Desulfovibrionaceae | Bilophila | wadsworthia |
| LEDD | 23 | ASV32 | ASV | Firmicutes | Oscillospiraceae | UCG-002 | NA |

| Ticker | DESeq2_baseMean | DESeq2_log2-FoldChange | DESeq2_lfcSE | DESeq2_stat | DESeq2_pvalue | DESeq2_padj | DESeq2 converged |
|---|---|---|---|---|---|---|---|
| 1 | 61,87979574 | -0,002711078 | 0,000767811 | -3,530916433 | 0,000414123 | 0,170204373 | YES |
| 2 | 82,13050064 | -0,001323145 | 0,000498954 | -2,651836209 | 0,008005537 | 0,999477707 | YES |
| 3 | 297,6477888 | -0,001361004 | 0,000736081 | -1,848986725 | 0,06445973 | 0,999477707 | YES |
| 4 | 26,08398385 | -0,002107162 | 0,001275452 | -1,6520909 | 0,098516022 | 0,999477707 | YES |
| 5 | 119,6318567 | -0,000981457 | 0,000627538 | -1,563981582 | 0,117821894 | 0,999477707 | YES |
| 6 | 392,023144 | -0,000737191 | 0,000498417 | -1,479065467 | 0,139122819 | 0,999477707 | YES |
| 7 | 214,0964186 | -0,001039448 | 0,000713463 | -1,456904549 | 0,14514273 | 0,999477707 | YES |
| 8 | 61,57773797 | -0,001334146 | 0,000923297 | -1,444981169 | 0,148463175 | 0,999477707 | YES |
| 9 | 646,9240633 | -0,001321354 | 0,001041389 | -1,268837253 | 0,204499115 | 0,999477707 | YES |
| 10 | 280,6266795 | -0,000682202 | 0,000575151 | -1,186126655 | 0,235572285 | 0,999477707 | YES |
| 11 | 79,94215441 | -0,00098147 | 0,000885382 | -1,108526496 | 0,267634503 | 0,999477707 | YES |
| 12 | 136,4336817 | -0,000561154 | 0,000543648 | -1,032200439 | 0,30197823 | 0,999477707 | YES |
| 13 | 329,5330742 | -0,002579203 | 0,002533587 | -1,018004517 | 0,308675809 | 0,999477707 | YES |
| 14 | 70,99439829 | 0,001619744 | 0,001658519 | 0,976620651 | 0,328756991 | 0,999477707 | YES |
| 15 | 219,9122333 | 0,000835081 | 0,000870263 | 0,959573367 | 0,337269979 | 0,999477707 | YES |
| 16 | 40,95303873 | -0,00109761 | 0,001170593 | -0,937652577 | 0,348422982 | 0,999477707 | YES |
| 17 | 171,7659937 | -0,000559494 | 0,000611835 | -0,914451646 | 0,360479575 | 0,999477707 | YES |
| 18 | 107,3283829 | -0,002165093 | 0,002524379 | -0,857673263 | 0,391072908 | 0,999477707 | YES |
| 19 | 61,62795417 | 0,001025164 | 0,001207258 | 0,849167683 | 0,395787993 | 0,999477707 | YES |
| 20 | 72,13617156 | -0,002187731 | 0,002604455 | -0,839995845 | 0,400910716 | 0,999477707 | YES |
| 21 | 411,9541167 | -0,00046656 | 0,000566499 | -0,823585204 | 0,410175286 | 0,999477707 | YES |
| 22 | 53,68000431 | 0,000593759 | 0,000734137 | 0,808784676 | 0,418639012 | 0,999477707 | YES |
| 23 | 616,792096 | -0,001785963 | 0,002214149 | -0,806613697 | 0,41988908 | 0,999477707 | YES |

| Medication | Ticker | ASVnr | taxLevel | Phylum | Family | Genus | Species |
|---|---|---|---|---|---|---|---|
| LEDD | 24 | ASV158 | ASV | Firmicutes | Lachnospiraceae | [Eubacterium] ventriosum group | NA |
| LEDD | 25 | ASV56 | ASV | Bacteroidota | Bacteroidaceae | Bacteroides | massiliensis |
| LEDD | 26 | ASV2 | ASV | Bacteroidota | Bacteroidaceae | Bacteroides | vulgatus |
| LEDD | 27 | ASV63 | ASV | Actinobacteriota | Eggerthellaceae | NA | NA |
| LEDD | 28 | ASV267 | ASV | Firmicutes | Lachnospiraceae | Lachnoclostridium | NA |
| LEDD | 29 | ASV7 | ASV | Firmicutes | Lachnospiraceae | Agathobacter | NA |
| LEDD | 30 | ASV89 | ASV | Firmicutes | Lachnospiraceae | Blautia | NA |
| LEDD | 31 | ASV31 | ASV | Bacteroidota | Rikenellaceae | Alistipes | putredinis |
| LEDD | 32 | ASV168 | ASV | Firmicutes | Lachnospiraceae | Lachnoclostridium | edouardi |
| LEDD | 33 | ASV24 | ASV | Bacteroidota | Bacteroidaceae | Bacteroides | uniformis |
| LEDD | 34 | ASV226 | ASV | Firmicutes | Lachnospiraceae | Lachnospiraceae UCG-001 | NA |
| LEDD | 35 | ASV116 | ASV | Firmicutes | Oscillospiraceae | UCG-003 | NA |
| LEDD | 36 | ASV41 | ASV | Firmicutes | Ruminococcaceae | Ruminococcus | bicirculans |
| LEDD | 37 | ASV6 | ASV | Firmicutes | Ruminococcaceae | Faecalibacterium | NA |
| LEDD | 38 | ASV219 | ASV | Firmicutes | Lachnospiraceae | Lachnospiraceae UCG-004 | NA |
| LEDD | 39 | ASV71 | ASV | Firmicutes | [Eubacterium] coprostanoligenes group | NA | NA |
| LEDD | 40 | ASV403 | ASV | Firmicutes | Oscillospiraceae | Colidextribacter | NA |
| LEDD | 41 | ASV221 | ASV | Firmicutes | Oscillospiraceae | Intestinimonas | NA |
| LEDD | 42 | ASV64 | ASV | Firmicutes | Oscillospiraceae | UCG-002 | NA |
| LEDD | 43 | ASV283 | ASV | Actinobacteriota | Eggerthellaceae | Slackia | isoflavoniconvertens |
| LEDD | 44 | ASV18 | ASV | Firmicutes | Lachnospiraceae | Dorea | longicatena |
| LEDD | 45 | ASV5 | ASV | Firmicutes | Lachnospiraceae | Blautia | NA |
| LEDD | 46 | ASV147 | ASV | Firmicutes | Oscillospiraceae | Oscillibacter | NA |
| LEDD | 47 | ASV512 | ASV | Proteobacteria | Oxalobacteraceae | Oxalobacter | formigenes |
| LEDD | 48 | ASV320 | ASV | Firmicutes | Christensenellaceae | Christensenellaceae R-7 group | NA |
| LEDD | 49 | ASV34 | ASV | Firmicutes | Lachnospiraceae | Anaerostipes | hadrus |
| LEDD | 50 | ASV393 | ASV | Bacteroidota | Rikenellaceae | Alistipes | indistinctus |
| LEDD | 51 | ASV626 | ASV | Firmicutes | NA | NA | NA |
| LEDD | 52 | ASV264 | ASV | Firmicutes | Oscillospiraceae | Colidextribacter | NA |
| LEDD | 53 | ASV17 | ASV | Actinobacteriota | Coriobacteriaceae | Collinsella | aerofaciens |

| Ticker | DESeq2_baseMean | DESeq2_log2-FoldChange | DESeq2_lfcSE | DESeq2_stat | DESeq2_pvalue | DESeq2_padj | DESeq2 converged |
|---|---|---|---|---|---|---|---|
| 24 | 60,2889301 | -0,000826825 | 0,001029039 | -0,80349224 | 0,42169028 | 0,999477707 | YES |
| 25 | 185,8878633 | -0,000952374 | 0,001257613 | -0,757287349 | 0,448877729 | 0,999477707 | YES |
| 26 | 1159,525526 | 0,000331007 | 0,000464654 | 0,712374316 | 0,476233016 | 0,999477707 | YES |
| 27 | 84,9391707 | -0,000400434 | 0,000563596 | -0,710497077 | 0,477395943 | 0,999477707 | YES |
| 28 | 21,37708722 | -0,001297533 | 0,001830203 | -0,708955587 | 0,478352038 | 0,999477707 | YES |
| 29 | 1686,904785 | -0,001650547 | 0,002420087 | -0,682019707 | 0,49522649 | 0,999477707 | YES |
| 30 | 64,70622657 | 0,001959986 | 0,002926216 | 0,669802419 | 0,502983751 | 0,999477707 | YES |
| 31 | 322,2845272 | -0,001435188 | 0,002215383 | -0,647828246 | 0,517096044 | 0,999477707 | YES |
| 32 | 58,01814875 | -0,000771748 | 0,001213598 | -0,635917429 | 0,524830242 | 0,999477707 | YES |
| 33 | 370,4736696 | -0,001726604 | 0,002814679 | -0,613428412 | 0,539593104 | 0,999477707 | YES |
| 34 | 49,5657396 | -0,001301298 | 0,002146623 | -0,606207023 | 0,54437729 | 0,999477707 | YES |
| 35 | 138,848875 | -0,001232015 | 0,002111747 | -0,583410197 | 0,559617195 | 0,999477707 | YES |
| 36 | 196,4702635 | -0,000849797 | 0,001460183 | -0,58198001 | 0,560580146 | 0,999477707 | YES |
| 37 | 819,0129138 | -0,001318808 | 0,002286877 | -0,576684967 | 0,564152289 | 0,999477707 | YES |
| 38 | 29,34100099 | -0,000979733 | 0,001709946 | -0,572961239 | 0,566670939 | 0,999477707 | YES |
| 39 | 55,27170415 | 0,001105296 | 0,001932968 | 0,571812765 | 0,567448828 | 0,999477707 | YES |
| 40 | 9,271850602 | -0,001474012 | 0,002613221 | -0,56405967 | 0,572713524 | 0,999477707 | YES |
| 41 | 34,09495391 | -0,000490434 | 0,000930438 | -0,52709979 | 0,598124293 | 0,999477707 | YES |
| 42 | 267,3251138 | -0,000982807 | 0,001929492 | -0,509360678 | 0,610499433 | 0,999477707 | YES |
| 43 | 9,822575183 | -0,001031641 | 0,002052731 | -0,50256985 | 0,615266731 | 0,999477707 | YES |
| 44 | 612,0135466 | -0,001071598 | 0,002147574 | -0,498980698 | 0,617792982 | 0,999477707 | YES |
| 45 | 492,9887856 | 0,000206389 | 0,00041984 | 0,491589524 | 0,623009551 | 0,999477707 | YES |
| 46 | 47,25557163 | 0,000526249 | 0,001073754 | 0,490101732 | 0,624061912 | 0,999477707 | YES |
| 47 | 12,74891559 | 0,000439131 | 0,000911474 | 0,481781577 | 0,629961118 | 0,999477707 | YES |
| 48 | 23,32683817 | 0,000718303 | 0,00152941 | 0,469660341 | 0,638597706 | 0,999477707 | YES |
| 49 | 135,9333701 | -0,000940165 | 0,002009155 | -0,467940591 | 0,639827075 | 0,999477707 | YES |
| 50 | 7,189307042 | 0,001301961 | 0,002852197 | 0,456476401 | 0,648047437 | 0,999477707 | YES |
| 51 | 14,3385944 | 0,00104283 | 0,002370184 | 0,439978376 | 0,659952769 | 0,999477707 | YES |
| 52 | 24,18771829 | 0,000980598 | 0,00222966 | 0,439797219 | 0,660083982 | 0,999477707 | YES |
| 53 | 207,6161908 | -0,00069945 | 0,001596788 | -0,438035203 | 0,661360765 | 0,999477707 | YES |

| Medication | Ticker | ASVnr | taxLevel | Phylum | Family | Genus | Species |
|---|---|---|---|---|---|---|---|
| LEDD | 54 | ASV29 | ASV | Firmicutes | Oscillospiraceae | UCG-002 | NA |
| LEDD | 55 | ASV46 | ASV | Bacteroidota | Bacteroidaceae | Bacteroides | NA |
| LEDD | 56 | ASV68 | ASV | Firmicutes | Ruminococcaceae | Faecalibacterium | NA |
| LEDD | 57 | ASV8 | ASV | Firmicutes | Ruminococcaceae | Faecalibacterium | prausnitzii |
| LEDD | 58 | ASV138 | ASV | Bacteroidota | Rikenellaceae | Alistipes | obesi |
| LEDD | 59 | ASV23 | ASV | Firmicutes | Streptococcaceae | Streptococcus | NA |
| LEDD | 60 | ASV33 | ASV | Firmicutes | Peptostreptococcaceae | Intestinibacter | bartlettii |
| LEDD | 61 | ASV77 | ASV | Firmicutes | Lachnospiraceae | [Ruminococcus] torques group | NA |
| LEDD | 62 | ASV190 | ASV | Firmicutes | Ruminococcaceae | UBA1819 | NA |
| LEDD | 63 | ASV123 | ASV | Firmicutes | Lachnospiraceae | [Ruminococcus] torques group | NA |
| LEDD | 64 | ASV126 | ASV | Bacteroidota | Tannerellaceae | Parabacteroides | distasonis |
| LEDD | 65 | ASV362 | ASV | Firmicutes | Ruminococcaceae | NA | NA |
| LEDD | 66 | ASV133 | ASV | Actinobacteriota | Eggerthellaceae | Senegalimassilia | anaerobia |
| LEDD | 67 | ASV287 | ASV | Firmicutes | Christensenellaceae | Christensenellaceae R-7 group | NA |
| LEDD | 68 | ASV357 | ASV | Firmicutes | Lachnospiraceae | Marvinbryantia | NA |
| LEDD | 69 | ASV16 | ASV | Firmicutes | Peptostreptococcaceae | Romboutsia | NA |
| LEDD | 70 | ASV315 | ASV | Firmicutes | Anaerovoracaceae | Family XIII UCG-001 | NA |
| LEDD | 71 | ASV3 | ASV | Firmicutes | Ruminococcaceae | Subdoligranulum | NA |
| LEDD | 72 | ASV129 | ASV | Firmicutes | Christensenellaceae | Christensenellaceae R-7 group | NA |
| LEDD | 73 | ASV128 | ASV | Firmicutes | Oscillospiraceae | NK4A214 group | NA |
| LEDD | 74 | ASV73 | ASV | Firmicutes | Oscillospiraceae | UCG-005 | NA |
| LEDD | 75 | ASV136 | ASV | Firmicutes | Ruminococcaceae | NA | NA |
| LEDD | 76 | ASV127 | ASV | Firmicutes | Lachnospiraceae | Lachnospiraceae NK4A136 group | bacterium |
| LEDD | 77 | ASV139 | ASV | Firmicutes | Christensenellaceae | Christensenellaceae R-7 group | NA |
| LEDD | 78 | ASV387 | ASV | Firmicutes | Lachnospiraceae | NA | NA |
| LEDD | 79 | ASV125 | ASV | Firmicutes | Oscillospiraceae | UCG-002 | NA |
| LEDD | 80 | ASV117 | ASV | Bacteroidota | Rikenellaceae | Alistipes | finegoldii |
| LEDD | 81 | ASV201 | ASV | Firmicutes | Oscillospiraceae | UCG-002 | NA |
| LEDD | 82 | ASV268 | ASV | Firmicutes | Ruminococcaceae | Incertae Sedis | NA |
| LEDD | 83 | ASV137 | ASV | Firmicutes | Butyricicoccaceae | Butyricicoccus | faecihominis |

| Ticker | DESeq2_baseMean | DESeq2_log2-FoldChange | DESeq2_lfcSE | DESeq2_stat | DESeq2_pvalue | DESeq2_padj | DESeq2 converged |
|---|---|---|---|---|---|---|---|
| 54 | 625,5412024 | -0,001211071 | 0,002820891 | -0,429322094 | 0,667688841 | 0,999477707 | YES |
| 55 | 124,2794829 | 0,000950927 | 0,00222088 | 0,428175635 | 0,668523254 | 0,999477707 | YES |
| 56 | 93,09663263 | -0,001179804 | 0,002832871 | -0,416469365 | 0,67706658 | 0,999477707 | YES |
| 57 | 972,1575633 | -0,001056044 | 0,00255199 | -0,413811854 | 0,679011902 | 0,999477707 | YES |
| 58 | 78,85312392 | 0,000400492 | 0,001013599 | 0,395118673 | 0,692755309 | 0,999477707 | YES |
| 59 | 205,3641771 | 0,000831869 | 0,002105979 | 0,395003494 | 6,93E-01 | 9,99E-01 | YES |
| 60 | 34,94531112 | 0,000572851 | 0,001481238 | 0,386737835 | 0,698950303 | 0,999477707 | YES |
| 61 | 140,4526504 | 0,001302464 | 0,003382508 | 0,385058642 | 0,700193969 | 0,999477707 | YES |
| 62 | 26,24405822 | 0,000637374 | 0,001665185 | 0,382764626 | 0,701894292 | 0,999477707 | YES |
| 63 | 121,3343881 | -0,001547903 | 0,004059083 | -0,381342922 | 0,702948808 | 0,999477707 | YES |
| 64 | 100,1236925 | 0,000987596 | 0,002701649 | 0,365553255 | 0,714698462 | 0,999477707 | YES |
| 65 | 14,09617392 | -0,000450269 | 0,001242865 | -0,362283384 | 0,717140274 | 0,999477707 | YES |
| 66 | 17,57410805 | -0,000449468 | 0,001245323 | -0,360924882 | 0,718155602 | 0,999477707 | YES |
| 67 | 28,75714711 | -0,000743131 | 0,002073525 | -0,358390112 | 0,72005139 | 0,999477707 | YES |
| 68 | 37,41460798 | 0,000979703 | 0,00281146 | 0,348467842 | 0,72748886 | 0,999477707 | YES |
| 69 | 37,6955578 | 0,000976815 | 0,002814618 | 0,347050815 | 0,728553144 | 0,999477707 | YES |
| 70 | 14,13608779 | -0,001040098 | 0,003002525 | -0,346407648 | 0,729036378 | 0,999477707 | YES |
| 71 | 488,2169575 | 0,000671453 | 0,001979474 | 0,339207937 | 0,73445309 | 0,999477707 | YES |
| 72 | 88,04840875 | -0,000297791 | 0,000887508 | -0,335535987 | 0,737220802 | 0,999477707 | YES |
| 73 | 102,520782 | 0,000534027 | 0,001602014 | 0,333347405 | 0,73887206 | 0,999477707 | YES |
| 74 | 196,5919775 | 0,000317851 | 0,000957721 | 0,331882172 | 0,739978233 | 0,999477707 | YES |
| 75 | 137,4812202 | -0,000800986 | 0,002460207 | -0,325576724 | 0,744744625 | 0,999477707 | YES |
| 76 | 76,66129975 | -0,001622572 | 0,004983812 | -0,325568472 | 0,744750869 | 0,999477707 | YES |
| 77 | 174,3923518 | 0,001077871 | 0,003510949 | 0,307002846 | 0,758841206 | 0,999477707 | YES |
| 78 | 12,36412939 | -0,001690132 | 0,005549316 | -0,304565834 | 0,760696845 | 0,999477707 | YES |
| 79 | 223,9543426 | 0,000776891 | 0,002563692 | 0,30303603 | 0,761862404 | 0,999477707 | YES |
| 80 | 42,22019237 | 0,001639266 | 0,005543188 | 0,295726128 | 0,767439242 | 0,999477707 | YES |
| 81 | 31,71809199 | 0,00159078 | 0,005551372 | 0,286556257 | 0,774452112 | 0,999477707 | YES |
| 82 | 13,17567418 | 0,001013976 | 0,003802333 | 0,266672146 | 0,789721601 | 0,999477707 | YES |
| 83 | 25,09711944 | 0,000598985 | 0,002274826 | 0,263310072 | 0,792311597 | 0,999477707 | YES |

| Medication | Ticker | ASVnr | taxLevel | Phylum | Family | Genus | Species |
|---|---|---|---|---|---|---|---|
| LEDD | 84 | ASV152 | ASV | Firmicutes | Lachnospiraceae | Roseburia | hominis |
| LEDD | 85 | ASV141 | ASV | Firmicutes | Lachnospiraceae | Lachnoclostridium | NA |
| LEDD | 86 | ASV203 | ASV | Firmicutes | NA | NA | NA |
| LEDD | 87 | ASV25 | ASV | Actinobacteriota | Bifidobacteriaceae | Bifidobacterium | NA |
| LEDD | 88 | ASV9 | ASV | Bacteroidota | Bacteroidaceae | Bacteroides | NA |
| LEDD | 89 | ASV57 | ASV | Firmicutes | Lachnospiraceae | Roseburia | inulinivorans |
| LEDD | 90 | ASV295 | ASV | Firmicutes | Ruminococcaceae | Negativibacillus | NA |
| LEDD | 91 | ASV93 | ASV | Firmicutes | Oscillospiraceae | NK4A214 group | NA |
| LEDD | 92 | ASV28 | ASV | Firmicutes | Clostridiaceae | Clostridium sensu stricto 1 | NA |
| LEDD | 93 | ASV91 | ASV | Firmicutes | Lachnospiraceae | [Eubacterium] eligens group | NA |
| LEDD | 94 | ASV1 | ASV | Actinobacteriota | Bifidobacteriaceae | Bifidobacterium | NA |
| LEDD | 95 | ASV192 | ASV | Bacteroidota | Marinifilaceae | Odoribacter | splanchnicus |
| LEDD | 96 | ASV47 | ASV | Bacteroidota | Rikenellaceae | Alistipes | NA |
| LEDD | 97 | ASV49 | ASV | Bacteroidota | Bacteroidaceae | Bacteroides | NA |
| LEDD | 98 | ASV222 | ASV | Firmicutes | Oscillospiraceae | NK4A214 group | NA |
| LEDD | 99 | ASV70 | ASV | Firmicutes | Lachnospiraceae | Roseburia | intestinalis |
| LEDD | 100 | ASV701 | ASV | Firmicutes | Lachnospiraceae | Lachnoclostridium | NA |
| LEDD | 101 | ASV26 | ASV | Euryarchaeota | Methanobacteriaceae | Methanobrevibacter | NA |
| LEDD | 102 | ASV39 | ASV | Bacteroidota | Tannerellaceae | Parabacteroides | merdae |
| LEDD | 103 | ASV94 | ASV | Bacteroidota | Rikenellaceae | Alistipes | shahii |
| LEDD | 104 | ASV48 | ASV | Bacteroidota | Bacteroidaceae | Bacteroides | caccae |
| LEDD | 105 | ASV35 | ASV | Firmicutes | Lachnospiraceae | Agathobacter | NA |
| LEDD | 106 | ASV303 | ASV | Firmicutes | Lachnospiraceae | GCA-900066575 | NA |
| LEDD | 107 | ASV86 | ASV | Firmicutes | Lachnospiraceae | [Eubacterium] hallii group | NA |
| LEDD | 108 | ASV213 | ASV | Firmicutes | NA | NA | NA |
| LEDD | 109 | ASV681 | ASV | Firmicutes | Oscillospiraceae | NA | NA |
| LEDD | 110 | ASV162 | ASV | Desulfobacterota | Desulfovibrionaceae | Desulfovibrio | NA |
| LEDD | 111 | ASV172 | ASV | Proteobacteria | Pasteurellaceae | Haemophilus | NA |
| LEDD | 112 | ASV130 | ASV | Proteobacteria | Enterobacteriaceae | Klebsiella | NA |
| LEDD | 113 | ASV487 | ASV | Firmicutes | Lachnospiraceae | Howardella | ureilytica |

| Ticker | DESeq2_baseMean | DESeq2_log2-FoldChange | DESeq2_lfcSE | DESeq2_stat | DESeq2_pvalue | DESeq2_padj | DESeq2 converged |
|---|---|---|---|---|---|---|---|
| 84 | 66,12943999 | -0,001009294 | 0,00387365 | -0,260553735 | 0,794436671 | 0,999477707 | YES |
| 85 | 56,93268433 | 0,000321287 | 0,001234553 | 0,26024576 | 0,794674209 | 0,999477707 | YES |
| 86 | 35,48121853 | -0,001442919 | 0,005544674 | -0,260235144 | 0,794682397 | 0,999477707 | YES |
| 87 | 314,5076205 | -0,000239213 | 0,000958966 | -0,249449202 | 0,80301333 | 0,999477707 | YES |
| 88 | 654,2550208 | 0,000660533 | 0,002650424 | 0,249217721 | 0,803192372 | 0,999477707 | YES |
| 89 | 164,1997714 | -0,000783244 | 0,003151995 | -0,248491447 | 0,803754187 | 0,999477707 | YES |
| 90 | 20,88080254 | 0,000809385 | 0,003281896 | 0,246621215 | 0,805201385 | 0,999477707 | YES |
| 91 | 97,03939266 | 0,000586965 | 0,002389143 | 0,245680122 | 0,805929861 | 0,999477707 | YES |
| 92 | 60,47003898 | 0,000338812 | 0,001379102 | 0,245675625 | 0,805933342 | 0,999477707 | YES |
| 93 | 72,73003574 | 0,000776309 | 0,003181545 | 0,244003648 | 0,807227994 | 0,999477707 | YES |
| 94 | 329,4609511 | 0,000730019 | 0,003010411 | 0,242498133 | 0,808394203 | 0,999477707 | YES |
| 95 | 31,05010717 | 0,000134611 | 0,000556115 | 0,242055983 | 0,808736785 | 0,999477707 | YES |
| 96 | 180,269199 | -0,000785294 | 0,003265509 | -0,240481295 | 0,809957162 | 0,999477707 | YES |
| 97 | 189,3186947 | -0,00092144 | 0,003838871 | -0,240029023 | 0,810307757 | 0,999477707 | YES |
| 98 | 52,54840517 | 0,00019057 | 0,000798123 | 0,238773218 | 0,811281438 | 0,999477707 | YES |
| 99 | 135,8032156 | -0,001295576 | 0,005478199 | -0,236496774 | 0,81304721 | 0,999477707 | YES |
| 100 | 5,023366844 | 0,00131302 | 0,00555287 | 0,236457961 | 0,813077325 | 0,999477707 | YES |
| 101 | 187,2991034 | 0,000487941 | 0,002080274 | 0,23455633 | 0,814553108 | 0,999477707 | YES |
| 102 | 339,1420055 | 0,000117373 | 0,000513253 | 0,228683832 | 0,819114662 | 0,999477707 | YES |
| 103 | 93,3536042 | 0,000736075 | 0,003354028 | 0,219459888 | 0,826291823 | 0,999477707 | YES |
| 104 | 271,0007549 | 0,000523821 | 0,002419582 | 0,216492309 | 0,82860402 | 0,999477707 | YES |
| 105 | 348,4599683 | -0,001142973 | 0,005379518 | -0,212467487 | 0,83174234 | 0,999477707 | YES |
| 106 | 8,981213672 | -0,000687542 | 0,003280193 | -0,209604072 | 0,833976701 | 0,999477707 | YES |
| 107 | 58,67428243 | -0,000226381 | 0,001080936 | -0,209430615 | 0,834112095 | 0,999477707 | YES |
| 108 | 21,48342194 | 0,001132797 | 0,005547053 | 0,204216092 | 0,838184636 | 0,999477707 | YES |
| 109 | 6,569822943 | -0,001129681 | 0,00555455 | -0,20337939 | 0,838838508 | 0,999477707 | YES |
| 110 | 172,3096971 | 0,000225989 | 0,001126951 | 0,20053109 | 0,841065246 | 0,999477707 | YES |
| 111 | 12,64520864 | -0,000989729 | 0,005117927 | -0,193384841 | 0,846657587 | 0,999477707 | YES |
| 112 | 60,97545019 | 0,001067339 | 0,005556281 | 0,192095916 | 0,847667074 | 0,999477707 | YES |
| 113 | 10,23380724 | -0,000443158 | 0,002327331 | -0,190414584 | 0,84898427 | 0,999477707 | YES |

| Medication | Ticker | ASVnr | taxLevel | Phylum | Family | Genus | Species |
|---|---|---|---|---|---|---|---|
| LEDD | 114 | ASV120 | ASV | Firmicutes | Lachnospiraceae | Anaerostipes | hadrus |
| LEDD | 115 | ASV276 | ASV | Firmicutes | Oscillospiraceae | NK4A214 group | NA |
| LEDD | 116 | ASV879 | ASV | Firmicutes | NA | NA | NA |
| LEDD | 117 | ASV415 | ASV | Firmicutes | Oscillospiraceae | Oscillospira | NA |
| LEDD | 118 | ASV81 | ASV | Firmicutes | Lachnospiraceae | Lachnospiraceae NK4A136 group | NA |
| LEDD | 119 | ASV270 | ASV | Firmicutes | Christensenellaceae | Christensenellaceae R-7 group | NA |
| LEDD | 120 | ASV159 | ASV | Firmicutes | Lachnospiraceae | Roseburia | NA |
| LEDD | 121 | ASV673 | ASV | Firmicutes | Christensenellaceae | Christensenellaceae R-7 group | NA |
| LEDD | 122 | ASV20 | ASV | Bacteroidota | Prevotellaceae | Prevotella | copri |
| LEDD | 123 | ASV355 | ASV | Firmicutes | NA | NA | NA |
| LEDD | 124 | ASV101 | ASV | Firmicutes | Lachnospiraceae | Roseburia | NA |
| LEDD | 125 | ASV593 | ASV | Firmicutes | Lachnospiraceae | NA | NA |
| LEDD | 126 | ASV145 | ASV | Bacteroidota | Muribaculaceae | NA | NA |
| LEDD | 127 | ASV143 | ASV | Firmicutes | Peptostreptococcaceae | Terrisporobacter | mayombei |
| LEDD | 128 | ASV436 | ASV | Firmicutes | Christensenellaceae | Christensenellaceae R-7 group | NA |
| LEDD | 129 | ASV630 | ASV | Firmicutes | Ruminococcaceae | Angelakisella | NA |
| LEDD | 130 | ASV100 | ASV | Firmicutes | Christensenellaceae | NA | NA |
| LEDD | 131 | ASV109 | ASV | Firmicutes | [Eubacterium] coprostanoligenes group | NA | NA |
| LEDD | 132 | ASV305 | ASV | Firmicutes | Lachnospiraceae | Lachnospiraceae UCG-004 | NA |
| LEDD | 133 | ASV256 | ASV | Firmicutes | Lachnospiraceae | Blautia | NA |
| LEDD | 134 | ASV52 | ASV | Firmicutes | Lachnospiraceae | Coprococcus | eutactus |
| LEDD | 135 | ASV338 | ASV | Firmicutes | Lachnospiraceae | Lachnotalea | NA |
| LEDD | 136 | ASV457 | ASV | Firmicutes | Lachnospiraceae | NA | NA |
| LEDD | 137 | ASV509 | ASV | Firmicutes | Lachnospiraceae | Marvinbryantia | NA |
| LEDD | 138 | ASV522 | ASV | Firmicutes | UCG-010 | NA | NA |
| LEDD | 139 | ASV98 | ASV | Firmicutes | Acidaminococcaceae | Phascolarctobacterium | succinatutens |
| LEDD | 140 | ASV307 | ASV | Firmicutes | Lachnospiraceae | Lachnoclostridium | NA |
| LEDD | 141 | ASV110 | ASV | Firmicutes | Ruminococcaceae | Faecalibacterium | prausnitzii |
| LEDD | 142 | ASV242 | ASV | Firmicutes | Lachnospiraceae | Tyzzerella | NA |
| LEDD | 143 | ASV533 | ASV | Firmicutes | Ruminococcaceae | Negativibacillus | massiliensis |

| Ticker | DESeq2_baseMean | DESeq2_log2-FoldChange | DESeq2_lfcSE | DESeq2_stat | DESeq2_pvalue | DESeq2_padj | DESeq2 converged |
|---|---|---|---|---|---|---|---|
| 114 | 18,72093584 | 0,001045971 | 0,005552073 | 0,188392929 | 0,850568642 | 0,999477707 | YES |
| 115 | 33,3035992 | 0,001030357 | 0,005557108 | 0,185412448 | 0,852905546 | 0,999477707 | YES |
| 116 | 1,709236849 | 0,001027438 | 0,005553322 | 0,185013248 | 0,853218644 | 0,999477707 | YES |
| 117 | 10,18965444 | 0,001013426 | 0,00554581 | 0,182737323 | 0,855004129 | 0,999477707 | YES |
| 118 | 113,9387832 | -0,000672727 | 0,003711411 | -0,18125896 | 0,856164318 | 0,999477707 | YES |
| 119 | 10,62271582 | 0,000830013 | 0,004644082 | 0,178724793 | 0,858153801 | 0,999477707 | YES |
| 120 | 53,37439601 | -0,000323068 | 0,001846034 | -0,175006221 | 0,861074745 | 0,999477707 | YES |
| 121 | 6,111053812 | 0,00096237 | 0,005553458 | 0,173292025 | 0,862421892 | 0,999477707 | YES |
| 122 | 453,818211 | -0,000849571 | 0,004906219 | -0,173161969 | 0,862524117 | 0,999477707 | YES |
| 123 | 23,11357952 | -0,000954495 | 0,005558969 | -0,17170358 | 0,863670571 | 0,999477707 | YES |
| 124 | 98,61254423 | 0,000907299 | 0,005402366 | 0,167944688 | 0,866626794 | 0,999477707 | YES |
| 125 | 5,084609489 | 0,00093092 | 0,005554163 | 0,167607523 | 0,866892052 | 0,999477707 | YES |
| 126 | 131,9175803 | -0,00092887 | 0,005547831 | -0,167429421 | 0,867032177 | 0,999477707 | YES |
| 127 | 7,024029205 | 0,000911852 | 0,005554016 | 0,164178887 | 0,869590322 | 0,999477707 | YES |
| 128 | 5,736317313 | 0,000907723 | 0,005547901 | 0,163615603 | 0,870033761 | 0,999477707 | YES |
| 129 | 2,985334318 | 0,000903061 | 0,00555203 | 0,162654239 | 0,870790679 | 0,999477707 | YES |
| 130 | 306,1393149 | 0,000394674 | 0,002442469 | 0,161588249 | 0,871630112 | 0,999477707 | YES |
| 131 | 88,6634389 | -0,000553887 | 0,003445831 | -0,160741319 | 0,872297145 | 0,999477707 | YES |
| 132 | 12,3174562 | -0,000882474 | 0,005549299 | -0,159024403 | 0,873649647 | 0,999477707 | YES |
| 133 | 8,884013475 | 0,000881567 | 0,00554888 | 0,158873014 | 0,873768922 | 0,999477707 | YES |
| 134 | 288,3266723 | -0,000670148 | 0,00428789 | -0,156288471 | 0,875805644 | 0,999477707 | YES |
| 135 | 4,808606648 | -0,000854277 | 0,005551166 | -0,153891432 | 0,87769534 | 0,999477707 | YES |
| 136 | 6,921613119 | -0,000842015 | 0,005550607 | -0,151697847 | 0,879425256 | 0,999477707 | YES |
| 137 | 8,021765729 | 0,000839443 | 0,005552001 | 0,151196523 | 0,879820693 | 0,999477707 | YES |
| 138 | 5,605550656 | 0,00083653 | 0,005551074 | 0,150697016 | 0,880214727 | 0,999477707 | YES |
| 139 | 200,6469766 | -0,000416144 | 0,002775267 | -0,149947397 | 0,880806117 | 0,999477707 | YES |
| 140 | 5,512274824 | 0,000808425 | 0,005552477 | 0,145597244 | 0,884239343 | 0,999477707 | YES |
| 141 | 45,96783544 | -0,000806918 | 0,005557007 | -0,145207382 | 0,884547138 | 0,999477707 | YES |
| 142 | 59,50731398 | 0,000701045 | 0,004829223 | 0,145167313 | 0,884578773 | 0,999477707 | YES |
| 143 | 9,123251396 | 0,000804598 | 0,005554794 | 0,144847418 | 0,884831343 | 0,999477707 | YES |

| Medication | Ticker | ASVnr | taxLevel | Phylum | Family | Genus | Species |
|---|---|---|---|---|---|---|---|
| LEDD | 144 | ASV257 | ASV | Bacteroidota | Bacteroidaceae | Bacteroides | ovatus |
| LEDD | 145 | ASV160 | ASV | Firmicutes | Ruminococcaceae | Ruminococcus | callidus |
| LEDD | 146 | ASV153 | ASV | Firmicutes | Streptococcaceae | Streptococcus | NA |
| LEDD | 147 | ASV218 | ASV | Firmicutes | Anaerovoracaceae | Family XIII AD3011 group | NA |
| LEDD | 148 | ASV450 | ASV | Firmicutes | NA | NA | NA |
| LEDD | 149 | ASV820 | ASV | Firmicutes | Lachnospiraceae | GCA-900066575 | NA |
| LEDD | 150 | ASV78 | ASV | Firmicutes | Lachnospiraceae | [Eubacterium] eligens group | NA |
| LEDD | 151 | ASV937 | ASV | Firmicutes | UCG-010 | NA | NA |
| LEDD | 152 | ASV75 | ASV | Firmicutes | Lachnospiraceae | [Ruminococcus] gauvreauii group | NA |
| LEDD | 153 | ASV261 | ASV | Firmicutes | Anaerovoracaceae | Family XIII UCG-001 | NA |
| LEDD | 154 | ASV215 | ASV | Firmicutes | Oscillospiraceae | UCG-002 | NA |
| LEDD | 155 | ASV328 | ASV | Firmicutes | Anaerovoracaceae | [Eubacterium] brachy group | NA |
| LEDD | 156 | ASV406 | ASV | Firmicutes | NA | NA | NA |
| LEDD | 157 | ASV413 | ASV | Firmicutes | Oscillospiraceae | Oscillibacter | NA |
| LEDD | 158 | ASV234 | ASV | Cyanobacteria | NA | NA | NA |
| LEDD | 159 | ASV444 | ASV | Firmicutes | Erysipelatoclostridiaceae | Coprobacillus | cateniformis |
| LEDD | 160 | ASV112 | ASV | Firmicutes | Oscillospiraceae | UCG-002 | NA |
| LEDD | 161 | ASV808 | ASV | Desulfobacterota | Desulfovibrionaceae | NA | NA |
| LEDD | 162 | ASV85 | ASV | Firmicutes | Lachnospiraceae | Coprococcus | NA |
| LEDD | 163 | ASV370 | ASV | Firmicutes | UCG-010 | NA | NA |
| LEDD | 164 | ASV485 | ASV | Firmicutes | Lachnospiraceae | NA | NA |
| LEDD | 165 | ASV451 | ASV | Firmicutes | Oscillospiraceae | UCG-005 | NA |
| LEDD | 166 | ASV180 | ASV | Firmicutes | Ruminococcaceae | Ruminococcus | NA |
| LEDD | 167 | ASV310 | ASV | Firmicutes | Lachnospiraceae | NA | NA |
| LEDD | 168 | ASV92 | ASV | Firmicutes | Erysipelotrichaceae | Turicibacter | sanguinis |
| LEDD | 169 | ASV134 | ASV | Actinobacteriota | Eggerthellaceae | Enterorhabdus | NA |
| LEDD | 170 | ASV207 | ASV | Firmicutes | Lachnospiraceae | Blautia | NA |
| LEDD | 171 | ASV243 | ASV | Firmicutes | NA | NA | NA |
| LEDD | 172 | ASV104 | ASV | Firmicutes | Christensenellaceae | Christensenellaceae R-7 group | NA |
| LEDD | 173 | ASV425 | ASV | Firmicutes | Oscillospiraceae | NK4A214 group | NA |

| Ticker | DESeq2_baseMean | DESeq2_log2-FoldChange | DESeq2_lfcSE | DESeq2_stat | DESeq2_pvalue | DESeq2_padj | DESeq2 converged |
|---|---|---|---|---|---|---|---|
| 144 | 16,17825249 | -0,000803919 | 0,005558835 | -0,144620129 | 0,885010803 | 0,999477707 | YES |
| 145 | 108,8714557 | -0,000787067 | 0,00555383 | -0,141716048 | 0,887304298 | 0,999477707 | YES |
| 146 | 19,28836149 | 0,000780176 | 0,005553165 | 0,140492128 | 0,888271171 | 0,999477707 | YES |
| 147 | 8,044960964 | -0,000356121 | 0,002563667 | -0,138910818 | 0,889520622 | 0,999477707 | YES |
| 148 | 3,918883104 | -0,000770852 | 0,00555147 | -0,138855527 | 0,889564315 | 0,999477707 | YES |
| 149 | 4,748954977 | 0,000759665 | 0,005555196 | 0,136748568 | 0,891229538 | 0,999477707 | YES |
| 150 | 99,71543913 | -0,000370579 | 0,002725318 | -0,135976439 | 0,891839907 | 0,999477707 | YES |
| 151 | 3,458344526 | -0,000754636 | 0,005555039 | -0,135847191 | 0,891942084 | 0,999477707 | YES |
| 152 | 43,5767171 | 0,000361968 | 0,002697408 | 0,134190994 | 0,893251547 | 0,999477707 | YES |
| 153 | 9,902869365 | 0,000743462 | 0,005553591 | 0,133870514 | 0,893504967 | 0,999477707 | YES |
| 154 | 130,7202466 | 0,000730893 | 0,005551186 | 0,131664298 | 0,895249826 | 0,999477707 | YES |
| 155 | 1,144020885 | 0,000703342 | 0,005558138 | 0,126542661 | 0,899302381 | 0,999477707 | YES |
| 156 | 9,058926154 | 0,000159256 | 0,001265741 | 0,125820361 | 0,899874124 | 0,999477707 | YES |
| 157 | 9,133748374 | 0,000688905 | 0,00555554 | 0,124003294 | 0,901312667 | 0,999477707 | YES |
| 158 | 9,97741108 | -0,000687484 | 0,005550639 | -0,123856773 | 0,90142868 | 0,999477707 | YES |
| 159 | 7,089198105 | 0,000680132 | 0,005551719 | 0,122508351 | 0,902496432 | 0,999477707 | YES |
| 160 | 40,77630395 | 0,000675833 | 0,005548736 | 0,121799508 | 0,903057804 | 0,999477707 | YES |
| 161 | 1,551220695 | 0,000658866 | 0,005553312 | 0,118643702 | 0,905557641 | 0,999477707 | YES |
| 162 | 293,4496484 | -0,000650239 | 0,005551011 | -0,117138874 | 0,906750005 | 0,999477707 | YES |
| 163 | 11,07180546 | -0,000645235 | 0,005551262 | -0,116232144 | 0,907468562 | 0,999477707 | YES |
| 164 | 3,691944363 | -0,000644798 | 0,005554732 | -0,116080909 | 0,907588419 | 0,999477707 | YES |
| 165 | 13,08387694 | -0,000642024 | 0,005554913 | -0,115577725 | 0,907987217 | 0,999477707 | YES |
| 166 | 57,87216551 | -0,000636713 | 0,005550483 | -0,114713022 | 0,908672592 | 0,999477707 | YES |
| 167 | 11,38247968 | -0,000625783 | 0,005559297 | -0,112565067 | 0,910375381 | 0,999477707 | YES |
| 168 | 24,93246918 | 0,00062305 | 0,005555717 | 0,112145719 | 0,910707867 | 0,999477707 | YES |
| 169 | 7,464465206 | -0,000201546 | 0,001802745 | -0,111799298 | 0,910982544 | 0,999477707 | YES |
| 170 | 46,96817061 | 0,000619751 | 0,005552992 | 0,11160666 | 0,911135291 | 0,999477707 | YES |
| 171 | 13,06580814 | -0,000604477 | 0,005553792 | -0,108840365 | 0,913329107 | 0,999477707 | YES |
| 172 | 53,48045156 | -0,000125193 | 0,001151794 | -0,108694166 | 0,91344507 | 0,999477707 | YES |
| 173 | 21,25846324 | -0,000602065 | 0,005552261 | -0,108435926 | 0,913649904 | 0,999477707 | YES |

| Medication | Ticker | ASVnr | taxLevel | Phylum | Family | Genus | Species |
|---|---|---|---|---|---|---|---|
| LEDD | 174 | ASV225 | ASV | Bacteroidota | Muribaculaceae | NA | NA |
| LEDD | 175 | ASV554 | ASV | Firmicutes | Lachnospiraceae | NA | NA |
| LEDD | 176 | ASV216 | ASV | Firmicutes | Lachnospiraceae | Lachnospira | pectinoschiza |
| LEDD | 177 | ASV299 | ASV | Firmicutes | Streptococcaceae | Lactococcus | NA |
| LEDD | 178 | ASV194 | ASV | Bacteroidota | Bacteroidaceae | Bacteroides | intestinalis |
| LEDD | 179 | ASV517 | ASV | Firmicutes | Lachnospiraceae | Blautia | NA |
| LEDD | 180 | ASV206 | ASV | Firmicutes | Oscillospiraceae | Intestinimonas | NA |
| LEDD | 181 | ASV249 | ASV | Proteobacteria | NA | NA | NA |
| LEDD | 182 | ASV417 | ASV | Firmicutes | UCG-010 | NA | NA |
| LEDD | 183 | ASV54 | ASV | Bacteroidota | Bacteroidaceae | Bacteroides | NA |
| LEDD | 184 | ASV61 | ASV | Firmicutes | Erysipelatoclostridiaceae | Catenibacterium | mitsuokai |
| LEDD | 185 | ASV187 | ASV | Firmicutes | NA | NA | NA |
| LEDD | 186 | ASV760 | ASV | Firmicutes | UCG-010 | NA | NA |
| LEDD | 187 | ASV363 | ASV | Bacteroidota | Rikenellaceae | Alistipes | NA |
| LEDD | 188 | ASV184 | ASV | Firmicutes | Ruminococcaceae | Ruminococcus | NA |
| LEDD | 189 | ASV154 | ASV | Firmicutes | NA | NA | NA |
| LEDD | 190 | ASV113 | ASV | Firmicutes | Lachnospiraceae | NA | NA |
| LEDD | 191 | ASV386 | ASV | Firmicutes | Ruminococcaceae | DTU089 | NA |
| LEDD | 192 | ASV477 | ASV | Firmicutes | Oscillospiraceae | Intestinimonas | massiliensis |
| LEDD | 193 | ASV82 | ASV | Firmicutes | Lachnospiraceae | Coprococcus | eutactus |
| LEDD | 194 | ASV150 | ASV | Proteobacteria | Sutterellaceae | Sutterella | NA |
| LEDD | 195 | ASV687 | ASV | Firmicutes | Ruminococcaceae | DTU089 | NA |
| LEDD | 196 | ASV867 | ASV | Firmicutes | NA | NA | NA |
| LEDD | 197 | ASV262 | ASV | Firmicutes | Ruminococcaceae | Incertae Sedis | NA |
| LEDD | 198 | ASV380 | ASV | Firmicutes | Christensenellaceae | Christensenellaceae R-7 group | NA |
| LEDD | 199 | ASV255 | ASV | Actinobacteriota | Coriobacteriales Incertae Sedis | NA | NA |
| LEDD | 200 | ASV66 | ASV | Firmicutes | Butyricicoccaceae | Butyricicoccus | NA |
| LEDD | 201 | ASV50 | ASV | Bacteroidota | Prevotellaceae | Prevotella | NA |
| LEDD | 202 | ASV132 | ASV | Firmicutes | Ruminococcaceae | CAG-352 | NA |
| LEDD | 203 | ASV339 | ASV | Firmicutes | Monoglobaceae | Monoglobus | pectinilyticus |

| Ticker | DESeq2_baseMean | DESeq2_log2-FoldChange | DESeq2_lfcSE | DESeq2_stat | DESeq2_pvalue | DESeq2_padj | DESeq2 converged |
|---|---|---|---|---|---|---|---|
| 174 | 74,19388671 | 0,000602006 | 0,005555163 | 0,108368743 | 0,913703195 | 0,999477707 | YES |
| 175 | 8,514583524 | 0,000600878 | 0,005554093 | 0,108186537 | 0,913847724 | 0,999477707 | YES |
| 176 | 28,20609117 | -0,000399478 | 0,003742584 | -0,106738478 | 0,914996456 | 0,999477707 | YES |
| 177 | 7,317008837 | 0,000588489 | 0,005554709 | 0,105944219 | 0,91562661 | 0,999477707 | YES |
| 178 | 58,01666222 | 0,000586651 | 0,005557747 | 0,105555585 | 0,915934966 | 0,999477707 | YES |
| 179 | 15,87663876 | 0,000580467 | 0,005557224 | 0,104452756 | 0,916810058 | 0,999477707 | YES |
| 180 | 31,19176193 | 0,00049201 | 0,004729624 | 0,104027391 | 0,917147611 | 0,999477707 | YES |
| 181 | 22,37698387 | 0,000569599 | 0,005556391 | 0,102512366 | 0,918349998 | 0,999477707 | YES |
| 182 | 16,24033686 | 0,000561752 | 0,005505653 | 0,10203177 | 0,918731458 | 0,999477707 | YES |
| 183 | 140,0576948 | 0,000318959 | 0,003144385 | 0,101437726 | 0,91920299 | 0,999477707 | YES |
| 184 | 64,3604178 | -0,000560701 | 0,005553417 | -0,100965074 | 0,919578185 | 0,999477707 | YES |
| 185 | 27,08985775 | -0,000552175 | 0,005556697 | -0,099371075 | 0,920843648 | 0,999477707 | YES |
| 186 | 4,627302076 | 0,000551122 | 0,005552349 | 0,099259312 | 0,920932383 | 0,999477707 | YES |
| 187 | 9,007958881 | 0,000543755 | 0,005552629 | 0,097927572 | 0,921989806 | 0,999477707 | YES |
| 188 | 88,29850228 | -0,000539742 | 0,005548532 | -0,097276459 | 0,92250685 | 0,999477707 | YES |
| 189 | 20,03721474 | -0,000535507 | 0,005558149 | -0,096346231 | 0,923245595 | 0,999477707 | YES |
| 190 | 146,3177638 | -0,000534091 | 0,005555237 | -0,096141904 | 0,923407871 | 0,999477707 | YES |
| 191 | 8,43598098 | 0,000254879 | 0,002706185 | 0,094183941 | 0,924963041 | 0,999477707 | YES |
| 192 | 9,488323889 | 0,000518422 | 0,005549704 | 0,093414329 | 0,925574407 | 0,999477707 | YES |
| 193 | 92,07779332 | -0,000518174 | 0,005548967 | -0,093382118 | 0,925599996 | 0,999477707 | YES |
| 194 | 37,1261767 | -0,000186758 | 0,002002364 | -0,09326862 | 0,925690161 | 0,999477707 | YES |
| 195 | 4,303921459 | 0,000518159 | 0,005557207 | 0,093240823 | 0,925712244 | 0,999477707 | YES |
| 196 | 3,347437588 | 0,000517245 | 0,005549994 | 0,093197409 | 0,925746732 | 0,999477707 | YES |
| 197 | 11,2416058 | 0,000499535 | 0,005554484 | 0,089933711 | 0,928339892 | 0,999477707 | YES |
| 198 | 2,659951872 | 0,000496278 | 0,005553116 | 0,089369264 | 0,92878845 | 0,999477707 | YES |
| 199 | 6,394636888 | 0,000491413 | 0,00555439 | 0,088472831 | 0,929500877 | 0,999477707 | YES |
| 200 | 78,55695416 | -6,04E-05 | 0,000695233 | -0,08689095 | 0,930758193 | 0,999477707 | YES |
| 201 | 110,5712201 | -0,000481354 | 0,005551548 | -0,08670622 | 0,930905032 | 0,999477707 | YES |
| 202 | 23,21558955 | -0,000472621 | 0,0055585 | -0,085026662 | 0,932240194 | 0,999477707 | YES |
| 203 | 9,728213387 | -0,000472161 | 0,005557859 | -0,084953745 | 0,932298164 | 0,999477707 | YES |

| Medication | Ticker | ASVnr | taxLevel | Phylum | Family | Genus | Species |
|---|---|---|---|---|---|---|---|
| LEDD | 204 | ASV230 | ASV | Firmicutes | Ruminococcaceae | [Eubacterium] siraeum group | NA |
| LEDD | 205 | ASV308 | ASV | Firmicutes | Christensenellaceae | NA | NA |
| LEDD | 206 | ASV106 | ASV | Bacteroidota | Bacteroidaceae | Bacteroides | NA |
| LEDD | 207 | ASV12 | ASV | Firmicutes | Veillonellaceae | Dialister | invisus |
| LEDD | 208 | ASV535 | ASV | Firmicutes | Erysipelatoclostridiaceae | Asteroleplasma | NA |
| LEDD | 209 | ASV653 | ASV | Firmicutes | Lachnospiraceae | NA | NA |
| LEDD | 210 | ASV237 | ASV | Bacteroidota | Bacteroidaceae | Bacteroides | clarus |
| LEDD | 211 | ASV797 | ASV | Firmicutes | Peptococcaceae | Peptococcus | NA |
| LEDD | 212 | ASV148 | ASV | Firmicutes | NA | NA | NA |
| LEDD | 213 | ASV95 | ASV | Bacteroidota | Prevotellaceae | Prevotella | NA |
| LEDD | 214 | ASV443 | ASV | Firmicutes | Veillonellaceae | Veillonella | NA |
| LEDD | 215 | ASV508 | ASV | Firmicutes | Ruminococcaceae | NA | NA |
| LEDD | 216 | ASV185 | ASV | Firmicutes | NA | NA | NA |
| LEDD | 217 | ASV438 | ASV | Firmicutes | UCG-010 | NA | NA |
| LEDD | 218 | ASV254 | ASV | Firmicutes | Lachnospiraceae | Lachnospira | NA |
| LEDD | 219 | ASV4 | ASV | Firmicutes | Ruminococcaceae | Ruminococcus | bromii |
| LEDD | 220 | ASV828 | ASV | Firmicutes | Christensenellaceae | Christensenellaceae R-7 group | NA |
| LEDD | 221 | ASV557 | ASV | Firmicutes | Oscillospiraceae | UCG-005 | NA |
| LEDD | 222 | ASV351 | ASV | Firmicutes | Lachnospiraceae | Roseburia | NA |
| LEDD | 223 | ASV625 | ASV | Firmicutes | UCG-010 | NA | NA |
| LEDD | 224 | ASV1158 | ASV | Firmicutes | Ruminococcaceae | NA | NA |
| LEDD | 225 | ASV414 | ASV | Firmicutes | Oscillospiraceae | UCG-003 | NA |
| LEDD | 226 | ASV40 | ASV | Firmicutes | Erysipelotrichaceae | Holdemanella | NA |
| LEDD | 227 | ASV695 | ASV | Firmicutes | UCG-010 | NA | NA |
| LEDD | 228 | ASV529 | ASV | Firmicutes | Lachnospiraceae | NA | NA |
| LEDD | 229 | ASV813 | ASV | Firmicutes | Ruminococcaceae | NA | NA |
| LEDD | 230 | ASV286 | ASV | Firmicutes | Lachnospiraceae | NA | NA |
| LEDD | 231 | ASV195 | ASV | Bacteroidota | Barnesiellaceae | Barnesiella | intestinihominis |
| LEDD | 232 | ASV244 | ASV | Bacteroidota | Bacteroidaceae | Bacteroides | coprophilus |
| LEDD | 233 | ASV427 | ASV | Firmicutes | Lactobacillaceae | Lactobacillus | NA |

| Ticker | DESeq2_baseMean | DESeq2_log2-FoldChange | DESeq2_lfcSE | DESeq2_stat | DESeq2_pvalue | DESeq2_padj | DESeq2 converged |
|---|---|---|---|---|---|---|---|
| 204 | 25,94039767 | -0,000471494 | 0,005553881 | -0,084894587 | 0,932345195 | 0,999477707 | YES |
| 205 | 11,68386395 | 0,000470588 | 0,005554463 | 0,084722508 | 0,932482001 | 0,999477707 | YES |
| 206 | 55,6836761 | 0,000338947 | 0,004073546 | 0,083206927 | 0,933687005 | 0,999477707 | YES |
| 207 | 353,6568712 | 0,00045897 | 0,005553451 | 0,082645874 | 0,934133124 | 0,999477707 | YES |
| 208 | 2,585148566 | -0,000458956 | 0,005557575 | -0,082582073 | 0,934183856 | 0,999477707 | YES |
| 209 | 7,037408263 | -0,000458787 | 0,005555607 | -0,082580978 | 0,934184727 | 0,999477707 | YES |
| 210 | 32,82183895 | 0,00045752 | 0,005556008 | 0,082346905 | 0,934370857 | 0,999477707 | YES |
| 211 | 4,027590161 | -0,000456401 | 0,00555714 | -0,082128839 | 0,93454426 | 0,999477707 | YES |
| 212 | 56,24238155 | 0,000429389 | 0,005325587 | 0,080627594 | 0,93573812 | 0,999477707 | YES |
| 213 | 36,03751168 | -0,000447425 | 0,005557147 | -0,080513457 | 0,935828894 | 0,999477707 | YES |
| 214 | 7,709181677 | 0,000441287 | 0,00555196 | 0,079483035 | 0,936648425 | 0,999477707 | YES |
| 215 | 2,656381379 | -0,000439772 | 0,005557961 | -0,079124713 | 0,936933427 | 0,999477707 | YES |
| 216 | 84,18084758 | -0,000435828 | 0,00555076 | -0,078516849 | 0,937416928 | 0,999477707 | YES |
| 217 | 12,24334777 | 0,000277133 | 0,003554298 | 0,077971303 | 0,93785088 | 0,999477707 | YES |
| 218 | 28,4383693 | -0,000432297 | 0,005546971 | -0,077933934 | 0,937880606 | 0,999477707 | YES |
| 219 | 1047,396357 | 0,000306553 | 0,004023499 | 0,07619071 | 0,939267374 | 0,999477707 | YES |
| 220 | 1,275801653 | 0,000420246 | 0,005554948 | 0,075652592 | 0,939695494 | 0,999477707 | YES |
| 221 | 25,39383483 | 0,000418124 | 0,005560686 | 0,075192915 | 0,940061222 | 0,999477707 | YES |
| 222 | 20,84532238 | -0,000416688 | 0,005554711 | -0,075015237 | 0,940202589 | 0,999477707 | YES |
| 223 | 2,748017918 | 0,0004136 | 0,005558403 | 0,07440991 | 0,940684223 | 0,999477707 | YES |
| 224 | 1,533889816 | -0,000413157 | 0,005554665 | -0,074380185 | 0,940707875 | 0,999477707 | YES |
| 225 | 3,501742822 | 0,000409186 | 0,005558494 | 0,073614585 | 0,941317065 | 0,999477707 | YES |
| 226 | 211,0214756 | 0,00013165 | 0,001792726 | 0,073435532 | 0,941459543 | 0,999477707 | YES |
| 227 | 4,674932431 | -0,00040618 | 0,005550885 | -0,073173869 | 0,94166776 | 0,999477707 | YES |
| 228 | 4,63217094 | -0,000404272 | 0,005553227 | -0,072799494 | 0,941965673 | 0,999477707 | YES |
| 229 | 1,593405793 | 0,000395187 | 0,005555326 | 0,071136596 | 0,943289042 | 0,999477707 | YES |
| 230 | 29,60002579 | -0,000394331 | 0,005550691 | -0,071041725 | 0,943364548 | 0,999477707 | YES |
| 231 | 60,04490399 | -0,000390802 | 0,005552558 | -0,070382361 | 0,94388933 | 0,999477707 | YES |
| 232 | 55,67549779 | 0,000388718 | 0,005554255 | 0,069985632 | 0,944205095 | 0,999477707 | YES |
| 233 | 8,257075044 | 0,000386841 | 0,005555977 | 0,069626069 | 0,944491287 | 0,999477707 | YES |

| Medication | Ticker | ASVnr | taxLevel | Phylum | Family | Genus | Species |
|---|---|---|---|---|---|---|---|
| LEDD | 234 | ASV251 | ASV | Firmicutes | Oscillospiraceae | NK4A214 group | NA |
| LEDD | 235 | ASV727 | ASV | Firmicutes | Ruminococcaceae | Angelakisella | NA |
| LEDD | 236 | ASV703 | ASV | Firmicutes | Hydrogenoanaerobacterium | NA | NA |
| LEDD | 237 | ASV332 | ASV | Firmicutes | Lachnospiraceae | [Eubacterium] ventriosum group | NA |
| LEDD | 238 | ASV383 | ASV | Firmicutes | NA | NA | NA |
| LEDD | 239 | ASV142 | ASV | Firmicutes | Ruminococcaceae | Faecalibacterium | prausnitzii |
| LEDD | 240 | ASV271 | ASV | Firmicutes | Oscillospiraceae | NK4A214 group | NA |
| LEDD | 241 | ASV99 | ASV | Bacteroidota | Bacteroidaceae | Bacteroides | plebeius |
| LEDD | 242 | ASV238 | ASV | Firmicutes | Butyricicoccaceae | NA | NA |
| LEDD | 243 | ASV217 | ASV | Firmicutes | Peptococcaceae | Peptococcus | NA |
| LEDD | 244 | ASV587 | ASV | Firmicutes | Anaerovoracaceae | Family XIII AD3011 group | NA |
| LEDD | 245 | ASV171 | ASV | Firmicutes | Oscillospiraceae | NA | NA |
| LEDD | 246 | ASV155 | ASV | Firmicutes | Ruminococcaceae | Ruminococcus | NA |
| LEDD | 247 | ASV730 | ASV | Firmicutes | Ruminococcaceae | NA | NA |
| LEDD | 248 | ASV801 | ASV | Firmicutes | Christensenellaceae | Christensenellaceae R-7 group | NA |
| LEDD | 249 | ASV459 | ASV | Bacteroidota | Marinifilaceae | Butyricimonas | paravirosa |
| LEDD | 250 | ASV536 | ASV | Bacteroidota | Marinifilaceae | Butyricimonas | NA |
| LEDD | 251 | ASV470 | ASV | Firmicutes | Christensenellaceae | Christensenellaceae R-7 group | NA |
| LEDD | 252 | ASV317 | ASV | Firmicutes | NA | NA | NA |
| LEDD | 253 | ASV291 | ASV | Firmicutes | Erysipelatoclostridiaceae | Erysipelatoclostridium | NA |
| LEDD | 254 | ASV454 | ASV | Actinobacteriota | Actinomycetaceae | Actinomyces | odontolyticus |
| LEDD | 255 | ASV494 | ASV | Firmicutes | Anaerovoracaceae | Family XIII AD3011 group | NA |
| LEDD | 256 | ASV191 | ASV | Firmicutes | Lachnospiraceae | Lachnospiraceae NK4A136 group | NA |
| LEDD | 257 | ASV167 | ASV | Firmicutes | Oscillospiraceae | Intestinimonas | NA |
| LEDD | 258 | ASV1464 | ASV | Firmicutes | Peptococcaceae | NA | NA |
| LEDD | 259 | ASV247 | ASV | Euryarchaeota | Methanobacteriaceae | Methanosphaera | NA |
| LEDD | 260 | ASV294 | ASV | Firmicutes | Anaerovoracaceae | Mogibacterium | NA |
| LEDD | 261 | ASV22 | ASV | Bacteroidota | Bacteroidaceae | Bacteroides | uniformis |
| LEDD | 262 | ASV149 | ASV | Firmicutes | Oscillospiraceae | UCG-002 | NA |
| LEDD | 263 | ASV296 | ASV | Proteobacteria | Sutterellaceae | Sutterella | NA |

| Ticker | DESeq2_baseMean | DESeq2_log2-FoldChange | DESeq2_lfcSE | DESeq2_stat | DESeq2_pvalue | DESeq2_padj | DESeq2 converged |
|---|---|---|---|---|---|---|---|
| 234 | 15,98302489 | 0,000386046 | 0,00555197 | 0,069533123 | 0,944565268 | 0,999477707 | YES |
| 235 | 2,767870925 | 0,000383187 | 0,005548204 | 0,06906503 | 0,944937856 | 0,999477707 | YES |
| 236 | 2,818052049 | -0,000380411 | 0,005553816 | -0,068495366 | 0,945391309 | 0,999477707 | YES |
| 237 | 13,96035869 | -0,00037999 | 0,005551021 | -0,068454026 | 0,945424216 | 0,999477707 | YES |
| 238 | 27,5064993 | 0,000378154 | 0,005555688 | 0,068066186 | 0,945732947 | 0,999477707 | YES |
| 239 | 99,35817288 | -0,000228278 | 0,003365571 | -0,067827533 | 0,945922926 | 0,999477707 | YES |
| 240 | 34,33950651 | 0,000374197 | 0,005555118 | 0,067360791 | 0,946294482 | 0,999477707 | YES |
| 241 | 56,73587672 | -0,000369565 | 0,005553151 | -0,066550498 | 0,946939556 | 0,999477707 | YES |
| 242 | 41,99365617 | 0,000166167 | 0,002508257 | 0,066248107 | 0,947180297 | 0,999477707 | YES |
| 243 | 54,36812989 | 0,000364802 | 0,005553951 | 0,065683332 | 9,48E-01 | 9,99E-01 | YES |
| 244 | 6,842526689 | 0,000362111 | 0,005553491 | 0,065204135 | 0,948011469 | 0,999477707 | YES |
| 245 | 15,5768961 | 0,00035895 | 0,005553276 | 0,064637473 | 0,948462648 | 0,999477707 | YES |
| 246 | 29,01572216 | -0,00035693 | 0,005553551 | -0,064270532 | 0,948754817 | 0,999477707 | YES |
| 247 | 2,641872796 | 0,000355719 | 0,005559006 | 0,063989703 | 0,948978426 | 0,999477707 | YES |
| 248 | 1,714775994 | 0,000354501 | 0,005553283 | 0,063836233 | 0,949100627 | 0,999477707 | YES |
| 249 | 5,716731909 | 0,000352994 | 0,005558486 | 0,063505362 | 0,94936409 | 0,999477707 | YES |
| 250 | 14,23384685 | 0,000343408 | 0,005553721 | 0,061833863 | 0,950695136 | 0,999477707 | YES |
| 251 | 8,025712213 | 0,00034153 | 0,005551544 | 0,061519815 | 0,950945234 | 0,999477707 | YES |
| 252 | 10,4207477 | 0,0003351 | 0,005549661 | 0,060381987 | 0,951851405 | 0,999477707 | YES |
| 253 | 2,397194588 | 0,000334772 | 0,00555507 | 0,060264199 | 0,951945215 | 0,999477707 | YES |
| 254 | 0,977468038 | 0,000334508 | 0,005557555 | 0,060189768 | 0,952004495 | 0,999477707 | YES |
| 255 | 2,717937618 | 0,000327147 | 0,005553662 | 0,058906569 | 0,953026525 | 0,999477707 | YES |
| 256 | 19,73124026 | 0,000315207 | 0,005556479 | 0,056727808 | 0,954762022 | 0,999477707 | YES |
| 257 | 38,10329659 | -6,16E-05 | 0,001097109 | -0,056135033 | 0,955234236 | 0,999477707 | YES |
| 258 | 1,34026348 | 0,00031021 | 0,005554577 | 0,055847656 | 0,95546317 | 0,999477707 | YES |
| 259 | 10,35634002 | 0,00030969 | 0,00555628 | 0,055736877 | 0,955551421 | 0,999477707 | YES |
| 260 | 25,74168808 | 0,000308518 | 0,005555014 | 0,055538699 | 0,9557093 | 0,999477707 | YES |
| 261 | 381,6499484 | -0,000131371 | 0,002382249 | -0,055145795 | 0,956022313 | 0,999477707 | YES |
| 262 | 70,30307183 | -0,000300051 | 0,00546298 | -0,054924387 | 0,956198703 | 0,999477707 | YES |
| 263 | 16,90005335 | -0,000299761 | 0,005548185 | -0,054028645 | 0,956912342 | 0,999477707 | YES |

| Medication | Ticker | ASVnr | taxLevel | Phylum | Family | Genus | Species |
|---|---|---|---|---|---|---|---|
| LEDD | 264 | ASV618 | ASV | Actinobacteriota | Atopobiaceae | NA | NA |
| LEDD | 265 | ASV538 | ASV | Bacteroidota | Marinifilaceae | Butyricimonas | NA |
| LEDD | 266 | ASV161 | ASV | Firmicutes | Oscillospiraceae | UCG-002 | NA |
| LEDD | 267 | ASV43 | ASV | Firmicutes | Lachnospiraceae | Blautia | faecis |
| LEDD | 268 | ASV709 | ASV | Firmicutes | Lachnospiraceae | NA | NA |
| LEDD | 269 | ASV231 | ASV | Proteobacteria | NA | NA | NA |
| LEDD | 270 | ASV193 | ASV | Actinobacteriota | Eggerthellaceae | Adlercreutzia | equolifaciens |
| LEDD | 271 | ASV505 | ASV | Bacteroidota | Bacteroidaceae | Bacteroides | nordii |
| LEDD | 272 | ASV836 | ASV | Verrucomicrobiota | Victivallaceae | Victivallis | vadensis |
| LEDD | 273 | ASV59 | ASV | Firmicutes | [Eubacterium] coprostanoligenes group | NA | NA |
| LEDD | 274 | ASV292 | ASV | Firmicutes | [Eubacterium] coprostanoligenes group | NA | NA |
| LEDD | 275 | ASV606 | ASV | Firmicutes | Lachnospiraceae | NA | NA |
| LEDD | 276 | ASV367 | ASV | Bacteroidota | Rikenellaceae | Alistipes | NA |
| LEDD | 277 | ASV151 | ASV | Firmicutes | Butyricicoccaceae | Butyricicoccus | NA |
| LEDD | 278 | ASV582 | ASV | Bacteroidota | NA | NA | NA |
| LEDD | 279 | ASV441 | ASV | Firmicutes | [Eubacterium] coprostanoligenes group | NA | NA |
| LEDD | 280 | ASV354 | ASV | Firmicutes | Ruminococcaceae | Negativibacillus | NA |
| LEDD | 281 | ASV97 | ASV | Firmicutes | Lachnospiraceae | [Ruminococcus] gnavus group | NA |
| LEDD | 282 | ASV546 | ASV | Firmicutes | Lachnospiraceae | NA | NA |
| LEDD | 283 | ASV324 | ASV | Proteobacteria | NA | NA | NA |
| LEDD | 284 | ASV337 | ASV | Firmicutes | Lachnospiraceae | Lachnoclostridium | NA |
| LEDD | 285 | ASV223 | ASV | Bacteroidota | Barnesiellaceae | Barnesiella | NA |
| LEDD | 286 | ASV435 | ASV | Firmicutes | Christensenellaceae | Christensenellaceae R-7 group | NA |
| LEDD | 287 | ASV15 | ASV | Firmicutes | Christensenellaceae | Christensenellaceae R-7 group | NA |
| LEDD | 288 | ASV114 | ASV | Bacteroidota | Bacteroidaceae | Bacteroides | coprocola |
| LEDD | 289 | ASV214 | ASV | Firmicutes | Lachnospiraceae | Lachnospiraceae UCG-010 | NA |
| LEDD | 290 | ASV726 | ASV | Verrucomicrobiota | vadinBE97 | NA | NA |
| LEDD | 291 | ASV202 | ASV | Firmicutes | Lachnospiraceae | [Eubacterium] hallii group | NA |
| LEDD | 292 | ASV422 | ASV | Firmicutes | Lachnospiraceae | Lachnospiraceae FCS020 group | bacterium |
| LEDD | 293 | ASV591 | ASV | Firmicutes | UCG-010 | NA | NA |

| Ticker | DESeq2_baseMean | DESeq2_log2-FoldChange | DESeq2_lfcSE | DESeq2_stat | DESeq2_pvalue | DESeq2_padj | DESeq2 converged |
|---|---|---|---|---|---|---|---|
| 264 | 1,405286744 | 0,000297863 | 0,005556937 | 0,053601923 | 0,957252324 | 0,999477707 | YES |
| 265 | 7,872578089 | 0,000294182 | 0,005552312 | 0,052983642 | 0,957744942 | 0,999477707 | YES |
| 266 | 16,25948563 | 0,000293374 | 0,005548943 | 0,05287026 | 0,95783528 | 0,999477707 | YES |
| 267 | 105,7539111 | -0,000118016 | 0,002242079 | -0,052636842 | 0,958021262 | 0,999477707 | YES |
| 268 | 3,728044901 | 0,00029024 | 0,005554421 | 0,052253865 | 0,958326414 | 0,999477707 | YES |
| 269 | 21,90960149 | 0,000287326 | 0,005551948 | 0,051752327 | 0,958726042 | 0,999477707 | YES |
| 270 | 8,784474931 | 0,000188261 | 0,003661871 | 0,051411283 | 0,958997794 | 0,999477707 | YES |
| 271 | 5,398458234 | 0,000279098 | 0,00555975 | 0,050199796 | 0,959963174 | 0,999477707 | YES |
| 272 | 1,209779611 | 0,000273462 | 0,005556816 | 0,049211976 | 0,960750367 | 0,999477707 | YES |
| 273 | 139,6694157 | -0,000198978 | 0,004097635 | -0,04855928 | 0,961270521 | 0,999477707 | YES |
| 274 | 32,88496859 | 0,000264257 | 0,005558043 | 0,047544932 | 0,96207892 | 0,999477707 | YES |
| 275 | 14,15018687 | 0,000262488 | 0,005556783 | 0,04723732 | 0,962324084 | 0,999477707 | YES |
| 276 | 14,02234981 | 0,000261487 | 0,00554872 | 0,047125698 | 0,962413046 | 0,999477707 | YES |
| 277 | 51,64233017 | 7,79E-05 | 0,001655999 | 0,047038709 | 0,962482376 | 0,999477707 | YES |
| 278 | 3,018899993 | 0,000260025 | 0,005558483 | 0,046779825 | 0,962688709 | 0,999477707 | YES |
| 279 | 9,85819851 | -0,000197859 | 0,00424491 | -0,046610968 | 0,962823291 | 0,999477707 | YES |
| 280 | 11,33772624 | 0,000247372 | 0,005549534 | 0,044575287 | 0,964445841 | 0,999477707 | YES |
| 281 | 57,72130721 | 0,000245536 | 0,005556284 | 0,04419067 | 0,964752419 | 0,999477707 | YES |
| 282 | 10,19948611 | 0,000243925 | 0,005553932 | 0,043919347 | 0,964968693 | 0,999477707 | YES |
| 283 | 11,12584357 | 0,000241367 | 0,00555401 | 0,043458179 | 0,965336302 | 0,999477707 | YES |
| 284 | 15,20955995 | -8,18E-05 | 0,001884554 | -0,043394838 | 0,965386793 | 0,999477707 | YES |
| 285 | 36,79977703 | 0,000240631 | 0,005560959 | 0,043271524 | 0,96548509 | 0,999477707 | YES |
| 286 | 10,95557651 | -0,000161134 | 0,003724283 | -0,043265735 | 0,965489705 | 0,999477707 | YES |
| 287 | 500,2723708 | -0,000116101 | 0,002684555 | -0,043247933 | 0,965503896 | 0,999477707 | YES |
| 288 | 56,2438255 | -0,000233942 | 0,005552037 | -0,04213625 | 0,966390083 | 0,999477707 | YES |
| 289 | 37,53411887 | -0,000169407 | 0,004124504 | -0,041073322 | 0,967237442 | 0,999477707 | YES |
| 290 | 1,504875068 | 0,000218416 | 0,005553365 | 0,039330405 | 0,968626966 | 0,999477707 | YES |
| 291 | 14,42897349 | 0,000216759 | 0,0055505 | 0,039052231 | 0,968848746 | 0,999477707 | YES |
| 292 | 4,351166029 | 0,000215421 | 0,005559339 | 0,038749353 | 0,969090225 | 0,999477707 | YES |
| 293 | 6,344996583 | 0,000213908 | 0,005557079 | 0,03849289 | 0,969294701 | 0,999477707 | YES |

| Medication | Ticker | ASVnr | taxLevel | Phylum | Family | Genus | Species |
|---|---|---|---|---|---|---|---|
| LEDD | 294 | ASV67 | ASV | Firmicutes | Erysipelotrichaceae | Holdemanella | biformis |
| LEDD | 295 | ASV360 | ASV | Firmicutes | [Eubacterium] coprostanoligenes group | NA | NA |
| LEDD | 296 | ASV347 | ASV | Bacteroidota | Tannerellaceae | Parabacteroides | NA |
| LEDD | 297 | ASV87 | ASV | Proteobacteria | Sutterellaceae | Sutterella | wadsworthensis |
| LEDD | 298 | ASV44 | ASV | Bacteroidota | Bacteroidaceae | Bacteroides | NA |
| LEDD | 299 | ASV156 | ASV | Bacteroidota | Prevotellaceae | Prevotella | NA |
| LEDD | 300 | ASV481 | ASV | Firmicutes | Lachnospiraceae | [Eubacterium] xylanophilum group | NA |
| LEDD | 301 | ASV124 | ASV | Firmicutes | NA | NA | NA |
| LEDD | 302 | ASV752 | ASV | Verrucomicrobiota | Puniceicoccaceae | NA | NA |
| LEDD | 303 | ASV200 | ASV | Firmicutes | Lachnospiraceae | [Eubacterium] xylanophilum group | NA |
| LEDD | 304 | ASV135 | ASV | Bacteroidota | Rikenellaceae | Alistipes | inops |
| LEDD | 305 | ASV174 | ASV | Actinobacteriota | Coriobacteriales Incertae Sedis | NA | NA |
| LEDD | 306 | ASV690 | ASV | Firmicutes | Ruminococcaceae | Fournierella | NA |
| LEDD | 307 | ASV480 | ASV | Firmicutes | UCG-010 | NA | NA |
| LEDD | 308 | ASV520 | ASV | Firmicutes | UCG-010 | NA | NA |
| LEDD | 309 | ASV58 | ASV | Bacteroidota | Bacteroidaceae | Bacteroides | NA |
| LEDD | 310 | ASV433 | ASV | Firmicutes | UCG-010 | NA | NA |
| LEDD | 311 | ASV288 | ASV | Firmicutes | Erysipelotrichaceae | NA | NA |
| LEDD | 312 | ASV252 | ASV | Firmicutes | Erysipelotrichaceae | Solobacterium | NA |
| LEDD | 313 | ASV329 | ASV | Firmicutes | Lachnospiraceae | NA | NA |
| LEDD | 314 | ASV83 | ASV | Firmicutes | Lachnospiraceae | Lachnospiraceae NK4A136 group | NA |
| LEDD | 315 | ASV62 | ASV | Firmicutes | NA | NA | NA |
| LEDD | 316 | ASV772 | ASV | Firmicutes | Ruminococcaceae | Fournierella | NA |
| LEDD | 317 | ASV341 | ASV | Firmicutes | Christensenellaceae | Christensenellaceae R-7 group | NA |
| LEDD | 318 | ASV27 | ASV | Firmicutes | Ruminococcaceae | CAG-352 | NA |
| LEDD | 319 | ASV675 | ASV | Bacteroidota | Prevotellaceae | NA | NA |
| LEDD | 320 | ASV38 | ASV | Firmicutes | Clostridiaceae | Clostridium sensu stricto 1 | NA |
| LEDD | 321 | ASV250 | ASV | Bacteroidota | Prevotellaceae | Paraprevotella | NA |
| LEDD | 322 | ASV666 | ASV | Firmicutes | Erysipelotrichaceae | Holdemania | filiformis |
| LEDD | 323 | ASV198 | ASV | Bacteroidota | Bacteroidaceae | Bacteroides | eggerthii |

| Ticker | DESeq2_baseMean | DESeq2_log2-FoldChange | DESeq2_lfcSE | DESeq2_stat | DESeq2_pvalue | DESeq2_padj | DESeq2 converged |
|---|---|---|---|---|---|---|---|
| 294 | 47,15928254 | -0,000213615 | 0,005557185 | -0,03843943 | 0,969337324 | 0,999477707 | YES |
| 295 | 16,07726368 | 0,000213163 | 0,005559819 | 0,038339975 | 0,969416619 | 0,999477707 | YES |
| 296 | 27,45099236 | 0,000212158 | 0,005559971 | 0,038158166 | 0,969561575 | 0,999477707 | YES |
| 297 | 36,31469707 | 0,000130944 | 0,003436894 | 0,038099547 | 0,969608312 | 0,999477707 | YES |
| 298 | 60,78089087 | -0,000210658 | 0,005548394 | -0,037967429 | 0,969713651 | 0,999477707 | YES |
| 299 | 18,45072649 | 0,000210695 | 0,005553354 | 0,037940211 | 0,969735353 | 0,999477707 | YES |
| 300 | 8,336489924 | -0,000210074 | 0,005557763 | -0,037798302 | 0,969848498 | 0,999477707 | YES |
| 301 | 48,88295812 | 0,000205795 | 0,00555279 | 0,037061577 | 0,970435908 | 0,999477707 | YES |
| 302 | 3,162923681 | 0,000203966 | 0,005558197 | 0,036696433 | 0,970727053 | 0,999477707 | YES |
| 303 | 29,3648638 | 8,01E-05 | 0,002209511 | 0,036235079 | 0,971094916 | 0,999477707 | YES |
| 304 | 66,61369828 | 0,000128127 | 0,003611079 | 0,035481566 | 0,971695745 | 0,999477707 | YES |
| 305 | 2,991697807 | 0,000196272 | 0,005556755 | 0,035321333 | 0,971823512 | 0,999477707 | YES |
| 306 | 7,105330285 | -0,000193652 | 0,00555723 | -0,034846871 | 0,972201846 | 0,999477707 | YES |
| 307 | 7,992442659 | 0,000191125 | 0,005553132 | 0,03441748 | 0,972544245 | 0,999477707 | YES |
| 308 | 5,640723253 | 0,000189463 | 0,005557337 | 0,034092383 | 0,972803482 | 0,999477707 | YES |
| 309 | 142,3397159 | 0,00011412 | 0,003351738 | 0,034048031 | 0,972838849 | 0,999477707 | YES |
| 310 | 12,92001934 | 0,000185436 | 0,005551084 | 0,03340542 | 0,973351287 | 0,999477707 | YES |
| 311 | 14,75712244 | 0,000167758 | 0,005059865 | 0,033154577 | 0,97355132 | 0,999477707 | YES |
| 312 | 15,98831435 | 0,000182511 | 0,005557643 | 0,032839554 | 0,973802535 | 0,999477707 | YES |
| 313 | 5,065534029 | -0,000174226 | 0,005552953 | -0,031375333 | 0,974970213 | 0,999477707 | YES |
| 314 | 55,95176432 | -0,000154136 | 0,004992675 | -0,030872348 | 0,975371343 | 0,999477707 | YES |
| 315 | 168,985979 | -0,00016043 | 0,005228486 | -0,03068391 | 0,975521623 | 0,999477707 | YES |
| 316 | 6,217452364 | -0,000169347 | 0,005554705 | -0,030487074 | 0,975678602 | 0,999477707 | YES |
| 317 | 8,747838125 | 0,000166111 | 0,005549226 | 0,029934155 | 0,976119566 | 0,999477707 | YES |
| 318 | 301,6970288 | -0,000165204 | 0,005546676 | -0,029784301 | 0,976239079 | 0,999477707 | YES |
| 319 | 15,53629147 | 0,000165008 | 0,00555303 | 0,029714872 | 0,976294451 | 0,999477707 | YES |
| 320 | 8,627364798 | 0,000164693 | 0,005552721 | 0,02965981 | 0,976338364 | 0,999477707 | YES |
| 321 | 26,68983111 | 0,000163915 | 0,005555825 | 0,029503322 | 0,97646317 | 0,999477707 | YES |
| 322 | 1,719887309 | 0,000162125 | 0,005550085 | 0,029211244 | 0,976696114 | 0,999477707 | YES |
| 323 | 96,40091483 | 0,000161623 | 0,005558758 | 0,029075445 | 0,97680442 | 0,999477707 | YES |

| Medication | Ticker | ASVnr | taxLevel | Phylum | Family | Genus | Species |
|---|---|---|---|---|---|---|---|
| LEDD | 324 | ASV42 | ASV | Actinobacteriota | Bifidobacteriaceae | Bifidobacterium | NA |
| LEDD | 325 | ASV272 | ASV | Firmicutes | Erysipelotrichaceae | Catenisphaera | NA |
| LEDD | 326 | ASV894 | ASV | Desulfobacterota | Desulfovibrionaceae | NA | NA |
| LEDD | 327 | ASV597 | ASV | Firmicutes | Oscillospiraceae | Oscillibacter | NA |
| LEDD | 328 | ASV84 | ASV | Bacteroidota | Bacteroidaceae | Bacteroides | NA |
| LEDD | 329 | ASV146 | ASV | Firmicutes | Erysipelotrichaceae | Turicibacter | sanguinis |
| LEDD | 330 | ASV437 | ASV | Firmicutes | Christensenellaceae | Christensenellaceae R-7 group | NA |
| LEDD | 331 | ASV421 | ASV | Firmicutes | NA | NA | NA |
| LEDD | 332 | ASV611 | ASV | Firmicutes | Lachnospiraceae | Lachnospiraceae UCG-010 | NA |
| LEDD | 333 | ASV13 | ASV | Firmicutes | Lachnospiraceae | Fusicatenibacter | saccharivorans |
| LEDD | 334 | ASV819 | ASV | Firmicutes | Oscillospiraceae | Colidextribacter | NA |
| LEDD | 335 | ASV189 | ASV | Firmicutes | Ruminococcaceae | Ruminococcus | champanellensis |
| LEDD | 336 | ASV385 | ASV | Firmicutes | Selenomonadaceae | Mitsuokella | jalaludinii |
| LEDD | 337 | ASV503 | ASV | Firmicutes | Erysipelatoclostridiaceae | Erysipelatoclostridium | NA |
| LEDD | 338 | ASV326 | ASV | Proteobacteria | NA | NA | NA |
| LEDD | 339 | ASV573 | ASV | Firmicutes | Erysipelatoclostridiaceae | Erysipelatoclostridium | NA |
| LEDD | 340 | ASV208 | ASV | Bacteroidota | Prevotellaceae | Prevotella | NA |
| LEDD | 341 | ASV615 | ASV | Firmicutes | Oscillospiraceae | Oscillibacter | NA |
| LEDD | 342 | ASV382 | ASV | Firmicutes | NA | NA | NA |
| LEDD | 343 | ASV164 | ASV | Bacteroidota | Tannerellaceae | Parabacteroides | distasonis |
| LEDD | 344 | ASV432 | ASV | Bacteroidota | Marinifilaceae | Butyricimonas | virosa |
| LEDD | 345 | ASV306 | ASV | Firmicutes | Veillonellaceae | Veillonella | NA |
| LEDD | 346 | ASV253 | ASV | Bacteroidota | Tannerellaceae | Parabacteroides | distasonis |
| LEDD | 347 | ASV196 | ASV | Firmicutes | Ruminococcaceae | Ruminococcus | NA |
| LEDD | 348 | ASV714 | ASV | Firmicutes | Oscillospiraceae | Oscillibacter | NA |
| LEDD | 349 | ASV335 | ASV | Bacteroidota | Barnesiellaceae | NA | NA |
| LEDD | 350 | ASV72 | ASV | Proteobacteria | Sutterellaceae | Parasutterella | excrementihominis |
| LEDD | 351 | ASV140 | ASV | Bacteroidota | Tannerellaceae | Parabacteroides | distasonis |
| LEDD | 352 | ASV373 | ASV | Firmicutes | NA | NA | NA |
| LEDD | 353 | ASV118 | ASV | Firmicutes | NA | NA | NA |

| Ticker | DESeq2_baseMean | DESeq2_log2-FoldChange | DESeq2_lfcSE | DESeq2_stat | DESeq2_pvalue | DESeq2_padj | DESeq2 converged |
|---|---|---|---|---|---|---|---|
| 324 | 46,7716175 | 0,000161564 | 0,005560285 | 0,02905677 | 0,976819314 | 0,999477707 | YES |
| 325 | 3,549088477 | 0,000160996 | 0,005558122 | 0,02896592 | 0,976891771 | 0,999477707 | YES |
| 326 | 3,275879807 | 0,000158508 | 0,005558781 | 0,02851485 | 0,977251524 | 0,999477707 | YES |
| 327 | 8,485709209 | -0,000158242 | 0,005556418 | -0,028479173 | 0,977279979 | 0,999477707 | YES |
| 328 | 42,67470703 | -0,000157552 | 0,005549273 | -0,028391459 | 0,977349936 | 0,999477707 | YES |
| 329 | 9,696741845 | 0,000156421 | 0,005553142 | 0,028168047 | 0,977528122 | 0,999477707 | YES |
| 330 | 4,824076391 | 0,000153444 | 0,005555635 | 0,02761951 | 0,977965621 | 0,999477707 | YES |
| 331 | 19,91706631 | -0,00015323 | 0,005554988 | -0,027584294 | 0,977993708 | 0,999477707 | YES |
| 332 | 4,081463827 | 0,000148878 | 0,005561463 | 0,026769526 | 0,978643559 | 0,999477707 | YES |
| 333 | 784,3099195 | 5,74E-05 | 0,002149652 | 0,026700189 | 0,978698862 | 0,999477707 | YES |
| 334 | 2,421772883 | 0,000146536 | 0,005557499 | 0,026367327 | 0,978964355 | 0,999477707 | YES |
| 335 | 76,71281636 | -0,000145148 | 0,00556133 | -0,026099553 | 0,979177934 | 0,999477707 | YES |
| 336 | 18,66242672 | 0,000141955 | 0,005557055 | 0,02554495 | 0,979620295 | 0,999477707 | YES |
| 337 | 3,609034086 | -0,000140996 | 0,005559473 | -0,025361403 | 0,979766697 | 0,999477707 | YES |
| 338 | 11,49770266 | -0,000131832 | 0,005551992 | -0,023744997 | 0,981056014 | 0,999477707 | YES |
| 339 | 3,091871824 | 0,000131793 | 0,005559041 | 0,02370792 | 0,981085589 | 0,999477707 | YES |
| 340 | 16,73438973 | 0,000129775 | 0,005558518 | 0,023347119 | 0,981373387 | 0,999477707 | YES |
| 341 | 1,09131054 | 0,000128858 | 0,005551605 | 0,023210863 | 0,981482073 | 0,999477707 | YES |
| 342 | 15,08492104 | 0,000128715 | 0,005557337 | 0,023161298 | 0,98152161 | 0,999477707 | YES |
| 343 | 21,94634572 | -0,000127973 | 0,005559028 | -0,023020809 | 0,981633674 | 0,999477707 | YES |
| 344 | 8,260028247 | 0,000126501 | 0,005550819 | 0,022789632 | 0,981818078 | 0,999477707 | YES |
| 345 | 18,53531193 | -0,000125557 | 0,005551321 | -0,022617487 | 0,981955395 | 0,999477707 | YES |
| 346 | 24,33668341 | -0,000124149 | 0,00555805 | -0,022336732 | 0,982179348 | 0,999477707 | YES |
| 347 | 45,20814381 | 0,000120919 | 0,005556761 | 0,021760717 | 0,98263883 | 0,999477707 | YES |
| 348 | 7,440181251 | 0,000118436 | 0,005553535 | 0,021326244 | 0,982985409 | 0,999477707 | YES |
| 349 | 24,57831886 | 6,74E-05 | 0,003189692 | 0,021141226 | 0,983132998 | 0,999477707 | YES |
| 350 | 51,05649708 | -3,00E-05 | 0,00144442 | -0,020802139 | 0,983403492 | 0,999477707 | YES |
| 351 | 52,28557923 | 9,72E-05 | 0,004725311 | 0,020576158 | 0,983583759 | 0,999477707 | YES |
| 352 | 16,80301954 | 0,000112367 | 0,005555994 | 0,020224377 | 0,983864382 | 0,999477707 | YES |
| 353 | 302,3165998 | 0,000110614 | 0,005554919 | 0,019912851 | 0,984112894 | 0,999477707 | YES |

| Medication | Ticker | ASVnr | taxLevel | Phylum | Family | Genus | Species |
|---|---|---|---|---|---|---|---|
| LEDD | 354 | ASV895 | ASV | Bacteroidota | Barnesiellaceae | Coprobacter | secundus |
| LEDD | 355 | ASV169 | ASV | Bacteroidota | Barnesiellaceae | Barnesiella | NA |
| LEDD | 356 | ASV241 | ASV | Firmicutes | [Eubacterium] coprostanoligenes group | NA | NA |
| LEDD | 357 | ASV119 | ASV | Firmicutes | Ruminococcaceae | Subdoligranulum | NA |
| LEDD | 358 | ASV448 | ASV | Firmicutes | Christensenellaceae | Christensenellaceae R-7 group | NA |
| LEDD | 359 | ASV258 | ASV | Firmicutes | Lachnospiraceae | Lachnospiraceae AC2044 group | NA |
| LEDD | 360 | ASV400 | ASV | Bacteroidota | Rikenellaceae | Rikenellaceae RC9 gut group | NA |
| LEDD | 361 | ASV175 | ASV | Firmicutes | NA | NA | NA |
| LEDD | 362 | ASV179 | ASV | Firmicutes | NA | NA | NA |
| LEDD | 363 | ASV232 | ASV | Firmicutes | Acholeplasmataceae | Anaeroplasma | NA |
| LEDD | 364 | ASV248 | ASV | Firmicutes | Ruminococcaceae | Subdoligranulum | NA |
| LEDD | 365 | ASV449 | ASV | Actinobacteriota | Atopobiaceae | Olsenella | NA |
| LEDD | 366 | ASV209 | ASV | Bacteroidota | Tannerellaceae | NA | NA |
| LEDD | 367 | ASV665 | ASV | Firmicutes | Lachnospiraceae | NA | NA |
| LEDD | 368 | ASV534 | ASV | Firmicutes | Anaerovoracaceae | Family XIII UCG-001 | NA |
| LEDD | 369 | ASV186 | ASV | Firmicutes | Lachnospiraceae | [Ruminococcus] torques group | NA |
| LEDD | 370 | ASV423 | ASV | Firmicutes | Christensenellaceae | Christensenellaceae R-7 group | NA |
| LEDD | 371 | ASV79 | ASV | Firmicutes | Lachnospiraceae | [Eubacterium] ruminantium group | NA |
| LEDD | 372 | ASV521 | ASV | Firmicutes | Lachnospiraceae | [Eubacterium] ventriosum group | NA |
| LEDD | 373 | ASV656 | ASV | Firmicutes | Oscillospiraceae | Flavonifractor | NA |
| LEDD | 374 | ASV1030 | ASV | Firmicutes | NA | NA | NA |
| LEDD | 375 | ASV530 | ASV | Firmicutes | Veillonellaceae | Allisonella | histaminiformans |
| LEDD | 376 | ASV507 | ASV | Firmicutes | Oscillospiraceae | Colidextribacter | NA |
| LEDD | 377 | ASV885 | ASV | Firmicutes | Lachnospiraceae | Shuttleworthia | NA |
| LEDD | 378 | ASV103 | ASV | Firmicutes | NA | NA | NA |
| LEDD | 379 | ASV446 | ASV | Firmicutes | Oscillospiraceae | UCG-005 | NA |
| LEDD | 380 | ASV608 | ASV | Firmicutes | Lachnospiraceae | NA | NA |
| LEDD | 381 | ASV284 | ASV | Firmicutes | Oscillospiraceae | NK4A214 group | NA |
| LEDD | 382 | ASV121 | ASV | Bacteroidota | Barnesiellaceae | Barnesiella | NA |
| LEDD | 383 | ASV205 | ASV | Firmicutes | NA | NA | NA |

| Ticker | DESeq2_baseMean | DESeq2_log2-FoldChange | DESeq2_lfcSE | DESeq2_stat | DESeq2_pvalue | DESeq2_padj | DESeq2 converged |
|---|---|---|---|---|---|---|---|
| 354 | 3,657989246 | 0,000109005 | 0,005552833 | 0,019630504 | 0,98433813 | 0,999477707 | YES |
| 355 | 41,58625889 | -0,000106036 | 0,005558636 | -0,019075921 | 0,98478054 | 0,999477707 | YES |
| 356 | 7,762635101 | -0,000105706 | 0,005557112 | -0,019021752 | 0,984823753 | 0,999477707 | YES |
| 357 | 52,62298781 | -6,22E-05 | 0,003278895 | -0,018973727 | 0,984862065 | 0,999477707 | YES |
| 358 | 14,5808246 | 9,75E-05 | 0,005555134 | 0,01754712 | 9,86E-01 | 0,999477707 | YES |
| 359 | 55,85242367 | 6,27E-05 | 0,003577496 | 0,017518741 | 0,986022782 | 0,999477707 | YES |
| 360 | 24,46505356 | 9,68E-05 | 0,005549569 | 0,017444006 | 0,986082403 | 0,999477707 | YES |
| 361 | 21,55097999 | 9,62E-05 | 0,005545372 | 0,017354226 | 0,986154026 | 0,999477707 | YES |
| 362 | 29,63800594 | 9,62E-05 | 0,00555083 | 0,017337661 | 0,986167241 | 0,999477707 | YES |
| 363 | 32,48546035 | -9,38E-05 | 0,00555924 | -0,016871612 | 9,87E-01 | 9,99E-01 | YES |
| 364 | 13,22637554 | -9,27E-05 | 0,005550242 | -0,016699069 | 0,98667669 | 0,999477707 | YES |
| 365 | 4,349924288 | -8,81E-05 | 0,005552085 | -0,015868769 | 0,987339085 | 0,999477707 | YES |
| 366 | 55,76378301 | -7,36E-05 | 0,004770325 | -0,01543295 | 0,987686776 | 0,999477707 | YES |
| 367 | 10,20531785 | 8,49E-05 | 0,005553239 | 0,015292481 | 0,987798841 | 0,999477707 | YES |
| 368 | 5,816478612 | 8,42E-05 | 0,005559072 | 0,015147791 | 0,987914274 | 0,999477707 | YES |
| 369 | 26,05244729 | -8,41E-05 | 0,005558133 | -0,015126295 | 0,987931423 | 0,999477707 | YES |
| 370 | 7,312103265 | 6,79E-05 | 0,004771187 | 0,014228458 | 0,988647716 | 0,999477707 | YES |
| 371 | 148,127431 | -7,02E-05 | 0,004962116 | -0,014140237 | 0,988718099 | 0,999477707 | YES |
| 372 | 4,140582892 | -7,83E-05 | 0,005559598 | -0,0140908 | 0,98875754 | 0,999477707 | YES |
| 373 | 1,249713349 | -7,78E-05 | 0,005559106 | -0,01398738 | 0,988840049 | 0,999477707 | YES |
| 374 | 0,982444278 | 7,38E-05 | 0,005554872 | 0,013283404 | 0,989401689 | 0,999477707 | YES |
| 375 | 9,225730262 | -7,31E-05 | 0,005553854 | -0,013161495 | 0,98949895 | 0,999477707 | YES |
| 376 | 12,01855857 | 7,28E-05 | 0,005550845 | 0,013116117 | 0,989535153 | 0,999477707 | YES |
| 377 | 2,990038708 | -7,17E-05 | 0,005559486 | -0,012892382 | 0,989713653 | 0,999477707 | YES |
| 378 | 250,8038323 | -7,05E-05 | 0,005550538 | -0,01270147 | 0,989865966 | 0,999477707 | YES |
| 379 | 14,02468503 | -6,97E-05 | 0,005553342 | -0,012543732 | 0,989991812 | 0,999477707 | YES |
| 380 | 6,128535647 | 6,87E-05 | 0,005552386 | 0,012366542 | 0,990133178 | 0,999477707 | YES |
| 381 | 6,375368608 | -6,68E-05 | 0,005553553 | -0,01203476 | 0,990397882 | 0,999477707 | YES |
| 382 | 41,60087519 | 6,61E-05 | 0,005555185 | 0,011891997 | 0,990511783 | 0,999477707 | YES |
| 383 | 47,05692284 | -6,43E-05 | 0,005554793 | -0,011580329 | 0,990760441 | 0,999477707 | YES |

| Medication | Ticker | ASVnr | taxLevel | Phylum | Family | Genus | Species |
|---|---|---|---|---|---|---|---|
| LEDD | 384 | ASV1086 | ASV | Firmicutes | UCG-010 | NA | NA |
| LEDD | 385 | ASV322 | ASV | Firmicutes | Anaerovoracaceae | Family XIII AD3011 group | NA |
| LEDD | 386 | ASV753 | ASV | Firmicutes | Ruminococcaceae | NA | NA |
| LEDD | 387 | ASV176 | ASV | Bacteroidota | Prevotellaceae | Paraprevotella | NA |
| LEDD | 388 | ASV453 | ASV | Verrucomicrobiota | Victivallaceae | Victivallis | NA |
| LEDD | 389 | ASV233 | ASV | Firmicutes | [Eubacterium] coprostanoligenes group | NA | NA |
| LEDD | 390 | ASV806 | ASV | Firmicutes | Peptococcaceae | NA | NA |
| LEDD | 391 | ASV1155 | ASV | Firmicutes | UCG-010 | NA | NA |
| LEDD | 392 | ASV620 | ASV | Firmicutes | UCG-010 | NA | NA |
| LEDD | 393 | ASV545 | ASV | Proteobacteria | Sutterellaceae | Sutterella | NA |
| LEDD | 394 | ASV60 | ASV | Firmicutes | Ruminococcaceae | [Eubacterium] siraeum group | NA |
| LEDD | 395 | ASV80 | ASV | Firmicutes | Acidaminococcaceae | Phascolarctobacterium | faecium |
| LEDD | 396 | ASV343 | ASV | Firmicutes | Lachnospiraceae | NA | NA |
| LEDD | 397 | ASV323 | ASV | Desulfobacterota | Desulfovibrionaceae | Desulfovibrio | piger |
| LEDD | 398 | ASV10 | ASV | Verrucomicrobiota | Akkermansiaceae | Akkermansia | muciniphila |
| LEDD | 399 | ASV376 | ASV | Bacteroidota | Barnesiellaceae | Coprobacter | fastidiosus |
| LEDD | 400 | ASV229 | ASV | Firmicutes | Oscillospiraceae | Flavonifractor | plautii |
| LEDD | 401 | ASV314 | ASV | Firmicutes | Ruminococcaceae | Subdoligranulum | NA |
| LEDD | 402 | ASV759 | ASV | Firmicutes | NA | NA | NA |
| LEDD | 403 | ASV460 | ASV | Firmicutes | NA | NA | NA |
| LEDD | 404 | ASV504 | ASV | Firmicutes | Christensenellaceae | Christensenellaceae R-7 group | NA |
| LEDD | 405 | ASV277 | ASV | Bacteroidota | Rikenellaceae | Alistipes | ihumii |
| LEDD | 406 | ASV353 | ASV | Firmicutes | Lachnospiraceae | [Eubacterium] xylanophilum group | NA |
| LEDD | 407 | ASV69 | ASV | Firmicutes | Christensenellaceae | NA | NA |
| LEDD | 408 | ASV240 | ASV | Bacteroidota | Prevotellaceae | Alloprevotella | NA |
| LEDD | 409 | ASV107 | ASV | Bacteroidota | Bacteroidaceae | Bacteroides | eggerthii |
| LEDD | 410 | ASV260 | ASV | Firmicutes | Lactobacillaceae | Lactobacillus | delbrueckii |
| LEDD | 411 | ASV499 | ASV | Firmicutes | Lachnospiraceae | Eisenbergiella | tayi |
| LEDD | 412 | ASV76 | Genus | Firmicutes | Lachnospiraceae | Lachnospiraceae ND3007 group | NA |
| LEDD | 413 | ASV7 | Genus | Firmicutes | Lachnospiraceae | Agathobacter | NA |

| Ticker | DESeq2_baseMean | DESeq2_log2-FoldChange | DESeq2_lfcSE | DESeq2_stat | DESeq2_pvalue | DESeq2_padj | DESeq2 converged |
|---|---|---|---|---|---|---|---|
| 384 | 1,129599922 | 6,28E-05 | 0,005558253 | 0,011290281 | 0,990991851 | 0,999477707 | YES |
| 385 | 27,13864507 | -1,25E-05 | 0,001146336 | -0,010930019 | 0,99127928 | 0,999477707 | YES |
| 386 | 5,313173559 | -5,79E-05 | 0,005556564 | -0,010422823 | 0,991683941 | 0,999477707 | YES |
| 387 | 37,41244117 | 5,73E-05 | 0,005550317 | 0,010317068 | 0,991768317 | 0,999477707 | YES |
| 388 | 5,186345754 | -4,69E-05 | 0,005548061 | -0,008453704 | 0,993255 | 0,999477707 | YES |
| 389 | 14,90640636 | 4,55E-05 | 0,005556832 | 0,008180882 | 0,993472673 | 0,999477707 | YES |
| 390 | 2,729887651 | 4,27E-05 | 0,005553738 | 0,007692414 | 0,993862402 | 0,999477707 | YES |
| 391 | 2,665945516 | -4,14E-05 | 0,005556945 | -0,007445336 | 0,994059536 | 0,999477707 | YES |
| 392 | 16,09675875 | 4,10E-05 | 0,005550265 | 0,007384626 | 0,994107975 | 0,999477707 | YES |
| 393 | 14,15500388 | 3,83E-05 | 0,005555083 | 0,006890449 | 0,994502261 | 0,999477707 | YES |
| 394 | 157,1480003 | -1,41E-05 | 0,002515239 | -0,005605431 | 0,995527536 | 0,999477707 | YES |
| 395 | 70,47815475 | -2,99E-05 | 0,005558424 | -0,005386527 | 0,995702194 | 0,999477707 | YES |
| 396 | 8,457088352 | 2,87E-05 | 0,005554961 | 0,005169616 | 0,995875262 | 0,999477707 | YES |
| 397 | 35,80719401 | -2,53E-05 | 0,005560244 | -0,004547562 | 0,996371583 | 0,999477707 | YES |
| 398 | 912,9538844 | 1,34E-05 | 0,003029327 | 0,004413034 | 0,99647892 | 0,999477707 | YES |
| 399 | 8,118634009 | -1,47E-05 | 0,00336054 | -0,004365745 | 0,99651665 | 0,999477707 | YES |
| 400 | 12,45080414 | -2,38E-05 | 0,005541543 | -0,004303277 | 0,996566492 | 0,999477707 | YES |
| 401 | 23,41177155 | -2,24E-05 | 0,005548859 | -0,0040292 | 0,996785172 | 0,999477707 | YES |
| 402 | 7,339116708 | 1,85E-05 | 0,005557796 | 0,003329434 | 0,997343501 | 0,999477707 | YES |
| 403 | 4,244930043 | 1,83E-05 | 0,005561114 | 0,003282742 | 0,997380755 | 0,999477707 | YES |
| 404 | 2,9824968 | 1,42E-05 | 0,005556259 | 0,002553736 | 0,997962416 | 0,999477707 | YES |
| 405 | 12,88364389 | -1,18E-05 | 0,005547998 | -0,002128393 | 0,99830179 | 0,999477707 | YES |
| 406 | 13,70410935 | 1,03E-05 | 0,005540358 | 0,00185634 | 0,998518856 | 0,999477707 | YES |
| 407 | 84,0393567 | -4,88E-06 | 0,002675397 | -0,001824226 | 0,998544479 | 0,999477707 | YES |
| 408 | 104,5073726 | 9,23E-06 | 0,005552413 | 0,001662122 | 0,998673819 | 0,999477707 | YES |
| 409 | 48,01217935 | -8,44E-06 | 0,005554133 | -0,001518833 | 0,998788147 | 0,999477707 | YES |
| 410 | 14,18298157 | -5,95E-06 | 0,005561012 | -0,001070615 | 0,999145773 | 0,999477707 | YES |
| 411 | 11,47156132 | -3,64E-06 | 0,005557368 | -0,000654597 | 0,999477707 | 0,999477707 | YES |
| 412 | 59,64700043 | -0,002677021 | 0,000664941 | -4,025951258 | 5,67E-05 | 0,007263417 | YES |
| 413 | 1987,419 | -0,001404089 | 0,000449226 | -3,125573648 | 0,001774586 | 0,098376655 | YES |

| Medication | Ticker | ASVnr | taxLevel | Phylum | Family | Genus | Species |
|---|---|---|---|---|---|---|---|
| LEDD | 414 | ASV96 | Genus | Firmicutes | Lachnospiraceae | CAG-56 | NA |
| LEDD | 415 | ASV65 | Genus | Firmicutes | Ruminococcaceae | Incertae Sedis | NA |
| LEDD | 416 | ASV295 | Genus | Firmicutes | Ruminococcaceae | Negativibacillus | NA |
| LEDD | 417 | ASV116 | Genus | Firmicutes | Oscillospiraceae | UCG-003 | NA |
| LEDD | 418 | ASV195 | Genus | Bacteroidota | Barnesiellaceae | Barnesiella | NA |
| LEDD | 419 | ASV144 | Genus | Desulfobacterota | Desulfovibrionaceae | Bilophila | NA |
| LEDD | 420 | ASV18 | Genus | Firmicutes | Lachnospiraceae | Dorea | NA |
| LEDD | 421 | ASV219 | Genus | Firmicutes | Lachnospiraceae | Lachnospiraceae UCG-004 | NA |
| LEDD | 422 | ASV85 | Genus | Firmicutes | Lachnospiraceae | Coprococcus | NA |
| LEDD | 423 | ASV536 | Genus | Bacteroidota | Marinifilaceae | Butyricimonas | NA |
| LEDD | 424 | ASV23 | Genus | Firmicutes | Streptococcaceae | Streptococcus | NA |
| LEDD | 425 | ASV211 | Genus | Firmicutes | Lachnospiraceae | Lachnospiraceae FCS020 group | NA |
| LEDD | 426 | ASV57 | Genus | Firmicutes | Lachnospiraceae | Roseburia | NA |
| LEDD | 427 | ASV17 | Genus | Actinobacteriota | Coriobacteriaceae | Collinsella | NA |
| LEDD | 428 | ASV357 | Genus | Firmicutes | Lachnospiraceae | Marvinbryantia | NA |
| LEDD | 429 | ASV4 | Genus | Firmicutes | Ruminococcaceae | Ruminococcus | NA |
| LEDD | 430 | ASV158 | Genus | Firmicutes | Lachnospiraceae | [Eubacterium] ventriosum group | NA |
| LEDD | 431 | ASV19 | Genus | Firmicutes | Erysipelatoclostridiaceae | Erysipelotrichaceae UCG-003 | NA |
| LEDD | 432 | ASV111 | Genus | Firmicutes | Lachnospiraceae | Lachnospira | NA |
| LEDD | 433 | ASV408 | Genus | Bacteroidota | Prevotellaceae | Paraprevotella | NA |
| LEDD | 434 | ASV226 | Genus | Firmicutes | Lachnospiraceae | Lachnospiraceae UCG-001 | NA |
| LEDD | 435 | ASV5 | Genus | Firmicutes | Lachnospiraceae | Blautia | NA |
| LEDD | 436 | ASV66 | Genus | Firmicutes | Butyricicoccaceae | Butyricicoccus | NA |
| LEDD | 437 | ASV309 | Genus | Firmicutes | Oscillospiraceae | Colidextribacter | NA |
| LEDD | 438 | ASV974 | Genus | Firmicutes | Butyricicoccaceae | UCG-009 | NA |
| LEDD | 439 | ASV512 | Genus | Proteobacteria | Oxalobacteraceae | Oxalobacter | NA |
| LEDD | 440 | ASV630 | Genus | Firmicutes | Ruminococcaceae | Angelakisella | NA |
| LEDD | 441 | ASV386 | Genus | Firmicutes | Ruminococcaceae | DTU089 | NA |
| LEDD | 442 | ASV37 | Genus | Firmicutes | Lachnospiraceae | [Eubacterium] hallii group | NA |
| LEDD | 443 | ASV214 | Genus | Firmicutes | Lachnospiraceae | Lachnospiraceae UCG-010 | NA |

| Ticker | DESeq2_baseMean | DESeq2_log2-FoldChange | DESeq2_lfcSE | DESeq2_stat | DESeq2_pvalue | DESeq2_padj | DESeq2 converged |
|---|---|---|---|---|---|---|---|
| 414 | 119,2328775 | -0,00286731 | 0,000940799 | -3,04773875 | 0,002305703 | 0,098376655 | YES |
| 415 | 120,0579137 | 0,001119054 | 0,000417941 | 2,677540388 | 0,007416491 | 0,237327714 | YES |
| 416 | 45,26405539 | 0,002009625 | 0,000780791 | 2,573831842 | 0,010057914 | 0,257482608 | YES |
| 417 | 149,7231774 | -0,001471954 | 0,000652972 | -2,254236155 | 0,024181315 | 0,515868062 | YES |
| 418 | 246,9722513 | -0,001161587 | 0,000550131 | -2,111472747 | 0,034731698 | 0,635093906 | YES |
| 419 | 64,32240255 | 0,000916827 | 0,000458069 | 2,001506786 | 0,045337803 | 0,656613357 | YES |
| 420 | 678,9890903 | -0,000707719 | 0,00035495 | -1,993852953 | 0,046168127 | 0,656613357 | YES |
| 421 | 44,44180757 | -0,001779335 | 0,000994239 | -1,789644783 | 0,073511034 | 0,905728798 | YES |
| 422 | 880,5389705 | -0,000737624 | 0,000433013 | -1,703470059 | 0,088480136 | 0,905728798 | YES |
| 423 | 49,26473663 | 0,000764803 | 0,000454321 | 1,683396861 | 0,092298291 | 0,905728798 | YES |
| 424 | 236,3988458 | 0,001204007 | 0,000724225 | 1,662475439 | 0,096417484 | 0,905728798 | YES |
| 425 | 41,87861749 | -0,001362856 | 0,00082627 | -1,649407949 | 0,099064087 | 0,905728798 | YES |
| 426 | 535,2124177 | -0,000769498 | 0,000490331 | -1,569343418 | 0,116567941 | 0,994713092 | YES |
| 427 | 233,933023 | -0,000626639 | 0,000467851 | -1,339397566 | 0,180441281 | 0,998407098 | YES |
| 428 | 63,59587409 | 0,001554695 | 0,001186816 | 1,309972211 | 0,190205237 | 0,998407098 | YES |
| 429 | 1802,317879 | -0,000607601 | 0,000487088 | -1,247414606 | 0,212245514 | 0,998407098 | YES |
| 430 | 101,4029537 | -0,000774761 | 0,000641441 | -1,207845022 | 0,22710688 | 0,998407098 | YES |
| 431 | 177,0676523 | -0,000625325 | 0,000530485 | -1,178778454 | 0,238486406 | 0,998407098 | YES |
| 432 | 123,7797096 | -0,002215551 | 0,001897749 | -1,167462449 | 0,243023664 | 0,998407098 | YES |
| 433 | 142,3554429 | -0,000636216 | 0,000598227 | -1,063502443 | 0,287554158 | 0,998407098 | YES |
| 434 | 57,93753735 | -0,001374795 | 0,001304746 | -1,053687983 | 0,292025793 | 0,998407098 | YES |
| 435 | 1358,368645 | -0,000269684 | 0,000268934 | -1,002791787 | 0,315961332 | 0,998407098 | YES |
| 436 | 151,7944076 | -0,000349864 | 0,000371945 | -0,940634893 | 0,346891994 | 0,998407098 | YES |
| 437 | 98,85286937 | -0,000331676 | 0,000370045 | -0,89631202 | 0,370086141 | 0,998407098 | YES |
| 438 | 11,79079998 | 0,002078132 | 0,002348971 | 0,884698827 | 0,376319089 | 0,998407098 | YES |
| 439 | 14,36227103 | 0,000608791 | 0,000711748 | 0,85534701 | 0,39235907 | 0,998407098 | YES |
| 440 | 7,049628943 | 0,002554922 | 0,00300797 | 0,849384105 | 0,395667595 | 0,998407098 | YES |
| 441 | 12,63647498 | 0,001078374 | 0,001346941 | 0,800609553 | 0,423357718 | 0,998407098 | YES |
| 442 | 208,3356985 | 0,000257926 | 0,000329507 | 0,782763185 | 0,43376619 | 0,998407098 | YES |
| 443 | 55,94278565 | -0,000616471 | 0,000802598 | -0,768093606 | 0,442431575 | 0,998407098 | YES |

| Medication | Ticker | ASVnr | taxLevel | Phylum | Family | Genus | Species |
|---|---|---|---|---|---|---|---|
| LEDD | 444 | ASV147 | Genus | Firmicutes | Oscillospiraceae | Oscillibacter | NA |
| LEDD | 445 | ASV53 | Genus | Firmicutes | Lachnospiraceae | [Ruminococcus] torques group | NA |
| LEDD | 446 | ASV55 | Genus | Firmicutes | Monoglobaceae | Monoglobus | NA |
| LEDD | 447 | ASV192 | Genus | Bacteroidota | Marinifilaceae | Odoribacter | NA |
| LEDD | 448 | ASV31 | Genus | Bacteroidota | Rikenellaceae | Alistipes | NA |
| LEDD | 449 | ASV81 | Genus | Firmicutes | Lachnospiraceae | Lachnospiraceae NK4A136 group | NA |
| LEDD | 450 | ASV283 | Genus | Actinobacteriota | Eggerthellaceae | Slackia | NA |
| LEDD | 451 | ASV315 | Genus | Firmicutes | Anaerovoracaceae | Family XIII UCG-001 | NA |
| LEDD | 452 | ASV141 | Genus | Firmicutes | Lachnospiraceae | Lachnoclostridium | NA |
| LEDD | 453 | ASV2 | Genus | Bacteroidota | Bacteroidaceae | Bacteroides | NA |
| LEDD | 454 | ASV20 | Genus | Bacteroidota | Prevotellaceae | Prevotella | NA |
| LEDD | 455 | ASV40 | Genus | Firmicutes | Erysipelotrichaceae | Holdemanella | NA |
| LEDD | 456 | ASV75 | Genus | Firmicutes | Lachnospiraceae | [Ruminococcus] gauvreauii group | NA |
| LEDD | 457 | ASV72 | Genus | Proteobacteria | Sutterellaceae | Parasutterella | NA |
| LEDD | 458 | ASV8 | Genus | Firmicutes | Ruminococcaceae | Faecalibacterium | NA |
| LEDD | 459 | ASV33 | Genus | Firmicutes | Peptostreptococcaceae | Intestinibacter | NA |
| LEDD | 460 | ASV133 | Genus | Actinobacteriota | Eggerthellaceae | Senegalimassilia | NA |
| LEDD | 461 | ASV128 | Genus | Firmicutes | Oscillospiraceae | NK4A214 group | NA |
| LEDD | 462 | ASV411 | Genus | Firmicutes | Lactobacillaceae | Lactobacillus | NA |
| LEDD | 463 | ASV190 | Genus | Firmicutes | Ruminococcaceae | UBA1819 | NA |
| LEDD | 464 | ASV415 | Genus | Firmicutes | Oscillospiraceae | Oscillospira | NA |
| LEDD | 465 | ASV16 | Genus | Firmicutes | Peptostreptococcaceae | Romboutsia | NA |
| LEDD | 466 | ASV487 | Genus | Firmicutes | Lachnospiraceae | Howardella | NA |
| LEDD | 467 | ASV376 | Genus | Bacteroidota | Barnesiellaceae | Coprobacter | NA |
| LEDD | 468 | ASV134 | Genus | Actinobacteriota | Eggerthellaceae | Enterorhabdus | NA |
| LEDD | 469 | ASV51 | Genus | Firmicutes | Oscillospiraceae | UCG-005 | NA |
| LEDD | 470 | ASV29 | Genus | Firmicutes | Oscillospiraceae | UCG-002 | NA |
| LEDD | 471 | ASV60 | Genus | Firmicutes | Ruminococcaceae | [Eubacterium] siraeum group | NA |
| LEDD | 472 | ASV3 | Genus | Firmicutes | Ruminococcaceae | Subdoligranulum | NA |
| LEDD | 473 | ASV30 | Genus | Proteobacteria | Enterobacteriaceae | Escherichia-Shigella | NA |

| Ticker | DESeq2_baseMean | DESeq2_log2-FoldChange | DESeq2_lfcSE | DESeq2_stat | DESeq2_pvalue | DESeq2_padj | DESeq2 converged |
|---|---|---|---|---|---|---|---|
| 444 | 82,17895313 | 0,000413505 | 0,000541117 | 0,764168575 | 0,444766791 | 0,998407098 | YES |
| 445 | 629,5657794 | -0,000264763 | 0,000372243 | -0,711263995 | 0,476920659 | 0,998407098 | YES |
| 446 | 134,5737112 | 0,00036449 | 0,000529287 | 0,688643852 | 0,491047418 | 0,998407098 | YES |
| 447 | 39,33887948 | 0,000326368 | 0,000478958 | 0,681411679 | 0,495611035 | 0,998407098 | YES |
| 448 | 1144,772964 | -0,000178658 | 0,00026738 | -0,668180193 | 0,504018577 | 0,998407098 | YES |
| 449 | 273,0660538 | -0,00123944 | 0,001891199 | -0,655372367 | 0,512228038 | 0,998407098 | YES |
| 450 | 9,730735359 | -0,001085031 | 0,001670705 | -0,649445026 | 0,516050769 | 0,998407098 | YES |
| 451 | 28,1853619 | -0,000413107 | 0,000665179 | -0,621046571 | 0,534568982 | 0,998407098 | YES |
| 452 | 182,5651877 | -0,000334104 | 0,000545616 | -0,612343435 | 0,540310558 | 0,998407098 | YES |
| 453 | 5032,644884 | -0,000146386 | 0,000242251 | -0,604272175 | 0,545662703 | 0,998407098 | YES |
| 454 | 1133,926116 | -0,001645297 | 0,002760742 | -0,595961774 | 0,551200764 | 0,998407098 | YES |
| 455 | 269,6096991 | -0,000366501 | 0,000618323 | -0,592734433 | 0,553358893 | 0,998407098 | YES |
| 456 | 47,70677293 | 0,000894229 | 0,001578829 | 0,566387399 | 0,571130459 | 0,998407098 | YES |
| 457 | 61,45386457 | 0,000443549 | 0,000849799 | 0,521945915 | 0,601707988 | 0,998407098 | YES |
| 458 | 2334,575011 | -0,00100342 | 0,002030627 | -0,494143121 | 0,621205106 | 0,998407098 | YES |
| 459 | 36,62497344 | 0,000612272 | 0,001241509 | 0,493167415 | 0,6218943 | 0,998407098 | YES |
| 460 | 19,54514968 | -0,000469098 | 0,000954145 | -0,49164174 | 0,622972631 | 0,998407098 | YES |
| 461 | 422,4623148 | 0,000760987 | 0,001613263 | 0,471706651 | 0,637136189 | 0,998407098 | YES |
| 462 | 164,6392328 | 0,001618643 | 0,003759068 | 0,430596932 | 0,666761472 | 0,998407098 | YES |
| 463 | 26,39412807 | 0,000595814 | 0,001502449 | 0,396561801 | 0,691690626 | 0,998407098 | YES |
| 464 | 34,73878558 | 0,000489927 | 0,001331999 | 0,367812967 | 0,7130127 | 0,998407098 | YES |
| 465 | 38,6635841 | 0,000909155 | 0,002508728 | 0,362396688 | 0,717055614 | 0,998407098 | YES |
| 466 | 9,850588669 | -0,00066026 | 0,001911363 | -0,345439479 | 0,729764002 | 0,998407098 | YES |
| 467 | 12,213473 | 0,000421521 | 0,001360291 | 0,30987569 | 0,75665549 | 0,998407098 | YES |
| 468 | 8,098004857 | -0,000419517 | 0,001453143 | -0,288696316 | 0,772813782 | 0,998407098 | YES |
| 469 | 668,0635607 | -0,000481547 | 0,001689918 | -0,284953193 | 0,775680002 | 0,998407098 | YES |
| 470 | 2032,576687 | -8,40E-05 | 0,000295751 | -0,284180092 | 0,776272371 | 0,998407098 | YES |
| 471 | 211,2297365 | -0,00020911 | 0,00079392 | -0,263388764 | 0,79225095 | 0,998407098 | YES |
| 472 | 1006,12734 | -9,18E-05 | 0,000355198 | -0,258405827 | 0,79609372 | 0,998407098 | YES |
| 473 | 661,0305271 | -0,000606606 | 0,002361188 | -0,256907226 | 0,797250392 | 0,998407098 | YES |

| Medication | Ticker | ASVnr | taxLevel | Phylum | Family | Genus | Species |
|---|---|---|---|---|---|---|---|
| LEDD | 474 | ASV328 | Genus | Firmicutes | Anaerovoracaceae | [Eubacterium] brachy group | NA |
| LEDD | 475 | ASV26 | Genus | Euryarchaeota | Methanobacteriaceae | Methanobrevibacter | NA |
| LEDD | 476 | ASV92 | Genus | Firmicutes | Erysipelotrichaceae | Turicibacter | NA |
| LEDD | 477 | ASV217 | Genus | Firmicutes | Peptococcaceae | Peptococcus | NA |
| LEDD | 478 | ASV98 | Genus | Firmicutes | Acidaminococcaceae | Phascolarctobacterium | NA |
| LEDD | 479 | ASV130 | Genus | Proteobacteria | Enterobacteriaceae | Klebsiella | NA |
| LEDD | 480 | ASV242 | Genus | Firmicutes | Lachnospiraceae | Tyzzerella | NA |
| LEDD | 481 | ASV28 | Genus | Firmicutes | Clostridiaceae | Clostridium sensu stricto 1 | NA |
| LEDD | 482 | ASV550 | Genus | Bacteroidota | Rikenellaceae | Rikenellaceae RC9 gut group | NA |
| LEDD | 483 | ASV172 | Genus | Proteobacteria | Pasteurellaceae | Haemophilus | NA |
| LEDD | 484 | ASV200 | Genus | Firmicutes | Lachnospiraceae | [Eubacterium] xylanophilum group | NA |
| LEDD | 485 | ASV143 | Genus | Firmicutes | Peptostreptococcaceae | Terrisporobacter | NA |
| LEDD | 486 | ASV338 | Genus | Firmicutes | Lachnospiraceae | Lachnotalea | NA |
| LEDD | 487 | ASV444 | Genus | Firmicutes | Erysipelatoclostridiaceae | Coprobacillus | NA |
| LEDD | 488 | ASV34 | Genus | Firmicutes | Lachnospiraceae | Anaerostipes | NA |
| LEDD | 489 | ASV322 | Genus | Firmicutes | Anaerovoracaceae | Family XIII AD3011 group | NA |
| LEDD | 490 | ASV299 | Genus | Firmicutes | Streptococcaceae | Lactococcus | NA |
| LEDD | 491 | ASV197 | Genus | Bacteroidota | Prevotellaceae | Prevotellaceae NK3B31 group | NA |
| LEDD | 492 | ASV229 | Genus | Firmicutes | Oscillospiraceae | Flavonifractor | NA |
| LEDD | 493 | ASV454 | Genus | Actinobacteriota | Actinomycetaceae | Actinomyces | NA |
| LEDD | 494 | ASV739 | Genus | Firmicutes | Ruminococcaceae | Anaerotruncus | NA |
| LEDD | 495 | ASV303 | Genus | Firmicutes | Lachnospiraceae | GCA-900066575 | NA |
| LEDD | 496 | ASV61 | Genus | Firmicutes | Erysipelatoclostridiaceae | Catenibacterium | NA |
| LEDD | 497 | ASV12 | Genus | Firmicutes | Veillonellaceae | Dialister | NA |
| LEDD | 498 | ASV690 | Genus | Firmicutes | Ruminococcaceae | Fournierella | NA |
| LEDD | 499 | ASV702 | Genus | Firmicutes | Lachnospiraceae | Hungatella | NA |
| LEDD | 500 | ASV27 | Genus | Firmicutes | Ruminococcaceae | CAG-352 | NA |
| LEDD | 501 | ASV1 | Genus | Actinobacteriota | Bifidobacteriaceae | Bifidobacterium | NA |
| LEDD | 502 | ASV1722 | Genus | Firmicutes | Erysipelatoclostridiaceae | Asteroleplasma | NA |
| LEDD | 503 | ASV193 | Genus | Actinobacteriota | Eggerthellaceae | Adlercreutzia | NA |

| Ticker | DESeq2_baseMean | DESeq2_log2-FoldChange | DESeq2_lfcSE | DESeq2_stat | DESeq2_pvalue | DESeq2_padj | DESeq2 converged |
|---|---|---|---|---|---|---|---|
| 474 | 1,595132946 | 0,001241571 | 0,005555242 | 0,22349545 | 0,823149924 | 0,998407098 | YES |
| 475 | 193,8582867 | 0,000453993 | 0,002061437 | 0,220231262 | 0,82569105 | 0,998407098 | YES |
| 476 | 32,84582589 | 0,000665751 | 0,003057593 | 0,217737035 | 0,827634006 | 0,998407098 | YES |
| 477 | 75,38353118 | -0,000234872 | 0,001088668 | -0,215742407 | 0,829188543 | 0,998407098 | YES |
| 478 | 346,0890663 | -0,00037459 | 0,001762602 | -0,212520971 | 0,831700619 | 0,998407098 | YES |
| 479 | 68,07885842 | 0,00116203 | 0,005556918 | 0,209114072 | 0,834359189 | 0,998407098 | YES |
| 480 | 54,98143302 | 0,000715968 | 0,003424441 | 0,209075794 | 0,834389071 | 0,998407098 | YES |
| 481 | 88,43123869 | 0,000131974 | 0,000680811 | 0,193848003 | 0,846294899 | 0,998407098 | YES |
| 482 | 207,2990445 | -0,000542391 | 0,00287587 | -0,188600542 | 0,850405908 | 0,998407098 | YES |
| 483 | 11,93705431 | -0,000966728 | 0,005218559 | -0,185248019 | 0,853034507 | 0,998407098 | YES |
| 484 | 54,39281101 | 0,000378763 | 0,002163283 | 0,175087321 | 0,861011021 | 0,998407098 | YES |
| 485 | 6,591681567 | 0,00085524 | 0,005553478 | 0,15400087 | 0,877609049 | 0,998407098 | YES |
| 486 | 4,438815253 | -0,000849144 | 0,005551812 | -0,152949031 | 0,87843847 | 0,998407098 | YES |
| 487 | 6,609277278 | 0,000740362 | 0,005551678 | 0,133358213 | 0,893910091 | 0,998407098 | YES |
| 488 | 165,7530311 | -0,000227507 | 0,001782256 | -0,127651266 | 0,898424959 | 0,998407098 | YES |
| 489 | 62,06718117 | 5,51E-05 | 0,000433267 | 0,127232363 | 0,898756492 | 0,998407098 | YES |
| 490 | 7,843484646 | 0,00065142 | 0,005554715 | 0,11727333 | 0,906643458 | 0,998407098 | YES |
| 491 | 51,69489275 | -0,000638789 | 0,005555505 | -0,114983045 | 0,908458561 | 0,998407098 | YES |
| 492 | 19,05663697 | -0,000305267 | 0,002676539 | -0,11405285 | 0,909195898 | 0,998407098 | YES |
| 493 | 1,080148438 | 0,000593581 | 0,005555091 | 0,10685344 | 0,914905251 | 0,998407098 | YES |
| 494 | 2,9255055 | 0,000553611 | 0,005556747 | 0,09962859 | 0,920639195 | 0,998407098 | YES |
| 495 | 16,11197157 | 0,000129641 | 0,001387994 | 0,093402073 | 0,925584144 | 0,998407098 | YES |
| 496 | 76,10366348 | -0,00051733 | 0,005551055 | -0,093194822 | 0,925748788 | 0,998407098 | YES |
| 497 | 564,3048754 | -0,000282323 | 0,003086364 | -0,091474217 | 0,927115792 | 0,998407098 | YES |
| 498 | 13,76967531 | -0,000438377 | 0,005550897 | -0,078974034 | 0,937053276 | 0,998407098 | YES |
| 499 | 13,17537301 | -0,000438939 | 0,005559796 | -0,07894878 | 0,937073363 | 0,998407098 | YES |
| 500 | 304,8295096 | -0,000424101 | 0,00554363 | -0,076502402 | 0,939019403 | 0,998407098 | YES |
| 501 | 721,2204328 | 0,000124833 | 0,001671497 | 0,07468341 | 0,940466608 | 0,998407098 | YES |
| 502 | 6,420258122 | -0,000385141 | 0,005557135 | -0,069305674 | 0,94474631 | 0,998407098 | YES |
| 503 | 8,504437492 | 0,000223324 | 0,003294381 | 0,067789401 | 0,945953281 | 0,998407098 | YES |

| Medication | Ticker | ASVnr | taxLevel | Phylum | Family | Genus | Species |
|---|---|---|---|---|---|---|---|
| LEDD | 504 | ASV10 | Genus | Verrucomicrobiota | Akkermansiaceae | Akkermansia | NA |
| LEDD | 505 | ASV247 | Genus | Euryarchaeota | Methanobacteriaceae | Methanosphaera | NA |
| LEDD | 506 | ASV306 | Genus | Firmicutes | Veillonellaceae | Veillonella | NA |
| LEDD | 507 | ASV258 | Genus | Firmicutes | Lachnospiraceae | Lachnospiraceae AC2044 group | NA |
| LEDD | 508 | ASV87 | Genus | Proteobacteria | Sutterellaceae | Sutterella | NA |
| LEDD | 509 | ASV294 | Genus | Firmicutes | Anaerovoracaceae | Mogibacterium | NA |
| LEDD | 510 | ASV167 | Genus | Firmicutes | Oscillospiraceae | Intestinimonas | NA |
| LEDD | 511 | ASV1506 | Genus | Firmicutes | Erysipelatoclostridiaceae | UCG-004 | NA |
| LEDD | 512 | ASV1403 | Genus | Bacteroidota | Marinifilaceae | Sanguibacteroides | NA |
| LEDD | 513 | ASV499 | Genus | Firmicutes | Lachnospiraceae | Eisenbergiella | NA |
| LEDD | 514 | ASV978 | Genus | Firmicutes | Erysipelotrichaceae | Dielma | NA |
| LEDD | 515 | ASV453 | Genus | Verrucomicrobiota | Victivallaceae | Victivallis | NA |
| LEDD | 516 | ASV761 | Genus | Firmicutes | Defluviitaleaceae | Defluviitaleaceae UCG-011 | NA |
| LEDD | 517 | ASV13 | Genus | Firmicutes | Lachnospiraceae | Fusicatenibacter | NA |
| LEDD | 518 | ASV1319 | Genus | Firmicutes | Ruminococcaceae | Candidatus Soleaferrea | NA |
| LEDD | 519 | ASV858 | Genus | Actinobacteriota | Atopobiaceae | Libanicoccus | NA |
| LEDD | 520 | ASV97 | Genus | Firmicutes | Lachnospiraceae | [Ruminococcus] gnavus group | NA |
| LEDD | 521 | ASV15 | Genus | Firmicutes | Christensenellaceae | Christensenellaceae R-7 group | NA |
| LEDD | 522 | ASV162 | Genus | Desulfobacterota | Desulfovibrionaceae | Desulfovibrio | NA |
| LEDD | 523 | ASV1213 | Genus | Desulfobacterota | Desulfovibrionaceae | Mailhella | NA |
| LEDD | 524 | ASV530 | Genus | Firmicutes | Veillonellaceae | Allisonella | NA |
| LEDD | 525 | ASV252 | Genus | Firmicutes | Erysipelotrichaceae | Solobacterium | NA |
| LEDD | 526 | ASV885 | Genus | Firmicutes | Lachnospiraceae | Shuttleworthia | NA |
| LEDD | 527 | ASV385 | Genus | Firmicutes | Selenomonadaceae | Mitsuokella | NA |
| LEDD | 528 | ASV293 | Genus | Firmicutes | Erysipelotrichaceae | Catenisphaera | NA |
| LEDD | 529 | ASV573 | Genus | Firmicutes | Erysipelatoclostridiaceae | Erysipelatoclostridium | NA |
| LEDD | 530 | ASV1631 | Genus | Firmicutes | Ruminococcaceae | Paludicola | NA |
| LEDD | 531 | ASV449 | Genus | Actinobacteriota | Atopobiaceae | Olsenella | NA |
| LEDD | 532 | ASV39 | Genus | Bacteroidota | Tannerellaceae | Parabacteroides | NA |
| LEDD | 533 | ASV240 | Genus | Bacteroidota | Prevotellaceae | Alloprevotella | NA |

| Ticker | DESeq2_baseMean | DESeq2_log2-FoldChange | DESeq2_lfcSE | DESeq2_stat | DESeq2_pvalue | DESeq2_padj | DESeq2 converged |
|---|---|---|---|---|---|---|---|
| 504 | 1143,268978 | 0,000184007 | 0,002957216 | 0,062223198 | 0,950385089 | 0,998407098 | YES |
| 505 | 11,31173121 | 0,000342572 | 0,00555601 | 0,061657843 | 0,950835313 | 0,998407098 | YES |
| 506 | 31,16545164 | -0,000323418 | 0,005245969 | -0,061650707 | 0,950840996 | 0,998407098 | YES |
| 507 | 65,84336143 | 0,000172821 | 0,002846305 | 0,060717616 | 0,951584102 | 0,998407098 | YES |
| 508 | 157,8381505 | 2,98E-05 | 0,000514576 | 0,05783467 | 0,953880322 | 0,998407098 | YES |
| 509 | 25,24876388 | 0,000319808 | 0,005554905 | 0,057572212 | 0,954089385 | 0,998407098 | YES |
| 510 | 131,2055575 | 7,00E-05 | 0,001303048 | 0,053745054 | 0,957138287 | 0,998407098 | YES |
| 511 | 8,366500089 | 0,000293122 | 0,005556102 | 0,05275679 | 0,95792569 | 0,998407098 | YES |
| 512 | 2,94952053 | 0,000286686 | 0,005551863 | 0,05163786 | 0,958817252 | 0,998407098 | YES |
| 513 | 16,69629512 | -0,000280447 | 0,005554103 | -0,050493729 | 0,959728947 | 0,998407098 | YES |
| 514 | 1,873897077 | -0,000277443 | 0,005557631 | -0,049921179 | 0,9601852 | 0,998407098 | YES |
| 515 | 7,171043458 | 0,000179426 | 0,003740535 | 0,047968114 | 0,961741655 | 0,998407098 | YES |
| 516 | 3,384136139 | 0,000264827 | 0,005556082 | 0,047664398 | 0,961983708 | 0,998407098 | YES |
| 517 | 768,4400643 | 9,58E-05 | 0,002085975 | 0,045918809 | 0,963374963 | 0,998407098 | YES |
| 518 | 6,094425354 | -0,000254897 | 0,005553838 | -0,045895575 | 0,963393481 | 0,998407098 | YES |
| 519 | 3,591584218 | 0,000252757 | 0,005556069 | 0,04549209 | 0,963715079 | 0,998407098 | YES |
| 520 | 59,7771096 | 0,000249972 | 0,005556293 | 0,044989024 | 0,964116058 | 0,998407098 | YES |
| 521 | 1021,268723 | 1,70E-05 | 0,000461962 | 0,03688775 | 0,970574507 | 0,998407098 | YES |
| 522 | 265,2572369 | -1,09E-05 | 0,000312116 | -0,034824614 | 0,972219594 | 0,998407098 | YES |
| 523 | 5,714723446 | 0,000180319 | 0,005556961 | 0,032449176 | 0,974113846 | 0,998407098 | YES |
| 524 | 9,50052128 | -0,000169891 | 0,005553368 | -0,030592401 | 0,975594603 | 0,998407098 | YES |
| 525 | 15,48373845 | 0,000162743 | 0,005557031 | 0,029285883 | 0,976636586 | 0,998407098 | YES |
| 526 | 8,078136836 | -0,000161335 | 0,005550616 | -0,029066221 | 0,976811776 | 0,998407098 | YES |
| 527 | 21,24036454 | 0,000155832 | 0,005556964 | 0,028042585 | 0,977628187 | 0,998407098 | YES |
| 528 | 20,19897597 | 0,000142906 | 0,005556295 | 0,025719641 | 0,979480958 | 0,998407098 | YES |
| 529 | 15,74390741 | 5,93E-05 | 0,002359556 | 0,02512933 | 0,979951805 | 0,998407098 | YES |
| 530 | 6,197525142 | -0,000125457 | 0,005547802 | -0,022613816 | 0,981958323 | 0,998407098 | YES |
| 531 | 4,974908538 | 0,000117503 | 0,005548879 | 0,021176004 | 0,983105256 | 0,998407098 | YES |
| 532 | 689,0294243 | -3,43E-05 | 0,001676055 | -0,02045705 | 0,983678774 | 0,998407098 | YES |
| 533 | 144,0791813 | 9,06E-05 | 0,005547693 | 0,01632326 | 0,986976501 | 0,998407098 | YES |

| Medication | Ticker | ASVnr | taxLevel | Phylum | Family | Genus | Species |
|---|---|---|---|---|---|---|---|
| LEDD | 534 | ASV79 | Genus | Firmicutes | Lachnospiraceae | [Eubacterium] ruminantium group | NA |
| LEDD | 535 | ASV78 | Genus | Firmicutes | Lachnospiraceae | [Eubacterium] eligens group | NA |
| LEDD | 536 | ASV428 | Genus | Bacteroidota | Prevotellaceae | Prevotellaceae UCG-001 | NA |
| LEDD | 537 | ASV232 | Genus | Firmicutes | Acholeplasmataceae | Anaeroplasma | NA |
| LEDD | 538 | ASV666 | Genus | Firmicutes | Erysipelotrichaceae | Holdemania | NA |
| LEDD | 539 | ASV892 | Genus | Synergistota | Synergistaceae | Cloacibacillus | NA |
| LEDD | 540 | ASV7 | Family | Firmicutes | Lachnospiraceae | NA | NA |
| LEDD | 541 | ASV4 | Family | Firmicutes | Ruminococcaceae | NA | NA |
| LEDD | 542 | ASV63 | Family | Actinobacteriota | Eggerthellaceae | NA | NA |
| LEDD | 543 | ASV31 | Family | Bacteroidota | Rikenellaceae | NA | NA |
| LEDD | 544 | ASV19 | Family | Firmicutes | Erysipelatoclostridiaceae | NA | NA |
| LEDD | 545 | ASV23 | Family | Firmicutes | Streptococcaceae | NA | NA |
| LEDD | 546 | ASV17 | Family | Actinobacteriota | Coriobacteriaceae | NA | NA |
| LEDD | 547 | ASV59 | Family | Firmicutes | [Eubacterium] coprostanoligenes group | NA | NA |
| LEDD | 548 | ASV16 | Family | Firmicutes | Peptostreptococcaceae | NA | NA |
| LEDD | 549 | ASV66 | Family | Firmicutes | Butyricicoccaceae | NA | NA |
| LEDD | 550 | ASV2 | Family | Bacteroidota | Bacteroidaceae | NA | NA |
| LEDD | 551 | ASV322 | Family | Firmicutes | Anaerovoracaceae | NA | NA |
| LEDD | 552 | ASV40 | Family | Firmicutes | Erysipelotrichaceae | NA | NA |
| LEDD | 553 | ASV512 | Family | Proteobacteria | Oxalobacteraceae | NA | NA |
| LEDD | 554 | ASV195 | Family | Bacteroidota | Barnesiellaceae | NA | NA |
| LEDD | 555 | ASV217 | Family | Firmicutes | Peptococcaceae | NA | NA |
| LEDD | 556 | ASV72 | Family | Proteobacteria | Sutterellaceae | NA | NA |
| LEDD | 557 | ASV20 | Family | Bacteroidota | Prevotellaceae | NA | NA |
| LEDD | 558 | ASV29 | Family | Firmicutes | Oscillospiraceae | NA | NA |
| LEDD | 559 | ASV192 | Family | Bacteroidota | Marinifilaceae | NA | NA |
| LEDD | 560 | ASV255 | Family | Actinobacteriota | Coriobacteriales Incertae Sedis | NA | NA |
| LEDD | 561 | ASV411 | Family | Firmicutes | Lactobacillaceae | NA | NA |
| LEDD | 562 | ASV55 | Family | Firmicutes | Monoglobaceae | NA | NA |
| LEDD | 563 | ASV449 | Family | Actinobacteriota | Atopobiaceae | NA | NA |

| Ticker | DESeq2_baseMean | DESeq2_log2-FoldChange | DESeq2_lfcSE | DESeq2_stat | DESeq2_pvalue | DESeq2_padj | DESeq2 converged |
|---|---|---|---|---|---|---|---|
| 534 | 144,1230944 | -7,37E-05 | 0,004939666 | -0,014929039 | 0,988088792 | 0,998407098 | YES |
| 535 | 197,9799092 | -2,74E-05 | 0,00206975 | -0,013214649 | 0,989456542 | 0,998407098 | YES |
| 536 | 81,0783181 | 6,82E-05 | 0,005555581 | 0,012279733 | 0,990202437 | 0,998407098 | YES |
| 537 | 36,74837665 | 5,34E-05 | 0,005557702 | 0,009616684 | 0,992327115 | 0,998407098 | YES |
| 538 | 2,556745844 | 2,07E-05 | 0,002853623 | 0,00724172 | 0,994221994 | 0,998407098 | YES |
| 539 | 12,2358954 | 1,11E-05 | 0,005555232 | 0,001996408 | 0,998407098 | 0,998407098 | YES |
| 540 | 10920,74407 | -0,000644468 | 0,000208232 | -3,09495098 | 0,001968454 | 0,093331699 | YES |
| 541 | 6452,345527 | -0,000474737 | 0,000164437 | -2,887042192 | 0,003888821 | 0,093331699 | YES |
| 542 | 141,9186143 | -0,000715377 | 0,000324642 | -2,203587086 | 0,027553396 | 0,35117265 | YES |
| 543 | 1311,209727 | -0,000541355 | 0,000248339 | -2,179906574 | 0,029264388 | 0,35117265 | YES |
| 544 | 295,1800348 | -0,000646069 | 0,000322197 | -2,005196244 | 0,044942071 | 0,431443886 | YES |
| 545 | 291,678827 | 0,001346479 | 0,000709612 | 1,897485686 | 0,057763866 | 0,462110929 | YES |
| 546 | 245,8784998 | -0,000783624 | 0,000467672 | -1,67558372 | 0,09381976 | 0,626924404 | YES |
| 547 | 428,7437323 | 0,000398644 | 0,000248736 | 1,60268155 | 0,109004979 | 0,626924404 | YES |
| 548 | 86,69618105 | 0,001103583 | 0,000705098 | 1,56514751 | 0,117548326 | 0,626924404 | YES |
| 549 | 220,0873183 | -0,000401912 | 0,000342079 | -1,174909455 | 0,240030943 | 0,997819615 | YES |
| 550 | 5171,570003 | -0,000241596 | 0,000233581 | -1,034314133 | 0,300989332 | 0,997819615 | YES |
| 551 | 117,7214411 | 0,000282381 | 0,000305406 | 0,924607381 | 0,355170172 | 0,997819615 | YES |
| 552 | 363,4789923 | -0,000357745 | 0,000393539 | -0,909046604 | 0,363325526 | 0,997819615 | YES |
| 553 | 13,04688119 | 0,000558045 | 0,000677571 | 0,823596243 | 0,410169011 | 0,997819615 | YES |
| 554 | 310,119534 | -0,001062574 | 0,001580893 | -0,672134925 | 0,501497805 | 0,997819615 | YES |
| 555 | 87,25824975 | -0,000401143 | 0,000607303 | -0,660531373 | 0,508912892 | 0,997819615 | YES |
| 556 | 202,7427138 | -0,000243024 | 0,000378857 | -0,64146744 | 0,521219029 | 0,997819615 | YES |
| 557 | 1582,448966 | -0,001403475 | 0,002325522 | -0,603509803 | 0,546169596 | 0,997819615 | YES |
| 558 | 3604,729134 | -0,00011416 | 0,000191373 | -0,596532963 | 0,55081924 | 0,997819615 | YES |
| 559 | 85,69374475 | 0,000174967 | 0,000321373 | 0,544434507 | 0,586142505 | 0,997819615 | YES |
| 560 | 24,05154352 | 0,000498341 | 0,001051924 | 0,473741839 | 0,635684014 | 0,997819615 | YES |
| 561 | 197,0007947 | 0,00173628 | 0,003751156 | 0,462865292 | 0,643460927 | 0,997819615 | YES |
| 562 | 145,7697528 | 0,000229439 | 0,000535153 | 0,428735078 | 0,66811603 | 0,997819615 | YES |
| 563 | 14,18352558 | -0,000377827 | 0,001592785 | -0,237211592 | 0,812492644 | 0,997819615 | YES |

| Medication | Ticker | ASVnr | taxLevel | Phylum | Family | Genus | Species |
|---|---|---|---|---|---|---|---|
| LEDD | 564 | ASV172 | Family | Proteobacteria | Pasteurellaceae | NA | NA |
| LEDD | 565 | ASV26 | Family | Euryarchaeota | Methanobacteriaceae | NA | NA |
| LEDD | 566 | ASV98 | Family | Firmicutes | Acidaminococcaceae | NA | NA |
| LEDD | 567 | ASV453 | Family | Verrucomicrobiota | Victivallaceae | NA | NA |
| LEDD | 568 | ASV39 | Family | Bacteroidota | Tannerellaceae | NA | NA |
| LEDD | 569 | ASV802 | Family | Firmicutes | Ruminiclostridium | NA | NA |
| LEDD | 570 | ASV30 | Family | Proteobacteria | Enterobacteriaceae | NA | NA |
| LEDD | 571 | ASV703 | Family | Firmicutes | Hydrogenoanaerobacterium | NA | NA |
| LEDD | 572 | ASV417 | Family | Firmicutes | UCG-010 | NA | NA |
| LEDD | 573 | ASV145 | Family | Bacteroidota | Muribaculaceae | NA | NA |
| LEDD | 574 | ASV1119 | Family | Verrucomicrobiota | vadinBE97 | NA | NA |
| LEDD | 575 | ASV28 | Family | Firmicutes | Clostridiaceae | NA | NA |
| LEDD | 576 | ASV162 | Family | Desulfobacterota | Desulfovibrionaceae | NA | NA |
| LEDD | 577 | ASV752 | Family | Verrucomicrobiota | Puniceicoccaceae | NA | NA |
| LEDD | 578 | ASV454 | Family | Actinobacteriota | Actinomycetaceae | NA | NA |
| LEDD | 579 | ASV15 | Family | Firmicutes | Christensenellaceae | NA | NA |
| LEDD | 580 | ASV761 | Family | Firmicutes | Defluviitaleaceae | NA | NA |
| LEDD | 581 | ASV10 | Family | Verrucomicrobiota | Akkermansiaceae | NA | NA |
| LEDD | 582 | ASV892 | Family | Synergistota | Synergistaceae | NA | NA |
| LEDD | 583 | ASV1052 | Family | Thermoplasmatota | Methanomethylophilaceae | NA | NA |
| LEDD | 584 | ASV12 | Family | Firmicutes | Veillonellaceae | NA | NA |
| LEDD | 585 | ASV232 | Family | Firmicutes | Acholeplasmataceae | NA | NA |
| LEDD | 586 | ASV385 | Family | Firmicutes | Selenomonadaceae | NA | NA |
| LEDD | 587 | ASV1 | Family | Actinobacteriota | Bifidobacteriaceae | NA | NA |

| Ticker | DESeq2_baseMean | DESeq2_log2-FoldChange | DESeq2_lfcSE | DESeq2_stat | DESeq2_pvalue | DESeq2_padj | DESeq2 converged |
|---|---|---|---|---|---|---|---|
| 564 | 13,90091425 | -0,000919637 | 0,004069706 | -0,225971376 | 0,821223683 | 0,997819615 | YES |
| 565 | 172,2411344 | 0,000406431 | 0,00196065 | 0,207294261 | 0,835780056 | 0,997819615 | YES |
| 566 | 393,6239555 | -0,000295198 | 0,001783756 | -0,165492355 | 0,868556467 | 0,997819615 | YES |
| 567 | 9,01369094 | -0,000259155 | 0,002118185 | -0,122347557 | 0,90262377 | 0,997819615 | YES |
| 568 | 715,5677576 | -0,000208885 | 0,001744552 | -0,119735498 | 0,904692679 | 0,997819615 | YES |
| 569 | 4,25990335 | 0,000548675 | 0,005553835 | 0,098792093 | 0,921303347 | 0,997819615 | YES |
| 570 | 816,1672056 | 0,00021035 | 0,002262824 | 0,092958976 | 0,925936152 | 0,997819615 | YES |
| 571 | 2,712422674 | -0,000512032 | 0,005553794 | -0,092194907 | 0,926543184 | 0,997819615 | YES |
| 572 | 163,9145478 | -0,00013716 | 0,001540162 | -0,089055668 | 0,929037669 | 0,997819615 | YES |
| 573 | 410,9691689 | -0,000204396 | 0,002445203 | -0,083590801 | 0,933381781 | 0,997819615 | YES |
| 574 | 9,106987731 | 0,000420817 | 0,00555074 | 0,075812813 | 0,939568022 | 0,997819615 | YES |
| 575 | 100,5839013 | -4,51E-05 | 0,000686752 | -0,065670252 | 0,947640357 | 0,997819615 | YES |
| 576 | 326,2821135 | -1,87E-05 | 0,000294269 | -0,063400467 | 0,949447615 | 0,997819615 | YES |
| 577 | 5,435988665 | -0,000335518 | 0,005547906 | -0,060476559 | 0,951776085 | 0,997819615 | YES |
| 578 | 1,250716078 | 0,000331611 | 0,005552294 | 0,059725008 | 0,952374653 | 0,997819615 | YES |
| 579 | 1338,01187 | -2,54E-05 | 0,000454375 | -0,055946102 | 0,955384745 | 0,997819615 | YES |
| 580 | 2,457580465 | 0,000208477 | 0,0055563 | 0,037520925 | 0,970069656 | 0,997819615 | YES |
| 581 | 923,0790758 | 7,74E-05 | 0,002801377 | 0,027626374 | 0,977960146 | 0,997819615 | YES |
| 582 | 9,128634694 | 9,11E-05 | 0,005550951 | 0,016418303 | 0,986900678 | 0,997819615 | YES |
| 583 | 3,550815016 | 8,19E-05 | 0,005558854 | 0,014734168 | 0,98824426 | 0,997819615 | YES |
| 584 | 739,0395625 | 1,27E-05 | 0,002381142 | 0,005346894 | 0,995733816 | 0,997819615 | YES |
| 585 | 27,48862548 | -2,68E-05 | 0,00555766 | -0,004830513 | 0,996145823 | 0,997819615 | YES |
| 586 | 18,38767109 | 1,93E-05 | 0,005556931 | 0,003478632 | 0,997224459 | 0,997819615 | YES |
| 587 | 719,8268243 | -4,62E-06 | 0,001689437 | -0,002732711 | 0,997819615 | 0,997819615 | YES |

**Supplementary Table 8b: Levodopa**

| Medication | Ticker | ASVnr | taxLevel | Phylum | Family | Genus | Species |
|---|---|---|---|---|---|---|---|
| Levodopa | 1 | ASV76 | ASV | Firmicutes | Lachnospiraceae | Lachnospiraceae ND3007 group | NA |
| Levodopa | 2 | ASV7 | ASV | Firmicutes | Lachnospiraceae | Agathobacter | NA |
| Levodopa | 3 | ASV6 | ASV | Firmicutes | Ruminococcaceae | Faecalibacterium | NA |
| Levodopa | 4 | ASV45 | ASV | Firmicutes | Lachnospiraceae | Coprococcus | comes |
| Levodopa | 5 | ASV36 | ASV | Firmicutes | Lachnospiraceae | Blautia | obeum |
| Levodopa | 6 | ASV309 | ASV | Firmicutes | Oscillospiraceae | Colidextribacter | massiliensis |
| Levodopa | 7 | ASV157 | ASV | Firmicutes | Oscillospiraceae | UCG-005 | NA |
| Levodopa | 8 | ASV37 | ASV | Firmicutes | Lachnospiraceae | [Eubacterium] hallii group | NA |
| Levodopa | 9 | ASV53 | ASV | Firmicutes | Lachnospiraceae | [Ruminococcus] torques group | NA |
| Levodopa | 10 | ASV144 | ASV | Desulfobacterota | Desulfovibrionaceae | Bilophila | wadsworthia |
| Levodopa | 11 | ASV19 | ASV | Firmicutes | Erysipelatoclostridiaceae | Erysipelotrichaceae UCG-003 | bacterium |
| Levodopa | 12 | ASV21 | ASV | Firmicutes | Ruminococcaceae | Faecalibacterium | prausnitzii |
| Levodopa | 13 | ASV3 | ASV | Firmicutes | Ruminococcaceae | Subdoligranulum | NA |
| Levodopa | 14 | ASV14 | ASV | Firmicutes | Lachnospiraceae | Blautia | massiliensis |
| Levodopa | 15 | ASV11 | ASV | Firmicutes | Ruminococcaceae | Subdoligranulum | NA |
| Levodopa | 16 | ASV43 | ASV | Firmicutes | Lachnospiraceae | Blautia | faecis |
| Levodopa | 17 | ASV211 | ASV | Firmicutes | Lachnospiraceae | Lachnospiraceae FCS020 group | NA |
| Levodopa | 18 | ASV74 | ASV | Firmicutes | Lachnospiraceae | Coprococcus | catus |
| Levodopa | 19 | ASV32 | ASV | Firmicutes | Oscillospiraceae | UCG-002 | NA |
| Levodopa | 20 | ASV86 | ASV | Firmicutes | Lachnospiraceae | [Eubacterium] hallii group | NA |
| Levodopa | 21 | ASV65 | ASV | Firmicutes | Ruminococcaceae | Incertae Sedis | NA |
| Levodopa | 22 | ASV28 | ASV | Firmicutes | Clostridiaceae | Clostridium sensu stricto 1 | NA |
| Levodopa | 23 | ASV96 | ASV | Firmicutes | Lachnospiraceae | CAG-56 | NA |
| Levodopa | 24 | ASV267 | ASV | Firmicutes | Lachnospiraceae | Lachnoclostridium | NA |
| Levodopa | 25 | ASV158 | ASV | Firmicutes | Lachnospiraceae | [Eubacterium] ventriosum group | NA |
| Levodopa | 26 | ASV13 | ASV | Firmicutes | Lachnospiraceae | Fusicatenibacter | saccharivorans |
| Levodopa | 27 | ASV47 | ASV | Bacteroidota | Rikenellaceae | Alistipes | NA |
| Levodopa | 28 | ASV39 | ASV | Bacteroidota | Tannerellaceae | Parabacteroides | merdae |
| Levodopa | 29 | ASV403 | ASV | Firmicutes | Oscillospiraceae | Colidextribacter | NA |

| Ticker | DESeq2_baseMean | DESeq2_log2-FoldChange | DESeq2_lfcSE | DESeq2_stat | DESeq2_pvalue | DESeq2_padj | DESeq2 converged |
|---|---|---|---|---|---|---|---|
| 1 | 72,59997327 | -0,003209906 | 0,001033188 | -3,10679712 | 0,001891261 | 0,777308457 | YES |
| 2 | 1602,321472 | -0,002336178 | 0,000956187 | -2,443222812 | 0,014556747 | 0,999413296 | YES |
| 3 | 779,6942869 | -0,001812214 | 0,000766084 | -2,365556341 | 0,018003004 | 0,999413296 | YES |
| 4 | 121,089678 | -0,001782772 | 0,000815972 | -2,184843168 | 0,028900348 | 0,999413296 | YES |
| 5 | 195,4902888 | -0,001713037 | 0,001003705 | -1,706713483 | 0,08787532 | 0,999413296 | YES |
| 6 | 26,1628603 | -0,003150012 | 0,001867867 | -1,686421702 | 0,091714604 | 0,999413296 | YES |
| 7 | 67,23477775 | -0,002098238 | 0,001261355 | -1,663479019 | 0,096216587 | 0,999413296 | YES |
| 8 | 143,4480555 | -0,001215348 | 0,000759151 | -1,600931462 | 0,1093921 | 0,999413296 | YES |
| 9 | 249,2766116 | -0,001577053 | 0,001104485 | -1,427862615 | 0,153331406 | 0,999413296 | YES |
| 10 | 50,9190978 | 0,001386706 | 0,001019807 | 1,359772211 | 0,173902019 | 0,999413296 | YES |
| 11 | 139,7331795 | -0,000916801 | 0,000726229 | -1,262413155 | 0,206800158 | 0,999413296 | YES |
| 12 | 347,8353987 | -0,003506224 | 0,003304625 | -1,061004954 | 0,28868765 | 0,999413296 | YES |
| 13 | 531,6351586 | 0,000875591 | 0,000827229 | 1,058462734 | 0,289844532 | 0,999413296 | YES |
| 14 | 367,1702874 | -0,00087766 | 0,000831665 | -1,055303801 | 0,291286403 | 0,999413296 | YES |
| 15 | 374,3666094 | -0,000815176 | 0,00077356 | -1,053797729 | 0,291975533 | 0,999413296 | YES |
| 16 | 78,55330611 | -0,001376503 | 0,001396222 | -0,985877372 | 0,324193292 | 0,999413296 | YES |
| 17 | 43,96681586 | -0,00165284 | 0,001795591 | -0,920499424 | 0,357311834 | 0,999413296 | YES |
| 18 | 87,92888506 | -0,000588681 | 0,000646932 | -0,909957822 | 0,362844754 | 0,999413296 | YES |
| 19 | 541,6644648 | -0,002762155 | 0,003052503 | -0,904881904 | 0,365527955 | 0,999413296 | YES |
| 20 | 56,64768015 | -0,001303089 | 0,001451432 | -0,897795837 | 0,369294405 | 0,999413296 | YES |
| 21 | 74,33547493 | 0,001948835 | 0,002225923 | 0,875517431 | 0,38129243 | 0,999413296 | YES |
| 22 | 67,08498444 | 0,001650364 | 0,001903309 | 0,867102224 | 0,385886002 | 0,999413296 | YES |
| 23 | 117,0625667 | -0,002537438 | 0,003161384 | -0,802635242 | 0,422185593 | 0,999413296 | YES |
| 24 | 23,88138539 | -0,001749812 | 0,002191596 | -0,798419125 | 0,424627308 | 0,999413296 | YES |
| 25 | 62,51801145 | -0,001102364 | 0,001413497 | -0,779884361 | 0,435458944 | 0,999413296 | YES |
| 26 | 737,9910079 | -0,000554114 | 0,000734623 | -0,754282971 | 0,450679325 | 0,999413296 | YES |
| 27 | 199,5822721 | -0,001491089 | 0,001998616 | -0,746060811 | 0,455630681 | 0,999413296 | YES |
| 28 | 331,2558741 | 0,000472887 | 0,000663689 | 0,712513351 | 0,476146947 | 0,999413296 | YES |
| 29 | 9,989651428 | -0,002045368 | 0,002939557 | -0,695808366 | 0,486548849 | 0,999413296 | YES |

| Medication | Ticker | ASVnr | taxLevel | Phylum | Family | Genus | Species |
|---|---|---|---|---|---|---|---|
| Levodopa | 30 | ASV5 | ASV | Firmicutes | Lachnospiraceae | Blautia | NA |
| Levodopa | 31 | ASV31 | ASV | Bacteroidota | Rikenellaceae | Alistipes | putredinis |
| Levodopa | 32 | ASV626 | ASV | Firmicutes | NA | NA | NA |
| Levodopa | 33 | ASV48 | ASV | Bacteroidota | Bacteroidaceae | Bacteroides | caccae |
| Levodopa | 34 | ASV88 | ASV | Firmicutes | Lachnospiraceae | Dorea | formicigenerans |
| Levodopa | 35 | ASV111 | ASV | Firmicutes | Lachnospiraceae | Lachnospira | pectinoschiza |
| Levodopa | 36 | ASV168 | ASV | Firmicutes | Lachnospiraceae | Lachnoclostridium | edouardi |
| Levodopa | 37 | ASV68 | ASV | Firmicutes | Ruminococcaceae | Faecalibacterium | NA |
| Levodopa | 38 | ASV226 | ASV | Firmicutes | Lachnospiraceae | Lachnospiraceae UCG-001 | NA |
| Levodopa | 39 | ASV406 | ASV | Firmicutes | NA | NA | NA |
| Levodopa | 40 | ASV24 | ASV | Bacteroidota | Bacteroidaceae | Bacteroides | uniformis |
| Levodopa | 41 | ASV192 | ASV | Bacteroidota | Marinifilaceae | Odoribacter | splanchnicus |
| Levodopa | 42 | ASV56 | ASV | Bacteroidota | Bacteroidaceae | Bacteroides | massiliensis |
| Levodopa | 43 | ASV34 | ASV | Firmicutes | Lachnospiraceae | Anaerostipes | hadrus |
| Levodopa | 44 | ASV2 | ASV | Bacteroidota | Bacteroidaceae | Bacteroides | vulgatus |
| Levodopa | 45 | ASV89 | ASV | Firmicutes | Lachnospiraceae | Blautia | NA |
| Levodopa | 46 | ASV357 | ASV | Firmicutes | Lachnospiraceae | Marvinbryantia | NA |
| Levodopa | 47 | ASV219 | ASV | Firmicutes | Lachnospiraceae | Lachnospiraceae UCG-004 | NA |
| Levodopa | 48 | ASV139 | ASV | Firmicutes | Christensenellaceae | Christensenellaceae R-7 group | NA |
| Levodopa | 49 | ASV264 | ASV | Firmicutes | Oscillospiraceae | Colidextribacter | NA |
| Levodopa | 50 | ASV222 | ASV | Firmicutes | Oscillospiraceae | NK4A214 group | NA |
| Levodopa | 51 | ASV125 | ASV | Firmicutes | Oscillospiraceae | UCG-002 | NA |
| Levodopa | 52 | ASV94 | ASV | Bacteroidota | Rikenellaceae | Alistipes | shahii |
| Levodopa | 53 | ASV63 | ASV | Actinobacteriota | Eggerthellaceae | NA | NA |
| Levodopa | 54 | ASV46 | ASV | Bacteroidota | Bacteroidaceae | Bacteroides | NA |
| Levodopa | 55 | ASV64 | ASV | Firmicutes | Oscillospiraceae | UCG-002 | NA |
| Levodopa | 56 | ASV33 | ASV | Firmicutes | Peptostreptococcaceae | Intestinibacter | bartlettii |
| Levodopa | 57 | ASV66 | ASV | Firmicutes | Butyricicoccaceae | Butyricicoccus | NA |
| Levodopa | 58 | ASV17 | ASV | Actinobacteriota | Coriobacteriaceae | Collinsella | aerofaciens |
| Levodopa | 59 | ASV116 | ASV | Firmicutes | Oscillospiraceae | UCG-003 | NA |

| Ticker | DESeq2_baseMean | DESeq2_log2-FoldChange | DESeq2_lfcSE | DESeq2_stat | DESeq2_pvalue | DESeq2_padj | DESeq2 converged |
|---|---|---|---|---|---|---|---|
| 30 | 415,0190944 | 0,000405373 | 0,000588301 | 0,689057315 | 0,4907872 | 0,999413296 | YES |
| 31 | 273,8873852 | -0,001940515 | 0,002864538 | -0,67742679 | 0,498135206 | 0,999413296 | YES |
| 32 | 15,32174136 | 0,001736377 | 0,002583267 | 0,672163279 | 0,501479756 | 0,999413296 | YES |
| 33 | 261,9482242 | 0,000959365 | 0,001509807 | 0,635422084 | 0,525153169 | 0,999413296 | YES |
| 34 | 42,28933085 | 0,001172178 | 0,001891628 | 0,619666139 | 0,535477613 | 0,999413296 | YES |
| 35 | 77,76089022 | -0,002195549 | 0,003573316 | -0,614429045 | 0,538931848 | 0,999413296 | YES |
| 36 | 49,92685806 | -0,001108316 | 0,001924856 | -0,575791403 | 0,564756185 | 0,999413296 | YES |
| 37 | 96,46549968 | -0,000852863 | 0,00149434 | -0,570729158 | 0,56818325 | 0,999413296 | YES |
| 38 | 50,61471211 | -0,001800059 | 0,003165775 | -0,568599819 | 0,569627748 | 0,999413296 | YES |
| 39 | 8,77073113 | -0,001032723 | 0,001827252 | -0,565178124 | 0,571952615 | 0,999413296 | YES |
| 40 | 412,0140052 | -0,001957058 | 0,003473175 | -0,563478115 | 0,573109358 | 0,999413296 | YES |
| 41 | 33,17019841 | 0,000443435 | 0,000788919 | 0,562079509 | 0,574061849 | 0,999413296 | YES |
| 42 | 164,1406568 | -0,000945538 | 0,001724638 | -0,548253258 | 0,583518018 | 0,999413296 | YES |
| 43 | 132,8820657 | -0,001528071 | 0,002864216 | -0,533504265 | 0,59368456 | 0,999413296 | YES |
| 44 | 1030,976376 | 0,000339978 | 0,000650241 | 0,522849901 | 0,601078711 | 0,999413296 | YES |
| 45 | 71,7924502 | 0,001794505 | 0,003483243 | 0,51518216 | 0,606425734 | 0,999413296 | YES |
| 46 | 35,49744459 | 0,001107587 | 0,002185304 | 0,506834533 | 0,612270924 | 0,999413296 | YES |
| 47 | 27,64704295 | -0,001316879 | 0,002605339 | -0,50545417 | 0,61323988 | 0,999413296 | YES |
| 48 | 227,287366 | 0,002007457 | 0,004057527 | 0,494748862 | 0,620777406 | 0,999413296 | YES |
| 49 | 26,3743812 | 0,001340521 | 0,002758893 | 0,485890915 | 0,62704451 | 0,999413296 | YES |
| 50 | 48,41260651 | 0,000600149 | 0,001249284 | 0,480394353 | 0,630947009 | 0,999413296 | YES |
| 51 | 248,9458341 | 0,000651249 | 0,0013622 | 0,478085865 | 0,632589092 | 0,999413296 | YES |
| 52 | 92,30897336 | 0,001931604 | 0,004084754 | 0,4728813 | 0,636297867 | 0,999413296 | YES |
| 53 | 83,76738392 | -0,00042977 | 0,000910134 | -0,472204915 | 0,636780532 | 0,999413296 | YES |
| 54 | 123,4362494 | 0,001452331 | 0,003092172 | 0,469680031 | 0,638583637 | 0,999413296 | YES |
| 55 | 297,2102856 | -0,001000108 | 0,002172946 | -0,460254418 | 0,645333615 | 0,999413296 | YES |
| 56 | 35,22493639 | 0,001014165 | 0,002226574 | 0,455482224 | 0,648762354 | 0,999413296 | YES |
| 57 | 72,12331995 | -0,000447842 | 0,000984976 | -0,454673068 | 0,649344459 | 0,999413296 | YES |
| 58 | 195,593335 | -0,000917493 | 0,002179373 | -0,42098933 | 0,673762873 | 0,999413296 | YES |
| 59 | 130,9450931 | -0,001171211 | 0,002868015 | -0,408369861 | 0,683002157 | 0,999413296 | YES |

| Medication | Ticker | ASVnr | taxLevel | Phylum | Family | Genus | Species |
|---|---|---|---|---|---|---|---|
| Levodopa | 60 | ASV283 | ASV | Actinobacteriota | Eggerthellaceae | Slackia | isoflavoniconvertens |
| Levodopa | 61 | ASV387 | ASV | Firmicutes | Lachnospiraceae | NA | NA |
| Levodopa | 62 | ASV71 | ASV | Firmicutes | [Eubacterium] coprostanoligenes group | NA | NA |
| Levodopa | 63 | ASV77 | ASV | Firmicutes | Lachnospiraceae | [Ruminococcus] torques group | NA |
| Levodopa | 64 | ASV123 | ASV | Firmicutes | Lachnospiraceae | [Ruminococcus] torques group | NA |
| Levodopa | 65 | ASV26 | ASV | Euryarchaeota | Methanobacteriaceae | Methanobrevibacter | NA |
| Levodopa | 66 | ASV137 | ASV | Firmicutes | Butyricicoccaceae | Butyricicoccus | faecihominis |
| Levodopa | 67 | ASV117 | ASV | Bacteroidota | Rikenellaceae | Alistipes | finegoldii |
| Levodopa | 68 | ASV268 | ASV | Firmicutes | Ruminococcaceae | Incertae Sedis | NA |
| Levodopa | 69 | ASV109 | ASV | Firmicutes | [Eubacterium] coprostanoligenes group | NA | NA |
| Levodopa | 70 | ASV30 | ASV | Proteobacteria | Enterobacteriaceae | Escherichia-Shigella | NA |
| Levodopa | 71 | ASV370 | ASV | Firmicutes | UCG-010 | NA | NA |
| Levodopa | 72 | ASV8 | ASV | Firmicutes | Ruminococcaceae | Faecalibacterium | prausnitzii |
| Levodopa | 73 | ASV119 | ASV | Firmicutes | Ruminococcaceae | Subdoligranulum | NA |
| Levodopa | 74 | ASV159 | ASV | Firmicutes | Lachnospiraceae | Roseburia | NA |
| Levodopa | 75 | ASV20 | ASV | Bacteroidota | Prevotellaceae | Prevotella | copri |
| Levodopa | 76 | ASV25 | ASV | Actinobacteriota | Bifidobacteriaceae | Bifidobacterium | NA |
| Levodopa | 77 | ASV127 | ASV | Firmicutes | Lachnospiraceae | Lachnospiraceae NK4A136 group | bacterium |
| Levodopa | 78 | ASV136 | ASV | Firmicutes | Ruminococcaceae | NA | NA |
| Levodopa | 79 | ASV73 | ASV | Firmicutes | Oscillospiraceae | UCG-005 | NA |
| Levodopa | 80 | ASV85 | ASV | Firmicutes | Lachnospiraceae | Coprococcus | NA |
| Levodopa | 81 | ASV487 | ASV | Firmicutes | Lachnospiraceae | Howardella | ureilytica |
| Levodopa | 82 | ASV101 | ASV | Firmicutes | Lachnospiraceae | Roseburia | NA |
| Levodopa | 83 | ASV1 | ASV | Actinobacteriota | Bifidobacteriaceae | Bifidobacterium | NA |
| Levodopa | 84 | ASV100 | ASV | Firmicutes | Christensenellaceae | NA | NA |
| Levodopa | 85 | ASV701 | ASV | Firmicutes | Lachnospiraceae | Lachnoclostridium | NA |
| Levodopa | 86 | ASV287 | ASV | Firmicutes | Christensenellaceae | Christensenellaceae R-7 group | NA |
| Levodopa | 87 | ASV415 | ASV | Firmicutes | Oscillospiraceae | Oscillospira | NA |
| Levodopa | 88 | ASV276 | ASV | Firmicutes | Oscillospiraceae | NK4A214 group | NA |
| Levodopa | 89 | ASV35 | ASV | Firmicutes | Lachnospiraceae | Agathobacter | NA |

| Ticker | DESeq2_baseMean | DESeq2_log2-FoldChange | DESeq2_lfcSE | DESeq2_stat | DESeq2_pvalue | DESeq2_padj | DESeq2 converged |
|---|---|---|---|---|---|---|---|
| 60 | 10,45285503 | -0,001093662 | 0,002696196 | -0,405631594 | 0,685013311 | 0,999413296 | YES |
| 61 | 12,27709941 | -0,002422047 | 0,006178395 | -0,392018831 | 0,6950443 | 0,999413296 | YES |
| 62 | 45,26333147 | 0,00114085 | 0,002925841 | 0,389922213 | 0,696594068 | 0,999413296 | YES |
| 63 | 125,0317041 | 0,001647393 | 0,004325425 | 0,380862696 | 0,703305134 | 0,999413296 | YES |
| 64 | 123,9750089 | -0,001865198 | 0,00505017 | -0,36933369 | 0,711879017 | 0,999413296 | YES |
| 65 | 218,7222979 | 0,001030202 | 0,002886368 | 0,35691988 | 0,721151787 | 0,999413296 | YES |
| 66 | 23,4462083 | 0,000988093 | 0,002857706 | 0,345764509 | 0,729519699 | 0,999413296 | YES |
| 67 | 33,60063308 | 0,002100295 | 0,006170921 | 0,340353582 | 0,73359027 | 0,999413296 | YES |
| 68 | 12,64849219 | 0,00145576 | 0,004301422 | 0,338436797 | 0,735034048 | 0,999413296 | YES |
| 69 | 100,9978767 | -0,00136885 | 0,004065959 | -0,336660918 | 0,736372528 | 0,999413296 | YES |
| 70 | 723,4835665 | -0,001052318 | 0,003138033 | -0,335343247 | 0,737366174 | 0,999413296 | YES |
| 71 | 12,43737457 | -0,001368916 | 0,004108748 | -0,333171086 | 0,739005143 | 0,999413296 | YES |
| 72 | 1045,211502 | -0,001101021 | 0,003498723 | -0,314692384 | 0,752995227 | 0,999413296 | YES |
| 73 | 59,90597499 | -0,001146622 | 0,003691565 | -0,310606016 | 0,756100153 | 0,999413296 | YES |
| 74 | 53,00377714 | -0,000748931 | 0,002534477 | -0,295497421 | 0,767613922 | 0,999413296 | YES |
| 75 | 292,2343386 | -0,001743154 | 0,006116732 | -0,284981261 | 0,775658499 | 0,999413296 | YES |
| 76 | 287,2858951 | 0,00041573 | 0,001472331 | 0,282361934 | 0,777666 | 0,999413296 | YES |
| 77 | 52,11686268 | -0,001729344 | 0,006166498 | -0,280441859 | 0,779138526 | 0,999413296 | YES |
| 78 | 136,9658738 | -0,000355688 | 0,001271397 | -0,27976196 | 0,779660138 | 0,999413296 | YES |
| 79 | 225,4569035 | 0,000772748 | 0,002793609 | 0,27661282 | 0,78207742 | 0,999413296 | YES |
| 80 | 235,0164518 | -0,001707382 | 0,006180239 | -0,276264801 | 0,782344689 | 0,999413296 | YES |
| 81 | 9,254855614 | -0,001179562 | 0,004309962 | -0,273682677 | 0,784328491 | 0,999413296 | YES |
| 82 | 88,91672255 | 0,00163798 | 0,006176424 | 0,265198794 | 0,790856324 | 0,999413296 | YES |
| 83 | 346,6327265 | -0,001024173 | 0,003960409 | -0,258602733 | 0,795941774 | 0,999413296 | YES |
| 84 | 357,1733653 | 0,000766743 | 0,00296535 | 0,258567603 | 0,795968883 | 0,999413296 | YES |
| 85 | 2,559200671 | 0,001566186 | 0,006184629 | 0,253238418 | 0,80008398 | 0,999413296 | YES |
| 86 | 36,29230641 | -0,000629868 | 0,002506671 | -0,25127685 | 0,801600072 | 0,999413296 | YES |
| 87 | 9,415592399 | 0,001547516 | 0,006175608 | 0,250585236 | 0,802134797 | 0,999413296 | YES |
| 88 | 27,83267671 | 0,001508231 | 0,006186142 | 0,243807986 | 0,807379535 | 0,999413296 | YES |
| 89 | 320,0109162 | -0,001499199 | 0,00616017 | -0,243369796 | 0,80771894 | 0,999413296 | YES |

| Medication | Ticker | ASVnr | taxLevel | Phylum | Family | Genus | Species |
|---|---|---|---|---|---|---|---|
| Levodopa | 90 | ASV362 | ASV | Firmicutes | Ruminococcaceae | NA | NA |
| Levodopa | 91 | ASV138 | ASV | Bacteroidota | Rikenellaceae | Alistipes | obesi |
| Levodopa | 92 | ASV104 | ASV | Firmicutes | Christensenellaceae | Christensenellaceae R-7 group | NA |
| Levodopa | 93 | ASV180 | ASV | Firmicutes | Ruminococcaceae | Ruminococcus | NA |
| Levodopa | 94 | ASV315 | ASV | Firmicutes | Anaerovoracaceae | Family XIII UCG-001 | NA |
| Levodopa | 95 | ASV451 | ASV | Firmicutes | Oscillospiraceae | UCG-005 | NA |
| Levodopa | 96 | ASV70 | ASV | Firmicutes | Lachnospiraceae | Roseburia | intestinalis |
| Levodopa | 97 | ASV436 | ASV | Firmicutes | Christensenellaceae | Christensenellaceae R-7 group | NA |
| Levodopa | 98 | ASV867 | ASV | Firmicutes | NA | NA | NA |
| Levodopa | 99 | ASV203 | ASV | Firmicutes | NA | NA | NA |
| Levodopa | 100 | ASV150 | ASV | Proteobacteria | Sutterellaceae | Sutterella | NA |
| Levodopa | 101 | ASV132 | ASV | Firmicutes | Ruminococcaceae | CAG-352 | NA |
| Levodopa | 102 | ASV681 | ASV | Firmicutes | Oscillospiraceae | NA | NA |
| Levodopa | 103 | ASV50 | ASV | Bacteroidota | Prevotellaceae | Prevotella | NA |
| Levodopa | 104 | ASV673 | ASV | Firmicutes | Christensenellaceae | Christensenellaceae R-7 group | NA |
| Levodopa | 105 | ASV69 | ASV | Firmicutes | Christensenellaceae | NA | NA |
| Levodopa | 106 | ASV201 | ASV | Firmicutes | Oscillospiraceae | UCG-002 | NA |
| Levodopa | 107 | ASV322 | ASV | Firmicutes | Anaerovoracaceae | Family XIII AD3011 group | NA |
| Levodopa | 108 | ASV52 | ASV | Firmicutes | Lachnospiraceae | Coprococcus | eutactus |
| Levodopa | 109 | ASV512 | ASV | Proteobacteria | Oxalobacteraceae | Oxalobacter | formigenes |
| Levodopa | 110 | ASV81 | ASV | Firmicutes | Lachnospiraceae | Lachnospiraceae NK4A136 group | NA |
| Levodopa | 111 | ASV218 | ASV | Firmicutes | Anaerovoracaceae | Family XIII AD3011 group | NA |
| Levodopa | 112 | ASV128 | ASV | Firmicutes | Oscillospiraceae | NK4A214 group | NA |
| Levodopa | 113 | ASV338 | ASV | Firmicutes | Lachnospiraceae | Lachnotalea | NA |
| Levodopa | 114 | ASV533 | ASV | Firmicutes | Ruminococcaceae | Negativibacillus | massiliensis |
| Levodopa | 115 | ASV143 | ASV | Firmicutes | Peptostreptococcaceae | Terrisporobacter | mayombei |
| Levodopa | 116 | ASV4 | ASV | Firmicutes | Ruminococcaceae | Ruminococcus | bromii |
| Levodopa | 117 | ASV29 | ASV | Firmicutes | Oscillospiraceae | UCG-002 | NA |
| Levodopa | 118 | ASV124 | ASV | Firmicutes | NA | NA | NA |
| Levodopa | 119 | ASV59 | ASV | Firmicutes | [Eubacterium] coprostanoligenes group | NA | NA |

| Ticker | DESeq2_baseMean | DESeq2_log2-FoldChange | DESeq2_lfcSE | DESeq2_stat | DESeq2_pvalue | DESeq2_padj | DESeq2 converged |
|---|---|---|---|---|---|---|---|
| 90 | 11,23850092 | -0,000622737 | 0,002566589 | -0,242632245 | 0,8082903 | 0,999413296 | YES |
| 91 | 76,68153906 | -0,000350461 | 0,001454047 | -0,241024345 | 0,809536248 | 0,999413296 | YES |
| 92 | 58,46974618 | -0,000341567 | 0,00142282 | -0,240063014 | 0,810281406 | 0,999413296 | YES |
| 93 | 68,35691256 | -0,001482013 | 0,006183688 | -0,239664913 | 0,810590038 | 0,999413296 | YES |
| 94 | 13,36751846 | -0,00094794 | 0,004027732 | -0,235353189 | 0,813934616 | 0,999413296 | YES |
| 95 | 12,31047961 | -0,001452715 | 0,006181031 | -0,235028019 | 0,814186986 | 0,999413296 | YES |
| 96 | 135,8232871 | -0,000739908 | 0,003167636 | -0,23358377 | 0,815308129 | 0,999413296 | YES |
| 97 | 5,638992417 | 0,001291816 | 0,005545523 | 0,232947636 | 0,815802067 | 0,999413296 | YES |
| 98 | 4,357255709 | 0,001003476 | 0,004326088 | 0,231959315 | 0,816569614 | 0,999413296 | YES |
| 99 | 34,35032394 | -0,001403532 | 0,006182595 | -0,227013335 | 0,820413373 | 0,999413296 | YES |
| 100 | 23,06248383 | -0,000781663 | 0,003447868 | -0,226709204 | 0,820649869 | 0,999413296 | YES |
| 101 | 29,67807042 | -0,001376501 | 0,006191054 | -0,2223371 | 0,824051474 | 0,999413296 | YES |
| 102 | 8,130540982 | -0,001343768 | 0,006188171 | -0,217151126 | 0,828090572 | 0,999413296 | YES |
| 103 | 45,54055903 | -0,001334522 | 0,006188224 | -0,215655061 | 0,829256633 | 0,999413296 | YES |
| 104 | 7,198649845 | 0,001325115 | 0,006180004 | 0,214419811 | 0,830219694 | 0,999413296 | YES |
| 105 | 88,23778836 | 0,000362621 | 0,001706156 | 0,212536703 | 0,831688347 | 0,999413296 | YES |
| 106 | 29,39239845 | 0,001310383 | 0,006173668 | 0,212253552 | 0,83190923 | 0,999413296 | YES |
| 107 | 30,22859143 | -0,000325866 | 0,001572876 | -0,207178596 | 0,835870383 | 0,999413296 | YES |
| 108 | 291,6553856 | -0,001000784 | 0,004906269 | -0,20398059 | 0,838368667 | 0,999413296 | YES |
| 109 | 13,73103523 | -0,00028316 | 0,001423967 | -0,198852702 | 0,842377969 | 0,999413296 | YES |
| 110 | 100,0852058 | -0,000864883 | 0,004350946 | -0,198780348 | 0,842434569 | 0,999413296 | YES |
| 111 | 7,697372797 | -0,0008328 | 0,004310926 | -0,193183603 | 0,84681518 | 0,999413296 | YES |
| 112 | 107,8757466 | 0,000408607 | 0,00212485 | 0,192299161 | 0,847507876 | 0,999413296 | YES |
| 113 | 3,92166846 | -0,001161062 | 0,006187629 | -0,187642458 | 0,85115694 | 0,999413296 | YES |
| 114 | 9,056758508 | 0,001155402 | 0,006182753 | 0,186875075 | 0,851758582 | 0,999413296 | YES |
| 115 | 7,13841624 | 0,001152299 | 0,006182603 | 0,186377681 | 0,852148594 | 0,999413296 | YES |
| 116 | 1141,850194 | 0,000890934 | 0,004802253 | 0,185524077 | 0,852817998 | 0,999413296 | YES |
| 117 | 631,2624844 | -0,000665605 | 0,003595505 | -0,185121356 | 0,853133852 | 0,999413296 | YES |
| 118 | 36,79873362 | 0,001112707 | 0,006187836 | 0,17982174 | 0,857292516 | 0,999413296 | YES |
| 119 | 164,9695729 | -0,000784989 | 0,004478727 | -0,175270676 | 0,860866952 | 0,999413296 | YES |

| Medication | Ticker | ASVnr | taxLevel | Phylum | Family | Genus | Species |
|---|---|---|---|---|---|---|---|
| Levodopa | 120 | ASV18 | ASV | Firmicutes | Lachnospiraceae | Dorea | longicatena |
| Levodopa | 121 | ASV485 | ASV | Firmicutes | Lachnospiraceae | NA | NA |
| Levodopa | 122 | ASV820 | ASV | Firmicutes | Lachnospiraceae | GCA-900066575 | NA |
| Levodopa | 123 | ASV154 | ASV | Firmicutes | NA | NA | NA |
| Levodopa | 124 | ASV243 | ASV | Firmicutes | NA | NA | NA |
| Levodopa | 125 | ASV310 | ASV | Firmicutes | Lachnospiraceae | NA | NA |
| Levodopa | 126 | ASV337 | ASV | Firmicutes | Lachnospiraceae | Lachnoclostridium | NA |
| Levodopa | 127 | ASV61 | ASV | Firmicutes | Erysipelatoclostridiaceae | Catenibacterium | mitsuokai |
| Levodopa | 128 | ASV320 | ASV | Firmicutes | Christensenellaceae | Christensenellaceae R-7 group | NA |
| Levodopa | 129 | ASV51 | ASV | Firmicutes | Oscillospiraceae | UCG-005 | NA |
| Levodopa | 130 | ASV470 | ASV | Firmicutes | Christensenellaceae | Christensenellaceae R-7 group | NA |
| Levodopa | 131 | ASV760 | ASV | Firmicutes | UCG-010 | NA | NA |
| Levodopa | 132 | ASV84 | ASV | Bacteroidota | Bacteroidaceae | Bacteroides | NA |
| Levodopa | 133 | ASV172 | ASV | Proteobacteria | Pasteurellaceae | Haemophilus | NA |
| Levodopa | 134 | ASV160 | ASV | Firmicutes | Ruminococcaceae | Ruminococcus | callidus |
| Levodopa | 135 | ASV417 | ASV | Firmicutes | UCG-010 | NA | NA |
| Levodopa | 136 | ASV393 | ASV | Bacteroidota | Rikenellaceae | Alistipes | indistinctus |
| Levodopa | 137 | ASV441 | ASV | Firmicutes | [Eubacterium] coprostanoligenes group | NA | NA |
| Levodopa | 138 | ASV161 | ASV | Firmicutes | Oscillospiraceae | UCG-002 | NA |
| Levodopa | 139 | ASV587 | ASV | Firmicutes | Anaerovoracaceae | Family XIII AD3011 group | NA |
| Levodopa | 140 | ASV305 | ASV | Firmicutes | Lachnospiraceae | Lachnospiraceae UCG-004 | NA |
| Levodopa | 141 | ASV15 | ASV | Firmicutes | Christensenellaceae | Christensenellaceae R-7 group | NA |
| Levodopa | 142 | ASV130 | ASV | Proteobacteria | Enterobacteriaceae | Klebsiella | NA |
| Levodopa | 143 | ASV303 | ASV | Firmicutes | Lachnospiraceae | GCA-900066575 | NA |
| Levodopa | 144 | ASV194 | ASV | Bacteroidota | Bacteroidaceae | Bacteroides | intestinalis |
| Levodopa | 145 | ASV308 | ASV | Firmicutes | Christensenellaceae | NA | NA |
| Levodopa | 146 | ASV49 | ASV | Bacteroidota | Bacteroidaceae | Bacteroides | NA |
| Levodopa | 147 | ASV41 | ASV | Firmicutes | Ruminococcaceae | Ruminococcus | bicirculans |
| Levodopa | 148 | ASV261 | ASV | Firmicutes | Anaerovoracaceae | Family XIII UCG-001 | NA |
| Levodopa | 149 | ASV57 | ASV | Firmicutes | Lachnospiraceae | Roseburia | inulinivorans |

| Ticker | DESeq2_baseMean | DESeq2_log2-FoldChange | DESeq2_lfcSE | DESeq2_stat | DESeq2_pvalue | DESeq2_padj | DESeq2 converged |
|---|---|---|---|---|---|---|---|
| 120 | 526,8625731 | -0,00048416 | 0,002762729 | -0,175247105 | 0,860885473 | 0,999413296 | YES |
| 121 | 4,052568483 | -0,001077215 | 0,006183358 | -0,174212033 | 0,86169883 | 0,999413296 | YES |
| 122 | 3,236851239 | 0,001068146 | 0,006184147 | 0,172723239 | 0,862868976 | 0,999413296 | YES |
| 123 | 22,58783515 | -0,001066941 | 0,006191314 | -0,172328627 | 0,86317918 | 0,999413296 | YES |
| 124 | 16,60882207 | -0,001057814 | 0,006179422 | -0,17118334 | 0,864079607 | 0,999413296 | YES |
| 125 | 8,651357003 | -0,001054015 | 0,006192363 | -0,170212148 | 0,864843299 | 0,999413296 | YES |
| 126 | 14,93406139 | -0,000468561 | 0,002775288 | -0,168833361 | 0,865927716 | 0,999413296 | YES |
| 127 | 67,01747497 | -0,001044825 | 0,006188534 | -0,168832394 | 0,865928477 | 0,999413296 | YES |
| 128 | 28,60355617 | 0,000288264 | 0,001732716 | 0,166365199 | 0,867869559 | 0,999413296 | YES |
| 129 | 302,0586381 | -0,000379346 | 0,002320144 | -0,16350114 | 0,870123876 | 0,999413296 | YES |
| 130 | 5,565362966 | 0,001009584 | 0,006181301 | 0,163328764 | 0,870259587 | 0,999413296 | YES |
| 131 | 6,197101774 | 0,000990055 | 0,0061775 | 0,160267953 | 0,872670002 | 0,999413296 | YES |
| 132 | 38,42992757 | 0,000958591 | 0,006020834 | 0,159212335 | 0,873501586 | 0,999413296 | YES |
| 133 | 8,543927258 | -0,00095392 | 0,006007535 | -0,1587872 | 0,873836534 | 0,999413296 | YES |
| 134 | 115,3116721 | -0,000977202 | 0,006183353 | -0,158037583 | 0,874427184 | 0,999413296 | YES |
| 135 | 20,03444959 | 0,000782967 | 0,00495471 | 0,158024732 | 0,87443731 | 0,999413296 | YES |
| 136 | 6,162876947 | 0,000725574 | 0,004611803 | 0,15732975 | 0,874984975 | 0,999413296 | YES |
| 137 | 11,04850758 | 0,000593818 | 0,003820843 | 0,155415458 | 0,876493799 | 0,999413296 | YES |
| 138 | 14,2452233 | 0,000936289 | 0,006178285 | 0,151545156 | 0,879545693 | 0,999413296 | YES |
| 139 | 7,920860219 | 0,000935293 | 0,006186881 | 0,151173537 | 0,879838824 | 0,999413296 | YES |
| 140 | 9,88773165 | -0,000918125 | 0,006187608 | -0,148381212 | 0,882041927 | 0,999413296 | YES |
| 141 | 637,6403702 | 0,000416764 | 0,002828847 | 0,147326406 | 0,882874391 | 0,999413296 | YES |
| 142 | 62,02797969 | 0,000887934 | 0,006191169 | 0,143419365 | 0,88595899 | 0,999413296 | YES |
| 143 | 7,912403662 | -0,000710055 | 0,004951104 | -0,143413535 | 0,885963594 | 0,999413296 | YES |
| 144 | 73,19466546 | 0,00088242 | 0,006186256 | 0,142642039 | 0,886572895 | 0,999413296 | YES |
| 145 | 12,76608642 | 0,000877943 | 0,006180992 | 0,142039163 | 0,887049072 | 0,999413296 | YES |
| 146 | 138,5637744 | -0,000722519 | 0,005097536 | -0,141738928 | 0,887286226 | 0,999413296 | YES |
| 147 | 199,656055 | -0,000286798 | 0,002067344 | -0,138727612 | 0,889665397 | 0,999413296 | YES |
| 148 | 8,730686227 | 0,000843949 | 0,00618905 | 0,136361703 | 0,891535348 | 0,999413296 | YES |
| 149 | 153,9674257 | -0,000552494 | 0,004086275 | -0,135207304 | 0,892447972 | 0,999413296 | YES |

| Medication | Ticker | ASVnr | taxLevel | Phylum | Family | Genus | Species |
|---|---|---|---|---|---|---|---|
| Levodopa | 150 | ASV141 | ASV | Firmicutes | Lachnospiraceae | Lachnoclostridium | NA |
| Levodopa | 151 | ASV83 | ASV | Firmicutes | Lachnospiraceae | Lachnospiraceae NK4A136 group | NA |
| Levodopa | 152 | ASV554 | ASV | Firmicutes | Lachnospiraceae | NA | NA |
| Levodopa | 153 | ASV443 | ASV | Firmicutes | Veillonellaceae | Veillonella | NA |
| Levodopa | 154 | ASV207 | ASV | Firmicutes | Lachnospiraceae | Blautia | NA |
| Levodopa | 155 | ASV307 | ASV | Firmicutes | Lachnospiraceae | Lachnoclostridium | NA |
| Levodopa | 156 | ASV196 | ASV | Firmicutes | Ruminococcaceae | Ruminococcus | NA |
| Levodopa | 157 | ASV836 | ASV | Verrucomicrobiota | Victivallaceae | Victivallis | vadensis |
| Levodopa | 158 | ASV93 | ASV | Firmicutes | Oscillospiraceae | NK4A214 group | NA |
| Levodopa | 159 | ASV103 | ASV | Firmicutes | NA | NA | NA |
| Levodopa | 160 | ASV113 | ASV | Firmicutes | Lachnospiraceae | NA | NA |
| Levodopa | 161 | ASV508 | ASV | Firmicutes | Ruminococcaceae | NA | NA |
| Levodopa | 162 | ASV134 | ASV | Actinobacteriota | Eggerthellaceae | Enterorhabdus | NA |
| Levodopa | 163 | ASV593 | ASV | Firmicutes | Lachnospiraceae | NA | NA |
| Levodopa | 164 | ASV630 | ASV | Firmicutes | Ruminococcaceae | Angelakisella | NA |
| Levodopa | 165 | ASV213 | ASV | Firmicutes | NA | NA | NA |
| Levodopa | 166 | ASV328 | ASV | Firmicutes | Anaerovoracaceae | [Eubacterium] brachy group | NA |
| Levodopa | 167 | ASV1158 | ASV | Firmicutes | Ruminococcaceae | NA | NA |
| Levodopa | 168 | ASV271 | ASV | Firmicutes | Oscillospiraceae | NK4A214 group | NA |
| Levodopa | 169 | ASV386 | ASV | Firmicutes | Ruminococcaceae | DTU089 | NA |
| Levodopa | 170 | ASV184 | ASV | Firmicutes | Ruminococcaceae | Ruminococcus | NA |
| Levodopa | 171 | ASV413 | ASV | Firmicutes | Oscillospiraceae | Oscillibacter | NA |
| Levodopa | 172 | ASV230 | ASV | Firmicutes | Ruminococcaceae | [Eubacterium] siraeum group | NA |
| Levodopa | 173 | ASV215 | ASV | Firmicutes | Oscillospiraceae | UCG-002 | NA |
| Levodopa | 174 | ASV606 | ASV | Firmicutes | Lachnospiraceae | NA | NA |
| Levodopa | 175 | ASV162 | ASV | Desulfobacterota | Desulfovibrionaceae | Desulfovibrio | NA |
| Levodopa | 176 | ASV351 | ASV | Firmicutes | Lachnospiraceae | Roseburia | NA |
| Levodopa | 177 | ASV27 | ASV | Firmicutes | Ruminococcaceae | CAG-352 | NA |
| Levodopa | 178 | ASV703 | ASV | Firmicutes | Hydrogenoanaerobacterium | NA | NA |
| Levodopa | 179 | ASV249 | ASV | Proteobacteria | NA | NA | NA |

| Ticker | DESeq2_baseMean | DESeq2_log2-FoldChange | DESeq2_lfcSE | DESeq2_stat | DESeq2_pvalue | DESeq2_padj | DESeq2 converged |
|---|---|---|---|---|---|---|---|
| 150 | 51,77060175 | -0,000244006 | 0,001810035 | -0,134807334 | 0,892764207 | 0,999413296 | YES |
| 151 | 52,16535846 | -0,000557141 | 0,004172012 | -0,133542613 | 0,893764266 | 0,999413296 | YES |
| 152 | 11,45333846 | 0,000817427 | 0,006180358 | 0,13226208 | 0,894777 | 0,999413296 | YES |
| 153 | 9,382984409 | 0,000812031 | 0,006179132 | 0,131415061 | 0,895446975 | 0,999413296 | YES |
| 154 | 62,35934014 | 0,000749031 | 0,0057284 | 0,130757491 | 0,895967152 | 0,999413296 | YES |
| 155 | 4,134367288 | 0,00080501 | 0,006184687 | 0,130161877 | 0,896438355 | 0,999413296 | YES |
| 156 | 47,69546804 | 0,000789661 | 0,006186155 | 0,127649788 | 0,898426128 | 0,999413296 | YES |
| 157 | 1,493976754 | 0,000785656 | 0,00618956 | 0,126932423 | 0,898993884 | 0,999413296 | YES |
| 158 | 104,516405 | 0,000414684 | 0,003285874 | 0,126202027 | 0,899572007 | 0,999413296 | YES |
| 159 | 256,2142209 | -0,000779783 | 0,006184472 | -0,12608716 | 0,899662931 | 0,999413296 | YES |
| 160 | 197,448331 | -0,000777388 | 0,006183644 | -0,125716861 | 0,899956054 | 0,999413296 | YES |
| 161 | 2,805795971 | -0,000765776 | 0,006191799 | -0,123675851 | 0,901571933 | 0,999413296 | YES |
| 162 | 7,454505117 | 0,000307756 | 0,002493935 | 0,12340198 | 0,901788789 | 0,999413296 | YES |
| 163 | 5,872426435 | 0,000759676 | 0,006188285 | 0,122760387 | 0,902296843 | 0,999413296 | YES |
| 164 | 1,756793572 | 0,000745159 | 0,006181949 | 0,120537945 | 0,904057023 | 0,999413296 | YES |
| 165 | 9,600944567 | 0,000743275 | 0,006184759 | 0,120178563 | 0,904341699 | 0,999413296 | YES |
| 166 | 0,554566966 | 0,000743034 | 0,006193312 | 0,11997356 | 0,904504093 | 0,999413296 | YES |
| 167 | 1,638703655 | -0,000742363 | 0,006188296 | -0,119962374 | 0,904512954 | 0,999413296 | YES |
| 168 | 42,70925267 | 0,000537086 | 0,004514854 | 0,118959845 | 0,905307169 | 0,999413296 | YES |
| 169 | 7,625424248 | 0,000501729 | 0,004242914 | 0,118251028 | 0,905868759 | 0,999413296 | YES |
| 170 | 66,43328135 | -0,00071967 | 0,006178538 | -0,116478988 | 0,907272938 | 0,999413296 | YES |
| 171 | 2,121682783 | 0,000716764 | 0,006194646 | 0,115707015 | 0,907884747 | 0,999413296 | YES |
| 172 | 32,01737841 | -0,000589057 | 0,00511511 | -0,115160231 | 0,90831812 | 0,999413296 | YES |
| 173 | 165,5478588 | 0,000707061 | 0,006179584 | 0,114418916 | 0,908905719 | 0,999413296 | YES |
| 174 | 17,93930574 | 0,000707765 | 0,006193501 | 0,114275494 | 0,909019408 | 0,999413296 | YES |
| 175 | 164,5704062 | 0,000211939 | 0,001857769 | 0,114082624 | 0,909172296 | 0,999413296 | YES |
| 176 | 20,64397058 | -0,000693071 | 0,006078716 | -0,114016073 | 0,909225052 | 0,999413296 | YES |
| 177 | 348,6736511 | -0,000696067 | 0,006167904 | -0,112853113 | 0,910147009 | 0,999413296 | YES |
| 178 | 2,721956617 | -0,000694283 | 0,006184298 | -0,112265472 | 0,910612918 | 0,999413296 | YES |
| 179 | 24,01745867 | 0,000691362 | 0,006191523 | 0,111662591 | 0,911090942 | 0,999413296 | YES |

| Medication | Ticker | ASVnr | taxLevel | Phylum | Family | Genus | Species |
|---|---|---|---|---|---|---|---|
| Levodopa | 180 | ASV666 | ASV | Firmicutes | Erysipelotrichaceae | Holdemania | filiformis |
| Levodopa | 181 | ASV808 | ASV | Desulfobacterota | Desulfovibrionaceae | NA | NA |
| Levodopa | 182 | ASV270 | ASV | Firmicutes | Christensenellaceae | Christensenellaceae R-7 group | NA |
| Levodopa | 183 | ASV937 | ASV | Firmicutes | UCG-010 | NA | NA |
| Levodopa | 184 | ASV185 | ASV | Firmicutes | NA | NA | NA |
| Levodopa | 185 | ASV120 | ASV | Firmicutes | Lachnospiraceae | Anaerostipes | hadrus |
| Levodopa | 186 | ASV140 | ASV | Bacteroidota | Tannerellaceae | Parabacteroides | distasonis |
| Levodopa | 187 | ASV58 | ASV | Bacteroidota | Bacteroidaceae | Bacteroides | NA |
| Levodopa | 188 | ASV242 | ASV | Firmicutes | Lachnospiraceae | Tyzzerella | NA |
| Levodopa | 189 | ASV256 | ASV | Firmicutes | Lachnospiraceae | Blautia | NA |
| Levodopa | 190 | ASV529 | ASV | Firmicutes | Lachnospiraceae | NA | NA |
| Levodopa | 191 | ASV363 | ASV | Bacteroidota | Rikenellaceae | Alistipes | NA |
| Levodopa | 192 | ASV78 | ASV | Firmicutes | Lachnospiraceae | [Eubacterium] eligens group | NA |
| Levodopa | 193 | ASV231 | ASV | Proteobacteria | NA | NA | NA |
| Levodopa | 194 | ASV517 | ASV | Firmicutes | Lachnospiraceae | Blautia | NA |
| Levodopa | 195 | ASV457 | ASV | Firmicutes | Lachnospiraceae | NA | NA |
| Levodopa | 196 | ASV155 | ASV | Firmicutes | Ruminococcaceae | Ruminococcus | NA |
| Levodopa | 197 | ASV522 | ASV | Firmicutes | UCG-010 | NA | NA |
| Levodopa | 198 | ASV12 | ASV | Firmicutes | Veillonellaceae | Dialister | invisus |
| Levodopa | 199 | ASV98 | ASV | Firmicutes | Acidaminococcaceae | Phascolarctobacterium | succinatutens |
| Levodopa | 200 | ASV195 | ASV | Bacteroidota | Barnesiellaceae | Barnesiella | intestinihominis |
| Levodopa | 201 | ASV450 | ASV | Firmicutes | NA | NA | NA |
| Levodopa | 202 | ASV477 | ASV | Firmicutes | Oscillospiraceae | Intestinimonas | massiliensis |
| Levodopa | 203 | ASV225 | ASV | Bacteroidota | Muribaculaceae | NA | NA |
| Levodopa | 204 | ASV459 | ASV | Bacteroidota | Marinifilaceae | Butyricimonas | paravirosa |
| Levodopa | 205 | ASV216 | ASV | Firmicutes | Lachnospiraceae | Lachnospira | pectinoschiza |
| Levodopa | 206 | ASV131 | ASV | Firmicutes | Lachnospiraceae | NA | NA |
| Levodopa | 207 | ASV97 | ASV | Firmicutes | Lachnospiraceae | [Ruminococcus] gnavus group | NA |
| Levodopa | 208 | ASV557 | ASV | Firmicutes | Oscillospiraceae | UCG-005 | NA |
| Levodopa | 209 | ASV16 | ASV | Firmicutes | Peptostreptococcaceae | Romboutsia | NA |

| Ticker | DESeq2_baseMean | DESeq2_log2-FoldChange | DESeq2_lfcSE | DESeq2_stat | DESeq2_pvalue | DESeq2_padj | DESeq2 converged |
|---|---|---|---|---|---|---|---|
| 180 | 1,219516932 | -0,000689632 | 0,006184647 | -0,111507038 | 0,911214285 | 0,999413296 | YES |
| 181 | 1,725099267 | 0,000676025 | 0,00618353 | 0,109326649 | 0,91294341 | 0,999413296 | YES |
| 182 | 7,869461939 | 0,000675803 | 0,006182 | 0,109317881 | 0,912950364 | 0,999413296 | YES |
| 183 | 4,561937719 | -0,000674763 | 0,006186227 | -0,109075061 | 0,913142956 | 0,999413296 | YES |
| 184 | 68,78971624 | -0,000673546 | 0,006185571 | -0,10888983 | 0,913289873 | 0,999413296 | YES |
| 185 | 9,332851223 | 0,000671162 | 0,006184761 | 0,10851865 | 0,913584287 | 0,999413296 | YES |
| 186 | 45,6144019 | 0,000475966 | 0,004398355 | 0,108214665 | 0,913825412 | 0,999413296 | YES |
| 187 | 151,8729563 | -0,000461958 | 0,004325427 | -0,106800663 | 0,914947122 | 0,999413296 | YES |
| 188 | 72,61139852 | 0,00052152 | 0,004896248 | 0,106514205 | 0,915174386 | 0,999413296 | YES |
| 189 | 9,246885654 | 0,000657603 | 0,006177005 | 0,106459898 | 0,915217471 | 0,999413296 | YES |
| 190 | 5,034264887 | -0,000652418 | 0,006176923 | -0,105621873 | 0,915882369 | 0,999413296 | YES |
| 191 | 8,9381144 | 0,000647089 | 0,006183838 | 0,104642007 | 0,916659881 | 0,999413296 | YES |
| 192 | 101,7224426 | -0,000387278 | 0,003703095 | -0,104582164 | 0,916707368 | 0,999413296 | YES |
| 193 | 18,57610302 | 0,000644977 | 0,006186251 | 0,104259687 | 0,916963269 | 0,999413296 | YES |
| 194 | 14,3654402 | 0,000637006 | 0,006195788 | 0,102812777 | 0,918111564 | 0,999413296 | YES |
| 195 | 4,728764316 | -0,000623088 | 0,006189434 | -0,100669555 | 0,91981278 | 0,999413296 | YES |
| 196 | 25,12366646 | -0,000620775 | 0,006187225 | -0,100331708 | 0,920080985 | 0,999413296 | YES |
| 197 | 6,437604221 | 0,000595154 | 0,005952666 | 0,099981059 | 0,920359362 | 0,999413296 | YES |
| 198 | 276,5548356 | 0,000464065 | 0,004706041 | 0,098610379 | 0,921447629 | 0,999413296 | YES |
| 199 | 185,5716817 | -0,000356988 | 0,003621922 | -0,098563093 | 0,921485175 | 0,999413296 | YES |
| 200 | 56,00017742 | -0,000599388 | 0,006179311 | -0,096999138 | 0,922727079 | 0,999413296 | YES |
| 201 | 5,002586655 | -0,000597294 | 0,006178949 | -0,096665942 | 0,922991687 | 0,999413296 | YES |
| 202 | 8,649431688 | 0,000596158 | 0,006181414 | 0,096443687 | 0,923168197 | 0,999413296 | YES |
| 203 | 67,31078843 | 0,000589896 | 0,0061865 | 0,095352063 | 0,92403519 | 0,999413296 | YES |
| 204 | 6,96757681 | 0,000580379 | 0,006191163 | 0,093743109 | 0,925313225 | 0,999413296 | YES |
| 205 | 23,3952391 | -0,000332194 | 0,003551089 | -0,093547131 | 0,925468909 | 0,999413296 | YES |
| 206 | 215,6612668 | -0,000295646 | 0,003187243 | -0,092759204 | 0,926094862 | 0,999413296 | YES |
| 207 | 55,700982 | 0,000573466 | 0,006185733 | 0,092707808 | 0,926135694 | 0,999413296 | YES |
| 208 | 30,91172048 | 0,000573201 | 0,006195279 | 0,092522297 | 0,926283077 | 0,999413296 | YES |
| 209 | 38,42899062 | -0,000341901 | 0,003744518 | -0,09130714 | 0,927248545 | 0,999413296 | YES |

| Medication | Ticker | ASVnr | taxLevel | Phylum | Family | Genus | Species |
|---|---|---|---|---|---|---|---|
| Levodopa | 210 | ASV535 | ASV | Firmicutes | Erysipelatoclostridiaceae | Asteroleplasma | NA |
| Levodopa | 211 | ASV326 | ASV | Proteobacteria | NA | NA | NA |
| Levodopa | 212 | ASV453 | ASV | Verrucomicrobiota | Victivallaceae | Victivallis | NA |
| Levodopa | 213 | ASV147 | ASV | Firmicutes | Oscillospiraceae | Oscillibacter | NA |
| Levodopa | 214 | ASV317 | ASV | Firmicutes | NA | NA | NA |
| Levodopa | 215 | ASV806 | ASV | Firmicutes | Peptococcaceae | NA | NA |
| Levodopa | 216 | ASV295 | ASV | Firmicutes | Ruminococcaceae | Negativibacillus | NA |
| Levodopa | 217 | ASV193 | ASV | Actinobacteriota | Eggerthellaceae | Adlercreutzia | equolifaciens |
| Levodopa | 218 | ASV187 | ASV | Firmicutes | NA | NA | NA |
| Levodopa | 219 | ASV148 | ASV | Firmicutes | NA | NA | NA |
| Levodopa | 220 | ASV145 | ASV | Bacteroidota | Muribaculaceae | NA | NA |
| Levodopa | 221 | ASV879 | ASV | Firmicutes | NA | NA | NA |
| Levodopa | 222 | ASV142 | ASV | Firmicutes | Ruminococcaceae | Faecalibacterium | prausnitzii |
| Levodopa | 223 | ASV237 | ASV | Bacteroidota | Bacteroidaceae | Bacteroides | clarus |
| Levodopa | 224 | ASV23 | ASV | Firmicutes | Streptococcaceae | Streptococcus | NA |
| Levodopa | 225 | ASV153 | ASV | Firmicutes | Streptococcaceae | Streptococcus | NA |
| Levodopa | 226 | ASV625 | ASV | Firmicutes | UCG-010 | NA | NA |
| Levodopa | 227 | ASV620 | ASV | Firmicutes | UCG-010 | NA | NA |
| Levodopa | 228 | ASV480 | ASV | Firmicutes | UCG-010 | NA | NA |
| Levodopa | 229 | ASV206 | ASV | Firmicutes | Oscillospiraceae | Intestinimonas | NA |
| Levodopa | 230 | ASV314 | ASV | Firmicutes | Ruminococcaceae | Subdoligranulum | NA |
| Levodopa | 231 | ASV238 | ASV | Firmicutes | Butyricicoccaceae | NA | NA |
| Levodopa | 232 | ASV92 | ASV | Firmicutes | Erysipelotrichaceae | Turicibacter | sanguinis |
| Levodopa | 233 | ASV146 | ASV | Firmicutes | Erysipelotrichaceae | Turicibacter | sanguinis |
| Levodopa | 234 | ASV79 | ASV | Firmicutes | Lachnospiraceae | [Eubacterium] ruminantium group | NA |
| Levodopa | 235 | ASV54 | ASV | Bacteroidota | Bacteroidaceae | Bacteroides | NA |
| Levodopa | 236 | ASV536 | ASV | Bacteroidota | Marinifilaceae | Butyricimonas | NA |
| Levodopa | 237 | ASV414 | ASV | Firmicutes | Oscillospiraceae | UCG-003 | NA |
| Levodopa | 238 | ASV425 | ASV | Firmicutes | Oscillospiraceae | NK4A214 group | NA |
| Levodopa | 239 | ASV895 | ASV | Bacteroidota | Barnesiellaceae | Coprobacter | secundus |

| Ticker | DESeq2_baseMean | DESeq2_log2-FoldChange | DESeq2_lfcSE | DESeq2_stat | DESeq2_pvalue | DESeq2_padj | DESeq2 converged |
|---|---|---|---|---|---|---|---|
| 210 | 3,166917521 | -0,00056443 | 0,006189799 | -0,091187143 | 0,927343891 | 0,999413296 | YES |
| 211 | 14,90153933 | -0,000555611 | 0,006175786 | -0,089965991 | 0,928314241 | 0,999413296 | YES |
| 212 | 5,775539756 | -0,000544281 | 0,006176016 | -0,08812812 | 0,929774846 | 0,999413296 | YES |
| 213 | 37,25355638 | 0,00014269 | 0,001645077 | 0,086737343 | 0,930880293 | 0,999413296 | YES |
| 214 | 13,39252942 | 0,000535047 | 0,006174321 | 0,086656792 | 0,930944322 | 0,999413296 | YES |
| 215 | 1,253800201 | 0,000532479 | 0,006193335 | 0,08597606 | 0,931485448 | 0,999413296 | YES |
| 216 | 13,81514126 | 0,000525824 | 0,006134157 | 0,08572073 | 0,931688423 | 0,999413296 | YES |
| 217 | 9,054189399 | -0,000377204 | 0,004428183 | -0,085182499 | 0,932116304 | 0,999413296 | YES |
| 218 | 30,59069723 | -0,000525842 | 0,00619182 | -0,084925236 | 0,932320829 | 0,999413296 | YES |
| 219 | 57,36799493 | -0,000466461 | 0,005501102 | -0,084794113 | 0,932425074 | 0,999413296 | YES |
| 220 | 87,98895088 | -0,000523549 | 0,006180889 | -0,084704421 | 0,932496381 | 0,999413296 | YES |
| 221 | 1,922600258 | 0,000523314 | 0,00618731 | 0,084578588 | 0,932596422 | 0,999413296 | YES |
| 222 | 114,1341006 | -0,000334569 | 0,004131498 | -0,08098011 | 0,93545777 | 0,999413296 | YES |
| 223 | 27,19033597 | 0,000499704 | 0,00618913 | 0,080738964 | 0,935649549 | 0,999413296 | YES |
| 224 | 160,1730331 | 0,000243538 | 0,003018443 | 0,080683453 | 0,935693696 | 0,999413296 | YES |
| 225 | 15,96534437 | 0,000497356 | 0,00618134 | 0,080460828 | 0,93587075 | 0,999413296 | YES |
| 226 | 3,211174268 | 0,000494624 | 0,006190971 | 0,079894362 | 0,936321274 | 0,999413296 | YES |
| 227 | 19,61685753 | 0,000491949 | 0,006182261 | 0,079574312 | 0,936575826 | 0,999413296 | YES |
| 228 | 9,327913783 | 0,000490431 | 0,006184312 | 0,079302395 | 0,936792102 | 0,999413296 | YES |
| 229 | 29,59105329 | 0,000451096 | 0,005763961 | 0,078261531 | 0,937620018 | 0,999413296 | YES |
| 230 | 16,743465 | 0,000482069 | 0,006179126 | 0,078015663 | 0,937815594 | 0,999413296 | YES |
| 231 | 52,03263594 | 0,000224231 | 0,002884306 | 0,077741603 | 0,9380336 | 0,999413296 | YES |
| 232 | 30,82796885 | 0,000479028 | 0,006185376 | 0,07744524 | 0,938269353 | 0,999413296 | YES |
| 233 | 11,42916362 | 0,000467751 | 0,006176698 | 0,075728315 | 0,939635249 | 0,999413296 | YES |
| 234 | 181,6773233 | -0,000417602 | 0,005587892 | -0,07473345 | 0,940426793 | 0,999413296 | YES |
| 235 | 144,2814565 | 0,000309538 | 0,004156562 | 0,07446961 | 0,940636722 | 0,999413296 | YES |
| 236 | 12,31516585 | 0,000458996 | 0,006182464 | 0,074241639 | 0,940818114 | 0,999413296 | YES |
| 237 | 4,265605972 | 0,000455405 | 0,006190135 | 0,073569519 | 0,941352926 | 0,999413296 | YES |
| 238 | 22,75111542 | -0,000453095 | 0,006180614 | -0,073309126 | 0,94156013 | 0,999413296 | YES |
| 239 | 3,875087249 | -0,000450106 | 0,006183545 | -0,072790938 | 0,941972483 | 0,999413296 | YES |

| Medication | Ticker | ASVnr | taxLevel | Phylum | Family | Genus | Species |
|---|---|---|---|---|---|---|---|
| Levodopa | 240 | ASV247 | ASV | Euryarchaeota | Methanobacteriaceae | Methanosphaera | NA |
| Levodopa | 241 | ASV730 | ASV | Firmicutes | Ruminococcaceae | NA | NA |
| Levodopa | 242 | ASV262 | ASV | Firmicutes | Ruminococcaceae | Incertae Sedis | NA |
| Levodopa | 243 | ASV801 | ASV | Firmicutes | Christensenellaceae | Christensenellaceae R-7 group | NA |
| Levodopa | 244 | ASV324 | ASV | Proteobacteria | NA | NA | NA |
| Levodopa | 245 | ASV714 | ASV | Firmicutes | Oscillospiraceae | Oscillibacter | NA |
| Levodopa | 246 | ASV95 | ASV | Bacteroidota | Prevotellaceae | Prevotella | NA |
| Levodopa | 247 | ASV618 | ASV | Actinobacteriota | Atopobiaceae | NA | NA |
| Levodopa | 248 | ASV67 | ASV | Firmicutes | Erysipelotrichaceae | Holdemanella | biformis |
| Levodopa | 249 | ASV1086 | ASV | Firmicutes | UCG-010 | NA | NA |
| Levodopa | 250 | ASV133 | ASV | Actinobacteriota | Eggerthellaceae | Senegalimassilia | anaerobia |
| Levodopa | 251 | ASV353 | ASV | Firmicutes | Lachnospiraceae | [Eubacterium] xylanophilum group | NA |
| Levodopa | 252 | ASV653 | ASV | Firmicutes | Lachnospiraceae | NA | NA |
| Levodopa | 253 | ASV202 | ASV | Firmicutes | Lachnospiraceae | [Eubacterium] hallii group | NA |
| Levodopa | 254 | ASV217 | ASV | Firmicutes | Peptococcaceae | Peptococcus | NA |
| Levodopa | 255 | ASV106 | ASV | Bacteroidota | Bacteroidaceae | Bacteroides | NA |
| Levodopa | 256 | ASV509 | ASV | Firmicutes | Lachnospiraceae | Marvinbryantia | NA |
| Levodopa | 257 | ASV421 | ASV | Firmicutes | NA | NA | NA |
| Levodopa | 258 | ASV179 | ASV | Firmicutes | NA | NA | NA |
| Levodopa | 259 | ASV695 | ASV | Firmicutes | UCG-010 | NA | NA |
| Levodopa | 260 | ASV291 | ASV | Firmicutes | Erysipelatoclostridiaceae | Erysipelatoclostridium | NA |
| Levodopa | 261 | ASV597 | ASV | Firmicutes | Oscillospiraceae | Oscillibacter | NA |
| Levodopa | 262 | ASV727 | ASV | Firmicutes | Ruminococcaceae | Angelakisella | NA |
| Levodopa | 263 | ASV538 | ASV | Bacteroidota | Marinifilaceae | Butyricimonas | NA |
| Levodopa | 264 | ASV169 | ASV | Bacteroidota | Barnesiellaceae | Barnesiella | NA |
| Levodopa | 265 | ASV135 | ASV | Bacteroidota | Rikenellaceae | Alistipes | inops |
| Levodopa | 266 | ASV233 | ASV | Firmicutes | [Eubacterium] coprostanoligenes group | NA | NA |
| Levodopa | 267 | ASV582 | ASV | Bacteroidota | NA | NA | NA |
| Levodopa | 268 | ASV383 | ASV | Firmicutes | NA | NA | NA |
| Levodopa | 269 | ASV152 | ASV | Firmicutes | Lachnospiraceae | Roseburia | hominis |

| Ticker | DESeq2_baseMean | DESeq2_log2-FoldChange | DESeq2_lfcSE | DESeq2_stat | DESeq2_pvalue | DESeq2_padj | DESeq2 converged |
|---|---|---|---|---|---|---|---|
| 240 | 10,34906432 | 0,000446837 | 0,006187294 | 0,072218526 | 0,942428002 | 0,999413296 | YES |
| 241 | 2,152719414 | 0,000443023 | 0,006197384 | 0,071485473 | 0,943011385 | 0,999413296 | YES |
| 242 | 7,341634343 | 0,000441075 | 0,006184387 | 0,07132075 | 0,943142481 | 0,999413296 | YES |
| 243 | 2,293529572 | 0,000440444 | 0,006179371 | 0,071276543 | 0,943177664 | 0,999413296 | YES |
| 244 | 13,18180794 | 0,000428111 | 0,006181154 | 0,069260618 | 0,944782173 | 0,999413296 | YES |
| 245 | 5,558682084 | 0,000424934 | 0,00619317 | 0,068613278 | 0,94529745 | 0,999413296 | YES |
| 246 | 18,93040995 | -0,000424135 | 0,00619331 | -0,068482763 | 0,945401341 | 0,999413296 | YES |
| 247 | 1,81793053 | 0,000423923 | 0,006190657 | 0,06847786 | 0,945405243 | 0,999413296 | YES |
| 248 | 49,83996399 | -0,000423491 | 0,006185791 | -0,068461875 | 0,945417968 | 0,999413296 | YES |
| 249 | 1,26539883 | 0,00042254 | 0,006193324 | 0,068225087 | 0,945606457 | 0,999413296 | YES |
| 250 | 19,55149837 | 0,000116354 | 0,001709923 | 0,068046115 | 0,945748925 | 0,999413296 | YES |
| 251 | 9,948554992 | -0,000419196 | 0,006169492 | -0,067946647 | 0,945828106 | 0,999413296 | YES |
| 252 | 7,072394613 | -0,00041199 | 0,006192452 | -0,066530991 | 0,946955085 | 0,999413296 | YES |
| 253 | 12,27098395 | -0,000407569 | 0,006179551 | -0,065954399 | 0,947414131 | 0,999413296 | YES |
| 254 | 44,56053753 | 0,000406667 | 0,006183773 | 0,065763647 | 0,947565999 | 0,999413296 | YES |
| 255 | 46,37553392 | 0,00035285 | 0,005373743 | 0,065661856 | 0,947647041 | 0,999413296 | YES |
| 256 | 7,050817322 | 0,000405153 | 0,006184987 | 0,065505882 | 0,947771223 | 0,999413296 | YES |
| 257 | 26,44148839 | -0,000399359 | 0,006181165 | -0,064609022 | 0,948485301 | 0,999413296 | YES |
| 258 | 37,65647991 | 0,000397063 | 0,006176572 | 0,064285284 | 0,948743071 | 0,999413296 | YES |
| 259 | 4,254865218 | -0,000388017 | 0,006182443 | -0,062761172 | 0,949956685 | 0,999413296 | YES |
| 260 | 1,948383656 | 0,000384848 | 0,00618651 | 0,062207671 | 0,950397454 | 0,999413296 | YES |
| 261 | 10,26691315 | -0,000324246 | 0,005239814 | -0,061881125 | 0,950657499 | 0,999413296 | YES |
| 262 | 2,641995519 | 0,000382016 | 0,006181323 | 0,061801698 | 0,950720751 | 0,999413296 | YES |
| 263 | 6,374459177 | 0,000381689 | 0,006181143 | 0,061750494 | 0,950761529 | 0,999413296 | YES |
| 264 | 50,95215862 | -0,000379334 | 0,006190132 | -0,0612805 | 0,95113582 | 0,999413296 | YES |
| 265 | 64,03685612 | 0,00029564 | 0,004869743 | 0,06070947 | 0,951590589 | 0,999413296 | YES |
| 266 | 16,72179855 | 0,000374583 | 0,006195565 | 0,060459924 | 0,951789334 | 0,999413296 | YES |
| 267 | 3,547781421 | 0,000372425 | 0,006193503 | 0,060131497 | 0,952050905 | 0,999413296 | YES |
| 268 | 29,78158179 | 0,000370573 | 0,006186082 | 0,059904364 | 0,952231804 | 0,999413296 | YES |
| 269 | 55,16735837 | -0,000153067 | 0,002578719 | -0,059357755 | 0,95266716 | 0,999413296 | YES |

| Medication | Ticker | ASVnr | taxLevel | Phylum | Family | Genus | Species |
|---|---|---|---|---|---|---|---|
| Levodopa | 270 | ASV44 | ASV | Bacteroidota | Bacteroidaceae | Bacteroides | NA |
| Levodopa | 271 | ASV1155 | ASV | Firmicutes | UCG-010 | NA | NA |
| Levodopa | 272 | ASV1464 | ASV | Firmicutes | Peptococcaceae | NA | NA |
| Levodopa | 273 | ASV244 | ASV | Bacteroidota | Bacteroidaceae | Bacteroides | coprophilus |
| Levodopa | 274 | ASV286 | ASV | Firmicutes | Lachnospiraceae | NA | NA |
| Levodopa | 275 | ASV9 | ASV | Bacteroidota | Bacteroidaceae | Bacteroides | NA |
| Levodopa | 276 | ASV214 | ASV | Firmicutes | Lachnospiraceae | Lachnospiraceae UCG-010 | NA |
| Levodopa | 277 | ASV292 | ASV | Firmicutes | [Eubacterium] coprostanoligenes group | NA | NA |
| Levodopa | 278 | ASV400 | ASV | Bacteroidota | Rikenellaceae | Rikenellaceae RC9 gut group | NA |
| Levodopa | 279 | ASV171 | ASV | Firmicutes | Oscillospiraceae | NA | NA |
| Levodopa | 280 | ASV176 | ASV | Bacteroidota | Prevotellaceae | Paraprevotella | NA |
| Levodopa | 281 | ASV156 | ASV | Bacteroidota | Prevotellaceae | Prevotella | NA |
| Levodopa | 282 | ASV355 | ASV | Firmicutes | NA | NA | NA |
| Levodopa | 283 | ASV376 | ASV | Bacteroidota | Barnesiellaceae | Coprobacter | fastidiosus |
| Levodopa | 284 | ASV454 | ASV | Actinobacteriota | Actinomycetaceae | Actinomyces | odontolyticus |
| Levodopa | 285 | ASV110 | ASV | Firmicutes | Ruminococcaceae | Faecalibacterium | prausnitzii |
| Levodopa | 286 | ASV422 | ASV | Firmicutes | Lachnospiraceae | Lachnospiraceae FCS020 group | bacterium |
| Levodopa | 287 | ASV726 | ASV | Verrucomicrobiota | vadinBE97 | NA | NA |
| Levodopa | 288 | ASV221 | ASV | Firmicutes | Oscillospiraceae | Intestinimonas | NA |
| Levodopa | 289 | ASV254 | ASV | Firmicutes | Lachnospiraceae | Lachnospira | NA |
| Levodopa | 290 | ASV55 | ASV | Firmicutes | Monoglobaceae | Monoglobus | NA |
| Levodopa | 291 | ASV813 | ASV | Firmicutes | Ruminococcaceae | NA | NA |
| Levodopa | 292 | ASV223 | ASV | Bacteroidota | Barnesiellaceae | Barnesiella | NA |
| Levodopa | 293 | ASV423 | ASV | Firmicutes | Christensenellaceae | Christensenellaceae R-7 group | NA |
| Levodopa | 294 | ASV373 | ASV | Firmicutes | NA | NA | NA |
| Levodopa | 295 | ASV591 | ASV | Firmicutes | UCG-010 | NA | NA |
| Levodopa | 296 | ASV772 | ASV | Firmicutes | Ruminococcaceae | Fournierella | NA |
| Levodopa | 297 | ASV499 | ASV | Firmicutes | Lachnospiraceae | Eisenbergiella | tayi |
| Levodopa | 298 | ASV894 | ASV | Desulfobacterota | Desulfovibrionaceae | NA | NA |
| Levodopa | 299 | ASV435 | ASV | Firmicutes | Christensenellaceae | Christensenellaceae R-7 group | NA |

| Ticker | DESeq2_baseMean | DESeq2_log2-FoldChange | DESeq2_lfcSE | DESeq2_stat | DESeq2_pvalue | DESeq2_padj | DESeq2 converged |
|---|---|---|---|---|---|---|---|
| 270 | 40,25734808 | -0,000366252 | 0,006175634 | -0,059305984 | 0,952708395 | 0,999413296 | YES |
| 271 | 3,02499994 | -0,000364889 | 0,006188711 | -0,05896041 | 0,952983642 | 0,999413296 | YES |
| 272 | 1,555478949 | 0,000363706 | 0,006181316 | 0,058839626 | 0,953079846 | 0,999413296 | YES |
| 273 | 21,11198514 | 0,000362208 | 0,00619067 | 0,058508709 | 0,953343426 | 0,999413296 | YES |
| 274 | 27,71648061 | -0,000357526 | 0,006187946 | -0,057777828 | 0,953925599 | 0,999413296 | YES |
| 275 | 661,8643954 | -0,000179212 | 0,00315993 | -0,056714018 | 0,954773007 | 0,999413296 | YES |
| 276 | 25,70293356 | -0,000349014 | 0,006181667 | -0,056459486 | 0,954975769 | 0,999413296 | YES |
| 277 | 42,35700937 | 0,000347461 | 0,006190196 | 0,056130898 | 0,95523753 | 0,999413296 | YES |
| 278 | 25,91588437 | 0,000326482 | 0,005821898 | 0,05607835 | 0,955279391 | 0,999413296 | YES |
| 279 | 15,72556168 | 0,000343609 | 0,006180311 | 0,055597307 | 0,95566261 | 0,999413296 | YES |
| 280 | 40,13409453 | 0,000343217 | 0,006183155 | 0,055508321 | 0,955733501 | 0,999413296 | YES |
| 281 | 24,01537456 | 0,000342001 | 0,006181737 | 0,055324405 | 0,95588002 | 0,999413296 | YES |
| 282 | 18,11799983 | -0,000342352 | 0,006192547 | -0,055284527 | 0,955911789 | 0,999413296 | YES |
| 283 | 7,537075577 | -0,000268456 | 0,004892915 | -0,054866223 | 0,956245041 | 0,999413296 | YES |
| 284 | 0,88183281 | 0,000338479 | 0,006194362 | 0,054643102 | 0,9564228 | 0,999413296 | YES |
| 285 | 50,51980185 | -0,000332191 | 0,00619292 | -0,053640483 | 0,957221602 | 0,999413296 | YES |
| 286 | 5,488811908 | 0,000329363 | 0,00619113 | 0,053199227 | 0,957573172 | 0,999413296 | YES |
| 287 | 1,801031007 | 0,000327518 | 0,006184832 | 0,052955072 | 0,957767705 | 0,999413296 | YES |
| 288 | 31,52528109 | 7,04E-05 | 0,001342064 | 0,052487728 | 0,958140074 | 0,999413296 | YES |
| 289 | 34,19366839 | -0,000312781 | 0,00596186 | -0,052463717 | 0,958159205 | 0,999413296 | YES |
| 290 | 78,31427244 | 0,000150217 | 0,002866806 | 0,05239868 | 0,958211026 | 0,999413296 | YES |
| 291 | 1,295754344 | 0,000322439 | 0,006188973 | 0,052098918 | 0,958449875 | 0,999413296 | YES |
| 292 | 45,82330375 | 0,000313671 | 0,006194675 | 0,050635645 | 0,959615859 | 0,999413296 | YES |
| 293 | 4,933840203 | -0,000311461 | 0,006175994 | -0,050430986 | 0,959778944 | 0,999413296 | YES |
| 294 | 17,72672335 | 0,000306813 | 0,006185791 | 0,049599639 | 0,960441435 | 0,999413296 | YES |
| 295 | 7,495680267 | 0,000304243 | 0,006187016 | 0,049174404 | 0,96078031 | 0,999413296 | YES |
| 296 | 7,879044875 | 0,000302512 | 0,006190889 | 0,048864049 | 0,96102764 | 0,999413296 | YES |
| 297 | 7,024069658 | 0,00030261 | 0,00619444 | 0,048851891 | 0,961037328 | 0,999413296 | YES |
| 298 | 1,374876307 | 0,000302639 | 0,006195684 | 0,048846781 | 0,961041401 | 0,999413296 | YES |
| 299 | 12,68864099 | 0,000189547 | 0,00392618 | 0,048277605 | 0,961495002 | 0,999413296 | YES |

| Medication | Ticker | ASVnr | taxLevel | Phylum | Family | Genus | Species |
|---|---|---|---|---|---|---|---|
| Levodopa | 300 | ASV234 | ASV | Cyanobacteria | NA | NA | NA |
| Levodopa | 301 | ASV675 | ASV | Bacteroidota | Prevotellaceae | NA | NA |
| Levodopa | 302 | ASV112 | ASV | Firmicutes | Oscillospiraceae | UCG-002 | NA |
| Levodopa | 303 | ASV200 | ASV | Firmicutes | Lachnospiraceae | [Eubacterium] xylanophilum group | NA |
| Levodopa | 304 | ASV60 | ASV | Firmicutes | Ruminococcaceae | [Eubacterium] siraeum group | NA |
| Levodopa | 305 | ASV343 | ASV | Firmicutes | Lachnospiraceae | NA | NA |
| Levodopa | 306 | ASV255 | ASV | Actinobacteriota | Coriobacteriales Incertae Sedis | NA | NA |
| Levodopa | 307 | ASV446 | ASV | Firmicutes | Oscillospiraceae | UCG-005 | NA |
| Levodopa | 308 | ASV329 | ASV | Firmicutes | Lachnospiraceae | NA | NA |
| Levodopa | 309 | ASV360 | ASV | Firmicutes | [Eubacterium] coprostanoligenes group | NA | NA |
| Levodopa | 310 | ASV191 | ASV | Firmicutes | Lachnospiraceae | Lachnospiraceae NK4A136 group | NA |
| Levodopa | 311 | ASV687 | ASV | Firmicutes | Ruminococcaceae | DTU089 | NA |
| Levodopa | 312 | ASV294 | ASV | Firmicutes | Anaerovoracaceae | Mogibacterium | NA |
| Levodopa | 313 | ASV91 | ASV | Firmicutes | Lachnospiraceae | [Eubacterium] eligens group | NA |
| Levodopa | 314 | ASV189 | ASV | Firmicutes | Ruminococcaceae | Ruminococcus | champanellensis |
| Levodopa | 315 | ASV380 | ASV | Firmicutes | Christensenellaceae | Christensenellaceae R-7 group | NA |
| Levodopa | 316 | ASV107 | ASV | Bacteroidota | Bacteroidaceae | Bacteroides | eggerthii |
| Levodopa | 317 | ASV665 | ASV | Firmicutes | Lachnospiraceae | NA | NA |
| Levodopa | 318 | ASV62 | ASV | Firmicutes | NA | NA | NA |
| Levodopa | 319 | ASV752 | ASV | Verrucomicrobiota | Puniceicoccaceae | NA | NA |
| Levodopa | 320 | ASV347 | ASV | Bacteroidota | Tannerellaceae | Parabacteroides | NA |
| Levodopa | 321 | ASV507 | ASV | Firmicutes | Oscillospiraceae | Colidextribacter | NA |
| Levodopa | 322 | ASV190 | ASV | Firmicutes | Ruminococcaceae | UBA1819 | NA |
| Levodopa | 323 | ASV299 | ASV | Firmicutes | Streptococcaceae | Lactococcus | NA |
| Levodopa | 324 | ASV520 | ASV | Firmicutes | UCG-010 | NA | NA |
| Levodopa | 325 | ASV339 | ASV | Firmicutes | Monoglobaceae | Monoglobus | pectinilyticus |
| Levodopa | 326 | ASV208 | ASV | Bacteroidota | Prevotellaceae | Prevotella | NA |
| Levodopa | 327 | ASV167 | ASV | Firmicutes | Oscillospiraceae | Intestinimonas | NA |
| Levodopa | 328 | ASV174 | ASV | Actinobacteriota | Coriobacteriales Incertae Sedis | NA | NA |
| Levodopa | 329 | ASV22 | ASV | Bacteroidota | Bacteroidaceae | Bacteroides | uniformis |

| Ticker | DESeq2_baseMean | DESeq2_log2-FoldChange | DESeq2_lfcSE | DESeq2_stat | DESeq2_pvalue | DESeq2_padj | DESeq2 converged |
|---|---|---|---|---|---|---|---|
| 300 | 10,56994452 | -0,0002978 | 0,006175829 | -0,04822018 | 0,961540767 | 0,999413296 | YES |
| 301 | 5,961837949 | 0,000285667 | 0,006187331 | 0,046169623 | 0,963175054 | 0,999413296 | YES |
| 302 | 28,85511555 | 0,00028198 | 0,006187832 | 0,045570021 | 0,963652964 | 0,999413296 | YES |
| 303 | 23,50210386 | -0,000199125 | 0,004395375 | -0,045303277 | 0,963865576 | 0,999413296 | YES |
| 304 | 115,647046 | 0,000163099 | 0,003620901 | 0,045043673 | 0,964072498 | 0,999413296 | YES |
| 305 | 2,705995867 | 0,000275308 | 0,006192728 | 0,044456695 | 0,96454037 | 0,999413296 | YES |
| 306 | 5,952421825 | 0,000274732 | 0,006187944 | 0,044397876 | 0,964587255 | 0,999413296 | YES |
| 307 | 13,76928275 | -0,000272705 | 0,006188181 | -0,04406864 | 0,964849691 | 0,999413296 | YES |
| 308 | 3,227624179 | -0,00027015 | 0,006187581 | -0,043660042 | 0,965175391 | 0,999413296 | YES |
| 309 | 22,52490053 | 0,000268504 | 0,006191712 | 0,043365018 | 0,965410563 | 0,999413296 | YES |
| 310 | 9,32575453 | 0,000266152 | 0,006185116 | 0,043031072 | 0,965676765 | 0,999413296 | YES |
| 311 | 3,018838706 | 0,000263939 | 0,006193946 | 0,042612359 | 0,966010544 | 0,999413296 | YES |
| 312 | 24,16835401 | 0,000262867 | 0,006183923 | 0,042508076 | 0,966093674 | 0,999413296 | YES |
| 313 | 79,09621748 | 0,000173454 | 0,004111075 | 0,042191926 | 0,966345699 | 0,999413296 | YES |
| 314 | 53,85886049 | 0,000260717 | 0,006197702 | 0,042066732 | 0,966445501 | 0,999413296 | YES |
| 315 | 3,42743513 | 0,000257179 | 0,006178472 | 0,041625097 | 0,966797566 | 0,999413296 | YES |
| 316 | 30,40443022 | -0,000256012 | 0,00618806 | -0,041371942 | 0,96699938 | 0,999413296 | YES |
| 317 | 9,362214561 | 0,000255662 | 0,006188433 | 0,041312942 | 0,967046415 | 0,999413296 | YES |
| 318 | 184,8812144 | -0,000237607 | 0,005820693 | -0,040821025 | 0,967438578 | 0,999413296 | YES |
| 319 | 4,171374726 | 0,000251286 | 0,006187501 | 0,040611891 | 0,967605304 | 0,999413296 | YES |
| 320 | 32,52636146 | 0,000250664 | 0,006192427 | 0,040479078 | 0,967711187 | 0,999413296 | YES |
| 321 | 10,98952418 | -0,000233993 | 0,006173904 | -0,037900334 | 0,969767147 | 0,999413296 | YES |
| 322 | 26,66292816 | -0,000102658 | 0,002718158 | -0,037767305 | 0,969873213 | 0,999413296 | YES |
| 323 | 8,470308428 | 0,000223329 | 0,006183815 | 0,036115142 | 0,971190548 | 0,999413296 | YES |
| 324 | 7,768918053 | 0,000219048 | 0,00618552 | 0,035412973 | 0,97175044 | 0,999413296 | YES |
| 325 | 9,771837182 | -0,000217727 | 0,006187007 | -0,035190938 | 0,971927488 | 0,999413296 | YES |
| 326 | 7,239848411 | 0,000213378 | 0,006190871 | 0,034466619 | 0,972505061 | 0,999413296 | YES |
| 327 | 36,744968 | -6,02E-05 | 0,001777068 | -0,033879335 | 0,972973372 | 0,999413296 | YES |
| 328 | 3,275222135 | 0,000195366 | 0,006192031 | 0,031551158 | 0,974829994 | 0,999413296 | YES |
| 329 | 339,2459582 | 0,000110552 | 0,00351951 | 0,031411107 | 0,974941683 | 0,999413296 | YES |

| Medication | Ticker | ASVnr | taxLevel | Phylum | Family | Genus | Species |
|---|---|---|---|---|---|---|---|
| Levodopa | 330 | ASV481 | ASV | Firmicutes | Lachnospiraceae | [Eubacterium] xylanophilum group | NA |
| Levodopa | 331 | ASV611 | ASV | Firmicutes | Lachnospiraceae | Lachnospiraceae UCG-010 | NA |
| Levodopa | 332 | ASV797 | ASV | Firmicutes | Peptococcaceae | Peptococcus | NA |
| Levodopa | 333 | ASV126 | ASV | Bacteroidota | Tannerellaceae | Parabacteroides | distasonis |
| Levodopa | 334 | ASV444 | ASV | Firmicutes | Erysipelatoclostridiaceae | Coprobacillus | cateniformis |
| Levodopa | 335 | ASV437 | ASV | Firmicutes | Christensenellaceae | Christensenellaceae R-7 group | NA |
| Levodopa | 336 | ASV288 | ASV | Firmicutes | Erysipelotrichaceae | NA | NA |
| Levodopa | 337 | ASV828 | ASV | Firmicutes | Christensenellaceae | Christensenellaceae R-7 group | NA |
| Levodopa | 338 | ASV72 | ASV | Proteobacteria | Sutterellaceae | Parasutterella | excrementihominis |
| Levodopa | 339 | ASV151 | ASV | Firmicutes | Butyricicoccaceae | Butyricicoccus | NA |
| Levodopa | 340 | ASV432 | ASV | Bacteroidota | Marinifilaceae | Butyricimonas | virosa |
| Levodopa | 341 | ASV505 | ASV | Bacteroidota | Bacteroidaceae | Bacteroides | nordii |
| Levodopa | 342 | ASV335 | ASV | Bacteroidota | Barnesiellaceae | NA | NA |
| Levodopa | 343 | ASV248 | ASV | Firmicutes | Ruminococcaceae | Subdoligranulum | NA |
| Levodopa | 344 | ASV252 | ASV | Firmicutes | Erysipelotrichaceae | Solobacterium | NA |
| Levodopa | 345 | ASV306 | ASV | Firmicutes | Veillonellaceae | Veillonella | NA |
| Levodopa | 346 | ASV114 | ASV | Bacteroidota | Bacteroidaceae | Bacteroides | coprocola |
| Levodopa | 347 | ASV503 | ASV | Firmicutes | Erysipelatoclostridiaceae | Erysipelatoclostridium | NA |
| Levodopa | 348 | ASV341 | ASV | Firmicutes | Christensenellaceae | Christensenellaceae R-7 group | NA |
| Levodopa | 349 | ASV284 | ASV | Firmicutes | Oscillospiraceae | NK4A214 group | NA |
| Levodopa | 350 | ASV205 | ASV | Firmicutes | NA | NA | NA |
| Levodopa | 351 | ASV530 | ASV | Firmicutes | Veillonellaceae | Allisonella | histaminiformans |
| Levodopa | 352 | ASV209 | ASV | Bacteroidota | Tannerellaceae | NA | NA |
| Levodopa | 353 | ASV573 | ASV | Firmicutes | Erysipelatoclostridiaceae | Erysipelatoclostridium | NA |
| Levodopa | 354 | ASV260 | ASV | Firmicutes | Lactobacillaceae | Lactobacillus | delbrueckii |
| Levodopa | 355 | ASV521 | ASV | Firmicutes | Lachnospiraceae | [Eubacterium] ventriosum group | NA |
| Levodopa | 356 | ASV885 | ASV | Firmicutes | Lachnospiraceae | Shuttleworthia | NA |
| Levodopa | 357 | ASV615 | ASV | Firmicutes | Oscillospiraceae | Oscillibacter | NA |
| Levodopa | 358 | ASV449 | ASV | Actinobacteriota | Atopobiaceae | Olsenella | NA |
| Levodopa | 359 | ASV149 | ASV | Firmicutes | Oscillospiraceae | UCG-002 | NA |

| Ticker | DESeq2_baseMean | DESeq2_log2-FoldChange | DESeq2_lfcSE | DESeq2_stat | DESeq2_pvalue | DESeq2_padj | DESeq2 converged |
|---|---|---|---|---|---|---|---|
| 330 | 8,325782108 | 0,000194515 | 0,006193363 | 0,031406984 | 0,974944971 | 0,999413296 | YES |
| 331 | 3,055112192 | 0,000192368 | 0,006197318 | 0,031040583 | 0,975237175 | 0,999413296 | YES |
| 332 | 4,546450581 | -0,000191112 | 0,006190627 | -0,030871127 | 0,975372316 | 0,999413296 | YES |
| 333 | 79,46445507 | 0,000102617 | 0,003348458 | 0,030646046 | 0,97555182 | 0,999413296 | YES |
| 334 | 8,923422661 | 0,00018947 | 0,006184132 | 0,030638155 | 0,975558113 | 0,999413296 | YES |
| 335 | 5,804203834 | 0,000183278 | 0,006186563 | 0,029625237 | 0,976365938 | 0,999413296 | YES |
| 336 | 16,83887458 | -0,000174601 | 0,006095948 | -0,028642134 | 0,977150008 | 0,999413296 | YES |
| 337 | 1,033120643 | 0,000175479 | 0,006190827 | 0,028345052 | 0,977386949 | 0,999413296 | YES |
| 338 | 52,42523433 | 5,68E-05 | 0,002013797 | 0,028188641 | 0,977511696 | 0,999413296 | YES |
| 339 | 52,62220893 | 6,72E-05 | 0,0023959 | 0,028052296 | 0,977620441 | 0,999413296 | YES |
| 340 | 7,24003739 | -0,000170467 | 0,006179221 | -0,027587104 | 0,977991467 | 0,999413296 | YES |
| 341 | 5,386847909 | 0,000170211 | 0,006195712 | 0,027472429 | 0,97808293 | 0,999413296 | YES |
| 342 | 21,92437397 | -0,000114499 | 0,004246772 | -0,026961442 | 0,978490488 | 0,999413296 | YES |
| 343 | 12,30986784 | -0,000166408 | 0,006181804 | -0,026918978 | 0,978524357 | 0,999413296 | YES |
| 344 | 18,9083293 | 0,000165615 | 0,006188671 | 0,026761033 | 0,978650333 | 0,999413296 | YES |
| 345 | 10,79563595 | -0,000162797 | 0,006179141 | -0,026346203 | 0,978981203 | 0,999413296 | YES |
| 346 | 54,90178431 | -0,000162818 | 0,006194385 | -0,026284845 | 0,979030143 | 0,999413296 | YES |
| 347 | 4,75739345 | -0,000158503 | 0,00619133 | -0,02560074 | 0,979575796 | 0,999413296 | YES |
| 348 | 8,220221102 | 0,000154004 | 0,006177937 | 0,024928029 | 0,98011237 | 0,999413296 | YES |
| 349 | 8,575826931 | -0,000149692 | 0,00618445 | -0,02420462 | 0,980689393 | 0,999413296 | YES |
| 350 | 36,01031628 | -0,000146982 | 0,006193172 | -0,023732991 | 0,98106559 | 0,999413296 | YES |
| 351 | 8,770524285 | -0,000142651 | 0,006181195 | -0,023078236 | 0,981587867 | 0,999413296 | YES |
| 352 | 63,88848232 | 0,00014199 | 0,006189003 | 0,022942383 | 0,981696233 | 0,999413296 | YES |
| 353 | 0,388289001 | 0,000142105 | 0,006194793 | 0,02293948 | 0,981698548 | 0,999413296 | YES |
| 354 | 8,552884092 | -0,000141455 | 0,006194861 | -0,022834294 | 0,981782452 | 0,999413296 | YES |
| 355 | 2,86496118 | 0,000138148 | 0,006192513 | 0,022308829 | 0,982201606 | 0,999413296 | YES |
| 356 | 2,894780216 | -0,00013063 | 0,006191885 | -0,021096923 | 0,98316834 | 0,999413296 | YES |
| 357 | 0,505963203 | -0,000120659 | 0,006188104 | -0,019498465 | 0,984443461 | 0,999413296 | YES |
| 358 | 4,677784239 | -0,000112644 | 0,006181592 | -0,018222452 | 0,985461392 | 0,999413296 | YES |
| 359 | 86,06796294 | -0,000109546 | 0,006068872 | -0,018050437 | 0,985598617 | 0,999413296 | YES |

| Medication | Ticker | ASVnr | taxLevel | Phylum | Family | Genus | Species |
|---|---|---|---|---|---|---|---|
| Levodopa | 360 | ASV240 | ASV | Bacteroidota | Prevotellaceae | Alloprevotella | NA |
| Levodopa | 361 | ASV82 | ASV | Firmicutes | Lachnospiraceae | Coprococcus | eutactus |
| Levodopa | 362 | ASV709 | ASV | Firmicutes | Lachnospiraceae | NA | NA |
| Levodopa | 363 | ASV175 | ASV | Firmicutes | NA | NA | NA |
| Levodopa | 364 | ASV121 | ASV | Bacteroidota | Barnesiellaceae | Barnesiella | NA |
| Levodopa | 365 | ASV656 | ASV | Firmicutes | Oscillospiraceae | Flavonifractor | NA |
| Levodopa | 366 | ASV427 | ASV | Firmicutes | Lactobacillaceae | Lactobacillus | NA |
| Levodopa | 367 | ASV40 | ASV | Firmicutes | Erysipelotrichaceae | Holdemanella | NA |
| Levodopa | 368 | ASV433 | ASV | Firmicutes | UCG-010 | NA | NA |
| Levodopa | 369 | ASV546 | ASV | Firmicutes | Lachnospiraceae | NA | NA |
| Levodopa | 370 | ASV504 | ASV | Firmicutes | Christensenellaceae | Christensenellaceae R-7 group | NA |
| Levodopa | 371 | ASV277 | ASV | Bacteroidota | Rikenellaceae | Alistipes | ihumii |
| Levodopa | 372 | ASV460 | ASV | Firmicutes | NA | NA | NA |
| Levodopa | 373 | ASV819 | ASV | Firmicutes | Oscillospiraceae | Colidextribacter | NA |
| Levodopa | 374 | ASV367 | ASV | Bacteroidota | Rikenellaceae | Alistipes | NA |
| Levodopa | 375 | ASV448 | ASV | Firmicutes | Christensenellaceae | Christensenellaceae R-7 group | NA |
| Levodopa | 376 | ASV1030 | ASV | Firmicutes | NA | NA | NA |
| Levodopa | 377 | ASV438 | ASV | Firmicutes | UCG-010 | NA | NA |
| Levodopa | 378 | ASV258 | ASV | Firmicutes | Lachnospiraceae | Lachnospiraceae AC2044 group | NA |
| Levodopa | 379 | ASV332 | ASV | Firmicutes | Lachnospiraceae | [Eubacterium] ventriosum group | NA |
| Levodopa | 380 | ASV759 | ASV | Firmicutes | NA | NA | NA |
| Levodopa | 381 | ASV253 | ASV | Bacteroidota | Tannerellaceae | Parabacteroides | distasonis |
| Levodopa | 382 | ASV164 | ASV | Bacteroidota | Tannerellaceae | Parabacteroides | distasonis |
| Levodopa | 383 | ASV494 | ASV | Firmicutes | Anaerovoracaceae | Family XIII AD3011 group | NA |
| Levodopa | 384 | ASV99 | ASV | Bacteroidota | Bacteroidaceae | Bacteroides | plebeius |
| Levodopa | 385 | ASV38 | ASV | Firmicutes | Clostridiaceae | Clostridium sensu stricto 1 | NA |
| Levodopa | 386 | ASV80 | ASV | Firmicutes | Acidaminococcaceae | Phascolarctobacterium | faecium |
| Levodopa | 387 | ASV198 | ASV | Bacteroidota | Bacteroidaceae | Bacteroides | eggerthii |
| Levodopa | 388 | ASV296 | ASV | Proteobacteria | Sutterellaceae | Sutterella | NA |
| Levodopa | 389 | ASV75 | ASV | Firmicutes | Lachnospiraceae | [Ruminococcus] gauvreauii group | NA |

| Ticker | DESeq2_baseMean | DESeq2_log2-FoldChange | DESeq2_lfcSE | DESeq2_stat | DESeq2_pvalue | DESeq2_padj | DESeq2 converged |
|---|---|---|---|---|---|---|---|
| 360 | 110,1361143 | 0,000111611 | 0,00618707 | 0,018039329 | 0,985607478 | 0,999413296 | YES |
| 361 | 119,8655212 | -0,000110072 | 0,006170703 | -0,017837789 | 0,985768259 | 0,999413296 | YES |
| 362 | 3,696366896 | 0,00010653 | 0,006184626 | 0,017224902 | 0,986257196 | 0,999413296 | YES |
| 363 | 25,94509867 | 0,000105245 | 0,006173004 | 0,017049244 | 0,98639733 | 0,999413296 | YES |
| 364 | 37,59825428 | 0,00010125 | 0,006196571 | 0,016339649 | 0,986963427 | 0,999413296 | YES |
| 365 | 0,930872367 | -9,90E-05 | 0,006194369 | -0,015989885 | 0,987242461 | 0,999413296 | YES |
| 366 | 9,990301438 | 9,84E-05 | 0,006188005 | 0,015905524 | 0,987309763 | 0,999413296 | YES |
| 367 | 223,2780932 | 3,60E-05 | 0,002265975 | 0,015900086 | 0,987314102 | 0,999413296 | YES |
| 368 | 15,37727118 | 9,68E-05 | 0,00617786 | 0,015675294 | 0,987493437 | 0,999413296 | YES |
| 369 | 11,48671386 | -9,67E-05 | 0,006186626 | -0,015633229 | 0,987526996 | 0,999413296 | YES |
| 370 | 3,183859222 | -9,65E-05 | 0,006189611 | -0,015593712 | 0,987558523 | 0,999413296 | YES |
| 371 | 14,3535372 | -9,33E-05 | 0,006092643 | -0,015315018 | 0,987780861 | 0,999413296 | YES |
| 372 | 5,371350046 | -8,52E-05 | 0,00619529 | -0,013748852 | 0,989030349 | 0,999413296 | YES |
| 373 | 1,608697539 | 7,98E-05 | 0,006193283 | 0,01288382 | 0,989720484 | 0,999413296 | YES |
| 374 | 16,41997627 | 7,54E-05 | 0,006180396 | 0,012202203 | 0,990264293 | 0,999413296 | YES |
| 375 | 18,63459123 | 7,19E-05 | 0,006183303 | 0,011624541 | 0,990725167 | 0,999413296 | YES |
| 376 | 1,13833772 | 6,78E-05 | 0,006186075 | 0,010956039 | 0,991258521 | 0,999413296 | YES |
| 377 | 10,03455377 | -6,48E-05 | 0,006178158 | -0,010485687 | 0,991633786 | 0,999413296 | YES |
| 378 | 74,80869052 | -3,25E-05 | 0,003140907 | -0,010346298 | 0,991744996 | 0,999413296 | YES |
| 379 | 12,47942199 | 6,32E-05 | 0,006181147 | 0,010222035 | 0,991844138 | 0,999413296 | YES |
| 380 | 8,554994248 | -6,32E-05 | 0,006191295 | -0,010210487 | 0,991853352 | 0,999413296 | YES |
| 381 | 30,01116406 | 6,20E-05 | 0,006189921 | 0,010019313 | 0,992005879 | 0,999413296 | YES |
| 382 | 27,56044501 | -6,20E-05 | 0,006193155 | -0,01000648 | 0,992016117 | 0,999413296 | YES |
| 383 | 2,806398495 | 6,16E-05 | 0,006186998 | 0,009954694 | 0,992057434 | 0,999413296 | YES |
| 384 | 40,62062215 | 6,07E-05 | 0,006193816 | 0,009793577 | 0,992185981 | 0,999413296 | YES |
| 385 | 9,162474875 | 5,64E-05 | 0,006181632 | 0,009119816 | 0,99272354 | 0,999413296 | YES |
| 386 | 91,33627835 | -5,53E-05 | 0,006190329 | -0,008926054 | 0,992878134 | 0,999413296 | YES |
| 387 | 101,360087 | 5,17E-05 | 0,006195208 | 0,008341363 | 0,993344632 | 0,999413296 | YES |
| 388 | 19,24472386 | -5,02E-05 | 0,006179492 | -0,008129735 | 0,993513482 | 0,999413296 | YES |
| 389 | 34,87997267 | -2,90E-05 | 0,004078156 | -0,007121743 | 0,994317719 | 0,999413296 | YES |

| Medication | Ticker | ASVnr | taxLevel | Phylum | Family | Genus | Species |
|---|---|---|---|---|---|---|---|
| Levodopa | 390 | ASV251 | ASV | Firmicutes | Oscillospiraceae | NK4A214 group | NA |
| Levodopa | 391 | ASV272 | ASV | Firmicutes | Erysipelotrichaceae | Catenisphaera | NA |
| Levodopa | 392 | ASV87 | ASV | Proteobacteria | Sutterellaceae | Sutterella | wadsworthensis |
| Levodopa | 393 | ASV118 | ASV | Firmicutes | NA | NA | NA |
| Levodopa | 394 | ASV10 | ASV | Verrucomicrobiota | Akkermansiaceae | Akkermansia | muciniphila |
| Levodopa | 395 | ASV385 | ASV | Firmicutes | Selenomonadaceae | Mitsuokella | jalaludinii |
| Levodopa | 396 | ASV690 | ASV | Firmicutes | Ruminococcaceae | Fournierella | NA |
| Levodopa | 397 | ASV42 | ASV | Actinobacteriota | Bifidobacteriaceae | Bifidobacterium | NA |
| Levodopa | 398 | ASV257 | ASV | Bacteroidota | Bacteroidaceae | Bacteroides | ovatus |
| Levodopa | 399 | ASV545 | ASV | Proteobacteria | Sutterellaceae | Sutterella | NA |
| Levodopa | 400 | ASV608 | ASV | Firmicutes | Lachnospiraceae | NA | NA |
| Levodopa | 401 | ASV232 | ASV | Firmicutes | Acholeplasmataceae | Anaeroplasma | NA |
| Levodopa | 402 | ASV382 | ASV | Firmicutes | NA | NA | NA |
| Levodopa | 403 | ASV323 | ASV | Desulfobacterota | Desulfovibrionaceae | Desulfovibrio | piger |
| Levodopa | 404 | ASV186 | ASV | Firmicutes | Lachnospiraceae | [Ruminococcus] torques group | NA |
| Levodopa | 405 | ASV129 | ASV | Firmicutes | Christensenellaceae | Christensenellaceae R-7 group | NA |
| Levodopa | 406 | ASV354 | ASV | Firmicutes | Ruminococcaceae | Negativibacillus | NA |
| Levodopa | 407 | ASV753 | ASV | Firmicutes | Ruminococcaceae | NA | NA |
| Levodopa | 408 | ASV534 | ASV | Firmicutes | Anaerovoracaceae | Family XIII UCG-001 | NA |
| Levodopa | 409 | ASV229 | ASV | Firmicutes | Oscillospiraceae | Flavonifractor | plautii |
| Levodopa | 410 | ASV250 | ASV | Bacteroidota | Prevotellaceae | Paraprevotella | NA |
| Levodopa | 411 | ASV241 | ASV | Firmicutes | [Eubacterium] coprostanoligenes group | NA | NA |
| Levodopa | 412 | ASV76 | Genus | Firmicutes | Lachnospiraceae | Lachnospiraceae ND3007 group | NA |
| Levodopa | 413 | ASV7 | Genus | Firmicutes | Lachnospiraceae | Agathobacter | NA |
| Levodopa | 414 | ASV144 | Genus | Desulfobacterota | Desulfovibrionaceae | Bilophila | NA |
| Levodopa | 415 | ASV96 | Genus | Firmicutes | Lachnospiraceae | CAG-56 | NA |
| Levodopa | 416 | ASV536 | Genus | Bacteroidota | Marinifilaceae | Butyricimonas | NA |
| Levodopa | 417 | ASV85 | Genus | Firmicutes | Lachnospiraceae | Coprococcus | NA |
| Levodopa | 418 | ASV195 | Genus | Bacteroidota | Barnesiellaceae | Barnesiella | NA |
| Levodopa | 419 | ASV65 | Genus | Firmicutes | Ruminococcaceae | Incertae Sedis | NA |

| Ticker | DESeq2_baseMean | DESeq2_log2-FoldChange | DESeq2_lfcSE | DESeq2_stat | DESeq2_pvalue | DESeq2_padj | DESeq2 converged |
|---|---|---|---|---|---|---|---|
| 390 | 15,84550348 | 4,24E-05 | 0,006181114 | 0,006863029 | 0,994524138 | 0,999413296 | YES |
| 391 | 2,3850677 | -3,63E-05 | 0,006193016 | -0,005867781 | 0,995318215 | 0,999413296 | YES |
| 392 | 41,68723249 | 2,55E-05 | 0,004506196 | 0,005654383 | 0,995488479 | 0,999413296 | YES |
| 393 | 398,3639327 | 3,49E-05 | 0,006180811 | 0,005642367 | 0,995498066 | 0,999413296 | YES |
| 394 | 1016,441608 | -2,18E-05 | 0,004101565 | -0,005307987 | 0,995764859 | 0,999413296 | YES |
| 395 | 19,59731936 | 3,28E-05 | 0,006188994 | 0,00530447 | 0,995767665 | 0,999413296 | YES |
| 396 | 8,946274281 | 3,15E-05 | 0,006195283 | 0,005079659 | 0,995947036 | 0,999413296 | YES |
| 397 | 45,15141618 | -3,02E-05 | 0,006194296 | -0,004870183 | 0,996114171 | 0,999413296 | YES |
| 398 | 12,87202367 | -2,68E-05 | 0,006192358 | -0,004326519 | 0,996547948 | 0,999413296 | YES |
| 399 | 2,523890074 | -2,53E-05 | 0,00618863 | -0,004090613 | 0,996736172 | 0,999413296 | YES |
| 400 | 4,914105369 | -2,43E-05 | 0,006189625 | -0,003928404 | 0,996865595 | 0,999413296 | YES |
| 401 | 34,67554047 | 2,30E-05 | 0,006193462 | 0,003709272 | 0,997040436 | 0,999413296 | YES |
| 402 | 20,82591292 | 2,05E-05 | 0,0061852 | 0,003317057 | 0,997353376 | 0,999413296 | YES |
| 403 | 42,22204903 | -1,73E-05 | 0,006193119 | -0,002799848 | 0,997766048 | 0,999413296 | YES |
| 404 | 27,80198025 | -1,43E-05 | 0,006188447 | -0,002308102 | 0,998158403 | 0,999413296 | YES |
| 405 | 85,01391949 | -2,92E-06 | 0,001323208 | -0,002207218 | 0,998238897 | 0,999413296 | YES |
| 406 | 11,61358952 | -1,12E-05 | 0,00618019 | -0,001817034 | 0,998550218 | 0,999413296 | YES |
| 407 | 5,988697938 | 1,12E-05 | 0,006186172 | 0,001804402 | 0,998560296 | 0,999413296 | YES |
| 408 | 7,625175334 | 7,58E-06 | 0,006190188 | 0,001225132 | 0,999022487 | 0,999413296 | YES |
| 409 | 5,424195007 | -6,84E-06 | 0,006171777 | -0,001108115 | 0,999115852 | 0,999413296 | YES |
| 410 | 25,80915858 | 6,81E-06 | 0,006190253 | 0,001100745 | 0,999121733 | 0,999413296 | YES |
| 411 | 9,596872634 | 4,55E-06 | 0,00619283 | 0,000735325 | 0,999413296 | 0,999413296 | YES |
| 412 | 70,42894352 | -0,00306688 | 0,000814292 | -3,766315563 | 0,000165674 | 0,021206319 | YES |
| 413 | 1903,075798 | -0,001819449 | 0,00059902 | -3,037374682 | 0,002386486 | 0,105046579 | YES |
| 414 | 59,72897528 | 0,001424381 | 0,000470408 | 3,027969518 | 0,002462029 | 0,105046579 | YES |
| 415 | 117,8471172 | -0,003256739 | 0,00116446 | -2,796781721 | 0,005161439 | 0,16516605 | YES |
| 416 | 48,10819097 | 0,001528146 | 0,000598513 | 2,553236024 | 0,010672715 | 0,273221493 | YES |
| 417 | 878,2528472 | -0,001267638 | 0,000552173 | -2,29572752 | 0,021691466 | 0,41186044 | YES |
| 418 | 256,6041676 | -0,001714296 | 0,000751416 | -2,281419794 | 0,022523618 | 0,41186044 | YES |
| 419 | 116,9962084 | 0,001110717 | 0,000551157 | 2,015245921 | 0,043878888 | 0,639021486 | YES |

| Medication | Ticker | ASVnr | taxLevel | Phylum | Family | Genus | Species |
|---|---|---|---|---|---|---|---|
| Levodopa | 420 | ASV111 | Genus | Firmicutes | Lachnospiraceae | Lachnospira | NA |
| Levodopa | 421 | ASV55 | Genus | Firmicutes | Monoglobaceae | Monoglobus | NA |
| Levodopa | 422 | ASV116 | Genus | Firmicutes | Oscillospiraceae | UCG-003 | NA |
| Levodopa | 423 | ASV211 | Genus | Firmicutes | Lachnospiraceae | Lachnospiraceae FCS020 group | NA |
| Levodopa | 424 | ASV28 | Genus | Firmicutes | Clostridiaceae | Clostridium sensu stricto 1 | NA |
| Levodopa | 425 | ASV295 | Genus | Firmicutes | Ruminococcaceae | Negativibacillus | NA |
| Levodopa | 426 | ASV19 | Genus | Firmicutes | Erysipelatoclostridiaceae | Erysipelotrichaceae UCG-003 | NA |
| Levodopa | 427 | ASV5 | Genus | Firmicutes | Lachnospiraceae | Blautia | NA |
| Levodopa | 428 | ASV309 | Genus | Firmicutes | Oscillospiraceae | Colidextribacter | NA |
| Levodopa | 429 | ASV219 | Genus | Firmicutes | Lachnospiraceae | Lachnospiraceae UCG-004 | NA |
| Levodopa | 430 | ASV51 | Genus | Firmicutes | Oscillospiraceae | UCG-005 | NA |
| Levodopa | 431 | ASV15 | Genus | Firmicutes | Christensenellaceae | Christensenellaceae R-7 group | NA |
| Levodopa | 432 | ASV18 | Genus | Firmicutes | Lachnospiraceae | Dorea | NA |
| Levodopa | 433 | ASV17 | Genus | Actinobacteriota | Coriobacteriaceae | Collinsella | NA |
| Levodopa | 434 | ASV81 | Genus | Firmicutes | Lachnospiraceae | Lachnospiraceae NK4A136 group | NA |
| Levodopa | 435 | ASV192 | Genus | Bacteroidota | Marinifilaceae | Odoribacter | NA |
| Levodopa | 436 | ASV66 | Genus | Firmicutes | Butyricicoccaceae | Butyricicoccus | NA |
| Levodopa | 437 | ASV408 | Genus | Bacteroidota | Prevotellaceae | Paraprevotella | NA |
| Levodopa | 438 | ASV226 | Genus | Firmicutes | Lachnospiraceae | Lachnospiraceae UCG-001 | NA |
| Levodopa | 439 | ASV13 | Genus | Firmicutes | Lachnospiraceae | Fusicatenibacter | NA |
| Levodopa | 440 | ASV12 | Genus | Firmicutes | Veillonellaceae | Dialister | NA |
| Levodopa | 441 | ASV37 | Genus | Firmicutes | Lachnospiraceae | [Eubacterium] hallii group | NA |
| Levodopa | 442 | ASV33 | Genus | Firmicutes | Peptostreptococcaceae | Intestinibacter | NA |
| Levodopa | 443 | ASV20 | Genus | Bacteroidota | Prevotellaceae | Prevotella | NA |
| Levodopa | 444 | ASV283 | Genus | Actinobacteriota | Eggerthellaceae | Slackia | NA |
| Levodopa | 445 | ASV974 | Genus | Firmicutes | Butyricicoccaceae | UCG-009 | NA |
| Levodopa | 446 | ASV87 | Genus | Proteobacteria | Sutterellaceae | Sutterella | NA |
| Levodopa | 447 | ASV8 | Genus | Firmicutes | Ruminococcaceae | Faecalibacterium | NA |
| Levodopa | 448 | ASV4 | Genus | Firmicutes | Ruminococcaceae | Ruminococcus | NA |
| Levodopa | 449 | ASV214 | Genus | Firmicutes | Lachnospiraceae | Lachnospiraceae UCG-010 | NA |

| Ticker | DESeq2_baseMean | DESeq2_log2-FoldChange | DESeq2_lfcSE | DESeq2_stat | DESeq2_pvalue | DESeq2_padj | DESeq2 converged |
|---|---|---|---|---|---|---|---|
| 420 | 130,5204589 | -0,002035276 | 0,001014949 | -2,005298002 | 0,044931198 | 0,639021486 | YES |
| 421 | 127,7908033 | 0,001204637 | 0,000680318 | 1,770696454 | 0,076611192 | 0,888030587 | YES |
| 422 | 141,0241001 | -0,001518871 | 0,000860216 | -1,765686062 | 0,077448532 | 0,888030587 | YES |
| 423 | 46,25841108 | -0,001852036 | 0,001102115 | -1,680438676 | 0,092871996 | 0,888030587 | YES |
| 424 | 97,80632938 | 0,001491879 | 0,000895877 | 1,665271347 | 0,095858632 | 0,888030587 | YES |
| 425 | 38,51324307 | 0,00184333 | 0,001118595 | 1,64789709 | 0,099373791 | 0,888030587 | YES |
| 426 | 142,8586301 | -0,000863554 | 0,00053127 | -1,625452936 | 0,104066084 | 0,888030587 | YES |
| 427 | 1223,163228 | -0,000484643 | 0,000331346 | -1,462649123 | 0,143563414 | 0,998341106 | YES |
| 428 | 92,9260607 | -0,000686408 | 0,000482458 | -1,422733032 | 0,154813561 | 0,998341106 | YES |
| 429 | 39,18375991 | -0,001981175 | 0,001412524 | -1,402577976 | 0,160742723 | 0,998341106 | YES |
| 430 | 722,6710778 | -0,00066486 | 0,000479796 | -1,385714764 | 0,165834018 | 0,998341106 | YES |
| 431 | 1241,920264 | 0,000680349 | 0,00053437 | 1,2731788 | 0,202954602 | 0,998341106 | YES |
| 432 | 563,3530868 | -0,000595886 | 0,00048669 | -1,224363675 | 0,220815066 | 0,998341106 | YES |
| 433 | 205,2048867 | -0,000768954 | 0,000629324 | -1,221872596 | 0,22175581 | 0,998341106 | YES |
| 434 | 207,5213105 | -0,001151475 | 0,000959513 | -1,200062966 | 0,230114887 | 0,998341106 | YES |
| 435 | 44,49373392 | 0,000641603 | 0,000613878 | 1,045163198 | 0,295947549 | 0,998341106 | YES |
| 436 | 145,9429848 | -0,000527551 | 0,000548812 | -0,961259864 | 0,336421523 | 0,998341106 | YES |
| 437 | 171,8329806 | -0,000687692 | 0,000764571 | -0,899448741 | 0,368413687 | 0,998341106 | YES |
| 438 | 61,35643696 | -0,001365195 | 0,001810973 | -0,753846312 | 0,450941512 | 0,998341106 | YES |
| 439 | 735,5686131 | -0,000428254 | 0,00058667 | -0,729974299 | 0,465405895 | 0,998341106 | YES |
| 440 | 486,5707601 | -0,000682819 | 0,000958126 | -0,71266062 | 0,47605579 | 0,998341106 | YES |
| 441 | 214,7423804 | -0,000331228 | 0,000467788 | -0,708072393 | 0,478900302 | 0,998341106 | YES |
| 442 | 34,28616114 | 0,001123136 | 0,001679174 | 0,668862604 | 0,503583127 | 0,998341106 | YES |
| 443 | 726,241955 | -0,002433896 | 0,003641446 | -0,668387401 | 0,503886336 | 0,998341106 | YES |
| 444 | 10,71585446 | -0,001292925 | 0,001981016 | -0,652657696 | 0,513976981 | 0,998341106 | YES |
| 445 | 13,40761928 | 0,002366901 | 0,003690912 | 0,641278098 | 0,521342016 | 0,998341106 | YES |
| 446 | 128,2324088 | -0,000465908 | 0,000750112 | -0,621117479 | 0,534522329 | 0,998341106 | YES |
| 447 | 2421,900823 | -0,001541994 | 0,002721862 | -0,566521911 | 0,571039042 | 0,998341106 | YES |
| 448 | 1932,526695 | -0,000273935 | 0,000553381 | -0,495021006 | 0,620585293 | 0,998341106 | YES |
| 449 | 42,29156391 | -0,000552295 | 0,001140653 | -0,484191972 | 0,628249634 | 0,998341106 | YES |

| Medication | Ticker | ASVnr | taxLevel | Phylum | Family | Genus | Species |
|---|---|---|---|---|---|---|---|
| Levodopa | 450 | ASV415 | Genus | Firmicutes | Oscillospiraceae | Oscillospira | NA |
| Levodopa | 451 | ASV550 | Genus | Bacteroidota | Rikenellaceae | Rikenellaceae RC9 gut group | NA |
| Levodopa | 452 | ASV23 | Genus | Firmicutes | Streptococcaceae | Streptococcus | NA |
| Levodopa | 453 | ASV40 | Genus | Firmicutes | Erysipelotrichaceae | Holdemanella | NA |
| Levodopa | 454 | ASV217 | Genus | Firmicutes | Peptococcaceae | Peptococcus | NA |
| Levodopa | 455 | ASV128 | Genus | Firmicutes | Oscillospiraceae | NK4A214 group | NA |
| Levodopa | 456 | ASV3 | Genus | Firmicutes | Ruminococcaceae | Subdoligranulum | NA |
| Levodopa | 457 | ASV26 | Genus | Euryarchaeota | Methanobacteriaceae | Methanobrevibacter | NA |
| Levodopa | 458 | ASV487 | Genus | Firmicutes | Lachnospiraceae | Howardella | NA |
| Levodopa | 459 | ASV133 | Genus | Actinobacteriota | Eggerthellaceae | Senegalimassilia | NA |
| Levodopa | 460 | ASV386 | Genus | Firmicutes | Ruminococcaceae | DTU089 | NA |
| Levodopa | 461 | ASV411 | Genus | Firmicutes | Lactobacillaceae | Lactobacillus | NA |
| Levodopa | 462 | ASV57 | Genus | Firmicutes | Lachnospiraceae | Roseburia | NA |
| Levodopa | 463 | ASV72 | Genus | Proteobacteria | Sutterellaceae | Parasutterella | NA |
| Levodopa | 464 | ASV53 | Genus | Firmicutes | Lachnospiraceae | [Ruminococcus] torques group | NA |
| Levodopa | 465 | ASV30 | Genus | Proteobacteria | Enterobacteriaceae | Escherichia-Shigella | NA |
| Levodopa | 466 | ASV328 | Genus | Firmicutes | Anaerovoracaceae | [Eubacterium] brachy group | NA |
| Levodopa | 467 | ASV630 | Genus | Firmicutes | Ruminococcaceae | Angelakisella | NA |
| Levodopa | 468 | ASV27 | Genus | Firmicutes | Ruminococcaceae | CAG-352 | NA |
| Levodopa | 469 | ASV315 | Genus | Firmicutes | Anaerovoracaceae | Family XIII UCG-001 | NA |
| Levodopa | 470 | ASV141 | Genus | Firmicutes | Lachnospiraceae | Lachnoclostridium | NA |
| Levodopa | 471 | ASV98 | Genus | Firmicutes | Acidaminococcaceae | Phascolarctobacterium | NA |
| Levodopa | 472 | ASV162 | Genus | Desulfobacterota | Desulfovibrionaceae | Desulfovibrio | NA |
| Levodopa | 473 | ASV34 | Genus | Firmicutes | Lachnospiraceae | Anaerostipes | NA |
| Levodopa | 474 | ASV197 | Genus | Bacteroidota | Prevotellaceae | Prevotellaceae NK3B31 group | NA |
| Levodopa | 475 | ASV303 | Genus | Firmicutes | Lachnospiraceae | GCA-900066575 | NA |
| Levodopa | 476 | ASV242 | Genus | Firmicutes | Lachnospiraceae | Tyzzerella | NA |
| Levodopa | 477 | ASV2 | Genus | Bacteroidota | Bacteroidaceae | Bacteroides | NA |
| Levodopa | 478 | ASV92 | Genus | Firmicutes | Erysipelotrichaceae | Turicibacter | NA |
| Levodopa | 479 | ASV338 | Genus | Firmicutes | Lachnospiraceae | Lachnotalea | NA |

| Ticker | DESeq2_baseMean | DESeq2_log2-FoldChange | DESeq2_lfcSE | DESeq2_stat | DESeq2_pvalue | DESeq2_padj | DESeq2 converged |
|---|---|---|---|---|---|---|---|
| 450 | 29,82142639 | 0,001036951 | 0,002226845 | 0,46565918 | 0,641459475 | 0,998341106 | YES |
| 451 | 187,5269105 | -0,000378723 | 0,00083262 | -0,454857653 | 0,64921165 | 0,998341106 | YES |
| 452 | 183,201132 | 0,000437939 | 0,00099804 | 0,438799393 | 0,6608069 | 0,998341106 | YES |
| 453 | 263,2560422 | -0,0002566 | 0,000587495 | -0,436769183 | 0,66227875 | 0,998341106 | YES |
| 454 | 74,43544401 | 0,00054722 | 0,001283891 | 0,42622024 | 0,66994737 | 0,998341106 | YES |
| 455 | 444,1717823 | 0,000891837 | 0,002094159 | 0,425868726 | 0,670203505 | 0,998341106 | YES |
| 456 | 1006,420478 | -0,000196077 | 0,000500008 | -0,392147723 | 0,694949068 | 0,998341106 | YES |
| 457 | 227,7305299 | 0,001086388 | 0,002803358 | 0,387530781 | 0,698363302 | 0,998341106 | YES |
| 458 | 9,07038158 | -0,001283889 | 0,00371847 | -0,345273482 | 0,729888781 | 0,998341106 | YES |
| 459 | 22,02411631 | 0,000399604 | 0,001161705 | 0,343980398 | 0,730861028 | 0,998341106 | YES |
| 460 | 10,14568357 | 0,000767645 | 0,002239427 | 0,3427865 | 0,731759083 | 0,998341106 | YES |
| 461 | 146,5234805 | 0,001369317 | 0,004184233 | 0,327256337 | 0,743474014 | 0,998341106 | YES |
| 462 | 502,1298847 | -0,000748128 | 0,002295054 | -0,325973959 | 0,744444058 | 0,998341106 | YES |
| 463 | 61,85286103 | 0,000344734 | 0,001076636 | 0,320195382 | 0,748820224 | 0,998341106 | YES |
| 464 | 559,9342945 | 0,000158572 | 0,000497665 | 0,318632766 | 0,750005004 | 0,998341106 | YES |
| 465 | 726,1129814 | -0,000947223 | 0,00302432 | -0,313201839 | 0,754127322 | 0,998341106 | YES |
| 466 | 0,876784564 | 0,001740765 | 0,006187577 | 0,281332306 | 0,778455534 | 0,998341106 | YES |
| 467 | 5,275101543 | 0,00157519 | 0,005706569 | 0,276030968 | 0,782524281 | 0,998341106 | YES |
| 468 | 344,5054983 | -0,001700525 | 0,00616321 | -0,275915437 | 0,782613017 | 0,998341106 | YES |
| 469 | 28,65779471 | -0,000249539 | 0,000919635 | -0,271345975 | 0,78612495 | 0,998341106 | YES |
| 470 | 168,1303521 | -0,000170167 | 0,000777121 | -0,218971004 | 0,826672635 | 0,998341106 | YES |
| 471 | 310,2423739 | -0,000470641 | 0,002376413 | -0,198047019 | 0,843008277 | 0,998341106 | YES |
| 472 | 267,8382523 | 7,36E-05 | 0,000385345 | 0,191052486 | 0,848484472 | 0,998341106 | YES |
| 473 | 151,3924744 | -0,000486642 | 0,002569621 | -0,189382801 | 0,849792803 | 0,998341106 | YES |
| 474 | 40,16815987 | -0,001153017 | 0,006192093 | -0,186207953 | 0,852281687 | 0,998341106 | YES |
| 475 | 14,47144605 | 0,000421332 | 0,002271728 | 0,185467769 | 0,852862158 | 0,998341106 | YES |
| 476 | 68,38846969 | 0,000578375 | 0,003315814 | 0,174429225 | 0,861528148 | 0,998341106 | YES |
| 477 | 4450,191083 | 5,18E-05 | 0,000298532 | 0,173430938 | 0,862312708 | 0,998341106 | YES |
| 478 | 39,53899495 | 0,000690547 | 0,00403241 | 0,171249167 | 0,864027849 | 0,998341106 | YES |
| 479 | 3,602630922 | -0,001058341 | 0,006187407 | -0,171047602 | 0,864186336 | 0,998341106 | YES |

| Medication | Ticker | ASVnr | taxLevel | Phylum | Family | Genus | Species |
|---|---|---|---|---|---|---|---|
| Levodopa | 480 | ASV143 | Genus | Firmicutes | Peptostreptococcaceae | Terrisporobacter | NA |
| Levodopa | 481 | ASV16 | Genus | Firmicutes | Peptostreptococcaceae | Romboutsia | NA |
| Levodopa | 482 | ASV61 | Genus | Firmicutes | Erysipelatoclostridiaceae | Catenibacterium | NA |
| Levodopa | 483 | ASV322 | Genus | Firmicutes | Anaerovoracaceae | Family XIII AD3011 group | NA |
| Levodopa | 484 | ASV130 | Genus | Proteobacteria | Enterobacteriaceae | Klebsiella | NA |
| Levodopa | 485 | ASV357 | Genus | Firmicutes | Lachnospiraceae | Marvinbryantia | NA |
| Levodopa | 486 | ASV75 | Genus | Firmicutes | Lachnospiraceae | [Ruminococcus] gauvreauii group | NA |
| Levodopa | 487 | ASV229 | Genus | Firmicutes | Oscillospiraceae | Flavonifractor | NA |
| Levodopa | 488 | ASV172 | Genus | Proteobacteria | Pasteurellaceae | Haemophilus | NA |
| Levodopa | 489 | ASV454 | Genus | Actinobacteriota | Actinomycetaceae | Actinomyces | NA |
| Levodopa | 490 | ASV306 | Genus | Firmicutes | Veillonellaceae | Veillonella | NA |
| Levodopa | 491 | ASV739 | Genus | Firmicutes | Ruminococcaceae | Anaerotruncus | NA |
| Levodopa | 492 | ASV147 | Genus | Firmicutes | Oscillospiraceae | Oscillibacter | NA |
| Levodopa | 493 | ASV512 | Genus | Proteobacteria | Oxalobacteraceae | Oxalobacter | NA |
| Levodopa | 494 | ASV97 | Genus | Firmicutes | Lachnospiraceae | [Ruminococcus] gnavus group | NA |
| Levodopa | 495 | ASV31 | Genus | Bacteroidota | Rikenellaceae | Alistipes | NA |
| Levodopa | 496 | ASV666 | Genus | Firmicutes | Erysipelotrichaceae | Holdemania | NA |
| Levodopa | 497 | ASV258 | Genus | Firmicutes | Lachnospiraceae | Lachnospiraceae AC2044 group | NA |
| Levodopa | 498 | ASV702 | Genus | Firmicutes | Lachnospiraceae | Hungatella | NA |
| Levodopa | 499 | ASV247 | Genus | Euryarchaeota | Methanobacteriaceae | Methanosphaera | NA |
| Levodopa | 500 | ASV444 | Genus | Firmicutes | Erysipelatoclostridiaceae | Coprobacillus | NA |
| Levodopa | 501 | ASV1 | Genus | Actinobacteriota | Bifidobacteriaceae | Bifidobacterium | NA |
| Levodopa | 502 | ASV240 | Genus | Bacteroidota | Prevotellaceae | Alloprevotella | NA |
| Levodopa | 503 | ASV190 | Genus | Firmicutes | Ruminococcaceae | UBA1819 | NA |
| Levodopa | 504 | ASV193 | Genus | Actinobacteriota | Eggerthellaceae | Adlercreutzia | NA |
| Levodopa | 505 | ASV1722 | Genus | Firmicutes | Erysipelatoclostridiaceae | Asteroleplasma | NA |
| Levodopa | 506 | ASV79 | Genus | Firmicutes | Lachnospiraceae | [Eubacterium] ruminantium group | NA |
| Levodopa | 507 | ASV1631 | Genus | Firmicutes | Ruminococcaceae | Paludicola | NA |
| Levodopa | 508 | ASV453 | Genus | Verrucomicrobiota | Victivallaceae | Victivallis | NA |
| Levodopa | 509 | ASV299 | Genus | Firmicutes | Streptococcaceae | Lactococcus | NA |

| Ticker | DESeq2_baseMean | DESeq2_log2-FoldChange | DESeq2_lfcSE | DESeq2_stat | DESeq2_pvalue | DESeq2_padj | DESeq2 converged |
|---|---|---|---|---|---|---|---|
| 480 | 6,236371299 | 0,001030556 | 0,006181248 | 0,166722981 | 0,867588023 | 0,998341106 | YES |
| 481 | 39,21052986 | 0,000263576 | 0,001632889 | 0,161416737 | 0,871765185 | 0,998341106 | YES |
| 482 | 77,10698596 | -0,000995218 | 0,006187186 | -0,160851434 | 0,872210415 | 0,998341106 | YES |
| 483 | 65,48340884 | -9,68E-05 | 0,000607976 | -0,159187135 | 0,87352144 | 0,998341106 | YES |
| 484 | 72,08342991 | 0,000934222 | 0,006191477 | 0,150888408 | 0,880063745 | 0,998341106 | YES |
| 485 | 68,44937291 | 0,000224059 | 0,001502966 | 0,149077684 | 0,881492335 | 0,998341106 | YES |
| 486 | 42,6394857 | 0,000326811 | 0,002207293 | 0,14805975 | 0,882295614 | 0,998341106 | YES |
| 487 | 7,524681296 | -0,000886328 | 0,006163501 | -0,143802611 | 0,885656341 | 0,998341106 | YES |
| 488 | 7,985578301 | -0,000849954 | 0,006174804 | -0,137648798 | 0,890517987 | 0,998341106 | YES |
| 489 | 0,9372743 | 0,000749959 | 0,006189369 | 0,121168837 | 0,903557306 | 0,998341106 | YES |
| 490 | 24,55111242 | -0,000721126 | 0,006175513 | -0,116771892 | 0,907040818 | 0,998341106 | YES |
| 491 | 3,394205931 | 0,000647344 | 0,006192697 | 0,104533501 | 0,916745984 | 0,998341106 | YES |
| 492 | 59,15597452 | 7,02E-05 | 0,000686921 | 0,102253483 | 0,918555477 | 0,998341106 | YES |
| 493 | 16,05310098 | 9,51E-05 | 0,000931018 | 0,102136003 | 0,918648724 | 0,998341106 | YES |
| 494 | 57,51803175 | 0,000618008 | 0,006185751 | 0,09990831 | 0,920417119 | 0,998341106 | YES |
| 495 | 1172,576683 | 2,97E-05 | 0,000309395 | 0,095867726 | 0,923625628 | 0,998341106 | YES |
| 496 | 1,872431237 | -0,000581868 | 0,006176535 | -0,094206246 | 0,924945324 | 0,998341106 | YES |
| 497 | 89,88255606 | 0,000213357 | 0,002275806 | 0,093750271 | 0,925307536 | 0,998341106 | YES |
| 498 | 16,87569309 | -0,000574401 | 0,006192279 | -0,09276081 | 0,926093586 | 0,998341106 | YES |
| 499 | 10,94265096 | 0,000559759 | 0,006186628 | 0,090478878 | 0,927906678 | 0,998341106 | YES |
| 500 | 8,161865312 | 0,000547646 | 0,006179024 | 0,088629918 | 0,929376031 | 0,998341106 | YES |
| 501 | 716,9067968 | -0,000192206 | 0,002182733 | -0,088057684 | 0,929830829 | 0,998341106 | YES |
| 502 | 125,4365284 | 0,000543127 | 0,006178713 | 0,087902993 | 0,929953778 | 0,998341106 | YES |
| 503 | 25,33044464 | -0,000192329 | 0,002219588 | -0,086650822 | 0,930949068 | 0,998341106 | YES |
| 504 | 9,224301864 | -0,000330711 | 0,00389074 | -0,084999556 | 0,932261743 | 0,998341106 | YES |
| 505 | 4,33460539 | -0,000432033 | 0,006189 | -0,069806605 | 0,94434759 | 0,998341106 | YES |
| 506 | 172,3895039 | -0,00036298 | 0,005544616 | -0,06546525 | 0,947803573 | 0,998341106 | YES |
| 507 | 6,081551098 | -0,000371236 | 0,006181344 | -0,060057564 | 0,952109788 | 0,998341106 | YES |
| 508 | 8,206624497 | 0,000322177 | 0,005377836 | 0,059908385 | 0,952228602 | 0,998341106 | YES |
| 509 | 8,931084815 | 0,000367011 | 0,006183712 | 0,059351212 | 0,952672372 | 0,998341106 | YES |

| Medication | Ticker | ASVnr | taxLevel | Phylum | Family | Genus | Species |
|---|---|---|---|---|---|---|---|
| Levodopa | 510 | ASV978 | Genus | Firmicutes | Erysipelotrichaceae | Dielma | NA |
| Levodopa | 511 | ASV294 | Genus | Firmicutes | Anaerovoracaceae | Mogibacterium | NA |
| Levodopa | 512 | ASV376 | Genus | Bacteroidota | Barnesiellaceae | Coprobacter | NA |
| Levodopa | 513 | ASV10 | Genus | Verrucomicrobiota | Akkermansiaceae | Akkermansia | NA |
| Levodopa | 514 | ASV134 | Genus | Actinobacteriota | Eggerthellaceae | Enterorhabdus | NA |
| Levodopa | 515 | ASV232 | Genus | Firmicutes | Acholeplasmataceae | Anaeroplasma | NA |
| Levodopa | 516 | ASV158 | Genus | Firmicutes | Lachnospiraceae | [Eubacterium] ventriosum group | NA |
| Levodopa | 517 | ASV885 | Genus | Firmicutes | Lachnospiraceae | Shuttleworthia | NA |
| Levodopa | 518 | ASV252 | Genus | Firmicutes | Erysipelotrichaceae | Solobacterium | NA |
| Levodopa | 519 | ASV892 | Genus | Synergistota | Synergistaceae | Cloacibacillus | NA |
| Levodopa | 520 | ASV858 | Genus | Actinobacteriota | Atopobiaceae | Libanicoccus | NA |
| Levodopa | 521 | ASV449 | Genus | Actinobacteriota | Atopobiaceae | Olsenella | NA |
| Levodopa | 522 | ASV29 | Genus | Firmicutes | Oscillospiraceae | UCG-002 | NA |
| Levodopa | 523 | ASV530 | Genus | Firmicutes | Veillonellaceae | Allisonella | NA |
| Levodopa | 524 | ASV1506 | Genus | Firmicutes | Erysipelatoclostridiaceae | UCG-004 | NA |
| Levodopa | 525 | ASV78 | Genus | Firmicutes | Lachnospiraceae | [Eubacterium] eligens group | NA |
| Levodopa | 526 | ASV499 | Genus | Firmicutes | Lachnospiraceae | Eisenbergiella | NA |
| Levodopa | 527 | ASV167 | Genus | Firmicutes | Oscillospiraceae | Intestinimonas | NA |
| Levodopa | 528 | ASV1319 | Genus | Firmicutes | Ruminococcaceae | Candidatus Soleaferrea | NA |
| Levodopa | 529 | ASV1213 | Genus | Desulfobacterota | Desulfovibrionaceae | Mailhella | NA |
| Levodopa | 530 | ASV385 | Genus | Firmicutes | Selenomonadaceae | Mitsuokella | NA |
| Levodopa | 531 | ASV1403 | Genus | Bacteroidota | Marinifilaceae | Sanguibacteroides | NA |
| Levodopa | 532 | ASV428 | Genus | Bacteroidota | Prevotellaceae | Prevotellaceae UCG-001 | NA |
| Levodopa | 533 | ASV60 | Genus | Firmicutes | Ruminococcaceae | [Eubacterium] siraeum group | NA |
| Levodopa | 534 | ASV761 | Genus | Firmicutes | Defluviitaleaceae | Defluviitaleaceae UCG-011 | NA |
| Levodopa | 535 | ASV39 | Genus | Bacteroidota | Tannerellaceae | Parabacteroides | NA |
| Levodopa | 536 | ASV690 | Genus | Firmicutes | Ruminococcaceae | Fournierella | NA |
| Levodopa | 537 | ASV293 | Genus | Firmicutes | Erysipelotrichaceae | Catenisphaera | NA |
| Levodopa | 538 | ASV573 | Genus | Firmicutes | Erysipelatoclostridiaceae | Erysipelatoclostridium | NA |
| Levodopa | 539 | ASV200 | Genus | Firmicutes | Lachnospiraceae | [Eubacterium] xylanophilum group | NA |

| Ticker | DESeq2_baseMean | DESeq2_log2-FoldChange | DESeq2_lfcSE | DESeq2_stat | DESeq2_pvalue | DESeq2_padj | DESeq2 converged |
|---|---|---|---|---|---|---|---|
| 510 | 2,467284572 | -0,000360403 | 0,006189256 | -0,058230472 | 0,953565049 | 0,998341106 | YES |
| 511 | 24,31824105 | 0,000338056 | 0,00618371 | 0,054668811 | 0,956402317 | 0,998341106 | YES |
| 512 | 12,23923207 | 9,25E-05 | 0,001731613 | 0,053424373 | 0,957393786 | 0,998341106 | YES |
| 513 | 1059,636449 | 0,000199107 | 0,003886828 | 0,051226117 | 0,959145341 | 0,998341106 | YES |
| 514 | 8,411446589 | -8,16E-05 | 0,001670129 | -0,048862415 | 0,961028942 | 0,998341106 | YES |
| 515 | 38,90305883 | 0,000298665 | 0,006190386 | 0,04824651 | 0,961519783 | 0,998341106 | YES |
| 516 | 78,28506593 | 4,16E-05 | 0,000867082 | 0,047990955 | 0,961723451 | 0,998341106 | YES |
| 517 | 7,490118204 | 0,000274435 | 0,006172826 | 0,04445865 | 0,964538812 | 0,998341106 | YES |
| 518 | 16,7696842 | 0,000271865 | 0,006187856 | 0,043935215 | 0,964956045 | 0,998341106 | YES |
| 519 | 13,57382377 | 0,00027071 | 0,006187299 | 0,043752499 | 0,965101691 | 0,998341106 | YES |
| 520 | 3,839444221 | 0,000270485 | 0,00618998 | 0,043697193 | 0,965145777 | 0,998341106 | YES |
| 521 | 5,319040721 | -0,000259104 | 0,006179987 | -0,041926226 | 0,96655751 | 0,998341106 | YES |
| 522 | 2038,191126 | 1,54E-05 | 0,000373096 | 0,041217575 | 0,967122443 | 0,998341106 | YES |
| 523 | 9,952250596 | -0,000253766 | 0,006179877 | -0,041063308 | 0,967245425 | 0,998341106 | YES |
| 524 | 11,24689325 | 0,000225893 | 0,006187091 | 0,036510386 | 0,970875398 | 0,998341106 | YES |
| 525 | 191,6833352 | 9,15E-05 | 0,002693434 | 0,033956594 | 0,972911764 | 0,998341106 | YES |
| 526 | 8,023429284 | -0,000205544 | 0,006189736 | -0,033207264 | 0,973509305 | 0,998341106 | YES |
| 527 | 126,1457446 | 5,32E-05 | 0,001801658 | 0,029548241 | 0,976427345 | 0,998341106 | YES |
| 528 | 6,99789109 | -0,000180349 | 0,006185097 | -0,029158556 | 0,976738135 | 0,998341106 | YES |
| 529 | 5,408764909 | 0,000157743 | 0,006191316 | 0,025478183 | 0,97967355 | 0,998341106 | YES |
| 530 | 25,868923 | 0,000142142 | 0,006188876 | 0,022967286 | 0,981676368 | 0,998341106 | YES |
| 531 | 2,994673597 | 0,000126331 | 0,006186941 | 0,020418962 | 0,983709157 | 0,998341106 | YES |
| 532 | 105,4778736 | 0,000117827 | 0,006193308 | 0,019024896 | 0,984821245 | 0,998341106 | YES |
| 533 | 174,2386528 | 1,64E-05 | 0,001067149 | 0,01534952 | 0,987753336 | 0,998341106 | YES |
| 534 | 4,440207473 | 7,58E-05 | 0,006183923 | 0,012261992 | 0,990216591 | 0,998341106 | YES |
| 535 | 632,4957271 | -2,21E-05 | 0,002011574 | -0,010986276 | 0,991234397 | 0,998341106 | YES |
| 536 | 16,90710766 | 4,89E-05 | 0,006184236 | 0,007905829 | 0,993692126 | 0,998341106 | YES |
| 537 | 23,74637616 | -4,61E-05 | 0,006189538 | -0,00744233 | 0,994061935 | 0,998341106 | YES |
| 538 | 8,972415919 | 9,19E-06 | 0,00406394 | 0,002261707 | 0,998195421 | 0,998341106 | YES |
| 539 | 43,51055513 | -2,80E-06 | 0,00134881 | -0,002079116 | 0,998341106 | 0,998341106 | YES |

| Medication | Ticker | ASVnr | taxLevel | Phylum | Family | Genus | Species |
|---|---|---|---|---|---|---|---|
| Levodopa | 540 | ASV7 | Family | Firmicutes | Lachnospiraceae | NA | NA |
| Levodopa | 541 | ASV19 | Family | Firmicutes | Erysipelatoclostridiaceae | NA | NA |
| Levodopa | 542 | ASV4 | Family | Firmicutes | Ruminococcaceae | NA | NA |
| Levodopa | 543 | ASV63 | Family | Actinobacteriota | Eggerthellaceae | NA | NA |
| Levodopa | 544 | ASV17 | Family | Actinobacteriota | Coriobacteriaceae | NA | NA |
| Levodopa | 545 | ASV31 | Family | Bacteroidota | Rikenellaceae | NA | NA |
| Levodopa | 546 | ASV40 | Family | Firmicutes | Erysipelotrichaceae | NA | NA |
| Levodopa | 547 | ASV55 | Family | Firmicutes | Monoglobaceae | NA | NA |
| Levodopa | 548 | ASV192 | Family | Bacteroidota | Marinifilaceae | NA | NA |
| Levodopa | 549 | ASV66 | Family | Firmicutes | Butyricicoccaceae | NA | NA |
| Levodopa | 550 | ASV28 | Family | Firmicutes | Clostridiaceae | NA | NA |
| Levodopa | 551 | ASV411 | Family | Firmicutes | Lactobacillaceae | NA | NA |
| Levodopa | 552 | ASV72 | Family | Proteobacteria | Sutterellaceae | NA | NA |
| Levodopa | 553 | ASV195 | Family | Bacteroidota | Barnesiellaceae | NA | NA |
| Levodopa | 554 | ASV59 | Family | Firmicutes | [Eubacterium] coprostanoligenes group | NA | NA |
| Levodopa | 555 | ASV20 | Family | Bacteroidota | Prevotellaceae | NA | NA |
| Levodopa | 556 | ASV2 | Family | Bacteroidota | Bacteroidaceae | NA | NA |
| Levodopa | 557 | ASV23 | Family | Firmicutes | Streptococcaceae | NA | NA |
| Levodopa | 558 | ASV162 | Family | Desulfobacterota | Desulfovibrionaceae | NA | NA |
| Levodopa | 559 | ASV15 | Family | Firmicutes | Christensenellaceae | NA | NA |
| Levodopa | 560 | ASV322 | Family | Firmicutes | Anaerovoracaceae | NA | NA |
| Levodopa | 561 | ASV217 | Family | Firmicutes | Peptococcaceae | NA | NA |
| Levodopa | 562 | ASV29 | Family | Firmicutes | Oscillospiraceae | NA | NA |
| Levodopa | 563 | ASV26 | Family | Euryarchaeota | Methanobacteriaceae | NA | NA |
| Levodopa | 564 | ASV449 | Family | Actinobacteriota | Atopobiaceae | NA | NA |
| Levodopa | 565 | ASV417 | Family | Firmicutes | UCG-010 | NA | NA |
| Levodopa | 566 | ASV16 | Family | Firmicutes | Peptostreptococcaceae | NA | NA |
| Levodopa | 567 | ASV172 | Family | Proteobacteria | Pasteurellaceae | NA | NA |
| Levodopa | 568 | ASV1 | Family | Actinobacteriota | Bifidobacteriaceae | NA | NA |
| Levodopa | 569 | ASV30 | Family | Proteobacteria | Enterobacteriaceae | NA | NA |

| Ticker | DESeq2_baseMean | DESeq2_log2-FoldChange | DESeq2_lfcSE | DESeq2_stat | DESeq2_pvalue | DESeq2_padj | DESeq2 converged |
|---|---|---|---|---|---|---|---|
| 540 | 10353,20266 | -0,000913081 | 0,000271599 | -3,361873826 | 0,000774155 | 0,037159435 | YES |
| 541 | 261,6799201 | -0,001166189 | 0,000391587 | -2,978109357 | 0,002900325 | 0,047110919 | YES |
| 542 | 6758,921304 | -0,000662556 | 0,000222822 | -2,973480365 | 0,002944432 | 0,047110919 | YES |
| 543 | 144,578283 | -0,001007844 | 0,000376778 | -2,674901038 | 0,007475131 | 0,089701574 | YES |
| 544 | 198,2351386 | -0,001083742 | 0,000619348 | -1,749811555 | 0,080150836 | 0,769448027 | YES |
| 545 | 1368,812 | -0,000438416 | 0,000297602 | -1,4731634 | 0,140706981 | 0,884526885 | YES |
| 546 | 380,6336525 | -0,000706337 | 0,00052298 | -1,35060136 | 0,176823165 | 0,884526885 | YES |
| 547 | 135,7952013 | 0,000929225 | 0,000695376 | 1,336290547 | 0,181454325 | 0,884526885 | YES |
| 548 | 93,33142425 | 0,000542298 | 0,00041239 | 1,315011257 | 0,188506119 | 0,884526885 | YES |
| 549 | 230,3853905 | -0,000615928 | 0,000481259 | -1,279825357 | 0,200606564 | 0,884526885 | YES |
| 550 | 118,888961 | 0,001152955 | 0,000905069 | 1,273885275 | 0,202704078 | 0,884526885 | YES |
| 551 | 185,0695562 | 0,002337256 | 0,002067503 | 1,130472899 | 0,258277012 | 0,97142824 | YES |
| 552 | 194,2738504 | -0,00055152 | 0,000492822 | -1,119105466 | 0,263095148 | 0,97142824 | YES |
| 553 | 328,3399431 | -0,001883894 | 0,002104026 | -0,895375735 | 0,370586267 | 0,996950525 | YES |
| 554 | 470,7983445 | 0,000239803 | 0,000327665 | 0,731855435 | 0,464256809 | 0,996950525 | YES |
| 555 | 1322,65234 | -0,002079487 | 0,003123615 | -0,665730908 | 0,505583114 | 0,996950525 | YES |
| 556 | 4686,872673 | -0,000170673 | 0,000309098 | -0,552163303 | 0,580836474 | 0,996950525 | YES |
| 557 | 224,4978492 | 0,00052195 | 0,000968739 | 0,538793941 | 0,590029045 | 0,996950525 | YES |
| 558 | 324,543189 | 0,000176358 | 0,000344538 | 0,511868066 | 0,608743349 | 0,996950525 | YES |
| 559 | 1661,879145 | 0,000224041 | 0,000534345 | 0,419281745 | 0,675010237 | 0,996950525 | YES |
| 560 | 121,6766104 | 0,000158769 | 0,000413161 | 0,384278229 | 0,700772243 | 0,996950525 | YES |
| 561 | 86,5675905 | 0,000272226 | 0,000721007 | 0,377564143 | 0,705754406 | 0,996950525 | YES |
| 562 | 3614,639811 | -9,27E-05 | 0,000255398 | -0,36292231 | 0,71666292 | 0,996950525 | YES |
| 563 | 205,3982853 | 0,000930275 | 0,002696483 | 0,344995686 | 0,730097614 | 0,996950525 | YES |
| 564 | 17,34544845 | -0,000644789 | 0,00187956 | -0,343052849 | 0,731558702 | 0,996950525 | YES |
| 565 | 194,0721117 | -0,000441499 | 0,001849143 | -0,238758439 | 0,811292899 | 0,996950525 | YES |
| 566 | 87,91799439 | 0,000211569 | 0,000943161 | 0,224319592 | 0,822508632 | 0,996950525 | YES |
| 567 | 9,306920299 | -0,001013439 | 0,005118162 | -0,198008348 | 0,843038533 | 0,996950525 | YES |
| 568 | 696,4177771 | -0,000393442 | 0,002218162 | -0,177373044 | 0,859215379 | 0,996950525 | YES |
| 569 | 964,6541405 | -0,000456714 | 0,002987071 | -0,152896867 | 0,878479607 | 0,996950525 | YES |

| Medication | Ticker | ASVnr | taxLevel | Phylum | Family | Genus | Species |
|---|---|---|---|---|---|---|---|
| Levodopa | 570 | ASV98 | Family | Firmicutes | Acidaminococcaceae | NA | NA |
| Levodopa | 571 | ASV703 | Family | Firmicutes | Hydrogenoanaerobacterium | NA | NA |
| Levodopa | 572 | ASV453 | Family | Verrucomicrobiota | Victivallaceae | NA | NA |
| Levodopa | 573 | ASV145 | Family | Bacteroidota | Muribaculaceae | NA | NA |
| Levodopa | 574 | ASV39 | Family | Bacteroidota | Tannerellaceae | NA | NA |
| Levodopa | 575 | ASV255 | Family | Actinobacteriota | Coriobacteriales Incertae Sedis | NA | NA |
| Levodopa | 576 | ASV1119 | Family | Verrucomicrobiota | vadinBE97 | NA | NA |
| Levodopa | 577 | ASV892 | Family | Synergistota | Synergistaceae | NA | NA |
| Levodopa | 578 | ASV752 | Family | Verrucomicrobiota | Puniceicoccaceae | NA | NA |
| Levodopa | 579 | ASV802 | Family | Firmicutes | Ruminiclostridium | NA | NA |
| Levodopa | 580 | ASV454 | Family | Actinobacteriota | Actinomycetaceae | NA | NA |
| Levodopa | 581 | ASV512 | Family | Proteobacteria | Oxalobacteraceae | NA | NA |
| Levodopa | 582 | ASV232 | Family | Firmicutes | Acholeplasmataceae | NA | NA |
| Levodopa | 583 | ASV10 | Family | Verrucomicrobiota | Akkermansiaceae | NA | NA |
| Levodopa | 584 | ASV1052 | Family | Thermoplasmatota | Methanomethylophilaceae | NA | NA |
| Levodopa | 585 | ASV385 | Family | Firmicutes | Selenomonadaceae | NA | NA |
| Levodopa | 586 | ASV761 | Family | Firmicutes | Defluviitaleaceae | NA | NA |
| Levodopa | 587 | ASV12 | Family | Firmicutes | Veillonellaceae | NA | NA |

| Ticker | DESeq2_baseMean | DESeq2_log2-FoldChange | DESeq2_lfcSE | DESeq2_stat | DESeq2_pvalue | DESeq2_padj | DESeq2 converged |
|---|---|---|---|---|---|---|---|
| 570 | 360,9905604 | -0,00033694 | 0,00237186 | -0,14205713 | 0,88703488 | 0,996950525 | YES |
| 571 | 2,829129513 | -0,000831702 | 0,006184309 | -0,134485829 | 0,893018416 | 0,996950525 | YES |
| 572 | 11,35129158 | -0,000350414 | 0,002662246 | -0,131623605 | 0,895282014 | 0,996950525 | YES |
| 573 | 382,535211 | -0,000384662 | 0,003117292 | -0,123396283 | 0,9017933 | 0,996950525 | YES |
| 574 | 702,7423541 | -0,000213751 | 0,002075293 | -0,102998244 | 0,917964365 | 0,996950525 | YES |
| 575 | 24,93678287 | 0,000159952 | 0,001602809 | 0,099795072 | 0,92050702 | 0,996950525 | YES |
| 576 | 8,370674172 | 0,000574002 | 0,006178254 | 0,092906773 | 0,925977625 | 0,996950525 | YES |
| 577 | 11,69017375 | 0,000490664 | 0,006176498 | 0,079440469 | 0,936682281 | 0,996950525 | YES |
| 578 | 6,964085125 | 0,000418823 | 0,006173594 | 0,067841036 | 0,945912177 | 0,996950525 | YES |
| 579 | 4,886888113 | 0,000240958 | 0,006182865 | 0,038971888 | 0,968912801 | 0,996950525 | YES |
| 580 | 1,22960968 | 0,000216576 | 0,0061821 | 0,035032733 | 0,97205364 | 0,996950525 | YES |
| 581 | 15,30891264 | 2,25E-05 | 0,000880651 | 0,025538071 | 0,979625782 | 0,996950525 | YES |
| 582 | 32,18864131 | 0,000133571 | 0,006190372 | 0,021577253 | 0,982785179 | 0,996950525 | YES |
| 583 | 911,7268497 | 6,52E-05 | 0,003694264 | 0,017653013 | 0,985915665 | 0,996950525 | YES |
| 584 | 4,633680537 | -6,93E-05 | 0,006193165 | -0,011196167 | 0,991066938 | 0,996950525 | YES |
| 585 | 19,43253785 | -3,93E-05 | 0,006188789 | -0,006358032 | 0,994927059 | 0,996950525 | YES |
| 586 | 3,371600749 | 3,73E-05 | 0,006184631 | 0,006036259 | 0,995183791 | 0,996950525 | YES |
| 587 | 642,4361392 | 1,26E-05 | 0,003288386 | 0,003821959 | 0,996950525 | 0,996950525 | YES |

**Supplementary Table 8c: Dopamine agonists (DA LEDD)**

| Medication | Ticker | ASVnr | taxLevel | Phylum | Family | Genus | Species |
|---|---|---|---|---|---|---|---|
| DA LEDD | 1 | ASV15 | ASV | Firmicutes | Christensenellaceae | Christensenellaceae R-7 group | NA |
| DA LEDD | 2 | ASV110 | ASV | Firmicutes | Ruminococcaceae | Faecalibacterium | prausnitzii |
| DA LEDD | 3 | ASV196 | ASV | Firmicutes | Ruminococcaceae | Ruminococcus | NA |
| DA LEDD | 4 | ASV13 | ASV | Firmicutes | Lachnospiraceae | Fusicatenibacter | saccharivorans |
| DA LEDD | 5 | ASV1 | ASV | Actinobacteriota | Bifidobacteriaceae | Bifidobacterium | NA |
| DA LEDD | 6 | ASV57 | ASV | Firmicutes | Lachnospiraceae | Roseburia | inulinivorans |
| DA LEDD | 7 | ASV193 | ASV | Actinobacteriota | Eggerthellaceae | Adlercreutzia | equolifaciens |
| DA LEDD | 8 | ASV138 | ASV | Bacteroidota | Rikenellaceae | Alistipes | obesi |
| DA LEDD | 9 | ASV35 | ASV | Firmicutes | Lachnospiraceae | Agathobacter | NA |
| DA LEDD | 10 | ASV16 | ASV | Firmicutes | Peptostreptococcaceae | Romboutsia | NA |
| DA LEDD | 11 | ASV328 | ASV | Firmicutes | Anaerovoracaceae | [Eubacterium] brachy group | NA |
| DA LEDD | 12 | ASV21 | ASV | Firmicutes | Ruminococcaceae | Faecalibacterium | prausnitzii |
| DA LEDD | 13 | ASV329 | ASV | Firmicutes | Lachnospiraceae | NA | NA |
| DA LEDD | 14 | ASV44 | ASV | Bacteroidota | Bacteroidaceae | Bacteroides | NA |
| DA LEDD | 15 | ASV92 | ASV | Firmicutes | Erysipelotrichaceae | Turicibacter | sanguinis |
| DA LEDD | 16 | ASV6 | ASV | Firmicutes | Ruminococcaceae | Faecalibacterium | NA |
| DA LEDD | 17 | ASV423 | ASV | Firmicutes | Christensenellaceae | Christensenellaceae R-7 group | NA |
| DA LEDD | 18 | ASV23 | ASV | Firmicutes | Streptococcaceae | Streptococcus | NA |
| DA LEDD | 19 | ASV5 | ASV | Firmicutes | Lachnospiraceae | Blautia | NA |
| DA LEDD | 20 | ASV512 | ASV | Proteobacteria | Oxalobacteraceae | Oxalobacter | formigenes |
| DA LEDD | 21 | ASV3 | ASV | Firmicutes | Ruminococcaceae | Subdoligranulum | NA |
| DA LEDD | 22 | ASV192 | ASV | Bacteroidota | Marinifilaceae | Odoribacter | splanchnicus |
| DA LEDD | 23 | ASV1030 | ASV | Firmicutes | NA | NA | NA |
| DA LEDD | 24 | ASV229 | ASV | Firmicutes | Oscillospiraceae | Flavonifractor | plautii |
| DA LEDD | 25 | ASV200 | ASV | Firmicutes | Lachnospiraceae | [Eubacterium] xylanophilum group | NA |
| DA LEDD | 26 | ASV25 | ASV | Actinobacteriota | Bifidobacteriaceae | Bifidobacterium | NA |
| DA LEDD | 27 | ASV97 | ASV | Firmicutes | Lachnospiraceae | [Ruminococcus] gnavus group | NA |
| DA LEDD | 28 | ASV284 | ASV | Firmicutes | Oscillospiraceae | NK4A214 group | NA |
| DA LEDD | 29 | ASV268 | ASV | Firmicutes | Ruminococcaceae | Incertae Sedis | NA |

| Ticker | DESeq2_baseMean | DESeq2_log2-FoldChange | DESeq2_lfcSE | DESeq2_stat | DESeq2_pvalue | DESeq2_padj | DESeq2 converged |
|---|---|---|---|---|---|---|---|
| 1 | 67,81235193 | -13,14477544 | 0,009284891 | -1415,716778 | 0 | 0 | YES |
| 2 | 27,57702312 | -9,304741586 | 0,009284895 | -1002,137495 | 0 | 0 | YES |
| 3 | 35,77101564 | -13,56396478 | 0,009284893 | -1460,86381 | 0 | 0 | YES |
| 4 | 717,8497493 | 0,008630894 | 0,005454512 | 1,582340315 | 0,113571902 | 0,997431792 | YES |
| 5 | 400,8590305 | 0,007996102 | 0,005293083 | 1,510670062 | 0,130872534 | 0,997431792 | YES |
| 6 | 165,9657635 | -0,008888785 | 0,006583915 | -1,35007586 | 0,176991651 | 0,997431792 | YES |
| 7 | 9,805297733 | 0,01022069 | 0,008002105 | 1,277250159 | 0,201513944 | 0,997431792 | YES |
| 8 | 80,9778734 | 0,007424547 | 0,006033104 | 1,230634594 | 0,218459561 | 0,997431792 | YES |
| 9 | 512,9968805 | 0,00850847 | 0,00700984 | 1,213789583 | 0,224828085 | 0,997431792 | YES |
| 10 | 44,10389525 | 0,007160083 | 0,005986139 | 1,196110331 | 0,231653507 | 0,997431792 | YES |
| 11 | 9,922552433 | 0,009064894 | 0,007727812 | 1,173022096 | 0,240786945 | 0,997431792 | YES |
| 12 | 460,9638656 | -0,006114912 | 0,005667414 | -1,078959798 | 0,28060565 | 0,997431792 | YES |
| 13 | 15,01808272 | -0,009353432 | 0,009004335 | -1,038769907 | 0,29891176 | 0,997431792 | YES |
| 14 | 246,6626698 | -0,007379815 | 0,007133781 | -1,034488552 | 0,300907826 | 0,997431792 | YES |
| 15 | 7,09932009 | 0,008614277 | 0,009036519 | 0,953273807 | 0,340451359 | 0,997431792 | YES |
| 16 | 977,2739625 | 0,004576026 | 0,004814752 | 0,95041786 | 0,341899972 | 0,997431792 | YES |
| 17 | 22,84262922 | 0,006645414 | 0,007207081 | 0,922067364 | 0,356493439 | 0,997431792 | YES |
| 18 | 655,5226161 | 0,003719285 | 0,004428159 | 0,839916748 | 0,400955067 | 0,997431792 | YES |
| 19 | 886,1689657 | -0,002957988 | 0,003626241 | -0,815717467 | 0,414661747 | 0,997431792 | YES |
| 20 | 9,924570874 | 0,005195733 | 0,007049302 | 0,737056413 | 0,461088044 | 0,997431792 | YES |
| 21 | 515,8006642 | 0,006543457 | 0,008908806 | 0,734493122 | 0,462648251 | 0,997431792 | YES |
| 22 | 22,93412291 | -0,003490271 | 0,004831749 | -0,722361943 | 0,470071979 | 0,997431792 | YES |
| 23 | 1,655977854 | 0,00634597 | 0,008959666 | 0,708282009 | 0,478770147 | 0,997431792 | YES |
| 24 | 73,16987435 | -0,004395815 | 0,006356908 | -0,69150212 | 0,489250049 | 0,997431792 | YES |
| 25 | 48,2493914 | -0,004803552 | 0,006971109 | -0,689065757 | 0,490781888 | 0,997431792 | YES |
| 26 | 597,1037513 | 0,002836353 | 0,004153858 | 0,682823755 | 0,494718217 | 0,997431792 | YES |
| 27 | 121,7579298 | -0,006094473 | 0,009059213 | -0,672737546 | 0,501114276 | 0,997431792 | YES |
| 28 | 3,415144542 | -0,006095756 | 0,009078088 | -0,671480193 | 0,501914673 | 0,997431792 | YES |
| 29 | 29,97171447 | 0,005142939 | 0,007814306 | 0,658143958 | 0,510445632 | 0,997431792 | YES |

| Medication | Ticker | ASVnr | taxLevel | Phylum | Family | Genus | Species |
|---|---|---|---|---|---|---|---|
| DA LEDD | 30 | ASV91 | ASV | Firmicutes | Lachnospiraceae | [Eubacterium] eligens group | NA |
| DA LEDD | 31 | ASV820 | ASV | Firmicutes | Lachnospiraceae | GCA-900066575 | NA |
| DA LEDD | 32 | ASV295 | ASV | Firmicutes | Ruminococcaceae | Negativibacillus | NA |
| DA LEDD | 33 | ASV413 | ASV | Firmicutes | Oscillospiraceae | Oscillibacter | NA |
| DA LEDD | 34 | ASV96 | ASV | Firmicutes | Lachnospiraceae | CAG-56 | NA |
| DA LEDD | 35 | ASV218 | ASV | Firmicutes | Anaerovoracaceae | Family XIII AD3011 group | NA |
| DA LEDD | 36 | ASV4 | ASV | Firmicutes | Ruminococcaceae | Ruminococcus | bromii |
| DA LEDD | 37 | ASV341 | ASV | Firmicutes | Christensenellaceae | Christensenellaceae R-7 group | NA |
| DA LEDD | 38 | ASV76 | ASV | Firmicutes | Lachnospiraceae | Lachnospiraceae ND3007 group | NA |
| DA LEDD | 39 | ASV60 | ASV | Firmicutes | Ruminococcaceae | [Eubacterium] siraeum group | NA |
| DA LEDD | 40 | ASV83 | ASV | Firmicutes | Lachnospiraceae | Lachnospiraceae NK4A136 group | NA |
| DA LEDD | 41 | ASV630 | ASV | Firmicutes | Ruminococcaceae | Angelakisella | NA |
| DA LEDD | 42 | ASV161 | ASV | Firmicutes | Oscillospiraceae | UCG-002 | NA |
| DA LEDD | 43 | ASV87 | ASV | Proteobacteria | Sutterellaceae | Sutterella | wadsworthensis |
| DA LEDD | 44 | ASV70 | ASV | Firmicutes | Lachnospiraceae | Roseburia | intestinalis |
| DA LEDD | 45 | ASV10 | ASV | Verrucomicrobiota | Akkermansiaceae | Akkermansia | muciniphila |
| DA LEDD | 46 | ASV226 | ASV | Firmicutes | Lachnospiraceae | Lachnospiraceae UCG-001 | NA |
| DA LEDD | 47 | ASV131 | ASV | Firmicutes | Lachnospiraceae | NA | NA |
| DA LEDD | 48 | ASV39 | ASV | Bacteroidota | Tannerellaceae | Parabacteroides | merdae |
| DA LEDD | 49 | ASV7 | ASV | Firmicutes | Lachnospiraceae | Agathobacter | NA |
| DA LEDD | 50 | ASV14 | ASV | Firmicutes | Lachnospiraceae | Blautia | massiliensis |
| DA LEDD | 51 | ASV27 | ASV | Firmicutes | Ruminococcaceae | CAG-352 | NA |
| DA LEDD | 52 | ASV879 | ASV | Firmicutes | NA | NA | NA |
| DA LEDD | 53 | ASV75 | ASV | Firmicutes | Lachnospiraceae | [Ruminococcus] gauvreauii group | NA |
| DA LEDD | 54 | ASV522 | ASV | Firmicutes | UCG-010 | NA | NA |
| DA LEDD | 55 | ASV357 | ASV | Firmicutes | Lachnospiraceae | Marvinbryantia | NA |
| DA LEDD | 56 | ASV101 | ASV | Firmicutes | Lachnospiraceae | Roseburia | NA |
| DA LEDD | 57 | ASV443 | ASV | Firmicutes | Veillonellaceae | Veillonella | NA |
| DA LEDD | 58 | ASV127 | ASV | Firmicutes | Lachnospiraceae | Lachnospiraceae NK4A136 group | bacterium |
| DA LEDD | 59 | ASV262 | ASV | Firmicutes | Ruminococcaceae | Incertae Sedis | NA |

| Ticker | DESeq2_baseMean | DESeq2_log2-FoldChange | DESeq2_lfcSE | DESeq2_stat | DESeq2_pvalue | DESeq2_padj | DESeq2 converged |
|---|---|---|---|---|---|---|---|
| 30 | 53,93957092 | -0,005165459 | 0,007864256 | -0,656827431 | 0,511291884 | 0,997431792 | YES |
| 31 | 28,66619448 | 0,004280382 | 0,006896969 | 0,620617742 | 0,534851163 | 0,997431792 | YES |
| 32 | 71,07282003 | 0,003511315 | 0,005665288 | 0,619794538 | 0,535393065 | 0,997431792 | YES |
| 33 | 74,22539422 | 0,004622938 | 0,007497701 | 0,616580762 | 0,537511287 | 0,997431792 | YES |
| 34 | 79,77909046 | -0,005472549 | 0,009034335 | -0,605750121 | 0,544680696 | 0,997431792 | YES |
| 35 | 6,803883342 | 0,005375143 | 0,009082694 | 0,591800449 | 0,553984223 | 0,997431792 | YES |
| 36 | 753,7235241 | -0,005167758 | 0,008980508 | -0,575441647 | 0,564992644 | 0,997431792 | YES |
| 37 | 25,10210339 | 0,003493407 | 0,00619101 | 0,56427094 | 0,572569755 | 0,997431792 | YES |
| 38 | 27,0844115 | -0,003649098 | 0,006656542 | -0,548197151 | 0,583556538 | 0,997431792 | YES |
| 39 | 356,4491153 | 0,002873873 | 0,005374868 | 0,534687114 | 0,592866234 | 0,997431792 | YES |
| 40 | 82,44934701 | 0,004223915 | 0,007995323 | 0,528298264 | 0,597292336 | 0,997431792 | YES |
| 41 | 19,16140334 | 0,004690326 | 0,008910806 | 0,52636385 | 0,598635428 | 0,997431792 | YES |
| 42 | 18,62244929 | -0,004621177 | 0,008930588 | -0,517454953 | 0,604838607 | 0,997431792 | YES |
| 43 | 25,69401646 | 0,003769924 | 0,007405006 | 0,509104863 | 0,610678723 | 0,997431792 | YES |
| 44 | 50,19902288 | -0,004571416 | 0,009081126 | -0,503397413 | 0,61468489 | 0,997431792 | YES |
| 45 | 391,9776858 | -0,003589797 | 0,007166349 | -0,50092407 | 0,616424562 | 0,997431792 | YES |
| 46 | 24,13944265 | -0,004521531 | 0,009039544 | -0,500194567 | 0,616938083 | 0,997431792 | YES |
| 47 | 205,1227669 | 0,004430066 | 0,008932741 | 0,495935789 | 0,619939715 | 0,997431792 | YES |
| 48 | 311,7696625 | -0,002737303 | 0,00558278 | -0,490311792 | 0,623913284 | 0,997431792 | YES |
| 49 | 2342,640596 | 0,002413527 | 0,005015764 | 0,481188265 | 0,6303827 | 0,997431792 | YES |
| 50 | 451,7473085 | 0,002065242 | 0,004326364 | 0,477362002 | 0,633104367 | 0,997431792 | YES |
| 51 | 194,2826734 | 0,004348437 | 0,009156245 | 0,47491493 | 0,634847609 | 0,997431792 | YES |
| 52 | 1,473218583 | 0,004284208 | 0,009156594 | 0,467882287 | 0,639868771 | 0,997431792 | YES |
| 53 | 23,15202526 | 0,004180387 | 0,009148827 | 0,456931503 | 0,64772028 | 0,997431792 | YES |
| 54 | 4,105164263 | 0,004146276 | 0,009120801 | 0,45459558 | 0,649400215 | 0,997431792 | YES |
| 55 | 24,70018565 | 0,004106244 | 0,009066688 | 0,452893532 | 0,650625408 | 0,997431792 | YES |
| 56 | 53,73004373 | 0,004028468 | 0,009018331 | 0,446697763 | 0,655093297 | 0,997431792 | YES |
| 57 | 5,086397915 | -0,004060594 | 0,009146536 | -0,443948761 | 0,65707963 | 0,997431792 | YES |
| 58 | 223,0800973 | -0,003941235 | 0,008883788 | -0,44364351 | 0,657300343 | 0,997431792 | YES |
| 59 | 74,61123849 | 0,003206079 | 0,007400859 | 0,433203599 | 0,664866864 | 0,997431792 | YES |

| Medication | Ticker | ASVnr | taxLevel | Phylum | Family | Genus | Species |
|---|---|---|---|---|---|---|---|
| DA LEDD | 60 | ASV46 | ASV | Bacteroidota | Bacteroidaceae | Bacteroides | NA |
| DA LEDD | 61 | ASV26 | ASV | Euryarchaeota | Methanobacteriaceae | Methanobrevibacter | NA |
| DA LEDD | 62 | ASV66 | ASV | Firmicutes | Butyricicoccaceae | Butyricicoccus | NA |
| DA LEDD | 63 | ASV124 | ASV | Firmicutes | NA | NA | NA |
| DA LEDD | 64 | ASV895 | ASV | Bacteroidota | Barnesiellaceae | Coprobacter | secundus |
| DA LEDD | 65 | ASV85 | ASV | Firmicutes | Lachnospiraceae | Coprococcus | NA |
| DA LEDD | 66 | ASV508 | ASV | Firmicutes | Ruminococcaceae | NA | NA |
| DA LEDD | 67 | ASV216 | ASV | Firmicutes | Lachnospiraceae | Lachnospira | pectinoschiza |
| DA LEDD | 68 | ASV63 | ASV | Actinobacteriota | Eggerthellaceae | NA | NA |
| DA LEDD | 69 | ASV55 | ASV | Firmicutes | Monoglobaceae | Monoglobus | NA |
| DA LEDD | 70 | ASV230 | ASV | Firmicutes | Ruminococcaceae | [Eubacterium] siraeum group | NA |
| DA LEDD | 71 | ASV154 | ASV | Firmicutes | NA | NA | NA |
| DA LEDD | 72 | ASV363 | ASV | Bacteroidota | Rikenellaceae | Alistipes | NA |
| DA LEDD | 73 | ASV257 | ASV | Bacteroidota | Bacteroidaceae | Bacteroides | ovatus |
| DA LEDD | 74 | ASV867 | ASV | Firmicutes | NA | NA | NA |
| DA LEDD | 75 | ASV64 | ASV | Firmicutes | Oscillospiraceae | UCG-002 | NA |
| DA LEDD | 76 | ASV84 | ASV | Bacteroidota | Bacteroidaceae | Bacteroides | NA |
| DA LEDD | 77 | ASV172 | ASV | Proteobacteria | Pasteurellaceae | Haemophilus | NA |
| DA LEDD | 78 | ASV242 | ASV | Firmicutes | Lachnospiraceae | Tyzzerella | NA |
| DA LEDD | 79 | ASV137 | ASV | Firmicutes | Butyricicoccaceae | Butyricicoccus | faecihominis |
| DA LEDD | 80 | ASV213 | ASV | Firmicutes | NA | NA | NA |
| DA LEDD | 81 | ASV189 | ASV | Firmicutes | Ruminococcaceae | Ruminococcus | champanellensis |
| DA LEDD | 82 | ASV48 | ASV | Bacteroidota | Bacteroidaceae | Bacteroides | caccae |
| DA LEDD | 83 | ASV33 | ASV | Firmicutes | Peptostreptococcaceae | Intestinibacter | bartlettii |
| DA LEDD | 84 | ASV470 | ASV | Firmicutes | Christensenellaceae | Christensenellaceae R-7 group | NA |
| DA LEDD | 85 | ASV457 | ASV | Firmicutes | Lachnospiraceae | NA | NA |
| DA LEDD | 86 | ASV665 | ASV | Firmicutes | Lachnospiraceae | NA | NA |
| DA LEDD | 87 | ASV128 | ASV | Firmicutes | Oscillospiraceae | NK4A214 group | NA |
| DA LEDD | 88 | ASV22 | ASV | Bacteroidota | Bacteroidaceae | Bacteroides | uniformis |
| DA LEDD | 89 | ASV100 | ASV | Firmicutes | Christensenellaceae | NA | NA |

| Ticker | DESeq2_baseMean | DESeq2_log2-FoldChange | DESeq2_lfcSE | DESeq2_stat | DESeq2_pvalue | DESeq2_padj | DESeq2 converged |
|---|---|---|---|---|---|---|---|
| 60 | 159,5331272 | -0,002165949 | 0,005032194 | -0,430418351 | 0,666891348 | 0,997431792 | YES |
| 61 | 78,94475221 | 0,002185361 | 0,005149877 | 0,424352129 | 0,671309027 | 0,997431792 | YES |
| 62 | 124,0117155 | 0,002557598 | 0,006111616 | 0,418481375 | 0,675595201 | 0,997431792 | YES |
| 63 | 168,5745109 | 0,0036386 | 0,009034295 | 0,402754199 | 0,687129058 | 0,997431792 | YES |
| 64 | 4,352557783 | 0,003667344 | 0,009172748 | 0,399808635 | 0,689297471 | 0,997431792 | YES |
| 65 | 333,6328387 | 0,003632033 | 0,009108994 | 0,398730365 | 0,690091891 | 0,997431792 | YES |
| 66 | 4,01270662 | 0,003647009 | 0,009155421 | 0,398344235 | 0,690376458 | 0,997431792 | YES |
| 67 | 68,96140702 | -0,002652929 | 0,006691866 | -0,396440856 | 0,691779831 | 0,997431792 | YES |
| 68 | 128,7041458 | 0,002181715 | 0,005756563 | 0,378996072 | 0,70469078 | 0,997431792 | YES |
| 69 | 117,3142266 | -0,00181519 | 0,004802697 | -0,377952286 | 0,70546604 | 0,997431792 | YES |
| 70 | 20,2441453 | -0,003414307 | 0,009052934 | -0,377149197 | 0,706062732 | 0,997431792 | YES |
| 71 | 47,83749254 | 0,003367552 | 0,009020141 | 0,373336953 | 0,708897677 | 0,997431792 | YES |
| 72 | 19,71155851 | 0,003304161 | 0,008968809 | 0,368405799 | 0,712570674 | 0,997431792 | YES |
| 73 | 50,21470141 | -0,003347093 | 0,009110995 | -0,367368547 | 0,71334413 | 0,997431792 | YES |
| 74 | 1,116911288 | -0,003352633 | 0,009206735 | -0,364150006 | 0,715745997 | 0,997431792 | YES |
| 75 | 145,5328527 | 0,002706798 | 0,007484138 | 0,361671315 | 0,717597666 | 0,997431792 | YES |
| 76 | 110,7148008 | -0,00322277 | 0,008984477 | -0,358704247 | 0,719816351 | 0,997431792 | YES |
| 77 | 37,11322796 | -0,003247726 | 0,009064983 | -0,358271605 | 0,720140066 | 0,997431792 | YES |
| 78 | 14,16800105 | 0,003254769 | 0,00915973 | 0,355334619 | 0,722338923 | 0,997431792 | YES |
| 79 | 11,06828922 | 0,003170188 | 0,009122406 | 0,347516686 | 0,728203185 | 0,997431792 | YES |
| 80 | 40,74832547 | 0,003101201 | 0,009034024 | 0,343280079 | 0,731387766 | 0,997431792 | YES |
| 81 | 267,4217133 | -0,003026536 | 0,009042622 | -0,334696778 | 0,737853832 | 0,997431792 | YES |
| 82 | 446,0773252 | -0,001704946 | 0,005123591 | -0,332763888 | 0,73931252 | 0,997431792 | YES |
| 83 | 47,48497571 | -0,002141738 | 0,00645926 | -0,331576357 | 0,740209175 | 0,997431792 | YES |
| 84 | 14,74533947 | -0,00293695 | 0,009040634 | -0,324861113 | 0,745286189 | 0,997431792 | YES |
| 85 | 2,447124746 | 0,002871766 | 0,009165939 | 0,31330842 | 0,754046354 | 0,997431792 | YES |
| 86 | 2,039270621 | 0,00286803 | 0,009167239 | 0,312856513 | 0,754389678 | 0,997431792 | YES |
| 87 | 78,55255695 | 0,002793412 | 0,00907881 | 0,307684784 | 0,758322199 | 0,997431792 | YES |
| 88 | 729,6551081 | -0,001408781 | 0,004601823 | -0,306135309 | 0,759501625 | 0,997431792 | YES |
| 89 | 145,4880729 | 0,002728631 | 0,009050751 | 0,301481151 | 0,763047621 | 0,997431792 | YES |

| Medication | Ticker | ASVnr | taxLevel | Phylum | Family | Genus | Species |
|---|---|---|---|---|---|---|---|
| DA LEDD | 90 | ASV47 | ASV | Bacteroidota | Rikenellaceae | Alistipes | NA |
| DA LEDD | 91 | ASV806 | ASV | Firmicutes | Peptococcaceae | NA | NA |
| DA LEDD | 92 | ASV53 | ASV | Firmicutes | Lachnospiraceae | [Ruminococcus] torques group | NA |
| DA LEDD | 93 | ASV687 | ASV | Firmicutes | Ruminococcaceae | DTU089 | NA |
| DA LEDD | 94 | ASV144 | ASV | Desulfobacterota | Desulfovibrionaceae | Bilophila | wadsworthia |
| DA LEDD | 95 | ASV141 | ASV | Firmicutes | Lachnospiraceae | Lachnoclostridium | NA |
| DA LEDD | 96 | ASV701 | ASV | Firmicutes | Lachnospiraceae | Lachnoclostridium | NA |
| DA LEDD | 97 | ASV86 | ASV | Firmicutes | Lachnospiraceae | [Eubacterium] hallii group | NA |
| DA LEDD | 98 | ASV406 | ASV | Firmicutes | NA | NA | NA |
| DA LEDD | 99 | ASV234 | ASV | Cyanobacteria | NA | NA | NA |
| DA LEDD | 100 | ASV135 | ASV | Bacteroidota | Rikenellaceae | Alistipes | inops |
| DA LEDD | 101 | ASV184 | ASV | Firmicutes | Ruminococcaceae | Ruminococcus | NA |
| DA LEDD | 102 | ASV77 | ASV | Firmicutes | Lachnospiraceae | [Ruminococcus] torques group | NA |
| DA LEDD | 103 | ASV521 | ASV | Firmicutes | Lachnospiraceae | [Eubacterium] ventriosum group | NA |
| DA LEDD | 104 | ASV56 | ASV | Bacteroidota | Bacteroidaceae | Bacteroides | massiliensis |
| DA LEDD | 105 | ASV134 | ASV | Actinobacteriota | Eggerthellaceae | Enterorhabdus | NA |
| DA LEDD | 106 | ASV238 | ASV | Firmicutes | Butyricicoccaceae | NA | NA |
| DA LEDD | 107 | ASV126 | ASV | Bacteroidota | Tannerellaceae | Parabacteroides | distasonis |
| DA LEDD | 108 | ASV74 | ASV | Firmicutes | Lachnospiraceae | Coprococcus | catus |
| DA LEDD | 109 | ASV42 | ASV | Actinobacteriota | Bifidobacteriaceae | Bifidobacterium | NA |
| DA LEDD | 110 | ASV337 | ASV | Firmicutes | Lachnospiraceae | Lachnoclostridium | NA |
| DA LEDD | 111 | ASV12 | ASV | Firmicutes | Veillonellaceae | Dialister | invisus |
| DA LEDD | 112 | ASV253 | ASV | Bacteroidota | Tannerellaceae | Parabacteroides | distasonis |
| DA LEDD | 113 | ASV503 | ASV | Firmicutes | Erysipelatoclostridiaceae | Erysipelatoclostridium | NA |
| DA LEDD | 114 | ASV133 | ASV | Actinobacteriota | Eggerthellaceae | Senegalimassilia | anaerobia |
| DA LEDD | 115 | ASV190 | ASV | Firmicutes | Ruminococcaceae | UBA1819 | NA |
| DA LEDD | 116 | ASV314 | ASV | Firmicutes | Ruminococcaceae | Subdoligranulum | NA |
| DA LEDD | 117 | ASV626 | ASV | Firmicutes | NA | NA | NA |
| DA LEDD | 118 | ASV119 | ASV | Firmicutes | Ruminococcaceae | Subdoligranulum | NA |
| DA LEDD | 119 | ASV143 | ASV | Firmicutes | Peptostreptococcaceae | Terrisporobacter | mayombei |

| Ticker | DESeq2_baseMean | DESeq2_log2-FoldChange | DESeq2_lfcSE | DESeq2_stat | DESeq2_pvalue | DESeq2_padj | DESeq2 converged |
|---|---|---|---|---|---|---|---|
| 90 | 232,3846727 | 0,002690558 | 0,008931029 | 0,301259525 | 0,763216602 | 0,997431792 | YES |
| 91 | 17,07699902 | -0,001966476 | 0,006575872 | -0,299044148 | 0,764906361 | 0,997431792 | YES |
| 92 | 403,5003596 | -0,001424798 | 0,004768578 | -0,298788799 | 0,765101198 | 0,997431792 | YES |
| 93 | 18,05574172 | 0,002717774 | 0,009113549 | 0,298212445 | 0,765541024 | 0,997431792 | YES |
| 94 | 54,48100851 | -0,001852217 | 0,006274933 | -0,29517719 | 0,767858526 | 0,997431792 | YES |
| 95 | 10,93357082 | 0,002680808 | 0,009100162 | 0,294589072 | 0,768307811 | 0,997431792 | YES |
| 96 | 37,13206487 | 0,002584108 | 0,008914267 | 0,289884496 | 0,771904603 | 0,997431792 | YES |
| 97 | 166,9880699 | 0,001570875 | 0,005431641 | 0,28920824 | 0,772422026 | 0,997431792 | YES |
| 98 | 5,98221351 | 0,002088658 | 0,007259031 | 0,287732357 | 0,77355162 | 0,997431792 | YES |
| 99 | 10,34967123 | -0,002575137 | 0,00911398 | -0,282547989 | 0,777523355 | 0,997431792 | YES |
| 100 | 141,1948633 | 0,002432668 | 0,008684927 | 0,280102337 | 0,779398992 | 0,997431792 | YES |
| 101 | 187,8251628 | 0,002511217 | 0,009127343 | 0,275131195 | 0,783215446 | 0,997431792 | YES |
| 102 | 250,8500642 | 0,002051861 | 0,00753117 | 0,272449157 | 0,78527668 | 0,997431792 | YES |
| 103 | 10,30099633 | -0,002454175 | 0,009209685 | -0,266477681 | 0,789871347 | 0,997431792 | YES |
| 104 | 143,7464759 | -0,002409226 | 0,009055819 | -0,266041786 | 0,790207028 | 0,997431792 | YES |
| 105 | 10,16755101 | -0,001891935 | 0,007114442 | -0,265928767 | 0,79029407 | 0,997431792 | YES |
| 106 | 6,050103979 | 0,002431423 | 0,009156779 | 0,265532508 | 0,790599271 | 0,997431792 | YES |
| 107 | 111,0222979 | 0,002047994 | 0,00771987 | 0,265288703 | 0,790787067 | 0,997431792 | YES |
| 108 | 28,27143501 | -0,001739824 | 0,006746172 | -0,257897909 | 0,796485699 | 0,997431792 | YES |
| 109 | 60,7130086 | 0,00238046 | 0,0092623 | 0,257005283 | 0,797174695 | 0,997431792 | YES |
| 110 | 22,39241141 | -0,002276016 | 0,009074957 | -0,250801822 | 0,801967333 | 0,997431792 | YES |
| 111 | 1017,182378 | 0,002235246 | 0,00895855 | 0,249509826 | 0,802966441 | 0,997431792 | YES |
| 112 | 27,91939325 | -0,002236073 | 0,009113982 | -0,245345334 | 0,806189053 | 0,997431792 | YES |
| 113 | 0,473939308 | -0,002243067 | 0,009187644 | -0,244139476 | 0,8071228 | 0,997431792 | YES |
| 114 | 6,284731383 | 0,002219964 | 0,009167946 | 0,242144109 | 0,808668501 | 0,997431792 | YES |
| 115 | 28,72913582 | 0,001787509 | 0,007470371 | 0,239279863 | 0,810888579 | 0,997431792 | YES |
| 116 | 11,10269959 | 0,002135844 | 0,009217121 | 0,231725705 | 0,816751065 | 0,997431792 | YES |
| 117 | 8,33456202 | -0,002074691 | 0,009161799 | -0,226450151 | 0,820851326 | 0,997431792 | YES |
| 118 | 10,53852819 | -0,002088515 | 0,009228656 | -0,2263076 | 0,820962188 | 0,997431792 | YES |
| 119 | 7,678192803 | -0,002086287 | 0,009229169 | -0,226053603 | 0,82115973 | 0,997431792 | YES |

| Medication | Ticker | ASVnr | taxLevel | Phylum | Family | Genus | Species |
|---|---|---|---|---|---|---|---|
| DA LEDD | 120 | ASV201 | ASV | Firmicutes | Oscillospiraceae | UCG-002 | NA |
| DA LEDD | 121 | ASV255 | ASV | Actinobacteriota | Coriobacteriales Incertae Sedis | NA | NA |
| DA LEDD | 122 | ASV427 | ASV | Firmicutes | Lactobacillaceae | Lactobacillus | NA |
| DA LEDD | 123 | ASV656 | ASV | Firmicutes | Oscillospiraceae | Flavonifractor | NA |
| DA LEDD | 124 | ASV249 | ASV | Proteobacteria | NA | NA | NA |
| DA LEDD | 125 | ASV505 | ASV | Bacteroidota | Bacteroidaceae | Bacteroides | nordii |
| DA LEDD | 126 | ASV191 | ASV | Firmicutes | Lachnospiraceae | Lachnospiraceae NK4A136 group | NA |
| DA LEDD | 127 | ASV339 | ASV | Firmicutes | Monoglobaceae | Monoglobus | pectinilyticus |
| DA LEDD | 128 | ASV206 | ASV | Firmicutes | Oscillospiraceae | Intestinimonas | NA |
| DA LEDD | 129 | ASV147 | ASV | Firmicutes | Oscillospiraceae | Oscillibacter | NA |
| DA LEDD | 130 | ASV730 | ASV | Firmicutes | Ruminococcaceae | NA | NA |
| DA LEDD | 131 | ASV477 | ASV | Firmicutes | Oscillospiraceae | Intestinimonas | massiliensis |
| DA LEDD | 132 | ASV270 | ASV | Firmicutes | Christensenellaceae | Christensenellaceae R-7 group | NA |
| DA LEDD | 133 | ASV283 | ASV | Actinobacteriota | Eggerthellaceae | Slackia | isoflavoniconvertens |
| DA LEDD | 134 | ASV52 | ASV | Firmicutes | Lachnospiraceae | Coprococcus | eutactus |
| DA LEDD | 135 | ASV557 | ASV | Firmicutes | Oscillospiraceae | UCG-005 | NA |
| DA LEDD | 136 | ASV421 | ASV | Firmicutes | NA | NA | NA |
| DA LEDD | 137 | ASV132 | ASV | Firmicutes | Ruminococcaceae | CAG-352 | NA |
| DA LEDD | 138 | ASV383 | ASV | Firmicutes | NA | NA | NA |
| DA LEDD | 139 | ASV382 | ASV | Firmicutes | NA | NA | NA |
| DA LEDD | 140 | ASV554 | ASV | Firmicutes | Lachnospiraceae | NA | NA |
| DA LEDD | 141 | ASV417 | ASV | Firmicutes | UCG-010 | NA | NA |
| DA LEDD | 142 | ASV202 | ASV | Firmicutes | Lachnospiraceae | [Eubacterium] hallii group | NA |
| DA LEDD | 143 | ASV690 | ASV | Firmicutes | Ruminococcaceae | Fournierella | NA |
| DA LEDD | 144 | ASV203 | ASV | Firmicutes | NA | NA | NA |
| DA LEDD | 145 | ASV19 | ASV | Firmicutes | Erysipelatoclostridiaceae | Erysipelotrichaceae UCG-003 | bacterium |
| DA LEDD | 146 | ASV1086 | ASV | Firmicutes | UCG-010 | NA | NA |
| DA LEDD | 147 | ASV1158 | ASV | Firmicutes | Ruminococcaceae | NA | NA |
| DA LEDD | 148 | ASV81 | ASV | Firmicutes | Lachnospiraceae | Lachnospiraceae NK4A136 group | NA |
| DA LEDD | 149 | ASV142 | ASV | Firmicutes | Ruminococcaceae | Faecalibacterium | prausnitzii |

| Ticker | DESeq2_baseMean | DESeq2_log2-FoldChange | DESeq2_lfcSE | DESeq2_stat | DESeq2_pvalue | DESeq2_padj | DESeq2 converged |
|---|---|---|---|---|---|---|---|
| 120 | 108,2671864 | 0,00200939 | 0,009076049 | 0,221394766 | 0,824785068 | 0,997431792 | YES |
| 121 | 14,05825884 | 0,002026462 | 0,009156522 | 0,22131353 | 0,824848316 | 0,997431792 | YES |
| 122 | 0,58311163 | 0,002029788 | 0,009178365 | 0,221149248 | 0,824976225 | 0,997431792 | YES |
| 123 | 5,053294801 | -0,002015201 | 0,009131183 | -0,220694379 | 0,825330408 | 0,997431792 | YES |
| 124 | 34,79510218 | 0,002031042 | 0,00922165 | 0,220247139 | 0,825678686 | 0,997431792 | YES |
| 125 | 3,435030008 | 0,002021002 | 0,009191822 | 0,219869536 | 0,825972763 | 0,997431792 | YES |
| 126 | 76,22456119 | 0,0020039 | 0,009155534 | 0,218873137 | 0,826748873 | 0,997431792 | YES |
| 127 | 27,54735273 | 0,00193085 | 0,0090338 | 0,213736141 | 0,830752826 | 0,997431792 | YES |
| 128 | 65,44470241 | 0,001690747 | 0,007917692 | 0,213540335 | 0,830905532 | 0,997431792 | YES |
| 129 | 147,7543168 | -0,001234561 | 0,005835601 | -0,211556763 | 0,832452844 | 0,997431792 | YES |
| 130 | 2,795814763 | 0,001931447 | 0,009247148 | 0,208869525 | 0,834550095 | 0,997431792 | YES |
| 131 | 21,72295123 | 0,001880393 | 0,009159313 | 0,20529845 | 0,837338955 | 0,997431792 | YES |
| 132 | 24,43761099 | 0,001085944 | 0,005335181 | 0,203544054 | 0,838709816 | 0,997431792 | YES |
| 133 | 4,32930549 | 0,001828184 | 0,009113445 | 0,200602967 | 0,841009038 | 0,997431792 | YES |
| 134 | 11,21988997 | 0,001840768 | 0,009224552 | 0,19955097 | 0,841831776 | 0,997431792 | YES |
| 135 | 8,378009947 | 0,001840398 | 0,009224675 | 0,199508124 | 0,841865289 | 0,997431792 | YES |
| 136 | 4,982516929 | 0,001839404 | 0,009225006 | 0,199393241 | 0,841955147 | 0,997431792 | YES |
| 137 | 4,576534068 | 0,001839189 | 0,009225078 | 0,199368352 | 0,841974615 | 0,997431792 | YES |
| 138 | 1,550116378 | 0,001834066 | 0,009226803 | 0,198775918 | 0,842438035 | 0,997431792 | YES |
| 139 | 1,291763648 | 0,001832533 | 0,009227322 | 0,198598575 | 0,842576769 | 0,997431792 | YES |
| 140 | 0,775058189 | 0,001826455 | 0,009229384 | 0,197895671 | 0,843126691 | 0,997431792 | YES |
| 141 | 0,627428058 | 0,00182292 | 0,009230588 | 0,197486871 | 0,843446554 | 0,997431792 | YES |
| 142 | 0,553612992 | 0,001820464 | 0,009231426 | 0,197202879 | 0,843668778 | 0,997431792 | YES |
| 143 | 0,332167795 | 0,001806801 | 0,009236112 | 0,195623591 | 0,844904794 | 0,997431792 | YES |
| 144 | 0,295260262 | 0,001802619 | 0,009237555 | 0,195140258 | 0,845283147 | 0,997431792 | YES |
| 145 | 143,7463084 | -0,001315713 | 0,006771177 | -0,194310893 | 0,845932457 | 0,997431792 | YES |
| 146 | 0,184537664 | 0,001780701 | 0,009245178 | 0,192608631 | 0,847265485 | 0,997431792 | YES |
| 147 | 2,083762464 | 0,001749348 | 0,009164692 | 0,190879031 | 0,848620369 | 0,997431792 | YES |
| 148 | 132,7178957 | 0,001708749 | 0,009170756 | 0,186325809 | 0,852189269 | 0,997431792 | YES |
| 149 | 60,19932348 | -0,001660571 | 0,009111075 | -0,182258477 | 0,855379884 | 0,997431792 | YES |

| Medication | Ticker | ASVnr | taxLevel | Phylum | Family | Genus | Species |
|---|---|---|---|---|---|---|---|
| DA LEDD | 150 | ASV59 | ASV | Firmicutes | [Eubacterium] coprostanoligenes group | NA | NA |
| DA LEDD | 151 | ASV149 | ASV | Firmicutes | Oscillospiraceae | UCG-002 | NA |
| DA LEDD | 152 | ASV529 | ASV | Firmicutes | Lachnospiraceae | NA | NA |
| DA LEDD | 153 | ASV351 | ASV | Firmicutes | Lachnospiraceae | Roseburia | NA |
| DA LEDD | 154 | ASV808 | ASV | Desulfobacterota | Desulfovibrionaceae | NA | NA |
| DA LEDD | 155 | ASV49 | ASV | Bacteroidota | Bacteroidaceae | Bacteroides | NA |
| DA LEDD | 156 | ASV291 | ASV | Firmicutes | Erysipelatoclostridiaceae | Erysipelatoclostridium | NA |
| DA LEDD | 157 | ASV24 | ASV | Bacteroidota | Bacteroidaceae | Bacteroides | uniformis |
| DA LEDD | 158 | ASV386 | ASV | Firmicutes | Ruminococcaceae | DTU089 | NA |
| DA LEDD | 159 | ASV93 | ASV | Firmicutes | Oscillospiraceae | NK4A214 group | NA |
| DA LEDD | 160 | ASV68 | ASV | Firmicutes | Ruminococcaceae | Faecalibacterium | NA |
| DA LEDD | 161 | ASV34 | ASV | Firmicutes | Lachnospiraceae | Anaerostipes | hadrus |
| DA LEDD | 162 | ASV533 | ASV | Firmicutes | Ruminococcaceae | Negativibacillus | massiliensis |
| DA LEDD | 163 | ASV615 | ASV | Firmicutes | Oscillospiraceae | Oscillibacter | NA |
| DA LEDD | 164 | ASV211 | ASV | Firmicutes | Lachnospiraceae | Lachnospiraceae FCS020 group | NA |
| DA LEDD | 165 | ASV449 | ASV | Actinobacteriota | Atopobiaceae | Olsenella | NA |
| DA LEDD | 166 | ASV120 | ASV | Firmicutes | Lachnospiraceae | Anaerostipes | hadrus |
| DA LEDD | 167 | ASV65 | ASV | Firmicutes | Ruminococcaceae | Incertae Sedis | NA |
| DA LEDD | 168 | ASV198 | ASV | Bacteroidota | Bacteroidaceae | Bacteroides | eggerthii |
| DA LEDD | 169 | ASV481 | ASV | Firmicutes | Lachnospiraceae | [Eubacterium] xylanophilum group | NA |
| DA LEDD | 170 | ASV62 | ASV | Firmicutes | NA | NA | NA |
| DA LEDD | 171 | ASV107 | ASV | Bacteroidota | Bacteroidaceae | Bacteroides | eggerthii |
| DA LEDD | 172 | ASV17 | ASV | Actinobacteriota | Coriobacteriaceae | Collinsella | aerofaciens |
| DA LEDD | 173 | ASV243 | ASV | Firmicutes | NA | NA | NA |
| DA LEDD | 174 | ASV123 | ASV | Firmicutes | Lachnospiraceae | [Ruminococcus] torques group | NA |
| DA LEDD | 175 | ASV435 | ASV | Firmicutes | Christensenellaceae | Christensenellaceae R-7 group | NA |
| DA LEDD | 176 | ASV451 | ASV | Firmicutes | Oscillospiraceae | UCG-005 | NA |
| DA LEDD | 177 | ASV415 | ASV | Firmicutes | Oscillospiraceae | Oscillospira | NA |
| DA LEDD | 178 | ASV45 | ASV | Firmicutes | Lachnospiraceae | Coprococcus | comes |
| DA LEDD | 179 | ASV219 | ASV | Firmicutes | Lachnospiraceae | Lachnospiraceae UCG-004 | NA |

| Ticker | DESeq2_baseMean | DESeq2_log2-FoldChange | DESeq2_lfcSE | DESeq2_stat | DESeq2_pvalue | DESeq2_padj | DESeq2 converged |
|---|---|---|---|---|---|---|---|
| 150 | 49,13549751 | 0,00162762 | 0,009110402 | 0,178655096 | 0,85820853 | 0,997431792 | YES |
| 151 | 46,70146365 | 0,001633654 | 0,009175657 | 0,178042154 | 0,858689871 | 0,997431792 | YES |
| 152 | 9,484818942 | -0,001620597 | 0,009216893 | -0,175828992 | 0,860428292 | 0,997431792 | YES |
| 153 | 19,79554004 | 0,001606597 | 0,009224383 | 0,17416849 | 0,861733048 | 0,997431792 | YES |
| 154 | 0,751315944 | 0,001608567 | 0,009263725 | 0,173641483 | 0,862147228 | 0,997431792 | YES |
| 155 | 782,0433039 | -0,001337514 | 0,007706397 | -0,173558922 | 0,862212118 | 0,997431792 | YES |
| 156 | 11,29170044 | 0,001530243 | 0,009017486 | 0,169697261 | 0,865248228 | 0,997431792 | YES |
| 157 | 208,0200067 | -0,001135098 | 0,006836152 | -0,166043374 | 0,868122816 | 0,997431792 | YES |
| 158 | 22,20999818 | 0,001264234 | 0,007667248 | 0,164887544 | 0,869032497 | 0,997431792 | YES |
| 159 | 55,39903495 | 0,001246756 | 0,007577867 | 0,164525966 | 0,869317107 | 0,997431792 | YES |
| 160 | 37,65228701 | -0,00150481 | 0,009160133 | -0,164278218 | 0,869512128 | 0,997431792 | YES |
| 161 | 104,897279 | 0,000845286 | 0,00517036 | 0,163486808 | 0,870135159 | 0,997431792 | YES |
| 162 | 23,55148509 | -0,001484418 | 0,0090835 | -0,16341919 | 0,870188394 | 0,997431792 | YES |
| 163 | 4,695124639 | -0,001495726 | 0,009161698 | -0,163258574 | 0,870314849 | 0,997431792 | YES |
| 164 | 27,74552205 | 0,000936506 | 0,005741297 | 0,16311754 | 0,870425889 | 0,997431792 | YES |
| 165 | 1,551705206 | 0,001490102 | 0,009245114 | 0,161177273 | 0,871953781 | 0,997431792 | YES |
| 166 | 74,08795427 | -0,001449 | 0,009075715 | -0,159656861 | 0,873151385 | 0,997431792 | YES |
| 167 | 57,80571486 | 0,001231586 | 0,007740824 | 0,159102663 | 0,87358799 | 0,997431792 | YES |
| 168 | 214,0966625 | -0,00145555 | 0,009155341 | -0,158983756 | 0,873681672 | 0,997431792 | YES |
| 169 | 10,75637349 | -0,001451482 | 0,009145101 | -0,15871686 | 0,873891954 | 0,997431792 | YES |
| 170 | 15,65657266 | -0,001449276 | 0,009240705 | -0,156836105 | 0,875374017 | 0,997431792 | YES |
| 171 | 134,4484627 | -0,001433167 | 0,009155359 | -0,15653863 | 0,875608472 | 0,997431792 | YES |
| 172 | 502,9478624 | 0,000664803 | 0,00424703 | 0,156533683 | 0,875612372 | 0,997431792 | YES |
| 173 | 2,634519437 | -0,001434101 | 0,009244415 | -0,155131651 | 0,876717531 | 0,997431792 | YES |
| 174 | 18,34931008 | 0,001410115 | 0,009146075 | 0,154177065 | 0,877470125 | 0,997431792 | YES |
| 175 | 1,354895711 | -0,001417795 | 0,009248499 | -0,153300024 | 0,878161681 | 0,997431792 | YES |
| 176 | 28,07579217 | 0,001399551 | 0,009170552 | 0,152613626 | 0,878702979 | 0,997431792 | YES |
| 177 | 29,91155672 | -0,000999307 | 0,006568398 | -0,152138682 | 0,879077555 | 0,997431792 | YES |
| 178 | 71,6824924 | -0,001101726 | 0,007300108 | -0,150919097 | 0,880039536 | 0,997431792 | YES |
| 179 | 30,95444734 | -0,001364287 | 0,009082669 | -0,150207714 | 0,88060074 | 0,997431792 | YES |

| Medication | Ticker | ASVnr | taxLevel | Phylum | Family | Genus | Species |
|---|---|---|---|---|---|---|---|
| DA LEDD | 180 | ASV666 | ASV | Firmicutes | Erysipelotrichaceae | Holdemania | filiformis |
| DA LEDD | 181 | ASV441 | ASV | Firmicutes | [Eubacterium] coprostanoligenes group | NA | NA |
| DA LEDD | 182 | ASV171 | ASV | Firmicutes | Oscillospiraceae | NA | NA |
| DA LEDD | 183 | ASV41 | ASV | Firmicutes | Ruminococcaceae | Ruminococcus | bicirculans |
| DA LEDD | 184 | ASV221 | ASV | Firmicutes | Oscillospiraceae | Intestinimonas | NA |
| DA LEDD | 185 | ASV214 | ASV | Firmicutes | Lachnospiraceae | Lachnospiraceae UCG-010 | NA |
| DA LEDD | 186 | ASV89 | ASV | Firmicutes | Lachnospiraceae | Blautia | NA |
| DA LEDD | 187 | ASV675 | ASV | Bacteroidota | Prevotellaceae | NA | NA |
| DA LEDD | 188 | ASV78 | ASV | Firmicutes | Lachnospiraceae | [Eubacterium] eligens group | NA |
| DA LEDD | 189 | ASV258 | ASV | Firmicutes | Lachnospiraceae | Lachnospiraceae AC2044 group | NA |
| DA LEDD | 190 | ASV139 | ASV | Firmicutes | Christensenellaceae | Christensenellaceae R-7 group | NA |
| DA LEDD | 191 | ASV125 | ASV | Firmicutes | Oscillospiraceae | UCG-002 | NA |
| DA LEDD | 192 | ASV367 | ASV | Bacteroidota | Rikenellaceae | Alistipes | NA |
| DA LEDD | 193 | ASV324 | ASV | Proteobacteria | NA | NA | NA |
| DA LEDD | 194 | ASV29 | ASV | Firmicutes | Oscillospiraceae | UCG-002 | NA |
| DA LEDD | 195 | ASV145 | ASV | Bacteroidota | Muribaculaceae | NA | NA |
| DA LEDD | 196 | ASV43 | ASV | Firmicutes | Lachnospiraceae | Blautia | faecis |
| DA LEDD | 197 | ASV813 | ASV | Firmicutes | Ruminococcaceae | NA | NA |
| DA LEDD | 198 | ASV28 | ASV | Firmicutes | Clostridiaceae | Clostridium sensu stricto 1 | NA |
| DA LEDD | 199 | ASV247 | ASV | Euryarchaeota | Methanobacteriaceae | Methanosphaera | NA |
| DA LEDD | 200 | ASV8 | ASV | Firmicutes | Ruminococcaceae | Faecalibacterium | prausnitzii |
| DA LEDD | 201 | ASV99 | ASV | Bacteroidota | Bacteroidaceae | Bacteroides | plebeius |
| DA LEDD | 202 | ASV296 | ASV | Proteobacteria | Sutterellaceae | Sutterella | NA |
| DA LEDD | 203 | ASV117 | ASV | Bacteroidota | Rikenellaceae | Alistipes | finegoldii |
| DA LEDD | 204 | ASV355 | ASV | Firmicutes | NA | NA | NA |
| DA LEDD | 205 | ASV509 | ASV | Firmicutes | Lachnospiraceae | Marvinbryantia | NA |
| DA LEDD | 206 | ASV836 | ASV | Verrucomicrobiota | Victivallaceae | Victivallis | vadensis |
| DA LEDD | 207 | ASV18 | ASV | Firmicutes | Lachnospiraceae | Dorea | longicatena |
| DA LEDD | 208 | ASV288 | ASV | Firmicutes | Erysipelotrichaceae | NA | NA |
| DA LEDD | 209 | ASV94 | ASV | Bacteroidota | Rikenellaceae | Alistipes | shahii |

| Ticker | DESeq2_baseMean | DESeq2_log2-FoldChange | DESeq2_lfcSE | DESeq2_stat | DESeq2_pvalue | DESeq2_padj | DESeq2 converged |
|---|---|---|---|---|---|---|---|
| 180 | 5,005656773 | 0,001206556 | 0,008111011 | 0,148755286 | 0,881746734 | 0,997431792 | YES |
| 181 | 9,544460345 | -0,001360496 | 0,009176922 | -0,148251913 | 0,882143964 | 0,997431792 | YES |
| 182 | 38,81620558 | 0,001339099 | 0,009113928 | 0,146928833 | 0,883188193 | 0,997431792 | YES |
| 183 | 200,0878474 | -0,001072871 | 0,007316746 | -0,146632206 | 0,883422332 | 0,997431792 | YES |
| 184 | 44,37254422 | -0,00085534 | 0,005847291 | -0,146279709 | 0,883700584 | 0,997431792 | YES |
| 185 | 31,64309664 | -0,001333213 | 0,009176986 | -0,145277869 | 0,884491487 | 0,997431792 | YES |
| 186 | 30,38632518 | -0,0013247 | 0,009249596 | -0,143217073 | 0,886118746 | 0,997431792 | YES |
| 187 | 50,04995218 | 0,00129001 | 0,00922426 | 0,139849672 | 0,888778766 | 0,997431792 | YES |
| 188 | 89,34026653 | -0,001007348 | 0,007228383 | -0,139360035 | 0,889165652 | 0,997431792 | YES |
| 189 | 5,116219409 | 0,00126799 | 0,009211171 | 0,137657828 | 0,89051085 | 0,997431792 | YES |
| 190 | 10,7821158 | 0,001263242 | 0,009211171 | 0,137142356 | 0,890918274 | 0,997431792 | YES |
| 191 | 17,2853955 | 0,001262558 | 0,00921129 | 0,137066342 | 0,890978357 | 0,997431792 | YES |
| 192 | 3,48410036 | 0,001252866 | 0,009273769 | 0,135097801 | 0,892534548 | 0,997431792 | YES |
| 193 | 3,371579368 | 0,001217487 | 0,00926153 | 0,131456302 | 0,895414352 | 0,997431792 | YES |
| 194 | 411,8775246 | 0,000882515 | 0,006820387 | 0,129393622 | 0,897046194 | 0,997431792 | YES |
| 195 | 304,0847259 | -0,001189417 | 0,009207145 | -0,12918415 | 0,897211938 | 0,997431792 | YES |
| 196 | 257,355953 | -0,001121434 | 0,008798301 | -0,12746023 | 0,898576148 | 0,997431792 | YES |
| 197 | 4,403870367 | 0,001165537 | 0,009257316 | 0,125904362 | 0,89980763 | 0,997431792 | YES |
| 198 | 41,82297384 | 0,000955184 | 0,007648837 | 0,124879678 | 0,900618807 | 0,997431792 | YES |
| 199 | 5,878889909 | 0,001150034 | 0,009213446 | 0,124821216 | 0,900665091 | 0,997431792 | YES |
| 200 | 605,778642 | -0,000716455 | 0,005781614 | -0,123919617 | 0,90137892 | 0,997431792 | YES |
| 201 | 81,97677146 | 0,001102663 | 0,009202024 | 0,119828321 | 0,904619146 | 0,997431792 | YES |
| 202 | 11,4823769 | 0,001095056 | 0,009160186 | 0,119545135 | 0,904843484 | 0,997431792 | YES |
| 203 | 28,32546676 | 0,001097134 | 0,00920576 | 0,119179048 | 0,905133505 | 0,997431792 | YES |
| 204 | 15,00686138 | 0,001089629 | 0,009254768 | 0,117737056 | 0,906276004 | 0,997431792 | YES |
| 205 | 5,305152038 | -0,00107782 | 0,009238849 | -0,116661668 | 0,907128167 | 0,997431792 | YES |
| 206 | 0,516255622 | -0,00106984 | 0,009256152 | -0,115581542 | 0,907984192 | 0,997431792 | YES |
| 207 | 577,6307374 | -0,000560184 | 0,00486512 | -0,115142877 | 0,908331875 | 0,997431792 | YES |
| 208 | 15,26354989 | 0,000853141 | 0,007505028 | 0,113675938 | 0,909494688 | 0,997431792 | YES |
| 209 | 82,21928101 | -0,000854424 | 0,007586787 | -0,112620043 | 0,910331794 | 0,997431792 | YES |

| Medication | Ticker | ASVnr | taxLevel | Phylum | Family | Genus | Species |
|---|---|---|---|---|---|---|---|
| DA LEDD | 210 | ASV307 | ASV | Firmicutes | Lachnospiraceae | Lachnoclostridium | NA |
| DA LEDD | 211 | ASV343 | ASV | Firmicutes | Lachnospiraceae | NA | NA |
| DA LEDD | 212 | ASV517 | ASV | Firmicutes | Lachnospiraceae | Blautia | NA |
| DA LEDD | 213 | ASV438 | ASV | Firmicutes | UCG-010 | NA | NA |
| DA LEDD | 214 | ASV73 | ASV | Firmicutes | Oscillospiraceae | UCG-005 | NA |
| DA LEDD | 215 | ASV267 | ASV | Firmicutes | Lachnospiraceae | Lachnoclostridium | NA |
| DA LEDD | 216 | ASV446 | ASV | Firmicutes | Oscillospiraceae | UCG-005 | NA |
| DA LEDD | 217 | ASV444 | ASV | Firmicutes | Erysipelatoclostridiaceae | Coprobacillus | cateniformis |
| DA LEDD | 218 | ASV534 | ASV | Firmicutes | Anaerovoracaceae | Family XIII UCG-001 | NA |
| DA LEDD | 219 | ASV50 | ASV | Bacteroidota | Prevotellaceae | Prevotella | NA |
| DA LEDD | 220 | ASV709 | ASV | Firmicutes | Lachnospiraceae | NA | NA |
| DA LEDD | 221 | ASV535 | ASV | Firmicutes | Erysipelatoclostridiaceae | Asteroleplasma | NA |
| DA LEDD | 222 | ASV233 | ASV | Firmicutes | [Eubacterium] coprostanoligenes group | NA | NA |
| DA LEDD | 223 | ASV587 | ASV | Firmicutes | Anaerovoracaceae | Family XIII AD3011 group | NA |
| DA LEDD | 224 | ASV160 | ASV | Firmicutes | Ruminococcaceae | Ruminococcus | callidus |
| DA LEDD | 225 | ASV167 | ASV | Firmicutes | Oscillospiraceae | Intestinimonas | NA |
| DA LEDD | 226 | ASV695 | ASV | Firmicutes | UCG-010 | NA | NA |
| DA LEDD | 227 | ASV530 | ASV | Firmicutes | Veillonellaceae | Allisonella | histaminiformans |
| DA LEDD | 228 | ASV545 | ASV | Proteobacteria | Sutterellaceae | Sutterella | NA |
| DA LEDD | 229 | ASV79 | ASV | Firmicutes | Lachnospiraceae | [Eubacterium] ruminantium group | NA |
| DA LEDD | 230 | ASV240 | ASV | Bacteroidota | Prevotellaceae | Alloprevotella | NA |
| DA LEDD | 231 | ASV116 | ASV | Firmicutes | Oscillospiraceae | UCG-003 | NA |
| DA LEDD | 232 | ASV376 | ASV | Bacteroidota | Barnesiellaceae | Coprobacter | fastidiosus |
| DA LEDD | 233 | ASV573 | ASV | Firmicutes | Erysipelatoclostridiaceae | Erysipelatoclostridium | NA |
| DA LEDD | 234 | ASV195 | ASV | Bacteroidota | Barnesiellaceae | Barnesiella | intestinihominis |
| DA LEDD | 235 | ASV703 | ASV | Firmicutes | Hydrogenoanaerobacterium | NA | NA |
| DA LEDD | 236 | ASV194 | ASV | Bacteroidota | Bacteroidaceae | Bacteroides | intestinalis |
| DA LEDD | 237 | ASV104 | ASV | Firmicutes | Christensenellaceae | Christensenellaceae R-7 group | NA |
| DA LEDD | 238 | ASV353 | ASV | Firmicutes | Lachnospiraceae | [Eubacterium] xylanophilum group | NA |
| DA LEDD | 239 | ASV71 | ASV | Firmicutes | [Eubacterium] coprostanoligenes group | NA | NA |

| Ticker | DESeq2_baseMean | DESeq2_log2-FoldChange | DESeq2_lfcSE | DESeq2_stat | DESeq2_pvalue | DESeq2_padj | DESeq2 converged |
|---|---|---|---|---|---|---|---|
| 210 | 10,58221923 | 0,000996073 | 0,009129473 | 0,109105222 | 0,913119033 | 0,997431792 | YES |
| 211 | 11,21380822 | -0,000992706 | 0,009175644 | -0,108189238 | 0,913845582 | 0,997431792 | YES |
| 212 | 1,886407998 | 0,000968858 | 0,009225976 | 0,105014136 | 0,916364592 | 0,997431792 | YES |
| 213 | 21,27090817 | 0,000953234 | 0,009220265 | 0,103384659 | 0,917657687 | 0,997431792 | YES |
| 214 | 131,7532683 | 0,000906801 | 0,009034626 | 0,10036952 | 0,920050967 | 0,997431792 | YES |
| 215 | 5,935439758 | -0,000895626 | 0,009211008 | -0,09723427 | 0,922540353 | 0,997431792 | YES |
| 216 | 3,470899527 | 0,000899758 | 0,009277221 | 0,096985735 | 0,922737723 | 0,997431792 | YES |
| 217 | 2,123983293 | 0,000898947 | 0,009277395 | 0,096896442 | 0,922808634 | 0,997431792 | YES |
| 218 | 2,020374352 | 0,000898838 | 0,009277418 | 0,096884475 | 0,922818138 | 0,997431792 | YES |
| 219 | 0,777067058 | 0,000895355 | 0,009278168 | 0,096501228 | 0,923122498 | 0,997431792 | YES |
| 220 | 0,777067058 | 0,000895355 | 0,009278168 | 0,096501228 | 0,923122498 | 0,997431792 | YES |
| 221 | 0,751458363 | 0,000868594 | 0,009220981 | 0,094197621 | 0,924952175 | 0,997431792 | YES |
| 222 | 1,988620903 | -0,000868685 | 0,009225931 | -0,094156915 | 0,92498451 | 0,997431792 | YES |
| 223 | 0,520829284 | -0,000858074 | 0,009230786 | -0,092957836 | 0,925937059 | 0,997431792 | YES |
| 224 | 28,8143168 | 0,000851881 | 0,009240287 | 0,092191998 | 0,926545495 | 0,997431792 | YES |
| 225 | 12,56394146 | -0,000837474 | 0,009161902 | -0,091408364 | 0,927168116 | 0,997431792 | YES |
| 226 | 1,529963724 | 0,000843512 | 0,009246422 | 0,091225814 | 0,927313164 | 0,997431792 | YES |
| 227 | 7,070697629 | -0,000831918 | 0,00920721 | -0,090355066 | 0,928005063 | 0,997431792 | YES |
| 228 | 53,05744211 | 0,000825534 | 0,009272728 | 0,089028192 | 0,929059505 | 0,997431792 | YES |
| 229 | 13,80818368 | 0,000814074 | 0,009277014 | 0,0877517 | 0,930074027 | 0,997431792 | YES |
| 230 | 118,5908202 | -0,000798151 | 0,009096516 | -0,08774248 | 0,930081356 | 0,997431792 | YES |
| 231 | 127,1856264 | -0,000607119 | 0,006936732 | -0,087522343 | 0,930256326 | 0,997431792 | YES |
| 232 | 4,848200515 | 0,000809813 | 0,009274256 | 0,087318403 | 0,930418426 | 0,997431792 | YES |
| 233 | 35,23305081 | 0,000638157 | 0,007431268 | 0,085874577 | 0,931566122 | 0,997431792 | YES |
| 234 | 137,8979738 | 0,000793739 | 0,009243463 | 0,085870293 | 0,931569527 | 0,997431792 | YES |
| 235 | 3,395585132 | -0,000789178 | 0,009256078 | -0,085260508 | 0,932054287 | 0,997431792 | YES |
| 236 | 14,23534299 | 0,000783876 | 0,009224421 | 0,084978296 | 0,932278646 | 0,997431792 | YES |
| 237 | 42,31758336 | -0,000783228 | 0,009225583 | -0,084897379 | 0,932342975 | 0,997431792 | YES |
| 238 | 22,80175238 | 0,000779722 | 0,009224333 | 0,08452881 | 0,932635998 | 0,997431792 | YES |
| 239 | 48,60197185 | -0,000771302 | 0,009143157 | -0,084358361 | 0,932771512 | 0,997431792 | YES |

| Medication | Ticker | ASVnr | taxLevel | Phylum | Family | Genus | Species |
|---|---|---|---|---|---|---|---|
| DA LEDD | 240 | ASV425 | ASV | Firmicutes | Oscillospiraceae | NK4A214 group | NA |
| DA LEDD | 241 | ASV80 | ASV | Firmicutes | Acidaminococcaceae | Phascolarctobacterium | faecium |
| DA LEDD | 242 | ASV260 | ASV | Firmicutes | Lactobacillaceae | Lactobacillus | delbrueckii |
| DA LEDD | 243 | ASV885 | ASV | Firmicutes | Lachnospiraceae | Shuttleworthia | NA |
| DA LEDD | 244 | ASV322 | ASV | Firmicutes | Anaerovoracaceae | Family XIII AD3011 group | NA |
| DA LEDD | 245 | ASV153 | ASV | Firmicutes | Streptococcaceae | Streptococcus | NA |
| DA LEDD | 246 | ASV403 | ASV | Firmicutes | Oscillospiraceae | Colidextribacter | NA |
| DA LEDD | 247 | ASV72 | ASV | Proteobacteria | Sutterellaceae | Parasutterella | excrementihominis |
| DA LEDD | 248 | ASV155 | ASV | Firmicutes | Ruminococcaceae | Ruminococcus | NA |
| DA LEDD | 249 | ASV303 | ASV | Firmicutes | Lachnospiraceae | GCA-900066575 | NA |
| DA LEDD | 250 | ASV538 | ASV | Bacteroidota | Marinifilaceae | Butyricimonas | NA |
| DA LEDD | 251 | ASV306 | ASV | Firmicutes | Veillonellaceae | Veillonella | NA |
| DA LEDD | 252 | ASV294 | ASV | Firmicutes | Anaerovoracaceae | Mogibacterium | NA |
| DA LEDD | 253 | ASV256 | ASV | Firmicutes | Lachnospiraceae | Blautia | NA |
| DA LEDD | 254 | ASV714 | ASV | Firmicutes | Oscillospiraceae | Oscillibacter | NA |
| DA LEDD | 255 | ASV606 | ASV | Firmicutes | Lachnospiraceae | NA | NA |
| DA LEDD | 256 | ASV752 | ASV | Verrucomicrobiota | Puniceicoccaceae | NA | NA |
| DA LEDD | 257 | ASV726 | ASV | Verrucomicrobiota | vadinBE97 | NA | NA |
| DA LEDD | 258 | ASV320 | ASV | Firmicutes | Christensenellaceae | Christensenellaceae R-7 group | NA |
| DA LEDD | 259 | ASV422 | ASV | Firmicutes | Lachnospiraceae | Lachnospiraceae FCS020 group | bacterium |
| DA LEDD | 260 | ASV232 | ASV | Firmicutes | Acholeplasmataceae | Anaeroplasma | NA |
| DA LEDD | 261 | ASV261 | ASV | Firmicutes | Anaerovoracaceae | Family XIII UCG-001 | NA |
| DA LEDD | 262 | ASV168 | ASV | Firmicutes | Lachnospiraceae | Lachnoclostridium | edouardi |
| DA LEDD | 263 | ASV432 | ASV | Bacteroidota | Marinifilaceae | Butyricimonas | virosa |
| DA LEDD | 264 | ASV894 | ASV | Desulfobacterota | Desulfovibrionaceae | NA | NA |
| DA LEDD | 265 | ASV347 | ASV | Bacteroidota | Tannerellaceae | Parabacteroides | NA |
| DA LEDD | 266 | ASV158 | ASV | Firmicutes | Lachnospiraceae | [Eubacterium] ventriosum group | NA |
| DA LEDD | 267 | ASV30 | ASV | Proteobacteria | Enterobacteriaceae | Escherichia-Shigella | NA |
| DA LEDD | 268 | ASV362 | ASV | Firmicutes | Ruminococcaceae | NA | NA |
| DA LEDD | 269 | ASV209 | ASV | Bacteroidota | Tannerellaceae | NA | NA |

| Ticker | DESeq2_baseMean | DESeq2_log2-FoldChange | DESeq2_lfcSE | DESeq2_stat | DESeq2_pvalue | DESeq2_padj | DESeq2 converged |
|---|---|---|---|---|---|---|---|
| 240 | 20,81801385 | -0,000762954 | 0,009174736 | -0,08315817 | 0,933725773 | 0,997431792 | YES |
| 241 | 4,627415686 | -0,000765324 | 0,009219762 | -0,083009058 | 0,933844337 | 0,997431792 | YES |
| 242 | 48,50443725 | 0,000753009 | 0,009220116 | 0,08167018 | 0,934908992 | 0,997431792 | YES |
| 243 | 9,595750395 | -0,000642093 | 0,008169245 | -0,078598772 | 0,937351765 | 0,997431792 | YES |
| 244 | 12,38549144 | 0,000709783 | 0,009168271 | 0,077417316 | 0,938291566 | 0,997431792 | YES |
| 245 | 50,75517682 | -0,000699573 | 0,009221794 | -0,075860865 | 0,939529792 | 0,997431792 | YES |
| 246 | 6,75389286 | -0,000683739 | 0,009087657 | -0,075238231 | 0,940025167 | 0,997431792 | YES |
| 247 | 101,9339438 | -0,000603611 | 0,008025788 | -0,075208991 | 0,940048431 | 0,997431792 | YES |
| 248 | 27,20871536 | -0,000682191 | 0,009145554 | -0,074592693 | 0,940538788 | 0,997431792 | YES |
| 249 | 4,522088092 | -0,000678886 | 0,00916781 | -0,074051061 | 0,940969756 | 0,997431792 | YES |
| 250 | 10,16390106 | -0,000678066 | 0,009238499 | -0,073395661 | 0,94149127 | 0,997431792 | YES |
| 251 | 56,75108944 | 0,000641638 | 0,009171938 | 0,069956614 | 0,944228192 | 0,997431792 | YES |
| 252 | 6,908737034 | 0,000642563 | 0,00927708 | 0,069263508 | 0,944779873 | 0,997431792 | YES |
| 253 | 2,208170287 | 0,000637564 | 0,009287794 | 0,068645415 | 0,945271868 | 0,997431792 | YES |
| 254 | 1,958188745 | 0,000637441 | 0,009287808 | 0,068632027 | 0,945282525 | 0,997431792 | YES |
| 255 | 1,791534384 | 0,000637339 | 0,009287819 | 0,068620948 | 0,945291344 | 0,997431792 | YES |
| 256 | 1,666543613 | 0,000637247 | 0,009287829 | 0,068610973 | 0,945299284 | 0,997431792 | YES |
| 257 | 0,291645132 | 0,000631262 | 0,009288489 | 0,067961715 | 0,94581611 | 0,997431792 | YES |
| 258 | 2,842283495 | -0,00061204 | 0,009177932 | -0,066686045 | 0,946831644 | 0,997431792 | YES |
| 259 | 4,417389267 | -0,000607804 | 0,009161367 | -0,066344291 | 0,947103722 | 0,997431792 | YES |
| 260 | 58,26968806 | 0,000606953 | 0,009276961 | 0,065425867 | 0,947834929 | 0,997431792 | YES |
| 261 | 18,70344525 | -0,000474323 | 0,007391129 | -0,064174643 | 0,948831168 | 0,997431792 | YES |
| 262 | 22,34143269 | -0,000511464 | 0,008082319 | -0,063281884 | 0,949542042 | 0,997431792 | YES |
| 263 | 13,37726803 | 0,000453812 | 0,007233867 | 0,062734356 | 0,949978039 | 0,997431792 | YES |
| 264 | 1,932859215 | 0,000565685 | 0,009287809 | 0,06090623 | 0,951433887 | 0,997431792 | YES |
| 265 | 2,322330941 | 0,000562944 | 0,009287788 | 0,060611243 | 0,951668819 | 0,997431792 | YES |
| 266 | 83,97160382 | 0,000410678 | 0,006974923 | 0,058879204 | 0,953048322 | 0,997431792 | YES |
| 267 | 133,001607 | 0,000385601 | 0,006625485 | 0,05819969 | 0,953589568 | 0,997431792 | YES |
| 268 | 27,99546171 | -0,000313601 | 0,005395538 | -0,058122239 | 0,95365126 | 0,997431792 | YES |
| 269 | 2,091095852 | 0,000532685 | 0,009225725 | 0,05773913 | 0,953956425 | 0,997431792 | YES |

| Medication | Ticker | ASVnr | taxLevel | Phylum | Family | Genus | Species |
|---|---|---|---|---|---|---|---|
| DA LEDD | 270 | ASV305 | ASV | Firmicutes | Lachnospiraceae | Lachnospiraceae UCG-004 | NA |
| DA LEDD | 271 | ASV2 | ASV | Bacteroidota | Bacteroidaceae | Bacteroides | vulgatus |
| DA LEDD | 272 | ASV40 | ASV | Firmicutes | Erysipelotrichaceae | Holdemanella | NA |
| DA LEDD | 273 | ASV1464 | ASV | Firmicutes | Peptococcaceae | NA | NA |
| DA LEDD | 274 | ASV176 | ASV | Bacteroidota | Prevotellaceae | Paraprevotella | NA |
| DA LEDD | 275 | ASV310 | ASV | Firmicutes | Lachnospiraceae | NA | NA |
| DA LEDD | 276 | ASV51 | ASV | Firmicutes | Oscillospiraceae | UCG-005 | NA |
| DA LEDD | 277 | ASV453 | ASV | Verrucomicrobiota | Victivallaceae | Victivallis | NA |
| DA LEDD | 278 | ASV272 | ASV | Firmicutes | Erysipelotrichaceae | Catenisphaera | NA |
| DA LEDD | 279 | ASV387 | ASV | Firmicutes | Lachnospiraceae | NA | NA |
| DA LEDD | 280 | ASV20 | ASV | Bacteroidota | Prevotellaceae | Prevotella | copri |
| DA LEDD | 281 | ASV146 | ASV | Firmicutes | Erysipelotrichaceae | Turicibacter | sanguinis |
| DA LEDD | 282 | ASV507 | ASV | Firmicutes | Oscillospiraceae | Colidextribacter | NA |
| DA LEDD | 283 | ASV454 | ASV | Actinobacteriota | Actinomycetaceae | Actinomyces | odontolyticus |
| DA LEDD | 284 | ASV152 | ASV | Firmicutes | Lachnospiraceae | Roseburia | hominis |
| DA LEDD | 285 | ASV111 | ASV | Firmicutes | Lachnospiraceae | Lachnospira | pectinoschiza |
| DA LEDD | 286 | ASV208 | ASV | Bacteroidota | Prevotellaceae | Prevotella | NA |
| DA LEDD | 287 | ASV608 | ASV | Firmicutes | Lachnospiraceae | NA | NA |
| DA LEDD | 288 | ASV32 | ASV | Firmicutes | Oscillospiraceae | UCG-002 | NA |
| DA LEDD | 289 | ASV162 | ASV | Desulfobacterota | Desulfovibrionaceae | Desulfovibrio | NA |
| DA LEDD | 290 | ASV276 | ASV | Firmicutes | Oscillospiraceae | NK4A214 group | NA |
| DA LEDD | 291 | ASV222 | ASV | Firmicutes | Oscillospiraceae | NK4A214 group | NA |
| DA LEDD | 292 | ASV106 | ASV | Bacteroidota | Bacteroidaceae | Bacteroides | NA |
| DA LEDD | 293 | ASV36 | ASV | Firmicutes | Lachnospiraceae | Blautia | obeum |
| DA LEDD | 294 | ASV58 | ASV | Bacteroidota | Bacteroidaceae | Bacteroides | NA |
| DA LEDD | 295 | ASV250 | ASV | Bacteroidota | Prevotellaceae | Paraprevotella | NA |
| DA LEDD | 296 | ASV88 | ASV | Firmicutes | Lachnospiraceae | Dorea | formicigenerans |
| DA LEDD | 297 | ASV217 | ASV | Firmicutes | Peptococcaceae | Peptococcus | NA |
| DA LEDD | 298 | ASV38 | ASV | Firmicutes | Clostridiaceae | Clostridium sensu stricto 1 | NA |
| DA LEDD | 299 | ASV309 | ASV | Firmicutes | Oscillospiraceae | Colidextribacter | massiliensis |

| Ticker | DESeq2_baseMean | DESeq2_log2-FoldChange | DESeq2_lfcSE | DESeq2_stat | DESeq2_pvalue | DESeq2_padj | DESeq2 converged |
|---|---|---|---|---|---|---|---|
| 270 | 15,76402688 | 0,000508257 | 0,009082898 | 0,055957624 | 0,955375566 | 0,997431792 | YES |
| 271 | 1053,515881 | 0,000388524 | 0,006945159 | 0,055941664 | 0,95538828 | 0,997431792 | YES |
| 272 | 120,8395767 | -0,000508636 | 0,009239381 | -0,055050823 | 0,956097974 | 0,997431792 | YES |
| 273 | 1,545222137 | 0,000503442 | 0,009287839 | 0,054204461 | 0,956772266 | 0,997431792 | YES |
| 274 | 15,47677946 | -0,000481822 | 0,009224401 | -0,05223344 | 0,958342688 | 0,997431792 | YES |
| 275 | 27,29458452 | -0,000478845 | 0,009209772 | -0,051993184 | 0,958534124 | 0,997431792 | YES |
| 276 | 112,3228646 | 0,000351742 | 0,006863218 | 0,051250311 | 0,959126062 | 0,997431792 | YES |
| 277 | 0,439374009 | -0,000465324 | 0,009238027 | -0,050370544 | 0,959827109 | 0,997431792 | YES |
| 278 | 6,715675586 | 0,000447698 | 0,009160673 | 0,048871748 | 0,961021504 | 0,997431792 | YES |
| 279 | 4,360669351 | -0,000438714 | 0,009184874 | -0,047764812 | 0,96190368 | 0,997431792 | YES |
| 280 | 781,5005243 | 0,000432267 | 0,009146865 | 0,047258518 | 0,962307189 | 0,997431792 | YES |
| 281 | 5,227121934 | -0,000435723 | 0,009235214 | -0,047180575 | 0,962369309 | 0,997431792 | YES |
| 282 | 42,29824705 | 0,000431993 | 0,009258329 | 0,046659971 | 0,962784234 | 0,997431792 | YES |
| 283 | 1,467775284 | 0,000431908 | 0,009258123 | 0,046651782 | 0,962790761 | 0,997431792 | YES |
| 284 | 87,97309925 | 0,000418951 | 0,009032889 | 0,046380606 | 0,963006894 | 0,997431792 | YES |
| 285 | 57,02858643 | -0,000287187 | 0,006260162 | -0,045875278 | 0,963409659 | 0,997431792 | YES |
| 286 | 18,95485928 | 0,000422224 | 0,009224358 | 0,045772702 | 0,963491416 | 0,997431792 | YES |
| 287 | 7,442933781 | -0,000417989 | 0,009160806 | -0,045628026 | 0,963606731 | 0,997431792 | YES |
| 288 | 425,2658718 | -0,000266665 | 0,00585034 | -0,045581044 | 0,963644178 | 0,997431792 | YES |
| 289 | 33,66228937 | 0,000409954 | 0,009287693 | 0,044139485 | 0,964793219 | 0,997431792 | YES |
| 290 | 141,6776611 | 0,00040532 | 0,009254076 | 0,0437991 | 0,965064544 | 0,997431792 | YES |
| 291 | 41,9580284 | 0,000403126 | 0,009205637 | 0,043791251 | 0,965070801 | 0,997431792 | YES |
| 292 | 50,52826394 | -0,000395677 | 0,009113499 | -0,04341657 | 0,965369469 | 0,997431792 | YES |
| 293 | 134,6497628 | -0,000249517 | 0,005993563 | -0,041630793 | 0,966793025 | 0,997431792 | YES |
| 294 | 151,3403235 | -0,000329683 | 0,007980336 | -0,041311958 | 0,9670472 | 0,997431792 | YES |
| 295 | 42,08744841 | -0,000329724 | 0,008060538 | -0,040905998 | 0,967370836 | 0,997431792 | YES |
| 296 | 89,42595153 | 0,000282237 | 0,006928978 | 0,040732804 | 0,967508909 | 0,997431792 | YES |
| 297 | 58,56256001 | 0,00037517 | 0,00928769 | 0,040394366 | 0,967778722 | 0,997431792 | YES |
| 298 | 7,461734711 | -0,000361567 | 0,009128948 | -0,039606613 | 0,968406755 | 0,997431792 | YES |
| 299 | 15,72089964 | -0,000347845 | 0,008967822 | -0,038788086 | 0,969059344 | 0,997431792 | YES |

| Medication | Ticker | ASVnr | taxLevel | Phylum | Family | Genus | Species |
|---|---|---|---|---|---|---|---|
| DA LEDD | 300 | ASV54 | ASV | Bacteroidota | Bacteroidaceae | Bacteroides | NA |
| DA LEDD | 301 | ASV140 | ASV | Bacteroidota | Tannerellaceae | Parabacteroides | distasonis |
| DA LEDD | 302 | ASV164 | ASV | Bacteroidota | Tannerellaceae | Parabacteroides | distasonis |
| DA LEDD | 303 | ASV223 | ASV | Bacteroidota | Barnesiellaceae | Barnesiella | NA |
| DA LEDD | 304 | ASV37 | ASV | Firmicutes | Lachnospiraceae | [Eubacterium] hallii group | NA |
| DA LEDD | 305 | ASV393 | ASV | Bacteroidota | Rikenellaceae | Alistipes | indistinctus |
| DA LEDD | 306 | ASV159 | ASV | Firmicutes | Lachnospiraceae | Roseburia | NA |
| DA LEDD | 307 | ASV819 | ASV | Firmicutes | Oscillospiraceae | Colidextribacter | NA |
| DA LEDD | 308 | ASV129 | ASV | Firmicutes | Christensenellaceae | Christensenellaceae R-7 group | NA |
| DA LEDD | 309 | ASV11 | ASV | Firmicutes | Ruminococcaceae | Subdoligranulum | NA |
| DA LEDD | 310 | ASV186 | ASV | Firmicutes | Lachnospiraceae | [Ruminococcus] torques group | NA |
| DA LEDD | 311 | ASV31 | ASV | Bacteroidota | Rikenellaceae | Alistipes | putredinis |
| DA LEDD | 312 | ASV61 | ASV | Firmicutes | Erysipelatoclostridiaceae | Catenibacterium | mitsuokai |
| DA LEDD | 313 | ASV175 | ASV | Firmicutes | NA | NA | NA |
| DA LEDD | 314 | ASV148 | ASV | Firmicutes | NA | NA | NA |
| DA LEDD | 315 | ASV373 | ASV | Firmicutes | NA | NA | NA |
| DA LEDD | 316 | ASV114 | ASV | Bacteroidota | Bacteroidaceae | Bacteroides | coprocola |
| DA LEDD | 317 | ASV179 | ASV | Firmicutes | NA | NA | NA |
| DA LEDD | 318 | ASV180 | ASV | Firmicutes | Ruminococcaceae | Ruminococcus | NA |
| DA LEDD | 319 | ASV252 | ASV | Firmicutes | Erysipelotrichaceae | Solobacterium | NA |
| DA LEDD | 320 | ASV205 | ASV | Firmicutes | NA | NA | NA |
| DA LEDD | 321 | ASV759 | ASV | Firmicutes | NA | NA | NA |
| DA LEDD | 322 | ASV546 | ASV | Firmicutes | Lachnospiraceae | NA | NA |
| DA LEDD | 323 | ASV437 | ASV | Firmicutes | Christensenellaceae | Christensenellaceae R-7 group | NA |
| DA LEDD | 324 | ASV597 | ASV | Firmicutes | Oscillospiraceae | Oscillibacter | NA |
| DA LEDD | 325 | ASV753 | ASV | Firmicutes | Ruminococcaceae | NA | NA |
| DA LEDD | 326 | ASV480 | ASV | Firmicutes | UCG-010 | NA | NA |
| DA LEDD | 327 | ASV370 | ASV | Firmicutes | UCG-010 | NA | NA |
| DA LEDD | 328 | ASV254 | ASV | Firmicutes | Lachnospiraceae | Lachnospira | NA |
| DA LEDD | 329 | ASV414 | ASV | Firmicutes | Oscillospiraceae | UCG-003 | NA |

| Ticker | DESeq2_baseMean | DESeq2_log2-FoldChange | DESeq2_lfcSE | DESeq2_stat | DESeq2_pvalue | DESeq2_padj | DESeq2 converged |
|---|---|---|---|---|---|---|---|
| 300 | 286,1460699 | 0,000259919 | 0,006760792 | 0,038445112 | 0,969332794 | 0,997431792 | YES |
| 301 | 36,88112669 | -0,000346135 | 0,009240206 | -0,037459706 | 0,970118468 | 0,997431792 | YES |
| 302 | 6,199235025 | 0,000340713 | 0,009287724 | 0,036684199 | 0,970736807 | 0,997431792 | YES |
| 303 | 13,68705374 | 0,000336273 | 0,009224421 | 0,03645468 | 0,970919815 | 0,997431792 | YES |
| 304 | 61,88242008 | 0,000204646 | 0,005719381 | 0,035781068 | 0,971456929 | 0,997431792 | YES |
| 305 | 15,81276091 | 0,000258167 | 0,007271131 | 0,035505735 | 0,971676473 | 0,997431792 | YES |
| 306 | 73,58395473 | -0,000254184 | 0,007160852 | -0,035496361 | 0,971683948 | 0,997431792 | YES |
| 307 | 7,484426045 | 0,000321212 | 0,009240417 | 0,034761664 | 0,97226979 | 0,997431792 | YES |
| 308 | 29,06788126 | -0,000322185 | 0,009273136 | -0,034743916 | 0,972283942 | 0,997431792 | YES |
| 309 | 312,1109785 | -0,000142816 | 0,004241328 | -0,033672518 | 0,973138294 | 0,997431792 | YES |
| 310 | 63,31194315 | 0,000308694 | 0,00925831 | 0,033342396 | 0,973401545 | 0,997431792 | YES |
| 311 | 438,7820828 | -0,000168359 | 0,005280725 | -0,031881882 | 0,974566247 | 0,997431792 | YES |
| 312 | 75,00824213 | -0,000270909 | 0,009272948 | -0,029214972 | 0,97669314 | 0,997431792 | YES |
| 313 | 3,986068578 | 0,000252141 | 0,009237925 | 0,027294115 | 0,978225151 | 0,997431792 | YES |
| 314 | 11,98018217 | -0,00025074 | 0,009277022 | -0,027028021 | 0,978437384 | 0,997431792 | YES |
| 315 | 8,193992525 | -0,000250586 | 0,009277058 | -0,027011321 | 0,978450704 | 0,997431792 | YES |
| 316 | 7,967951352 | -0,000250569 | 0,009277061 | -0,027009572 | 0,9784521 | 0,997431792 | YES |
| 317 | 7,515869006 | -0,000250541 | 0,009277068 | -0,027006503 | 0,978454547 | 0,997431792 | YES |
| 318 | 4,520823462 | -0,000250196 | 0,009277149 | -0,026969052 | 0,978484418 | 0,997431792 | YES |
| 319 | 4,407802875 | -0,000250174 | 0,009277155 | -0,026966681 | 0,978486309 | 0,997431792 | YES |
| 320 | 3,27759701 | -0,000249869 | 0,009277227 | -0,026933598 | 0,978512696 | 0,997431792 | YES |
| 321 | 2,995045544 | -0,000249759 | 0,009277253 | -0,026921635 | 0,978522238 | 0,997431792 | YES |
| 322 | 2,599473491 | -0,000249562 | 0,0092773 | -0,026900251 | 0,978539293 | 0,997431792 | YES |
| 323 | 2,316922024 | -0,000249382 | 0,009277343 | -0,026880772 | 0,97855483 | 0,997431792 | YES |
| 324 | 2,316922024 | -0,000249382 | 0,009277343 | -0,026880772 | 0,97855483 | 0,997431792 | YES |
| 325 | 2,316922024 | -0,000249382 | 0,009277343 | -0,026880772 | 0,97855483 | 0,997431792 | YES |
| 326 | 1,921349971 | -0,00024904 | 0,009277425 | -0,026843667 | 0,978584424 | 0,997431792 | YES |
| 327 | 1,469267625 | -0,000248427 | 0,009277572 | -0,026777137 | 0,978637489 | 0,997431792 | YES |
| 328 | 1,299736745 | -0,000248089 | 0,009277653 | -0,026740488 | 0,97866672 | 0,997431792 | YES |
| 329 | 0,621613226 | -0,000244913 | 0,009278417 | -0,026396025 | 0,978941465 | 0,997431792 | YES |

| Medication | Ticker | ASVnr | taxLevel | Phylum | Family | Genus | Species |
|---|---|---|---|---|---|---|---|
| DA LEDD | 330 | ASV241 | ASV | Firmicutes | [Eubacterium] coprostanoligenes group | NA | NA |
| DA LEDD | 331 | ASV150 | ASV | Proteobacteria | Sutterellaceae | Sutterella | NA |
| DA LEDD | 332 | ASV828 | ASV | Firmicutes | Christensenellaceae | Christensenellaceae R-7 group | NA |
| DA LEDD | 333 | ASV937 | ASV | Firmicutes | UCG-010 | NA | NA |
| DA LEDD | 334 | ASV118 | ASV | Firmicutes | NA | NA | NA |
| DA LEDD | 335 | ASV286 | ASV | Firmicutes | Lachnospiraceae | NA | NA |
| DA LEDD | 336 | ASV277 | ASV | Bacteroidota | Rikenellaceae | Alistipes | ihumii |
| DA LEDD | 337 | ASV727 | ASV | Firmicutes | Ruminococcaceae | Angelakisella | NA |
| DA LEDD | 338 | ASV231 | ASV | Proteobacteria | NA | NA | NA |
| DA LEDD | 339 | ASV433 | ASV | Firmicutes | UCG-010 | NA | NA |
| DA LEDD | 340 | ASV174 | ASV | Actinobacteriota | Coriobacteriales Incertae Sedis | NA | NA |
| DA LEDD | 341 | ASV136 | ASV | Firmicutes | Ruminococcaceae | NA | NA |
| DA LEDD | 342 | ASV332 | ASV | Firmicutes | Lachnospiraceae | [Eubacterium] ventriosum group | NA |
| DA LEDD | 343 | ASV98 | ASV | Firmicutes | Acidaminococcaceae | Phascolarctobacterium | succinatutens |
| DA LEDD | 344 | ASV157 | ASV | Firmicutes | Oscillospiraceae | UCG-005 | NA |
| DA LEDD | 345 | ASV109 | ASV | Firmicutes | [Eubacterium] coprostanoligenes group | NA | NA |
| DA LEDD | 346 | ASV251 | ASV | Firmicutes | Oscillospiraceae | NK4A214 group | NA |
| DA LEDD | 347 | ASV151 | ASV | Firmicutes | Butyricicoccaceae | Butyricicoccus | NA |
| DA LEDD | 348 | ASV308 | ASV | Firmicutes | Christensenellaceae | NA | NA |
| DA LEDD | 349 | ASV69 | ASV | Firmicutes | Christensenellaceae | NA | NA |
| DA LEDD | 350 | ASV326 | ASV | Proteobacteria | NA | NA | NA |
| DA LEDD | 351 | ASV237 | ASV | Bacteroidota | Bacteroidaceae | Bacteroides | clarus |
| DA LEDD | 352 | ASV287 | ASV | Firmicutes | Christensenellaceae | Christensenellaceae R-7 group | NA |
| DA LEDD | 353 | ASV436 | ASV | Firmicutes | Christensenellaceae | Christensenellaceae R-7 group | NA |
| DA LEDD | 354 | ASV299 | ASV | Firmicutes | Streptococcaceae | Lactococcus | NA |
| DA LEDD | 355 | ASV9 | ASV | Bacteroidota | Bacteroidaceae | Bacteroides | NA |
| DA LEDD | 356 | ASV338 | ASV | Firmicutes | Lachnospiraceae | Lachnotalea | NA |
| DA LEDD | 357 | ASV264 | ASV | Firmicutes | Oscillospiraceae | Colidextribacter | NA |
| DA LEDD | 358 | ASV536 | ASV | Bacteroidota | Marinifilaceae | Butyricimonas | NA |
| DA LEDD | 359 | ASV494 | ASV | Firmicutes | Anaerovoracaceae | Family XIII AD3011 group | NA |

| Ticker | DESeq2_baseMean | DESeq2_log2-FoldChange | DESeq2_lfcSE | DESeq2_stat | DESeq2_pvalue | DESeq2_padj | DESeq2 converged |
|---|---|---|---|---|---|---|---|
| 330 | 0,452082346 | -0,000242658 | 0,00927896 | -0,026151426 | 0,979136559 | 0,997431792 | YES |
| 331 | 8,128584484 | -0,000240584 | 0,009223941 | -0,02608254 | 0,979191504 | 0,997431792 | YES |
| 332 | 0,33906176 | -0,000239926 | 0,009279619 | -0,025855168 | 0,979372859 | 0,997431792 | YES |
| 333 | 0,282551466 | -0,000237762 | 0,009280141 | -0,025620491 | 0,979560042 | 0,997431792 | YES |
| 334 | 63,10985334 | -0,000200476 | 0,009277342 | -0,021609232 | 0,982759669 | 0,997431792 | YES |
| 335 | 5,191965365 | 0,000198552 | 0,009211057 | 0,021555879 | 0,982802229 | 0,997431792 | YES |
| 336 | 3,374096179 | 0,000173817 | 0,009255596 | 0,018779629 | 0,985016905 | 0,997431792 | YES |
| 337 | 6,234247042 | -0,000169727 | 0,009258661 | -0,018331726 | 0,985374218 | 0,997431792 | YES |
| 338 | 4,980552695 | -0,000155186 | 0,009254873 | -0,016768008 | 0,986621692 | 0,997431792 | YES |
| 339 | 8,287123424 | -0,00015298 | 0,009210803 | -0,016608807 | 0,986748698 | 0,997431792 | YES |
| 340 | 7,097758825 | 0,000150615 | 0,009160537 | 0,016441726 | 0,986881992 | 0,997431792 | YES |
| 341 | 64,64348299 | 0,000149459 | 0,009157839 | 0,016320318 | 0,986978849 | 0,997431792 | YES |
| 342 | 31,85861736 | -0,000145276 | 0,009095097 | -0,015972977 | 0,98725595 | 0,997431792 | YES |
| 343 | 39,74120698 | -0,000132684 | 0,009272738 | -0,014309083 | 0,988583393 | 0,997431792 | YES |
| 344 | 17,46368906 | -9,98E-05 | 0,008213991 | -0,012144208 | 0,990310562 | 0,997431792 | YES |
| 345 | 56,50189389 | 9,81E-05 | 0,009034402 | 0,010853522 | 0,991340312 | 0,997431792 | YES |
| 346 | 13,88456112 | -9,97E-05 | 0,00922042 | -0,010815159 | 0,99137092 | 0,997431792 | YES |
| 347 | 19,94635241 | -9,72E-05 | 0,00920999 | -0,010552916 | 0,991580147 | 0,997431792 | YES |
| 348 | 25,82786753 | 9,68E-05 | 0,009258365 | 0,010450044 | 0,991662223 | 0,997431792 | YES |
| 349 | 105,3457468 | -8,18E-05 | 0,009273006 | -0,008823906 | 0,992959633 | 0,997431792 | YES |
| 350 | 1,43990671 | -7,49E-05 | 0,009165477 | -0,008176046 | 0,993476532 | 0,997431792 | YES |
| 351 | 110,0502031 | -7,30E-05 | 0,009207012 | -0,007932119 | 0,993671151 | 0,997431792 | YES |
| 352 | 7,240776119 | -7,23E-05 | 0,009272995 | -0,007791737 | 0,993783156 | 0,997431792 | YES |
| 353 | 1,154331945 | 7,00E-05 | 0,009194986 | 0,00761176 | 0,993926753 | 0,997431792 | YES |
| 354 | 11,46938317 | 4,98E-05 | 0,009258482 | 0,005373581 | 0,995712523 | 0,997431792 | YES |
| 355 | 1397,971376 | 4,28E-05 | 0,008768302 | 0,004876477 | 0,99610915 | 0,997431792 | YES |
| 356 | 6,454170329 | -4,39E-05 | 0,009224682 | -0,004755307 | 0,996205829 | 0,997431792 | YES |
| 357 | 20,01693115 | 3,92E-05 | 0,009098805 | 0,004304726 | 0,996565336 | 0,997431792 | YES |
| 358 | 1,235586066 | 3,69E-05 | 0,009287875 | 0,003970395 | 0,996832091 | 0,997431792 | YES |
| 359 | 1,404075075 | 3,67E-05 | 0,009287852 | 0,00395374 | 0,99684538 | 0,997431792 | YES |

| Medication | Ticker | ASVnr | taxLevel | Phylum | Family | Genus | Species |
|---|---|---|---|---|---|---|---|
| DA LEDD | 360 | ASV187 | ASV | Firmicutes | NA | NA | NA |
| DA LEDD | 361 | ASV317 | ASV | Firmicutes | NA | NA | NA |
| DA LEDD | 362 | ASV499 | ASV | Firmicutes | Lachnospiraceae | Eisenbergiella | tayi |
| DA LEDD | 363 | ASV487 | ASV | Firmicutes | Lachnospiraceae | Howardella | ureilytica |
| DA LEDD | 364 | ASV335 | ASV | Bacteroidota | Barnesiellaceae | NA | NA |
| DA LEDD | 365 | ASV611 | ASV | Firmicutes | Lachnospiraceae | Lachnospiraceae UCG-010 | NA |
| DA LEDD | 366 | ASV411 | Genus | Firmicutes | Lactobacillaceae | Lactobacillus | NA |
| DA LEDD | 367 | ASV295 | Genus | Firmicutes | Ruminococcaceae | Negativibacillus | NA |
| DA LEDD | 368 | ASV13 | Genus | Firmicutes | Lachnospiraceae | Fusicatenibacter | NA |
| DA LEDD | 369 | ASV96 | Genus | Firmicutes | Lachnospiraceae | CAG-56 | NA |
| DA LEDD | 370 | ASV51 | Genus | Firmicutes | Oscillospiraceae | UCG-005 | NA |
| DA LEDD | 371 | ASV1 | Genus | Actinobacteriota | Bifidobacteriaceae | Bifidobacterium | NA |
| DA LEDD | 372 | ASV322 | Genus | Firmicutes | Anaerovoracaceae | Family XIII AD3011 group | NA |
| DA LEDD | 373 | ASV16 | Genus | Firmicutes | Peptostreptococcaceae | Romboutsia | NA |
| DA LEDD | 374 | ASV2 | Genus | Bacteroidota | Bacteroidaceae | Bacteroides | NA |
| DA LEDD | 375 | ASV192 | Genus | Bacteroidota | Marinifilaceae | Odoribacter | NA |
| DA LEDD | 376 | ASV386 | Genus | Firmicutes | Ruminococcaceae | DTU089 | NA |
| DA LEDD | 377 | ASV141 | Genus | Firmicutes | Lachnospiraceae | Lachnoclostridium | NA |
| DA LEDD | 378 | ASV92 | Genus | Firmicutes | Erysipelotrichaceae | Turicibacter | NA |
| DA LEDD | 379 | ASV65 | Genus | Firmicutes | Ruminococcaceae | Incertae Sedis | NA |
| DA LEDD | 380 | ASV328 | Genus | Firmicutes | Anaerovoracaceae | [Eubacterium] brachy group | NA |
| DA LEDD | 381 | ASV5 | Genus | Firmicutes | Lachnospiraceae | Blautia | NA |
| DA LEDD | 382 | ASV111 | Genus | Firmicutes | Lachnospiraceae | Lachnospira | NA |
| DA LEDD | 383 | ASV53 | Genus | Firmicutes | Lachnospiraceae | [Ruminococcus] torques group | NA |
| DA LEDD | 384 | ASV193 | Genus | Actinobacteriota | Eggerthellaceae | Adlercreutzia | NA |
| DA LEDD | 385 | ASV200 | Genus | Firmicutes | Lachnospiraceae | [Eubacterium] xylanophilum group | NA |
| DA LEDD | 386 | ASV23 | Genus | Firmicutes | Streptococcaceae | Streptococcus | NA |
| DA LEDD | 387 | ASV357 | Genus | Firmicutes | Lachnospiraceae | Marvinbryantia | NA |
| DA LEDD | 388 | ASV37 | Genus | Firmicutes | Lachnospiraceae | [Eubacterium] hallii group | NA |
| DA LEDD | 389 | ASV536 | Genus | Bacteroidota | Marinifilaceae | Butyricimonas | NA |

| Ticker | DESeq2_baseMean | DESeq2_log2-FoldChange | DESeq2_lfcSE | DESeq2_stat | DESeq2_pvalue | DESeq2_padj | DESeq2 converged |
|---|---|---|---|---|---|---|---|
| 360 | 3,088965164 | 3,61E-05 | 0,009287762 | 0,003887062 | 0,996898581 | 0,997431792 | YES |
| 361 | 3,538269188 | 3,60E-05 | 0,009287752 | 0,003880129 | 0,996904113 | 0,997431792 | YES |
| 362 | 3,931410209 | 3,60E-05 | 0,009287745 | 0,003874996 | 0,996908208 | 0,997431792 | YES |
| 363 | 5,909439436 | -3,42E-05 | 0,00921752 | -0,003712813 | 0,997037611 | 0,997431792 | YES |
| 364 | 52,8323071 | 3,38E-05 | 0,009208149 | 0,0036731 | 0,997069297 | 0,997431792 | YES |
| 365 | 19,70269215 | 2,94E-05 | 0,009128237 | 0,003218777 | 0,997431792 | 0,997431792 | YES |
| 366 | 92,84598104 | 0,009863515 | 0,003864749 | 2,552174799 | 0,01070528 | 0,703863289 | YES |
| 367 | 92,12266094 | 0,006338793 | 0,002709426 | 2,339533679 | 0,019307831 | 0,703863289 | YES |
| 368 | 711,149347 | 0,008451691 | 0,003711314 | 2,277277426 | 0,022769664 | 0,703863289 | YES |
| 369 | 178,255087 | -0,013519642 | 0,006131073 | -2,205102199 | 0,027446924 | 0,703863289 | YES |
| 370 | 408,1887532 | 0,007029587 | 0,00322114 | 2,182329004 | 0,02908526 | 0,703863289 | YES |
| 371 | 1054,994047 | 0,00342136 | 0,00172641 | 1,981777182 | 0,047504184 | 0,890375585 | YES |
| 372 | 53,55976405 | 0,003506055 | 0,001800551 | 1,947211935 | 0,051509331 | 0,890375585 | YES |
| 373 | 45,56525839 | 0,007716292 | 0,004205948 | 1,834614388 | 0,066562845 | 0,998311993 | YES |
| 374 | 7247,818069 | -0,001610687 | 0,001049605 | -1,534564301 | 0,124890887 | 0,998311993 | YES |
| 375 | 22,25984237 | -0,00369658 | 0,002428306 | -1,522287681 | 0,127937013 | 0,998311993 | YES |
| 376 | 34,94914423 | 0,003824327 | 0,002536768 | 1,507559175 | 0,131667388 | 0,998311993 | YES |
| 377 | 215,256091 | -0,002125429 | 0,001481486 | -1,434660143 | 0,151383967 | 0,998311993 | YES |
| 378 | 12,0624508 | 0,007142764 | 0,005005418 | 1,427006623 | 0,153577986 | 0,998311993 | YES |
| 379 | 206,4588675 | 0,002243164 | 0,001624306 | 1,380999085 | 0,167279242 | 0,998311993 | YES |
| 380 | 9,443006777 | 0,007499683 | 0,005463916 | 1,372583949 | 0,169881725 | 0,998311993 | YES |
| 381 | 1879,779624 | -0,002169299 | 0,001580508 | -1,372532452 | 0,169897744 | 0,998311993 | YES |
| 382 | 133,9734081 | -0,004112696 | 0,003152459 | -1,304599236 | 0,192029349 | 0,998311993 | YES |
| 383 | 675,4032774 | -0,002226195 | 0,00172889 | -1,28764377 | 0,197869996 | 0,998311993 | YES |
| 384 | 8,913992309 | 0,009789335 | 0,007925109 | 1,235230199 | 0,216744843 | 0,998311993 | YES |
| 385 | 100,5747759 | -0,003616553 | 0,003001482 | -1,204922415 | 0,228233255 | 0,998311993 | YES |
| 386 | 723,5098522 | 0,003116642 | 0,002617093 | 1,190879315 | 0,233700966 | 0,998311993 | YES |
| 387 | 33,45446218 | 0,007428935 | 0,00628795 | 1,181455717 | 0,237421735 | 0,998311993 | YES |
| 388 | 236,0654733 | 0,000893406 | 0,000786128 | 1,136464159 | 0,255762357 | 0,998311993 | YES |
| 389 | 26,30575282 | -0,001888874 | 0,001750125 | -1,079279415 | 0,280463186 | 0,998311993 | YES |

| Medication | Ticker | ASVnr | taxLevel | Phylum | Family | Genus | Species |
|---|---|---|---|---|---|---|---|
| DA LEDD | 390 | ASV3 | Genus | Firmicutes | Ruminococcaceae | Subdoligranulum | NA |
| DA LEDD | 391 | ASV512 | Genus | Proteobacteria | Oxalobacteraceae | Oxalobacter | NA |
| DA LEDD | 392 | ASV10 | Genus | Verrucomicrobiota | Akkermansiaceae | Akkermansia | NA |
| DA LEDD | 393 | ASV309 | Genus | Firmicutes | Oscillospiraceae | Colidextribacter | NA |
| DA LEDD | 394 | ASV158 | Genus | Firmicutes | Lachnospiraceae | [Eubacterium] ventriosum group | NA |
| DA LEDD | 395 | ASV408 | Genus | Bacteroidota | Prevotellaceae | Paraprevotella | NA |
| DA LEDD | 396 | ASV57 | Genus | Firmicutes | Lachnospiraceae | Roseburia | NA |
| DA LEDD | 397 | ASV28 | Genus | Firmicutes | Clostridiaceae | Clostridium sensu stricto 1 | NA |
| DA LEDD | 398 | ASV630 | Genus | Firmicutes | Ruminococcaceae | Angelakisella | NA |
| DA LEDD | 399 | ASV76 | Genus | Firmicutes | Lachnospiraceae | Lachnospiraceae ND3007 group | NA |
| DA LEDD | 400 | ASV128 | Genus | Firmicutes | Oscillospiraceae | NK4A214 group | NA |
| DA LEDD | 401 | ASV303 | Genus | Firmicutes | Lachnospiraceae | GCA-900066575 | NA |
| DA LEDD | 402 | ASV15 | Genus | Firmicutes | Christensenellaceae | Christensenellaceae R-7 group | NA |
| DA LEDD | 403 | ASV97 | Genus | Firmicutes | Lachnospiraceae | [Ruminococcus] gnavus group | NA |
| DA LEDD | 404 | ASV226 | Genus | Firmicutes | Lachnospiraceae | Lachnospiraceae UCG-001 | NA |
| DA LEDD | 405 | ASV55 | Genus | Firmicutes | Monoglobaceae | Monoglobus | NA |
| DA LEDD | 406 | ASV144 | Genus | Desulfobacterota | Desulfovibrionaceae | Bilophila | NA |
| DA LEDD | 407 | ASV20 | Genus | Bacteroidota | Prevotellaceae | Prevotella | NA |
| DA LEDD | 408 | ASV33 | Genus | Firmicutes | Peptostreptococcaceae | Intestinibacter | NA |
| DA LEDD | 409 | ASV415 | Genus | Firmicutes | Oscillospiraceae | Oscillospira | NA |
| DA LEDD | 410 | ASV8 | Genus | Firmicutes | Ruminococcaceae | Faecalibacterium | NA |
| DA LEDD | 411 | ASV18 | Genus | Firmicutes | Lachnospiraceae | Dorea | NA |
| DA LEDD | 412 | ASV573 | Genus | Firmicutes | Erysipelatoclostridiaceae | Erysipelatoclostridium | NA |
| DA LEDD | 413 | ASV217 | Genus | Firmicutes | Peptococcaceae | Peptococcus | NA |
| DA LEDD | 414 | ASV87 | Genus | Proteobacteria | Sutterellaceae | Sutterella | NA |
| DA LEDD | 415 | ASV78 | Genus | Firmicutes | Lachnospiraceae | [Eubacterium] eligens group | NA |
| DA LEDD | 416 | ASV31 | Genus | Bacteroidota | Rikenellaceae | Alistipes | NA |
| DA LEDD | 417 | ASV75 | Genus | Firmicutes | Lachnospiraceae | [Ruminococcus] gauvreauii group | NA |
| DA LEDD | 418 | ASV7 | Genus | Firmicutes | Lachnospiraceae | Agathobacter | NA |
| DA LEDD | 419 | ASV81 | Genus | Firmicutes | Lachnospiraceae | Lachnospiraceae NK4A136 group | NA |

| Ticker | DESeq2_baseMean | DESeq2_log2-FoldChange | DESeq2_lfcSE | DESeq2_stat | DESeq2_pvalue | DESeq2_padj | DESeq2 converged |
|---|---|---|---|---|---|---|---|
| 390 | 1065,856619 | 0,001648814 | 0,001629284 | 1,011987179 | 0,311544183 | 0,998311993 | YES |
| 391 | 10,23479096 | 0,005152244 | 0,005127607 | 1,0048048 | 0,314990852 | 0,998311993 | YES |
| 392 | 861,3548571 | -0,003825228 | 0,003835124 | -0,997419548 | 0,318560907 | 0,998311993 | YES |
| 393 | 207,9260896 | 0,001662518 | 0,001709389 | 0,972580681 | 0,330761748 | 0,998311993 | YES |
| 394 | 251,7753773 | -0,002580404 | 0,002851794 | -0,904835422 | 0,365552583 | 0,998311993 | YES |
| 395 | 76,89042138 | -0,002908832 | 0,003217109 | -0,904175739 | 0,365902223 | 0,998311993 | YES |
| 396 | 546,2324732 | -0,002959976 | 0,003339995 | -0,88622182 | 0,375498008 | 0,998311993 | YES |
| 397 | 56,51096338 | -0,002799161 | 0,003173608 | -0,882012214 | 0,377770202 | 0,998311993 | YES |
| 398 | 24,46009323 | 0,004549159 | 0,005389158 | 0,84413163 | 0,398595855 | 0,998311993 | YES |
| 399 | 26,88123588 | -0,00415771 | 0,004971964 | -0,83623081 | 0,403025069 | 0,998311993 | YES |
| 400 | 384,9475 | 0,002024491 | 0,002502544 | 0,808973159 | 0,418530586 | 0,998311993 | YES |
| 401 | 29,27683927 | 0,002934564 | 0,003733296 | 0,786051802 | 0,431837139 | 0,998311993 | YES |
| 402 | 306,551642 | 0,002349472 | 0,003359202 | 0,699413787 | 0,484293474 | 0,998311993 | YES |
| 403 | 132,4461467 | -0,006318061 | 0,009054533 | -0,697778746 | 0,485315573 | 0,998311993 | YES |
| 404 | 35,98088275 | -0,005551617 | 0,008138272 | -0,68216158 | 0,495136786 | 0,998311993 | YES |
| 405 | 225,7522721 | 0,001152504 | 0,001890307 | 0,609691698 | 0,542066055 | 0,998311993 | YES |
| 406 | 65,01195307 | -0,002026793 | 0,003354053 | -0,604281866 | 0,545656261 | 0,998311993 | YES |
| 407 | 1202,055699 | -0,002469684 | 0,004157574 | -0,594020396 | 0,552498472 | 0,998311993 | YES |
| 408 | 46,54954651 | -0,002510289 | 0,004247239 | -0,591040265 | 0,554493443 | 0,998311993 | YES |
| 409 | 33,72458445 | -0,001144797 | 0,001982334 | -0,577499543 | 0,563602047 | 0,998311993 | YES |
| 410 | 2320,25665 | -0,000520968 | 0,000927258 | -0,561837036 | 0,574227056 | 0,998311993 | YES |
| 411 | 758,8763265 | -0,000801361 | 0,001467495 | -0,546073644 | 0,585015316 | 0,998311993 | YES |
| 412 | 75,75037649 | -0,001563335 | 0,002871626 | -0,544407608 | 0,586161012 | 0,998311993 | YES |
| 413 | 63,6478755 | -0,002963155 | 0,005491228 | -0,539616095 | 0,589461815 | 0,998311993 | YES |
| 414 | 123,5047879 | 0,001466231 | 0,002722634 | 0,53853386 | 0,590208535 | 0,998311993 | YES |
| 415 | 133,936392 | -0,002456912 | 0,004613837 | -0,532509386 | 0,594373242 | 0,998311993 | YES |
| 416 | 1042,659174 | 0,001089433 | 0,002046115 | 0,532439651 | 0,594421529 | 0,998311993 | YES |
| 417 | 27,28353221 | 0,004213281 | 0,009042067 | 0,465964401 | 0,641240982 | 0,998311993 | YES |
| 418 | 2890,414456 | 0,000576462 | 0,001311668 | 0,439487611 | 0,660308257 | 0,998311993 | YES |
| 419 | 642,5453446 | 0,001150522 | 0,002657844 | 0,432877974 | 0,665103422 | 0,998311993 | YES |

| Medication | Ticker | ASVnr | taxLevel | Phylum | Family | Genus | Species |
|---|---|---|---|---|---|---|---|
| DA LEDD | 420 | ASV147 | Genus | Firmicutes | Oscillospiraceae | Oscillibacter | NA |
| DA LEDD | 421 | ASV172 | Genus | Proteobacteria | Pasteurellaceae | Haemophilus | NA |
| DA LEDD | 422 | ASV134 | Genus | Actinobacteriota | Eggerthellaceae | Enterorhabdus | NA |
| DA LEDD | 423 | ASV27 | Genus | Firmicutes | Ruminococcaceae | CAG-352 | NA |
| DA LEDD | 424 | ASV376 | Genus | Bacteroidota | Barnesiellaceae | Coprobacter | NA |
| DA LEDD | 425 | ASV26 | Genus | Euryarchaeota | Methanobacteriaceae | Methanobrevibacter | NA |
| DA LEDD | 426 | ASV242 | Genus | Firmicutes | Lachnospiraceae | Tyzzerella | NA |
| DA LEDD | 427 | ASV690 | Genus | Firmicutes | Ruminococcaceae | Fournierella | NA |
| DA LEDD | 428 | ASV974 | Genus | Firmicutes | Butyricicoccaceae | UCG-009 | NA |
| DA LEDD | 429 | ASV34 | Genus | Firmicutes | Lachnospiraceae | Anaerostipes | NA |
| DA LEDD | 430 | ASV885 | Genus | Firmicutes | Lachnospiraceae | Shuttleworthia | NA |
| DA LEDD | 431 | ASV60 | Genus | Firmicutes | Ruminococcaceae | [Eubacterium] siraeum group | NA |
| DA LEDD | 432 | ASV19 | Genus | Firmicutes | Erysipelatoclostridiaceae | Erysipelotrichaceae UCG-003 | NA |
| DA LEDD | 433 | ASV229 | Genus | Firmicutes | Oscillospiraceae | Flavonifractor | NA |
| DA LEDD | 434 | ASV190 | Genus | Firmicutes | Ruminococcaceae | UBA1819 | NA |
| DA LEDD | 435 | ASV143 | Genus | Firmicutes | Peptostreptococcaceae | Terrisporobacter | NA |
| DA LEDD | 436 | ASV39 | Genus | Bacteroidota | Tannerellaceae | Parabacteroides | NA |
| DA LEDD | 437 | ASV453 | Genus | Verrucomicrobiota | Victivallaceae | Victivallis | NA |
| DA LEDD | 438 | ASV12 | Genus | Firmicutes | Veillonellaceae | Dialister | NA |
| DA LEDD | 439 | ASV133 | Genus | Actinobacteriota | Eggerthellaceae | Senegalimassilia | NA |
| DA LEDD | 440 | ASV17 | Genus | Actinobacteriota | Coriobacteriaceae | Collinsella | NA |
| DA LEDD | 441 | ASV29 | Genus | Firmicutes | Oscillospiraceae | UCG-002 | NA |
| DA LEDD | 442 | ASV283 | Genus | Actinobacteriota | Eggerthellaceae | Slackia | NA |
| DA LEDD | 443 | ASV116 | Genus | Firmicutes | Oscillospiraceae | UCG-003 | NA |
| DA LEDD | 444 | ASV1319 | Genus | Firmicutes | Ruminococcaceae | Candidatus Soleaferrea | NA |
| DA LEDD | 445 | ASV214 | Genus | Firmicutes | Lachnospiraceae | Lachnospiraceae UCG-010 | NA |
| DA LEDD | 446 | ASV1631 | Genus | Firmicutes | Ruminococcaceae | Paludicola | NA |
| DA LEDD | 447 | ASV98 | Genus | Firmicutes | Acidaminococcaceae | Phascolarctobacterium | NA |
| DA LEDD | 448 | ASV252 | Genus | Firmicutes | Erysipelotrichaceae | Solobacterium | NA |
| DA LEDD | 449 | ASV72 | Genus | Proteobacteria | Sutterellaceae | Parasutterella | NA |

| Ticker | DESeq2_baseMean | DESeq2_log2-FoldChange | DESeq2_lfcSE | DESeq2_stat | DESeq2_pvalue | DESeq2_padj | DESeq2 converged |
|---|---|---|---|---|---|---|---|
| 420 | 267,8988818 | 0,000907459 | 0,002110256 | 0,430023195 | 0,667178769 | 0,998311993 | YES |
| 421 | 40,05658099 | -0,003654967 | 0,008533225 | -0,428321855 | 0,668416809 | 0,998311993 | YES |
| 422 | 9,34864251 | -0,002317835 | 0,005541073 | -0,418300675 | 0,675727296 | 0,998311993 | YES |
| 423 | 183,1689752 | 0,003721026 | 0,009081881 | 0,409719717 | 0,682011565 | 0,998311993 | YES |
| 424 | 7,703336528 | 0,003640727 | 0,00917902 | 0,396635733 | 0,691636098 | 0,998311993 | YES |
| 425 | 84,42452394 | 0,00127808 | 0,003346262 | 0,381942507 | 0,70250401 | 0,998311993 | YES |
| 426 | 11,54643079 | 0,003173699 | 0,009160603 | 0,346450866 | 0,729003904 | 0,998311993 | YES |
| 427 | 1,672806929 | 0,00308319 | 0,009093012 | 0,339072504 | 0,734555111 | 0,998311993 | YES |
| 428 | 13,40497275 | 0,001744216 | 0,005257417 | 0,331762988 | 0,740068234 | 0,998311993 | YES |
| 429 | 166,5056402 | 0,000610088 | 0,001998278 | 0,305306966 | 0,760132371 | 0,998311993 | YES |
| 430 | 19,52623002 | -0,001141282 | 0,003813732 | -0,299255995 | 0,764744727 | 0,998311993 | YES |
| 431 | 486,2965926 | 0,000847139 | 0,002838422 | 0,298454314 | 0,76535644 | 0,998311993 | YES |
| 432 | 133,7131249 | -0,001456771 | 0,005101032 | -0,285583657 | 0,775197022 | 0,998311993 | YES |
| 433 | 108,9075689 | -0,000988997 | 0,003566813 | -0,277277526 | 0,781567015 | 0,998311993 | YES |
| 434 | 24,55110178 | 0,001505685 | 0,005756587 | 0,26155868 | 0,793661703 | 0,998311993 | YES |
| 435 | 6,873461442 | -0,002392828 | 0,009229427 | -0,259260708 | 0,795434093 | 0,998311993 | YES |
| 436 | 662,4121459 | 0,000709758 | 0,002767332 | 0,256477275 | 0,797582326 | 0,998311993 | YES |
| 437 | 1,446560666 | -0,00226511 | 0,00919469 | -0,246349751 | 0,8054115 | 0,998311993 | YES |
| 438 | 1196,04593 | 0,002175837 | 0,008958852 | 0,242870116 | 0,808106016 | 0,998311993 | YES |
| 439 | 6,290512682 | 0,002092959 | 0,00907484 | 0,230633181 | 0,817599788 | 0,998311993 | YES |
| 440 | 438,0900912 | 0,000444047 | 0,002214596 | 0,200509074 | 0,841082462 | 0,998311993 | YES |
| 441 | 1408,342623 | 0,000465926 | 0,002489741 | 0,18713818 | 0,851552294 | 0,998311993 | YES |
| 442 | 3,66075168 | 0,001703298 | 0,009117071 | 0,186825107 | 0,851797761 | 0,998311993 | YES |
| 443 | 183,3027975 | -0,000794301 | 0,004394437 | -0,180751542 | 0,856562601 | 0,998311993 | YES |
| 444 | 1,072416593 | -0,001617864 | 0,009070438 | -0,178366663 | 0,858435029 | 0,998311993 | YES |
| 445 | 83,90158914 | -0,000511103 | 0,0030115 | -0,169717055 | 0,865232661 | 0,998311993 | YES |
| 446 | 4,924808752 | 0,00147514 | 0,00902684 | 0,16341712 | 0,870190024 | 0,998311993 | YES |
| 447 | 347,7782958 | -0,000595873 | 0,003936657 | -0,151365253 | 0,879687597 | 0,998311993 | YES |
| 448 | 13,59437992 | -0,001301132 | 0,009210422 | -0,141267345 | 0,887658746 | 0,998311993 | YES |
| 449 | 117,353991 | -0,000478392 | 0,003422766 | -0,139767621 | 0,888843597 | 0,998311993 | YES |

| Medication | Ticker | ASVnr | taxLevel | Phylum | Family | Genus | Species |
|---|---|---|---|---|---|---|---|
| DA LEDD | 450 | ASV449 | Genus | Actinobacteriota | Atopobiaceae | Olsenella | NA |
| DA LEDD | 451 | ASV167 | Genus | Firmicutes | Oscillospiraceae | Intestinimonas | NA |
| DA LEDD | 452 | ASV211 | Genus | Firmicutes | Lachnospiraceae | Lachnospiraceae FCS020 group | NA |
| DA LEDD | 453 | ASV258 | Genus | Firmicutes | Lachnospiraceae | Lachnospiraceae AC2044 group | NA |
| DA LEDD | 454 | ASV162 | Genus | Desulfobacterota | Desulfovibrionaceae | Desulfovibrio | NA |
| DA LEDD | 455 | ASV550 | Genus | Bacteroidota | Rikenellaceae | Rikenellaceae RC9 gut group | NA |
| DA LEDD | 456 | ASV240 | Genus | Bacteroidota | Prevotellaceae | Alloprevotella | NA |
| DA LEDD | 457 | ASV195 | Genus | Bacteroidota | Barnesiellaceae | Barnesiella | NA |
| DA LEDD | 458 | ASV85 | Genus | Firmicutes | Lachnospiraceae | Coprococcus | NA |
| DA LEDD | 459 | ASV530 | Genus | Firmicutes | Veillonellaceae | Allisonella | NA |
| DA LEDD | 460 | ASV306 | Genus | Firmicutes | Veillonellaceae | Veillonella | NA |
| DA LEDD | 461 | ASV739 | Genus | Firmicutes | Ruminococcaceae | Anaerotruncus | NA |
| DA LEDD | 462 | ASV247 | Genus | Euryarchaeota | Methanobacteriaceae | Methanosphaera | NA |
| DA LEDD | 463 | ASV293 | Genus | Firmicutes | Erysipelotrichaceae | Catenisphaera | NA |
| DA LEDD | 464 | ASV499 | Genus | Firmicutes | Lachnospiraceae | Eisenbergiella | NA |
| DA LEDD | 465 | ASV444 | Genus | Firmicutes | Erysipelatoclostridiaceae | Coprobacillus | NA |
| DA LEDD | 466 | ASV40 | Genus | Firmicutes | Erysipelotrichaceae | Holdemanella | NA |
| DA LEDD | 467 | ASV79 | Genus | Firmicutes | Lachnospiraceae | [Eubacterium] ruminantium group | NA |
| DA LEDD | 468 | ASV1722 | Genus | Firmicutes | Erysipelatoclostridiaceae | Asteroleplasma | NA |
| DA LEDD | 469 | ASV219 | Genus | Firmicutes | Lachnospiraceae | Lachnospiraceae UCG-004 | NA |
| DA LEDD | 470 | ASV30 | Genus | Proteobacteria | Enterobacteriaceae | Escherichia-Shigella | NA |
| DA LEDD | 471 | ASV294 | Genus | Firmicutes | Anaerovoracaceae | Mogibacterium | NA |
| DA LEDD | 472 | ASV61 | Genus | Firmicutes | Erysipelatoclostridiaceae | Catenibacterium | NA |
| DA LEDD | 473 | ASV978 | Genus | Firmicutes | Erysipelotrichaceae | Dielma | NA |
| DA LEDD | 474 | ASV1506 | Genus | Firmicutes | Erysipelatoclostridiaceae | UCG-004 | NA |
| DA LEDD | 475 | ASV232 | Genus | Firmicutes | Acholeplasmataceae | Anaeroplasma | NA |
| DA LEDD | 476 | ASV666 | Genus | Firmicutes | Erysipelotrichaceae | Holdemania | NA |
| DA LEDD | 477 | ASV66 | Genus | Firmicutes | Butyricicoccaceae | Butyricicoccus | NA |
| DA LEDD | 478 | ASV338 | Genus | Firmicutes | Lachnospiraceae | Lachnotalea | NA |
| DA LEDD | 479 | ASV299 | Genus | Firmicutes | Streptococcaceae | Lactococcus | NA |

| Ticker | DESeq2_baseMean | DESeq2_log2-FoldChange | DESeq2_lfcSE | DESeq2_stat | DESeq2_pvalue | DESeq2_padj | DESeq2 converged |
|---|---|---|---|---|---|---|---|
| 450 | 1,366993497 | 0,001234158 | 0,009244955 | 0,133495324 | 0,893801662 | 0,998311993 | YES |
| 451 | 177,2590394 | -0,000252835 | 0,001979354 | -0,127736363 | 0,898357612 | 0,998311993 | YES |
| 452 | 32,84736195 | 0,000262697 | 0,00207873 | 0,126373779 | 0,899436057 | 0,998311993 | YES |
| 453 | 7,831182968 | 0,001131242 | 0,009211075 | 0,122813219 | 0,902255006 | 0,998311993 | YES |
| 454 | 72,4324127 | 0,000784699 | 0,006404359 | 0,122525841 | 0,902482582 | 0,998311993 | YES |
| 455 | 102,0607186 | -0,001072953 | 0,009274908 | -0,115683408 | 0,907903457 | 0,998311993 | YES |
| 456 | 190,3388284 | -0,001032936 | 0,00909646 | -0,113553582 | 0,909591686 | 0,998311993 | YES |
| 457 | 227,6580989 | 0,000262759 | 0,002323449 | 0,113090207 | 0,909959039 | 0,998311993 | YES |
| 458 | 495,238952 | 0,000292951 | 0,002695501 | 0,10868148 | 0,913455131 | 0,998311993 | YES |
| 459 | 9,31522785 | -0,000979386 | 0,009207146 | -0,106372402 | 0,915286889 | 0,998311993 | YES |
| 460 | 67,34805537 | -0,000927845 | 0,009101363 | -0,101945706 | 0,918799771 | 0,998311993 | YES |
| 461 | 2,34582764 | -0,000929545 | 0,009138754 | -0,101714642 | 0,91898318 | 0,998311993 | YES |
| 462 | 6,87489613 | 0,000898512 | 0,009213444 | 0,09752188 | 0,922311959 | 0,998311993 | YES |
| 463 | 8,934247857 | 0,000890204 | 0,00914662 | 0,097326025 | 0,922467489 | 0,998311993 | YES |
| 464 | 26,77826566 | 0,000815494 | 0,009123937 | 0,089379581 | 0,928780251 | 0,998311993 | YES |
| 465 | 1,947881622 | 0,000753818 | 0,00927751 | 0,081252212 | 0,935241377 | 0,998311993 | YES |
| 466 | 147,8981584 | -0,000681339 | 0,009239348 | -0,0737432 | 0,941214723 | 0,998311993 | YES |
| 467 | 13,09952607 | 0,000633645 | 0,009277028 | 0,068302603 | 0,945544752 | 0,998311993 | YES |
| 468 | 26,50145184 | 0,000600749 | 0,009218794 | 0,065165672 | 0,948042093 | 0,998311993 | YES |
| 469 | 67,12358874 | 0,000302751 | 0,00542907 | 0,055764779 | 0,955529193 | 0,998311993 | YES |
| 470 | 116,2531639 | 0,000249362 | 0,005186371 | 0,048080301 | 0,961652245 | 0,998311993 | YES |
| 471 | 7,003045608 | 0,000414907 | 0,009277098 | 0,044723761 | 0,964327494 | 0,998311993 | YES |
| 472 | 64,06289774 | -0,000414557 | 0,009272908 | -0,044706212 | 0,964341482 | 0,998311993 | YES |
| 473 | 0,502482894 | 0,000388023 | 0,009262234 | 0,041893002 | 0,966583995 | 0,998311993 | YES |
| 474 | 0,407412491 | -0,000376879 | 0,009235823 | -0,040806228 | 0,967450374 | 0,998311993 | YES |
| 475 | 59,87875326 | 0,000372804 | 0,009276963 | 0,040185964 | 0,967944868 | 0,998311993 | YES |
| 476 | 5,55987889 | 0,000121928 | 0,003183584 | 0,038298832 | 0,969449422 | 0,998311993 | YES |
| 477 | 193,8532185 | -3,73E-05 | 0,001065571 | -0,035044202 | 0,972044494 | 0,998311993 | YES |
| 478 | 10,50194988 | -0,000284767 | 0,009224682 | -0,03087008 | 0,975373151 | 0,998311993 | YES |
| 479 | 9,575547846 | -0,000190652 | 0,00925848 | -0,020592094 | 0,983571047 | 0,998311993 | YES |

| Medication | Ticker | ASVnr | taxLevel | Phylum | Family | Genus | Species |
|---|---|---|---|---|---|---|---|
| DA LEDD | 480 | ASV454 | Genus | Actinobacteriota | Actinomycetaceae | Actinomyces | NA |
| DA LEDD | 481 | ASV487 | Genus | Firmicutes | Lachnospiraceae | Howardella | NA |
| DA LEDD | 482 | ASV858 | Genus | Actinobacteriota | Atopobiaceae | Libanicoccus | NA |
| DA LEDD | 483 | ASV315 | Genus | Firmicutes | Anaerovoracaceae | Family XIII UCG-001 | NA |
| DA LEDD | 484 | ASV4 | Genus | Firmicutes | Ruminococcaceae | Ruminococcus | NA |
| DA LEDD | 485 | ASV197 | Genus | Bacteroidota | Prevotellaceae | Prevotellaceae NK3B31 group | NA |
| DA LEDD | 486 | ASV761 | Genus | Firmicutes | Defluviitaleaceae | Defluviitaleaceae UCG-011 | NA |
| DA LEDD | 487 | ASV411 | Family | Firmicutes | Lactobacillaceae | NA | NA |
| DA LEDD | 488 | ASV1 | Family | Actinobacteriota | Bifidobacteriaceae | NA | NA |
| DA LEDD | 489 | ASV322 | Family | Firmicutes | Anaerovoracaceae | NA | NA |
| DA LEDD | 490 | ASV192 | Family | Bacteroidota | Marinifilaceae | NA | NA |
| DA LEDD | 491 | ASV12 | Family | Firmicutes | Veillonellaceae | NA | NA |
| DA LEDD | 492 | ASV63 | Family | Actinobacteriota | Eggerthellaceae | NA | NA |
| DA LEDD | 493 | ASV15 | Family | Firmicutes | Christensenellaceae | NA | NA |
| DA LEDD | 494 | ASV59 | Family | Firmicutes | [Eubacterium] coprostanoligenes group | NA | NA |
| DA LEDD | 495 | ASV2 | Family | Bacteroidota | Bacteroidaceae | NA | NA |
| DA LEDD | 496 | ASV29 | Family | Firmicutes | Oscillospiraceae | NA | NA |
| DA LEDD | 497 | ASV20 | Family | Bacteroidota | Prevotellaceae | NA | NA |
| DA LEDD | 498 | ASV217 | Family | Firmicutes | Peptococcaceae | NA | NA |
| DA LEDD | 499 | ASV66 | Family | Firmicutes | Butyricicoccaceae | NA | NA |
| DA LEDD | 500 | ASV23 | Family | Firmicutes | Streptococcaceae | NA | NA |
| DA LEDD | 501 | ASV55 | Family | Firmicutes | Monoglobaceae | NA | NA |
| DA LEDD | 502 | ASV512 | Family | Proteobacteria | Oxalobacteraceae | NA | NA |
| DA LEDD | 503 | ASV4 | Family | Firmicutes | Ruminococcaceae | NA | NA |
| DA LEDD | 504 | ASV31 | Family | Bacteroidota | Rikenellaceae | NA | NA |
| DA LEDD | 505 | ASV16 | Family | Firmicutes | Peptostreptococcaceae | NA | NA |
| DA LEDD | 506 | ASV19 | Family | Firmicutes | Erysipelatoclostridiaceae | NA | NA |
| DA LEDD | 507 | ASV28 | Family | Firmicutes | Clostridiaceae | NA | NA |
| DA LEDD | 508 | ASV255 | Family | Actinobacteriota | Coriobacteriales Incertae Sedis | NA | NA |
| DA LEDD | 509 | ASV417 | Family | Firmicutes | UCG-010 | NA | NA |

| Ticker | DESeq2_baseMean | DESeq2_log2-FoldChange | DESeq2_lfcSE | DESeq2_stat | DESeq2_pvalue | DESeq2_padj | DESeq2 converged |
|---|---|---|---|---|---|---|---|
| 480 | 1,21009094 | 0,000190269 | 0,009258138 | 0,020551504 | 0,983603427 | 0,998311993 | YES |
| 481 | 5,48928889 | -0,000179886 | 0,009217315 | -0,019516091 | 0,984429401 | 0,998311993 | YES |
| 482 | 4,806469958 | -0,000106656 | 0,009287726 | -0,011483584 | 0,990837627 | 0,998311993 | YES |
| 483 | 19,59074966 | 5,89E-05 | 0,00533119 | 0,01105417 | 0,991180228 | 0,998311993 | YES |
| 484 | 1506,084107 | 9,38E-06 | 0,002597336 | 0,003610767 | 0,997119031 | 0,998311993 | YES |
| 485 | 127,0218344 | -3,04E-05 | 0,009258287 | -0,003288243 | 0,997376366 | 0,998311993 | YES |
| 486 | 0,534033279 | -1,96E-05 | 0,00925421 | -0,002115604 | 0,998311993 | 0,998311993 | YES |
| 487 | 105,0044887 | 0,010019525 | 0,003425014 | 2,925396744 | 0,003440173 | 0,158247968 | YES |
| 488 | 1051,128543 | 0,003886717 | 0,001510568 | 2,573016774 | 0,010081632 | 0,23187754 | YES |
| 489 | 97,51950525 | 0,00303579 | 0,00125881 | 2,411635119 | 0,015881166 | 0,243511214 | YES |
| 490 | 45,35172578 | -0,002508558 | 0,001214526 | -2,065461912 | 0,038879316 | 0,445696897 | YES |
| 491 | 1608,066586 | 0,003997955 | 0,00202588 | 1,973441223 | 0,048445315 | 0,445696897 | YES |
| 492 | 154,3967646 | 0,003504022 | 0,001859192 | 1,884701568 | 0,059470141 | 0,455937745 | YES |
| 493 | 502,0114562 | 0,004687524 | 0,002656671 | 1,764435087 | 0,077658755 | 0,510328963 | YES |
| 494 | 328,8566557 | 0,002154005 | 0,001312088 | 1,641661689 | 0,100660135 | 0,578795778 | YES |
| 495 | 7180,909544 | -0,001133064 | 0,000728444 | -1,555458397 | 0,119836935 | 0,612499892 | YES |
| 496 | 3327,623644 | 0,001451884 | 0,00098456 | 1,474652158 | 0,140306084 | 0,645407988 | YES |
| 497 | 1620,824833 | -0,002312538 | 0,001757442 | -1,315854037 | 0,188223038 | 0,737627961 | YES |
| 498 | 72,25285468 | -0,002385959 | 0,001846723 | -1,291996586 | 0,196358327 | 0,737627961 | YES |
| 499 | 213,8474286 | 0,000882867 | 0,000714154 | 1,236241644 | 0,216368755 | 0,737627961 | YES |
| 500 | 781,8006256 | 0,003073779 | 0,002530565 | 1,21466086 | 0,224495466 | 0,737627961 | YES |
| 501 | 244,6838494 | 0,00172969 | 0,001834631 | 0,942799867 | 0,34578328 | 0,899211575 | YES |
| 502 | 8,282395154 | 0,005495772 | 0,005898099 | 0,931786999 | 0,351446615 | 0,899211575 | YES |
| 503 | 6687,853491 | 0,000715162 | 0,000792838 | 0,902027916 | 0,367042038 | 0,899211575 | YES |
| 504 | 1099,133854 | 0,001365258 | 0,001546356 | 0,882887119 | 0,377297264 | 0,899211575 | YES |
| 505 | 101,3902113 | 0,002214934 | 0,002525708 | 0,876955525 | 0,380510795 | 0,899211575 | YES |
| 506 | 281,8983195 | -0,001624064 | 0,001988997 | -0,816524379 | 0,414200286 | 0,899211575 | YES |
| 507 | 56,99955518 | -0,002565104 | 0,003169662 | -0,809267396 | 0,418361356 | 0,899211575 | YES |
| 508 | 40,1387451 | 0,002873298 | 0,003641267 | 0,789092903 | 0,43005771 | 0,899211575 | YES |
| 509 | 77,65154318 | 0,002817654 | 0,004229454 | 0,666198005 | 0,505284548 | 0,965151096 | YES |

| Medication | Ticker | ASVnr | taxLevel | Phylum | Family | Genus | Species |
|---|---|---|---|---|---|---|---|
| DA LEDD | 510 | ASV39 | Family | Bacteroidota | Tannerellaceae | NA | NA |
| DA LEDD | 511 | ASV195 | Family | Bacteroidota | Barnesiellaceae | NA | NA |
| DA LEDD | 512 | ASV26 | Family | Euryarchaeota | Methanobacteriaceae | NA | NA |
| DA LEDD | 513 | ASV162 | Family | Desulfobacterota | Desulfovibrionaceae | NA | NA |
| DA LEDD | 514 | ASV17 | Family | Actinobacteriota | Coriobacteriaceae | NA | NA |
| DA LEDD | 515 | ASV10 | Family | Verrucomicrobiota | Akkermansiaceae | NA | NA |
| DA LEDD | 516 | ASV40 | Family | Firmicutes | Erysipelotrichaceae | NA | NA |
| DA LEDD | 517 | ASV449 | Family | Actinobacteriota | Atopobiaceae | NA | NA |
| DA LEDD | 518 | ASV172 | Family | Proteobacteria | Pasteurellaceae | NA | NA |
| DA LEDD | 519 | ASV30 | Family | Proteobacteria | Enterobacteriaceae | NA | NA |
| DA LEDD | 520 | ASV453 | Family | Verrucomicrobiota | Victivallaceae | NA | NA |
| DA LEDD | 521 | ASV72 | Family | Proteobacteria | Sutterellaceae | NA | NA |
| DA LEDD | 522 | ASV145 | Family | Bacteroidota | Muribaculaceae | NA | NA |
| DA LEDD | 523 | ASV7 | Family | Firmicutes | Lachnospiraceae | NA | NA |
| DA LEDD | 524 | ASV1119 | Family | Verrucomicrobiota | vadinBE97 | NA | NA |
| DA LEDD | 525 | ASV232 | Family | Firmicutes | Acholeplasmataceae | NA | NA |
| DA LEDD | 526 | ASV1052 | Family | Thermoplasmatota | Methanomethylophilaceae | NA | NA |
| DA LEDD | 527 | ASV454 | Family | Actinobacteriota | Actinomycetaceae | NA | NA |
| DA LEDD | 528 | ASV761 | Family | Firmicutes | Defluviitaleaceae | NA | NA |
| DA LEDD | 529 | ASV98 | Family | Firmicutes | Acidaminococcaceae | NA | NA |
| DA LEDD | 530 | ASV802 | Family | Firmicutes | Ruminiclostridium | NA | NA |
| DA LEDD | 531 | ASV703 | Family | Firmicutes | Hydrogenoanaerobacterium | NA | NA |
| DA LEDD | 532 | ASV752 | Family | Verrucomicrobiota | Puniceicoccaceae | NA | NA |

| Ticker | DESeq2_baseMean | DESeq2_log2-FoldChange | DESeq2_lfcSE | DESeq2_stat | DESeq2_pvalue | DESeq2_padj | DESeq2 converged |
|---|---|---|---|---|---|---|---|
| 510 | 611,6429564 | 0,001427555 | 0,002351902 | 0,60697879 | 0,543864988 | 0,965151096 | YES |
| 511 | 252,5257347 | 0,001324583 | 0,002205967 | 0,600454627 | 0,548203291 | 0,965151096 | YES |
| 512 | 79,20511498 | 0,001659193 | 0,003159348 | 0,525169381 | 0,599465448 | 0,965151096 | YES |
| 513 | 137,0403272 | -0,001278115 | 0,002458847 | -0,51980253 | 0,603201216 | 0,965151096 | YES |
| 514 | 521,0394889 | 0,001033638 | 0,00211353 | 0,489057669 | 0,624800869 | 0,965151096 | YES |
| 515 | 761,6767525 | -0,004110012 | 0,008947942 | -0,459324899 | 0,64600087 | 0,965151096 | YES |
| 516 | 171,3521872 | -0,000790737 | 0,001772039 | -0,44623004 | 0,655431085 | 0,965151096 | YES |
| 517 | 6,066628147 | 0,003977243 | 0,009155653 | 0,434402982 | 0,663995833 | 0,965151096 | YES |
| 518 | 44,08973922 | -0,003115145 | 0,007343326 | -0,424214402 | 0,671409458 | 0,965151096 | YES |
| 519 | 133,7261228 | 0,001493258 | 0,004837111 | 0,308708535 | 0,757543251 | 0,98276578 | YES |
| 520 | 2,060516529 | -0,001985186 | 0,009191385 | -0,215983332 | 0,82900074 | 0,98276578 | YES |
| 521 | 223,3608361 | 0,000370896 | 0,001959165 | 0,189313492 | 0,849847121 | 0,98276578 | YES |
| 522 | 517,5474959 | -0,001254753 | 0,009174456 | -0,13676597 | 0,891215783 | 0,98276578 | YES |
| 523 | 11942,20647 | 8,77E-05 | 0,000814472 | 0,107662368 | 0,914263522 | 0,98276578 | YES |
| 524 | 0,147824868 | 0,000910901 | 0,009288679 | 0,098065732 | 0,921880098 | 0,98276578 | YES |
| 525 | 43,66029827 | 0,00084763 | 0,009276964 | 0,091369304 | 0,927199152 | 0,98276578 | YES |
| 526 | 2,948528745 | 0,000751791 | 0,009277219 | 0,081036206 | 0,935413159 | 0,98276578 | YES |
| 527 | 0,969820909 | 0,000658153 | 0,009258198 | 0,071088654 | 0,943327198 | 0,98276578 | YES |
| 528 | 0,432160341 | 0,000563399 | 0,009254242 | 0,060880073 | 0,951454719 | 0,98276578 | YES |
| 529 | 383,4330824 | 0,000211143 | 0,003630728 | 0,05815441 | 0,953625635 | 0,98276578 | YES |
| 530 | 4,965768924 | 0,000466658 | 0,009287715 | 0,050244693 | 0,959927397 | 0,98276578 | YES |
| 531 | 2,490877974 | -0,000423028 | 0,009255876 | -0,045703732 | 0,963546389 | 0,98276578 | YES |
| 532 | 1,494746013 | -0,000199847 | 0,009251519 | -0,021601571 | 0,98276578 | 0,98276578 | YES |

**Supplementary Table 8d: Pramipexole**

| Medication | Ticker | ASVnr | taxLevel | Phylum | Family | Genus | Species |
|---|---|---|---|---|---|---|---|
| Pramipexole | 1 | ASV15 | ASV | Firmicutes | Christensenellaceae | Christensenellaceae R-7 group | NA |
| Pramipexole | 2 | ASV145 | ASV | Bacteroidota | Muribaculaceae | NA | NA |
| Pramipexole | 3 | ASV42 | ASV | Actinobacteriota | Bifidobacteriaceae | Bifidobacterium | NA |
| Pramipexole | 4 | ASV27 | ASV | Firmicutes | Ruminococcaceae | CAG-352 | NA |
| Pramipexole | 5 | ASV13 | ASV | Firmicutes | Lachnospiraceae | Fusicatenibacter | saccharivorans |
| Pramipexole | 6 | ASV35 | ASV | Firmicutes | Lachnospiraceae | Agathobacter | NA |
| Pramipexole | 7 | ASV99 | ASV | Bacteroidota | Bacteroidaceae | Bacteroides | plebeius |
| Pramipexole | 8 | ASV44 | ASV | Bacteroidota | Bacteroidaceae | Bacteroides | NA |
| Pramipexole | 9 | ASV1 | ASV | Actinobacteriota | Bifidobacteriaceae | Bifidobacterium | NA |
| Pramipexole | 10 | ASV328 | ASV | Firmicutes | Anaerovoracaceae | [Eubacterium] brachy group | NA |
| Pramipexole | 11 | ASV3 | ASV | Firmicutes | Ruminococcaceae | Subdoligranulum | NA |
| Pramipexole | 12 | ASV57 | ASV | Firmicutes | Lachnospiraceae | Roseburia | inulinivorans |
| Pramipexole | 13 | ASV329 | ASV | Firmicutes | Lachnospiraceae | NA | NA |
| Pramipexole | 14 | ASV193 | ASV | Actinobacteriota | Eggerthellaceae | Adlercreutzia | equolifaciens |
| Pramipexole | 15 | ASV268 | ASV | Firmicutes | Ruminococcaceae | Incertae Sedis | NA |
| Pramipexole | 16 | ASV6 | ASV | Firmicutes | Ruminococcaceae | Faecalibacterium | NA |
| Pramipexole | 17 | ASV21 | ASV | Firmicutes | Ruminococcaceae | Faecalibacterium | prausnitzii |
| Pramipexole | 18 | ASV284 | ASV | Firmicutes | Oscillospiraceae | NK4A214 group | NA |
| Pramipexole | 19 | ASV92 | ASV | Firmicutes | Erysipelotrichaceae | Turicibacter | sanguinis |
| Pramipexole | 20 | ASV1030 | ASV | Firmicutes | NA | NA | NA |
| Pramipexole | 21 | ASV97 | ASV | Firmicutes | Lachnospiraceae | [Ruminococcus] gnavus group | NA |
| Pramipexole | 22 | ASV23 | ASV | Firmicutes | Streptococcaceae | Streptococcus | NA |
| Pramipexole | 23 | ASV630 | ASV | Firmicutes | Ruminococcaceae | Angelakisella | NA |
| Pramipexole | 24 | ASV138 | ASV | Bacteroidota | Rikenellaceae | Alistipes | obesi |
| Pramipexole | 25 | ASV10 | ASV | Verrucomicrobiota | Akkermansiaceae | Akkermansia | muciniphila |
| Pramipexole | 26 | ASV218 | ASV | Firmicutes | Anaerovoracaceae | Family XIII AD3011 group | NA |
| Pramipexole | 27 | ASV16 | ASV | Firmicutes | Peptostreptococcaceae | Romboutsia | NA |
| Pramipexole | 28 | ASV47 | ASV | Bacteroidota | Rikenellaceae | Alistipes | NA |
| Pramipexole | 29 | ASV216 | ASV | Firmicutes | Lachnospiraceae | Lachnospira | pectinoschiza |

| Ticker | DESeq2_baseMean | DESeq2_log2-FoldChange | DESeq2_lfcSE | DESeq2_stat | DESeq2_pvalue | DESeq2_padj | DESeq2 converged |
|---|---|---|---|---|---|---|---|
| 1 | 81,53264531 | -19,31687639 | 0,855405015 | -22,58214069 | 6,49E-113 | 1,97E-110 | YES |
| 2 | 60,60593301 | -17,62282477 | 0,855405216 | -20,60172704 | 2,65E-94 | 4,03E-92 | YES |
| 3 | 96,71036381 | 5,671565686 | 0,853809519 | 6,642659233 | 3,08E-11 | 3,12E-09 | YES |
| 4 | 411,9532093 | 3,619606302 | 0,842670837 | 4,295397611 | 1,74E-05 | 0,001325292 | YES |
| 5 | 902,9077777 | 2,090809601 | 0,60145344 | 3,476261773 | 0,000508456 | 0,030914108 | YES |
| 6 | 487,2873051 | 2,288004274 | 0,807539999 | 2,833301481 | 0,004606991 | 0,233420895 | YES |
| 7 | 99,52452745 | 1,857934806 | 0,842961526 | 2,204056471 | 0,027520373 | 0,999171269 | YES |
| 8 | 143,4182641 | -1,508405127 | 0,686450918 | -2,197396911 | 0,027992112 | 0,999171269 | YES |
| 9 | 538,6681246 | 1,032761311 | 0,487017313 | 2,120584391 | 0,033956793 | 0,999171269 | YES |
| 10 | 17,56213136 | 1,207685152 | 0,688117478 | 1,755056645 | 0,079249619 | 0,999171269 | YES |
| 11 | 872,8219076 | 1,190458548 | 0,78303217 | 1,520318823 | 0,128430866 | 0,999171269 | YES |
| 12 | 127,5742219 | -1,013931297 | 0,693775046 | -1,461469827 | 0,14388655 | 0,999171269 | YES |
| 13 | 21,61991235 | -1,118273951 | 0,816676212 | -1,369299039 | 0,170905818 | 0,999171269 | YES |
| 14 | 12,52817079 | 1,111588512 | 0,812512066 | 1,36808862 | 0,171284338 | 0,999171269 | YES |
| 15 | 44,02602912 | 0,900322185 | 0,660744708 | 1,362587053 | 0,173012691 | 0,999171269 | YES |
| 16 | 1287,39372 | 0,787563882 | 0,58528239 | 1,345613496 | 0,178427197 | 0,999171269 | YES |
| 17 | 688,2219105 | -0,800140263 | 0,634827177 | -1,260406442 | 0,207522779 | 0,999171269 | YES |
| 18 | 5,966074393 | -1,000160751 | 0,825345626 | -1,211808386 | 0,225585737 | 0,999171269 | YES |
| 19 | 7,956798089 | 0,989387183 | 0,820067941 | 1,206469773 | 0,227636407 | 0,999171269 | YES |
| 20 | 2,316101286 | 0,947552826 | 0,811249383 | 1,168016699 | 0,242800029 | 0,999171269 | YES |
| 21 | 2,792784839 | -0,905953753 | 0,830602758 | -1,09071845 | 0,27539679 | 0,999171269 | YES |
| 22 | 546,0398678 | 0,565468523 | 0,520803485 | 1,085761789 | 0,277584396 | 0,999171269 | YES |
| 23 | 35,12840033 | 0,678852221 | 0,655161975 | 1,036159372 | 0,300127797 | 0,999171269 | YES |
| 24 | 73,45126475 | 0,613755851 | 0,594096243 | 1,033091621 | 0,301561024 | 0,999171269 | YES |
| 25 | 435,6769742 | -0,721012679 | 0,703493306 | -1,024903396 | 0,305408772 | 0,999171269 | YES |
| 26 | 11,8953349 | 0,686735561 | 0,682326697 | 1,006461515 | 0,314193616 | 0,999171269 | YES |
| 27 | 35,47408363 | 0,679201291 | 0,683269582 | 0,994045849 | 0,320200547 | 0,999171269 | YES |
| 28 | 331,7576695 | 0,681902283 | 0,698996012 | 0,975545312 | 0,329289837 | 0,999171269 | YES |
| 29 | 83,48713658 | -0,656550161 | 0,679207332 | -0,966641747 | 0,333723158 | 0,999171269 | YES |

| Medication | Ticker | ASVnr | taxLevel | Phylum | Family | Genus | Species |
|---|---|---|---|---|---|---|---|
| Pramipexole | 30 | ASV25 | ASV | Actinobacteriota | Bifidobacteriaceae | Bifidobacterium | NA |
| Pramipexole | 31 | ASV820 | ASV | Firmicutes | Lachnospiraceae | GCA-900066575 | NA |
| Pramipexole | 32 | ASV7 | ASV | Firmicutes | Lachnospiraceae | Agathobacter | NA |
| Pramipexole | 33 | ASV221 | ASV | Firmicutes | Oscillospiraceae | Intestinimonas | NA |
| Pramipexole | 34 | ASV295 | ASV | Firmicutes | Ruminococcaceae | Negativibacillus | NA |
| Pramipexole | 35 | ASV413 | ASV | Firmicutes | Oscillospiraceae | Oscillibacter | NA |
| Pramipexole | 36 | ASV131 | ASV | Firmicutes | Lachnospiraceae | NA | NA |
| Pramipexole | 37 | ASV363 | ASV | Bacteroidota | Rikenellaceae | Alistipes | NA |
| Pramipexole | 38 | ASV341 | ASV | Firmicutes | Christensenellaceae | Christensenellaceae R-7 group | NA |
| Pramipexole | 39 | ASV26 | ASV | Euryarchaeota | Methanobacteriaceae | Methanobrevibacter | NA |
| Pramipexole | 40 | ASV4 | ASV | Firmicutes | Ruminococcaceae | Ruminococcus | bromii |
| Pramipexole | 41 | ASV30 | ASV | Proteobacteria | Enterobacteriaceae | Escherichia-Shigella | NA |
| Pramipexole | 42 | ASV879 | ASV | Firmicutes | NA | NA | NA |
| Pramipexole | 43 | ASV5 | ASV | Firmicutes | Lachnospiraceae | Blautia | NA |
| Pramipexole | 44 | ASV154 | ASV | Firmicutes | NA | NA | NA |
| Pramipexole | 45 | ASV14 | ASV | Firmicutes | Lachnospiraceae | Blautia | massiliensis |
| Pramipexole | 46 | ASV386 | ASV | Firmicutes | Ruminococcaceae | DTU089 | NA |
| Pramipexole | 47 | ASV423 | ASV | Firmicutes | Christensenellaceae | Christensenellaceae R-7 group | NA |
| Pramipexole | 48 | ASV262 | ASV | Firmicutes | Ruminococcaceae | Incertae Sedis | NA |
| Pramipexole | 49 | ASV283 | ASV | Actinobacteriota | Eggerthellaceae | Slackia | isoflavoniconvertens |
| Pramipexole | 50 | ASV508 | ASV | Firmicutes | Ruminococcaceae | NA | NA |
| Pramipexole | 51 | ASV124 | ASV | Firmicutes | NA | NA | NA |
| Pramipexole | 52 | ASV895 | ASV | Bacteroidota | Barnesiellaceae | Coprobacter | secundus |
| Pramipexole | 53 | ASV101 | ASV | Firmicutes | Lachnospiraceae | Roseburia | NA |
| Pramipexole | 54 | ASV172 | ASV | Proteobacteria | Pasteurellaceae | Haemophilus | NA |
| Pramipexole | 55 | ASV270 | ASV | Firmicutes | Christensenellaceae | Christensenellaceae R-7 group | NA |
| Pramipexole | 56 | ASV86 | ASV | Firmicutes | Lachnospiraceae | [Eubacterium] hallii group | NA |
| Pramipexole | 57 | ASV17 | ASV | Actinobacteriota | Coriobacteriaceae | Collinsella | aerofaciens |
| Pramipexole | 58 | ASV83 | ASV | Firmicutes | Lachnospiraceae | Lachnospiraceae NK4A136 group | NA |
| Pramipexole | 59 | ASV137 | ASV | Firmicutes | Butyricicoccaceae | Butyricicoccus | faecihominis |

| Ticker | DESeq2_baseMean | DESeq2_log2-FoldChange | DESeq2_lfcSE | DESeq2_stat | DESeq2_pvalue | DESeq2_padj | DESeq2 converged |
|---|---|---|---|---|---|---|---|
| 30 | 594,797018 | 0,406492494 | 0,433785125 | 0,937082603 | 0,34871607 | 0,999171269 | YES |
| 31 | 44,77586329 | 0,581340046 | 0,638435185 | 0,910570187 | 0,362521885 | 0,999171269 | YES |
| 32 | 3345,312936 | 0,457741684 | 0,526209918 | 0,869884182 | 0,384363701 | 0,999171269 | YES |
| 33 | 40,29552417 | -0,556343346 | 0,645484742 | -0,861900073 | 0,388742509 | 0,999171269 | YES |
| 34 | 95,31389306 | 0,434683329 | 0,517656297 | 0,839714172 | 0,401068666 | 0,999171269 | YES |
| 35 | 128,8129072 | 0,570434922 | 0,68463151 | 0,833199924 | 0,404731985 | 0,999171269 | YES |
| 36 | 187,7566819 | 0,580264133 | 0,702090537 | 0,826480493 | 0,408531579 | 0,999171269 | YES |
| 37 | 24,16993243 | 0,665871504 | 0,809272128 | 0,822802962 | 0,410620052 | 0,999171269 | YES |
| 38 | 40,84893866 | 0,464098106 | 0,564795011 | 0,821710704 | 0,411241566 | 0,999171269 | YES |
| 39 | 57,00131746 | 0,439054762 | 0,560288651 | 0,783622444 | 0,433261683 | 0,999171269 | YES |
| 40 | 829,5920807 | -0,614864583 | 0,807516875 | -0,761426296 | 0,446402486 | 0,999171269 | YES |
| 41 | 148,7567609 | 0,482008368 | 0,642061753 | 0,750719639 | 0,452821401 | 0,999171269 | YES |
| 42 | 2,790106318 | 0,609313044 | 0,833646753 | 0,730900758 | 0,464839773 | 0,999171269 | YES |
| 43 | 1308,754837 | -0,310480951 | 0,429362998 | -0,723119953 | 0,469606192 | 0,999171269 | YES |
| 44 | 66,5504132 | 0,570070776 | 0,808325856 | 0,705248721 | 0,480655466 | 0,999171269 | YES |
| 45 | 654,9178766 | 0,360815998 | 0,520657117 | 0,693001184 | 0,488308808 | 0,999171269 | YES |
| 46 | 37,23194595 | 0,439236065 | 0,648370446 | 0,677446154 | 0,498122923 | 0,999171269 | YES |
| 47 | 40,54368899 | 0,430623297 | 0,660111765 | 0,652349071 | 0,514176011 | 0,999171269 | YES |
| 48 | 123,2049149 | 0,43925504 | 0,681174404 | 0,644849597 | 0,519024657 | 0,999171269 | YES |
| 49 | 5,521707361 | 0,520757116 | 0,833172614 | 0,625029085 | 0,531951969 | 0,999171269 | YES |
| 50 | 6,352552955 | 0,515245199 | 0,83334528 | 0,618285375 | 0,536387241 | 0,999171269 | YES |
| 51 | 346,05527 | 0,499986589 | 0,810294105 | 0,61704335 | 0,537206133 | 0,999171269 | YES |
| 52 | 2,93437005 | 0,506960038 | 0,836672508 | 0,605924102 | 0,544565154 | 0,999171269 | YES |
| 53 | 75,89357853 | 0,48847622 | 0,808166395 | 0,604425305 | 0,545560916 | 0,999171269 | YES |
| 54 | 13,88186595 | -0,491710856 | 0,824375453 | -0,596464699 | 0,55086483 | 0,999171269 | YES |
| 55 | 21,80343709 | 0,333190247 | 0,566070102 | 0,588602447 | 0,556127992 | 0,999171269 | YES |
| 56 | 278,2596982 | 0,291727229 | 0,508834081 | 0,573324861 | 0,566424755 | 0,999171269 | YES |
| 57 | 835,9405041 | 0,244328334 | 0,442365859 | 0,552321861 | 0,580727856 | 0,999171269 | YES |
| 58 | 41,34021869 | 0,454523899 | 0,830300495 | 0,547420966 | 0,584089554 | 0,999171269 | YES |
| 59 | 17,18883172 | 0,450278212 | 0,82605229 | 0,5450965 | 0,58568715 | 9,99E-01 | YES |

| Medication | Ticker | ASVnr | taxLevel | Phylum | Family | Genus | Species |
|---|---|---|---|---|---|---|---|
| Pramipexole | 60 | ASV213 | ASV | Firmicutes | NA | NA | NA |
| Pramipexole | 61 | ASV470 | ASV | Firmicutes | Christensenellaceae | Christensenellaceae R-7 group | NA |
| Pramipexole | 62 | ASV100 | ASV | Firmicutes | Christensenellaceae | NA | NA |
| Pramipexole | 63 | ASV111 | ASV | Firmicutes | Lachnospiraceae | Lachnospira | pectinoschiza |
| Pramipexole | 64 | ASV126 | ASV | Bacteroidota | Tannerellaceae | Parabacteroides | distasonis |
| Pramipexole | 65 | ASV88 | ASV | Firmicutes | Lachnospiraceae | Dorea | formicigenerans |
| Pramipexole | 66 | ASV867 | ASV | Firmicutes | NA | NA | NA |
| Pramipexole | 67 | ASV201 | ASV | Firmicutes | Oscillospiraceae | UCG-002 | NA |
| Pramipexole | 68 | ASV87 | ASV | Proteobacteria | Sutterellaceae | Sutterella | wadsworthensis |
| Pramipexole | 69 | ASV512 | ASV | Proteobacteria | Oxalobacteraceae | Oxalobacter | formigenes |
| Pramipexole | 70 | ASV242 | ASV | Firmicutes | Lachnospiraceae | Tyzzerella | NA |
| Pramipexole | 71 | ASV141 | ASV | Firmicutes | Lachnospiraceae | Lachnoclostridium | NA |
| Pramipexole | 72 | ASV457 | ASV | Firmicutes | Lachnospiraceae | NA | NA |
| Pramipexole | 73 | ASV665 | ASV | Firmicutes | Lachnospiraceae | NA | NA |
| Pramipexole | 74 | ASV403 | ASV | Firmicutes | Oscillospiraceae | Colidextribacter | NA |
| Pramipexole | 75 | ASV229 | ASV | Firmicutes | Oscillospiraceae | Flavonifractor | plautii |
| Pramipexole | 76 | ASV70 | ASV | Firmicutes | Lachnospiraceae | Roseburia | intestinalis |
| Pramipexole | 77 | ASV63 | ASV | Actinobacteriota | Eggerthellaceae | NA | NA |
| Pramipexole | 78 | ASV77 | ASV | Firmicutes | Lachnospiraceae | [Ruminococcus] torques group | NA |
| Pramipexole | 79 | ASV184 | ASV | Firmicutes | Ruminococcaceae | Ruminococcus | NA |
| Pramipexole | 80 | ASV626 | ASV | Firmicutes | NA | NA | NA |
| Pramipexole | 81 | ASV238 | ASV | Firmicutes | Butyricicoccaceae | NA | NA |
| Pramipexole | 82 | ASV701 | ASV | Firmicutes | Lachnospiraceae | Lachnoclostridium | NA |
| Pramipexole | 83 | ASV28 | ASV | Firmicutes | Clostridiaceae | Clostridium sensu stricto 1 | NA |
| Pramipexole | 84 | ASV200 | ASV | Firmicutes | Lachnospiraceae | [Eubacterium] xylanophilum group | NA |
| Pramipexole | 85 | ASV503 | ASV | Firmicutes | Erysipelatoclostridiaceae | Erysipelatoclostridium | NA |
| Pramipexole | 86 | ASV190 | ASV | Firmicutes | Ruminococcaceae | UBA1819 | NA |
| Pramipexole | 87 | ASV12 | ASV | Firmicutes | Veillonellaceae | Dialister | invisus |
| Pramipexole | 88 | ASV806 | ASV | Firmicutes | Peptococcaceae | NA | NA |
| Pramipexole | 89 | ASV75 | ASV | Firmicutes | Lachnospiraceae | [Ruminococcus] gauvreauii group | NA |

| Ticker | DESeq2_baseMean | DESeq2_log2-FoldChange | DESeq2_lfcSE | DESeq2_stat | DESeq2_pvalue | DESeq2_padj | DESeq2 converged |
|---|---|---|---|---|---|---|---|
| 60 | 78,20423814 | 0,433044553 | 0,810220693 | 0,534477281 | 0,593011364 | 0,999171269 | YES |
| 61 | 7,038863537 | -0,429779824 | 0,813055618 | -0,528598308 | 0,597084134 | 0,999171269 | YES |
| 62 | 137,3160995 | 0,429769756 | 0,824137137 | 0,521478449 | 0,602033515 | 0,999171269 | YES |
| 63 | 72,2808885 | -0,343117689 | 0,670512841 | -0,511724263 | 0,608844003 | 0,999171269 | YES |
| 64 | 162,320947 | 0,373041079 | 0,731012879 | 0,510307122 | 0,609836314 | 0,999171269 | YES |
| 65 | 111,3930899 | 0,348118012 | 0,682786658 | 0,509848879 | 0,610157339 | 0,999171269 | YES |
| 66 | 1,895833441 | -0,427576375 | 0,841924027 | -0,507856245 | 0,611554162 | 0,999171269 | YES |
| 67 | 156,8534097 | 0,42154405 | 0,830080363 | 0,507835228 | 0,611568902 | 0,999171269 | YES |
| 68 | 36,89237627 | 0,410899433 | 0,812337489 | 0,505823551 | 0,612980524 | 0,999171269 | YES |
| 69 | 12,25359827 | 0,35048469 | 0,714651062 | 0,490427719 | 0,623831266 | 0,999171269 | YES |
| 70 | 7,906390886 | 0,398997442 | 0,833138588 | 0,478908848 | 0,632003476 | 0,999171269 | YES |
| 71 | 5,575019215 | 0,39773191 | 0,833487843 | 0,477189816 | 0,633226962 | 0,999171269 | YES |
| 72 | 4,865471315 | 0,397114714 | 0,833658959 | 0,476351522 | 0,633823966 | 0,999171269 | YES |
| 73 | 4,054559429 | 0,396148153 | 0,833925725 | 0,475040092 | 0,634758397 | 0,999171269 | YES |
| 74 | 7,860171156 | -0,377492421 | 0,831438498 | -0,454023264 | 0,649812082 | 0,999171269 | YES |
| 75 | 118,9162928 | -0,257014271 | 0,568622636 | -0,451994442 | 0,651272987 | 0,999171269 | YES |
| 76 | 39,39869301 | -0,37004877 | 0,830165947 | -0,44575277 | 0,655775842 | 0,999171269 | YES |
| 77 | 177,8807931 | 0,275428786 | 0,628840538 | 0,437994642 | 0,661390168 | 0,999171269 | YES |
| 78 | 302,2008219 | 0,29770689 | 0,700555278 | 0,424958457 | 0,67086696 | 0,999171269 | YES |
| 79 | 324,1512848 | 0,350959563 | 0,826558802 | 0,424603262 | 0,671125915 | 0,999171269 | YES |
| 80 | 3,625895907 | -0,348509087 | 0,834321334 | -0,41771566 | 0,676155021 | 0,999171269 | YES |
| 81 | 8,684758312 | 0,345307019 | 0,832136619 | 0,414964335 | 0,678168012 | 0,999171269 | YES |
| 82 | 61,12933745 | 0,28042052 | 0,676685398 | 0,41440309 | 0,678578926 | 0,999171269 | YES |
| 83 | 11,63613081 | 0,333983845 | 0,830756824 | 0,402023595 | 0,687666662 | 0,999171269 | YES |
| 84 | 51,36103563 | -0,271358143 | 0,685698235 | -0,395739889 | 0,692296925 | 0,999171269 | YES |
| 85 | 0,848613936 | -0,327424877 | 0,840044969 | -0,389770655 | 0,696706145 | 0,999171269 | YES |
| 86 | 12,78580628 | 0,267439648 | 0,689575968 | 0,387832031 | 0,69814034 | 0,999171269 | YES |
| 87 | 1313,509776 | 0,295044289 | 0,780868224 | 0,377841331 | 0,705548468 | 0,999171269 | YES |
| 88 | 26,27030935 | -0,222053856 | 0,595478991 | -0,372899563 | 0,709223197 | 0,999171269 | YES |
| 89 | 6,794697969 | 0,306136802 | 0,830860862 | 0,368457363 | 0,712532232 | 0,999171269 | YES |

| Medication | Ticker | ASVnr | taxLevel | Phylum | Family | Genus | Species |
|---|---|---|---|---|---|---|---|
| Pramipexole | 90 | ASV357 | ASV | Firmicutes | Lachnospiraceae | Marvinbryantia | NA |
| Pramipexole | 91 | ASV128 | ASV | Firmicutes | Oscillospiraceae | NK4A214 group | NA |
| Pramipexole | 92 | ASV147 | ASV | Firmicutes | Oscillospiraceae | Oscillibacter | NA |
| Pramipexole | 93 | ASV533 | ASV | Firmicutes | Ruminococcaceae | Negativibacillus | massiliensis |
| Pramipexole | 94 | ASV135 | ASV | Bacteroidota | Rikenellaceae | Alistipes | inops |
| Pramipexole | 95 | ASV656 | ASV | Firmicutes | Oscillospiraceae | Flavonifractor | NA |
| Pramipexole | 96 | ASV477 | ASV | Firmicutes | Oscillospiraceae | Intestinimonas | massiliensis |
| Pramipexole | 97 | ASV309 | ASV | Firmicutes | Oscillospiraceae | Colidextribacter | massiliensis |
| Pramipexole | 98 | ASV127 | ASV | Firmicutes | Lachnospiraceae | Lachnospiraceae NK4A136 group | bacterium |
| Pramipexole | 99 | ASV427 | ASV | Firmicutes | Lactobacillaceae | Lactobacillus | NA |
| Pramipexole | 100 | ASV34 | ASV | Firmicutes | Lachnospiraceae | Anaerostipes | hadrus |
| Pramipexole | 101 | ASV91 | ASV | Firmicutes | Lachnospiraceae | [Eubacterium] eligens group | NA |
| Pramipexole | 102 | ASV339 | ASV | Firmicutes | Monoglobaceae | Monoglobus | pectinilyticus |
| Pramipexole | 103 | ASV730 | ASV | Firmicutes | Ruminococcaceae | NA | NA |
| Pramipexole | 104 | ASV522 | ASV | Firmicutes | UCG-010 | NA | NA |
| Pramipexole | 105 | ASV66 | ASV | Firmicutes | Butyricicoccaceae | Butyricicoccus | NA |
| Pramipexole | 106 | ASV189 | ASV | Firmicutes | Ruminococcaceae | Ruminococcus | champanellensis |
| Pramipexole | 107 | ASV255 | ASV | Actinobacteriota | Coriobacteriales Incertae Sedis | NA | NA |
| Pramipexole | 108 | ASV615 | ASV | Firmicutes | Oscillospiraceae | Oscillibacter | NA |
| Pramipexole | 109 | ASV198 | ASV | Bacteroidota | Bacteroidaceae | Bacteroides | eggerthii |
| Pramipexole | 110 | ASV230 | ASV | Firmicutes | Ruminococcaceae | [Eubacterium] siraeum group | NA |
| Pramipexole | 111 | ASV107 | ASV | Bacteroidota | Bacteroidaceae | Bacteroides | eggerthii |
| Pramipexole | 112 | ASV443 | ASV | Firmicutes | Veillonellaceae | Veillonella | NA |
| Pramipexole | 113 | ASV1158 | ASV | Firmicutes | Ruminococcaceae | NA | NA |
| Pramipexole | 114 | ASV687 | ASV | Firmicutes | Ruminococcaceae | DTU089 | NA |
| Pramipexole | 115 | ASV291 | ASV | Firmicutes | Erysipelatoclostridiaceae | Erysipelatoclostridium | NA |
| Pramipexole | 116 | ASV406 | ASV | Firmicutes | NA | NA | NA |
| Pramipexole | 117 | ASV362 | ASV | Firmicutes | Ruminococcaceae | NA | NA |
| Pramipexole | 118 | ASV249 | ASV | Proteobacteria | NA | NA | NA |
| Pramipexole | 119 | ASV666 | ASV | Firmicutes | Erysipelotrichaceae | Holdemania | filiformis |

| Ticker | DESeq2_baseMean | DESeq2_log2-FoldChange | DESeq2_lfcSE | DESeq2_stat | DESeq2_pvalue | DESeq2_padj | DESeq2 converged |
|---|---|---|---|---|---|---|---|
| 90 | 36,87430144 | 0,302422691 | 0,826830453 | 0,365761433 | 0,7145431 | 0,999171269 | YES |
| 91 | 43,08507005 | 0,301038707 | 0,830135986 | 0,362637823 | 0,716875452 | 0,999171269 | YES |
| 92 | 226,320627 | 0,18537333 | 0,516001703 | 0,359249455 | 0,719408483 | 0,999171269 | YES |
| 93 | 30,8501947 | -0,295203071 | 0,827736893 | -0,356638775 | 0,721362246 | 0,999171269 | YES |
| 94 | 236,5477575 | 0,288102418 | 0,809946921 | 0,355705307 | 0,72206127 | 0,999171269 | YES |
| 95 | 5,603909884 | -0,291784432 | 0,828295638 | -0,352270878 | 0,724635126 | 0,999171269 | YES |
| 96 | 5,74610761 | 0,293220038 | 0,833129549 | 0,351950112 | 0,724875676 | 0,999171269 | YES |
| 97 | 17,18629522 | -0,282832068 | 0,811038854 | -0,348728136 | 0,72729342 | 0,999171269 | YES |
| 98 | 89,10619009 | -0,240336951 | 0,695825256 | -0,345398429 | 0,729794858 | 0,999171269 | YES |
| 99 | 1,242401646 | 0,285795433 | 0,836073656 | 0,341830449 | 0,732478493 | 0,999171269 | YES |
| 100 | 167,2686241 | -0,172447831 | 0,515864352 | -0,334289101 | 0,738161414 | 0,999171269 | YES |
| 101 | 70,83354964 | -0,261555571 | 0,806779141 | -0,32419724 | 0,745788711 | 0,999171269 | YES |
| 102 | 52,2071447 | 0,261847291 | 0,810141162 | 0,323211932 | 0,746534745 | 0,999171269 | YES |
| 103 | 3,942889868 | 0,2719427 | 0,848432731 | 0,320523584 | 0,748571456 | 0,999171269 | YES |
| 104 | 4,763700512 | 0,253551581 | 0,833583361 | 0,304170636 | 0,760997895 | 0,999171269 | YES |
| 105 | 132,1223433 | 0,244518218 | 0,808936912 | 0,30227106 | 0,762445438 | 0,999171269 | YES |
| 106 | 62,3688652 | -0,249438825 | 0,83235932 | -0,299676857 | 0,764423653 | 0,999171269 | YES |
| 107 | 22,99427955 | 0,247978174 | 0,83249176 | 0,297874629 | 0,765798853 | 0,999171269 | YES |
| 108 | 7,40906732 | -0,247745175 | 0,832906708 | -0,297446488 | 0,766125657 | 0,999171269 | YES |
| 109 | 223,3965113 | -0,240733411 | 0,83230799 | -0,289235973 | 0,772400805 | 0,999171269 | YES |
| 110 | 20,52132053 | -0,238508807 | 0,824616826 | -0,289235921 | 0,772400845 | 0,999171269 | YES |
| 111 | 194,604676 | -0,239932849 | 0,832311342 | -0,288272953 | 0,773137809 | 0,999171269 | YES |
| 112 | 0,986165204 | -0,241306241 | 0,83742776 | -0,288151709 | 0,773230612 | 0,999171269 | YES |
| 113 | 3,999290986 | 0,23686486 | 0,833291934 | 0,284251953 | 0,776217304 | 0,999171269 | YES |
| 114 | 20,82580503 | 0,233907228 | 0,832477617 | 0,280977198 | 0,778727889 | 0,999171269 | YES |
| 115 | 18,36670979 | 0,187007635 | 0,668297537 | 0,27982691 | 0,779610305 | 0,999171269 | YES |
| 116 | 10,63970657 | 0,169195544 | 0,623047248 | 0,271561337 | 0,785959331 | 0,999171269 | YES |
| 117 | 28,99652836 | 0,161388396 | 0,597938884 | 0,269907845 | 0,787231151 | 0,999171269 | YES |
| 118 | 56,81768963 | 0,223966456 | 0,844727165 | 0,265134668 | 0,790905721 | 0,999171269 | YES |
| 119 | 8,314560698 | 0,161604725 | 0,621066209 | 0,260205308 | 0,79470541 | 0,999171269 | YES |

| Medication | Ticker | ASVnr | taxLevel | Phylum | Family | Genus | Species |
|---|---|---|---|---|---|---|---|
| Pramipexole | 120 | ASV206 | ASV | Firmicutes | Oscillospiraceae | Intestinimonas | NA |
| Pramipexole | 121 | ASV24 | ASV | Bacteroidota | Bacteroidaceae | Bacteroides | uniformis |
| Pramipexole | 122 | ASV36 | ASV | Firmicutes | Lachnospiraceae | Blautia | obeum |
| Pramipexole | 123 | ASV171 | ASV | Firmicutes | Oscillospiraceae | NA | NA |
| Pramipexole | 124 | ASV234 | ASV | Cyanobacteria | NA | NA | NA |
| Pramipexole | 125 | ASV64 | ASV | Firmicutes | Oscillospiraceae | UCG-002 | NA |
| Pramipexole | 126 | ASV192 | ASV | Bacteroidota | Marinifilaceae | Odoribacter | splanchnicus |
| Pramipexole | 127 | ASV158 | ASV | Firmicutes | Lachnospiraceae | [Eubacterium] ventriosum group | NA |
| Pramipexole | 128 | ASV94 | ASV | Bacteroidota | Rikenellaceae | Alistipes | shahii |
| Pramipexole | 129 | ASV96 | ASV | Firmicutes | Lachnospiraceae | CAG-56 | NA |
| Pramipexole | 130 | ASV123 | ASV | Firmicutes | Lachnospiraceae | [Ruminococcus] torques group | NA |
| Pramipexole | 131 | ASV305 | ASV | Firmicutes | Lachnospiraceae | Lachnospiraceae UCG-004 | NA |
| Pramipexole | 132 | ASV159 | ASV | Firmicutes | Lachnospiraceae | Roseburia | NA |
| Pramipexole | 133 | ASV39 | ASV | Bacteroidota | Tannerellaceae | Parabacteroides | merdae |
| Pramipexole | 134 | ASV211 | ASV | Firmicutes | Lachnospiraceae | Lachnospiraceae FCS020 group | NA |
| Pramipexole | 135 | ASV84 | ASV | Bacteroidota | Bacteroidaceae | Bacteroides | NA |
| Pramipexole | 136 | ASV415 | ASV | Firmicutes | Oscillospiraceae | Oscillospira | NA |
| Pramipexole | 137 | ASV813 | ASV | Firmicutes | Ruminococcaceae | NA | NA |
| Pramipexole | 138 | ASV219 | ASV | Firmicutes | Lachnospiraceae | Lachnospiraceae UCG-004 | NA |
| Pramipexole | 139 | ASV19 | ASV | Firmicutes | Erysipelatoclostridiaceae | Erysipelotrichaceae UCG-003 | bacterium |
| Pramipexole | 140 | ASV324 | ASV | Proteobacteria | NA | NA | NA |
| Pramipexole | 141 | ASV257 | ASV | Bacteroidota | Bacteroidaceae | Bacteroides | ovatus |
| Pramipexole | 142 | ASV808 | ASV | Desulfobacterota | Desulfovibrionaceae | NA | NA |
| Pramipexole | 143 | ASV56 | ASV | Bacteroidota | Bacteroidaceae | Bacteroides | massiliensis |
| Pramipexole | 144 | ASV31 | ASV | Bacteroidota | Rikenellaceae | Alistipes | putredinis |
| Pramipexole | 145 | ASV306 | ASV | Firmicutes | Veillonellaceae | Veillonella | NA |
| Pramipexole | 146 | ASV226 | ASV | Firmicutes | Lachnospiraceae | Lachnospiraceae UCG-001 | NA |
| Pramipexole | 147 | ASV22 | ASV | Bacteroidota | Bacteroidaceae | Bacteroides | uniformis |
| Pramipexole | 148 | ASV93 | ASV | Firmicutes | Oscillospiraceae | NK4A214 group | NA |
| Pramipexole | 149 | ASV296 | ASV | Proteobacteria | Sutterellaceae | Sutterella | NA |

| Ticker | DESeq2_baseMean | DESeq2_log2-FoldChange | DESeq2_lfcSE | DESeq2_stat | DESeq2_pvalue | DESeq2_padj | DESeq2 converged |
|---|---|---|---|---|---|---|---|
| 120 | 69,02539648 | 0,210043058 | 0,810061279 | 0,259292801 | 0,795409334 | 0,999171269 | YES |
| 121 | 167,3961647 | -0,168016708 | 0,64867127 | -0,259016725 | 0,795622336 | 0,999171269 | YES |
| 122 | 154,2226588 | 0,177720816 | 0,69709074 | 0,25494646 | 0,798764452 | 0,999171269 | YES |
| 123 | 65,60773752 | 0,181665261 | 0,714214909 | 0,254356579 | 0,799220093 | 0,999171269 | YES |
| 124 | 2,183770884 | -0,209591977 | 0,832360133 | -0,25180444 | 0,801192225 | 0,999171269 | YES |
| 125 | 82,02825286 | 0,204183353 | 0,829787155 | 0,246067141 | 0,805630259 | 0,999171269 | YES |
| 126 | 22,98590963 | -0,128160036 | 0,531780544 | -0,241001739 | 0,809553769 | 0,999171269 | YES |
| 127 | 65,37327323 | 0,196353245 | 0,829792114 | 0,236629442 | 0,812944278 | 0,999171269 | YES |
| 128 | 65,95549915 | -0,167628843 | 0,709259301 | -0,236343525 | 0,813166115 | 0,999171269 | YES |
| 129 | 97,32724809 | -0,196078009 | 0,829769404 | -0,236304217 | 0,813196615 | 0,999171269 | YES |
| 130 | 31,3318211 | 0,195121961 | 0,829913043 | 0,235111332 | 0,814122324 | 0,999171269 | YES |
| 131 | 15,4038418 | 0,194650987 | 0,830160824 | 0,234473829 | 0,814617149 | 0,999171269 | YES |
| 132 | 63,25682592 | 0,168333551 | 0,723256132 | 0,232744036 | 0,815960172 | 0,999171269 | YES |
| 133 | 401,239006 | -0,137632047 | 0,618815932 | -0,222411932 | 0,823993224 | 0,999171269 | YES |
| 134 | 35,15341408 | 0,137827977 | 0,632655142 | 0,217856409 | 0,827540992 | 0,999171269 | YES |
| 135 | 55,29048009 | -0,175825781 | 0,810408216 | -0,216959525 | 0,828239888 | 0,999171269 | YES |
| 136 | 52,05245337 | -0,110243945 | 0,508219893 | -0,216921744 | 0,828269332 | 0,999171269 | YES |
| 137 | 1,306896808 | 0,18097086 | 0,85107969 | 0,212636798 | 0,831610267 | 0,999171269 | YES |
| 138 | 41,25216796 | -0,175756571 | 0,82990101 | -0,211780163 | 0,832278545 | 0,999171269 | YES |
| 139 | 227,8322627 | -0,129062832 | 0,61649131 | -0,20935061 | 0,834174546 | 0,999171269 | YES |
| 140 | 0,560098632 | 0,177453424 | 0,851953043 | 0,208290147 | 0,835002425 | 0,999171269 | YES |
| 141 | 25,44965612 | -0,172515368 | 0,832455506 | -0,207236744 | 0,835824973 | 0,999171269 | YES |
| 142 | 0,46674886 | 0,176286247 | 0,852246373 | 0,206848926 | 0,836127845 | 0,999171269 | YES |
| 143 | 207,8403903 | -0,165825598 | 0,832305461 | -0,199236466 | 0,842077776 | 0,999171269 | YES |
| 144 | 290,3412161 | 0,127216497 | 0,661355503 | 0,192357206 | 0,847462411 | 0,999171269 | YES |
| 145 | 14,6533267 | -0,155854487 | 0,831675426 | -0,187398211 | 0,851348425 | 0,999171269 | YES |
| 146 | 11,8970432 | -0,155453689 | 0,830243661 | -0,187238634 | 0,851473535 | 0,999171269 | YES |
| 147 | 542,3847711 | -0,103369331 | 0,554393668 | -0,186454746 | 0,852088164 | 0,999171269 | YES |
| 148 | 42,16748708 | 0,150848059 | 0,829870131 | 0,181773091 | 0,855760803 | 0,999171269 | YES |
| 149 | 21,27744894 | 0,151260318 | 0,83244179 | 0,18170678 | 0,855812845 | 0,999171269 | YES |

| Medication | Ticker | ASVnr | taxLevel | Phylum | Family | Genus | Species |
|---|---|---|---|---|---|---|---|
| Pramipexole | 150 | ASV85 | ASV | Firmicutes | Lachnospiraceae | Coprococcus | NA |
| Pramipexole | 151 | ASV41 | ASV | Firmicutes | Ruminococcaceae | Ruminococcus | bicirculans |
| Pramipexole | 152 | ASV71 | ASV | Firmicutes | [Eubacterium] coprostanoligenes group | NA | NA |
| Pramipexole | 153 | ASV2 | ASV | Bacteroidota | Bacteroidaceae | Bacteroides | vulgatus |
| Pramipexole | 154 | ASV59 | ASV | Firmicutes | [Eubacterium] coprostanoligenes group | NA | NA |
| Pramipexole | 155 | ASV43 | ASV | Firmicutes | Lachnospiraceae | Blautia | faecis |
| Pramipexole | 156 | ASV307 | ASV | Firmicutes | Lachnospiraceae | Lachnoclostridium | NA |
| Pramipexole | 157 | ASV836 | ASV | Verrucomicrobiota | Victivallaceae | Victivallis | vadensis |
| Pramipexole | 158 | ASV78 | ASV | Firmicutes | Lachnospiraceae | [Eubacterium] eligens group | NA |
| Pramipexole | 159 | ASV33 | ASV | Firmicutes | Peptostreptococcaceae | Intestinibacter | bartlettii |
| Pramipexole | 160 | ASV9 | ASV | Bacteroidota | Bacteroidaceae | Bacteroides | NA |
| Pramipexole | 161 | ASV73 | ASV | Firmicutes | Oscillospiraceae | UCG-005 | NA |
| Pramipexole | 162 | ASV80 | ASV | Firmicutes | Acidaminococcaceae | Phascolarctobacterium | faecium |
| Pramipexole | 163 | ASV573 | ASV | Firmicutes | Erysipelatoclostridiaceae | Erysipelatoclostridium | NA |
| Pramipexole | 164 | ASV74 | ASV | Firmicutes | Lachnospiraceae | Coprococcus | catus |
| Pramipexole | 165 | ASV191 | ASV | Firmicutes | Lachnospiraceae | Lachnospiraceae NK4A136 group | NA |
| Pramipexole | 166 | ASV45 | ASV | Firmicutes | Lachnospiraceae | Coprococcus | comes |
| Pramipexole | 167 | ASV214 | ASV | Firmicutes | Lachnospiraceae | Lachnospiraceae UCG-010 | NA |
| Pramipexole | 168 | ASV117 | ASV | Bacteroidota | Rikenellaceae | Alistipes | finegoldii |
| Pramipexole | 169 | ASV446 | ASV | Firmicutes | Oscillospiraceae | UCG-005 | NA |
| Pramipexole | 170 | ASV444 | ASV | Firmicutes | Erysipelatoclostridiaceae | Coprobacillus | cateniformis |
| Pramipexole | 171 | ASV534 | ASV | Firmicutes | Anaerovoracaceae | Family XIII UCG-001 | NA |
| Pramipexole | 172 | ASV367 | ASV | Bacteroidota | Rikenellaceae | Alistipes | NA |
| Pramipexole | 173 | ASV89 | ASV | Firmicutes | Lachnospiraceae | Blautia | NA |
| Pramipexole | 174 | ASV50 | ASV | Bacteroidota | Prevotellaceae | Prevotella | NA |
| Pramipexole | 175 | ASV709 | ASV | Firmicutes | Lachnospiraceae | NA | NA |
| Pramipexole | 176 | ASV376 | ASV | Bacteroidota | Barnesiellaceae | Coprobacter | fastidiosus |
| Pramipexole | 177 | ASV72 | ASV | Proteobacteria | Sutterellaceae | Parasutterella | excrementihominis |
| Pramipexole | 178 | ASV134 | ASV | Actinobacteriota | Eggerthellaceae | Enterorhabdus | NA |
| Pramipexole | 179 | ASV535 | ASV | Firmicutes | Erysipelatoclostridiaceae | Asteroleplasma | NA |

| Ticker | DESeq2_baseMean | DESeq2_log2-FoldChange | DESeq2_lfcSE | DESeq2_stat | DESeq2_pvalue | DESeq2_padj | DESeq2 converged |
|---|---|---|---|---|---|---|---|
| 150 | 507,9557155 | 0,150509725 | 0,829711107 | 0,181400157 | 0,856053496 | 0,999171269 | YES |
| 151 | 224,0627518 | 0,12980584 | 0,723752428 | 0,179351163 | 0,857661976 | 0,999171269 | YES |
| 152 | 12,14282493 | -0,148542851 | 0,830196632 | -0,178924902 | 0,857996669 | 0,999171269 | YES |
| 153 | 1305,812412 | 0,143749036 | 0,847632184 | 0,169588931 | 0,865333428 | 0,999171269 | YES |
| 154 | 59,57505941 | 0,141118709 | 0,832339979 | 0,169544553 | 0,865368332 | 0,999171269 | YES |
| 155 | 268,157392 | 0,072610684 | 0,435681837 | 0,166659884 | 0,867637672 | 0,999171269 | YES |
| 156 | 18,22978649 | 0,137050387 | 0,826948459 | 0,165730265 | 0,868369228 | 0,999171269 | YES |
| 157 | 0,784639242 | -0,140676394 | 0,850172034 | -0,16546815 | 0,868575517 | 0,999171269 | YES |
| 158 | 140,623728 | -0,113169806 | 0,692542848 | -0,163411991 | 0,870194062 | 0,999171269 | YES |
| 159 | 42,4840573 | -0,099725405 | 0,617252466 | -0,161563397 | 0,871649684 | 0,999171269 | YES |
| 160 | 1524,367925 | 0,094750421 | 0,603285892 | 0,157057246 | 0,875199732 | 0,999171269 | YES |
| 161 | 153,5235801 | -0,127081783 | 0,829743236 | -0,153157962 | 0,878273708 | 0,999171269 | YES |
| 162 | 6,109153174 | 0,128395009 | 0,850524838 | 0,150959741 | 0,880007474 | 0,999171269 | YES |
| 163 | 58,45840405 | 0,103401329 | 0,685111917 | 0,150926186 | 0,880033943 | 0,999171269 | YES |
| 164 | 15,47441742 | -0,125174708 | 0,83013207 | -0,150788907 | 0,880142237 | 0,999171269 | YES |
| 165 | 23,57099048 | 0,125278712 | 0,853881337 | 0,146716771 | 0,883355581 | 0,999171269 | YES |
| 166 | 48,39972217 | -0,121691333 | 0,829810123 | -0,146649612 | 0,883408592 | 0,999171269 | YES |
| 167 | 9,483372262 | 0,125202704 | 0,853896795 | 0,146625101 | 0,88342794 | 0,999171269 | YES |
| 168 | 7,009449063 | 0,125157609 | 0,85390591 | 0,146570725 | 0,883470862 | 0,999171269 | YES |
| 169 | 4,604245953 | 0,125067636 | 0,853924129 | 0,146462234 | 0,883556501 | 0,999171269 | YES |
| 170 | 2,817523643 | 0,124901447 | 0,853957689 | 0,146261868 | 0,883714667 | 0,999171269 | YES |
| 171 | 2,680083465 | 0,124879545 | 0,853962113 | 0,146235463 | 0,883735511 | 0,999171269 | YES |
| 172 | 1,305681688 | 0,12440977 | 0,854056973 | 0,145669169 | 0,884182561 | 0,999171269 | YES |
| 173 | 1,099521422 | 0,124239455 | 0,854091369 | 0,145463892 | 0,884344622 | 0,999171269 | YES |
| 174 | 1,030801333 | 0,124167845 | 0,854105846 | 0,145377584 | 0,884412762 | 0,999171269 | YES |
| 175 | 1,030801333 | 0,124167845 | 0,854105846 | 0,145377584 | 0,884412762 | 0,999171269 | YES |
| 176 | 0,824641066 | 0,123882815 | 0,854163485 | 0,14503408 | 0,884683964 | 0,999171269 | YES |
| 177 | 162,2978453 | -0,091421497 | 0,631818905 | -0,14469573 | 0,884951111 | 0,999171269 | YES |
| 178 | 10,07662933 | 0,103265062 | 0,714040718 | 0,144620691 | 0,88501036 | 0,999171269 | YES |
| 179 | 0,549760711 | 0,123180221 | 0,854305727 | 0,144187516 | 0,885352398 | 0,999171269 | YES |

| Medication | Ticker | ASVnr | taxLevel | Phylum | Family | Genus | Species |
|---|---|---|---|---|---|---|---|
| Pramipexole | 180 | ASV133 | ASV | Actinobacteriota | Eggerthellaceae | Senegalimassilia | anaerobia |
| Pramipexole | 181 | ASV146 | ASV | Firmicutes | Erysipelotrichaceae | Turicibacter | sanguinis |
| Pramipexole | 182 | ASV32 | ASV | Firmicutes | Oscillospiraceae | UCG-002 | NA |
| Pramipexole | 183 | ASV261 | ASV | Firmicutes | Anaerovoracaceae | Family XIII UCG-001 | NA |
| Pramipexole | 184 | ASV8 | ASV | Firmicutes | Ruminococcaceae | Faecalibacterium | prausnitzii |
| Pramipexole | 185 | ASV79 | ASV | Firmicutes | Lachnospiraceae | [Eubacterium] ruminantium group | NA |
| Pramipexole | 186 | ASV195 | ASV | Bacteroidota | Barnesiellaceae | Barnesiella | intestinihominis |
| Pramipexole | 187 | ASV18 | ASV | Firmicutes | Lachnospiraceae | Dorea | longicatena |
| Pramipexole | 188 | ASV37 | ASV | Firmicutes | Lachnospiraceae | [Eubacterium] hallii group | NA |
| Pramipexole | 189 | ASV885 | ASV | Firmicutes | Lachnospiraceae | Shuttleworthia | NA |
| Pramipexole | 190 | ASV355 | ASV | Firmicutes | NA | NA | NA |
| Pramipexole | 191 | ASV288 | ASV | Firmicutes | Erysipelotrichaceae | NA | NA |
| Pramipexole | 192 | ASV65 | ASV | Firmicutes | Ruminococcaceae | Incertae Sedis | NA |
| Pramipexole | 193 | ASV155 | ASV | Firmicutes | Ruminococcaceae | Ruminococcus | NA |
| Pramipexole | 194 | ASV153 | ASV | Firmicutes | Streptococcaceae | Streptococcus | NA |
| Pramipexole | 195 | ASV294 | ASV | Firmicutes | Anaerovoracaceae | Mogibacterium | NA |
| Pramipexole | 196 | ASV232 | ASV | Firmicutes | Acholeplasmataceae | Anaeroplasma | NA |
| Pramipexole | 197 | ASV335 | ASV | Bacteroidota | Barnesiellaceae | NA | NA |
| Pramipexole | 198 | ASV393 | ASV | Bacteroidota | Rikenellaceae | Alistipes | indistinctus |
| Pramipexole | 199 | ASV314 | ASV | Firmicutes | Ruminococcaceae | Subdoligranulum | NA |
| Pramipexole | 200 | ASV40 | ASV | Firmicutes | Erysipelotrichaceae | Holdemanella | NA |
| Pramipexole | 201 | ASV819 | ASV | Firmicutes | Oscillospiraceae | Colidextribacter | NA |
| Pramipexole | 202 | ASV422 | ASV | Firmicutes | Lachnospiraceae | Lachnospiraceae FCS020 group | bacterium |
| Pramipexole | 203 | ASV545 | ASV | Proteobacteria | Sutterellaceae | Sutterella | NA |
| Pramipexole | 204 | ASV272 | ASV | Firmicutes | Erysipelotrichaceae | Catenisphaera | NA |
| Pramipexole | 205 | ASV106 | ASV | Bacteroidota | Bacteroidaceae | Bacteroides | NA |
| Pramipexole | 206 | ASV337 | ASV | Firmicutes | Lachnospiraceae | Lachnoclostridium | NA |
| Pramipexole | 207 | ASV454 | ASV | Actinobacteriota | Actinomycetaceae | Actinomyces | odontolyticus |
| Pramipexole | 208 | ASV247 | ASV | Euryarchaeota | Methanobacteriaceae | Methanosphaera | NA |
| Pramipexole | 209 | ASV104 | ASV | Firmicutes | Christensenellaceae | Christensenellaceae R-7 group | NA |

| Ticker | DESeq2_baseMean | DESeq2_log2-FoldChange | DESeq2_lfcSE | DESeq2_stat | DESeq2_pvalue | DESeq2_padj | DESeq2 converged |
|---|---|---|---|---|---|---|---|
| 180 | 0,412320533 | 0,122491812 | 0,854445385 | 0,143358269 | 0,886007239 | 0,999171269 | YES |
| 181 | 0,274880355 | 0,121155541 | 0,85471726 | 0,141749262 | 0,887278062 | 0,999171269 | YES |
| 182 | 90,69184796 | 0,113450377 | 0,831738282 | 0,136401533 | 0,891503862 | 0,999171269 | YES |
| 183 | 17,45796301 | 0,092767131 | 0,688361678 | 0,1347651 | 0,8927976 | 0,999171269 | YES |
| 184 | 462,9451701 | 0,099947243 | 0,743215291 | 0,134479531 | 0,893023396 | 0,999171269 | YES |
| 185 | 18,19762785 | 0,114083541 | 0,853884223 | 0,133605397 | 0,893714616 | 0,999171269 | YES |
| 186 | 202,7115034 | 0,109805844 | 0,84764033 | 0,129542967 | 0,896928028 | 0,999171269 | YES |
| 187 | 512,566191 | -0,078974773 | 0,612988948 | -0,128835558 | 0,897487769 | 0,999171269 | YES |
| 188 | 75,61948249 | 0,094660709 | 0,744230084 | 0,1271928 | 0,898787803 | 0,999171269 | YES |
| 189 | 9,167173499 | -0,102271658 | 0,827326285 | -0,123617078 | 0,90161847 | 0,999171269 | YES |
| 190 | 5,733651328 | 0,104962949 | 0,850529551 | 0,123408939 | 0,901783279 | 0,999171269 | YES |
| 191 | 8,386122009 | 0,084039076 | 0,699971974 | 0,12006063 | 0,904435119 | 0,999171269 | YES |
| 192 | 14,33377851 | 0,095264396 | 0,853887467 | 0,111565516 | 0,911167915 | 0,999171269 | YES |
| 193 | 45,98798335 | -0,090525641 | 0,829823963 | -0,109090174 | 0,913130968 | 0,999171269 | YES |
| 194 | 2,125665448 | 0,092331774 | 0,853981624 | 0,108119158 | 0,913901171 | 0,999171269 | YES |
| 195 | 8,982149978 | 0,091767004 | 0,853897228 | 0,107468441 | 0,91441736 | 0,999171269 | YES |
| 196 | 75,53470726 | 0,087123105 | 0,853874033 | 0,102032737 | 0,91873069 | 0,999171269 | YES |
| 197 | 2,947201643 | 0,086182579 | 0,853950469 | 0,100922222 | 0,919612203 | 0,999171269 | YES |
| 198 | 21,25255595 | -0,069561494 | 0,717833126 | -0,096904825 | 0,922801977 | 0,999171269 | YES |
| 199 | 13,63571058 | 0,082560357 | 0,853888118 | 0,096687559 | 0,92297452 | 0,999171269 | YES |
| 200 | 74,37586975 | -0,081726261 | 0,847667303 | -0,096413133 | 0,923192462 | 0,999171269 | YES |
| 201 | 8,789093928 | 0,081716165 | 0,84783921 | 0,096381677 | 0,923217444 | 0,999171269 | YES |
| 202 | 8,430134655 | -0,069804904 | 0,832694872 | -0,083830112 | 0,933191507 | 0,999171269 | YES |
| 203 | 51,7567803 | 0,070512824 | 0,853875409 | 0,082579757 | 0,934185698 | 0,999171269 | YES |
| 204 | 11,91508582 | 0,066119919 | 0,832536971 | 0,0794198 | 0,936698721 | 0,999171269 | YES |
| 205 | 79,02373633 | -0,058112914 | 0,736083409 | -0,078948816 | 0,937073335 | 0,999171269 | YES |
| 206 | 38,6579463 | -0,056582319 | 0,717256118 | -0,078887188 | 0,937122354 | 0,999171269 | YES |
| 207 | 1,024744735 | 0,065289661 | 0,85114492 | 0,076708043 | 0,938855806 | 0,999171269 | YES |
| 208 | 6,36457686 | 0,06549221 | 0,853907298 | 0,076697096 | 0,938864515 | 0,999171269 | YES |
| 209 | 10,88069728 | 0,064664985 | 0,853892205 | 0,075729682 | 0,939634161 | 0,999171269 | YES |

| Medication | Ticker | ASVnr | taxLevel | Phylum | Family | Genus | Species |
|---|---|---|---|---|---|---|---|
| Pramipexole | 210 | ASV231 | ASV | Proteobacteria | NA | NA | NA |
| Pramipexole | 211 | ASV521 | ASV | Firmicutes | Lachnospiraceae | [Eubacterium] ventriosum group | NA |
| Pramipexole | 212 | ASV481 | ASV | Firmicutes | Lachnospiraceae | [Eubacterium] xylanophilum group | NA |
| Pramipexole | 213 | ASV507 | ASV | Firmicutes | Oscillospiraceae | Colidextribacter | NA |
| Pramipexole | 214 | ASV703 | ASV | Firmicutes | Hydrogenoanaerobacterium | NA | NA |
| Pramipexole | 215 | ASV250 | ASV | Bacteroidota | Prevotellaceae | Paraprevotella | NA |
| Pramipexole | 216 | ASV20 | ASV | Bacteroidota | Prevotellaceae | Prevotella | copri |
| Pramipexole | 217 | ASV38 | ASV | Firmicutes | Clostridiaceae | Clostridium sensu stricto 1 | NA |
| Pramipexole | 218 | ASV253 | ASV | Bacteroidota | Tannerellaceae | Parabacteroides | distasonis |
| Pramipexole | 219 | ASV116 | ASV | Firmicutes | Oscillospiraceae | UCG-003 | NA |
| Pramipexole | 220 | ASV608 | ASV | Firmicutes | Lachnospiraceae | NA | NA |
| Pramipexole | 221 | ASV161 | ASV | Firmicutes | Oscillospiraceae | UCG-002 | NA |
| Pramipexole | 222 | ASV55 | ASV | Firmicutes | Monoglobaceae | Monoglobus | NA |
| Pramipexole | 223 | ASV29 | ASV | Firmicutes | Oscillospiraceae | UCG-002 | NA |
| Pramipexole | 224 | ASV120 | ASV | Firmicutes | Lachnospiraceae | Anaerostipes | hadrus |
| Pramipexole | 225 | ASV144 | ASV | Desulfobacterota | Desulfovibrionaceae | Bilophila | wadsworthia |
| Pramipexole | 226 | ASV276 | ASV | Firmicutes | Oscillospiraceae | NK4A214 group | NA |
| Pramipexole | 227 | ASV168 | ASV | Firmicutes | Lachnospiraceae | Lachnoclostridium | edouardi |
| Pramipexole | 228 | ASV186 | ASV | Firmicutes | Lachnospiraceae | [Ruminococcus] torques group | NA |
| Pramipexole | 229 | ASV322 | ASV | Firmicutes | Anaerovoracaceae | Family XIII AD3011 group | NA |
| Pramipexole | 230 | ASV142 | ASV | Firmicutes | Ruminococcaceae | Faecalibacterium | prausnitzii |
| Pramipexole | 231 | ASV109 | ASV | Firmicutes | [Eubacterium] coprostanoligenes group | NA | NA |
| Pramipexole | 232 | ASV196 | ASV | Firmicutes | Ruminococcaceae | Ruminococcus | NA |
| Pramipexole | 233 | ASV110 | ASV | Firmicutes | Ruminococcaceae | Faecalibacterium | prausnitzii |
| Pramipexole | 234 | ASV98 | ASV | Firmicutes | Acidaminococcaceae | Phascolarctobacterium | succinatutens |
| Pramipexole | 235 | ASV125 | ASV | Firmicutes | Oscillospiraceae | UCG-002 | NA |
| Pramipexole | 236 | ASV81 | ASV | Firmicutes | Lachnospiraceae | Lachnospiraceae NK4A136 group | NA |
| Pramipexole | 237 | ASV148 | ASV | Firmicutes | NA | NA | NA |
| Pramipexole | 238 | ASV222 | ASV | Firmicutes | Oscillospiraceae | NK4A214 group | NA |
| Pramipexole | 239 | ASV139 | ASV | Firmicutes | Christensenellaceae | Christensenellaceae R-7 group | NA |

| Ticker | DESeq2_baseMean | DESeq2_log2-FoldChange | DESeq2_lfcSE | DESeq2_stat | DESeq2_pvalue | DESeq2_padj | DESeq2 converged |
|---|---|---|---|---|---|---|---|
| 210 | 6,388883913 | -0,063126159 | 0,850515085 | -0,074221093 | 0,940834463 | 0,999171269 | YES |
| 211 | 4,646461028 | -0,062977036 | 0,850562832 | -0,074041604 | 0,940977281 | 0,999171269 | YES |
| 212 | 3,252522719 | -0,062743844 | 0,850637627 | -0,073760955 | 0,941200596 | 0,999171269 | YES |
| 213 | 68,95655279 | 0,062388185 | 0,850398638 | 0,073363458 | 0,941516896 | 0,999171269 | YES |
| 214 | 2,207068988 | -0,062378071 | 0,85075514 | -0,073320828 | 0,941550818 | 0,999171269 | YES |
| 215 | 53,74541723 | -0,060226511 | 0,829882427 | -0,072572342 | 0,942146437 | 0,999171269 | YES |
| 216 | 1275,5993 | 0,058298746 | 0,830046639 | 0,070235507 | 0,944006214 | 0,999171269 | YES |
| 217 | 6,150366803 | -0,055725223 | 0,827727185 | -0,067323176 | 0,946324427 | 0,999171269 | YES |
| 218 | 41,34844229 | -0,053379564 | 0,832359351 | -0,064130431 | 0,948866371 | 0,999171269 | YES |
| 219 | 64,248212 | -0,053643222 | 0,847665275 | -0,063283496 | 0,949540757 | 0,999171269 | YES |
| 220 | 11,96956381 | -0,05260272 | 0,832578873 | -0,063180464 | 0,949622801 | 0,999171269 | YES |
| 221 | 17,09603952 | -0,050466666 | 0,808393918 | -0,06242831 | 0,950221751 | 0,999171269 | YES |
| 222 | 132,3023306 | -0,031871674 | 0,540601636 | -0,058955933 | 0,952987208 | 0,999171269 | YES |
| 223 | 118,4631776 | 0,049951255 | 0,853872857 | 0,05849964 | 0,953350649 | 0,999171269 | YES |
| 224 | 130,5332795 | 0,04082912 | 0,712988936 | 0,057264731 | 0,954334315 | 0,999171269 | YES |
| 225 | 73,02845104 | 0,03714871 | 0,657035723 | 0,056539864 | 0,954911739 | 0,999171269 | YES |
| 226 | 211,4863622 | 0,047236109 | 0,850390891 | 0,055546349 | 0,955703206 | 0,999171269 | YES |
| 227 | 13,39462051 | 0,046558825 | 0,853888072 | 0,054525677 | 0,956516352 | 0,999171269 | YES |
| 228 | 103,5982391 | 0,045764999 | 0,85039475 | 0,053816182 | 0,957081617 | 0,999171269 | YES |
| 229 | 6,284213504 | 0,040088011 | 0,853907378 | 0,046946556 | 0,962555822 | 0,999171269 | YES |
| 230 | 13,66365008 | -0,035915449 | 0,832538544 | -0,043139684 | 0,965590186 | 0,999171269 | YES |
| 231 | 72,43561471 | 0,033813081 | 0,829843656 | 0,040746327 | 0,967498129 | 0,999171269 | YES |
| 232 | 43,0084704 | -0,032487325 | 0,85387635 | -0,038046872 | 0,96965031 | 0,999171269 | YES |
| 233 | 33,15660909 | -0,03247898 | 0,853877965 | -0,038037028 | 0,969658159 | 0,999171269 | YES |
| 234 | 30,98240522 | -0,032476404 | 0,85387846 | -0,03803399 | 0,969660582 | 0,999171269 | YES |
| 235 | 18,20895745 | -0,032449768 | 0,853883749 | -0,038002559 | 0,969685642 | 0,999171269 | YES |
| 236 | 14,8117639 | -0,032435102 | 0,853886691 | -0,037985253 | 0,96969944 | 0,999171269 | YES |
| 237 | 14,40410067 | -0,032432933 | 0,853887137 | -0,037982693 | 0,969701481 | 0,999171269 | YES |
| 238 | 11,55045809 | -0,032413102 | 0,85389114 | -0,037959291 | 0,96972014 | 0,999171269 | YES |
| 239 | 10,12363679 | -0,032399264 | 0,853893987 | -0,037942958 | 0,969733162 | 0,999171269 | YES |

| Medication | Ticker | ASVnr | taxLevel | Phylum | Family | Genus | Species |
|---|---|---|---|---|---|---|---|
| Pramipexole | 240 | ASV373 | ASV | Firmicutes | NA | NA | NA |
| Pramipexole | 241 | ASV136 | ASV | Firmicutes | Ruminococcaceae | NA | NA |
| Pramipexole | 242 | ASV114 | ASV | Bacteroidota | Bacteroidaceae | Bacteroides | coprocola |
| Pramipexole | 243 | ASV179 | ASV | Firmicutes | NA | NA | NA |
| Pramipexole | 244 | ASV433 | ASV | Firmicutes | UCG-010 | NA | NA |
| Pramipexole | 245 | ASV68 | ASV | Firmicutes | Ruminococcaceae | Faecalibacterium | NA |
| Pramipexole | 246 | ASV180 | ASV | Firmicutes | Ruminococcaceae | Ruminococcus | NA |
| Pramipexole | 247 | ASV252 | ASV | Firmicutes | Erysipelotrichaceae | Solobacterium | NA |
| Pramipexole | 248 | ASV287 | ASV | Firmicutes | Christensenellaceae | Christensenellaceae R-7 group | NA |
| Pramipexole | 249 | ASV61 | ASV | Firmicutes | Erysipelatoclostridiaceae | Catenibacterium | mitsuokai |
| Pramipexole | 250 | ASV205 | ASV | Firmicutes | NA | NA | NA |
| Pramipexole | 251 | ASV175 | ASV | Firmicutes | NA | NA | NA |
| Pramipexole | 252 | ASV759 | ASV | Firmicutes | NA | NA | NA |
| Pramipexole | 253 | ASV151 | ASV | Firmicutes | Butyricicoccaceae | Butyricicoccus | NA |
| Pramipexole | 254 | ASV530 | ASV | Firmicutes | Veillonellaceae | Allisonella | histaminiformans |
| Pramipexole | 255 | ASV118 | ASV | Firmicutes | NA | NA | NA |
| Pramipexole | 256 | ASV546 | ASV | Firmicutes | Lachnospiraceae | NA | NA |
| Pramipexole | 257 | ASV437 | ASV | Firmicutes | Christensenellaceae | Christensenellaceae R-7 group | NA |
| Pramipexole | 258 | ASV597 | ASV | Firmicutes | Oscillospiraceae | Oscillibacter | NA |
| Pramipexole | 259 | ASV753 | ASV | Firmicutes | Ruminococcaceae | NA | NA |
| Pramipexole | 260 | ASV129 | ASV | Firmicutes | Christensenellaceae | Christensenellaceae R-7 group | NA |
| Pramipexole | 261 | ASV480 | ASV | Firmicutes | UCG-010 | NA | NA |
| Pramipexole | 262 | ASV303 | ASV | Firmicutes | Lachnospiraceae | GCA-900066575 | NA |
| Pramipexole | 263 | ASV370 | ASV | Firmicutes | UCG-010 | NA | NA |
| Pramipexole | 264 | ASV254 | ASV | Firmicutes | Lachnospiraceae | Lachnospira | NA |
| Pramipexole | 265 | ASV258 | ASV | Firmicutes | Lachnospiraceae | Lachnospiraceae AC2044 group | NA |
| Pramipexole | 266 | ASV267 | ASV | Firmicutes | Lachnospiraceae | Lachnoclostridium | NA |
| Pramipexole | 267 | ASV286 | ASV | Firmicutes | Lachnospiraceae | NA | NA |
| Pramipexole | 268 | ASV167 | ASV | Firmicutes | Oscillospiraceae | Intestinimonas | NA |
| Pramipexole | 269 | ASV174 | ASV | Actinobacteriota | Coriobacteriales Incertae Sedis | NA | NA |

| Ticker | DESeq2_baseMean | DESeq2_log2-FoldChange | DESeq2_lfcSE | DESeq2_stat | DESeq2_pvalue | DESeq2_padj | DESeq2 converged |
|---|---|---|---|---|---|---|---|
| 240 | 9,851861308 | -0,032396026 | 0,853894622 | -0,037939138 | 0,969736208 | 0,999171269 | YES |
| 241 | 9,648029695 | -0,03239361 | 0,853895122 | -0,037936287 | 0,969738481 | 0,999171269 | YES |
| 242 | 9,580085824 | -0,03239278 | 0,853895294 | -0,037935307 | 0,969739262 | 0,999171269 | YES |
| 243 | 9,036534855 | -0,03238567 | 0,853896758 | -0,037926915 | 0,969745953 | 9,99E-01 | YES |
| 244 | 6,658499367 | -0,032340713 | 0,853905968 | -0,037873858 | 0,969788257 | 0,999171269 | YES |
| 245 | 6,047004527 | -0,032323581 | 0,853909505 | -0,037853637 | 0,969804379 | 0,999171269 | YES |
| 246 | 5,435509687 | -0,032302496 | 0,853913835 | -0,037828754 | 0,969824218 | 0,999171269 | YES |
| 247 | 5,299621945 | -0,032297192 | 0,853914932 | -0,037822494 | 0,96982921 | 0,999171269 | YES |
| 248 | 4,552239363 | -0,032262208 | 0,853922137 | -0,037781206 | 0,969862129 | 0,999171269 | YES |
| 249 | 4,280463879 | -0,03224658 | 0,853925378 | -0,037762761 | 0,969876836 | 0,999171269 | YES |
| 250 | 3,940744523 | -0,032223996 | 0,853930056 | -0,037736107 | 0,969898087 | 0,999171269 | YES |
| 251 | 3,872800652 | -0,032219005 | 0,85393109 | -0,037730216 | 0,969902784 | 0,999171269 | YES |
| 252 | 3,601025168 | -0,032197164 | 0,853935613 | -0,03770444 | 0,969923336 | 0,999171269 | YES |
| 253 | 3,465137426 | -0,032184847 | 0,85393814 | -0,037689905 | 0,969934925 | 0,999171269 | YES |
| 254 | 3,329249684 | -0,032171688 | 0,853940872 | -0,037674374 | 0,969947308 | 0,999171269 | YES |
| 255 | 3,193361941 | -0,032157407 | 0,853943836 | -0,03765752 | 0,969960746 | 0,999171269 | YES |
| 256 | 3,12541807 | -0,032149802 | 0,853945414 | -0,037648545 | 0,969967902 | 0,999171269 | YES |
| 257 | 2,785698715 | -0,03210624 | 0,853954453 | -0,037597134 | 0,970008893 | 0,999171269 | YES |
| 258 | 2,785698715 | -0,03210624 | 0,853954453 | -0,037597134 | 0,970008893 | 0,999171269 | YES |
| 259 | 2,785698715 | -0,03210624 | 0,853954453 | -0,037597134 | 0,970008893 | 0,999171269 | YES |
| 260 | 2,51392323 | -0,032062873 | 0,853963432 | -0,037545955 | 0,970049699 | 0,999171269 | YES |
| 261 | 2,310091617 | -0,032023839 | 0,853971545 | -0,03749989 | 0,970086428 | 0,999171269 | YES |
| 262 | 1,834484519 | -0,031899195 | 0,853997432 | -0,0373528 | 0,970203707 | 0,999171269 | YES |
| 263 | 1,766540648 | -0,031876009 | 0,854002259 | -0,037325439 | 0,970225522 | 0,999171269 | YES |
| 264 | 1,562709035 | -0,031794465 | 0,854019234 | -0,037229214 | 0,970302245 | 0,999171269 | YES |
| 265 | 1,358877422 | -0,031688745 | 0,854041249 | -0,037104467 | 0,970401711 | 0,999171269 | YES |
| 266 | 1,358877422 | -0,031688745 | 0,854041249 | -0,037104467 | 0,970401711 | 0,999171269 | YES |
| 267 | 1,358877422 | -0,031688745 | 0,854041249 | -0,037104467 | 0,970401711 | 0,999171269 | YES |
| 268 | 1,087101937 | -0,031486981 | 0,854083277 | -0,036866406 | 0,970591525 | 0,999171269 | YES |
| 269 | 14,09708202 | 0,030684058 | 0,832514763 | 0,036857074 | 0,970598967 | 0,999171269 | YES |

| Medication | Ticker | ASVnr | taxLevel | Phylum | Family | Genus | Species |
|---|---|---|---|---|---|---|---|
| Pramipexole | 270 | ASV414 | ASV | Firmicutes | Oscillospiraceae | UCG-003 | NA |
| Pramipexole | 271 | ASV487 | ASV | Firmicutes | Lachnospiraceae | Howardella | ureilytica |
| Pramipexole | 272 | ASV119 | ASV | Firmicutes | Ruminococcaceae | Subdoligranulum | NA |
| Pramipexole | 273 | ASV143 | ASV | Firmicutes | Peptostreptococcaceae | Terrisporobacter | mayombei |
| Pramipexole | 274 | ASV241 | ASV | Firmicutes | [Eubacterium] coprostanoligenes group | NA | NA |
| Pramipexole | 275 | ASV828 | ASV | Firmicutes | Christensenellaceae | Christensenellaceae R-7 group | NA |
| Pramipexole | 276 | ASV237 | ASV | Bacteroidota | Bacteroidaceae | Bacteroides | clarus |
| Pramipexole | 277 | ASV320 | ASV | Firmicutes | Christensenellaceae | Christensenellaceae R-7 group | NA |
| Pramipexole | 278 | ASV937 | ASV | Firmicutes | UCG-010 | NA | NA |
| Pramipexole | 279 | ASV240 | ASV | Bacteroidota | Prevotellaceae | Alloprevotella | NA |
| Pramipexole | 280 | ASV76 | ASV | Firmicutes | Lachnospiraceae | Lachnospiraceae ND3007 group | NA |
| Pramipexole | 281 | ASV54 | ASV | Bacteroidota | Bacteroidaceae | Bacteroides | NA |
| Pramipexole | 282 | ASV505 | ASV | Bacteroidota | Bacteroidaceae | Bacteroides | nordii |
| Pramipexole | 283 | ASV529 | ASV | Firmicutes | Lachnospiraceae | NA | NA |
| Pramipexole | 284 | ASV727 | ASV | Firmicutes | Ruminococcaceae | Angelakisella | NA |
| Pramipexole | 285 | ASV308 | ASV | Firmicutes | Christensenellaceae | NA | NA |
| Pramipexole | 286 | ASV277 | ASV | Bacteroidota | Rikenellaceae | Alistipes | ihumii |
| Pramipexole | 287 | ASV51 | ASV | Firmicutes | Oscillospiraceae | UCG-005 | NA |
| Pramipexole | 288 | ASV69 | ASV | Firmicutes | Christensenellaceae | NA | NA |
| Pramipexole | 289 | ASV49 | ASV | Bacteroidota | Bacteroidaceae | Bacteroides | NA |
| Pramipexole | 290 | ASV152 | ASV | Firmicutes | Lachnospiraceae | Roseburia | hominis |
| Pramipexole | 291 | ASV264 | ASV | Firmicutes | Oscillospiraceae | Colidextribacter | NA |
| Pramipexole | 292 | ASV157 | ASV | Firmicutes | Oscillospiraceae | UCG-005 | NA |
| Pramipexole | 293 | ASV299 | ASV | Firmicutes | Streptococcaceae | Lactococcus | NA |
| Pramipexole | 294 | ASV11 | ASV | Firmicutes | Ruminococcaceae | Subdoligranulum | NA |
| Pramipexole | 295 | ASV611 | ASV | Firmicutes | Lachnospiraceae | Lachnospiraceae UCG-010 | NA |
| Pramipexole | 296 | ASV48 | ASV | Bacteroidota | Bacteroidaceae | Bacteroides | caccae |
| Pramipexole | 297 | ASV432 | ASV | Bacteroidota | Marinifilaceae | Butyricimonas | virosa |
| Pramipexole | 298 | ASV46 | ASV | Bacteroidota | Bacteroidaceae | Bacteroides | NA |
| Pramipexole | 299 | ASV332 | ASV | Firmicutes | Lachnospiraceae | [Eubacterium] ventriosum group | NA |

| Ticker | DESeq2_baseMean | DESeq2_log2-FoldChange | DESeq2_lfcSE | DESeq2_stat | DESeq2_pvalue | DESeq2_padj | DESeq2 converged |
|---|---|---|---|---|---|---|---|
| 270 | 0,747382582 | -0,031033164 | 0,854177986 | -0,036331027 | 0,97101841 | 0,999171269 | YES |
| 271 | 0,747382582 | -0,031033164 | 0,854177986 | -0,036331027 | 0,97101841 | 0,999171269 | YES |
| 272 | 0,543550969 | -0,030496367 | 0,854290179 | -0,035697902 | 0,971523244 | 0,999171269 | YES |
| 273 | 0,543550969 | -0,030496367 | 0,854290179 | -0,035697902 | 0,971523244 | 0,999171269 | YES |
| 274 | 0,543550969 | -0,030496367 | 0,854290179 | -0,035697902 | 0,971523244 | 0,999171269 | YES |
| 275 | 0,407663227 | -0,029851532 | 0,854425192 | -0,034937561 | 0,97212953 | 0,999171269 | YES |
| 276 | 80,22179945 | 0,0296199 | 0,85039692 | 0,034830676 | 0,97221476 | 0,999171269 | YES |
| 277 | 0,339719355 | -0,029344135 | 0,854531571 | -0,034339439 | 0,972606476 | 0,999171269 | YES |
| 278 | 0,339719355 | -0,029344135 | 0,854531571 | -0,034339439 | 0,972606476 | 0,999171269 | YES |
| 279 | 7,307610916 | -0,028588105 | 0,832696566 | -0,034331959 | 0,97261244 | 0,999171269 | YES |
| 280 | 14,46116544 | 0,025856222 | 0,809500996 | 0,031940939 | 0,974519151 | 0,999171269 | YES |
| 281 | 415,0059752 | -0,022006646 | 0,826520285 | -0,026625657 | 0,978758309 | 0,999171269 | YES |
| 282 | 1,931225187 | 0,018961968 | 0,850787729 | 0,022287543 | 0,982218585 | 0,999171269 | YES |
| 283 | 13,63473783 | 0,017971793 | 0,850444391 | 0,021132237 | 0,983140169 | 0,999171269 | YES |
| 284 | 10,34678682 | -0,017727611 | 0,85046333 | -0,02084465 | 0,98336958 | 0,999171269 | YES |
| 285 | 42,53257762 | 0,017509156 | 0,850405539 | 0,020589184 | 0,983573369 | 0,999171269 | YES |
| 286 | 5,115312645 | -0,017097067 | 0,850540877 | -0,020101406 | 0,983962479 | 0,999171269 | YES |
| 287 | 21,20583703 | 0,016540295 | 0,853881715 | 0,019370709 | 0,984545377 | 0,999171269 | YES |
| 288 | 3,545500781 | 0,015998067 | 0,853935321 | 0,018734519 | 0,985052891 | 0,999171269 | YES |
| 289 | 1273,945506 | -0,014713629 | 0,809874194 | -0,018167796 | 0,985504993 | 0,999171269 | YES |
| 290 | 34,40256681 | -0,014199448 | 0,829854867 | -0,017110761 | 0,986348254 | 0,999171269 | YES |
| 291 | 33,00466302 | 0,013889807 | 0,830102853 | 0,016732634 | 0,986649913 | 0,999171269 | YES |
| 292 | 6,191430665 | -0,012877364 | 0,853908162 | -0,015080503 | 0,987967956 | 0,999171269 | YES |
| 293 | 18,91393827 | 0,011287234 | 0,850428464 | 0,013272408 | 0,989410461 | 0,999171269 | YES |
| 294 | 334,2045594 | 0,006745949 | 0,516578547 | 0,013058903 | 0,989580799 | 0,999171269 | YES |
| 295 | 32,94686015 | 0,010588977 | 0,826723027 | 0,012808373 | 0,989780677 | 0,999171269 | YES |
| 296 | 536,4100119 | -0,005622911 | 0,54296336 | -0,010355968 | 0,991737281 | 0,999171269 | YES |
| 297 | 13,44926126 | 0,008462655 | 0,830129622 | 0,010194378 | 0,991866204 | 0,999171269 | YES |
| 298 | 80,22945882 | 0,006215891 | 0,622208378 | 0,009990047 | 0,992029228 | 0,999171269 | YES |
| 299 | 42,81919195 | 0,008223216 | 0,832360798 | 0,009879388 | 0,992117517 | 0,999171269 | YES |

| Medication | Ticker | ASVnr | taxLevel | Phylum | Family | Genus | Species |
|---|---|---|---|---|---|---|---|
| Pramipexole | 300 | ASV53 | ASV | Firmicutes | Lachnospiraceae | [Ruminococcus] torques group | NA |
| Pramipexole | 301 | ASV58 | ASV | Bacteroidota | Bacteroidaceae | Bacteroides | NA |
| Pramipexole | 302 | ASV326 | ASV | Proteobacteria | NA | NA | NA |
| Pramipexole | 303 | ASV310 | ASV | Firmicutes | Lachnospiraceae | NA | NA |
| Pramipexole | 304 | ASV60 | ASV | Firmicutes | Ruminococcaceae | [Eubacterium] siraeum group | NA |
| Pramipexole | 305 | ASV550 | Genus | Bacteroidota | Rikenellaceae | Rikenellaceae RC9 gut group | NA |
| Pramipexole | 306 | ASV13 | Genus | Firmicutes | Lachnospiraceae | Fusicatenibacter | NA |
| Pramipexole | 307 | ASV328 | Genus | Firmicutes | Anaerovoracaceae | [Eubacterium] brachy group | NA |
| Pramipexole | 308 | ASV111 | Genus | Firmicutes | Lachnospiraceae | Lachnospira | NA |
| Pramipexole | 309 | ASV1 | Genus | Actinobacteriota | Bifidobacteriaceae | Bifidobacterium | NA |
| Pramipexole | 310 | ASV51 | Genus | Firmicutes | Oscillospiraceae | UCG-005 | NA |
| Pramipexole | 311 | ASV322 | Genus | Firmicutes | Anaerovoracaceae | Family XIII AD3011 group | NA |
| Pramipexole | 312 | ASV386 | Genus | Firmicutes | Ruminococcaceae | DTU089 | NA |
| Pramipexole | 313 | ASV10 | Genus | Verrucomicrobiota | Akkermansiaceae | Akkermansia | NA |
| Pramipexole | 314 | ASV147 | Genus | Firmicutes | Oscillospiraceae | Oscillibacter | NA |
| Pramipexole | 315 | ASV295 | Genus | Firmicutes | Ruminococcaceae | Negativibacillus | NA |
| Pramipexole | 316 | ASV65 | Genus | Firmicutes | Ruminococcaceae | Incertae Sedis | NA |
| Pramipexole | 317 | ASV23 | Genus | Firmicutes | Streptococcaceae | Streptococcus | NA |
| Pramipexole | 318 | ASV357 | Genus | Firmicutes | Lachnospiraceae | Marvinbryantia | NA |
| Pramipexole | 319 | ASV411 | Genus | Firmicutes | Lactobacillaceae | Lactobacillus | NA |
| Pramipexole | 320 | ASV193 | Genus | Actinobacteriota | Eggerthellaceae | Adlercreutzia | NA |
| Pramipexole | 321 | ASV630 | Genus | Firmicutes | Ruminococcaceae | Angelakisella | NA |
| Pramipexole | 322 | ASV96 | Genus | Firmicutes | Lachnospiraceae | CAG-56 | NA |
| Pramipexole | 323 | ASV92 | Genus | Firmicutes | Erysipelotrichaceae | Turicibacter | NA |
| Pramipexole | 324 | ASV97 | Genus | Firmicutes | Lachnospiraceae | [Ruminococcus] gnavus group | NA |
| Pramipexole | 325 | ASV16 | Genus | Firmicutes | Peptostreptococcaceae | Romboutsia | NA |
| Pramipexole | 326 | ASV57 | Genus | Firmicutes | Lachnospiraceae | Roseburia | NA |
| Pramipexole | 327 | ASV200 | Genus | Firmicutes | Lachnospiraceae | [Eubacterium] xylanophilum group | NA |
| Pramipexole | 328 | ASV60 | Genus | Firmicutes | Ruminococcaceae | [Eubacterium] siraeum group | NA |
| Pramipexole | 329 | ASV5 | Genus | Firmicutes | Lachnospiraceae | Blautia | NA |

| Ticker | DESeq2_baseMean | DESeq2_log2-FoldChange | DESeq2_lfcSE | DESeq2_stat | DESeq2_pvalue | DESeq2_padj | DESeq2 converged |
|---|---|---|---|---|---|---|---|
| 300 | 603,4481863 | 0,005036613 | 0,519467491 | 0,009695723 | 0,992264054 | 0,999171269 | YES |
| 301 | 174,7509689 | 0,005755905 | 0,829729639 | 0,006937085 | 0,994465051 | 0,999171269 | YES |
| 302 | 2,460439192 | -0,004965352 | 0,833487226 | -0,005957322 | 0,995246773 | 0,999171269 | YES |
| 303 | 10,48431403 | -0,004404388 | 0,85389285 | -0,00515801 | 0,995884522 | 0,999171269 | YES |
| 304 | 277,2992052 | 0,00021833 | 0,636245048 | 0,000343154 | 0,999726202 | 0,999726202 | YES |
| 305 | 116,3580989 | -17,72637817 | 0,855404985 | -20,72279036 | 2,16E-95 | 2,55E-93 | YES |
| 306 | 899,3581077 | 1,877663729 | 0,462147085 | 4,062913714 | 4,85E-05 | 0,002859373 | YES |
| 307 | 15,82505315 | 0,968220818 | 0,405417388 | 2,388207429 | 0,016930783 | 0,665944125 | YES |
| 308 | 177,5815805 | -0,636274252 | 0,291651014 | -2,181628799 | 0,02913694 | 0,824488376 | YES |
| 309 | 1344,355805 | 0,449556684 | 0,213150954 | 2,109100032 | 0,034935948 | 0,824488376 | YES |
| 310 | 207,6260688 | 1,037013027 | 0,517002143 | 2,005819591 | 0,0448755 | 0,882551493 | YES |
| 311 | 66,47623354 | 0,350003267 | 0,19512341 | 1,793753331 | 0,072852549 | 0,994962331 | YES |
| 312 | 52,03740731 | 0,521631132 | 0,298327636 | 1,74851763 | 0,080374435 | 0,994962331 | YES |
| 313 | 1364,830344 | -0,77772757 | 0,450310519 | -1,727091724 | 0,084151192 | 0,994962331 | YES |
| 314 | 409,7871718 | 0,276357426 | 0,165359195 | 1,671255268 | 0,094671268 | 0,994962331 | YES |
| 315 | 117,8970404 | 0,357779455 | 0,227956148 | 1,569510009 | 0,116529149 | 0,994962331 | YES |
| 316 | 281,134366 | 0,290670902 | 0,203655088 | 1,427270516 | 0,153501936 | 0,994962331 | YES |
| 317 | 531,702743 | 0,469944422 | 0,341008953 | 1,378099951 | 0,168172431 | 0,994962331 | YES |
| 318 | 41,68597386 | 0,698627711 | 0,510111782 | 1,369558076 | 0,170824894 | 0,994962331 | YES |
| 319 | 4,952380139 | 1,074701307 | 0,82321932 | 1,305486013 | 0,191727407 | 0,994962331 | YES |
| 320 | 11,47978911 | 1,021798862 | 0,799694447 | 1,277736598 | 0,201342316 | 0,994962331 | YES |
| 321 | 41,88704901 | 0,521434385 | 0,424953944 | 1,227037406 | 0,219808532 | 0,994962331 | YES |
| 322 | 343,2845611 | -0,719958231 | 0,587440054 | -1,225585874 | 0,220354556 | 0,994962331 | YES |
| 323 | 7,336055277 | 0,990397012 | 0,821566242 | 1,20549867 | 0,228010851 | 0,994962331 | YES |
| 324 | 2,812958118 | -0,963761703 | 0,831597139 | -1,158928594 | 0,246485292 | 0,994962331 | YES |
| 325 | 33,54813725 | 0,663032159 | 0,579963827 | 1,143230195 | 0,252943029 | 0,994962331 | YES |
| 326 | 371,9546907 | -0,427864597 | 0,384256092 | -1,113488128 | 0,265498844 | 0,994962331 | YES |
| 327 | 57,28847948 | -0,533532333 | 0,499741857 | -1,06761586 | 0,285693826 | 0,994962331 | YES |
| 328 | 349,2160858 | -0,200444944 | 0,189724705 | -1,056504177 | 0,290737933 | 0,994962331 | YES |
| 329 | 2676,734993 | -0,168886274 | 0,165625428 | -1,019688073 | 0,30787642 | 0,994962331 | YES |

| Medication | Ticker | ASVnr | taxLevel | Phylum | Family | Genus | Species |
|---|---|---|---|---|---|---|---|
| Pramipexole | 330 | ASV37 | Genus | Firmicutes | Lachnospiraceae | [Eubacterium] hallii group | NA |
| Pramipexole | 331 | ASV31 | Genus | Bacteroidota | Rikenellaceae | Alistipes | NA |
| Pramipexole | 332 | ASV30 | Genus | Proteobacteria | Enterobacteriaceae | Escherichia-Shigella | NA |
| Pramipexole | 333 | ASV26 | Genus | Euryarchaeota | Methanobacteriaceae | Methanobrevibacter | NA |
| Pramipexole | 334 | ASV303 | Genus | Firmicutes | Lachnospiraceae | GCA-900066575 | NA |
| Pramipexole | 335 | ASV18 | Genus | Firmicutes | Lachnospiraceae | Dorea | NA |
| Pramipexole | 336 | ASV7 | Genus | Firmicutes | Lachnospiraceae | Agathobacter | NA |
| Pramipexole | 337 | ASV4 | Genus | Firmicutes | Ruminococcaceae | Ruminococcus | NA |
| Pramipexole | 338 | ASV128 | Genus | Firmicutes | Oscillospiraceae | NK4A214 group | NA |
| Pramipexole | 339 | ASV192 | Genus | Bacteroidota | Marinifilaceae | Odoribacter | NA |
| Pramipexole | 340 | ASV17 | Genus | Actinobacteriota | Coriobacteriaceae | Collinsella | NA |
| Pramipexole | 341 | ASV87 | Genus | Proteobacteria | Sutterellaceae | Sutterella | NA |
| Pramipexole | 342 | ASV3 | Genus | Firmicutes | Ruminococcaceae | Subdoligranulum | NA |
| Pramipexole | 343 | ASV141 | Genus | Firmicutes | Lachnospiraceae | Lachnoclostridium | NA |
| Pramipexole | 344 | ASV53 | Genus | Firmicutes | Lachnospiraceae | [Ruminococcus] torques group | NA |
| Pramipexole | 345 | ASV20 | Genus | Bacteroidota | Prevotellaceae | Prevotella | NA |
| Pramipexole | 346 | ASV55 | Genus | Firmicutes | Monoglobaceae | Monoglobus | NA |
| Pramipexole | 347 | ASV172 | Genus | Proteobacteria | Pasteurellaceae | Haemophilus | NA |
| Pramipexole | 348 | ASV309 | Genus | Firmicutes | Oscillospiraceae | Colidextribacter | NA |
| Pramipexole | 349 | ASV15 | Genus | Firmicutes | Christensenellaceae | Christensenellaceae R-7 group | NA |
| Pramipexole | 350 | ASV28 | Genus | Firmicutes | Clostridiaceae | Clostridium sensu stricto 1 | NA |
| Pramipexole | 351 | ASV85 | Genus | Firmicutes | Lachnospiraceae | Coprococcus | NA |
| Pramipexole | 352 | ASV195 | Genus | Bacteroidota | Barnesiellaceae | Barnesiella | NA |
| Pramipexole | 353 | ASV283 | Genus | Actinobacteriota | Eggerthellaceae | Slackia | NA |
| Pramipexole | 354 | ASV2 | Genus | Bacteroidota | Bacteroidaceae | Bacteroides | NA |
| Pramipexole | 355 | ASV666 | Genus | Firmicutes | Erysipelotrichaceae | Holdemania | NA |
| Pramipexole | 356 | ASV33 | Genus | Firmicutes | Peptostreptococcaceae | Intestinibacter | NA |
| Pramipexole | 357 | ASV376 | Genus | Bacteroidota | Barnesiellaceae | Coprobacter | NA |
| Pramipexole | 358 | ASV512 | Genus | Proteobacteria | Oxalobacteraceae | Oxalobacter | NA |
| Pramipexole | 359 | ASV415 | Genus | Firmicutes | Oscillospiraceae | Oscillospira | NA |

| Ticker | DESeq2_baseMean | DESeq2_log2-FoldChange | DESeq2_lfcSE | DESeq2_stat | DESeq2_pvalue | DESeq2_padj | DESeq2 converged |
|---|---|---|---|---|---|---|---|
| 330 | 384,917426 | 0,13183838 | 0,137709243 | 0,957367693 | 0,338381699 | 0,994962331 | YES |
| 331 | 1102,079131 | 0,258644605 | 0,271927374 | 0,951153248 | 0,341526587 | 0,994962331 | YES |
| 332 | 130,0618911 | 0,490411845 | 0,521853337 | 0,939750328 | 0,347345643 | 0,994962331 | YES |
| 333 | 80,54366585 | 0,343918206 | 0,367354846 | 0,936201629 | 0,349169384 | 0,994962331 | YES |
| 334 | 44,19644707 | 0,336794977 | 0,36764565 | 0,916085848 | 0,35962187 | 0,994962331 | YES |
| 335 | 677,4623792 | -0,185855213 | 0,204716587 | -0,907865926 | 0,363949062 | 0,994962331 | YES |
| 336 | 4103,809375 | 0,153151764 | 0,173841921 | 0,880982922 | 0,378327063 | 0,994962331 | YES |
| 337 | 1502,742616 | -0,261909215 | 0,30793129 | -0,850544338 | 0,395022521 | 0,994962331 | YES |
| 338 | 307,3640933 | 0,294473746 | 0,348380093 | 0,845265707 | 0,397962503 | 0,994962331 | YES |
| 339 | 24,07193252 | -0,218501263 | 0,267884896 | -0,815653538 | 0,41469832 | 0,994962331 | YES |
| 340 | 807,8694594 | 0,171544125 | 0,211803043 | 0,809922852 | 0,417984517 | 0,994962331 | YES |
| 341 | 145,7481738 | 0,208168938 | 0,262479105 | 0,793087658 | 0,427726753 | 0,994962331 | YES |
| 342 | 1353,359098 | 0,143293592 | 0,182249762 | 0,786248445 | 0,43172195 | 0,994962331 | YES |
| 343 | 308,8576131 | -0,14571551 | 0,196888532 | -0,740091404 | 0,459244534 | 0,994962331 | YES |
| 344 | 1058,1215 | -0,121670945 | 0,165768217 | -0,733982346 | 0,462959498 | 0,994962331 | YES |
| 345 | 1982,176061 | -0,30463421 | 0,419665798 | -0,725897159 | 0,467901818 | 0,994962331 | YES |
| 346 | 218,6629912 | 0,182988645 | 0,262708359 | 0,696546716 | 0,486086512 | 0,994962331 | YES |
| 347 | 15,46153561 | -0,537628179 | 0,824166322 | -0,652329711 | 0,514188497 | 0,994962331 | YES |
| 348 | 331,4686411 | 0,102225059 | 0,161530058 | 0,632854715 | 0,526828523 | 0,994962331 | YES |
| 349 | 286,0527492 | 0,220641622 | 0,355123094 | 0,62131026 | 0,534395504 | 0,994962331 | YES |
| 350 | 34,84101479 | -0,261429218 | 0,431948636 | -0,605232187 | 0,545024732 | 0,994962331 | YES |
| 351 | 574,4731389 | -0,144569651 | 0,248290521 | -0,582260051 | 0,56039153 | 0,994962331 | YES |
| 352 | 259,4575144 | 0,195232878 | 0,364728869 | 0,535282218 | 0,592454721 | 0,994962331 | YES |
| 353 | 5,414722143 | 0,44457136 | 0,832898079 | 0,53376442 | 0,593504533 | 0,994962331 | YES |
| 354 | 7941,910738 | -0,071206157 | 0,134270721 | -0,530317823 | 0,595891591 | 0,994962331 | YES |
| 355 | 9,418012638 | 0,138066158 | 0,262926167 | 0,525113795 | 0,599504087 | 0,994962331 | YES |
| 356 | 42,5719079 | -0,189561195 | 0,365322706 | -0,518886977 | 0,60383956 | 0,994962331 | YES |
| 357 | 3,742647479 | 0,431755371 | 0,836093107 | 0,516396281 | 0,605577663 | 0,994962331 | YES |
| 358 | 12,48260945 | 0,308954811 | 0,602552709 | 0,51274321 | 0,608130959 | 0,994962331 | YES |
| 359 | 55,07116979 | -0,105607934 | 0,221911224 | -0,475901725 | 0,634144393 | 0,994962331 | YES |

| Medication | Ticker | ASVnr | taxLevel | Phylum | Family | Genus | Species |
|---|---|---|---|---|---|---|---|
| Pramipexole | 360 | ASV72 | Genus | Proteobacteria | Sutterellaceae | Parasutterella | NA |
| Pramipexole | 361 | ASV39 | Genus | Bacteroidota | Tannerellaceae | Parabacteroides | NA |
| Pramipexole | 362 | ASV214 | Genus | Firmicutes | Lachnospiraceae | Lachnospiraceae UCG-010 | NA |
| Pramipexole | 363 | ASV19 | Genus | Firmicutes | Erysipelatoclostridiaceae | Erysipelotrichaceae UCG-003 | NA |
| Pramipexole | 364 | ASV217 | Genus | Firmicutes | Peptococcaceae | Peptococcus | NA |
| Pramipexole | 365 | ASV167 | Genus | Firmicutes | Oscillospiraceae | Intestinimonas | NA |
| Pramipexole | 366 | ASV1319 | Genus | Firmicutes | Ruminococcaceae | Candidatus Soleaferrea | NA |
| Pramipexole | 367 | ASV242 | Genus | Firmicutes | Lachnospiraceae | Tyzzerella | NA |
| Pramipexole | 368 | ASV690 | Genus | Firmicutes | Ruminococcaceae | Fournierella | NA |
| Pramipexole | 369 | ASV66 | Genus | Firmicutes | Butyricicoccaceae | Butyricicoccus | NA |
| Pramipexole | 370 | ASV78 | Genus | Firmicutes | Lachnospiraceae | [Eubacterium] eligens group | NA |
| Pramipexole | 371 | ASV226 | Genus | Firmicutes | Lachnospiraceae | Lachnospiraceae UCG-001 | NA |
| Pramipexole | 372 | ASV12 | Genus | Firmicutes | Veillonellaceae | Dialister | NA |
| Pramipexole | 373 | ASV190 | Genus | Firmicutes | Ruminococcaceae | UBA1819 | NA |
| Pramipexole | 374 | ASV739 | Genus | Firmicutes | Ruminococcaceae | Anaerotruncus | NA |
| Pramipexole | 375 | ASV75 | Genus | Firmicutes | Lachnospiraceae | [Ruminococcus] gauvreauii group | NA |
| Pramipexole | 376 | ASV158 | Genus | Firmicutes | Lachnospiraceae | [Eubacterium] ventriosum group | NA |
| Pramipexole | 377 | ASV81 | Genus | Firmicutes | Lachnospiraceae | Lachnospiraceae NK4A136 group | NA |
| Pramipexole | 378 | ASV453 | Genus | Verrucomicrobiota | Victivallaceae | Victivallis | NA |
| Pramipexole | 379 | ASV408 | Genus | Bacteroidota | Prevotellaceae | Paraprevotella | NA |
| Pramipexole | 380 | ASV974 | Genus | Firmicutes | Butyricicoccaceae | UCG-009 | NA |
| Pramipexole | 381 | ASV306 | Genus | Firmicutes | Veillonellaceae | Veillonella | NA |
| Pramipexole | 382 | ASV211 | Genus | Firmicutes | Lachnospiraceae | Lachnospiraceae FCS020 group | NA |
| Pramipexole | 383 | ASV98 | Genus | Firmicutes | Acidaminococcaceae | Phascolarctobacterium | NA |
| Pramipexole | 384 | ASV885 | Genus | Firmicutes | Lachnospiraceae | Shuttleworthia | NA |
| Pramipexole | 385 | ASV8 | Genus | Firmicutes | Ruminococcaceae | Faecalibacterium | NA |
| Pramipexole | 386 | ASV499 | Genus | Firmicutes | Lachnospiraceae | Eisenbergiella | NA |
| Pramipexole | 387 | ASV1631 | Genus | Firmicutes | Ruminococcaceae | Paludicola | NA |
| Pramipexole | 388 | ASV315 | Genus | Firmicutes | Anaerovoracaceae | Family XIII UCG-001 | NA |
| Pramipexole | 389 | ASV116 | Genus | Firmicutes | Oscillospiraceae | UCG-003 | NA |

| Ticker | DESeq2_baseMean | DESeq2_log2-FoldChange | DESeq2_lfcSE | DESeq2_stat | DESeq2_pvalue | DESeq2_padj | DESeq2 converged |
|---|---|---|---|---|---|---|---|
| 360 | 218,5495706 | -0,119259884 | 0,251298834 | -0,474573962 | 0,635090669 | 0,994962331 | YES |
| 361 | 834,0789902 | 0,177957222 | 0,384367243 | 0,462987482 | 0,64337334 | 0,994962331 | YES |
| 362 | 71,03005293 | 0,114712081 | 0,264160869 | 0,434250847 | 0,664106293 | 0,994962331 | YES |
| 363 | 241,6263661 | -0,18997209 | 0,461161795 | -0,411942385 | 0,680381653 | 0,994962331 | YES |
| 364 | 10,76653394 | -0,337145426 | 0,833416519 | -0,404534129 | 0,685819986 | 0,994962331 | YES |
| 365 | 170,7079586 | -0,104801296 | 0,260852234 | -0,401764992 | 0,687856988 | 0,994962331 | YES |
| 366 | 1,731469796 | -0,320690257 | 0,820544461 | -0,39082618 | 0,695925722 | 0,994962331 | YES |
| 367 | 7,349290314 | 0,313194526 | 0,833050864 | 0,375960868 | 0,706945988 | 0,994962331 | YES |
| 368 | 1,790211743 | 0,30537128 | 0,835308824 | 0,365578898 | 0,714679323 | 0,994962331 | YES |
| 369 | 196,1325531 | -0,049410722 | 0,135684608 | -0,364158641 | 0,71573955 | 0,994962331 | YES |
| 370 | 184,1701242 | -0,209188193 | 0,623369976 | -0,335576305 | 0,737190395 | 0,994962331 | YES |
| 371 | 29,79850509 | -0,275296079 | 0,830176213 | -0,33161162 | 0,740182544 | 0,994962331 | YES |
| 372 | 1236,026384 | 0,194115689 | 0,641742808 | 0,302482064 | 0,762284604 | 0,994962331 | YES |
| 373 | 12,83772834 | 0,168287602 | 0,564486487 | 0,298125121 | 0,765607669 | 0,994962331 | YES |
| 374 | 4,452740537 | -0,241049761 | 0,829736499 | -0,290513629 | 0,771423326 | 0,994962331 | YES |
| 375 | 6,575366094 | 0,228797214 | 0,830770991 | 0,27540347 | 0,783006278 | 0,994962331 | YES |
| 376 | 111,1692141 | -0,09930479 | 0,370475504 | -0,268046845 | 0,788663261 | 0,994962331 | YES |
| 377 | 180,0333742 | 0,109184175 | 0,414460645 | 0,263436773 | 0,792213951 | 0,994962331 | YES |
| 378 | 0,778211941 | -0,219920045 | 0,850287159 | -0,258642087 | 0,795911407 | 0,994962331 | YES |
| 379 | 73,12775143 | -0,15037776 | 0,585322872 | -0,256914205 | 0,797245005 | 0,994962331 | YES |
| 380 | 12,89337023 | 0,136733633 | 0,55986178 | 0,244227482 | 0,807054644 | 0,994962331 | YES |
| 381 | 17,04352115 | -0,201886302 | 0,830910662 | -0,242969926 | 0,808028695 | 0,994962331 | YES |
| 382 | 43,80668629 | -0,053662368 | 0,222770387 | -0,24088645 | 0,809643124 | 0,994962331 | YES |
| 383 | 587,9129411 | -0,119595086 | 0,497807771 | -0,240243509 | 0,810141486 | 0,994962331 | YES |
| 384 | 8,044064653 | -0,178746386 | 0,754093911 | -0,23703465 | 0,81262991 | 0,994962331 | YES |
| 385 | 2498,489811 | 0,031609089 | 0,134857629 | 0,234388587 | 0,814683318 | 0,994962331 | YES |
| 386 | 42,40951613 | 0,137775961 | 0,64019551 | 0,215209196 | 0,829604222 | 0,994962331 | YES |
| 387 | 6,285305421 | 0,168413042 | 0,809974677 | 0,207923836 | 0,835288438 | 0,994962331 | YES |
| 388 | 19,80434916 | 0,108587898 | 0,540395695 | 0,200941457 | 0,840744351 | 0,994962331 | YES |
| 389 | 77,4927846 | -0,135567187 | 0,685895104 | -0,197650029 | 0,843318889 | 0,994962331 | YES |

| Medication | Ticker | ASVnr | taxLevel | Phylum | Family | Genus | Species |
|---|---|---|---|---|---|---|---|
| Pramipexole | 390 | ASV29 | Genus | Firmicutes | Oscillospiraceae | UCG-002 | NA |
| Pramipexole | 391 | ASV40 | Genus | Firmicutes | Erysipelotrichaceae | Holdemanella | NA |
| Pramipexole | 392 | ASV573 | Genus | Firmicutes | Erysipelatoclostridiaceae | Erysipelatoclostridium | NA |
| Pramipexole | 393 | ASV27 | Genus | Firmicutes | Ruminococcaceae | CAG-352 | NA |
| Pramipexole | 394 | ASV229 | Genus | Firmicutes | Oscillospiraceae | Flavonifractor | NA |
| Pramipexole | 395 | ASV240 | Genus | Bacteroidota | Prevotellaceae | Alloprevotella | NA |
| Pramipexole | 396 | ASV252 | Genus | Firmicutes | Erysipelotrichaceae | Solobacterium | NA |
| Pramipexole | 397 | ASV61 | Genus | Firmicutes | Erysipelatoclostridiaceae | Catenibacterium | NA |
| Pramipexole | 398 | ASV162 | Genus | Desulfobacterota | Desulfovibrionaceae | Desulfovibrio | NA |
| Pramipexole | 399 | ASV530 | Genus | Firmicutes | Veillonellaceae | Allisonella | NA |
| Pramipexole | 400 | ASV258 | Genus | Firmicutes | Lachnospiraceae | Lachnospiraceae AC2044 group | NA |
| Pramipexole | 401 | ASV487 | Genus | Firmicutes | Lachnospiraceae | Howardella | NA |
| Pramipexole | 402 | ASV143 | Genus | Firmicutes | Peptostreptococcaceae | Terrisporobacter | NA |
| Pramipexole | 403 | ASV536 | Genus | Bacteroidota | Marinifilaceae | Butyricimonas | NA |
| Pramipexole | 404 | ASV293 | Genus | Firmicutes | Erysipelotrichaceae | Catenisphaera | NA |
| Pramipexole | 405 | ASV299 | Genus | Firmicutes | Streptococcaceae | Lactococcus | NA |
| Pramipexole | 406 | ASV761 | Genus | Firmicutes | Defluviitaleaceae | Defluviitaleaceae UCG-011 | NA |
| Pramipexole | 407 | ASV219 | Genus | Firmicutes | Lachnospiraceae | Lachnospiraceae UCG-004 | NA |
| Pramipexole | 408 | ASV444 | Genus | Firmicutes | Erysipelatoclostridiaceae | Coprobacillus | NA |
| Pramipexole | 409 | ASV133 | Genus | Actinobacteriota | Eggerthellaceae | Senegalimassilia | NA |
| Pramipexole | 410 | ASV134 | Genus | Actinobacteriota | Eggerthellaceae | Enterorhabdus | NA |
| Pramipexole | 411 | ASV1506 | Genus | Firmicutes | Erysipelatoclostridiaceae | UCG-004 | NA |
| Pramipexole | 412 | ASV79 | Genus | Firmicutes | Lachnospiraceae | [Eubacterium] ruminantium group | NA |
| Pramipexole | 413 | ASV1722 | Genus | Firmicutes | Erysipelatoclostridiaceae | Asteroleplasma | NA |
| Pramipexole | 414 | ASV144 | Genus | Desulfobacterota | Desulfovibrionaceae | Bilophila | NA |
| Pramipexole | 415 | ASV197 | Genus | Bacteroidota | Prevotellaceae | Prevotellaceae NK3B31 group | NA |
| Pramipexole | 416 | ASV76 | Genus | Firmicutes | Lachnospiraceae | Lachnospiraceae ND3007 group | NA |
| Pramipexole | 417 | ASV34 | Genus | Firmicutes | Lachnospiraceae | Anaerostipes | NA |
| Pramipexole | 418 | ASV294 | Genus | Firmicutes | Anaerovoracaceae | Mogibacterium | NA |
| Pramipexole | 419 | ASV978 | Genus | Firmicutes | Erysipelotrichaceae | Dielma | NA |

| Ticker | DESeq2_baseMean | DESeq2_log2-FoldChange | DESeq2_lfcSE | DESeq2_stat | DESeq2_pvalue | DESeq2_padj | DESeq2 converged |
|---|---|---|---|---|---|---|---|
| 390 | 526,5545115 | 0,0676718 | 0,345922734 | 0,195626923 | 0,844902186 | 0,994962331 | YES |
| 391 | 87,88670941 | -0,164061411 | 0,84766313 | -0,193545532 | 0,846531751 | 0,994962331 | YES |
| 392 | 110,1501044 | -0,065489156 | 0,382171962 | -0,171360441 | 0,863940358 | 0,994962331 | YES |
| 393 | 312,4986088 | 0,135230439 | 0,829778017 | 0,162971827 | 0,870540617 | 0,994962331 | YES |
| 394 | 161,6784889 | -0,034994682 | 0,276787286 | -0,126431683 | 0,899390224 | 0,994962331 | YES |
| 395 | 7,419443565 | -0,10044452 | 0,8327031 | -0,12062465 | 0,903988343 | 0,994962331 | YES |
| 396 | 7,174649577 | -0,098424682 | 0,85391074 | -0,115263432 | 0,908236322 | 0,994962331 | YES |
| 397 | 5,794909274 | -0,098383235 | 0,853920197 | -0,115213618 | 0,908275805 | 0,994962331 | YES |
| 398 | 91,9738297 | 0,066530013 | 0,577643814 | 0,115174803 | 0,90830657 | 0,994962331 | YES |
| 399 | 4,507151657 | -0,098322158 | 0,853934229 | -0,1151402 | 0,908333997 | 0,994962331 | YES |
| 400 | 1,839653738 | -0,097927471 | 0,854025257 | -0,114665779 | 0,90871004 | 0,994962331 | YES |
| 401 | 1,011809556 | -0,097389864 | 0,854149603 | -0,11401968 | 0,909222193 | 0,994962331 | YES |
| 402 | 0,735861495 | -0,096947829 | 0,854251922 | -0,11348857 | 0,909643224 | 0,994962331 | YES |
| 403 | 18,7865398 | -0,04060173 | 0,359641176 | -0,112895109 | 0,910113714 | 0,994962331 | YES |
| 404 | 16,85469065 | 0,077459038 | 0,83000381 | 0,093323713 | 0,925646394 | 0,994962331 | YES |
| 405 | 17,17654912 | -0,060626043 | 0,850430047 | -0,071288688 | 0,943167998 | 0,994962331 | YES |
| 406 | 0,855235155 | -0,059412415 | 0,849778708 | -0,069915161 | 0,944261186 | 0,994962331 | YES |
| 407 | 89,92750812 | 0,042160593 | 0,605075582 | 0,069678225 | 0,944449773 | 0,994962331 | YES |
| 408 | 2,89436445 | 0,058715932 | 0,85396815 | 0,068756583 | 0,945183378 | 0,994962331 | YES |
| 409 | 0,423565529 | 0,055794179 | 0,854510057 | 0,065293765 | 0,947940107 | 0,994962331 | YES |
| 410 | 9,98438168 | 0,03965784 | 0,626906475 | 0,063259579 | 0,949559803 | 0,994962331 | YES |
| 411 | 0,14118851 | 0,049859674 | 0,855634694 | 0,058272151 | 0,95353185 | 0,994962331 | YES |
| 412 | 19,07108089 | 0,045661599 | 0,853885579 | 0,053475079 | 0,957353386 | 0,994962331 | YES |
| 413 | 37,71216728 | 0,044357301 | 0,853878325 | 0,051948035 | 0,9585701 | 0,994962331 | YES |
| 414 | 98,93991685 | -0,015963707 | 0,341147965 | -0,046794086 | 0,962677342 | 0,994962331 | YES |
| 415 | 231,8491692 | -0,037813805 | 0,850390325 | -0,04446641 | 0,964532626 | 0,994962331 | YES |
| 416 | 16,3345729 | -0,032530282 | 0,809683273 | -0,040176552 | 0,967952371 | 0,994962331 | YES |
| 417 | 281,498817 | 0,00408487 | 0,169564276 | 0,024090391 | 0,980780508 | 0,994962331 | YES |
| 418 | 9,803946621 | 0,020059138 | 0,853898955 | 0,023491232 | 0,981258432 | 0,994962331 | YES |
| 419 | 0,959063134 | 0,017395597 | 0,851134923 | 0,020438119 | 0,983693876 | 0,994962331 | YES |

| Medication | Ticker | ASVnr | taxLevel | Phylum | Family | Genus | Species |
|---|---|---|---|---|---|---|---|
| Pramipexole | 420 | ASV232 | Genus | Firmicutes | Acholeplasmataceae | Anaeroplasma | NA |
| Pramipexole | 421 | ASV247 | Genus | Euryarchaeota | Methanobacteriaceae | Methanosphaera | NA |
| Pramipexole | 422 | ASV454 | Genus | Actinobacteriota | Actinomycetaceae | Actinomyces | NA |
| Pramipexole | 423 | ASV1 | Family | Actinobacteriota | Bifidobacteriaceae | NA | NA |
| Pramipexole | 424 | ASV63 | Family | Actinobacteriota | Eggerthellaceae | NA | NA |
| Pramipexole | 425 | ASV322 | Family | Firmicutes | Anaerovoracaceae | NA | NA |
| Pramipexole | 426 | ASV10 | Family | Verrucomicrobiota | Akkermansiaceae | NA | NA |
| Pramipexole | 427 | ASV145 | Family | Bacteroidota | Muribaculaceae | NA | NA |
| Pramipexole | 428 | ASV15 | Family | Firmicutes | Christensenellaceae | NA | NA |
| Pramipexole | 429 | ASV59 | Family | Firmicutes | [Eubacterium] coprostanoligenes group | NA | NA |
| Pramipexole | 430 | ASV411 | Family | Firmicutes | Lactobacillaceae | NA | NA |
| Pramipexole | 431 | ASV30 | Family | Proteobacteria | Enterobacteriaceae | NA | NA |
| Pramipexole | 432 | ASV23 | Family | Firmicutes | Streptococcaceae | NA | NA |
| Pramipexole | 433 | ASV217 | Family | Firmicutes | Peptococcaceae | NA | NA |
| Pramipexole | 434 | ASV19 | Family | Firmicutes | Erysipelatoclostridiaceae | NA | NA |
| Pramipexole | 435 | ASV192 | Family | Bacteroidota | Marinifilaceae | NA | NA |
| Pramipexole | 436 | ASV20 | Family | Bacteroidota | Prevotellaceae | NA | NA |
| Pramipexole | 437 | ASV255 | Family | Actinobacteriota | Coriobacteriales Incertae Sedis | NA | NA |
| Pramipexole | 438 | ASV17 | Family | Actinobacteriota | Coriobacteriaceae | NA | NA |
| Pramipexole | 439 | ASV26 | Family | Euryarchaeota | Methanobacteriaceae | NA | NA |
| Pramipexole | 440 | ASV12 | Family | Firmicutes | Veillonellaceae | NA | NA |
| Pramipexole | 441 | ASV31 | Family | Bacteroidota | Rikenellaceae | NA | NA |
| Pramipexole | 442 | ASV55 | Family | Firmicutes | Monoglobaceae | NA | NA |
| Pramipexole | 443 | ASV172 | Family | Proteobacteria | Pasteurellaceae | NA | NA |
| Pramipexole | 444 | ASV16 | Family | Firmicutes | Peptostreptococcaceae | NA | NA |
| Pramipexole | 445 | ASV29 | Family | Firmicutes | Oscillospiraceae | NA | NA |
| Pramipexole | 446 | ASV195 | Family | Bacteroidota | Barnesiellaceae | NA | NA |
| Pramipexole | 447 | ASV39 | Family | Bacteroidota | Tannerellaceae | NA | NA |
| Pramipexole | 448 | ASV28 | Family | Firmicutes | Clostridiaceae | NA | NA |
| Pramipexole | 449 | ASV512 | Family | Proteobacteria | Oxalobacteraceae | NA | NA |

| Ticker | DESeq2_baseMean | DESeq2_log2-FoldChange | DESeq2_lfcSE | DESeq2_stat | DESeq2_pvalue | DESeq2_padj | DESeq2 converged |
|---|---|---|---|---|---|---|---|
| 420 | 83,16336531 | 0,014921875 | 0,853874216 | 0,017475495 | 0,986057282 | 0,994962331 | YES |
| 421 | 7,292472392 | -0,008145482 | 0,853908298 | -0,00953906 | 0,992389047 | 0,994962331 | YES |
| 422 | 0,96555808 | -0,005373857 | 0,851125545 | -0,006313824 | 0,994962331 | 0,994962331 | YES |
| 423 | 1452,852193 | 0,462530538 | 0,197291758 | 2,344398685 | 0,019057788 | 0,483050418 | YES |
| 424 | 184,6983295 | 0,524817141 | 0,242068642 | 2,168050915 | 0,030154813 | 0,483050418 | YES |
| 425 | 117,5343183 | 0,377760088 | 0,17711337 | 2,132871659 | 0,032935256 | 0,483050418 | YES |
| 426 | 1064,304049 | -0,724420652 | 0,409082501 | -1,770842435 | 0,076586907 | 0,687496851 | YES |
| 427 | 127,7645415 | -1,33620113 | 0,7584844 | -1,761672527 | 0,078124642 | 0,687496851 | YES |
| 428 | 468,487708 | 0,462718905 | 0,3290139 | 1,406381024 | 0,159611012 | 0,765344169 | YES |
| 429 | 395,6115501 | 0,273498945 | 0,195188578 | 1,401203637 | 0,161153187 | 0,765344169 | YES |
| 430 | 4,047915222 | 1,0509616 | 0,824730829 | 1,274308614 | 0,202554065 | 0,765344169 | YES |
| 431 | 143,5553921 | 0,588504577 | 0,480709971 | 1,224240419 | 0,220861546 | 0,765344169 | YES |
| 432 | 477,6137919 | 0,391526574 | 0,323607746 | 1,209880106 | 0,226324902 | 0,765344169 | YES |
| 433 | 49,90636662 | -0,31397795 | 0,264923358 | -1,185165218 | 0,235952129 | 0,765344169 | YES |
| 434 | 361,249562 | -0,287116055 | 0,248025863 | -1,157605305 | 0,247025143 | 0,765344169 | YES |
| 435 | 41,71305716 | -0,216996782 | 0,188667756 | -1,150152983 | 0,250080867 | 0,765344169 | YES |
| 436 | 2183,684499 | -0,287428392 | 0,251090076 | -1,144722231 | 0,252324239 | 0,765344169 | YES |
| 437 | 41,06695359 | 0,468315938 | 0,416563608 | 1,124236321 | 0,260912785 | 0,765344169 | YES |
| 438 | 874,8211621 | 0,19013499 | 0,206145046 | 0,922335961 | 0,356353362 | 0,85957437 | YES |
| 439 | 70,49045809 | 0,320297238 | 0,347752182 | 0,921050262 | 0,357024186 | 0,85957437 | YES |
| 440 | 1819,828224 | 0,243146269 | 0,271645021 | 0,895088261 | 0,370739909 | 0,85957437 | YES |
| 441 | 1198,635133 | 0,210502788 | 0,241538963 | 0,871506546 | 0,383477633 | 0,85957437 | YES |
| 442 | 243,3811586 | 0,202866376 | 0,262285642 | 0,773455894 | 0,439252622 | 0,85957437 | YES |
| 443 | 15,2039086 | -0,561428255 | 0,73138181 | -0,767626768 | 0,442708955 | 0,85957437 | YES |
| 444 | 89,80318698 | 0,165189976 | 0,217724798 | 0,758709977 | 0,448026067 | 0,85957437 | YES |
| 445 | 2443,524326 | 0,128859424 | 0,17032636 | 0,756544227 | 0,449322966 | 0,85957437 | YES |
| 446 | 232,0328804 | 0,238321251 | 0,33788922 | 0,70532363 | 0,480608858 | 0,88111624 | YES |
| 447 | 741,2548036 | 0,21695002 | 0,344112094 | 0,630463224 | 0,528391557 | 0,929064567 | YES |
| 448 | 36,31313935 | -0,246591188 | 0,4114858 | -0,599270227 | 0,548992699 | 0,929064567 | YES |
| 449 | 12,30161991 | 0,298329385 | 0,588385076 | 0,507030849 | 0,612133173 | 0,997550357 | YES |

| Medication | Ticker | ASVnr | taxLevel | Phylum | Family | Genus | Species |
|---|---|---|---|---|---|---|---|
| Pramipexole | 450 | ASV2 | Family | Bacteroidota | Bacteroidaceae | NA | NA |
| Pramipexole | 451 | ASV417 | Family | Firmicutes | UCG-010 | NA | NA |
| Pramipexole | 452 | ASV453 | Family | Verrucomicrobiota | Victivallaceae | NA | NA |
| Pramipexole | 453 | ASV752 | Family | Verrucomicrobiota | Puniceicoccaceae | NA | NA |
| Pramipexole | 454 | ASV98 | Family | Firmicutes | Acidaminococcaceae | NA | NA |
| Pramipexole | 455 | ASV4 | Family | Firmicutes | Ruminococcaceae | NA | NA |
| Pramipexole | 456 | ASV449 | Family | Actinobacteriota | Atopobiaceae | NA | NA |
| Pramipexole | 457 | ASV66 | Family | Firmicutes | Butyricicoccaceae | NA | NA |
| Pramipexole | 458 | ASV162 | Family | Desulfobacterota | Desulfovibrionaceae | NA | NA |
| Pramipexole | 459 | ASV703 | Family | Firmicutes | Hydrogenoanaerobacterium | NA | NA |
| Pramipexole | 460 | ASV40 | Family | Firmicutes | Erysipelotrichaceae | NA | NA |
| Pramipexole | 461 | ASV761 | Family | Firmicutes | Defluviitaleaceae | NA | NA |
| Pramipexole | 462 | ASV232 | Family | Firmicutes | Acholeplasmataceae | NA | NA |
| Pramipexole | 463 | ASV7 | Family | Firmicutes | Lachnospiraceae | NA | NA |
| Pramipexole | 464 | ASV72 | Family | Proteobacteria | Sutterellaceae | NA | NA |
| Pramipexole | 465 | ASV1052 | Family | Thermoplasmatota | Methanomethylophilaceae | NA | NA |
| Pramipexole | 466 | ASV454 | Family | Actinobacteriota | Actinomycetaceae | NA | NA |

| Ticker | DESeq2_baseMean | DESeq2_log2-FoldChange | DESeq2_lfcSE | DESeq2_stat | DESeq2_pvalue | DESeq2_padj | DESeq2 converged |
|---|---|---|---|---|---|---|---|
| 450 | 8015,177685 | -0,064037164 | 0,141079158 | -0,453909456 | 0,649893997 | 0,999732848 | YES |
| 451 | 32,48961552 | 0,174705291 | 0,713432803 | 0,244879813 | 0,806549493 | 0,999732848 | YES |
| 452 | 1,73190797 | -0,199863207 | 0,849290708 | -0,235329558 | 0,813952955 | 0,999732848 | YES |
| 453 | 1,060703435 | -0,196157766 | 0,850323469 | -0,230686055 | 0,817558708 | 0,999732848 | YES |
| 454 | 571,4730027 | -0,101312544 | 0,465671051 | -0,21756247 | 0,827770028 | 0,999732848 | YES |
| 455 | 7957,067181 | 0,030132869 | 0,141012269 | 0,213689695 | 0,830789048 | 0,999732848 | YES |
| 456 | 1,36035841 | 0,174593446 | 0,836545039 | 0,208707766 | 0,834676377 | 0,999732848 | YES |
| 457 | 222,8593832 | 0,022961498 | 0,134605385 | 0,170583802 | 0,864551035 | 0,999732848 | YES |
| 458 | 174,7569869 | 0,04933051 | 0,318121603 | 0,155068091 | 0,876767637 | 0,999732848 | YES |
| 459 | 1,256263016 | -0,128756336 | 0,850757672 | -0,151343138 | 0,879705042 | 0,999732848 | YES |
| 460 | 137,2007408 | -0,032048157 | 0,223615634 | -0,143318051 | 0,886039001 | 0,999732848 | YES |
| 461 | 0,690960416 | -0,035635578 | 0,849674821 | -0,041940254 | 0,966546326 | 0,999732848 | YES |
| 462 | 88,39021296 | 0,023085968 | 0,780434886 | 0,029580902 | 0,976401297 | 0,999732848 | YES |
| 463 | 14073,51074 | -0,003959063 | 0,139945779 | -0,02828998 | 0,977430872 | 0,999732848 | YES |
| 464 | 364,3547965 | 0,00293217 | 0,197199277 | 0,014869072 | 0,988136634 | 0,999732848 | YES |
| 465 | 5,867307413 | 0,009680236 | 0,853922444 | 0,011336201 | 0,990955214 | 0,999732848 | YES |
| 466 | 0,556230961 | -0,000284985 | 0,851143884 | -0,000334825 | 0,999732848 | 0,999732848 | YES |

**Supplementary Table 8e: Ropinirole**

| Medication | Ticker | ASVnr | taxLevel | Phylum | Family | Genus | Species |
|---|---|---|---|---|---|---|---|
| Ropinirole | 1 | ASV70 | ASV | Firmicutes | Lachnospiraceae | Roseburia | intestinalis |
| Ropinirole | 2 | ASV118 | ASV | Firmicutes | NA | NA | NA |
| Ropinirole | 3 | ASV201 | ASV | Firmicutes | Oscillospiraceae | UCG-002 | NA |
| Ropinirole | 4 | ASV230 | ASV | Firmicutes | Ruminococcaceae | [Eubacterium] siraeum group | NA |
| Ropinirole | 5 | ASV147 | ASV | Firmicutes | Oscillospiraceae | Oscillibacter | NA |
| Ropinirole | 6 | ASV75 | ASV | Firmicutes | Lachnospiraceae | [Ruminococcus] gauvreauii group | NA |
| Ropinirole | 7 | ASV138 | ASV | Bacteroidota | Rikenellaceae | Alistipes | obesi |
| Ropinirole | 8 | ASV136 | ASV | Firmicutes | Ruminococcaceae | NA | NA |
| Ropinirole | 9 | ASV216 | ASV | Firmicutes | Lachnospiraceae | Lachnospira | pectinoschiza |
| Ropinirole | 10 | ASV96 | ASV | Firmicutes | Lachnospiraceae | CAG-56 | NA |
| Ropinirole | 11 | ASV73 | ASV | Firmicutes | Oscillospiraceae | UCG-005 | NA |
| Ropinirole | 12 | ASV451 | ASV | Firmicutes | Oscillospiraceae | UCG-005 | NA |
| Ropinirole | 13 | ASV60 | ASV | Firmicutes | Ruminococcaceae | [Eubacterium] siraeum group | NA |
| Ropinirole | 14 | ASV481 | ASV | Firmicutes | Lachnospiraceae | [Eubacterium] xylanophilum group | NA |
| Ropinirole | 15 | ASV226 | ASV | Firmicutes | Lachnospiraceae | Lachnospiraceae UCG-001 | NA |
| Ropinirole | 16 | ASV83 | ASV | Firmicutes | Lachnospiraceae | Lachnospiraceae NK4A136 group | NA |
| Ropinirole | 17 | ASV229 | ASV | Firmicutes | Oscillospiraceae | Flavonifractor | plautii |
| Ropinirole | 18 | ASV152 | ASV | Firmicutes | Lachnospiraceae | Roseburia | hominis |
| Ropinirole | 19 | ASV47 | ASV | Bacteroidota | Rikenellaceae | Alistipes | NA |
| Ropinirole | 20 | ASV52 | ASV | Firmicutes | Lachnospiraceae | Coprococcus | eutactus |
| Ropinirole | 21 | ASV192 | ASV | Bacteroidota | Marinifilaceae | Odoribacter | splanchnicus |
| Ropinirole | 22 | ASV557 | ASV | Firmicutes | Oscillospiraceae | UCG-005 | NA |
| Ropinirole | 23 | ASV39 | ASV | Bacteroidota | Tannerellaceae | Parabacteroides | merdae |
| Ropinirole | 24 | ASV34 | ASV | Firmicutes | Lachnospiraceae | Anaerostipes | hadrus |
| Ropinirole | 25 | ASV21 | ASV | Firmicutes | Ruminococcaceae | Faecalibacterium | prausnitzii |
| Ropinirole | 26 | ASV505 | ASV | Bacteroidota | Bacteroidaceae | Bacteroides | nordii |
| Ropinirole | 27 | ASV133 | ASV | Actinobacteriota | Eggerthellaceae | Senegalimassilia | anaerobia |
| Ropinirole | 28 | ASV421 | ASV | Firmicutes | NA | NA | NA |
| Ropinirole | 29 | ASV16 | ASV | Firmicutes | Peptostreptococcaceae | Romboutsia | NA |

| Ticker | DESeq2_baseMean | DESeq2_log2-FoldChange | DESeq2_lfcSE | DESeq2_stat | DESeq2_pvalue | DESeq2_padj | DESeq2 converged |
|---|---|---|---|---|---|---|---|
| 1 | 87,08091928 | -3,562899511 | 0,318803939 | -11,17583276 | 5,35E-29 | 1,55E-26 | YES |
| 2 | 133,3288744 | 1,968888031 | 0,311200883 | 6,326743077 | 2,50E-10 | 3,62E-08 | YES |
| 3 | 45,25323134 | -1,747668758 | 0,318887769 | -5,480513611 | 4,24E-08 | 4,09E-06 | YES |
| 4 | 33,71455881 | -1,728823993 | 0,318893397 | -5,421322643 | 5,92E-08 | 4,27E-06 | YES |
| 5 | 34,97471262 | -0,692507835 | 0,27510609 | -2,5172392 | 0,01182785 | 0,683649722 | YES |
| 6 | 47,46343103 | 0,655561327 | 0,302669142 | 2,165933805 | 0,030316246 | 0,997060958 | YES |
| 7 | 78,51327234 | 0,50254187 | 0,248714248 | 2,020559236 | 0,043325412 | 0,997060958 | YES |
| 8 | 133,4255727 | 0,390135455 | 0,216769613 | 1,799770042 | 0,071896956 | 0,997060958 | YES |
| 9 | 34,31389825 | 0,426669068 | 0,241103724 | 1,769649426 | 0,07678556 | 0,997060958 | YES |
| 10 | 32,54258343 | -0,48137888 | 0,274300777 | -1,754930793 | 0,079271146 | 0,997060958 | YES |
| 11 | 126,6456263 | 0,395391273 | 0,240709126 | 1,642610234 | 0,100463602 | 0,997060958 | YES |
| 12 | 65,99196383 | 0,344769295 | 0,212275859 | 1,624156869 | 0,104342333 | 0,997060958 | YES |
| 13 | 450,5266071 | 0,295895666 | 0,184422496 | 1,604444535 | 0,108616101 | 0,997060958 | YES |
| 14 | 28,75569958 | -0,446136361 | 0,299253638 | -1,490830201 | 0,136006081 | 0,997060958 | YES |
| 15 | 37,99489292 | -0,370950392 | 0,25114101 | -1,477060207 | 0,139659499 | 0,997060958 | YES |
| 16 | 126,9372717 | 0,340563699 | 0,241693506 | 1,409072608 | 0,158813699 | 0,997060958 | YES |
| 17 | 8,325923641 | -0,430471282 | 0,305766193 | -1,407844594 | 0,159177092 | 0,997060958 | YES |
| 18 | 154,8795821 | 0,342161959 | 0,249115834 | 1,373505464 | 0,169595264 | 0,997060958 | YES |
| 19 | 76,84794037 | -0,271299397 | 0,206931846 | -1,311056771 | 0,189838584 | 0,997060958 | YES |
| 20 | 29,2088876 | 0,396197662 | 0,304713892 | 1,300228419 | 0,193522693 | 0,997060958 | YES |
| 21 | 19,62201318 | -0,281988135 | 0,218768716 | -1,288978333 | 0,197405619 | 0,997060958 | YES |
| 22 | 21,81058383 | 0,38421406 | 0,307775357 | 1,248358749 | 0,211899708 | 0,997060958 | YES |
| 23 | 147,9798323 | -0,266132321 | 0,215075922 | -1,237387794 | 0,215943148 | 0,997060958 | YES |
| 24 | 29,58458004 | 0,286778448 | 0,240278952 | 1,193522964 | 0,232664624 | 0,997060958 | YES |
| 25 | 90,15691841 | -0,242009776 | 0,209089211 | -1,157447459 | 0,247089594 | 0,997060958 | YES |
| 26 | 5,722969127 | 0,359277145 | 0,311777879 | 1,152349697 | 0,249177403 | 0,997060958 | YES |
| 27 | 14,24401045 | 0,254330144 | 0,221048645 | 1,15056188 | 0,249912523 | 0,997060958 | YES |
| 28 | 12,97105206 | 0,361784207 | 0,315924221 | 1,145161349 | 0,252142326 | 0,997060958 | YES |
| 29 | 49,26327331 | 0,24166693 | 0,212668497 | 1,136355092 | 0,255807983 | 0,997060958 | YES |

| Medication | Ticker | ASVnr | taxLevel | Phylum | Family | Genus | Species |
|---|---|---|---|---|---|---|---|
| Ropinirole | 30 | ASV132 | ASV | Firmicutes | Ruminococcaceae | CAG-352 | NA |
| Ropinirole | 31 | ASV149 | ASV | Firmicutes | Oscillospiraceae | UCG-002 | NA |
| Ropinirole | 32 | ASV258 | ASV | Firmicutes | Lachnospiraceae | Lachnospiraceae AC2044 group | NA |
| Ropinirole | 33 | ASV357 | ASV | Firmicutes | Lachnospiraceae | Marvinbryantia | NA |
| Ropinirole | 34 | ASV109 | ASV | Firmicutes | [Eubacterium] coprostanoligenes group | NA | NA |
| Ropinirole | 35 | ASV134 | ASV | Actinobacteriota | Eggerthellaceae | Enterorhabdus | NA |
| Ropinirole | 36 | ASV233 | ASV | Firmicutes | [Eubacterium] coprostanoligenes group | NA | NA |
| Ropinirole | 37 | ASV139 | ASV | Firmicutes | Christensenellaceae | Christensenellaceae R-7 group | NA |
| Ropinirole | 38 | ASV19 | ASV | Firmicutes | Erysipelatoclostridiaceae | Erysipelotrichaceae UCG-003 | bacterium |
| Ropinirole | 39 | ASV125 | ASV | Firmicutes | Oscillospiraceae | UCG-002 | NA |
| Ropinirole | 40 | ASV432 | ASV | Bacteroidota | Marinifilaceae | Butyricimonas | virosa |
| Ropinirole | 41 | ASV127 | ASV | Firmicutes | Lachnospiraceae | Lachnospiraceae NK4A136 group | bacterium |
| Ropinirole | 42 | ASV200 | ASV | Firmicutes | Lachnospiraceae | [Eubacterium] xylanophilum group | NA |
| Ropinirole | 43 | ASV383 | ASV | Firmicutes | NA | NA | NA |
| Ropinirole | 44 | ASV49 | ASV | Bacteroidota | Bacteroidaceae | Bacteroides | NA |
| Ropinirole | 45 | ASV17 | ASV | Actinobacteriota | Coriobacteriaceae | Collinsella | aerofaciens |
| Ropinirole | 46 | ASV382 | ASV | Firmicutes | NA | NA | NA |
| Ropinirole | 47 | ASV522 | ASV | Firmicutes | UCG-010 | NA | NA |
| Ropinirole | 48 | ASV362 | ASV | Firmicutes | Ruminococcaceae | NA | NA |
| Ropinirole | 49 | ASV91 | ASV | Firmicutes | Lachnospiraceae | [Eubacterium] eligens group | NA |
| Ropinirole | 50 | ASV144 | ASV | Desulfobacterota | Desulfovibrionaceae | Bilophila | wadsworthia |
| Ropinirole | 51 | ASV687 | ASV | Firmicutes | Ruminococcaceae | DTU089 | NA |
| Ropinirole | 52 | ASV320 | ASV | Firmicutes | Christensenellaceae | Christensenellaceae R-7 group | NA |
| Ropinirole | 53 | ASV131 | ASV | Firmicutes | Lachnospiraceae | NA | NA |
| Ropinirole | 54 | ASV403 | ASV | Firmicutes | Oscillospiraceae | Colidextribacter | NA |
| Ropinirole | 55 | ASV161 | ASV | Firmicutes | Oscillospiraceae | UCG-002 | NA |
| Ropinirole | 56 | ASV189 | ASV | Firmicutes | Ruminococcaceae | Ruminococcus | champanellensis |
| Ropinirole | 57 | ASV76 | ASV | Firmicutes | Lachnospiraceae | Lachnospiraceae ND3007 group | NA |
| Ropinirole | 58 | ASV88 | ASV | Firmicutes | Lachnospiraceae | Dorea | formicigenerans |
| Ropinirole | 59 | ASV81 | ASV | Firmicutes | Lachnospiraceae | Lachnospiraceae NK4A136 group | NA |

| Ticker | DESeq2_baseMean | DESeq2_log2-FoldChange | DESeq2_lfcSE | DESeq2_stat | DESeq2_pvalue | DESeq2_padj | DESeq2 converged |
|---|---|---|---|---|---|---|---|
| 30 | 11,91415152 | 0,359049007 | 0,316920303 | 1,132931539 | 0,257243004 | 0,997060958 | YES |
| 31 | 111,5309248 | 0,233502436 | 0,207859803 | 1,123365043 | 0,26128249 | 0,997060958 | YES |
| 32 | 10,37684165 | 0,356004897 | 0,317587621 | 1,12096591 | 0,262302373 | 0,997060958 | YES |
| 33 | 8,359122438 | 0,350938375 | 0,318727545 | 1,101060705 | 0,270870238 | 0,997060958 | YES |
| 34 | 29,90846617 | 0,283430583 | 0,259629606 | 1,091672816 | 0,274976938 | 0,997060958 | YES |
| 35 | 9,290547146 | -0,275989366 | 0,259442412 | -1,063778908 | 0,287428869 | 0,997060958 | YES |
| 36 | 7,57225385 | -0,32526925 | 0,309482505 | -1,051010135 | 0,293253935 | 0,997060958 | YES |
| 37 | 6,149239495 | 0,335008283 | 0,318985666 | 1,050229894 | 0,293612429 | 0,997060958 | YES |
| 38 | 7,958833976 | 0,311126042 | 0,300887781 | 1,034026844 | 0,301123614 | 0,997060958 | YES |
| 39 | 5,572748292 | 0,32948839 | 0,318996002 | 1,032891909 | 0,301654486 | 0,997060958 | YES |
| 40 | 12,10162028 | 0,262058705 | 0,254731697 | 1,028763628 | 0,303590762 | 0,997060958 | YES |
| 41 | 450,6191727 | -0,251558102 | 0,247221713 | -1,017540483 | 0,308896384 | 0,997060958 | YES |
| 42 | 36,92033723 | -0,25483087 | 0,25667323 | -0,992822157 | 0,32079663 | 0,997060958 | YES |
| 43 | 4,035438419 | 0,311444633 | 0,319036756 | 0,976202983 | 0,328963885 | 0,997060958 | YES |
| 44 | 156,0403863 | -0,250536138 | 0,259661908 | -0,964855185 | 0,334617351 | 0,997060958 | YES |
| 45 | 68,43117881 | -0,204078311 | 0,212435434 | -0,960660411 | 0,336722943 | 0,997060958 | YES |
| 46 | 3,362865349 | 0,301291537 | 0,319065284 | 0,944294325 | 0,345019265 | 0,997060958 | YES |
| 47 | 3,362865349 | 0,301291537 | 0,319065284 | 0,944294325 | 0,345019265 | 0,997060958 | YES |
| 48 | 25,97374053 | -0,195839338 | 0,211423853 | -0,92628781 | 0,354296429 | 0,997060958 | YES |
| 49 | 39,02358076 | -0,225518135 | 0,245424803 | -0,91888893 | 0,358153671 | 0,997060958 | YES |
| 50 | 26,14121155 | -0,217885791 | 0,237729389 | -0,916528629 | 0,359389699 | 0,997060958 | YES |
| 51 | 16,64121602 | 0,239949909 | 0,263028483 | 0,912258274 | 0,361632774 | 0,997060958 | YES |
| 52 | 6,25502432 | 0,278062671 | 0,306122325 | 0,908338425 | 0,363699447 | 0,997060958 | YES |
| 53 | 242,7791617 | -0,187490063 | 0,206510982 | -0,90789391 | 0,363934276 | 0,997060958 | YES |
| 54 | 2,498128545 | 0,284802385 | 0,319122448 | 0,892454877 | 0,372149168 | 0,997060958 | YES |
| 55 | 27,96510718 | -0,215677022 | 0,243439461 | -0,885957525 | 0,375640416 | 0,997060958 | YES |
| 56 | 564,9063736 | -0,187778183 | 0,216607317 | -0,866905997 | 0,385993517 | 0,997060958 | YES |
| 57 | 40,87751298 | -0,177955437 | 0,206385835 | -0,862246371 | 0,388551957 | 0,997060958 | YES |
| 58 | 42,0828179 | -0,220776697 | 0,257349288 | -0,857887347 | 0,390954672 | 0,997060958 | YES |
| 59 | 334,3969195 | 0,188219798 | 0,219762083 | 0,85647076 | 0,39173744 | 0,997060958 | YES |

| Medication | Ticker | ASVnr | taxLevel | Phylum | Family | Genus | Species |
|---|---|---|---|---|---|---|---|
| Ropinirole | 60 | ASV554 | ASV | Firmicutes | Lachnospiraceae | NA | NA |
| Ropinirole | 61 | ASV441 | ASV | Firmicutes | [Eubacterium] coprostanoligenes group | NA | NA |
| Ropinirole | 62 | ASV417 | ASV | Firmicutes | UCG-010 | NA | NA |
| Ropinirole | 63 | ASV159 | ASV | Firmicutes | Lachnospiraceae | Roseburia | NA |
| Ropinirole | 64 | ASV423 | ASV | Firmicutes | Christensenellaceae | Christensenellaceae R-7 group | NA |
| Ropinirole | 65 | ASV202 | ASV | Firmicutes | Lachnospiraceae | [Eubacterium] hallii group | NA |
| Ropinirole | 66 | ASV314 | ASV | Firmicutes | Ruminococcaceae | Subdoligranulum | NA |
| Ropinirole | 67 | ASV53 | ASV | Firmicutes | Lachnospiraceae | [Ruminococcus] torques group | NA |
| Ropinirole | 68 | ASV587 | ASV | Firmicutes | Anaerovoracaceae | Family XIII AD3011 group | NA |
| Ropinirole | 69 | ASV46 | ASV | Bacteroidota | Bacteroidaceae | Bacteroides | NA |
| Ropinirole | 70 | ASV257 | ASV | Bacteroidota | Bacteroidaceae | Bacteroides | ovatus |
| Ropinirole | 71 | ASV9 | ASV | Bacteroidota | Bacteroidaceae | Bacteroides | NA |
| Ropinirole | 72 | ASV84 | ASV | Bacteroidota | Bacteroidaceae | Bacteroides | NA |
| Ropinirole | 73 | ASV62 | ASV | Firmicutes | NA | NA | NA |
| Ropinirole | 74 | ASV154 | ASV | Firmicutes | NA | NA | NA |
| Ropinirole | 75 | ASV43 | ASV | Firmicutes | Lachnospiraceae | Blautia | faecis |
| Ropinirole | 76 | ASV26 | ASV | Euryarchaeota | Methanobacteriaceae | Methanobrevibacter | NA |
| Ropinirole | 77 | ASV10 | ASV | Verrucomicrobiota | Akkermansiaceae | Akkermansia | muciniphila |
| Ropinirole | 78 | ASV343 | ASV | Firmicutes | Lachnospiraceae | NA | NA |
| Ropinirole | 79 | ASV41 | ASV | Firmicutes | Ruminococcaceae | Ruminococcus | bicirculans |
| Ropinirole | 80 | ASV214 | ASV | Firmicutes | Lachnospiraceae | Lachnospiraceae UCG-010 | NA |
| Ropinirole | 81 | ASV119 | ASV | Firmicutes | Ruminococcaceae | Subdoligranulum | NA |
| Ropinirole | 82 | ASV64 | ASV | Firmicutes | Oscillospiraceae | UCG-002 | NA |
| Ropinirole | 83 | ASV521 | ASV | Firmicutes | Lachnospiraceae | [Eubacterium] ventriosum group | NA |
| Ropinirole | 84 | ASV143 | ASV | Firmicutes | Peptostreptococcaceae | Terrisporobacter | mayombei |
| Ropinirole | 85 | ASV690 | ASV | Firmicutes | Ruminococcaceae | Fournierella | NA |
| Ropinirole | 86 | ASV443 | ASV | Firmicutes | Veillonellaceae | Veillonella | NA |
| Ropinirole | 87 | ASV203 | ASV | Firmicutes | NA | NA | NA |
| Ropinirole | 88 | ASV512 | ASV | Proteobacteria | Oxalobacteraceae | Oxalobacter | formigenes |
| Ropinirole | 89 | ASV253 | ASV | Bacteroidota | Tannerellaceae | Parabacteroides | distasonis |

| Ticker | DESeq2_baseMean | DESeq2_log2-FoldChange | DESeq2_lfcSE | DESeq2_stat | DESeq2_pvalue | DESeq2_padj | DESeq2 converged |
|---|---|---|---|---|---|---|---|
| 60 | 2,017719209 | 0,273006613 | 0,319173391 | 0,855355181 | 0,392354548 | 0,997060958 | YES |
| 61 | 26,38571486 | -0,208215322 | 0,247455151 | -0,841426502 | 0,400109044 | 0,997060958 | YES |
| 62 | 1,633391741 | 0,26138091 | 0,319233676 | 0,81877612 | 0,412914158 | 0,997060958 | YES |
| 63 | 106,2770123 | -0,196717323 | 0,242357016 | -0,811684045 | 0,416972952 | 0,997060958 | YES |
| 64 | 1,411867977 | 0,257283351 | 0,31874037 | 0,807187841 | 0,41955827 | 0,997060958 | YES |
| 65 | 1,441228007 | 0,254516904 | 0,319274759 | 0,797172018 | 0,425351135 | 0,997060958 | YES |
| 66 | 1,441228007 | 0,254516904 | 0,319274759 | 0,797172018 | 0,425351135 | 0,997060958 | YES |
| 67 | 124,8641022 | -0,163452808 | 0,206403687 | -0,791908374 | 0,428414104 | 0,997060958 | YES |
| 68 | 1,983209342 | -0,24593354 | 0,312599518 | -0,786736786 | 0,431435965 | 0,997060958 | YES |
| 69 | 271,7800089 | -0,139358142 | 0,177306259 | -0,785974184 | 0,431882612 | 0,997060958 | YES |
| 70 | 67,59191608 | -0,238816914 | 0,306036752 | -0,780353708 | 0,435182708 | 0,997060958 | YES |
| 71 | 1396,578398 | -0,138028953 | 0,177473466 | -0,777744168 | 0,436719849 | 0,997060958 | YES |
| 72 | 168,508339 | -0,203926042 | 0,262727478 | -0,77618848 | 0,437637707 | 0,997060958 | YES |
| 73 | 31,31206803 | -0,233699277 | 0,308491992 | -0,757553789 | 0,448718153 | 0,997060958 | YES |
| 74 | 29,0539862 | -0,233148722 | 0,308941494 | -0,754669497 | 0,450447313 | 0,997060958 | YES |
| 75 | 222,9423771 | 0,186223042 | 0,24749016 | 0,752446246 | 0,451782745 | 0,997060958 | YES |
| 76 | 101,9192515 | -0,158223285 | 0,210586428 | -0,751346071 | 0,452444409 | 0,997060958 | YES |
| 77 | 216,0470614 | 0,190612693 | 0,254207766 | 0,749830329 | 0,4533569 | 0,997060958 | YES |
| 78 | 27,80701959 | -0,184517032 | 0,248318281 | -0,743066643 | 0,457441334 | 0,997060958 | YES |
| 79 | 167,9029641 | -0,191204148 | 0,257860526 | -0,741502205 | 0,458388995 | 0,997060958 | YES |
| 80 | 85,24482502 | -0,176259022 | 0,238714847 | -0,738366399 | 0,460291828 | 0,997060958 | YES |
| 81 | 20,17219767 | -0,229983323 | 0,311931166 | -0,7372887 | 0,460946803 | 0,997060958 | YES |
| 82 | 261,6859641 | 0,159745618 | 0,217276308 | 0,735218763 | 0,462206274 | 0,997060958 | YES |
| 83 | 15,05387886 | -0,226836627 | 0,315524515 | -0,718919185 | 0,472190715 | 0,997060958 | YES |
| 84 | 14,4517237 | -0,226354382 | 0,316125459 | -0,716027058 | 0,473974641 | 0,997060958 | YES |
| 85 | 0,864736804 | 0,226687693 | 0,319494667 | 0,709519491 | 0,478002161 | 0,997060958 | YES |
| 86 | 8,881788527 | -0,220275328 | 0,318177617 | -0,692303029 | 0,488747048 | 0,997060958 | YES |
| 87 | 0,768654937 | 0,220316565 | 0,319559886 | 0,689437486 | 0,490548001 | 0,997060958 | YES |
| 88 | 4,166803908 | 0,201651722 | 0,292823389 | 0,688646225 | 0,491045924 | 0,997060958 | YES |
| 89 | 6,322629121 | -0,215036432 | 0,319487037 | -0,673067785 | 0,500904168 | 0,997060958 | YES |

| Medication | Ticker | ASVnr | taxLevel | Phylum | Family | Genus | Species |
|---|---|---|---|---|---|---|---|
| Ropinirole | 90 | ASV268 | ASV | Firmicutes | Ruminococcaceae | Incertae Sedis | NA |
| Ropinirole | 91 | ASV351 | ASV | Firmicutes | Lachnospiraceae | Roseburia | NA |
| Ropinirole | 92 | ASV243 | ASV | Firmicutes | NA | NA | NA |
| Ropinirole | 93 | ASV386 | ASV | Firmicutes | Ruminococcaceae | DTU089 | NA |
| Ropinirole | 94 | ASV89 | ASV | Firmicutes | Lachnospiraceae | Blautia | NA |
| Ropinirole | 95 | ASV303 | ASV | Firmicutes | Lachnospiraceae | GCA-900066575 | NA |
| Ropinirole | 96 | ASV36 | ASV | Firmicutes | Lachnospiraceae | Blautia | obeum |
| Ropinirole | 97 | ASV1086 | ASV | Firmicutes | UCG-010 | NA | NA |
| Ropinirole | 98 | ASV191 | ASV | Firmicutes | Lachnospiraceae | Lachnospiraceae NK4A136 group | NA |
| Ropinirole | 99 | ASV509 | ASV | Firmicutes | Lachnospiraceae | Marvinbryantia | NA |
| Ropinirole | 100 | ASV337 | ASV | Firmicutes | Lachnospiraceae | Lachnoclostridium | NA |
| Ropinirole | 101 | ASV435 | ASV | Firmicutes | Christensenellaceae | Christensenellaceae R-7 group | NA |
| Ropinirole | 102 | ASV529 | ASV | Firmicutes | Lachnospiraceae | NA | NA |
| Ropinirole | 103 | ASV56 | ASV | Bacteroidota | Bacteroidaceae | Bacteroides | massiliensis |
| Ropinirole | 104 | ASV8 | ASV | Firmicutes | Ruminococcaceae | Faecalibacterium | prausnitzii |
| Ropinirole | 105 | ASV30 | ASV | Proteobacteria | Enterobacteriaceae | Escherichia-Shigella | NA |
| Ropinirole | 106 | ASV65 | ASV | Firmicutes | Ruminococcaceae | Incertae Sedis | NA |
| Ropinirole | 107 | ASV449 | ASV | Actinobacteriota | Atopobiaceae | Olsenella | NA |
| Ropinirole | 108 | ASV120 | ASV | Firmicutes | Lachnospiraceae | Anaerostipes | hadrus |
| Ropinirole | 109 | ASV221 | ASV | Firmicutes | Oscillospiraceae | Intestinimonas | NA |
| Ropinirole | 110 | ASV80 | ASV | Firmicutes | Acidaminococcaceae | Phascolarctobacterium | faecium |
| Ropinirole | 111 | ASV283 | ASV | Actinobacteriota | Eggerthellaceae | Slackia | isoflavoniconvertens |
| Ropinirole | 112 | ASV48 | ASV | Bacteroidota | Bacteroidaceae | Bacteroides | caccae |
| Ropinirole | 113 | ASV675 | ASV | Bacteroidota | Prevotellaceae | NA | NA |
| Ropinirole | 114 | ASV146 | ASV | Firmicutes | Erysipelotrichaceae | Turicibacter | sanguinis |
| Ropinirole | 115 | ASV7 | ASV | Firmicutes | Lachnospiraceae | Agathobacter | NA |
| Ropinirole | 116 | ASV193 | ASV | Actinobacteriota | Eggerthellaceae | Adlercreutzia | equolifaciens |
| Ropinirole | 117 | ASV436 | ASV | Firmicutes | Christensenellaceae | Christensenellaceae R-7 group | NA |
| Ropinirole | 118 | ASV33 | ASV | Firmicutes | Peptostreptococcaceae | Intestinibacter | bartlettii |
| Ropinirole | 119 | ASV328 | ASV | Firmicutes | Anaerovoracaceae | [Eubacterium] brachy group | NA |

| Ticker | DESeq2_baseMean | DESeq2_log2-FoldChange | DESeq2_lfcSE | DESeq2_stat | DESeq2_pvalue | DESeq2_padj | DESeq2 converged |
|---|---|---|---|---|---|---|---|
| 90 | 6,172090332 | -0,214631965 | 0,319590622 | -0,671584054 | 0,501848533 | 0,997060958 | YES |
| 91 | 53,01676487 | 0,179349394 | 0,267657465 | 0,670070586 | 0,502812794 | 0,997060958 | YES |
| 92 | 5,268857601 | -0,211868439 | 0,320308256 | -0,661451695 | 0,508322683 | 0,997060958 | YES |
| 93 | 4,666702446 | -0,209239837 | 0,320846311 | -0,652149737 | 0,514304581 | 0,997060958 | YES |
| 94 | 59,54103669 | -0,191981378 | 0,29547562 | -0,649736779 | 0,515862262 | 0,997060958 | YES |
| 95 | 6,437801 | 0,192132581 | 0,305674154 | 0,628553569 | 0,529641366 | 0,997060958 | YES |
| 96 | 98,89352133 | -0,128974764 | 0,206832112 | -0,623572245 | 0,532908543 | 0,997060958 | YES |
| 97 | 0,480409336 | 0,195086232 | 0,319891972 | 0,609850354 | 0,541960942 | 0,997060958 | YES |
| 98 | 163,0389885 | 0,128069245 | 0,210328487 | 0,608901089 | 0,542590003 | 0,997060958 | YES |
| 99 | 10,92677978 | -0,176347328 | 0,306184082 | -0,575951977 | 0,564647642 | 0,997060958 | YES |
| 100 | 2,709698195 | -0,181508905 | 0,320955436 | -0,565526814 | 0,571715492 | 0,997060958 | YES |
| 101 | 2,709698195 | -0,181508905 | 0,320955436 | -0,565526814 | 0,571715492 | 0,997060958 | YES |
| 102 | 2,408620617 | -0,175553902 | 0,320986051 | -0,546920659 | 0,584433242 | 0,997060958 | YES |
| 103 | 31,16965165 | -0,12551468 | 0,229724391 | -0,546370715 | 0,584811136 | 0,997060958 | YES |
| 104 | 767,4170769 | -0,098327726 | 0,180494962 | -0,544767153 | 0,585913673 | 0,997060958 | YES |
| 105 | 105,8925823 | -0,142002468 | 0,261673714 | -0,542669975 | 0,58735705 | 0,997060958 | YES |
| 106 | 111,8646858 | 0,113728461 | 0,215917116 | 0,526722767 | 0,598386123 | 0,997060958 | YES |
| 107 | 3,719904428 | 0,165742175 | 0,319188773 | 0,519260666 | 0,603578979 | 0,997060958 | YES |
| 108 | 1,957004252 | -0,165106046 | 0,321047511 | -0,514272935 | 0,607061181 | 0,997060958 | YES |
| 109 | 46,64306583 | 0,108321966 | 0,213807945 | 0,50663209 | 0,612412988 | 0,997060958 | YES |
| 110 | 1,655926674 | -0,156749366 | 0,321104795 | -0,488156416 | 0,625439053 | 0,997060958 | YES |
| 111 | 1,655926674 | -0,156749366 | 0,321104795 | -0,488156416 | 0,625439053 | 0,997060958 | YES |
| 112 | 203,6058767 | -0,108741925 | 0,228182562 | -0,476556683 | 0,633677835 | 0,997060958 | YES |
| 113 | 139,9907637 | 0,121542464 | 0,264864141 | 0,45888607 | 0,646315983 | 0,997060958 | YES |
| 114 | 10,87009923 | -0,139161934 | 0,309959606 | -0,448967969 | 0,653454764 | 0,997060958 | YES |
| 115 | 649,4738201 | -0,097022471 | 0,216140001 | -0,448887159 | 0,653513059 | 0,997060958 | YES |
| 116 | 7,048892715 | -0,12362502 | 0,288222592 | -0,428922032 | 0,667979966 | 0,997060958 | YES |
| 117 | 2,687163261 | 0,127326873 | 0,30513406 | 0,417281746 | 0,676472339 | 0,997060958 | YES |
| 118 | 45,61025965 | -0,109212994 | 0,2634112 | -0,414610288 | 0,678427215 | 0,997060958 | YES |
| 119 | 2,391027292 | -0,131091276 | 0,320243718 | -0,40934847 | 0,68228395 | 0,997060958 | YES |

| Medication | Ticker | ASVnr | taxLevel | Phylum | Family | Genus | Species |
|---|---|---|---|---|---|---|---|
| Ropinirole | 120 | ASV329 | ASV | Firmicutes | Lachnospiraceae | NA | NA |
| Ropinirole | 121 | ASV66 | ASV | Firmicutes | Butyricicoccaceae | Butyricicoccus | NA |
| Ropinirole | 122 | ASV270 | ASV | Firmicutes | Christensenellaceae | Christensenellaceae R-7 group | NA |
| Ropinirole | 123 | ASV29 | ASV | Firmicutes | Oscillospiraceae | UCG-002 | NA |
| Ropinirole | 124 | ASV322 | ASV | Firmicutes | Anaerovoracaceae | Family XIII AD3011 group | NA |
| Ropinirole | 125 | ASV142 | ASV | Firmicutes | Ruminococcaceae | Faecalibacterium | prausnitzii |
| Ropinirole | 126 | ASV32 | ASV | Firmicutes | Oscillospiraceae | UCG-002 | NA |
| Ropinirole | 127 | ASV68 | ASV | Firmicutes | Ruminococcaceae | Faecalibacterium | NA |
| Ropinirole | 128 | ASV31 | ASV | Bacteroidota | Rikenellaceae | Alistipes | putredinis |
| Ropinirole | 129 | ASV85 | ASV | Firmicutes | Lachnospiraceae | Coprococcus | NA |
| Ropinirole | 130 | ASV306 | ASV | Firmicutes | Veillonellaceae | Veillonella | NA |
| Ropinirole | 131 | ASV160 | ASV | Firmicutes | Ruminococcaceae | Ruminococcus | callidus |
| Ropinirole | 132 | ASV59 | ASV | Firmicutes | [Eubacterium] coprostanoligenes group | NA | NA |
| Ropinirole | 133 | ASV533 | ASV | Firmicutes | Ruminococcaceae | Negativibacillus | massiliensis |
| Ropinirole | 134 | ASV153 | ASV | Firmicutes | Streptococcaceae | Streptococcus | NA |
| Ropinirole | 135 | ASV87 | ASV | Proteobacteria | Sutterellaceae | Sutterella | wadsworthensis |
| Ropinirole | 136 | ASV393 | ASV | Bacteroidota | Rikenellaceae | Alistipes | indistinctus |
| Ropinirole | 137 | ASV695 | ASV | Firmicutes | UCG-010 | NA | NA |
| Ropinirole | 138 | ASV222 | ASV | Firmicutes | Oscillospiraceae | NK4A214 group | NA |
| Ropinirole | 139 | ASV55 | ASV | Firmicutes | Monoglobaceae | Monoglobus | NA |
| Ropinirole | 140 | ASV438 | ASV | Firmicutes | UCG-010 | NA | NA |
| Ropinirole | 141 | ASV28 | ASV | Firmicutes | Clostridiaceae | Clostridium sensu stricto 1 | NA |
| Ropinirole | 142 | ASV175 | ASV | Firmicutes | NA | NA | NA |
| Ropinirole | 143 | ASV111 | ASV | Firmicutes | Lachnospiraceae | Lachnospira | pectinoschiza |
| Ropinirole | 144 | ASV104 | ASV | Firmicutes | Christensenellaceae | Christensenellaceae R-7 group | NA |
| Ropinirole | 145 | ASV425 | ASV | Firmicutes | Oscillospiraceae | NK4A214 group | NA |
| Ropinirole | 146 | ASV172 | ASV | Proteobacteria | Pasteurellaceae | Haemophilus | NA |
| Ropinirole | 147 | ASV54 | ASV | Bacteroidota | Bacteroidaceae | Bacteroides | NA |
| Ropinirole | 148 | ASV44 | ASV | Bacteroidota | Bacteroidaceae | Bacteroides | NA |
| Ropinirole | 149 | ASV51 | ASV | Firmicutes | Oscillospiraceae | UCG-005 | NA |

| Ticker | DESeq2_baseMean | DESeq2_log2-FoldChange | DESeq2_lfcSE | DESeq2_stat | DESeq2_pvalue | DESeq2_padj | DESeq2 converged |
|---|---|---|---|---|---|---|---|
| 120 | 3,034240331 | -0,130787691 | 0,320173041 | -0,408490641 | 0,6829135 | 0,997060958 | YES |
| 121 | 113,1693386 | 0,070421005 | 0,178220534 | 0,395134068 | 0,692743948 | 0,997060958 | YES |
| 122 | 26,57182091 | -0,091913425 | 0,233858006 | -0,393030911 | 0,694296651 | 0,997060958 | YES |
| 123 | 832,7630492 | 0,069707734 | 0,178255278 | 0,391055654 | 0,695756099 | 0,997060958 | YES |
| 124 | 22,60629182 | 0,094048213 | 0,241425373 | 0,389553974 | 0,696866392 | 0,997060958 | YES |
| 125 | 108,3279621 | -0,104565218 | 0,269352473 | -0,388209607 | 0,697860924 | 0,997060958 | YES |
| 126 | 1048,595926 | -0,070197844 | 0,182924811 | -0,383752448 | 0,701161936 | 0,997060958 | YES |
| 127 | 98,9610722 | -0,080010994 | 0,211507765 | -0,378288684 | 0,705216151 | 0,997060958 | YES |
| 128 | 596,6671214 | -0,066926352 | 0,179129301 | -0,373620349 | 0,708686792 | 0,997060958 | YES |
| 129 | 122,0142076 | 0,116806268 | 0,313892671 | 0,37212168 | 0,709802256 | 0,997060958 | YES |
| 130 | 120,6733921 | 0,116804657 | 0,313893424 | 0,372115654 | 0,709806742 | 0,997060958 | YES |
| 131 | 75,75607392 | 0,116724333 | 0,313908757 | 0,371841596 | 0,710010792 | 0,997060958 | YES |
| 132 | 44,24691043 | 0,116558916 | 0,314011591 | 0,371193038 | 0,710493759 | 0,997060958 | YES |
| 133 | 12,73774694 | 0,115636914 | 0,314469303 | 0,367720831 | 0,713081406 | 0,997060958 | YES |
| 134 | 107,4417965 | -0,096849125 | 0,264767105 | -0,365789868 | 0,714521881 | 0,997060958 | YES |
| 135 | 18,95763385 | 0,085697732 | 0,238240993 | 0,359710269 | 0,719063812 | 0,997060958 | YES |
| 136 | 6,750517999 | 0,101078324 | 0,281545102 | 0,359012901 | 0,719585438 | 0,997060958 | YES |
| 137 | 4,022446403 | 0,11298752 | 0,315827234 | 0,357751036 | 0,720529636 | 0,997060958 | YES |
| 138 | 84,44170634 | 0,080399303 | 0,225250578 | 0,356932726 | 0,721142169 | 0,997060958 | YES |
| 139 | 98,03847048 | -0,072002802 | 0,201939758 | -0,356555851 | 0,721424334 | 0,997060958 | YES |
| 140 | 58,50497648 | 0,07878317 | 0,22412239 | 0,351518515 | 0,725199384 | 0,997060958 | YES |
| 141 | 74,57984974 | -0,078917473 | 0,228405592 | -0,345514625 | 0,729707517 | 0,997060958 | YES |
| 142 | 2,011223201 | 0,109432341 | 0,317729494 | 0,34441984 | 0,73053057 | 0,997060958 | YES |
| 143 | 28,63969775 | 0,080328194 | 0,2374139 | 0,338346637 | 0,735101983 | 0,997060958 | YES |
| 144 | 71,77510376 | -0,087976254 | 0,270351964 | -0,325413779 | 0,744867928 | 0,997060958 | YES |
| 145 | 61,49209071 | -0,064960782 | 0,212847136 | -0,305199229 | 0,76021442 | 0,997060958 | YES |
| 146 | 69,915044 | 0,080970512 | 0,272737132 | 0,296881146 | 0,766557251 | 0,997060958 | YES |
| 147 | 112,8103918 | 0,066025752 | 0,222615205 | 0,296591384 | 0,76677849 | 0,997060958 | YES |
| 148 | 350,1142862 | -0,079171956 | 0,267766804 | -0,295675022 | 0,767478274 | 0,997060958 | YES |
| 149 | 223,8674951 | 0,052269881 | 0,178339941 | 0,293091277 | 0,769452384 | 0,997060958 | YES |

| Medication | Ticker | ASVnr | taxLevel | Phylum | Family | Genus | Species |
|---|---|---|---|---|---|---|---|
| Ropinirole | 150 | ASV158 | ASV | Firmicutes | Lachnospiraceae | [Eubacterium] ventriosum group | NA |
| Ropinirole | 151 | ASV309 | ASV | Firmicutes | Oscillospiraceae | Colidextribacter | massiliensis |
| Ropinirole | 152 | ASV247 | ASV | Euryarchaeota | Methanobacteriaceae | Methanosphaera | NA |
| Ropinirole | 153 | ASV517 | ASV | Firmicutes | Lachnospiraceae | Blautia | NA |
| Ropinirole | 154 | ASV387 | ASV | Firmicutes | Lachnospiraceae | NA | NA |
| Ropinirole | 155 | ASV128 | ASV | Firmicutes | Oscillospiraceae | NK4A214 group | NA |
| Ropinirole | 156 | ASV145 | ASV | Bacteroidota | Muribaculaceae | NA | NA |
| Ropinirole | 157 | ASV260 | ASV | Firmicutes | Lactobacillaceae | Lactobacillus | delbrueckii |
| Ropinirole | 158 | ASV12 | ASV | Firmicutes | Veillonellaceae | Dialister | invisus |
| Ropinirole | 159 | ASV14 | ASV | Firmicutes | Lachnospiraceae | Blautia | massiliensis |
| Ropinirole | 160 | ASV538 | ASV | Bacteroidota | Marinifilaceae | Butyricimonas | NA |
| Ropinirole | 161 | ASV194 | ASV | Bacteroidota | Bacteroidaceae | Bacteroides | intestinalis |
| Ropinirole | 162 | ASV353 | ASV | Firmicutes | Lachnospiraceae | [Eubacterium] xylanophilum group | NA |
| Ropinirole | 163 | ASV92 | ASV | Firmicutes | Erysipelotrichaceae | Turicibacter | sanguinis |
| Ropinirole | 164 | ASV3 | ASV | Firmicutes | Ruminococcaceae | Subdoligranulum | NA |
| Ropinirole | 165 | ASV261 | ASV | Firmicutes | Anaerovoracaceae | Family XIII UCG-001 | NA |
| Ropinirole | 166 | ASV286 | ASV | Firmicutes | Lachnospiraceae | NA | NA |
| Ropinirole | 167 | ASV2 | ASV | Bacteroidota | Bacteroidaceae | Bacteroides | vulgatus |
| Ropinirole | 168 | ASV35 | ASV | Firmicutes | Lachnospiraceae | Agathobacter | NA |
| Ropinirole | 169 | ASV176 | ASV | Bacteroidota | Prevotellaceae | Paraprevotella | NA |
| Ropinirole | 170 | ASV107 | ASV | Bacteroidota | Bacteroidaceae | Bacteroides | eggerthii |
| Ropinirole | 171 | ASV413 | ASV | Firmicutes | Oscillospiraceae | Oscillibacter | NA |
| Ropinirole | 172 | ASV198 | ASV | Bacteroidota | Bacteroidaceae | Bacteroides | eggerthii |
| Ropinirole | 173 | ASV1030 | ASV | Firmicutes | NA | NA | NA |
| Ropinirole | 174 | ASV168 | ASV | Firmicutes | Lachnospiraceae | Lachnoclostridium | edouardi |
| Ropinirole | 175 | ASV242 | ASV | Firmicutes | Lachnospiraceae | Tyzzerella | NA |
| Ropinirole | 176 | ASV545 | ASV | Proteobacteria | Sutterellaceae | Sutterella | NA |
| Ropinirole | 177 | ASV355 | ASV | Firmicutes | NA | NA | NA |
| Ropinirole | 178 | ASV701 | ASV | Firmicutes | Lachnospiraceae | Lachnoclostridium | NA |
| Ropinirole | 179 | ASV101 | ASV | Firmicutes | Lachnospiraceae | Roseburia | NA |

| Ticker | DESeq2_baseMean | DESeq2_log2-FoldChange | DESeq2_lfcSE | DESeq2_stat | DESeq2_pvalue | DESeq2_padj | DESeq2 converged |
|---|---|---|---|---|---|---|---|
| 150 | 97,5878392 | -0,067626673 | 0,230877979 | -0,292910883 | 0,76959027 | 0,997060958 | YES |
| 151 | 13,9719109 | 0,080820908 | 0,27894594 | 0,289736813 | 0,772017591 | 0,997060958 | YES |
| 152 | 2,6309324 | 0,089107137 | 0,31145215 | 0,286102175 | 0,774799866 | 0,997060958 | YES |
| 153 | 5,538238425 | 0,085194302 | 0,300976066 | 0,283060055 | 0,777130801 | 0,997060958 | YES |
| 154 | 11,54856209 | -0,071969871 | 0,256960168 | -0,280081819 | 0,779414733 | 0,997060958 | YES |
| 155 | 139,2105032 | 0,060820752 | 0,222320739 | 0,273572103 | 0,784413475 | 0,997060958 | YES |
| 156 | 797,6950536 | 0,063128651 | 0,234249076 | 0,269493703 | 0,787549788 | 0,997060958 | YES |
| 157 | 135,3358851 | 0,058905272 | 0,222925646 | 0,264237305 | 0,791597067 | 0,997060958 | YES |
| 158 | 860,0452301 | 0,068687454 | 0,263563769 | 0,260610381 | 0,794392983 | 0,997060958 | YES |
| 159 | 216,6989455 | -0,04642825 | 0,179750167 | -0,25829322 | 0,796180618 | 0,997060958 | YES |
| 160 | 23,2449133 | -0,058661578 | 0,23470823 | -0,249934047 | 0,802638353 | 0,997060958 | YES |
| 161 | 43,04718327 | 0,065840123 | 0,268488214 | 0,245225376 | 0,806281929 | 0,997060958 | YES |
| 162 | 68,99813587 | 0,065343511 | 0,266554469 | 0,245141306 | 0,806347021 | 0,997060958 | YES |
| 163 | 7,231184968 | 0,076721806 | 0,314225049 | 0,244161967 | 0,807105381 | 0,997060958 | YES |
| 164 | 156,836539 | 0,049720331 | 0,212184844 | 0,234325553 | 0,81473225 | 0,997060958 | YES |
| 165 | 19,19879679 | -0,062661919 | 0,278790381 | -0,224763563 | 0,822163214 | 0,997060958 | YES |
| 166 | 12,3989851 | 0,06099297 | 0,280810683 | 0,217203167 | 0,828050017 | 0,997060958 | YES |
| 167 | 629,0234291 | -0,048517981 | 0,223772912 | -0,216817935 | 0,828350234 | 0,997060958 | YES |
| 168 | 532,6939736 | -0,048055166 | 0,225683809 | -0,212931384 | 0,831380483 | 0,997060958 | YES |
| 169 | 56,48736206 | -0,057261734 | 0,269595803 | -0,212398463 | 0,831796185 | 0,997060958 | YES |
| 170 | 60,78666764 | 0,06026809 | 0,286953496 | 0,210027378 | 0,833646305 | 0,997060958 | YES |
| 171 | 18,01623432 | 0,061532838 | 0,295726772 | 0,208073275 | 0,835171754 | 0,997060958 | YES |
| 172 | 202,8205626 | 0,058273131 | 0,283717679 | 0,205391257 | 0,83726645 | 0,997060958 | YES |
| 173 | 0,819166674 | -0,065436587 | 0,321326318 | -0,203645275 | 0,83863071 | 0,997060958 | YES |
| 174 | 34,48027654 | -0,043946078 | 0,215872433 | -0,203574294 | 0,838686183 | 0,997060958 | YES |
| 175 | 22,65841788 | 0,058451557 | 0,291828251 | 0,200294376 | 0,841250361 | 0,997060958 | YES |
| 176 | 28,02571102 | 0,065205272 | 0,325598173 | 0,200263017 | 0,841274885 | 0,997060958 | YES |
| 177 | 25,26908371 | 0,065201566 | 0,325598822 | 0,200251234 | 0,8412841 | 0,997060958 | YES |
| 178 | 7,902331633 | 0,065117485 | 0,325613268 | 0,199984128 | 0,841492995 | 0,997060958 | YES |
| 179 | 5,788917359 | 0,065072931 | 0,325620909 | 0,199842605 | 0,841603679 | 0,997060958 | YES |

| Medication | Ticker | ASVnr | taxLevel | Phylum | Family | Genus | Species |
|---|---|---|---|---|---|---|---|
| Ropinirole | 180 | ASV367 | ASV | Bacteroidota | Rikenellaceae | Alistipes | NA |
| Ropinirole | 181 | ASV256 | ASV | Firmicutes | Lachnospiraceae | Blautia | NA |
| Ropinirole | 182 | ASV714 | ASV | Firmicutes | Oscillospiraceae | Oscillibacter | NA |
| Ropinirole | 183 | ASV606 | ASV | Firmicutes | Lachnospiraceae | NA | NA |
| Ropinirole | 184 | ASV752 | ASV | Verrucomicrobiota | Puniceicoccaceae | NA | NA |
| Ropinirole | 185 | ASV231 | ASV | Proteobacteria | NA | NA | NA |
| Ropinirole | 186 | ASV249 | ASV | Proteobacteria | NA | NA | NA |
| Ropinirole | 187 | ASV808 | ASV | Desulfobacterota | Desulfovibrionaceae | NA | NA |
| Ropinirole | 188 | ASV277 | ASV | Bacteroidota | Rikenellaceae | Alistipes | ihumii |
| Ropinirole | 189 | ASV726 | ASV | Verrucomicrobiota | vadinBE97 | NA | NA |
| Ropinirole | 190 | ASV406 | ASV | Firmicutes | NA | NA | NA |
| Ropinirole | 191 | ASV4 | ASV | Firmicutes | Ruminococcaceae | Ruminococcus | bromii |
| Ropinirole | 192 | ASV18 | ASV | Firmicutes | Lachnospiraceae | Dorea | longicatena |
| Ropinirole | 193 | ASV135 | ASV | Bacteroidota | Rikenellaceae | Alistipes | inops |
| Ropinirole | 194 | ASV93 | ASV | Firmicutes | Oscillospiraceae | NK4A214 group | NA |
| Ropinirole | 195 | ASV885 | ASV | Firmicutes | Lachnospiraceae | Shuttleworthia | NA |
| Ropinirole | 196 | ASV347 | ASV | Bacteroidota | Tannerellaceae | Parabacteroides | NA |
| Ropinirole | 197 | ASV894 | ASV | Desulfobacterota | Desulfovibrionaceae | NA | NA |
| Ropinirole | 198 | ASV117 | ASV | Bacteroidota | Rikenellaceae | Alistipes | finegoldii |
| Ropinirole | 199 | ASV288 | ASV | Firmicutes | Erysipelotrichaceae | NA | NA |
| Ropinirole | 200 | ASV335 | ASV | Bacteroidota | Barnesiellaceae | NA | NA |
| Ropinirole | 201 | ASV305 | ASV | Firmicutes | Lachnospiraceae | Lachnospiraceae UCG-004 | NA |
| Ropinirole | 202 | ASV97 | ASV | Firmicutes | Lachnospiraceae | [Ruminococcus] gnavus group | NA |
| Ropinirole | 203 | ASV295 | ASV | Firmicutes | Ruminococcaceae | Negativibacillus | NA |
| Ropinirole | 204 | ASV626 | ASV | Firmicutes | NA | NA | NA |
| Ropinirole | 205 | ASV487 | ASV | Firmicutes | Lachnospiraceae | Howardella | ureilytica |
| Ropinirole | 206 | ASV74 | ASV | Firmicutes | Lachnospiraceae | Coprococcus | catus |
| Ropinirole | 207 | ASV656 | ASV | Firmicutes | Oscillospiraceae | Flavonifractor | NA |
| Ropinirole | 208 | ASV453 | ASV | Verrucomicrobiota | Victivallaceae | Victivallis | NA |
| Ropinirole | 209 | ASV1464 | ASV | Firmicutes | Peptococcaceae | NA | NA |

| Ticker | DESeq2_baseMean | DESeq2_log2-FoldChange | DESeq2_lfcSE | DESeq2_stat | DESeq2_pvalue | DESeq2_padj | DESeq2 converged |
|---|---|---|---|---|---|---|---|
| 180 | 5,513254628 | 0,065064574 | 0,325622335 | 0,199816066 | 0,841624436 | 0,997060958 | YES |
| 181 | 4,870041588 | 0,06504156 | 0,325626286 | 0,199742966 | 0,841681609 | 0,997060958 | YES |
| 182 | 4,318716125 | 0,065016404 | 0,325630601 | 0,199663066 | 0,841744101 | 0,997060958 | YES |
| 183 | 3,951165816 | 0,064995727 | 0,325634142 | 0,199597397 | 0,841795464 | 0,997060958 | YES |
| 184 | 3,675503085 | 0,064977575 | 0,325637259 | 0,199539743 | 0,841840558 | 0,997060958 | YES |
| 185 | 2,57285216 | 0,064866553 | 0,325656318 | 0,199187146 | 0,842116354 | 0,997060958 | YES |
| 186 | 1,837751543 | 0,064720064 | 0,325681509 | 0,19872195 | 0,842480253 | 0,997060958 | YES |
| 187 | 1,010763348 | 0,064310364 | 0,325752289 | 0,197421064 | 0,843498047 | 0,997060958 | YES |
| 188 | 0,64321304 | 0,063810003 | 0,325839399 | 0,19583268 | 0,84474113 | 0,997060958 | YES |
| 189 | 0,64321304 | 0,063810003 | 0,325839399 | 0,19583268 | 0,84474113 | 0,997060958 | YES |
| 190 | 0,275662731 | 0,062142431 | 0,326135177 | 0,190541946 | 0,848884477 | 0,997060958 | YES |
| 191 | 668,7963802 | 0,042629582 | 0,230645198 | 0,184827528 | 0,853364316 | 0,997060958 | YES |
| 192 | 700,6026635 | 0,032643964 | 0,177000116 | 0,184429055 | 0,853676878 | 0,997060958 | YES |
| 193 | 24,70129956 | 0,053149 | 0,289629618 | 0,183506784 | 0,854400396 | 0,997060958 | YES |
| 194 | 69,29626908 | 0,046794776 | 0,2596561 | 0,180218282 | 0,856981206 | 0,997060958 | YES |
| 195 | 10,13780504 | 0,053093912 | 0,301025303 | 0,176376906 | 0,859997847 | 0,997060958 | YES |
| 196 | 5,169606671 | 0,056239209 | 0,319408047 | 0,176073238 | 0,860236406 | 0,997060958 | YES |
| 197 | 4,301082976 | 0,056387137 | 0,325629425 | 0,173163519 | 0,862522898 | 0,997060958 | YES |
| 198 | 73,38945966 | 0,039029746 | 0,226646392 | 0,172205458 | 0,863276007 | 0,997060958 | YES |
| 199 | 24,06435572 | 0,049451292 | 0,289247594 | 0,170965268 | 0,864251076 | 0,997060958 | YES |
| 200 | 114,1415579 | -0,039829203 | 0,233581063 | -0,170515549 | 0,864604707 | 0,997060958 | YES |
| 201 | 23,32776262 | -0,046685407 | 0,274914798 | -0,16981773 | 0,865153483 | 0,997060958 | YES |
| 202 | 300,8338418 | 0,044181837 | 0,2684976 | 0,164552073 | 0,869296557 | 0,997060958 | YES |
| 203 | 40,62664904 | 0,04591486 | 0,285450201 | 0,160850683 | 0,872211006 | 0,997060958 | YES |
| 204 | 14,20221341 | 0,046822251 | 0,294653311 | 0,158906243 | 0,873742742 | 0,997060958 | YES |
| 205 | 12,3594249 | 0,037869374 | 0,238573773 | 0,158732343 | 0,873879755 | 0,997060958 | YES |
| 206 | 41,35040522 | -0,033914977 | 0,213993072 | -0,158486333 | 0,874073589 | 0,997060958 | YES |
| 207 | 4,333801902 | 0,050448524 | 0,324587856 | 0,155423325 | 0,876487597 | 0,997060958 | YES |
| 208 | 1,272492496 | -0,048563321 | 0,313684946 | -0,154815592 | 0,876966699 | 0,997060958 | YES |
| 209 | 3,465278207 | 0,049423413 | 0,325637231 | 0,151774453 | 0,879364832 | 0,997060958 | YES |

| Medication | Ticker | ASVnr | taxLevel | Phylum | Family | Genus | Species |
|---|---|---|---|---|---|---|---|
| Ropinirole | 210 | ASV209 | ASV | Bacteroidota | Tannerellaceae | NA | NA |
| Ropinirole | 211 | ASV287 | ASV | Firmicutes | Christensenellaceae | Christensenellaceae R-7 group | NA |
| Ropinirole | 212 | ASV63 | ASV | Actinobacteriota | Eggerthellaceae | NA | NA |
| Ropinirole | 213 | ASV206 | ASV | Firmicutes | Oscillospiraceae | Intestinimonas | NA |
| Ropinirole | 214 | ASV255 | ASV | Actinobacteriota | Coriobacteriales Incertae Sedis | NA | NA |
| Ropinirole | 215 | ASV234 | ASV | Cyanobacteria | NA | NA | NA |
| Ropinirole | 216 | ASV94 | ASV | Bacteroidota | Rikenellaceae | Alistipes | shahii |
| Ropinirole | 217 | ASV250 | ASV | Bacteroidota | Prevotellaceae | Paraprevotella | NA |
| Ropinirole | 218 | ASV363 | ASV | Bacteroidota | Rikenellaceae | Alistipes | NA |
| Ropinirole | 219 | ASV162 | ASV | Desulfobacterota | Desulfovibrionaceae | Desulfovibrio | NA |
| Ropinirole | 220 | ASV98 | ASV | Firmicutes | Acidaminococcaceae | Phascolarctobacterium | succinatutens |
| Ropinirole | 221 | ASV40 | ASV | Firmicutes | Erysipelotrichaceae | Holdemanella | NA |
| Ropinirole | 222 | ASV150 | ASV | Proteobacteria | Sutterellaceae | Sutterella | NA |
| Ropinirole | 223 | ASV86 | ASV | Firmicutes | Lachnospiraceae | [Eubacterium] hallii group | NA |
| Ropinirole | 224 | ASV217 | ASV | Firmicutes | Peptococcaceae | Peptococcus | NA |
| Ropinirole | 225 | ASV276 | ASV | Firmicutes | Oscillospiraceae | NK4A214 group | NA |
| Ropinirole | 226 | ASV151 | ASV | Firmicutes | Butyricicoccaceae | Butyricicoccus | NA |
| Ropinirole | 227 | ASV267 | ASV | Firmicutes | Lachnospiraceae | Lachnoclostridium | NA |
| Ropinirole | 228 | ASV208 | ASV | Bacteroidota | Prevotellaceae | Prevotella | NA |
| Ropinirole | 229 | ASV22 | ASV | Bacteroidota | Bacteroidaceae | Bacteroides | uniformis |
| Ropinirole | 230 | ASV530 | ASV | Firmicutes | Veillonellaceae | Allisonella | histaminiformans |
| Ropinirole | 231 | ASV264 | ASV | Firmicutes | Oscillospiraceae | Colidextribacter | NA |
| Ropinirole | 232 | ASV38 | ASV | Firmicutes | Clostridiaceae | Clostridium sensu stricto 1 | NA |
| Ropinirole | 233 | ASV820 | ASV | Firmicutes | Lachnospiraceae | GCA-900066575 | NA |
| Ropinirole | 234 | ASV164 | ASV | Bacteroidota | Tannerellaceae | Parabacteroides | distasonis |
| Ropinirole | 235 | ASV167 | ASV | Firmicutes | Oscillospiraceae | Intestinimonas | NA |
| Ropinirole | 236 | ASV240 | ASV | Bacteroidota | Prevotellaceae | Alloprevotella | NA |
| Ropinirole | 237 | ASV433 | ASV | Firmicutes | UCG-010 | NA | NA |
| Ropinirole | 238 | ASV157 | ASV | Firmicutes | Oscillospiraceae | UCG-005 | NA |
| Ropinirole | 239 | ASV71 | ASV | Firmicutes | [Eubacterium] coprostanoligenes group | NA | NA |

| Ticker | DESeq2_baseMean | DESeq2_log2-FoldChange | DESeq2_lfcSE | DESeq2_stat | DESeq2_pvalue | DESeq2_padj | DESeq2 converged |
|---|---|---|---|---|---|---|---|
| 210 | 6,584373047 | 0,04460205 | 0,295281306 | 0,151049352 | 0,879936785 | 0,997060958 | YES |
| 211 | 7,78144696 | 0,045984604 | 0,305623016 | 0,150461849 | 0,880400248 | 0,997060958 | YES |
| 212 | 56,86391424 | 0,033451537 | 0,224095271 | 0,149273732 | 0,881337642 | 0,997060958 | YES |
| 213 | 66,19255201 | 0,041792584 | 0,283232559 | 0,147555718 | 0,882693404 | 0,997060958 | YES |
| 214 | 3,097727899 | 0,047691766 | 0,325642258 | 0,146454474 | 0,883562627 | 0,997060958 | YES |
| 215 | 19,46971616 | -0,044234523 | 0,302664345 | -0,146150427 | 0,883802639 | 0,997060958 | YES |
| 216 | 85,09983947 | 0,041237955 | 0,282518077 | 0,145965722 | 0,883948448 | 0,997060958 | YES |
| 217 | 6,320062301 | 0,044311646 | 0,311043213 | 0,142461381 | 0,886715582 | 0,997060958 | YES |
| 218 | 13,10899267 | -0,040166249 | 0,28392261 | -0,141469003 | 0,887499445 | 0,997060958 | YES |
| 219 | 76,38188359 | 0,039969473 | 0,282741162 | 0,141364183 | 0,887582248 | 0,997060958 | YES |
| 220 | 31,70473749 | 0,039924188 | 0,286466642 | 0,139367668 | 0,88915962 | 0,997060958 | YES |
| 221 | 167,5637378 | 0,038019511 | 0,2812187 | 0,135195531 | 0,892457281 | 0,997060958 | YES |
| 222 | 19,36741831 | -0,031691184 | 0,236023205 | -0,134271477 | 0,893187907 | 0,997060958 | YES |
| 223 | 19,6223459 | -0,036521773 | 0,278464988 | -0,131153915 | 0,895653551 | 0,997060958 | YES |
| 224 | 133,4585594 | 0,036248631 | 0,281480133 | 0,128778647 | 0,897532802 | 0,997060958 | YES |
| 225 | 28,21300956 | 0,036861458 | 0,287177046 | 0,128357955 | 0,897865703 | 0,997060958 | YES |
| 226 | 54,57508989 | 0,034308631 | 0,267477843 | 0,128267188 | 0,897937531 | 0,997060958 | YES |
| 227 | 17,18597086 | -0,035050377 | 0,277674842 | -0,126228133 | 0,899551342 | 0,997060958 | YES |
| 228 | 60,75139152 | 0,033097648 | 0,267071986 | 0,123927817 | 0,901372428 | 0,997060958 | YES |
| 229 | 912,4786994 | -0,022510994 | 0,182497586 | -0,123349546 | 0,901830308 | 0,997060958 | YES |
| 230 | 12,95684549 | -0,02910092 | 0,243251145 | -0,119633228 | 0,904773697 | 0,997060958 | YES |
| 231 | 8,079533426 | 0,033575363 | 0,281416578 | 0,119308405 | 0,905031025 | 0,997060958 | YES |
| 232 | 8,914269441 | 0,035329972 | 0,302176524 | 0,116918322 | 0,906924779 | 0,997060958 | YES |
| 233 | 10,98360084 | 0,033257573 | 0,298135025 | 0,111552048 | 0,911178595 | 0,997060958 | YES |
| 234 | 14,18830209 | 0,032652861 | 0,294141072 | 0,111010884 | 0,911607716 | 0,997060958 | YES |
| 235 | 29,42991897 | -0,023805184 | 0,21580514 | -0,110308698 | 0,912164559 | 0,997060958 | YES |
| 236 | 404,511955 | -0,028704214 | 0,265068032 | -0,108289987 | 0,913765665 | 0,997060958 | YES |
| 237 | 8,867699966 | 0,031101982 | 0,287452664 | 0,108198621 | 0,913838139 | 0,997060958 | YES |
| 238 | 38,52111495 | 0,02330207 | 0,227335452 | 0,10250082 | 0,918359162 | 0,997060958 | YES |
| 239 | 93,82993685 | 0,027579745 | 0,28230812 | 0,097693773 | 0,922175461 | 0,997060958 | YES |

| Medication | Ticker | ASVnr | taxLevel | Phylum | Family | Genus | Species |
|---|---|---|---|---|---|---|---|
| Ropinirole | 240 | ASV1 | ASV | Actinobacteriota | Bifidobacteriaceae | Bifidobacterium | NA |
| Ropinirole | 241 | ASV140 | ASV | Bacteroidota | Tannerellaceae | Parabacteroides | distasonis |
| Ropinirole | 242 | ASV223 | ASV | Bacteroidota | Barnesiellaceae | Barnesiella | NA |
| Ropinirole | 243 | ASV470 | ASV | Firmicutes | Christensenellaceae | Christensenellaceae R-7 group | NA |
| Ropinirole | 244 | ASV45 | ASV | Firmicutes | Lachnospiraceae | Coprococcus | comes |
| Ropinirole | 245 | ASV61 | ASV | Firmicutes | Erysipelatoclostridiaceae | Catenibacterium | mitsuokai |
| Ropinirole | 246 | ASV262 | ASV | Firmicutes | Ruminococcaceae | Incertae Sedis | NA |
| Ropinirole | 247 | ASV251 | ASV | Firmicutes | Oscillospiraceae | NK4A214 group | NA |
| Ropinirole | 248 | ASV895 | ASV | Bacteroidota | Barnesiellaceae | Coprobacter | secundus |
| Ropinirole | 249 | ASV11 | ASV | Firmicutes | Ruminococcaceae | Subdoligranulum | NA |
| Ropinirole | 250 | ASV5 | ASV | Firmicutes | Lachnospiraceae | Blautia | NA |
| Ropinirole | 251 | ASV211 | ASV | Firmicutes | Lachnospiraceae | Lachnospiraceae FCS020 group | NA |
| Ropinirole | 252 | ASV324 | ASV | Proteobacteria | NA | NA | NA |
| Ropinirole | 253 | ASV454 | ASV | Actinobacteriota | Actinomycetaceae | Actinomyces | odontolyticus |
| Ropinirole | 254 | ASV341 | ASV | Firmicutes | Christensenellaceae | Christensenellaceae R-7 group | NA |
| Ropinirole | 255 | ASV535 | ASV | Firmicutes | Erysipelatoclostridiaceae | Asteroleplasma | NA |
| Ropinirole | 256 | ASV126 | ASV | Bacteroidota | Tannerellaceae | Parabacteroides | distasonis |
| Ropinirole | 257 | ASV37 | ASV | Firmicutes | Lachnospiraceae | [Eubacterium] hallii group | NA |
| Ropinirole | 258 | ASV129 | ASV | Firmicutes | Christensenellaceae | Christensenellaceae R-7 group | NA |
| Ropinirole | 259 | ASV116 | ASV | Firmicutes | Oscillospiraceae | UCG-003 | NA |
| Ropinirole | 260 | ASV23 | ASV | Firmicutes | Streptococcaceae | Streptococcus | NA |
| Ropinirole | 261 | ASV77 | ASV | Firmicutes | Lachnospiraceae | [Ruminococcus] torques group | NA |
| Ropinirole | 262 | ASV376 | ASV | Bacteroidota | Barnesiellaceae | Coprobacter | fastidiosus |
| Ropinirole | 263 | ASV813 | ASV | Firmicutes | Ruminococcaceae | NA | NA |
| Ropinirole | 264 | ASV806 | ASV | Firmicutes | Peptococcaceae | NA | NA |
| Ropinirole | 265 | ASV13 | ASV | Firmicutes | Lachnospiraceae | Fusicatenibacter | saccharivorans |
| Ropinirole | 266 | ASV310 | ASV | Firmicutes | Lachnospiraceae | NA | NA |
| Ropinirole | 267 | ASV24 | ASV | Bacteroidota | Bacteroidaceae | Bacteroides | uniformis |
| Ropinirole | 268 | ASV573 | ASV | Firmicutes | Erysipelatoclostridiaceae | Erysipelatoclostridium | NA |
| Ropinirole | 269 | ASV6 | ASV | Firmicutes | Ruminococcaceae | Faecalibacterium | NA |

| Ticker | DESeq2_baseMean | DESeq2_log2-FoldChange | DESeq2_lfcSE | DESeq2_stat | DESeq2_pvalue | DESeq2_padj | DESeq2 converged |
|---|---|---|---|---|---|---|---|
| 240 | 229,8495876 | 0,027242002 | 0,281027285 | 0,096937212 | 0,922776257 | 0,997060958 | YES |
| 241 | 84,50901957 | -0,025889666 | 0,26981357 | -0,095953906 | 0,923557182 | 0,997060958 | YES |
| 242 | 44,47130369 | 0,025665646 | 0,268433324 | 0,095612742 | 0,923828145 | 0,997060958 | YES |
| 243 | 24,64193245 | 0,026911301 | 0,288396521 | 0,093313544 | 0,925654473 | 0,997060958 | YES |
| 244 | 91,7422339 | 0,019632654 | 0,210505321 | 0,093264407 | 0,925693508 | 0,997060958 | YES |
| 245 | 164,8782222 | 0,025736845 | 0,281466831 | 0,091438287 | 0,92714434 | 0,997060958 | YES |
| 246 | 20,4025655 | 0,026498901 | 0,290069576 | 0,091353603 | 0,927211627 | 0,997060958 | YES |
| 247 | 46,98216282 | -0,020224923 | 0,226278924 | -0,0893805 | 0,92877952 | 0,997060958 | YES |
| 248 | 5,632766388 | 0,027751845 | 0,314710174 | 0,088182231 | 0,92973184 | 0,997060958 | YES |
| 249 | 245,2198831 | 0,015868132 | 0,180699031 | 0,087815257 | 0,930023511 | 0,997060958 | YES |
| 250 | 298,3218128 | -0,015595425 | 0,178987286 | -0,08713147 | 0,930567011 | 0,997060958 | YES |
| 251 | 15,84385141 | 0,018748854 | 0,223773202 | 0,083785074 | 0,933227315 | 0,997060958 | YES |
| 252 | 6,975813746 | 0,025393183 | 0,308937951 | 0,082195091 | 0,934491577 | 0,997060958 | YES |
| 253 | 1,935908323 | 0,026276758 | 0,325668465 | 0,080685609 | 0,935691982 | 0,997060958 | YES |
| 254 | 8,318861105 | 0,023760231 | 0,304207105 | 0,078105444 | 0,937744177 | 0,997060958 | YES |
| 255 | 1,105494502 | 0,024276678 | 0,312449431 | 0,077697942 | 0,938068331 | 0,997060958 | YES |
| 256 | 48,51559059 | -0,020863017 | 0,268811529 | -0,077612063 | 0,938136646 | 0,997060958 | YES |
| 257 | 40,35163701 | -0,013662128 | 0,181687368 | -0,075195805 | 0,940058922 | 0,997060958 | YES |
| 258 | 62,64125852 | 0,020706472 | 0,283938898 | 0,072925802 | 0,941865161 | 0,997060958 | YES |
| 259 | 254,1192167 | 0,015314708 | 0,211151352 | 0,072529526 | 0,942180509 | 0,997060958 | YES |
| 260 | 813,8455384 | 0,013388168 | 0,187432521 | 0,071429269 | 0,943056116 | 0,997060958 | YES |
| 261 | 157,5810915 | 0,019171896 | 0,282158605 | 0,067947232 | 0,94582764 | 0,997060958 | YES |
| 262 | 9,828050468 | 0,020008993 | 0,300885411 | 0,066500378 | 0,946979457 | 0,997060958 | YES |
| 263 | 8,342763457 | 0,01913325 | 0,304422884 | 0,062850891 | 0,949885241 | 0,997060958 | YES |
| 264 | 6,198242023 | 0,01851627 | 0,301333509 | 0,061447763 | 0,951002615 | 0,997060958 | YES |
| 265 | 436,2633886 | 0,012236093 | 0,210658558 | 0,058084957 | 0,953680957 | 0,997060958 | YES |
| 266 | 65,09911417 | -0,0153828 | 0,267403747 | -0,057526494 | 0,954125802 | 0,997060958 | YES |
| 267 | 332,30271 | 0,015077113 | 0,264157645 | 0,057076193 | 0,954484501 | 0,997060958 | YES |
| 268 | 3,362026684 | 0,018549559 | 0,325636062 | 0,056964081 | 0,954573808 | 0,997060958 | YES |
| 269 | 629,0680022 | 0,009123221 | 0,177550766 | 0,051383733 | 0,959019747 | 0,997060958 | YES |

| Medication | Ticker | ASVnr | taxLevel | Phylum | Family | Genus | Species |
|---|---|---|---|---|---|---|---|
| Ropinirole | 270 | ASV477 | ASV | Firmicutes | Oscillospiraceae | Intestinimonas | massiliensis |
| Ropinirole | 271 | ASV69 | ASV | Firmicutes | Christensenellaceae | NA | NA |
| Ropinirole | 272 | ASV332 | ASV | Firmicutes | Lachnospiraceae | [Eubacterium] ventriosum group | NA |
| Ropinirole | 273 | ASV100 | ASV | Firmicutes | Christensenellaceae | NA | NA |
| Ropinirole | 274 | ASV237 | ASV | Bacteroidota | Bacteroidaceae | Bacteroides | clarus |
| Ropinirole | 275 | ASV141 | ASV | Firmicutes | Lachnospiraceae | Lachnoclostridium | NA |
| Ropinirole | 276 | ASV338 | ASV | Firmicutes | Lachnospiraceae | Lachnotalea | NA |
| Ropinirole | 277 | ASV190 | ASV | Firmicutes | Ruminococcaceae | UBA1819 | NA |
| Ropinirole | 278 | ASV57 | ASV | Firmicutes | Lachnospiraceae | Roseburia | inulinivorans |
| Ropinirole | 279 | ASV58 | ASV | Bacteroidota | Bacteroidaceae | Bacteroides | NA |
| Ropinirole | 280 | ASV78 | ASV | Firmicutes | Lachnospiraceae | [Eubacterium] eligens group | NA |
| Ropinirole | 281 | ASV536 | ASV | Bacteroidota | Marinifilaceae | Butyricimonas | NA |
| Ropinirole | 282 | ASV494 | ASV | Firmicutes | Anaerovoracaceae | Family XIII AD3011 group | NA |
| Ropinirole | 283 | ASV819 | ASV | Firmicutes | Oscillospiraceae | Colidextribacter | NA |
| Ropinirole | 284 | ASV703 | ASV | Firmicutes | Hydrogenoanaerobacterium | NA | NA |
| Ropinirole | 285 | ASV187 | ASV | Firmicutes | NA | NA | NA |
| Ropinirole | 286 | ASV317 | ASV | Firmicutes | NA | NA | NA |
| Ropinirole | 287 | ASV499 | ASV | Firmicutes | Lachnospiraceae | Eisenbergiella | tayi |
| Ropinirole | 288 | ASV25 | ASV | Actinobacteriota | Bifidobacteriaceae | Bifidobacterium | NA |
| Ropinirole | 289 | ASV219 | ASV | Firmicutes | Lachnospiraceae | Lachnospiraceae UCG-004 | NA |
| Ropinirole | 290 | ASV295 | Genus | Firmicutes | Ruminococcaceae | Negativibacillus | NA |
| Ropinirole | 291 | ASV229 | Genus | Firmicutes | Oscillospiraceae | Flavonifractor | NA |
| Ropinirole | 292 | ASV60 | Genus | Firmicutes | Ruminococcaceae | [Eubacterium] siraeum group | NA |
| Ropinirole | 293 | ASV96 | Genus | Firmicutes | Lachnospiraceae | CAG-56 | NA |
| Ropinirole | 294 | ASV453 | Genus | Verrucomicrobiota | Victivallaceae | Victivallis | NA |
| Ropinirole | 295 | ASV226 | Genus | Firmicutes | Lachnospiraceae | Lachnospiraceae UCG-001 | NA |
| Ropinirole | 296 | ASV16 | Genus | Firmicutes | Peptostreptococcaceae | Romboutsia | NA |
| Ropinirole | 297 | ASV357 | Genus | Firmicutes | Lachnospiraceae | Marvinbryantia | NA |
| Ropinirole | 298 | ASV133 | Genus | Actinobacteriota | Eggerthellaceae | Senegalimassilia | NA |
| Ropinirole | 299 | ASV4 | Genus | Firmicutes | Ruminococcaceae | Ruminococcus | NA |

| Ticker | DESeq2_baseMean | DESeq2_log2-FoldChange | DESeq2_lfcSE | DESeq2_stat | DESeq2_pvalue | DESeq2_padj | DESeq2 converged |
|---|---|---|---|---|---|---|---|
| 270 | 44,56095926 | 0,013789605 | 0,286455406 | 0,048138751 | 0,961605663 | 0,997060958 | YES |
| 271 | 240,6254946 | 0,011607759 | 0,283313638 | 0,040971411 | 0,967318688 | 0,997060958 | YES |
| 272 | 33,71868837 | 0,010726584 | 0,270079275 | 0,039716427 | 0,968319205 | 0,997060958 | YES |
| 273 | 166,1995977 | 0,008823477 | 0,225074291 | 0,03920251 | 0,968728932 | 0,997060958 | YES |
| 274 | 147,2683699 | 0,009522614 | 0,26886544 | 0,035417767 | 0,971746617 | 0,997060958 | YES |
| 275 | 27,26601311 | 0,00793631 | 0,271622952 | 0,029218112 | 0,976690636 | 0,997060958 | YES |
| 276 | 22,220056 | -0,007879208 | 0,273654729 | -0,028792514 | 0,977030071 | 0,997060958 | YES |
| 277 | 50,89218706 | 0,00793007 | 0,288131791 | 0,027522369 | 0,978043098 | 0,997060958 | YES |
| 278 | 173,2381547 | -0,003831808 | 0,22326282 | -0,01716277 | 0,986306763 | 0,997060958 | YES |
| 279 | 184,355587 | -0,004451297 | 0,26481291 | -0,016809214 | 0,986588819 | 0,997060958 | YES |
| 280 | 20,78455532 | -0,003896979 | 0,277902615 | -0,014022824 | 0,988811772 | 0,997060958 | YES |
| 281 | 2,935307725 | 0,004145847 | 0,325642602 | 0,01273128 | 0,989842183 | 0,997060958 | YES |
| 282 | 3,33557696 | 0,004123126 | 0,325636589 | 0,012661742 | 0,989897662 | 0,997060958 | YES |
| 283 | 4,002692352 | 0,00409535 | 0,325629228 | 0,012576727 | 0,989965488 | 0,997060958 | YES |
| 284 | 4,9366539 | 0,004068957 | 0,325622252 | 0,012495943 | 0,990029939 | 0,997060958 | YES |
| 285 | 7,338269311 | 0,004031808 | 0,325612448 | 0,01238223 | 0,990120662 | 0,997060958 | YES |
| 286 | 8,405653939 | 0,004022033 | 0,325609885 | 0,012352305 | 0,990144537 | 0,997060958 | YES |
| 287 | 9,339615487 | 0,00401534 | 0,325608122 | 0,012331819 | 0,990160882 | 0,997060958 | YES |
| 288 | 578,8214631 | -0,001728086 | 0,226181171 | -0,007640272 | 0,993904004 | 0,99735506 | YES |
| 289 | 7,234308225 | 0,000137184 | 0,292851396 | 0,000468441 | 0,999626238 | 0,999626238 | YES |
| 290 | 63,70001016 | 0,490833209 | 0,178954912 | 2,742775836 | 0,006092226 | 0,670144846 | YES |
| 291 | 42,70432203 | -0,599070274 | 0,243215579 | -2,463124586 | 0,013773205 | 0,757526262 | YES |
| 292 | 587,3382827 | 0,282628232 | 0,12902055 | 2,190567573 | 0,0284831 | 0,978192336 | YES |
| 293 | 28,8579116 | -0,463719254 | 0,242792527 | -1,909940389 | 0,056140889 | 0,978192336 | YES |
| 294 | 2,113081886 | -0,518324395 | 0,318544774 | -1,627163391 | 0,103702401 | 0,978192336 | YES |
| 295 | 35,57741409 | -0,340384507 | 0,212309493 | -1,603246762 | 0,108880185 | 0,978192336 | YES |
| 296 | 54,040719 | 0,252356875 | 0,163774062 | 1,540884267 | 0,123344956 | 0,978192336 | YES |
| 297 | 22,76436955 | 0,407079908 | 0,266676463 | 1,526493578 | 0,126886989 | 0,978192336 | YES |
| 298 | 14,42728858 | 0,262446809 | 0,192293891 | 1,364821356 | 0,172309205 | 0,978192336 | YES |
| 299 | 1513,493292 | 0,160188377 | 0,118003796 | 1,357484948 | 0,174627173 | 0,978192336 | YES |

| Medication | Ticker | ASVnr | taxLevel | Phylum | Family | Genus | Species |
|---|---|---|---|---|---|---|---|
| Ropinirole | 300 | ASV192 | Genus | Bacteroidota | Marinifilaceae | Odoribacter | NA |
| Ropinirole | 301 | ASV10 | Genus | Verrucomicrobiota | Akkermansiaceae | Akkermansia | NA |
| Ropinirole | 302 | ASV111 | Genus | Firmicutes | Lachnospiraceae | Lachnospira | NA |
| Ropinirole | 303 | ASV15 | Genus | Firmicutes | Christensenellaceae | Christensenellaceae R-7 group | NA |
| Ropinirole | 304 | ASV158 | Genus | Firmicutes | Lachnospiraceae | [Eubacterium] ventriosum group | NA |
| Ropinirole | 305 | ASV27 | Genus | Firmicutes | Ruminococcaceae | CAG-352 | NA |
| Ropinirole | 306 | ASV258 | Genus | Firmicutes | Lachnospiraceae | Lachnospiraceae AC2044 group | NA |
| Ropinirole | 307 | ASV411 | Genus | Firmicutes | Lactobacillaceae | Lactobacillus | NA |
| Ropinirole | 308 | ASV53 | Genus | Firmicutes | Lachnospiraceae | [Ruminococcus] torques group | NA |
| Ropinirole | 309 | ASV75 | Genus | Firmicutes | Lachnospiraceae | [Ruminococcus] gauvreauii group | NA |
| Ropinirole | 310 | ASV19 | Genus | Firmicutes | Erysipelatoclostridiaceae | Erysipelotrichaceae UCG-003 | NA |
| Ropinirole | 311 | ASV147 | Genus | Firmicutes | Oscillospiraceae | Oscillibacter | NA |
| Ropinirole | 312 | ASV76 | Genus | Firmicutes | Lachnospiraceae | Lachnospiraceae ND3007 group | NA |
| Ropinirole | 313 | ASV17 | Genus | Actinobacteriota | Coriobacteriaceae | Collinsella | NA |
| Ropinirole | 314 | ASV134 | Genus | Actinobacteriota | Eggerthellaceae | Enterorhabdus | NA |
| Ropinirole | 315 | ASV144 | Genus | Desulfobacterota | Desulfovibrionaceae | Bilophila | NA |
| Ropinirole | 316 | ASV322 | Genus | Firmicutes | Anaerovoracaceae | Family XIII AD3011 group | NA |
| Ropinirole | 317 | ASV51 | Genus | Firmicutes | Oscillospiraceae | UCG-005 | NA |
| Ropinirole | 318 | ASV214 | Genus | Firmicutes | Lachnospiraceae | Lachnospiraceae UCG-010 | NA |
| Ropinirole | 319 | ASV85 | Genus | Firmicutes | Lachnospiraceae | Coprococcus | NA |
| Ropinirole | 320 | ASV309 | Genus | Firmicutes | Oscillospiraceae | Colidextribacter | NA |
| Ropinirole | 321 | ASV26 | Genus | Euryarchaeota | Methanobacteriaceae | Methanobrevibacter | NA |
| Ropinirole | 322 | ASV512 | Genus | Proteobacteria | Oxalobacteraceae | Oxalobacter | NA |
| Ropinirole | 323 | ASV2 | Genus | Bacteroidota | Bacteroidaceae | Bacteroides | NA |
| Ropinirole | 324 | ASV143 | Genus | Firmicutes | Peptostreptococcaceae | Terrisporobacter | NA |
| Ropinirole | 325 | ASV3 | Genus | Firmicutes | Ruminococcaceae | Subdoligranulum | NA |
| Ropinirole | 326 | ASV30 | Genus | Proteobacteria | Enterobacteriaceae | Escherichia-Shigella | NA |
| Ropinirole | 327 | ASV449 | Genus | Actinobacteriota | Atopobiaceae | Olsenella | NA |
| Ropinirole | 328 | ASV5 | Genus | Firmicutes | Lachnospiraceae | Blautia | NA |
| Ropinirole | 329 | ASV303 | Genus | Firmicutes | Lachnospiraceae | GCA-900066575 | NA |

| Ticker | DESeq2_baseMean | DESeq2_log2-FoldChange | DESeq2_lfcSE | DESeq2_stat | DESeq2_pvalue | DESeq2_padj | DESeq2 converged |
|---|---|---|---|---|---|---|---|
| 300 | 18,06683066 | -0,253208032 | 0,186992093 | -1,354110905 | 0,175700998 | 0,978192336 | YES |
| 301 | 220,900072 | 0,19347762 | 0,146609071 | 1,319683827 | 0,186940602 | 0,978192336 | YES |
| 302 | 66,9975868 | 0,213259706 | 0,171977022 | 1,240047675 | 0,214957761 | 0,978192336 | YES |
| 303 | 262,6169075 | 0,202990945 | 0,16383818 | 1,238972168 | 0,215355805 | 0,978192336 | YES |
| 304 | 391,2471127 | -0,153520941 | 0,125791898 | -1,220435849 | 0,222299695 | 0,978192336 | YES |
| 305 | 15,81206508 | 0,372528545 | 0,316212426 | 1,178095844 | 0,238758399 | 0,978192336 | YES |
| 306 | 13,77179862 | 0,369659748 | 0,316827801 | 1,166752877 | 0,243310182 | 0,978192336 | YES |
| 307 | 208,3182409 | 0,142574621 | 0,127283953 | 1,120130366 | 0,262658212 | 0,978192336 | YES |
| 308 | 276,9588331 | -0,165384267 | 0,148748439 | -1,111838676 | 0,266207519 | 0,978192336 | YES |
| 309 | 62,18513984 | 0,238238858 | 0,214617836 | 1,11006085 | 0,266972806 | 0,978192336 | YES |
| 310 | 10,2251921 | 0,338421182 | 0,306764277 | 1,103196192 | 0,269941975 | 0,978192336 | YES |
| 311 | 88,48221631 | -0,15479302 | 0,154915588 | -0,999208811 | 0,317693548 | 0,978192336 | YES |
| 312 | 38,49542381 | -0,154077938 | 0,155464836 | -0,991079024 | 0,321646995 | 0,978192336 | YES |
| 313 | 69,23958923 | -0,162349303 | 0,165112687 | -0,983263644 | 0,325477699 | 0,978192336 | YES |
| 314 | 8,715536085 | -0,258831379 | 0,265314906 | -0,975562902 | 0,329281116 | 0,978192336 | YES |
| 315 | 26,56160531 | -0,19316885 | 0,204042309 | -0,946709784 | 0,343786683 | 0,978192336 | YES |
| 316 | 38,56080584 | 0,156063675 | 0,164943987 | 0,946161656 | 0,34406614 | 0,978192336 | YES |
| 317 | 614,1585699 | 0,097136163 | 0,113804899 | 0,853532348 | 0,393364157 | 0,978192336 | YES |
| 318 | 100,7158558 | -0,162470723 | 0,192839678 | -0,842517084 | 0,399498581 | 0,978192336 | YES |
| 319 | 297,4399249 | 0,124889767 | 0,151613837 | 0,823735944 | 0,410089611 | 0,978192336 | YES |
| 320 | 62,92704697 | 0,109715987 | 0,133444874 | 0,822182101 | 0,410973264 | 0,978192336 | YES |
| 321 | 98,19313528 | -0,124579671 | 0,162618436 | -0,76608578 | 0,443625263 | 0,978192336 | YES |
| 322 | 3,869289193 | 0,219487933 | 0,319036949 | 0,687970261 | 0,491471509 | 0,978192336 | YES |
| 323 | 5783,119515 | -0,076069811 | 0,110836511 | -0,686324483 | 0,492508513 | 0,978192336 | YES |
| 324 | 12,69970671 | -0,206380743 | 0,316610616 | -0,651844039 | 0,514501788 | 0,978192336 | YES |
| 325 | 623,727588 | 0,071742653 | 0,119419792 | 0,600760153 | 0,547999748 | 0,978192336 | YES |
| 326 | 95,82908542 | -0,127529889 | 0,216948478 | -0,587834909 | 0,55664311 | 0,978192336 | YES |
| 327 | 3,760878694 | 0,186466776 | 0,319209558 | 0,584151606 | 0,559118317 | 0,978192336 | YES |
| 328 | 861,989844 | -0,070448407 | 0,121076048 | -0,581852544 | 0,560666008 | 0,978192336 | YES |
| 329 | 16,59031394 | 0,11783789 | 0,221450861 | 0,532117553 | 0,594644581 | 0,978192336 | YES |

| Medication | Ticker | ASVnr | taxLevel | Phylum | Family | Genus | Species |
|---|---|---|---|---|---|---|---|
| Ropinirole | 330 | ASV141 | Genus | Firmicutes | Lachnospiraceae | Lachnoclostridium | NA |
| Ropinirole | 331 | ASV1506 | Genus | Firmicutes | Erysipelatoclostridiaceae | UCG-004 | NA |
| Ropinirole | 332 | ASV306 | Genus | Firmicutes | Veillonellaceae | Veillonella | NA |
| Ropinirole | 333 | ASV172 | Genus | Proteobacteria | Pasteurellaceae | Haemophilus | NA |
| Ropinirole | 334 | ASV34 | Genus | Firmicutes | Lachnospiraceae | Anaerostipes | NA |
| Ropinirole | 335 | ASV98 | Genus | Firmicutes | Acidaminococcaceae | Phascolarctobacterium | NA |
| Ropinirole | 336 | ASV8 | Genus | Firmicutes | Ruminococcaceae | Faecalibacterium | NA |
| Ropinirole | 337 | ASV283 | Genus | Actinobacteriota | Eggerthellaceae | Slackia | NA |
| Ropinirole | 338 | ASV167 | Genus | Firmicutes | Oscillospiraceae | Intestinimonas | NA |
| Ropinirole | 339 | ASV12 | Genus | Firmicutes | Veillonellaceae | Dialister | NA |
| Ropinirole | 340 | ASV195 | Genus | Bacteroidota | Barnesiellaceae | Barnesiella | NA |
| Ropinirole | 341 | ASV66 | Genus | Firmicutes | Butyricicoccaceae | Butyricicoccus | NA |
| Ropinirole | 342 | ASV211 | Genus | Firmicutes | Lachnospiraceae | Lachnospiraceae FCS020 group | NA |
| Ropinirole | 343 | ASV33 | Genus | Firmicutes | Peptostreptococcaceae | Intestinibacter | NA |
| Ropinirole | 344 | ASV536 | Genus | Bacteroidota | Marinifilaceae | Butyricimonas | NA |
| Ropinirole | 345 | ASV193 | Genus | Actinobacteriota | Eggerthellaceae | Adlercreutzia | NA |
| Ropinirole | 346 | ASV18 | Genus | Firmicutes | Lachnospiraceae | Dorea | NA |
| Ropinirole | 347 | ASV666 | Genus | Firmicutes | Erysipelotrichaceae | Holdemania | NA |
| Ropinirole | 348 | ASV81 | Genus | Firmicutes | Lachnospiraceae | Lachnospiraceae NK4A136 group | NA |
| Ropinirole | 349 | ASV974 | Genus | Firmicutes | Butyricicoccaceae | UCG-009 | NA |
| Ropinirole | 350 | ASV408 | Genus | Bacteroidota | Prevotellaceae | Paraprevotella | NA |
| Ropinirole | 351 | ASV97 | Genus | Firmicutes | Lachnospiraceae | [Ruminococcus] gnavus group | NA |
| Ropinirole | 352 | ASV573 | Genus | Firmicutes | Erysipelatoclostridiaceae | Erysipelatoclostridium | NA |
| Ropinirole | 353 | ASV7 | Genus | Firmicutes | Lachnospiraceae | Agathobacter | NA |
| Ropinirole | 354 | ASV328 | Genus | Firmicutes | Anaerovoracaceae | [Eubacterium] brachy group | NA |
| Ropinirole | 355 | ASV247 | Genus | Euryarchaeota | Methanobacteriaceae | Methanosphaera | NA |
| Ropinirole | 356 | ASV28 | Genus | Firmicutes | Clostridiaceae | Clostridium sensu stricto 1 | NA |
| Ropinirole | 357 | ASV690 | Genus | Firmicutes | Ruminococcaceae | Fournierella | NA |
| Ropinirole | 358 | ASV92 | Genus | Firmicutes | Erysipelotrichaceae | Turicibacter | NA |
| Ropinirole | 359 | ASV40 | Genus | Firmicutes | Erysipelotrichaceae | Holdemanella | NA |

| Ticker | DESeq2_baseMean | DESeq2_log2-FoldChange | DESeq2_lfcSE | DESeq2_stat | DESeq2_pvalue | DESeq2_padj | DESeq2 converged |
|---|---|---|---|---|---|---|---|
| 330 | 110,8206978 | -0,065036836 | 0,122808251 | -0,529580345 | 0,596402925 | 0,978192336 | YES |
| 331 | 0,728481251 | -0,165662036 | 0,31698177 | -0,522623228 | 0,601236473 | 0,978192336 | YES |
| 332 | 130,0522936 | 0,110314504 | 0,212804841 | 0,518383435 | 0,60419077 | 0,978192336 | YES |
| 333 | 70,26092568 | 0,112073318 | 0,221879151 | 0,505109731 | 0,613481767 | 0,978192336 | YES |
| 334 | 36,53435864 | 0,081467404 | 0,166702231 | 0,488700139 | 0,625054006 | 0,978192336 | YES |
| 335 | 73,55906196 | 0,075901119 | 0,16996536 | 0,446568163 | 0,655186887 | 0,978192336 | YES |
| 336 | 1830,650072 | -0,04972186 | 0,111972396 | -0,444054619 | 0,657003095 | 0,978192336 | YES |
| 337 | 1,455174728 | -0,139066767 | 0,321109458 | -0,433082126 | 0,664955108 | 0,978192336 | YES |
| 338 | 185,3406644 | 0,049506289 | 0,114514371 | 0,432315077 | 0,665512431 | 0,978192336 | YES |
| 339 | 1025,394869 | 0,083323631 | 0,195070536 | 0,427146166 | 0,669272869 | 0,978192336 | YES |
| 340 | 167,7390849 | -0,051036665 | 0,119492891 | -0,427110472 | 0,669298866 | 0,978192336 | YES |
| 341 | 178,694455 | 0,048714918 | 0,114886608 | 0,424026081 | 0,671546793 | 0,978192336 | YES |
| 342 | 16,85277489 | 0,073777492 | 0,180873625 | 0,407895247 | 0,683350582 | 0,978192336 | YES |
| 343 | 40,5772326 | -0,091726187 | 0,226685108 | -0,404641432 | 0,685741099 | 0,978192336 | YES |
| 344 | 36,72177525 | -0,054234206 | 0,134395248 | -0,403542588 | 0,686549111 | 0,978192336 | YES |
| 345 | 7,152143617 | -0,1188896 | 0,304293977 | -0,390706386 | 0,696014278 | 0,978192336 | YES |
| 346 | 789,0232489 | 0,043186667 | 0,111911759 | 0,385899275 | 0,699571267 | 0,978192336 | YES |
| 347 | 1,058308893 | -0,123233916 | 0,321243263 | -0,383615564 | 0,701263403 | 0,978192336 | YES |
| 348 | 1188,883235 | 0,040994967 | 0,11084867 | 0,369828222 | 0,711510486 | 0,978192336 | YES |
| 349 | 16,13403659 | 0,090505818 | 0,249485008 | 0,362770567 | 0,71677628 | 0,978192336 | YES |
| 350 | 65,6993896 | -0,061749072 | 0,170242488 | -0,362712462 | 0,716819689 | 0,978192336 | YES |
| 351 | 298,3901702 | 0,073499324 | 0,20457793 | 0,359272988 | 0,719390879 | 0,978192336 | YES |
| 352 | 40,97346121 | -0,063603642 | 0,181538446 | -0,350359076 | 0,726069235 | 0,978192336 | YES |
| 353 | 1254,197603 | -0,040286349 | 0,115139767 | -0,349890834 | 0,726420626 | 0,978192336 | YES |
| 354 | 2,087173488 | -0,11163019 | 0,320265748 | -0,348554884 | 0,727423503 | 0,978192336 | YES |
| 355 | 3,205901334 | 0,107685244 | 0,311430333 | 0,345776351 | 0,729510799 | 0,978192336 | YES |
| 356 | 77,0983509 | -0,057622108 | 0,169332555 | -0,340289603 | 0,733638446 | 0,978192336 | YES |
| 357 | 2,073670166 | 0,10297538 | 0,306385071 | 0,336097905 | 0,736797038 | 0,978192336 | YES |
| 358 | 16,63640348 | 0,064930477 | 0,2076982 | 0,312619354 | 0,754569872 | 0,978192336 | YES |
| 359 | 225,0410883 | 0,06532483 | 0,221968391 | 0,294297894 | 0,768530282 | 0,978192336 | YES |

| Medication | Ticker | ASVnr | taxLevel | Phylum | Family | Genus | Species |
|---|---|---|---|---|---|---|---|
| Ropinirole | 360 | ASV242 | Genus | Firmicutes | Lachnospiraceae | Tyzzerella | NA |
| Ropinirole | 361 | ASV885 | Genus | Firmicutes | Lachnospiraceae | Shuttleworthia | NA |
| Ropinirole | 362 | ASV116 | Genus | Firmicutes | Oscillospiraceae | UCG-003 | NA |
| Ropinirole | 363 | ASV550 | Genus | Bacteroidota | Rikenellaceae | Rikenellaceae RC9 gut group | NA |
| Ropinirole | 364 | ASV630 | Genus | Firmicutes | Ruminococcaceae | Angelakisella | NA |
| Ropinirole | 365 | ASV72 | Genus | Proteobacteria | Sutterellaceae | Parasutterella | NA |
| Ropinirole | 366 | ASV162 | Genus | Desulfobacterota | Desulfovibrionaceae | Desulfovibrio | NA |
| Ropinirole | 367 | ASV31 | Genus | Bacteroidota | Rikenellaceae | Alistipes | NA |
| Ropinirole | 368 | ASV487 | Genus | Firmicutes | Lachnospiraceae | Howardella | NA |
| Ropinirole | 369 | ASV128 | Genus | Firmicutes | Oscillospiraceae | NK4A214 group | NA |
| Ropinirole | 370 | ASV13 | Genus | Firmicutes | Lachnospiraceae | Fusicatenibacter | NA |
| Ropinirole | 371 | ASV252 | Genus | Firmicutes | Erysipelotrichaceae | Solobacterium | NA |
| Ropinirole | 372 | ASV87 | Genus | Proteobacteria | Sutterellaceae | Sutterella | NA |
| Ropinirole | 373 | ASV39 | Genus | Bacteroidota | Tannerellaceae | Parabacteroides | NA |
| Ropinirole | 374 | ASV1 | Genus | Actinobacteriota | Bifidobacteriaceae | Bifidobacterium | NA |
| Ropinirole | 375 | ASV78 | Genus | Firmicutes | Lachnospiraceae | [Eubacterium] eligens group | NA |
| Ropinirole | 376 | ASV65 | Genus | Firmicutes | Ruminococcaceae | Incertae Sedis | NA |
| Ropinirole | 377 | ASV61 | Genus | Firmicutes | Erysipelatoclostridiaceae | Catenibacterium | NA |
| Ropinirole | 378 | ASV315 | Genus | Firmicutes | Anaerovoracaceae | Family XIII UCG-001 | NA |
| Ropinirole | 379 | ASV217 | Genus | Firmicutes | Peptococcaceae | Peptococcus | NA |
| Ropinirole | 380 | ASV376 | Genus | Bacteroidota | Barnesiellaceae | Coprobacter | NA |
| Ropinirole | 381 | ASV415 | Genus | Firmicutes | Oscillospiraceae | Oscillospira | NA |
| Ropinirole | 382 | ASV454 | Genus | Actinobacteriota | Actinomycetaceae | Actinomyces | NA |
| Ropinirole | 383 | ASV1722 | Genus | Firmicutes | Erysipelatoclostridiaceae | Asteroleplasma | NA |
| Ropinirole | 384 | ASV23 | Genus | Firmicutes | Streptococcaceae | Streptococcus | NA |
| Ropinirole | 385 | ASV1631 | Genus | Firmicutes | Ruminococcaceae | Paludicola | NA |
| Ropinirole | 386 | ASV240 | Genus | Bacteroidota | Prevotellaceae | Alloprevotella | NA |
| Ropinirole | 387 | ASV858 | Genus | Actinobacteriota | Atopobiaceae | Libanicoccus | NA |
| Ropinirole | 388 | ASV190 | Genus | Firmicutes | Ruminococcaceae | UBA1819 | NA |
| Ropinirole | 389 | ASV200 | Genus | Firmicutes | Lachnospiraceae | [Eubacterium] xylanophilum group | NA |

| Ticker | DESeq2_baseMean | DESeq2_log2-FoldChange | DESeq2_lfcSE | DESeq2_stat | DESeq2_pvalue | DESeq2_padj | DESeq2 converged |
|---|---|---|---|---|---|---|---|
| 360 | 19,77158324 | 0,078113848 | 0,274704353 | 0,284356061 | 0,776137528 | 0,978192336 | YES |
| 361 | 34,44927347 | 0,048988729 | 0,179040157 | 0,273618667 | 0,784377687 | 0,978192336 | YES |
| 362 | 292,1278078 | 0,038233514 | 0,143590988 | 0,266266811 | 0,790033732 | 0,978192336 | YES |
| 363 | 8,926982486 | 0,085421378 | 0,325610272 | 0,262342392 | 0,793057482 | 0,978192336 | YES |
| 364 | 5,450989306 | 0,085350387 | 0,325621711 | 0,26211516 | 0,793232659 | 0,978192336 | YES |
| 365 | 0,710998605 | 0,084196892 | 0,325809458 | 0,258423721 | 0,796079912 | 0,978192336 | YES |
| 366 | 70,36519532 | 0,058383238 | 0,232895976 | 0,250683755 | 0,802058622 | 0,978192336 | YES |
| 367 | 940,0463924 | -0,028677455 | 0,114902805 | -0,249580111 | 0,802912081 | 0,978192336 | YES |
| 368 | 11,54581472 | 0,053808099 | 0,219589936 | 0,245039001 | 0,806426233 | 0,978192336 | YES |
| 369 | 477,7791718 | 0,02639102 | 0,113683621 | 0,232144431 | 0,816425837 | 0,978192336 | YES |
| 370 | 443,5395504 | 0,032382289 | 0,142364443 | 0,227460511 | 0,820065672 | 0,978192336 | YES |
| 371 | 17,50218372 | -0,056100767 | 0,265129959 | -0,211597239 | 0,832421264 | 0,978192336 | YES |
| 372 | 62,12576903 | 0,029695345 | 0,150423775 | 0,197411247 | 0,843505729 | 0,978192336 | YES |
| 373 | 398,1486777 | -0,022751357 | 0,116903604 | -0,194616387 | 0,845693274 | 0,978192336 | YES |
| 374 | 775,0952342 | -0,030707015 | 0,158732267 | -0,193451623 | 0,84660529 | 0,978192336 | YES |
| 375 | 61,52353179 | -0,028523691 | 0,153539559 | -0,185774214 | 0,852621827 | 0,978192336 | YES |
| 376 | 132,880875 | 0,023399919 | 0,126055688 | 0,185631597 | 0,852733674 | 0,978192336 | YES |
| 377 | 153,459711 | 0,041389827 | 0,224910079 | 0,184028335 | 0,853991226 | 0,978192336 | YES |
| 378 | 17,77146951 | -0,044989646 | 0,263987787 | -0,170423209 | 0,864677321 | 0,978192336 | YES |
| 379 | 137,8624935 | -0,027033991 | 0,162431576 | -0,1664331 | 0,867816127 | 0,978192336 | YES |
| 380 | 14,48752516 | 0,040451677 | 0,288146011 | 0,140386039 | 0,888354988 | 0,978192336 | YES |
| 381 | 13,572396 | 0,040029473 | 0,292234979 | 0,136977008 | 0,891048969 | 0,978192336 | YES |
| 382 | 1,802291109 | 0,044175282 | 0,32566815 | 0,135645078 | 0,892101868 | 0,978192336 | YES |
| 383 | 1,238547867 | 0,042291051 | 0,312448064 | 0,13535386 | 0,892332103 | 0,978192336 | YES |
| 384 | 994,4670852 | 0,017126134 | 0,134113679 | 0,12769864 | 0,898387467 | 0,978192336 | YES |
| 385 | 4,983906894 | 0,039529572 | 0,325619628 | 0,121398001 | 0,9033758 | 0,978192336 | YES |
| 386 | 429,5712851 | -0,02400286 | 0,199055536 | -0,120583735 | 0,904020753 | 0,978192336 | YES |
| 387 | 13,20882232 | 0,031514952 | 0,2971797 | 0,106046787 | 0,915545231 | 0,978192336 | YES |
| 388 | 49,07126391 | 0,024696657 | 0,249219189 | 0,09909613 | 0,921061945 | 0,978192336 | YES |
| 389 | 145,1226248 | -0,010698753 | 0,12024815 | -0,088972288 | 0,929103934 | 0,978192336 | YES |

| Medication | Ticker | ASVnr | taxLevel | Phylum | Family | Genus | Species |
|---|---|---|---|---|---|---|---|
| Ropinirole | 390 | ASV219 | Genus | Firmicutes | Lachnospiraceae | Lachnospiraceae UCG-004 | NA |
| Ropinirole | 391 | ASV499 | Genus | Firmicutes | Lachnospiraceae | Eisenbergiella | NA |
| Ropinirole | 392 | ASV530 | Genus | Firmicutes | Veillonellaceae | Allisonella | NA |
| Ropinirole | 393 | ASV37 | Genus | Firmicutes | Lachnospiraceae | [Eubacterium] hallii group | NA |
| Ropinirole | 394 | ASV55 | Genus | Firmicutes | Monoglobaceae | Monoglobus | NA |
| Ropinirole | 395 | ASV29 | Genus | Firmicutes | Oscillospiraceae | UCG-002 | NA |
| Ropinirole | 396 | ASV57 | Genus | Firmicutes | Lachnospiraceae | Roseburia | NA |
| Ropinirole | 397 | ASV386 | Genus | Firmicutes | Ruminococcaceae | DTU089 | NA |
| Ropinirole | 398 | ASV20 | Genus | Bacteroidota | Prevotellaceae | Prevotella | NA |
| Ropinirole | 399 | ASV338 | Genus | Firmicutes | Lachnospiraceae | Lachnotalea | NA |
| Ropinirole | 400 | ASV12 | Family | Firmicutes | Veillonellaceae | NA | NA |
| Ropinirole | 401 | ASV10 | Family | Verrucomicrobiota | Akkermansiaceae | NA | NA |
| Ropinirole | 402 | ASV411 | Family | Firmicutes | Lactobacillaceae | NA | NA |
| Ropinirole | 403 | ASV16 | Family | Firmicutes | Peptostreptococcaceae | NA | NA |
| Ropinirole | 404 | ASV162 | Family | Desulfobacterota | Desulfovibrionaceae | NA | NA |
| Ropinirole | 405 | ASV417 | Family | Firmicutes | UCG-010 | NA | NA |
| Ropinirole | 406 | ASV2 | Family | Bacteroidota | Bacteroidaceae | NA | NA |
| Ropinirole | 407 | ASV66 | Family | Firmicutes | Butyricicoccaceae | NA | NA |
| Ropinirole | 408 | ASV17 | Family | Actinobacteriota | Coriobacteriaceae | NA | NA |
| Ropinirole | 409 | ASV192 | Family | Bacteroidota | Marinifilaceae | NA | NA |
| Ropinirole | 410 | ASV98 | Family | Firmicutes | Acidaminococcaceae | NA | NA |
| Ropinirole | 411 | ASV15 | Family | Firmicutes | Christensenellaceae | NA | NA |
| Ropinirole | 412 | ASV449 | Family | Actinobacteriota | Atopobiaceae | NA | NA |
| Ropinirole | 413 | ASV172 | Family | Proteobacteria | Pasteurellaceae | NA | NA |
| Ropinirole | 414 | ASV19 | Family | Firmicutes | Erysipelatoclostridiaceae | NA | NA |
| Ropinirole | 415 | ASV4 | Family | Firmicutes | Ruminococcaceae | NA | NA |
| Ropinirole | 416 | ASV26 | Family | Euryarchaeota | Methanobacteriaceae | NA | NA |
| Ropinirole | 417 | ASV512 | Family | Proteobacteria | Oxalobacteraceae | NA | NA |
| Ropinirole | 418 | ASV29 | Family | Firmicutes | Oscillospiraceae | NA | NA |
| Ropinirole | 419 | ASV40 | Family | Firmicutes | Erysipelotrichaceae | NA | NA |

| Ticker | DESeq2_baseMean | DESeq2_log2-FoldChange | DESeq2_lfcSE | DESeq2_stat | DESeq2_pvalue | DESeq2_padj | DESeq2 converged |
|---|---|---|---|---|---|---|---|
| 390 | 33,30020942 | 0,017743865 | 0,221898623 | 0,079963838 | 0,936266017 | 0,978192336 | YES |
| 391 | 9,102273821 | 0,024298607 | 0,325607057 | 0,074625555 | 0,940512641 | 0,978192336 | YES |
| 392 | 12,69300113 | -0,016569636 | 0,224781025 | -0,073714568 | 0,941237507 | 0,978192336 | YES |
| 393 | 61,15688542 | 0,007184145 | 0,122325838 | 0,058729578 | 0,9531675 | 0,978192336 | YES |
| 394 | 231,8641466 | -0,006125193 | 0,114925591 | -0,05329703 | 0,957495247 | 0,978192336 | YES |
| 395 | 2408,052753 | 0,005964917 | 0,112693916 | 0,052930247 | 0,957787485 | 0,978192336 | YES |
| 396 | 708,3171437 | -0,005027781 | 0,156432147 | -0,032140331 | 0,97436014 | 0,978192336 | YES |
| 397 | 19,23929244 | 0,006008007 | 0,204490725 | 0,02938034 | 0,976561253 | 0,978192336 | YES |
| 398 | 153,4665116 | 0,005877531 | 0,203445042 | 0,028890017 | 0,976952307 | 0,978192336 | YES |
| 399 | 24,05981237 | 0,006708954 | 0,24543228 | 0,027335257 | 0,978192336 | 0,978192336 | YES |
| 400 | 1398,278448 | 0,184806652 | 0,085321841 | 2,165994649 | 0,030311596 | 0,971722462 | YES |
| 401 | 178,1127135 | 0,189727202 | 0,103138993 | 1,839529321 | 0,065837369 | 0,971722462 | YES |
| 402 | 233,2572654 | 0,158376905 | 0,087015845 | 1,820092707 | 0,068744888 | 0,971722462 | YES |
| 403 | 114,2054329 | 0,212267885 | 0,133786997 | 1,586610734 | 0,112600835 | 0,971722462 | YES |
| 404 | 87,17576814 | -0,154057626 | 0,101772265 | -1,513748633 | 0,130089605 | 0,971722462 | YES |
| 405 | 135,7797637 | 0,093219587 | 0,069621635 | 1,338945673 | 0,180588359 | 0,971722462 | YES |
| 406 | 6003,305767 | -0,061368645 | 0,055271785 | -1,110306916 | 0,266866794 | 0,971722462 | YES |
| 407 | 211,8057314 | 0,062269119 | 0,059488756 | 1,046737613 | 0,295220611 | 0,971722462 | YES |
| 408 | 62,97320948 | -0,131533629 | 0,137955577 | -0,95344916 | 0,340362543 | 0,971722462 | YES |
| 409 | 51,53716572 | -0,08081837 | 0,08503162 | -0,950450781 | 0,341883252 | 0,971722462 | YES |
| 410 | 72,1926898 | 0,106595891 | 0,117513331 | 0,907096159 | 0,364355949 | 0,971722462 | YES |
| 411 | 556,4461087 | 0,094334166 | 0,104460119 | 0,903063939 | 0,366491961 | 0,971722462 | YES |
| 412 | 13,32550939 | 0,204316889 | 0,244883498 | 0,834343232 | 0,404087597 | 0,971722462 | YES |
| 413 | 77,66308689 | 0,146582406 | 0,177420918 | 0,826184468 | 0,408699458 | 0,971722462 | YES |
| 414 | 179,2542659 | 0,071978301 | 0,088655854 | 0,811884363 | 0,416857988 | 0,971722462 | YES |
| 415 | 5266,685092 | 0,042379173 | 0,054164978 | 0,782409136 | 0,433974166 | 0,971722462 | YES |
| 416 | 86,15911518 | -0,102386057 | 0,134123991 | -0,763368706 | 0,445243538 | 0,971722462 | YES |
| 417 | 3,344068027 | 0,23502458 | 0,319093192 | 0,736538999 | 0,461402743 | 0,971722462 | YES |
| 418 | 4447,37526 | 0,037096286 | 0,053497803 | 0,693416998 | 0,488047898 | 0,971722462 | YES |
| 419 | 230,3135919 | -0,058286153 | 0,088430028 | -0,659121729 | 0,509817603 | 0,971722462 | YES |

| Medication | Ticker | ASVnr | taxLevel | Phylum | Family | Genus | Species |
|---|---|---|---|---|---|---|---|
| Ropinirole | 420 | ASV30 | Family | Proteobacteria | Enterobacteriaceae | NA | NA |
| Ropinirole | 421 | ASV255 | Family | Actinobacteriota | Coriobacteriales Incertae Sedis | NA | NA |
| Ropinirole | 422 | ASV145 | Family | Bacteroidota | Muribaculaceae | NA | NA |
| Ropinirole | 423 | ASV28 | Family | Firmicutes | Clostridiaceae | NA | NA |
| Ropinirole | 424 | ASV195 | Family | Bacteroidota | Barnesiellaceae | NA | NA |
| Ropinirole | 425 | ASV63 | Family | Actinobacteriota | Eggerthellaceae | NA | NA |
| Ropinirole | 426 | ASV453 | Family | Verrucomicrobiota | Victivallaceae | NA | NA |
| Ropinirole | 427 | ASV72 | Family | Proteobacteria | Sutterellaceae | NA | NA |
| Ropinirole | 428 | ASV23 | Family | Firmicutes | Streptococcaceae | NA | NA |
| Ropinirole | 429 | ASV752 | Family | Verrucomicrobiota | Puniceicoccaceae | NA | NA |
| Ropinirole | 430 | ASV1119 | Family | Verrucomicrobiota | vadinBE97 | NA | NA |
| Ropinirole | 431 | ASV1 | Family | Actinobacteriota | Bifidobacteriaceae | NA | NA |
| Ropinirole | 432 | ASV31 | Family | Bacteroidota | Rikenellaceae | NA | NA |
| Ropinirole | 433 | ASV454 | Family | Actinobacteriota | Actinomycetaceae | NA | NA |
| Ropinirole | 434 | ASV802 | Family | Firmicutes | Ruminiclostridium | NA | NA |
| Ropinirole | 435 | ASV7 | Family | Firmicutes | Lachnospiraceae | NA | NA |
| Ropinirole | 436 | ASV55 | Family | Firmicutes | Monoglobaceae | NA | NA |
| Ropinirole | 437 | ASV703 | Family | Firmicutes | Hydrogenoanaerobacterium | NA | NA |
| Ropinirole | 438 | ASV59 | Family | Firmicutes | [Eubacterium] coprostanoligenes group | NA | NA |
| Ropinirole | 439 | ASV39 | Family | Bacteroidota | Tannerellaceae | NA | NA |
| Ropinirole | 440 | ASV322 | Family | Firmicutes | Anaerovoracaceae | NA | NA |
| Ropinirole | 441 | ASV20 | Family | Bacteroidota | Prevotellaceae | NA | NA |
| Ropinirole | 442 | ASV217 | Family | Firmicutes | Peptococcaceae | NA | NA |

| Ticker | DESeq2_baseMean | DESeq2_log2-FoldChange | DESeq2_lfcSE | DESeq2_stat | DESeq2_pvalue | DESeq2_padj | DESeq2 converged |
|---|---|---|---|---|---|---|---|
| 420 | 103,8794616 | -0,121186104 | 0,188495071 | -0,642913918 | 0,520279958 | 0,971722462 | YES |
| 421 | 36,4048039 | -0,096945596 | 0,151380736 | -0,640409067 | 0,521906689 | 0,971722462 | YES |
| 422 | 1021,270773 | 0,08692214 | 0,144350116 | 0,6021619 | 0,547066374 | 0,971722462 | YES |
| 423 | 79,13344444 | -0,054562342 | 0,115519637 | -0,472320927 | 0,636697735 | 0,971722462 | YES |
| 424 | 274,8745053 | -0,037557971 | 0,080834572 | -0,464627576 | 0,642198181 | 0,971722462 | YES |
| 425 | 89,52129243 | -0,038173918 | 0,083037215 | -0,459720594 | 0,645716785 | 0,971722462 | YES |
| 426 | 2,509140312 | -0,132944373 | 0,311206835 | -0,42718976 | 0,669241119 | 0,971722462 | YES |
| 427 | 56,93635731 | 0,043578358 | 0,123920768 | 0,351663069 | 0,72509096 | 0,971722462 | YES |
| 428 | 1041,929649 | 0,032144215 | 0,100286646 | 0,32052338 | 0,74857161 | 0,971722462 | YES |
| 429 | 2,542214907 | 0,099758393 | 0,325642468 | 0,306343316 | 0,759343262 | 0,971722462 | YES |
| 430 | 0,444887609 | 0,09839602 | 0,325866025 | 0,301952374 | 0,762688369 | 0,971722462 | YES |
| 431 | 626,50076 | -0,023971286 | 0,093853936 | -0,255410561 | 0,798406015 | 0,971722462 | YES |
| 432 | 978,8666317 | -0,01311997 | 0,068657736 | -0,191092385 | 0,848453214 | 0,971722462 | YES |
| 433 | 1,402128322 | 0,05886292 | 0,325668169 | 0,180745082 | 0,856567671 | 0,971722462 | YES |
| 434 | 12,9845155 | 0,054153868 | 0,300782467 | 0,180043301 | 0,857118574 | 0,971722462 | YES |
| 435 | 8883,985597 | 0,009405721 | 0,056784088 | 0,165640091 | 0,868440195 | 0,971722462 | YES |
| 436 | 246,1453133 | 0,008771076 | 0,065040612 | 0,13485538 | 0,892726219 | 0,971722462 | YES |
| 437 | 3,674039732 | 0,038880338 | 0,325620369 | 0,119403887 | 0,904955382 | 0,971722462 | YES |
| 438 | 242,0207145 | -0,005529785 | 0,068076707 | -0,081228741 | 0,935260043 | 0,971722462 | YES |
| 439 | 421,0129823 | -0,004768953 | 0,066353136 | -0,071872311 | 0,942703525 | 0,971722462 | YES |
| 440 | 67,10174575 | 0,004542863 | 0,078483565 | 0,057882984 | 0,953841837 | 0,971722462 | YES |
| 441 | 922,5244736 | 0,004528455 | 0,089956153 | 0,050340695 | 0,959850895 | 0,971722462 | YES |
| 442 | 123,4033056 | 0,003114271 | 0,087854472 | 0,035448061 | 0,971722462 | 0,971722462 | YES |

### Supplementary Figure 2: outlier check dose-effect relationships dopaminergic medication and gut microbiota

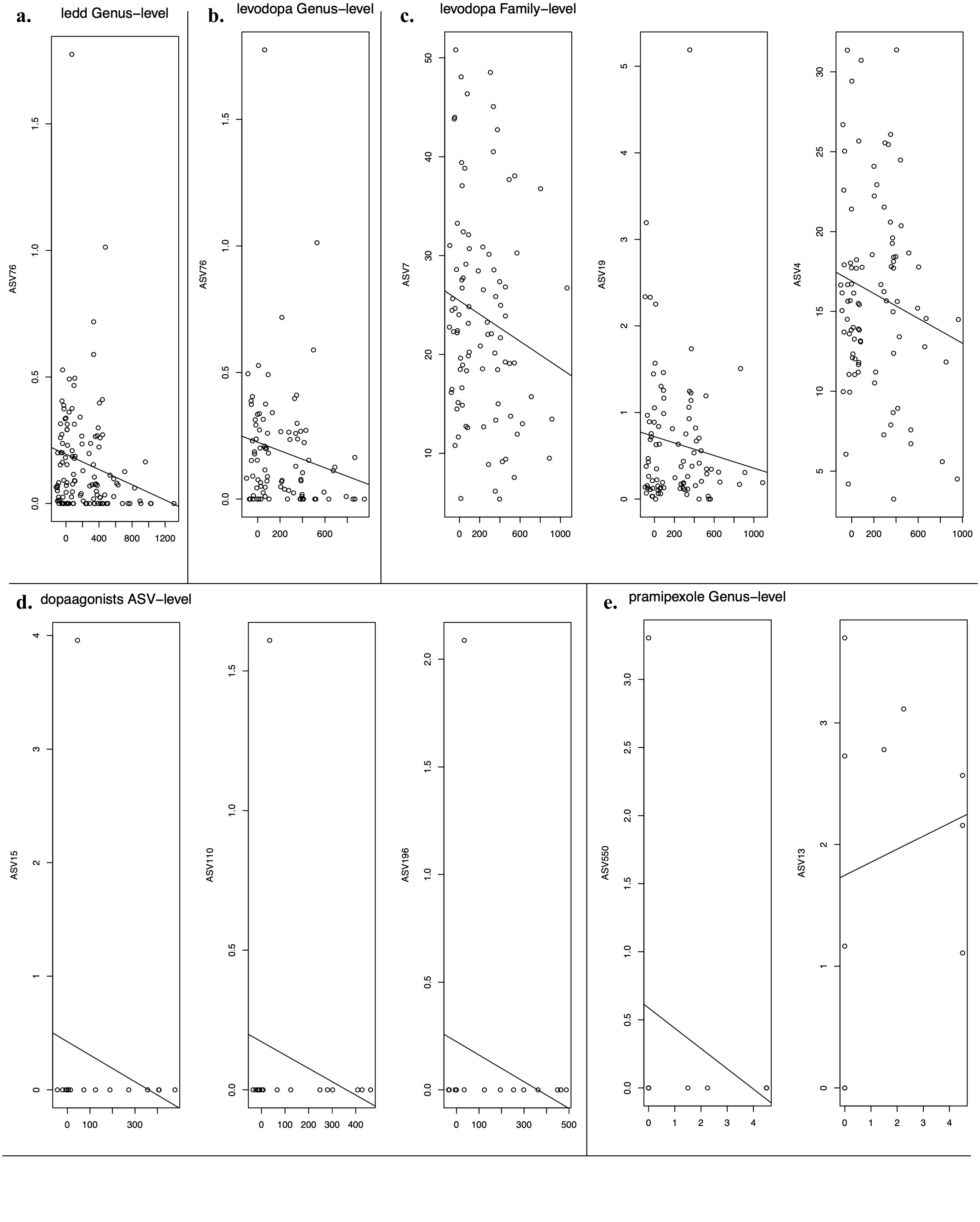

**f.**

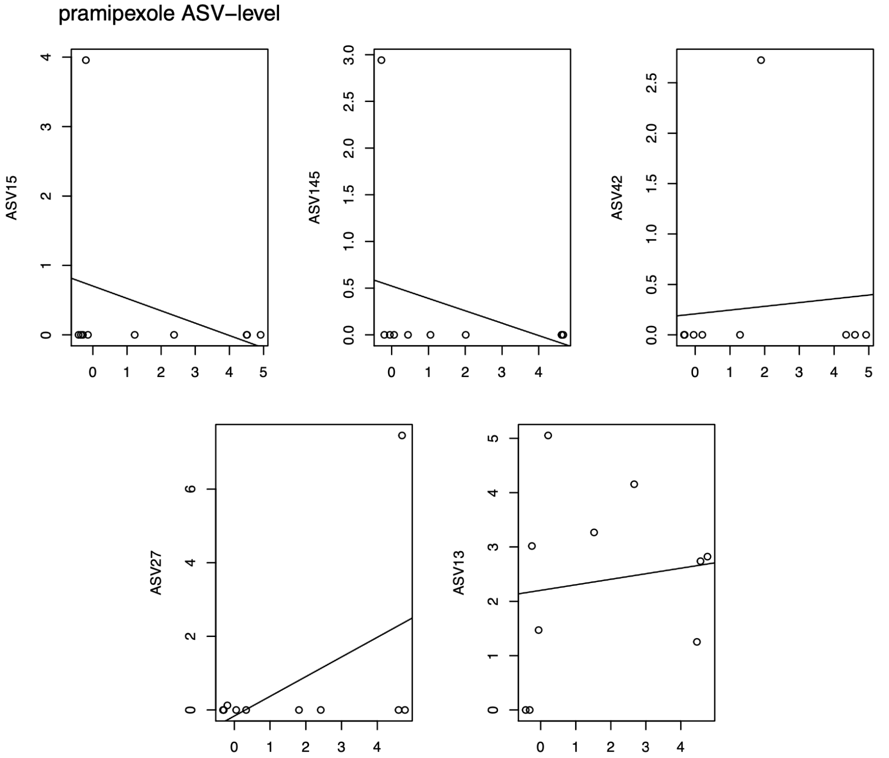


---

**g.**

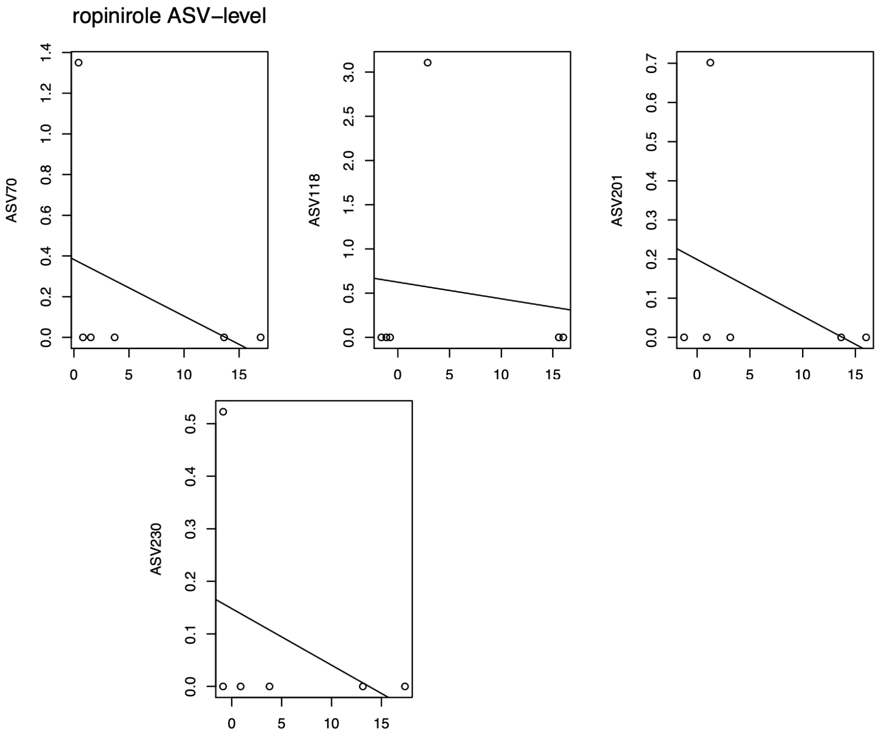


---

Associations were removed if based on the presence of a taxon in less than three biological samples. Associations that might have been driven by an outlier were re-assessed after pairwise removal of baseline and follow-up samples of the participant that contained the outlier value.

**a.** The association between ASV76 (genus-level) and LEDD remained statistically significant after outlier removal *(p*=8.1E-05; *padj*=0.01*)*. **b.** The association between ASV76 (genus-level) and levodopa dosage remained statistically significant after outlier removal *(p*=0.00025; *padj*=0.032*)*. **c.** The association between ASV19 (family-level) and levodopa dosage remained statistically significant after outlier removal *(p*=0.0014; *padj*=0.034*)*. **d.** No associations with the compound dopamine agonist dosage (LEDD) were kept in the final results. **e.** The association between ASV550 (genus-level) and pramipexol dosage was removed from the final results. **f.** The associations between ASV15, ASV145, ASV42, ASV27 (ASV-level) and pramipexole dosage were removed from the final results. **g.** No associations with ropinirole dosage were kept in the final results.
